# ‘One Health’ Genomic Surveillance of Avian and Human Influenza A Viruses Through Environmental Wastewater Monitoring

**DOI:** 10.1101/2023.08.08.23293833

**Authors:** Andrew J. Lee, Stephen Carson, Marina I. Reyne, Andrew Marshall, Daniel Moody, Danielle M. Allen, Pearce Allingham, Ashley Levickas, Arthur Fitzgerald, Stephen H. Bell, Jonathan Lock, Jonathon D. Coey, Cormac McSparron, Behnam F. Nejad, David G. Courtney, Gisli G. Einarsson, James P. McKenna, Derek J. Fairley, Tanya Curran, Jennifer M. McKinley, Deirdre F. Gilpin, Ken Lemon, John W. McGrath, Connor G. G. Bamford

**Affiliations:** School of Biological Sciences, Queen’s University Belfast, 19 Chlorine Gardens, Belfast, BT9 5DL, Northern Ireland (UK); Geography, School of Natural and Built Environment, Queen’s University Belfast, Elmwood Avenue, Belfast, BT9 6AZ, Northern Ireland (UK); Wellcome-Wolfson Institute for Experimental Medicine, School of Medicine, Dentistry and Biomedical Sciences, Queen’s University Belfast, 97 Lisburn Road, Belfast, BT9 7BL, Northern Ireland (UK); School of Pharmacy, Queen’s University Belfast, 97 Lisburn Road, Belfast, BT9 7BL, Northern Ireland (UK); Regional Virology Laboratory (RVL), Belfast Health and Social Care Trust (BHSCT), Royal Victoria Hospital (RVH), 274 Grosvenor Road, Belfast, BT12 6BA, Northern Ireland (UK); Veterinary Sciences Division, Agri-Food and Biosciences Institute (AFBI), Stormont, 12 Stoney Road, Belfast, BT4 3SD, Northern Ireland (UK); Institute for Global Food Security (IGFS), Queen’s University Belfast, 19 Chlorine Gardens, Belfast, BT9 5DL, Northern Ireland (UK)

**Keywords:** wastewater, epidemiology, surveillance, influenza, virus, avian, human, genomic, nanopore, sequencing

## Abstract

**Background:** Influenza A viruses (IAV) are significant pathogens of humans and other animals. Although endemic in humans and birds, novel IAV strains can emerge, jump species, and cause epidemics, like the latest variant of H5N1. Wastewater-based epidemiology (WBE) has very recently been shown to detect human IAV but whether it can detect avian-origin IAV, and if whole genome sequencing (WGS) can be used to discriminate circulating strains of IAV in wastewater remains unknown.

**Methods:** Using a pan-IAV RT-qPCR assay, six wastewater treatment works (WWTWs) across Northern Ireland (NI), were screened from August to December 2022. A WGS approach using Oxford Nanopore technology was employed to sequence positive samples. Phylogenetic analysis of sequences relative to currently circulating human and avian IAVs was performed.

**Findings:** We detected a dynamic IAV signal in wastewater from September 2022 onwards across NI. “Meta” whole genome sequences were generated displaying homology to both human and avian IAV strains. The relative proportion of human versus avian-origin IAV reads differed across time and sample site. A diversity in subtypes and lineages was detected (e.g. H1N1, H3N2, and several avian). Avian segment 8 related to those found in recent H5N1 clade 2.3.4.4b was identified.

**Interpretation:** WBE affords a means to monitor circulating human and avian IAV strains and provide crucial genetic information. As such WBE can provide rapid, cost-effective, year-round “one-health” IAV surveillance to help control epidemic and pandemic threats.

**Funding:** This study was funded by the Department of Health for Northern Ireland as part of the Northern Ireland Wastewater Surveillance Programme.

**Highlights:**

- Dynamic IAV RT-qPCR signal in wastewater detected across NI.
- Nanopore-based WGS reveals presence of both human and avian IAVs in wastewater.
- Avian IAV sequence similarity to gull-associated H13/H16 and recent H5N1 isolates.
- Co-detection of distinct clades of human H1N1 and H3N2 subtypes.

**Author Summary:** Influenza A virus (IAV) is a major pathogen of humans and other animals and causes regular epidemics and devastating pandemics. Recently, a novel variant of highly-pathogenic H5N1 avian influenza has emerged spreading across the world killing millions of birds and infecting mammals, enhancing its pandemic potential. Strengthening global surveillance systems for human and animal IAV is thus a major priority. Wastewater-based epidemiology (WBE) has been applied to track SARS-CoV-2 and IAV in humans but whether this approach could work for avian IAV is not known. Here, we develop a “one-health” method to survey pan-IAV levels and genetically characterise the viruses. Through this we highlight co-detection of human and avian IAVs in wastewater, with homology to recent H5N1 isolates. Our work demonstrates the potential for WBE to help defend against not only human infections but emerging, zoonotic IAVs of pandemic potential.

**Graphical Abstract:** 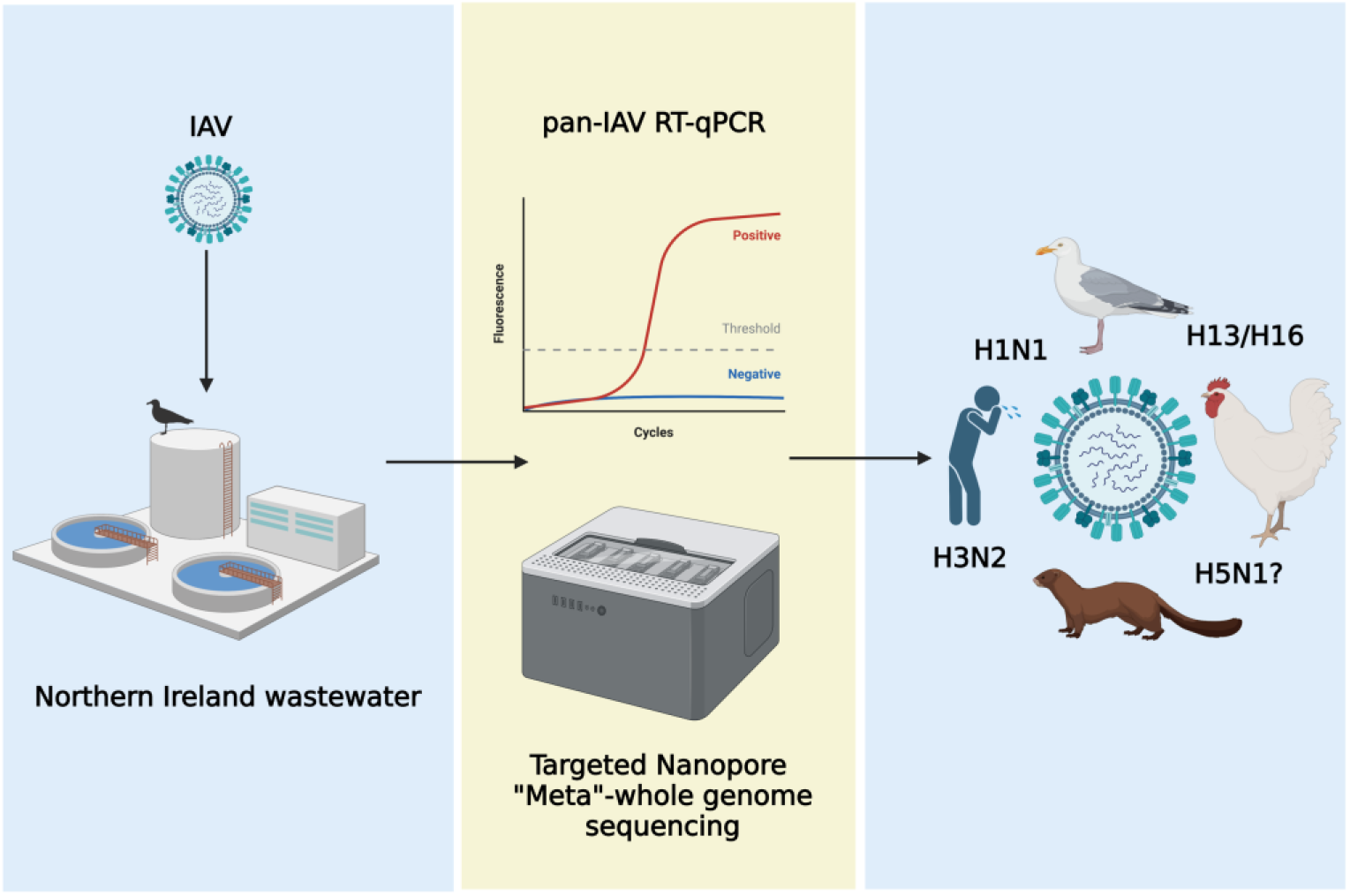

## Introduction

Influenza A virus (IAV) (Family *Orthomyxoviridae*; Genus *Influenzavirus*) is an enveloped virus with a genome comprising 8 segments (named 1-8) of single-stranded negative sense RNA (Krammer et al., 2018). The IAV genome encodes at least 10 proteins, including the hemagglutinin (HA) and neuraminidase (NA) glycoproteins, and the innate immune antagonist non-structural protein 1 (NS1). IAV is a major pathogen of humans, livestock, and wildlife, and is the causative agent of several pandemics over the last century following zoonosis from animal reservoirs like pigs and birds. Clinical burden in humans and animals is extensive (1,2,3) but can be managed through prophylactic vaccination focused towards circulating HA and NA sequences (4).

IAV has many subtypes and although some are endemic in humans (e.g., H1N1 and H3N2) and other mammals, wild birds belonging to the orders Anseriformes (e.g., ducks) and Charadriiformes (e.g., gulls [5]) are widely acknowledged as the major IAV genetic reservoirs. Most IAVs are considered low pathogenicity avian influenza viruses (LPAIVs) e.g., H13Nx and H16Nx that infect gulls (6,7,8). Others (e.g., H5 and H7) can be high pathogenicity avian influenza viruses (HPAIVs) due to the acquisition of a “multibasic cleavage sequence” in HA (9). A novel strain of H5Nx (clade 2.3.4.4b) HPAIV was first detected in 2016 in Eurasia (10). This variant spread globally killing over 50 million birds and infecting wild and farmed mammal species. Additional evidence suggesting mammal-to-mammal transmission (e.g., mink) increases the pandemic potential of such circulating HPAIVs, emphasising the need for heightened, active surveillance in both humans and animals (11,12).

Current human IAV monitoring usually entails clinical surveillance via primary care (sentinel GP services and in-hospital testing) with onward analysis of positive samples by PCR and sequencing. Avian IAV surveillance incorporates active (capture and swabbing of live birds) and passive (dead birds) monitoring. Environmental sampling (e.g., faeces or mud) has been explored but has not played a significant role in surveillance efforts globally (13). Complementing established clinical surveillance methods, WBE has recently proven an effective approach to track levels and identify variants of SARS-CoV-2 circulating in the human population (14,15,16). Likewise, this method has proven useful for the monitoring of enteric pathogens like adenoviruses (17), and other viruses such as respiratory syncytial virus (18,19). WBE studies have also demonstrated the detection of human IAV and influenza B virus (IBV) in wastewater samples, using targeted PCR but with limited sequencing (20,21,22,23,24,25,26).

Whether WBE surveillance can be repurposed as a single sampling approach to monitor both human and animal viral reservoirs and thus help identify and characterise pathogens such as avian influenza with zoonotic potential, remains unknown. Here, we describe an integrated pan-IAV genomic WBE surveillance approach, using an RT-qPCR assay and nanopore WGS to rapidly sequence and subtype influenza positive samples detected and distinguish between human and avian sequences.

## Methods

### Sample Collection and Processing

Composite (24 hour) wastewater samples comprising primary untreated influent were collected using an Isco Glacier autosampler (Isco; Lincoln, USA) from 6 WWTW covering key population centres from each health trust of NI. Sampling was carried out by Northern Ireland Water Ltd and the Northern Ireland Environment Agency (NIEA). Once received, samples were stored at 4°C before pre-processing the same day. Sample processing, viral concentration, and extraction was carried out as previously described (17).

### Wastewater Screening for IAV - Matrix Protein (MP) segment RT-qPCR

Screening of wastewater for the presence of IAV employed the SVIP-MPv2 assay (27). Universal IAV detection was confirmed against avian (H6N1), human (H1N1 and H3N2) and swine (pH1N1 and H1N2) nucleic acid extracts prepared in-house from inactivated virus stocks supplied by the Agri-Food and Biosciences Institute (AFBI, Northern Ireland) and QUB. Raw wastewater samples spiked with cultured human H1N1 virus, inactivated using Triton X (1% w/v), were prepared. These were processed to determine the SVIP-MPv2 assays suitability for use against wastewater, to identify any sample matrix effects and to determine if processing methods were applicable for the recovery of IAV. Replicate PCR reactions (comprising 2x 6µl and 2x 3µl purified nucleic acid template volumes) per wastewater sample were performed on a LightCycler 480 II Real-Time PCR System (Roche Diagnostic Limited). RT-PCR was carried out in a reaction volume of 25µL using AgPath-ID™ One-Step Reagents (ThermoFisher Scientific) with primers and probe at a final concentration of 1.4µM and 0.4µM respectively, and supplemented with bovine serum albumin (0.2mg/ml, ThermoFisher Scientific) with the following thermo-profile: 50°C for 10mins, 95°C for 10mins followed by 45 cycles of 95°C for 10s and 60°C for 30s (Table S1 and S2). A standard curve and limit of detection (LOD) were calculated for the SVIP-MPv2 assay (Figure S1) using synthetic RNA respiratory virus controls H1N1 and H3N2 (103001, 103002, Twist Bioscience) serially diluted in 0.1X TE buffer (ThermoFisher Scientific) supplemented with a blocking nucleic acid (0.1µg/µl yeast tRNA, Sigma-Aldrich). IAV MP segment concentrations were normalised using wastewater flow rate and expressed as gene copies (g.c.) per 100,000 population equivalents (p.e.) per day as previously described (Reyne et al., 2022). Clinical data by epidemiological week covering molecular tests carried out for IAV was supplied by the Regional Virus Laboratory (RVL), Belfast Health and Social Care Trust (BHSCT), NI.

### Amplicon Generation for Sequencing - Multi-segment RT-PCR Approach (M-RTPCR)

A M-RTPCR approach to simultaneously amplify all eight segments of the IAV genome from IAV MP positive wastewater samples within a single PCR reaction was employed (28). Only those IAV positive WW samples with a Ct value of ≤37 were selected for amplicon generation. One step RT-PCR was carried out using SuperScript III One-Step RT-PCR reagents (ThermoFisher Scientific) with final concentrations of primers MBTuni-12 and MBTuni-13 (0.2µM each [Table S1]) in a total volume of 50µl (40µl reaction mix and 10µl of purified nucleic acid). Reactions were run in duplicate on a C1000 Touch thermal cycler (Biorad) using a 96 deep-well reaction module, following thermal cycling conditions as previously described (Table S2). Duplicate reaction mixtures were pooled, and amplified DNA purified using a 0.8X ratio of KAPA Pure Beads (Roche Diagnostic Limited). Quantification was carried out using a Qubit high-sensitivity double-stranded DNA kit (Qubit Flex, ThermoFisher Scientific), and amplicons analysed using D5000 ScreenTape assays (TapeStation 4200 System, Agilent Technologies) both according to the manufacturers’ instructions (Figure S2). As an additional approach to further quantify generated amplicons prior to library preparation, 2 µl of purified amplicon mix was re-tested using the SVIP-MPv2 assay. A significant decrease in Ct value versus original screening Ct value was concomitant with the presence of increased amounts of matrix protein (segment 7) amplicons signifying successful genomic segment amplification (Table S3). To monitor for potential operator and environmental contamination as well as background nucleic acid present in reagents and consumables, negative process controls consisting of DEPC-treated nuclease free water (ThermoFisher Scientific) were included alongside and treated in the same manner.

### Nanopore Library Preparation and Sequencing

Samples were prepared for sequencing following the suggested protocol for ligation sequencing of gDNA with the SQK-LSK109 kit (Oxford Nanopore Technologies). To mitigate any problems of cross-barcode contamination samples were not multiplexed, instead a single sample per FLO-MIN106D R9 flow cell was sequenced on a GridION device (Oxford Nanopore Technologies) with sequencing proceeding for 24 hours. Negative process controls were also sequenced in the same way. Flow cells were only used once and not reused for subsequent IAV sequencing runs.

### Genomic Analysis

Nanopore reads were base called using Guppy (version 6.4.2). Reads were taxonomically classified using Centrifuge (29). A total IAV read number of greater than 20 was arbitrarily chosen as a cut-off for downstream analysis. Identified IAV reads were mapped using Minimap2 (30). For each sample, reference genomes encompassing potential originating viruses were utilised for mapping: human H1N1 (A/California/07/2009), H3N2 (A/Barry/3792/2022; EPI_ISL_15391170), and avian H5N1 (A/Mute_Swan/Netherlands/2/2022; EPI_ESL_15364797) and H13N6 (A/black0headed gull/Netherlands/31/2014). Samtools was used for coverage calculations and BCFtools to generate consensus sequences (31). Alignment length and percentage coverage of raw reads was investigated using Basic Local Alignment Search Tool (BLAST+, version 2.13.0) against NCBI’s available precompiled nucleotide database (32). Generated raw sequences are in the process of being submitted but are available on request.

### Phylogenetic Analyses

Generated IAV consensus sequences were used for phylogenetic analysis. Consensus sequence incorporation differed depending on the analysis used with only homogenous consensus sequences used for interrogation of databases and phylogenetics of avian segments 5, 7 and 8, while all consensus sequences available were used for human segment 4 and 6 analysis. For human segment 4 and 6 analysis, Nextclade was used to remove low quality consensus sequences. Similarity to known sequences in the GISAID database was determined using their integrated BLAST protocol. All gull H13/16-like sequences since 2013, and H5N1 European sequences (2022) were used (downloaded January 2023). Select human H1N1 and H3N2 segment 4 and 6 sequences were used, including available recent sequences from Northern Ireland. Multiple sequence alignments were performed in MAFFT [https://mafft.cbrc.jp/], and maximum likelihood phylogenies inferred (1000 ultrafast bootstrap replicates) using IQ-TREE [http://iqtree.cibiv.univie.ac.at/] (33). The best substitution model was identified using IQ-TREE, ranked by BIC and implemented. Trees were visualised using FigTree v1.4.4 [http://tree.bio.ed.ac.uk/software/figtree/]. Table S4 provides metadata on the GISAID genome sequences used in this study.

## Results

### Pan-IAV wastewater screening by RT-qPCR

144 wastewater samples, from 6 WWTW across NI (**Figure 1 A**), were screened for IAV between 1^st^ August 2022 and 5^th^ December 2022 (epi weeks 31 - 49) using our pan-IAV assay. A consistent IAV signal above the LOD was first detected from the 5^th^ of September 2022 onwards (epi week 36), corresponding to an average weekly concentration of 5.03 x 10^12^ g.c. per 100k p.e. across the WWTW screened (**Figure 1 B**). IAV positivity and concentrations in wastewater varied across the sites sampled during the period investigated, with three peaks observed during weeks 37, 42 and 48, with an average of 5.74 x 10^12^, 8.67 x 10^12^, and 1.13 x 10^13^ g.c. per 100k p.e. respectively. Clinical data by epi week covering molecular tests carried out for IAV was supplied by the Regional Virus Laboratory, Royal Victoria Hospital. Clinical percentage positivity remained below 1% until week 35 (1.39%) and did not sustain an upwards trajectory until week 42 (1.44%), steadily increasing to 3.99% during week 49 before rising markedly from week 50 onwards (6.42%) (**Figure 1 B**). The peak of the clinical influenza 2022/23 season was during week 52 (14.42% [data not shown]) outside of our study period.

**Fig 1.**
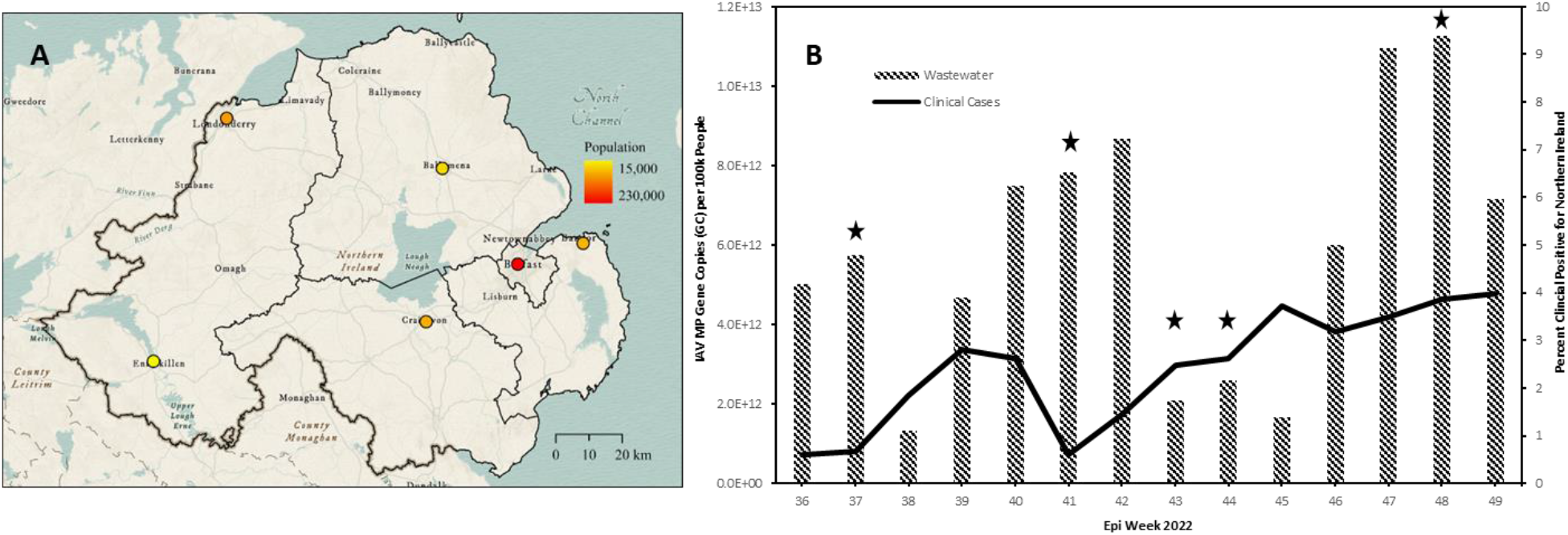
Dynamic IAV wastewater signal across NI. Map showing geographical locations of WWTW across NI screened for IAV (A). West-to-East: Enniskillen, Derry/Londonderry, Craigavon, Ballymena, Belfast, and North Down. Normalised and averaged levels of IAV MP gene copies detected (g.c. per 100k p.e.) per epi week across the 6 WWTW sampled (B). Clinical data (black line) represents percentage IAV positive PCR and point of care tests carried out by the Regional Virus Laboratory for NI. ★ indicates epi weeks where IAV sequence was obtained.

### Meta-whole genome sequencing of human and avian IAV from wastewater

The SVIP-MPv2 assay targets a highly conserved region of the IAV MP segment regardless of subtype and host. To resolve the genomic composition of detected IAV samples, a whole-genome amplification approach was employed. Overall, 12 of 36 IAV positive (<Ct 38) wastewater samples were successfully processed and sequenced (>20 IAV reads) following the full-segment amplicon generation method adopted (Zhou et al., 2009). IAV sequence was generated from samples collected on 12/09/2022, 10/10/2022, 24/10/2022, 31/10/2022, and 28/11/2022 (epi weeks 37-48 inclusive) covering a range of geographical locations (Fig 1A): West-to-East: Enniskillen (ENN), Craigavon (CRG), Ballymena (BYM), Belfast (BEL), and North Down (NDN). No sequence data was obtained for Derry/Londonderry.

Identified IAV reads in these IAV RT-qPCR-positive samples ranged from 65 to 1,440, with 7,044 sequences detected overall. Reads were generally biased towards the smaller segments (7 and 8) (**Table 1**). No single sample produced reads for every segment. However, across the samples successfully sequenced, reads mapping to all 8 segments were generated and referred to here as “meta”-WGS” to reflect sequencing all segments and to distinguish from true WGS from clinical samples containing single strains. On average, percentage coverage of segments varied considerably from 25.7% (PB1) up to 99.6% (NP). Sequences mapping to segments 4 and 6, encoding HA and NA, were generated for 9 samples (75%). Average read number and coverage was 203/29.5% and 10.2/57.9% for HA and NA segments respectively.

**Table 1.** Description of IAV WGS in wastewater. Read number and average percentage coverage per segment, per sample. Identity and alignment length of raw reads investigated using Basic Local Alignment Search Tool against NCBI’s available precompiled nucleotide database.

| WwTW | Sample Date | Segment 1 PB2 (2.3kb) |  | Segment 2 PB1 (2.3kb) |  | Segment 3 PA (2.2kb) |  | Segment 4 HA (1.8kb) |  | Segment 5 NP (1.6kb) |  | Segment 6 NA (1.4kb) |  | Segment 7 MP (1.0kb) |  | Segment 8 NS (0.9kb) |  | Total Reads per Sample |
| --- | --- | --- | --- | --- | --- | --- | --- | --- | --- | --- | --- | --- | --- | --- | --- | --- | --- | --- |
|  |  | Reads | % Cover | Reads | % Cover | Reads | % Cover | Reads | % Cover | Reads | % Cover | Reads | % Cover | Reads | % Cover | Reads | % Cover |  |
| Ballymena | 12/09/2022 | 3 | 50.5 | 0 | 0 | 0 | 0 | 0 | 0 | 25 | 32.3 | 0 | 0 | 67 | 90.7 | 74 | 84.9 | 169 |
| Belfast | 12/09/2022 | 2 | 58 | 0 | 0 | 0 | 0 | 6 | 76.5 | 26 | 87 | 1 | 96.8 | 216 | 91.7 | 326 | 89.9 | 577 |
| Craigavon | 12/09/2022 | 2 | 91.6 | 35 | 25.7 | 0 | 0 | 112 | 33.3 | 13 | 88.2 | 4 | 76.4 | 277 | 92.7 | 167 | 91.1 | 610 |
| Enniskillen | 12/09/2022 | 0 | 0 | 0 | 0 | 0 | 0 | 0 | 0 | 18 | 78.2 | 31 | 95.7 | 347 | 96.1 | 228 | 93.8 | 624 |
| North Down | 12/09/2022 | 0 | 0 | 0 | 0 | 0 | 0 | 3 | 66 | 14 | 82.6 | 3 | 96 | 120 | 89.2 | 508 | 90.1 | 648 |
| Belfast | 10/10/2022 | 0 | 0 | 0 | 0 | 0 | 0 | 0 | 0 | 9 | 48.4 | 27 | 58.3 | 47 | 77.7 | 48 | 76.8 | 131 |
| North Down | 10/10/2022 | 0 | 0 | 0 | 0 | 0 | 0 | 0 | 0 | 5 | 86.7 | 0 | 0 | 60 | 88.7 | 0 | 0 | 65 |
| Belfast | 24/10/2022 | 0 | 0 | 0 | 0 | 0 | 0 | 0 | 0 | 0 | 0 | 12 | 87.1 | 507 | 96.7 | 676 | 92.1 | 1195 |
| Belfast | 31/10/2022 | 0 | 0 | 0 | 0 | 0 | 0 | 9 | 60.4 | 1 | 99.6 | 26 | 89.4 | 98 | 93.9 | 1306 | 90.4 | 1440 |
| North Down | 31/10/2022 | 0 | 0 | 0 | 0 | 0 | 0 | 0 | 0 | 0 | 0 | 0 | 0 | 96 | 93.4 | 0 | 0 | 96 |
| Ballymena | 28/11/2022 | 3 | 39.8 | 0 | 0 | 2 | 68.9 | 67 | 77.1 | 85 | 88.3 | 18 | 95.1 | 279 | 89.4 | 364 | 91.3 | 818 |
| Belfast | 28/11/2022 | 0 | 0 | 0 | 0 | 0 | 0 | 191 | 40.2 | 12 | 87.6 | 0 | 0 | 186 | 95.4 | 282 | 91.2 | 671 |
| Per Segment Total Reads & Average % Coverage |  | 10 | 60 | 35 | 25.7 | 2 | 68.9 | 388 | 65.6 | 208 | 86.7 | 140 | 86.9 | 2,300 | 91.3 | 3,979 | 89.2 |  |

Initial read interrogation using a taxonomic classification first approach suggested the presence of diverse IAV strains, possibly consisting of both human-like and avian-like viruses (**Figure 2 A and B**). The relative abundance of these differed by sampling time and site. At first heterogenous mixes predominated, but the relative abundance of human-like IAV reads increased as the clinical influenza season commenced for Belfast and Ballymena (**Figure 2 C**). However, North Down showed a more stable mix of human and avian-like sequences. Furthermore, we generated consensus sequences by mapping, focussing our downstream analysis on sequences where the consensus generated by each reference was similar to each other. This resulted in 25 consensus sequences spanning all IAV segments (described in **Table 2 and found in supplement).** Derived consensus sequences were compared to known IAV sequences by BLAST, revealing diverse origins corroborating the original analysis by Centrifuge, including human subtypes (H1N1 and H3N2) but also avian subtypes, which are predominantly circulating in gull populations (H13N6), and even H5N1 clade 2.3.4.4b (Belfast 10/10/2022) from a Eurasian curlew (a member of the Charadriiformes) (**Table 2**). Similar to the Centrifuge analysis, we detect here that the proportion of avian-like sequences compared to human-like decreases at later sampling dates.

**Fig 2.**
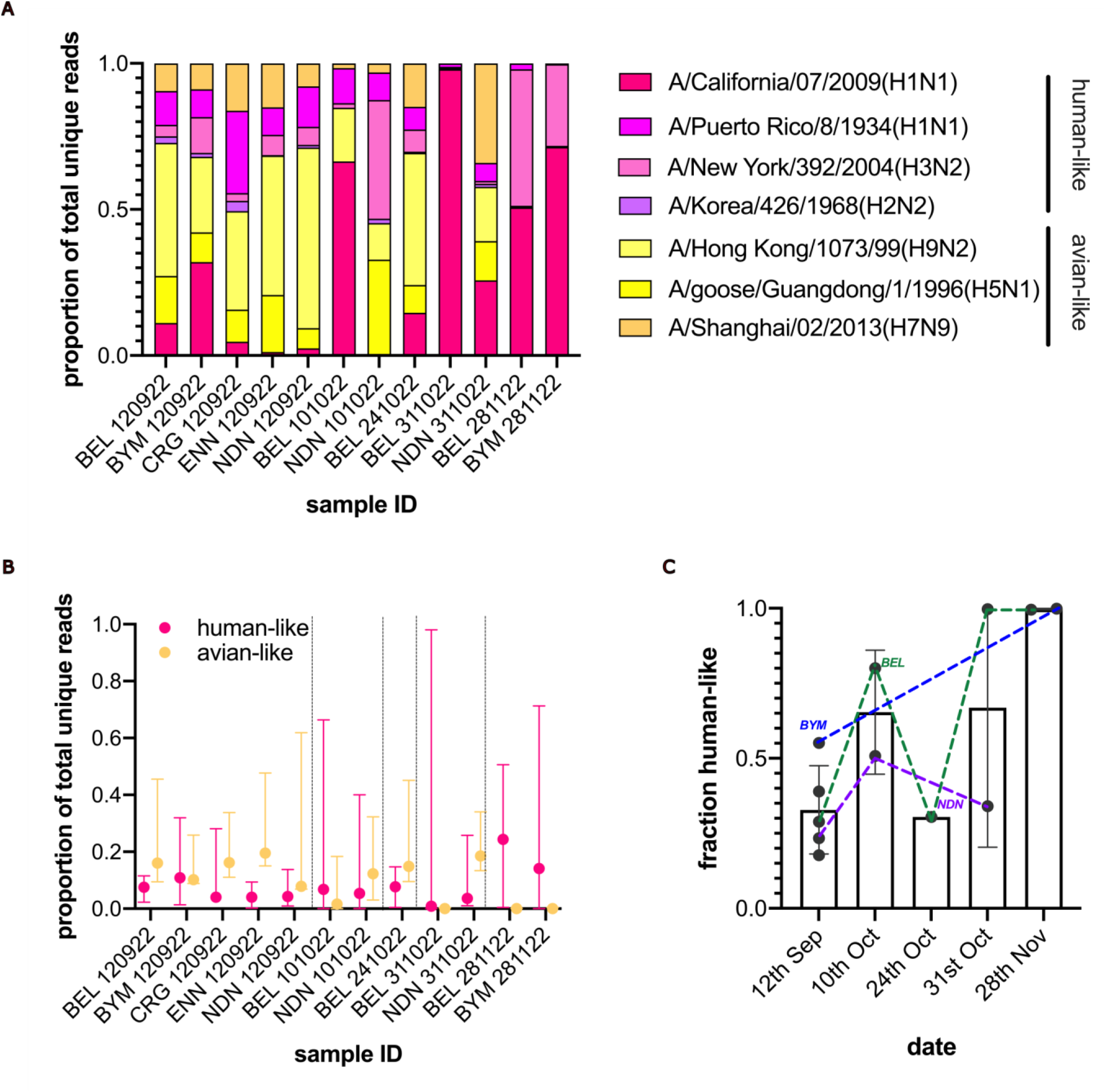
Taxonomic classification-first interrogation of IAV reads from wastewater. Proportion unique reads from samples (site: BEL = Belfast; BYM = Ballymena; CRG = Craigavon; NDN = North Down) were assigned a similarity to 1 of 7 IAV strains (A/California/07/2009(H1N1); A/Puerto Rico/8/1934(H1N1); A/New York/392/2004(H3N2); A/Korea/426/1968(H2N2); A/Hong Kong/1073/99(H9N2); A/goose/Guangdong/1/1996(H5N1); A/Shanghai/02/2013(H7N9)) in the Centrifuge database. These strains were grouped into human-like (H1N1, H3N2 and H2N2; purple shades) and avian-like (H9N2, H5N1 and H7N9; yellow shades) (A). Data from A shown aggregated by human-like and avian-like for each sample with median and range illustrated (B). Fractional proportion of aggregate human-like sequences shown sample date only for all sites with links for those for which multidate data is available (BEL in green, BYM in blue and NDN in purple) are shown (C). Graphs generated in Graphpad Prism v9.0.

**Table 2:**
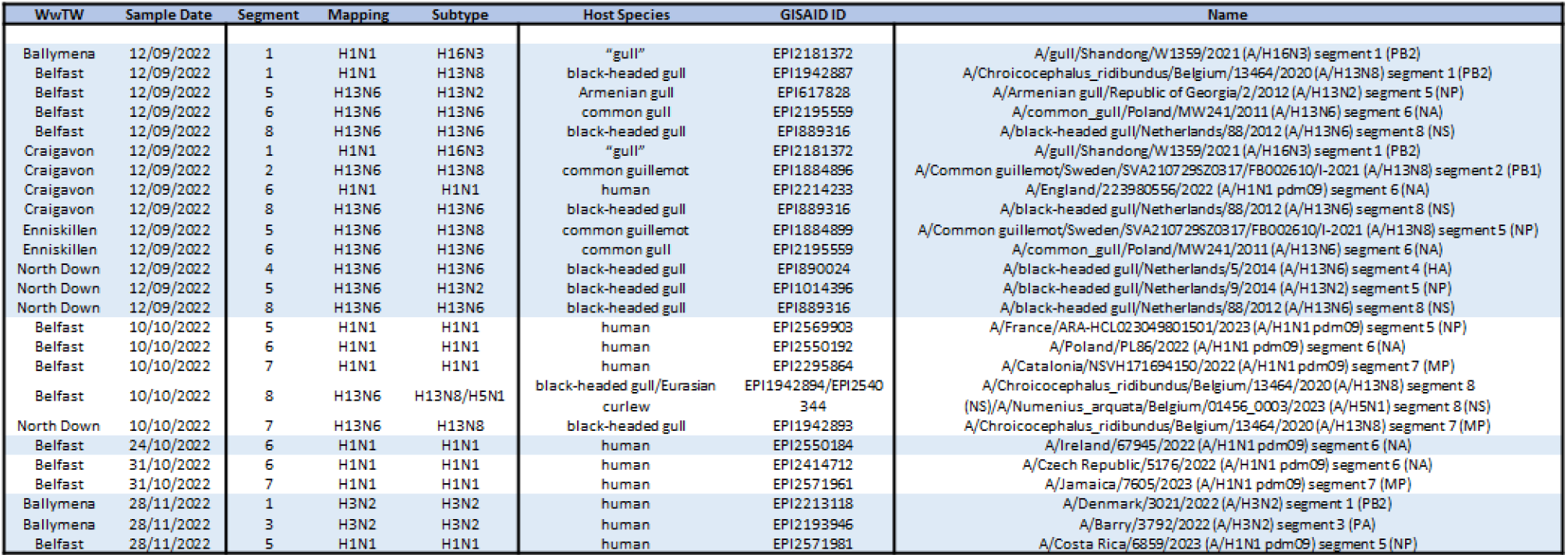
Similarity of wastewater IAV consensus sequences to known sequences. Table describing BLAST (GISAID) results for consensus sequences from samples sites and dates. Sequences spanned all segments and interrogation of GISAID database allowed low resolution sub-typing by identifying known subtype of the top-ranked similarity hit. For each top hit, the subtype, host species, GISAID ID and name for each sequence is shown. For Belfast 10/10/2022 two hits are shown, including similarity to H5N1.

### Detection of diverse lineages of gull-associated IAV in wastewater

To explore the avian IAV sequences further, we determined the phylogenetic relatedness of our derived consensus sequences relative to the two gull-associated sub-types (H13 and H16 Nx) as well as recent (2022) H5N1 clade 2.3.4.4b viruses found across Europe. Phylogenetic analysis of segment 8 (encoding the virulence factor NS1, and the essential NEP) confirmed clear avian association but disclosed two distinct clades found in our samples (**Figure 3**). One clade (gull 2; sequences found on 12-09-22) was closely related to - but distinct from - H16Nx viruses identified in Europe in 2013/2015 (Netherlands). One of these clades (gull 3; marked by Belfast 10-10-23) was more closely related to recent European H5N1 viruses (clade 2.3.4.4b), which include viruses sequenced from birds and mammals (mink). However, phylogenetically our H5N1-related sequence sat at a basal position to the H5N1 sub-clade along with other gull-associated sequences (found in Europe and China in 2020/21), suggesting that it is unlikely to be from an H5N1 viruses but from the ancestor of the lineage that donated its segment 8 to H5N1 clade 2.3.4.4b. No other H5-like sequences were detected. Interrogation of the three segment 5 sequences and the single segment 7 sequence, identified similarity to sequences from Europe (2020/2021), China (2021) and Alaska (2020) (Supplementary Figures 3 and 4).

**Figure 3.**
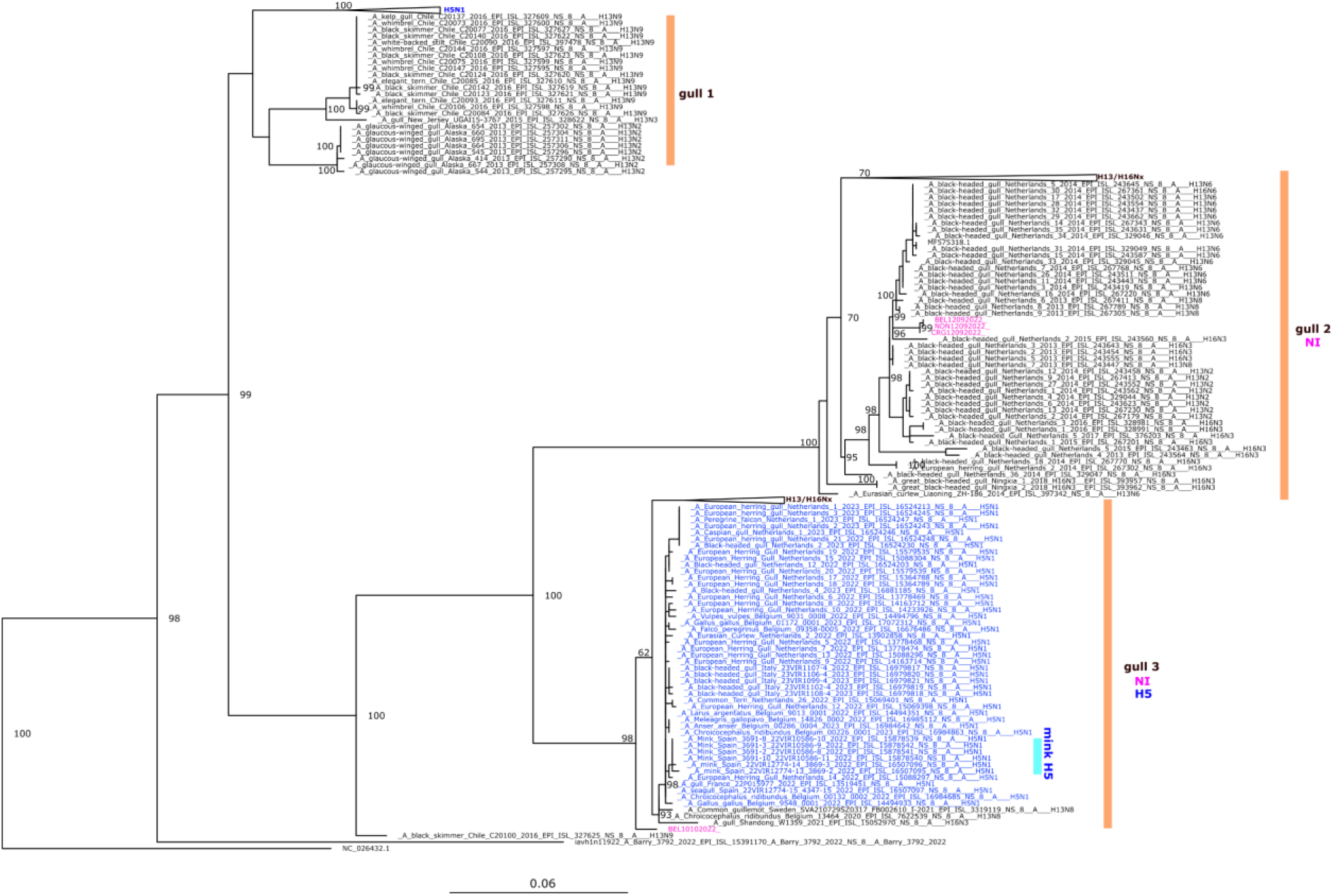
Maximum-likelihood phylogeny of wastewater-derived avian-like segment 8 sequences. Phylogenetic tree of the 4 avian-like segment 8 sequences (in pink) alongside all GISAID gull H13/16-like sequences (black), and 2022 H5N1 European sequences (in blue) (accessed January 2023). Three clades are highlighted (gull 1 -3) with presence of NI (pink) and H5 (blue) annotated. Mink H5N1 sequences are highlighted in cyan. For illustrative purposes, several clades are shown collapsed in cartoon form. Relevant bootstrap replicate values are shown on the tree.

### WBE reveals co-circulation of multiple lineages of human associated H1N1 and H3N2

To determine whether our derived sequences might also provide information on the evolution of the human epidemic, derived human segments 4 and 6 consensus sequences were analysed further. To maximise the genomic information available, for this analysis, we included all generated consensus sequences at our disposal, using Nextclade to identify sequences of low quality that were subsequently removed from analysis. This resulted in 11 sequences from 7 samples (available in supplement). Phylogenetic analyses focused on our segment 4 and 6 sequences compared to those from recently circulating IAVs from around the world and 2022 Northern Ireland, demonstrated a diversity in WBE sequences, from at least one H1N1 clade (6B.1A.5a.2a.1 [5a.2a.1]) and three H3N2 clades (all ‘Bangladesh-like’ 3C.2a1b.2a.2a.1 [2a.1], 3C.2a1b.2a.2a.3a.1 [2a.3a.1], and 3C.2a1b.2a.2b [2b]) as suggested by our initial analysis with Centrifuge (Figure 4 **A - D**). Our sequences were similar to recently circulating strains in NI but phylogenetically distinct from those HA sequences incorporated into the utilised vaccines (5a.2 for H1N1 and 3C.2a1b.2a.2 for H3N2) for that season.

**Figure 4.**
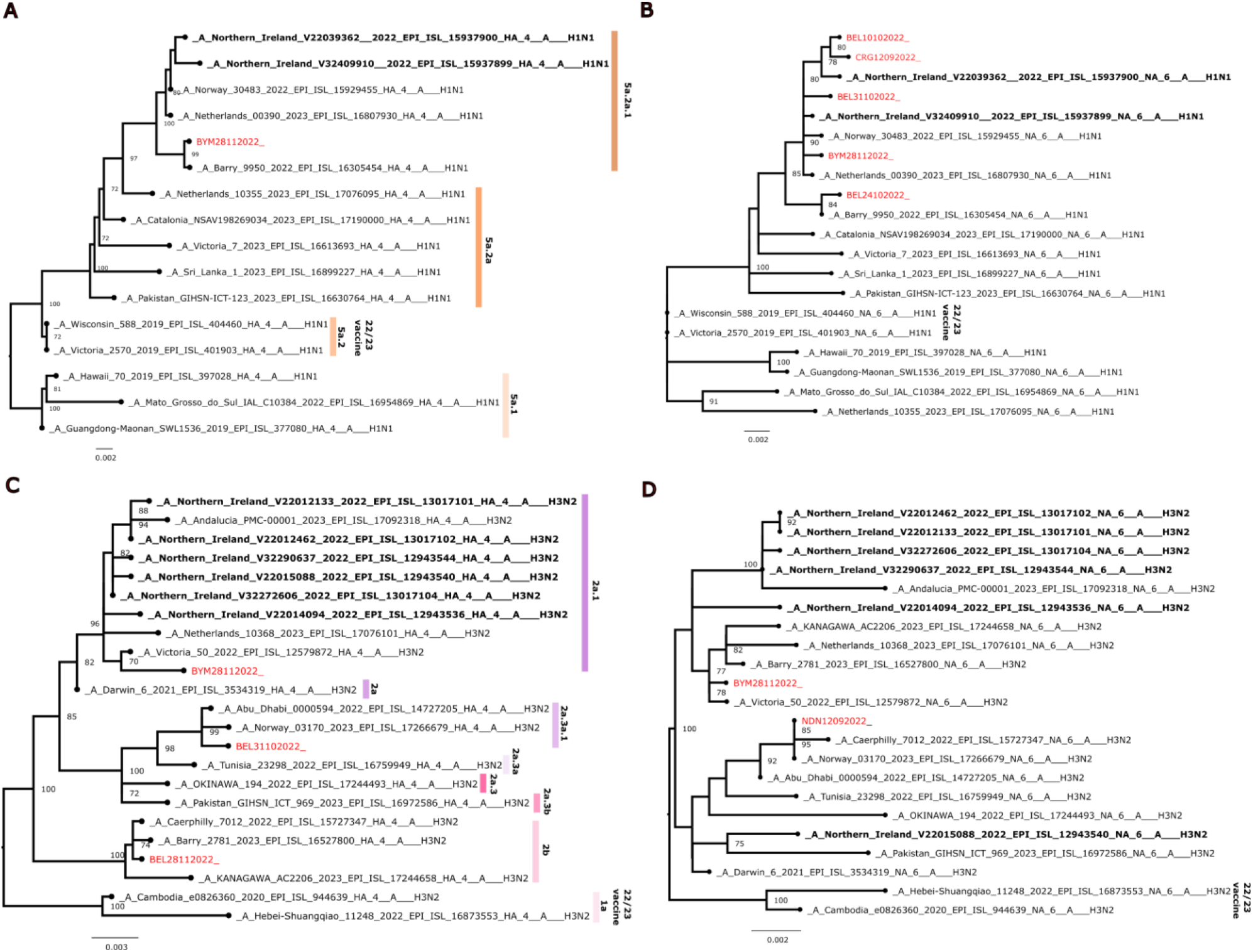
Maximum-likelihood phylogeny of wastewater-derived human-like segment 4 (HA) and 6 (NA) sequences from H1N1 and H3N2. Phylogenetic trees of the 11 human-like segment 4 and 6 sequences (in red) for H1N1 4 (A) and 6 (B) as well as H3N2 4 (C) and 6 (D) alongside select recent worldwide sequences and all available relevant Northern Ireland (NI) clinical sequences (in bold black). H1N1 NI clinical sequences were sampled during the 2022-2023 season, while the H3N2 NI clinical sequences were sampled in Spring 2022 during the end of the 2021-2022 season. Established HA sequence clades are shown alongside sequence groupings. Northern Hemisphere 2022/2023 winter season vaccine strain sequences highlighted. Bootstrap replicate values >69 are only shown on the tree.

## Discussion

Here we show that WBE, a surveillance approach traditionally focused on monitoring human infections, can also be repurposed and through WGS be used to distinguish between circulating strains of the virus and characterise animal pathogens, in this instance avian IAV (Figure S5).

Like others, we detect a clear and dynamic IAV signal in wastewater samples, highlighting the utility of WBE for monitoring influenza (20,21,22,23,24,25,26). However, comparing our wastewater IAV viral load data from the 6 WWTW to the clinical % positivity rate across all of NI did not show a clear positive relationship. Like the rest of the UK (34), NI clinical positivity increased over time, while our IAV signal showed several clear peaks and troughs. WGS suggested that earlier in the season more avian IAV dominated before a later shift to human. Thus, considering our genomics findings, a lack of correlation may be expected given the overwhelming non-human avian IAV signal detected. This could potentially reflect dynamics of avian IAVs in wild avian reservoirs. Indeed, comparing peaks in early September and early October, distinct avian IAV strains were even identified. Alternatively, the lack of correlation could be explained by the lower % population coverage for the wastewater surveillance compared to clinical testing, which covers all NI.

Wastewater sequencing of IAV detected diverse avian IAV lineages, notably recent H13 and H16-like viruses, which are LPAIV subtypes infecting gulls and closely related species belonging to the order Charadriiformes, poultry and mammals (6,7,35,36,8). Remarkably, we identified one sequence (segment 8 BEL10102022) with close affinity to contemporaneous HPAI H5N1 viruses found across Europe, proven capable of spreading between mammals (12). Although recently in NI, gulls have been confirmed infected with H5N1 (37), it is unlikely that we detected the HPAIV H5N1 clade 2.3.4.4b as H5N1 homology was identified only for one segment (8; NS1/NEP), and phylogenetic analysis suggested that they fell outside of the recent H5N1 group. Indeed, we only found non-H5 avian H13 segments with no H5 identified. Notably, recent European H5N1 viruses have reassorted with local gull-associated viruses, acquiring segments 3, 5 and 8 (12). Gulls have been shown to be important in spreading HPAIV H5 (38).

Previous WBE IAV studies, sampling at both the catchment and near source level, and using a variety of processing methods, have proven that molecular detection of IAV/IBV from both solid and liquid WW fractions is possible, highlighting how effective this medium is for tracking influenza dynamics within human communities. However, these studies have usually adopted a human-centric screening and sequencing approach, possibly explaining why convincing evidence of avian IAV has not been reported thus far. It is also likely that timing and geography are factors impacting avian IAV detection. The stable mixture of human and avian IAV seen throughout the study period at the North Down WWTW may reflect to the coastal location of this site, which is close to important seabird colonies in Belfast Lough, the Copeland Islands and Strangford Lough (39). The other sampled WWTW sites (with the exception of Derry/Londonderry, from which no sequences were recovered) are located much further inland. Regarding the precise origin of our avian influenza sequences, WWTW in general are recognised as providing artificial habitats for birds year-round (40,41). However, without in-depth ecological studies at WWTW, other mechanisms must be considered - such as the role of mammal vectors (42) or that true human infection exists. However, we consider these scenarios less likely than direct avian deposition, as no human infections with H13 or H16 subtypes have been noted.

Our protocol generated sequence information from the early stages of the human 2022/23 IAV epidemic in NI, in particular covering segments 4 (HA) and 6 (NA) that facilitate subtyping and antigenic comparison. Given the dearth of available clinical IAV sequences from NI, an accurate comparison between wastewater and the clinical would be challenging, although wastewater-derived sequences generally matched contemporary globally circulating viruses. Comparing our sequences with those of the chosen vaccine strains for 2022/2023 showed phylogenetic distinctness, which may impact antigenicity although vaccine match was generally considered good for that year (43). This suggests that sensitive and effective real-time WBE IAV sequencing could be as a tool to inform in-season estimates of vaccine effectiveness and clinical burden. However, this would require further in-depth molecular and serological analysis of derived HA and NA sequences, and development of more specific RT-qPCR assays

We show that “meta” WGS of IAV from WW is possible, despite the unique challenges such complex sample matrices pose. The heterogenous mix of PCR inhibitors, fragmented state of shed virus and variations in viral titres present can negatively impact sequencing workflows. Capturing more virus material, reducing inhibitors, enhancing nucleic acid extraction/purification steps, using more robust enzyme systems, and updated sequencing reagent chemistries/technologies could improve sequencing outcomes. The choice of bioinformatic workflow, and attendant reference databases, should also be carefully considered to enhance sensitivity and accuracy. Recently developed, influenza focussed bioinformatics tools, such as those provided by “EPI2ME Labs” (44) and “INSaFLU” (45), promise a streamlined approach to deal with primary data, but it remains to be seen how suitable these workflows are for analysing highly complex mixed environmental samples such as those from wastewater.

In conclusion, we demonstrate that human-centric WBE can be refocussed as a useful tool for combined avian and human IAV genomic surveillance. WBE may augment established veterinary and clinical testing structures, presenting opportunities to give advance warning on the incidence and spread of IAV and provide a cost-effective means to develop a global and sustained year-round “one health” sentinel service for IAV.

## Supporting information

GISAID Acknowledgements

## Data Availability

All data produced in the present study are available upon reasonable request to the authors.

## Notes

### Competing Interest Statement

The authors have declared no competing interest.

