## Supplementary material for "‘One Health’ Genomic Surveillance of Avian and Human Influenza A Viruses Through Environmental Wastewater Monitoring": GISAID Acknowledgements

| Segment ID | Segment | Country | Collection date | Isolate-ID | Isolate name | Originating Lab | Submitting Lab | Authors |
|---|---|---|---|---|---|---|---|---|
| EPI2181379 | NS | China | 2021-Oct-10 | EPI_ISL_15052970 | A/gull/Shandong/W1359/2021 | College of Agronomy, Liaocheng University | Liaocheng University | |
| EPI2181378 | MP | China | 2021-Oct-10 | EPI_ISL_15052970 | A/gull/Shandong/W1359/2021 | College of Agronomy, Liaocheng University | Liaocheng University | |
| EPI2181376 | NP | China | 2021-Oct-10 | EPI_ISL_15052970 | A/gull/Shandong/W1359/2021 | College of Agronomy, Liaocheng University | Liaocheng University | |
| EPI2311576 | NP | United States | 2021-Aug-01 | EPI_ISL_16641800 | A/gull/AK/21MB00111/2021 | Icahn School of Medicine at Mount Sinai | Icahn School of Medicine at Mount Sinai | Hill, Nichola; Bishop, MaryAnne; Puryear, Wendy; Foss, Alexa; Runstadler, Jonathan van de Guchte,Adriana; Silvia Gonzalez-Reiche,Ana ; Khalil,Zain ; van Bakel,Harm |
| EPI2311577 | NS | United States | 2021-Aug-01 | EPI_ISL_16641800 | A/gull/AK/21MB00111/2021 | Icahn School of Medicine at Mount Sinai | Icahn School of Medicine at Mount Sinai | Hill, Nichola; Bishop, MaryAnne; Puryear, Wendy; Foss, Alexa; Runstadler, Jonathan van de Guchte,Adriana; Silvia Gonzalez-Reiche,Ana ; Khalil,Zain ; van Bakel,Harm |
| EPI2311578 | MP | United States | 2021-Aug-01 | EPI_ISL_16641800 | A/gull/AK/21MB00111/2021 | Icahn School of Medicine at Mount Sinai | Icahn School of Medicine at Mount Sinai | Hill, Nichola; Bishop, MaryAnne; Puryear, Wendy; Foss, Alexa; Runstadler, Jonathan van de Guchte,Adriana; Silvia Gonzalez-Reiche,Ana ; Khalil,Zain ; van Bakel,Harm |
| EPI823673 | NP | Russian Federation | 2015-Sep-20 | EPI_ISL_231640 | A/black-headed gull/Kamchatka/007/2015 | Research Institute of Experimental and Clinical Medicine | Research Institute of Experimental and Clinical Medicine | Sharshov, K.A.; Sobolev, I.A.; Kabilov, M.R.; Shestopalov, A.M. |
| EPI823675 | MP | Russian Federation | 2015-Sep-20 | EPI_ISL_231640 | A/black-headed gull/Kamchatka/007/2015 | Research Institute of Experimental and Clinical Medicine | Research Institute of Experimental and Clinical Medicine | Sharshov, K.A.; Sobolev, I.A.; Kabilov, M.R.; Shestopalov, A.M. |
| EPI823676 | NS | Russian Federation | 2015-Sep-20 | EPI_ISL_231640 | A/black-headed gull/Kamchatka/007/2015 | Research Institute of Experimental and Clinical Medicine | Research Institute of Experimental and Clinical Medicine | Sharshov, K.A.; Sobolev, I.A.; Kabilov, M.R.; Shestopalov, A.M. |
| EPI2498183 | NP | United States | 2017-Aug-30 | EPI_ISL_17373344 | A/gull/USA/4_MNAI1365/2017 | | Import from public-domain | Rasmussen,E.A.; Cuthbert,F.J.; Tan,G.S.; Lemey,P.; Nelson,M.I.; Culhane,M.R.; Singh,V. |
| EPI2498189 | MP | United States | 2017-Aug-30 | EPI_ISL_17373344 | A/gull/USA/4_MNAI1365/2017 | | Import from public-domain | Rasmussen,E.A.; Cuthbert,F.J.; Tan,G.S.; Lemey,P.; Nelson,M.I.; Culhane,M.R.; Singh,V. |
| EPI2498192 | NS | United States | 2017-Aug-30 | EPI_ISL_17373344 | A/gull/USA/4_MNAI1365/2017 | | Import from public-domain | Rasmussen,E.A.; Cuthbert,F.J.; Tan,G.S.; Lemey,P.; Nelson,M.I.; Culhane,M.R.; Singh,V. |
| EPI2498182 | NP | United States | 2017-Aug-16 | EPI_ISL_17373343 | A/gull/USA/3_MNAI1172/2017 | | Import from public-domain | Rasmussen,E.A.; Cuthbert,F.J.; Tan,G.S.; Lemey,P.; Nelson,M.I.; Culhane,M.R.; Singh,V. |
| EPI2498188 | MP | United States | 2017-Aug-16 | EPI_ISL_17373343 | A/gull/USA/3_MNAI1172/2017 | | Import from public-domain | Rasmussen,E.A.; Cuthbert,F.J.; Tan,G.S.; Lemey,P.; Nelson,M.I.; Culhane,M.R.; Singh,V. |
| EPI2498191 | NS | United States | 2017-Aug-16 | EPI_ISL_17373343 | A/gull/USA/3_MNAI1172/2017 | | Import from public-domain | Rasmussen,E.A.; Cuthbert,F.J.; Tan,G.S.; Lemey,P.; Nelson,M.I.; Culhane,M.R.; Singh,V. |
| EPI2498181 | NP | United States | 2017-Aug-15 | EPI_ISL_17373342 | A/gull/USA/2_MNAI1144/2017 | | Import from public-domain | Rasmussen,E.A.; Cuthbert,F.J.; Tan,G.S.; Lemey,P.; Nelson,M.I.; Culhane,M.R.; Singh,V. |
| EPI2498187 | MP | United States | 2017-Aug-15 | EPI_ISL_17373342 | A/gull/USA/2_MNAI1144/2017 | | Import from public-domain | Rasmussen,E.A.; Cuthbert,F.J.; Tan,G.S.; Lemey,P.; Nelson,M.I.; Culhane,M.R.; Singh,V. |
| EPI2498190 | NS | United States | 2017-Aug-15 | EPI_ISL_17373342 | A/gull/USA/2_MNAI1144/2017 | | Import from public-domain | Rasmussen,E.A.; Cuthbert,F.J.; Tan,G.S.; Lemey,P.; Nelson,M.I.; Culhane,M.R.; Singh,V. |
| EPI1305481 | MP | United States | 2015-May-24 | EPI_ISL_328622 | A/gull/New Jersey/UGAI15-3767/2015 | | Import from public-domain | Tan,G.; Pickett,B.; Fedorova,N.; Amedeo,P.; Hu,L.; Christensen,J.; Miller,J.; Durbin,A.; Williams,T.; Poulson,R.; Stallknecht,D. |
| EPI1305483 | NP | United States | 2015-May-24 | EPI_ISL_328622 | A/gull/New Jersey/UGAI15-3767/2015 | | Import from public-domain | Tan,G.; Pickett,B.; Fedorova,N.; Amedeo,P.; Hu,L.; Christensen,J.; Miller,J.; Durbin,A.; Williams,T.; Poulson,R.; Stallknecht,D. |
| EPI1305484 | NS | United States | 2015-May-24 | EPI_ISL_328622 | A/gull/New Jersey/UGAI15-3767/2015 | | Import from public-domain | Tan,G.; Pickett,B.; Fedorova,N.; Amedeo,P.; Hu,L.; Christensen,J.; Miller,J.; Durbin,A.; Williams,T.; Poulson,R.; Stallknecht,D. |
| EPI1305473 | MP | United States | 2015-May-24 | EPI_ISL_328621 | A/gull/New Jersey/UGAI15-3728/2015 | | Import from public-domain | Tan,G.; Pickett,B.; Fedorova,N.; Amedeo,P.; Hu,L.; Christensen,J.; Miller,J.; Durbin,A.; Williams,T.; Poulson,R.; Stallknecht,D. |
| EPI1305475 | NP | United States | 2015-May-24 | EPI_ISL_328621 | A/gull/New Jersey/UGAI15-3728/2015 | | Import from public-domain | Tan,G.; Pickett,B.; Fedorova,N.; Amedeo,P.; Hu,L.; Christensen,J.; Miller,J.; Durbin,A.; Williams,T.; Poulson,R.; Stallknecht,D. |
| EPI1305476 | NS | United States | 2015-May-24 | EPI_ISL_328621 | A/gull/New Jersey/UGAI15-3728/2015 | | Import from public-domain | Tan,G.; Pickett,B.; Fedorova,N.; Amedeo,P.; Hu,L.; Christensen,J.; Miller,J.; Durbin,A.; Williams,T.; Poulson,R.; Stallknecht,D. |
| EPI1305465 | MP | United States | 2015-May-20 | EPI_ISL_328620 | A/gull/New Jersey/UGAI15-3459/2015 | | Import from public-domain | Tan,G.; Pickett,B.; Fedorova,N.; Amedeo,P.; Hu,L.; Christensen,J.; Miller,J.; Durbin,A.; Williams,T.; Poulson,R.; Stallknecht,D. |
| EPI1305467 | NP | United States | 2015-May-20 | EPI_ISL_328620 | A/gull/New Jersey/UGAI15-3459/2015 | | Import from public-domain | Tan,G.; Pickett,B.; Fedorova,N.; Amedeo,P.; Hu,L.; Christensen,J.; Miller,J.; Durbin,A.; Williams,T.; Poulson,R.; Stallknecht,D. |
| EPI1305468 | NS | United States | 2015-May-20 | EPI_ISL_328620 | A/gull/New Jersey/UGAI15-3459/2015 | | Import from public-domain | Tan,G.; Pickett,B.; Fedorova,N.; Amedeo,P.; Hu,L.; Christensen,J.; Miller,J.; Durbin,A.; Williams,T.; Poulson,R.; Stallknecht,D. |
| EPI1305457 | MP | United States | 2015-May-20 | EPI_ISL_328619 | A/gull/New Jersey/UGAI15-3416/2015 | | Import from public-domain | Tan,G.; Pickett,B.; Fedorova,N.; Amedeo,P.; Hu,L.; Christensen,J.; Miller,J.; Durbin,A.; Williams,T.; Poulson,R.; Stallknecht,D. |
| EPI1305459 | NP | United States | 2015-May-20 | EPI_ISL_328619 | A/gull/New Jersey/UGAI15-3416/2015 | | Import from public-domain | Tan,G.; Pickett,B.; Fedorova,N.; Amedeo,P.; Hu,L.; Christensen,J.; Miller,J.; Durbin,A.; Williams,T.; Poulson,R.; Stallknecht,D. |
| EPI1305460 | NS | United States | 2015-May-20 | EPI_ISL_328619 | A/gull/New Jersey/UGAI15-3416/2015 | | Import from public-domain | Tan,G.; Pickett,B.; Fedorova,N.; Amedeo,P.; Hu,L.; Christensen,J.; Miller,J.; Durbin,A.; Williams,T.; Poulson,R.; Stallknecht,D. |
| EPI1305449 | MP | United States | 2015-May-20 | EPI_ISL_328618 | A/gull/New Jersey/UGAI15-3414/2015 | | Import from public-domain | Tan,G.; Pickett,B.; Fedorova,N.; Amedeo,P.; Hu,L.; Christensen,J.; Miller,J.; Durbin,A.; Williams,T.; Poulson,R.; Stallknecht,D. |
| EPI1305451 | NP | United States | 2015-May-20 | EPI_ISL_328618 | A/gull/New Jersey/UGAI15-3414/2015 | | Import from public-domain | Tan,G.; Pickett,B.; Fedorova,N.; Amedeo,P.; Hu,L.; Christensen,J.; Miller,J.; Durbin,A.; Williams,T.; Poulson,R.; Stallknecht,D. |
| EPI1305452 | NS | United States | 2015-May-20 | EPI_ISL_328618 | A/gull/New Jersey/UGAI15-3414/2015 | | Import from public-domain | Tan,G.; Pickett,B.; Fedorova,N.; Amedeo,P.; Hu,L.; Christensen,J.; Miller,J.; Durbin,A.; Williams,T.; Poulson,R.; Stallknecht,D. |
| EPI699167 | MP | United States | 2013-Jul-03 | EPI_ISL_208103 | A/gull/Massachusetts/13JR03320/2013 | | Import from public-domain | Das,S.R.; Halpin,R.A.; Lin,X.; Simenauer,A.; Akopov,A.; Fedorova,N.; Puri,V.; Stockwell,T.; Amedeo,P.; Katzel,D.; Schobel,S.; Shrivastava,S.; Hill,N.; Puryear,W.; Bao,Y.; Sanders,R.; Zhdanov,S.; Kiryutin,B.; Lipman,D.J.; Tatusova,T.; Runstadler,J. |

| | | | | | | | | |
|---|---|---|---|---|---|---|---|---|
| EPI699169 | NP | United States | 2013-Jul-03 | EPI_ISL_208103 | A/gull/Massachusetts/13JR03320/2013 | | Import from public-domain | Das,S.R.; Halpin,R.A.; Lin,X.; Simenauer,A.; Akopov,A.; Fedorova,N.; Puri,V.; Stockwell,T.; Amedeo,P.; Katzel,D.; Schobel,S.; Shrivastava,S.; Hill,N.; Puryear,W.; Bao,Y.; Sanders,R.; Zhdanov,S.; Kiryutin,B.; Lipman,D.J.; Tatusova,T.; Runstadler,J. |
| EPI699170 | NS | United States | 2013-Jul-03 | EPI_ISL_208103 | A/gull/Massachusetts/13JR03320/2013 | | Import from public-domain | Das,S.R.; Halpin,R.A.; Lin,X.; Simenauer,A.; Akopov,A.; Fedorova,N.; Puri,V.; Stockwell,T.; Amedeo,P.; Katzel,D.; Schobel,S.; Shrivastava,S.; Hill,N.; Puryear,W.; Bao,Y.; Sanders,R.; Zhdanov,S.; Kiryutin,B.; Lipman,D.J.; Tatusova,T.; Runstadler,J. |
| EPI1057458 | MP | United States | 2014-Sep-03 | EPI_ISL_277379 | A/glaucous-winged gull/Southcentral Alaska/14MB02094/2014 | | Import from public-domain | Pickett,B.; Tan,G.; Fedorova,N.; Puri,V.; Shrivastava,S.; Amedeo,P.; Isom,R.; Hu,L.; Christensen,J.; Novotny,M.; Durbin,A.; Rocchi,I.; Williams,T.; Hill,N.; Bao,Y.; Sanders,R.; Zhdanov,S.; Kiryutin,B.; Lipman,D.J.; Tatusova,T.; Hatcher,E.; Runstadler,J. |
| EPI1057589 | NP | United States | 2014-Sep-03 | EPI_ISL_277379 | A/glaucous-winged gull/Southcentral Alaska/14MB02094/2014 | | Import from public-domain | Pickett,B.; Tan,G.; Fedorova,N.; Puri,V.; Shrivastava,S.; Amedeo,P.; Isom,R.; Hu,L.; Christensen,J.; Novotny,M.; Durbin,A.; Rocchi,I.; Williams,T.; Hill,N.; Bao,Y.; Sanders,R.; Zhdanov,S.; Kiryutin,B.; Lipman,D.J.; Tatusova,T.; Hatcher,E.; Runstadler,J. |
| EPI1057590 | NS | United States | 2014-Sep-03 | EPI_ISL_277379 | A/glaucous-winged gull/Southcentral Alaska/14MB02094/2014 | | Import from public-domain | Pickett,B.; Tan,G.; Fedorova,N.; Puri,V.; Shrivastava,S.; Amedeo,P.; Isom,R.; Hu,L.; Christensen,J.; Novotny,M.; Durbin,A.; Rocchi,I.; Williams,T.; Hill,N.; Bao,Y.; Sanders,R.; Zhdanov,S.; Kiryutin,B.; Lipman,D.J.; Tatusova,T.; Hatcher,E.; Runstadler,J. |
| EPI1353409 | NP | Mongolia | 2017-May-31 | EPI_ISL_337414 | A/herring gull/Uvs-Nuur Lake/56/2017 | National Institute of Animal Health | National Institute of Animal Health | Yoshida,E. |
| EPI1353410 | NS | Mongolia | 2017-May-31 | EPI_ISL_337414 | A/herring gull/Uvs-Nuur Lake/56/2017 | National Institute of Animal Health | National Institute of Animal Health | Yoshida,E. |
| EPI1353411 | MP | Mongolia | 2017-May-31 | EPI_ISL_337414 | A/herring gull/Uvs-Nuur Lake/56/2017 | National Institute of Animal Health | National Institute of Animal Health | Yoshida,E. |
| EPI1774601 | MP | Belgium | 2016-Oct-28 | EPI_ISL_399571 | A/Larus argentatus/Belgium/U1610336_H907447/2016 | | Import from public-domain | Steensels,M.; Lambrecht,B. |
| EPI1774604 | NS | Belgium | 2016-Oct-28 | EPI_ISL_399571 | A/Larus argentatus/Belgium/U1610336_H907447/2016 | | Import from public-domain | Steensels,M.; Lambrecht,B. |
| EPI1774603 | NP | Belgium | 2016-Oct-28 | EPI_ISL_399571 | A/Larus argentatus/Belgium/U1610336_H907447/2016 | | Import from public-domain | Steensels,M.; Lambrecht,B. |
| EPI1307160 | NP | Netherlands | 2015-Nov-26 | EPI_ISL_328987 | A/European herring gull/Netherlands/3/2015 | | Import from public-domain | Poen,M.J.; Verhagen,J.H.; Vuong,O.; Scheuer,R.D.; Pas,S.D.; Fouchier,R.A.M. |
| EPI1307162 | MP | Netherlands | 2015-Nov-26 | EPI_ISL_328987 | A/European herring gull/Netherlands/3/2015 | | Import from public-domain | Poen,M.J.; Verhagen,J.H.; Vuong,O.; Scheuer,R.D.; Pas,S.D.; Fouchier,R.A.M. |
| EPI1307176 | NS | Netherlands | 2015-Nov-26 | EPI_ISL_328987 | A/European herring gull/Netherlands/3/2015 | | Import from public-domain | Poen,M.J.; Verhagen,J.H.; Vuong,O.; Scheuer,R.D.; Pas,S.D.; Fouchier,R.A.M. |
| EPI1011048 | NP | Netherlands | 2014-Dec-24 | EPI_ISL_267302 | A/European herring gull/Netherlands/2/2014 | | Import from public-domain | |
| EPI1014519 | NS | Netherlands | 2014-Dec-24 | EPI_ISL_267302 | A/European herring gull/Netherlands/2/2014 | | Import from public-domain | |
| EPI1014955 | MP | Netherlands | 2014-Dec-24 | EPI_ISL_267302 | A/European herring gull/Netherlands/2/2014 | | Import from public-domain | |
| EPI542112 | NP | United States | 2013-Sep-03 | EPI_ISL_166438 | A/herring gull/Maine/ND0000047/2013 | | Import from public-domain | Sinnett,D.R.; Pedersen,K.; Killian,M.L. |
| EPI542114 | MP | United States | 2013-Sep-03 | EPI_ISL_166438 | A/herring gull/Maine/ND0000047/2013 | | Import from public-domain | Sinnett,D.R.; Pedersen,K.; Killian,M.L. |
| EPI542136 | NS | United States | 2013-Sep-03 | EPI_ISL_166438 | A/herring gull/Maine/ND0000047/2013 | | Import from public-domain | Sinnett,D.R.; Pedersen,K.; Killian,M.L. |
| EPI1223781 | NP | China | 2016-Dec-28 | EPI_ISL_308785 | A/black-tailed gull/Weihai/17/2016 | | Import from public-domain | Yu,Z.; Cheng,K. |
| EPI1223783 | MP | China | 2016-Dec-28 | EPI_ISL_308785 | A/black-tailed gull/Weihai/17/2016 | | Import from public-domain | Yu,Z.; Cheng,K. |
| EPI1223784 | NS | China | 2016-Dec-28 | EPI_ISL_308785 | A/black-tailed gull/Weihai/17/2016 | | Import from public-domain | Yu,Z.; Cheng,K. |
| EPI1223774 | NP | China | 2016-Dec-28 | EPI_ISL_308784 | A/black-tailed gull/Weihai/115/2016 | | Import from public-domain | Yu,Z.; Cheng,K. |
| EPI1223776 | MP | China | 2016-Dec-28 | EPI_ISL_308784 | A/black-tailed gull/Weihai/115/2016 | | Import from public-domain | Yu,Z.; Cheng,K. |
| EPI1223777 | NS | China | 2016-Dec-28 | EPI_ISL_308784 | A/black-tailed gull/Weihai/115/2016 | | Import from public-domain | Yu,Z.; Cheng,K. |
| EPI699203 | NP | United States | 2013-Dec-19 | EPI_ISL_208108 | A/ring-billed gull/Massachusetts/13DC30736-2/2013 | | Import from public-domain | Das,S.R.; Halpin,R.A.; Lin,X.; Simenauer,A.; Akopov,A.; Fedorova,N.; Puri,V.; Stockwell,T.; Amedeo,P.; Katzel,D.; Schobel,S.; Shrivastava,S.; Hill,N.; Puryear,W.; Bao,Y.; Sanders,R.; Zhdanov,S.; Kiryutin,B.; Lipman,D.J.; Tatusova,T.; Runstadler,J. |
| EPI699204 | NS | United States | 2013-Dec-19 | EPI_ISL_208108 | A/ring-billed gull/Massachusetts/13DC30736-2/2013 | | Import from public-domain | Das,S.R.; Halpin,R.A.; Lin,X.; Simenauer,A.; Akopov,A.; Fedorova,N.; Puri,V.; Stockwell,T.; Amedeo,P.; Katzel,D.; Schobel,S.; Shrivastava,S.; Hill,N.; Puryear,W.; Bao,Y.; Sanders,R.; Zhdanov,S.; Kiryutin,B.; Lipman,D.J.; Tatusova,T.; Runstadler,J. |
| EPI699193 | MP | United States | 2013-Dec-19 | EPI_ISL_208108 | A/ring-billed gull/Massachusetts/13DC30736-2/2013 | | Import from public-domain | Das,S.R.; Halpin,R.A.; Lin,X.; Simenauer,A.; Akopov,A.; Fedorova,N.; Puri,V.; Stockwell,T.; Amedeo,P.; Katzel,D.; Schobel,S.; Shrivastava,S.; Hill,N.; Puryear,W.; Bao,Y.; Sanders,R.; Zhdanov,S.; Kiryutin,B.; Lipman,D.J.; Tatusova,T.; Runstadler,J. |
| EPI699194 | MP | United States | 2013-Dec-19 | EPI_ISL_208107 | A/ring-billed gull/Massachusetts/13DC30736-1/2013 | | Import from public-domain | Das,S.R.; Halpin,R.A.; Lin,X.; Simenauer,A.; Akopov,A.; Fedorova,N.; Puri,V.; Stockwell,T.; Amedeo,P.; Katzel,D.; Schobel,S.; Shrivastava,S.; Hill,N.; Puryear,W.; Bao,Y.; Sanders,R.; Zhdanov,S.; Kiryutin,B.; Lipman,D.J.; Tatusova,T.; Runstadler,J. |
| EPI699196 | NP | United States | 2013-Dec-19 | EPI_ISL_208107 | A/ring-billed gull/Massachusetts/13DC30736-1/2013 | | Import from public-domain | Das,S.R.; Halpin,R.A.; Lin,X.; Simenauer,A.; Akopov,A.; Fedorova,N.; Puri,V.; Stockwell,T.; Amedeo,P.; Katzel,D.; Schobel,S.; Shrivastava,S.; Hill,N.; Puryear,W.; Bao,Y.; Sanders,R.; Zhdanov,S.; Kiryutin,B.; Lipman,D.J.; Tatusova,T.; Runstadler,J. |
| EPI699197 | NS | United States | 2013-Dec-19 | EPI_ISL_208107 | A/ring-billed gull/Massachusetts/13DC30736-1/2013 | | Import from public-domain | Das,S.R.; Halpin,R.A.; Lin,X.; Simenauer,A.; Akopov,A.; Fedorova,N.; Puri,V.; Stockwell,T.; Amedeo,P.; Katzel,D.; Schobel,S.; Shrivastava,S.; Hill,N.; Puryear,W.; Bao,Y.; Sanders,R.; Zhdanov,S.; Kiryutin,B.; Lipman,D.J.; Tatusova,T.; Runstadler,J. |
| EPI1301196 | NP | Chile | 2016-Dec-13 | EPI_ISL_327609 | A/kelp gull/Chile/C20137/2016 | | Import from public-domain | Bravo-Vasquez,N.; Bahl,J.; Hamilton-West,C.; Schultz-Cherry,S. |
| EPI1301198 | MP | Chile | 2016-Dec-13 | EPI_ISL_327609 | A/kelp gull/Chile/C20137/2016 | | Import from public-domain | Bravo-Vasquez,N.; Bahl,J.; Hamilton-West,C.; Schultz-Cherry,S. |
| EPI1301200 | NS | Chile | 2016-Dec-13 | EPI_ISL_327609 | A/kelp gull/Chile/C20137/2016 | | Import from public-domain | Bravo-Vasquez,N.; Bahl,J.; Hamilton-West,C.; Schultz-Cherry,S. |

| | | | | | | | |
|---|---|---|---|---|---|---|---|
| EPI1301159 | MP | Chile | 2017-Jun-23 | EPI_ISL_327607 | A/kelp gull/Chile/C27733/2017 | Import from public-domain | Bravo-Vasquez,N.; Bahl,J.; Hamilton-West,C.; Schultz-Cherry,S. |
| EPI1301169 | NS | Chile | 2017-Jun-23 | EPI_ISL_327607 | A/kelp gull/Chile/C27733/2017 | Import from public-domain | Bravo-Vasquez,N.; Bahl,J.; Hamilton-West,C.; Schultz-Cherry,S. |
| EPI1301171 | NP | Chile | 2017-Jun-23 | EPI_ISL_327607 | A/kelp gull/Chile/C27733/2017 | Import from public-domain | Bravo-Vasquez,N.; Bahl,J.; Hamilton-West,C.; Schultz-Cherry,S. |
| EPI1301180 | NP | Chile | 2016-Mar-10 | EPI_ISL_327606 | A/kelp gull/Chile/C8602/2016 | Import from public-domain | Bravo-Vasquez,N.; Bahl,J.; Hamilton-West,C.; Schultz-Cherry,S. |
| EPI1301154 | NS | Chile | 2016-Mar-10 | EPI_ISL_327606 | A/kelp gull/Chile/C8602/2016 | Import from public-domain | Bravo-Vasquez,N.; Bahl,J.; Hamilton-West,C.; Schultz-Cherry,S. |
| EPI1301166 | MP | Chile | 2016-Mar-10 | EPI_ISL_327606 | A/kelp gull/Chile/C8602/2016 | Import from public-domain | Bravo-Vasquez,N.; Bahl,J.; Hamilton-West,C.; Schultz-Cherry,S. |
| EPI1301150 | NP | Chile | 2016-Mar-10 | EPI_ISL_327605 | A/kelp gull/Chile/C8599/2016 | Import from public-domain | Bravo-Vasquez,N.; Bahl,J.; Hamilton-West,C.; Schultz-Cherry,S. |
| EPI1301153 | NS | Chile | 2016-Mar-10 | EPI_ISL_327605 | A/kelp gull/Chile/C8599/2016 | Import from public-domain | Bravo-Vasquez,N.; Bahl,J.; Hamilton-West,C.; Schultz-Cherry,S. |
| EPI1301174 | MP | Chile | 2016-Mar-10 | EPI_ISL_327605 | A/kelp gull/Chile/C8599/2016 | Import from public-domain | Bravo-Vasquez,N.; Bahl,J.; Hamilton-West,C.; Schultz-Cherry,S. |
| EPI1301188 | MP | Chile | 2016-Mar-10 | EPI_ISL_327604 | A/kelp gull/Chile/C8595/2016 | Import from public-domain | Bravo-Vasquez,N.; Bahl,J.; Hamilton-West,C.; Schultz-Cherry,S. |
| EPI1301149 | NP | Chile | 2016-Mar-10 | EPI_ISL_327604 | A/kelp gull/Chile/C8595/2016 | Import from public-domain | Bravo-Vasquez,N.; Bahl,J.; Hamilton-West,C.; Schultz-Cherry,S. |
| EPI1301177 | NS | Chile | 2016-Mar-10 | EPI_ISL_327604 | A/kelp gull/Chile/C8595/2016 | Import from public-domain | Bravo-Vasquez,N.; Bahl,J.; Hamilton-West,C.; Schultz-Cherry,S. |
| EPI1301146 | MP | Chile | 2016-Mar-10 | EPI_ISL_327603 | A/kelp gull/Chile/C8594/2016 | Import from public-domain | Bravo-Vasquez,N.; Bahl,J.; Hamilton-West,C.; Schultz-Cherry,S. |
| EPI1301147 | NP | Chile | 2016-Mar-10 | EPI_ISL_327603 | A/kelp gull/Chile/C8594/2016 | Import from public-domain | Bravo-Vasquez,N.; Bahl,J.; Hamilton-West,C.; Schultz-Cherry,S. |
| EPI1301168 | NS | Chile | 2016-Mar-10 | EPI_ISL_327603 | A/kelp gull/Chile/C8594/2016 | Import from public-domain | Bravo-Vasquez,N.; Bahl,J.; Hamilton-West,C.; Schultz-Cherry,S. |
| EPI1301128 | NS | Chile | 2016-Mar-10 | EPI_ISL_327602 | A/kelp gull/Chile/C8609/2016 | Import from public-domain | Bravo-Vasquez,N.; Bahl,J.; Hamilton-West,C.; Schultz-Cherry,S. |
| EPI1301129 | MP | Chile | 2016-Mar-10 | EPI_ISL_327602 | A/kelp gull/Chile/C8609/2016 | Import from public-domain | Bravo-Vasquez,N.; Bahl,J.; Hamilton-West,C.; Schultz-Cherry,S. |
| EPI1301140 | NP | Chile | 2016-Mar-10 | EPI_ISL_327602 | A/kelp gull/Chile/C8609/2016 | Import from public-domain | Bravo-Vasquez,N.; Bahl,J.; Hamilton-West,C.; Schultz-Cherry,S. |
| EPI1301112 | MP | Chile | 2016-Mar-15 | EPI_ISL_327601 | A/kelp gull/Chile/C8939/2016 | Import from public-domain | Bravo-Vasquez,N.; Bahl,J.; Hamilton-West,C.; Schultz-Cherry,S. |
| EPI1301137 | NP | Chile | 2016-Mar-15 | EPI_ISL_327601 | A/kelp gull/Chile/C8939/2016 | Import from public-domain | Bravo-Vasquez,N.; Bahl,J.; Hamilton-West,C.; Schultz-Cherry,S. |
| EPI1301142 | NS | Chile | 2016-Mar-15 | EPI_ISL_327601 | A/kelp gull/Chile/C8939/2016 | Import from public-domain | Bravo-Vasquez,N.; Bahl,J.; Hamilton-West,C.; Schultz-Cherry,S. |
| EPI1298538 | NP | Chile | 2016-Apr-27 | EPI_ISL_326977 | A/kelp gull/Chile/C10791/2016 | Import from public-domain | Jimenez-Bluhm,P.; Hamilton,C.; van Bakel,H.; Kriti,D.; Schultz-Cherry,S. |
| EPI1298584 | NS | Chile | 2016-Apr-27 | EPI_ISL_326977 | A/kelp gull/Chile/C10791/2016 | Import from public-domain | Jimenez-Bluhm,P.; Hamilton,C.; van Bakel,H.; Kriti,D.; Schultz-Cherry,S. |
| EPI1298628 | MP | Chile | 2016-Apr-27 | EPI_ISL_326977 | A/kelp gull/Chile/C10791/2016 | Import from public-domain | Jimenez-Bluhm,P.; Hamilton,C.; van Bakel,H.; Kriti,D.; Schultz-Cherry,S. |
| EPI1298465 | NS | Chile | 2016-Apr-27 | EPI_ISL_326972 | A/kelp gull/Chile/C10798/2016 | Import from public-domain | Jimenez-Bluhm,P.; Hamilton,C.; van Bakel,H.; Kriti,D.; Schultz-Cherry,S. |
| EPI1298578 | NP | Chile | 2016-Apr-27 | EPI_ISL_326972 | A/kelp gull/Chile/C10798/2016 | Import from public-domain | Jimenez-Bluhm,P.; Hamilton,C.; van Bakel,H.; Kriti,D.; Schultz-Cherry,S. |
| EPI1298590 | MP | Chile | 2016-Apr-27 | EPI_ISL_326972 | A/kelp gull/Chile/C10798/2016 | Import from public-domain | Jimenez-Bluhm,P.; Hamilton,C.; van Bakel,H.; Kriti,D.; Schultz-Cherry,S. |
| EPI1298457 | NP | Chile | 2016-Apr-22 | EPI_ISL_326967 | A/kelp gull/Chile/C10579/2016 | Import from public-domain | Jimenez-Bluhm,P.; Hamilton,C.; van Bakel,H.; Kriti,D.; Schultz-Cherry,S. |
| EPI1298526 | NS | Chile | 2016-Apr-22 | EPI_ISL_326967 | A/kelp gull/Chile/C10579/2016 | Import from public-domain | Jimenez-Bluhm,P.; Hamilton,C.; van Bakel,H.; Kriti,D.; Schultz-Cherry,S. |
| EPI1298619 | MP | Chile | 2016-Apr-22 | EPI_ISL_326967 | A/kelp gull/Chile/C10579/2016 | Import from public-domain | Jimenez-Bluhm,P.; Hamilton,C.; van Bakel,H.; Kriti,D.; Schultz-Cherry,S. |
| EPI1298455 | NS | Chile | 2016-May-31 | EPI_ISL_326965 | A/kelp gull/Chile/C12508/2016 | Import from public-domain | Jimenez-Bluhm,P.; Hamilton,C.; van Bakel,H.; Kriti,D.; Schultz-Cherry,S. |
| EPI1298591 | NP | Chile | 2016-May-31 | EPI_ISL_326965 | A/kelp gull/Chile/C12508/2016 | Import from public-domain | Jimenez-Bluhm,P.; Hamilton,C.; van Bakel,H.; Kriti,D.; Schultz-Cherry,S. |
| EPI1298604 | MP | Chile | 2016-May-31 | EPI_ISL_326965 | A/kelp gull/Chile/C12508/2016 | Import from public-domain | Jimenez-Bluhm,P.; Hamilton,C.; van Bakel,H.; Kriti,D.; Schultz-Cherry,S. |
| EPI1298450 | MP | Chile | 2016-Apr-27 | EPI_ISL_326961 | A/kelp gull/Chile/C10826/2016 | Import from public-domain | Jimenez-Bluhm,P.; Hamilton,C.; van Bakel,H.; Kriti,D.; Schultz-Cherry,S. |
| EPI1298514 | NP | Chile | 2016-Apr-27 | EPI_ISL_326961 | A/kelp gull/Chile/C10826/2016 | Import from public-domain | Jimenez-Bluhm,P.; Hamilton,C.; van Bakel,H.; Kriti,D.; Schultz-Cherry,S. |

| | | | | | | | |
|---|---|---|---|---|---|---|---|
| EPI1298603 | NS | Chile | 2016-Apr-27 | EPI_ISL_326961 | A/kelp gull/Chile/C10826/2016 | Import from public-domain | Jimenez-Bluhm,P.; Hamilton,C.; van Bakel,H.; Kriti,D.; Schultz-Cherry,S. |
| EPI1298540 | MP | Chile | 2016-Apr-27 | EPI_ISL_326959 | A/kelp gull/Chile/C10779/2016 | Import from public-domain | Jimenez-Bluhm,P.; Hamilton,C.; van Bakel,H.; Kriti,D.; Schultz-Cherry,S. |
| EPI1298521 | NP | Chile | 2016-Apr-27 | EPI_ISL_326959 | A/kelp gull/Chile/C10779/2016 | Import from public-domain | Jimenez-Bluhm,P.; Hamilton,C.; van Bakel,H.; Kriti,D.; Schultz-Cherry,S. |
| EPI1298609 | NS | Chile | 2016-Apr-27 | EPI_ISL_326959 | A/kelp gull/Chile/C10779/2016 | Import from public-domain | Jimenez-Bluhm,P.; Hamilton,C.; van Bakel,H.; Kriti,D.; Schultz-Cherry,S. |
| EPI1530530 | MP | United States | 2018-May-22 | EPI_ISL_373078 | A/kelp gull/Delaware Bay/441/2018 | Import from public-domain | Woodard,K.; Turner,J.C.; Franks,J.; Friedman,K.; Kercher,L.; Webby,R.J. |
| EPI1530531 | NS | United States | 2018-May-22 | EPI_ISL_373078 | A/kelp gull/Delaware Bay/441/2018 | Import from public-domain | Woodard,K.; Turner,J.C.; Franks,J.; Friedman,K.; Kercher,L.; Webby,R.J. |
| EPI1530533 | NP | United States | 2018-May-22 | EPI_ISL_373078 | A/kelp gull/Delaware Bay/441/2018 | Import from public-domain | Woodard,K.; Turner,J.C.; Franks,J.; Friedman,K.; Kercher,L.; Webby,R.J. |
| EPI1307067 | MP | Netherlands | 2015-Sep-18 | EPI_ISL_328974 | A/lesser black-backed gull/Netherlands/1/2015 | Import from public-domain | Poen,M.J.; Verhagen,J.H.; Vuong,O.; Scheuer,R.D.; Pas,S.D.; Fouchier,R.A.M. |
| EPI1307081 | NP | Netherlands | 2015-Sep-18 | EPI_ISL_328974 | A/lesser black-backed gull/Netherlands/1/2015 | Import from public-domain | Poen,M.J.; Verhagen,J.H.; Vuong,O.; Scheuer,R.D.; Pas,S.D.; Fouchier,R.A.M. |
| EPI1307082 | NS | Netherlands | 2015-Sep-18 | EPI_ISL_328974 | A/lesser black-backed gull/Netherlands/1/2015 | Import from public-domain | Poen,M.J.; Verhagen,J.H.; Vuong,O.; Scheuer,R.D.; Pas,S.D.; Fouchier,R.A.M. |
| EPI1975122 | NP | United States | 2020-Sep-04 | EPI_ISL_9589576 | A/Glaucous-Winged Gull/Alaska/20MB02820/2020 | Import from public-domain | Hill,N.; Runstadler,J. |
| EPI1975123 | NS | United States | 2020-Sep-04 | EPI_ISL_9589576 | A/Glaucous-Winged Gull/Alaska/20MB02820/2020 | Import from public-domain | Hill,N.; Runstadler,J. |
| EPI1975126 | MP | United States | 2020-Sep-04 | EPI_ISL_9589576 | A/Glaucous-Winged Gull/Alaska/20MB02820/2020 | Import from public-domain | Hill,N.; Runstadler,J. |
| EPI1975114 | MP | United States | 2020-Sep-04 | EPI_ISL_9589575 | A/Glaucous-Winged Gull/Alaska/20MB02834/2020 | Import from public-domain | Hill,N.; Runstadler,J. |
| EPI1975118 | NS | United States | 2020-Sep-04 | EPI_ISL_9589575 | A/Glaucous-Winged Gull/Alaska/20MB02834/2020 | Import from public-domain | Hill,N.; Runstadler,J. |
| EPI1975120 | NP | United States | 2020-Sep-04 | EPI_ISL_9589575 | A/Glaucous-Winged Gull/Alaska/20MB02834/2020 | Import from public-domain | Hill,N.; Runstadler,J. |
| EPI1975108 | NP | United States | 2020-Aug-10 | EPI_ISL_9589574 | A/Glaucous-Winged Gull/Alaska/20MB02534/2020 | Import from public-domain | Hill,N.; Runstadler,J. |
| EPI1975110 | NS | United States | 2020-Aug-10 | EPI_ISL_9589574 | A/Glaucous-Winged Gull/Alaska/20MB02534/2020 | Import from public-domain | Hill,N.; Runstadler,J. |
| EPI1975111 | MP | United States | 2020-Aug-10 | EPI_ISL_9589574 | A/Glaucous-Winged Gull/Alaska/20MB02534/2020 | Import from public-domain | Hill,N.; Runstadler,J. |
| EPI1975103 | NS | United States | 2020-Sep-04 | EPI_ISL_9589573 | A/Glaucous-Winged Gull/Alaska/20MB02826/2020 | Import from public-domain | Hill,N.; Runstadler,J. |
| EPI1975100 | MP | United States | 2020-Sep-04 | EPI_ISL_9589573 | A/Glaucous-Winged Gull/Alaska/20MB02826/2020 | Import from public-domain | Hill,N.; Runstadler,J. |
| EPI1975101 | NP | United States | 2020-Sep-04 | EPI_ISL_9589573 | A/Glaucous-Winged Gull/Alaska/20MB02826/2020 | Import from public-domain | Hill,N.; Runstadler,J. |
| EPI1975095 | NP | United States | 2020-Sep-04 | EPI_ISL_9589507 | A/Glaucous-Winged Gull/Alaska/20MB02836/2020 | Import from public-domain | Hill,N.; Runstadler,J. |
| EPI1975097 | MP | United States | 2020-Sep-04 | EPI_ISL_9589507 | A/Glaucous-Winged Gull/Alaska/20MB02836/2020 | Import from public-domain | Hill,N.; Runstadler,J. |
| EPI1975098 | NS | United States | 2020-Sep-04 | EPI_ISL_9589507 | A/Glaucous-Winged Gull/Alaska/20MB02836/2020 | Import from public-domain | Hill,N.; Runstadler,J. |
| EPI1975084 | NS | United States | 2020-Sep-04 | EPI_ISL_9589474 | A/Glaucous-Winged Gull/Alaska/20MB02837/2020 | Import from public-domain | Hill,N.; Runstadler,J. |
| EPI1975085 | MP | United States | 2020-Sep-04 | EPI_ISL_9589474 | A/Glaucous-Winged Gull/Alaska/20MB02837/2020 | Import from public-domain | Hill,N.; Runstadler,J. |
| EPI1975090 | NP | United States | 2020-Sep-04 | EPI_ISL_9589474 | A/Glaucous-Winged Gull/Alaska/20MB02837/2020 | Import from public-domain | Hill,N.; Runstadler,J. |
| EPI1975076 | MP | United States | 2020-Aug-07 | EPI_ISL_9589464 | A/Glaucous-Winged Gull/Alaska/20MB02498/2020 | Import from public-domain | Hill,N.; Runstadler,J. |
| EPI1975081 | NP | United States | 2020-Aug-07 | EPI_ISL_9589464 | A/Glaucous-Winged Gull/Alaska/20MB02498/2020 | Import from public-domain | Hill,N.; Runstadler,J. |
| EPI1975082 | NS | United States | 2020-Aug-07 | EPI_ISL_9589464 | A/Glaucous-Winged Gull/Alaska/20MB02498/2020 | Import from public-domain | Hill,N.; Runstadler,J. |
| EPI1975060 | NP | United States | 2020-Sep-05 | EPI_ISL_9589443 | A/Glaucous-Winged Gull/Alaska/20MB02858/2020 | Import from public-domain | Hill,N.; Runstadler,J. |
| EPI1975064 | MP | United States | 2020-Sep-05 | EPI_ISL_9589443 | A/Glaucous-Winged Gull/Alaska/20MB02858/2020 | Import from public-domain | Hill,N.; Runstadler,J. |
| EPI1975067 | NS | United States | 2020-Sep-05 | EPI_ISL_9589443 | A/Glaucous-Winged Gull/Alaska/20MB02858/2020 | Import from public-domain | Hill,N.; Runstadler,J. |
| EPI1975058 | NP | United States | 2020-Aug-21 | EPI_ISL_9589438 | A/Glaucous-Winged Gull/Alaska/20MB02698/2020 | Import from public-domain | Hill,N.; Runstadler,J. |

| | | | | | | | |
|---|---|---|---|---|---|---|---|
| EPI1975059 | NS | United States | 2020-Aug-21 | EPI_ISL_9589438 | A/Glaucous-Winged Gull/Alaska/20MB02698/2020 | Import from public-domain | Hill,N.; Runstadler,J. |
| EPI1975055 | MP | United States | 2020-Aug-21 | EPI_ISL_9589438 | A/Glaucous-Winged Gull/Alaska/20MB02698/2020 | Import from public-domain | Hill,N.; Runstadler,J. |
| EPI1975051 | NS | United States | 2020-Sep-04 | EPI_ISL_9589403 | A/Glaucous-Winged Gull/Alaska/20MB02816/2020 | Import from public-domain | Hill,N.; Runstadler,J. |
| EPI1975052 | MP | United States | 2020-Sep-04 | EPI_ISL_9589403 | A/Glaucous-Winged Gull/Alaska/20MB02816/2020 | Import from public-domain | Hill,N.; Runstadler,J. |
| EPI1975054 | NP | United States | 2020-Sep-04 | EPI_ISL_9589403 | A/Glaucous-Winged Gull/Alaska/20MB02816/2020 | Import from public-domain | Hill,N.; Runstadler,J. |
| EPI1975041 | MP | United States | 2020-Sep-04 | EPI_ISL_9589378 | A/Glaucous-Winged Gull/Alaska/20MB02831/2020 | Import from public-domain | Hill,N.; Runstadler,J. |
| EPI1975042 | NP | United States | 2020-Sep-04 | EPI_ISL_9589378 | A/Glaucous-Winged Gull/Alaska/20MB02831/2020 | Import from public-domain | Hill,N.; Runstadler,J. |
| EPI1975046 | NS | United States | 2020-Sep-04 | EPI_ISL_9589378 | A/Glaucous-Winged Gull/Alaska/20MB02831/2020 | Import from public-domain | Hill,N.; Runstadler,J. |
| EPI1975037 | NP | United States | 2020-Jul-21 | EPI_ISL_9589331 | A/Glaucous-Winged Gull/Alaska/20MB02182/2020 | Import from public-domain | Hill,N.; Runstadler,J. |
| EPI1975032 | MP | United States | 2020-Jul-21 | EPI_ISL_9589331 | A/Glaucous-Winged Gull/Alaska/20MB02182/2020 | Import from public-domain | Hill,N.; Runstadler,J. |
| EPI1975035 | NS | United States | 2020-Jul-21 | EPI_ISL_9589331 | A/Glaucous-Winged Gull/Alaska/20MB02182/2020 | Import from public-domain | Hill,N.; Runstadler,J. |
| EPI1894717 | MP | United States | 2019-Sep-09 | EPI_ISL_4056593 | A/Glaucous-Winged Gull/Alaska/19MB02557/2019 | Import from public-domain | Hill,N.; Runstadler,J. |
| EPI1894720 | NS | United States | 2019-Sep-09 | EPI_ISL_4056593 | A/Glaucous-Winged Gull/Alaska/19MB02557/2019 | Import from public-domain | Hill,N.; Runstadler,J. |
| EPI1894722 | NP | United States | 2019-Sep-09 | EPI_ISL_4056593 | A/Glaucous-Winged Gull/Alaska/19MB02557/2019 | Import from public-domain | Hill,N.; Runstadler,J. |
| EPI1894701 | NP | United States | 2019-Sep-05 | EPI_ISL_4056527 | A/Glaucous-Winged Gull/Alaska/19MB02458/2019 | Import from public-domain | Hill,N.; Runstadler,J. |
| EPI1894706 | MP | United States | 2019-Sep-05 | EPI_ISL_4056527 | A/Glaucous-Winged Gull/Alaska/19MB02458/2019 | Import from public-domain | Hill,N.; Runstadler,J. |
| EPI1894707 | NS | United States | 2019-Sep-05 | EPI_ISL_4056527 | A/Glaucous-Winged Gull/Alaska/19MB02458/2019 | Import from public-domain | Hill,N.; Runstadler,J. |
| EPI1894699 | MP | United States | 2019-Sep-05 | EPI_ISL_4056514 | A/Glaucous-Winged Gull/Alaska/19MB02464/2019 | Import from public-domain | Hill,N.; Runstadler,J. |
| EPI1894696 | NP | United States | 2019-Sep-05 | EPI_ISL_4056514 | A/Glaucous-Winged Gull/Alaska/19MB02464/2019 | Import from public-domain | Hill,N.; Runstadler,J. |
| EPI1894698 | NS | United States | 2019-Sep-05 | EPI_ISL_4056514 | A/Glaucous-Winged Gull/Alaska/19MB02464/2019 | Import from public-domain | Hill,N.; Runstadler,J. |
| EPI1894689 | MP | United States | 2019-Sep-05 | EPI_ISL_4056513 | A/Glaucous-Winged Gull/Alaska/19MB02477/2019 | Import from public-domain | Hill,N.; Runstadler,J. |
| EPI1894690 | NS | United States | 2019-Sep-05 | EPI_ISL_4056513 | A/Glaucous-Winged Gull/Alaska/19MB02477/2019 | Import from public-domain | Hill,N.; Runstadler,J. |
| EPI1894691 | NP | United States | 2019-Sep-05 | EPI_ISL_4056513 | A/Glaucous-Winged Gull/Alaska/19MB02477/2019 | Import from public-domain | Hill,N.; Runstadler,J. |
| EPI1894681 | NP | United States | 2019-Sep-06 | EPI_ISL_4056512 | A/Glaucous-Winged Gull/Alaska/19MB02504/2019 | Import from public-domain | Hill,N.; Runstadler,J. |
| EPI1894682 | MP | United States | 2019-Sep-06 | EPI_ISL_4056512 | A/Glaucous-Winged Gull/Alaska/19MB02504/2019 | Import from public-domain | Hill,N.; Runstadler,J. |
| EPI1894684 | NS | United States | 2019-Sep-06 | EPI_ISL_4056512 | A/Glaucous-Winged Gull/Alaska/19MB02504/2019 | Import from public-domain | Hill,N.; Runstadler,J. |
| EPI1894670 | MP | United States | 2019-Sep-06 | EPI_ISL_4056511 | A/Glaucous-Winged Gull/Alaska/19MB02547/2019 | Import from public-domain | Hill,N.; Runstadler,J. |
| EPI1894672 | NP | United States | 2019-Sep-06 | EPI_ISL_4056511 | A/Glaucous-Winged Gull/Alaska/19MB02547/2019 | Import from public-domain | Hill,N.; Runstadler,J. |
| EPI1894673 | NS | United States | 2019-Sep-06 | EPI_ISL_4056511 | A/Glaucous-Winged Gull/Alaska/19MB02547/2019 | Import from public-domain | Hill,N.; Runstadler,J. |
| EPI1894662 | NP | United States | 2019-Sep-05 | EPI_ISL_4056510 | A/Glaucous-Winged Gull/Alaska/19MB02493/2019 | Import from public-domain | Hill,N.; Runstadler,J. |
| EPI1894666 | MP | United States | 2019-Sep-05 | EPI_ISL_4056510 | A/Glaucous-Winged Gull/Alaska/19MB02493/2019 | Import from public-domain | Hill,N.; Runstadler,J. |
| EPI1894668 | NS | United States | 2019-Sep-05 | EPI_ISL_4056510 | A/Glaucous-Winged Gull/Alaska/19MB02493/2019 | Import from public-domain | Hill,N.; Runstadler,J. |
| EPI1894639 | NP | United States | 2019-Sep-05 | EPI_ISL_4056507 | A/Glaucous-Winged Gull/Alaska/19MB02474/2019 | Import from public-domain | Hill,N.; Runstadler,J. |
| EPI1894640 | MP | United States | 2019-Sep-05 | EPI_ISL_4056507 | A/Glaucous-Winged Gull/Alaska/19MB02474/2019 | Import from public-domain | Hill,N.; Runstadler,J. |
| EPI1894644 | NS | United States | 2019-Sep-05 | EPI_ISL_4056507 | A/Glaucous-Winged Gull/Alaska/19MB02474/2019 | Import from public-domain | Hill,N.; Runstadler,J. |

| | | | | | | | |
|---|---|---|---|---|---|---|---|
| EPI1894623 | NS | United States | 2019-Sep-05 | EPI_ISL_4056465 | A/Glaucous-Winged Gull/Alaska/19MB02459/2019 | Import from public-domain | Hill,N.; Runstadler,J. |
| EPI1894627 | NP | United States | 2019-Sep-05 | EPI_ISL_4056465 | A/Glaucous-Winged Gull/Alaska/19MB02459/2019 | Import from public-domain | Hill,N.; Runstadler,J. |
| EPI1894629 | MP | United States | 2019-Sep-05 | EPI_ISL_4056465 | A/Glaucous-Winged Gull/Alaska/19MB02459/2019 | Import from public-domain | Hill,N.; Runstadler,J. |
| EPI1894592 | NS | United States | 2019-Sep-05 | EPI_ISL_4056416 | A/Glaucous-Winged Gull/Alaska/19MB02461/2019 | Import from public-domain | Hill,N.; Runstadler,J. |
| EPI1894598 | MP | United States | 2019-Sep-05 | EPI_ISL_4056416 | A/Glaucous-Winged Gull/Alaska/19MB02461/2019 | Import from public-domain | Hill,N.; Runstadler,J. |
| EPI1894599 | NP | United States | 2019-Sep-05 | EPI_ISL_4056416 | A/Glaucous-Winged Gull/Alaska/19MB02461/2019 | Import from public-domain | Hill,N.; Runstadler,J. |
| EPI1894585 | NP | United States | 2019-Sep-05 | EPI_ISL_4056415 | A/Glaucous-Winged Gull/Alaska/19MB02467/2019 | Import from public-domain | Hill,N.; Runstadler,J. |
| EPI1894589 | MP | United States | 2019-Sep-05 | EPI_ISL_4056415 | A/Glaucous-Winged Gull/Alaska/19MB02467/2019 | Import from public-domain | Hill,N.; Runstadler,J. |
| EPI1894591 | NS | United States | 2019-Sep-05 | EPI_ISL_4056415 | A/Glaucous-Winged Gull/Alaska/19MB02467/2019 | Import from public-domain | Hill,N.; Runstadler,J. |
| EPI1894580 | NP | United States | 2019-Sep-09 | EPI_ISL_4056414 | A/Glaucous-Winged Gull/Alaska/19MB02555/2019 | Import from public-domain | Hill,N.; Runstadler,J. |
| EPI1894581 | NS | United States | 2019-Sep-09 | EPI_ISL_4056414 | A/Glaucous-Winged Gull/Alaska/19MB02555/2019 | Import from public-domain | Hill,N.; Runstadler,J. |
| EPI1894582 | MP | United States | 2019-Sep-09 | EPI_ISL_4056414 | A/Glaucous-Winged Gull/Alaska/19MB02555/2019 | Import from public-domain | Hill,N.; Runstadler,J. |
| EPI967006 | NP | United States | 2013-Sep-17 | EPI_ISL_257311 | A/glaucous-winged gull/Alaska/695/2013 | Import from public-domain | Reeves,A.B.; Ramey,A.M.; Hall,J.S. |
| EPI967010 | MP | United States | 2013-Sep-17 | EPI_ISL_257311 | A/glaucous-winged gull/Alaska/695/2013 | Import from public-domain | Reeves,A.B.; Ramey,A.M.; Hall,J.S. |
| EPI967012 | NS | United States | 2013-Sep-17 | EPI_ISL_257311 | A/glaucous-winged gull/Alaska/695/2013 | Import from public-domain | Reeves,A.B.; Ramey,A.M.; Hall,J.S. |
| EPI966990 | NP | United States | 2013-Sep-18 | EPI_ISL_257308 | A/glaucous-winged gull/Alaska/667/2013 | Import from public-domain | Reeves,A.B.; Ramey,A.M.; Hall,J.S. |
| EPI966994 | MP | United States | 2013-Sep-18 | EPI_ISL_257308 | A/glaucous-winged gull/Alaska/667/2013 | Import from public-domain | Reeves,A.B.; Ramey,A.M.; Hall,J.S. |
| EPI966996 | NS | United States | 2013-Sep-18 | EPI_ISL_257308 | A/glaucous-winged gull/Alaska/667/2013 | Import from public-domain | Reeves,A.B.; Ramey,A.M.; Hall,J.S. |
| EPI966974 | NP | United States | 2013-Sep-17 | EPI_ISL_257306 | A/glaucous-winged gull/Alaska/664/2013 | Import from public-domain | Reeves,A.B.; Ramey,A.M.; Hall,J.S. |
| EPI966978 | MP | United States | 2013-Sep-17 | EPI_ISL_257306 | A/glaucous-winged gull/Alaska/664/2013 | Import from public-domain | Reeves,A.B.; Ramey,A.M.; Hall,J.S. |
| EPI966980 | NS | United States | 2013-Sep-17 | EPI_ISL_257306 | A/glaucous-winged gull/Alaska/664/2013 | Import from public-domain | Reeves,A.B.; Ramey,A.M.; Hall,J.S. |
| EPI966959 | NP | United States | 2013-Sep-21 | EPI_ISL_257304 | A/glaucous-winged gull/Alaska/660/2013 | Import from public-domain | Reeves,A.B.; Ramey,A.M.; Hall,J.S. |
| EPI966963 | MP | United States | 2013-Sep-21 | EPI_ISL_257304 | A/glaucous-winged gull/Alaska/660/2013 | Import from public-domain | Reeves,A.B.; Ramey,A.M.; Hall,J.S. |
| EPI966965 | NS | United States | 2013-Sep-21 | EPI_ISL_257304 | A/glaucous-winged gull/Alaska/660/2013 | Import from public-domain | Reeves,A.B.; Ramey,A.M.; Hall,J.S. |
| EPI966943 | NP | United States | 2013-Sep-17 | EPI_ISL_257302 | A/glaucous-winged gull/Alaska/654/2013 | Import from public-domain | Reeves,A.B.; Ramey,A.M.; Hall,J.S. |
| EPI966947 | MP | United States | 2013-Sep-17 | EPI_ISL_257302 | A/glaucous-winged gull/Alaska/654/2013 | Import from public-domain | Reeves,A.B.; Ramey,A.M.; Hall,J.S. |
| EPI966949 | NS | United States | 2013-Sep-17 | EPI_ISL_257302 | A/glaucous-winged gull/Alaska/654/2013 | Import from public-domain | Reeves,A.B.; Ramey,A.M.; Hall,J.S. |
| EPI966904 | NP | United States | 2013-Sep-18 | EPI_ISL_257297 | A/glaucous-winged gull/Alaska/567/2013 | Import from public-domain | Reeves,A.B.; Ramey,A.M.; Hall,J.S. |
| EPI966906 | MP | United States | 2013-Sep-18 | EPI_ISL_257297 | A/glaucous-winged gull/Alaska/567/2013 | Import from public-domain | Reeves,A.B.; Ramey,A.M.; Hall,J.S. |
| EPI966907 | NS | United States | 2013-Sep-18 | EPI_ISL_257297 | A/glaucous-winged gull/Alaska/567/2013 | Import from public-domain | Reeves,A.B.; Ramey,A.M.; Hall,J.S. |
| EPI966896 | NP | United States | 2013-Sep-17 | EPI_ISL_257296 | A/glaucous-winged gull/Alaska/545/2013 | Import from public-domain | Reeves,A.B.; Ramey,A.M.; Hall,J.S. |
| EPI966898 | MP | United States | 2013-Sep-17 | EPI_ISL_257296 | A/glaucous-winged gull/Alaska/545/2013 | Import from public-domain | Reeves,A.B.; Ramey,A.M.; Hall,J.S. |
| EPI966899 | NS | United States | 2013-Sep-17 | EPI_ISL_257296 | A/glaucous-winged gull/Alaska/545/2013 | Import from public-domain | Reeves,A.B.; Ramey,A.M.; Hall,J.S. |
| EPI966888 | NP | United States | 2013-Sep-18 | EPI_ISL_257295 | A/glaucous-winged gull/Alaska/544/2013 | Import from public-domain | Reeves,A.B.; Ramey,A.M.; Hall,J.S. |
| EPI966890 | MP | United States | 2013-Sep-18 | EPI_ISL_257295 | A/glaucous-winged gull/Alaska/544/2013 | Import from public-domain | Reeves,A.B.; Ramey,A.M.; Hall,J.S. |

| | | | | | | | |
|---|---|---|---|---|---|---|---|
| EPI966891 | NS | United States | 2013-Sep-18 | EPI_ISL_257295 | A/glaucous-winged gull/Alaska/544/2013 | Import from public-domain | Reeves,A.B.; Ramey,A.M.; Hall,J.S. |
| EPI966871 | NP | United States | 2013-Sep-18 | EPI_ISL_257293 | A/glaucous-winged gull/Alaska/458/2013 | Import from public-domain | Reeves,A.B.; Ramey,A.M.; Hall,J.S. |
| EPI966873 | MP | United States | 2013-Sep-18 | EPI_ISL_257293 | A/glaucous-winged gull/Alaska/458/2013 | Import from public-domain | Reeves,A.B.; Ramey,A.M.; Hall,J.S. |
| EPI966874 | NS | United States | 2013-Sep-18 | EPI_ISL_257293 | A/glaucous-winged gull/Alaska/458/2013 | Import from public-domain | Reeves,A.B.; Ramey,A.M.; Hall,J.S. |
| EPI966851 | NP | United States | 2013-Sep-18 | EPI_ISL_257290 | A/glaucous-winged gull/Alaska/414/2013 | Import from public-domain | Reeves,A.B.; Ramey,A.M.; Hall,J.S. |
| EPI966855 | MP | United States | 2013-Sep-18 | EPI_ISL_257290 | A/glaucous-winged gull/Alaska/414/2013 | Import from public-domain | Reeves,A.B.; Ramey,A.M.; Hall,J.S. |
| EPI966857 | NS | United States | 2013-Sep-18 | EPI_ISL_257290 | A/glaucous-winged gull/Alaska/414/2013 | Import from public-domain | Reeves,A.B.; Ramey,A.M.; Hall,J.S. |
| EPI966805 | NP | United States | 2013-Sep-18 | EPI_ISL_257285 | A/glaucous-winged gull/Alaska/387/2013 | Import from public-domain | Reeves,A.B.; Ramey,A.M.; Hall,J.S. |
| EPI966809 | MP | United States | 2013-Sep-18 | EPI_ISL_257285 | A/glaucous-winged gull/Alaska/387/2013 | Import from public-domain | Reeves,A.B.; Ramey,A.M.; Hall,J.S. |
| EPI966811 | NS | United States | 2013-Sep-18 | EPI_ISL_257285 | A/glaucous-winged gull/Alaska/387/2013 | Import from public-domain | Reeves,A.B.; Ramey,A.M.; Hall,J.S. |
| EPI729767 | NP | United States | 2014-Sep-24 | EPI_ISL_215575 | A/glaucous-winged gull/Alaska/915/2014 | Import from public-domain | Ramey,A.M.; Reeves,A.B.; TeSlaa,J.L.; Nashold,S.; Donnelly,T.; Bahl,J.; Hall,J.S.; Donnelly,T.F. |
| EPI729769 | MP | United States | 2014-Sep-24 | EPI_ISL_215575 | A/glaucous-winged gull/Alaska/915/2014 | Import from public-domain | Ramey,A.M.; Reeves,A.B.; TeSlaa,J.L.; Nashold,S.; Donnelly,T.; Bahl,J.; Hall,J.S.; Donnelly,T.F. |
| EPI729770 | NS | United States | 2014-Sep-24 | EPI_ISL_215575 | A/glaucous-winged gull/Alaska/915/2014 | Import from public-domain | Ramey,A.M.; Reeves,A.B.; TeSlaa,J.L.; Nashold,S.; Donnelly,T.; Bahl,J.; Hall,J.S.; Donnelly,T.F. |
| EPI1057666 | MP | United States | 2016-Aug-27 | EPI_ISL_277416 | A/glaucous-winged gull/Southcentral Alaska/16MB03160/2016 | Import from public-domain | Pickett,B.; Tan,G.; Fedorova,N.; Puri,V.; Shrivastava,S.; Amedeo,P.; Isom,R.; Hu,L.; Christensen,J.; Novotny,M.; Durbin,A.; Rocchi,I.; Williams,T.; Hill,N.; Bao,Y.; Sanders,R.; Zhdanov,S.; Kiryutin,B.; Lipman,D.J.; Tatusova,T.; Hatcher,E.; Runstadler,J. |
| EPI1057668 | NP | United States | 2016-Aug-27 | EPI_ISL_277416 | A/glaucous-winged gull/Southcentral Alaska/16MB03160/2016 | Import from public-domain | Pickett,B.; Tan,G.; Fedorova,N.; Puri,V.; Shrivastava,S.; Amedeo,P.; Isom,R.; Hu,L.; Christensen,J.; Novotny,M.; Durbin,A.; Rocchi,I.; Williams,T.; Hill,N.; Bao,Y.; Sanders,R.; Zhdanov,S.; Kiryutin,B.; Lipman,D.J.; Tatusova,T.; Hatcher,E.; Runstadler,J. |
| EPI1057669 | NS | United States | 2016-Aug-27 | EPI_ISL_277416 | A/glaucous-winged gull/Southcentral Alaska/16MB03160/2016 | Import from public-domain | Pickett,B.; Tan,G.; Fedorova,N.; Puri,V.; Shrivastava,S.; Amedeo,P.; Isom,R.; Hu,L.; Christensen,J.; Novotny,M.; Durbin,A.; Rocchi,I.; Williams,T.; Hill,N.; Bao,Y.; Sanders,R.; Zhdanov,S.; Kiryutin,B.; Lipman,D.J.; Tatusova,T.; Hatcher,E.; Runstadler,J. |
| EPI1057614 | MP | United States | 2016-Aug-19 | EPI_ISL_277415 | A/glaucous-winged gull/Southcentral Alaska/16MB03089/2016 | Import from public-domain | Pickett,B.; Tan,G.; Fedorova,N.; Puri,V.; Shrivastava,S.; Amedeo,P.; Isom,R.; Hu,L.; Christensen,J.; Novotny,M.; Durbin,A.; Rocchi,I.; Williams,T.; Hill,N.; Bao,Y.; Sanders,R.; Zhdanov,S.; Kiryutin,B.; Lipman,D.J.; Tatusova,T.; Hatcher,E.; Runstadler,J. |
| EPI1057661 | NP | United States | 2016-Aug-19 | EPI_ISL_277415 | A/glaucous-winged gull/Southcentral Alaska/16MB03089/2016 | Import from public-domain | Pickett,B.; Tan,G.; Fedorova,N.; Puri,V.; Shrivastava,S.; Amedeo,P.; Isom,R.; Hu,L.; Christensen,J.; Novotny,M.; Durbin,A.; Rocchi,I.; Williams,T.; Hill,N.; Bao,Y.; Sanders,R.; Zhdanov,S.; Kiryutin,B.; Lipman,D.J.; Tatusova,T.; Hatcher,E.; Runstadler,J. |
| EPI1057662 | NS | United States | 2016-Aug-19 | EPI_ISL_277415 | A/glaucous-winged gull/Southcentral Alaska/16MB03089/2016 | Import from public-domain | Pickett,B.; Tan,G.; Fedorova,N.; Puri,V.; Shrivastava,S.; Amedeo,P.; Isom,R.; Hu,L.; Christensen,J.; Novotny,M.; Durbin,A.; Rocchi,I.; Williams,T.; Hill,N.; Bao,Y.; Sanders,R.; Zhdanov,S.; Kiryutin,B.; Lipman,D.J.; Tatusova,T.; Hatcher,E.; Runstadler,J. |
| EPI1057652 | MP | United States | 2016-May-04 | EPI_ISL_277414 | A/glaucous-winged gull/Southcentral Alaska/16MB00031/2016 | Import from public-domain | Pickett,B.; Tan,G.; Fedorova,N.; Puri,V.; Shrivastava,S.; Amedeo,P.; Isom,R.; Hu,L.; Christensen,J.; Novotny,M.; Durbin,A.; Rocchi,I.; Williams,T.; Hill,N.; Bao,Y.; Sanders,R.; Zhdanov,S.; Kiryutin,B.; Lipman,D.J.; Tatusova,T.; Hatcher,E.; Runstadler,J. |
| EPI1057654 | NP | United States | 2016-May-04 | EPI_ISL_277414 | A/glaucous-winged gull/Southcentral Alaska/16MB00031/2016 | Import from public-domain | Pickett,B.; Tan,G.; Fedorova,N.; Puri,V.; Shrivastava,S.; Amedeo,P.; Isom,R.; Hu,L.; Christensen,J.; Novotny,M.; Durbin,A.; Rocchi,I.; Williams,T.; Hill,N.; Bao,Y.; Sanders,R.; Zhdanov,S.; Kiryutin,B.; Lipman,D.J.; Tatusova,T.; Hatcher,E.; Runstadler,J. |
| EPI1057655 | NS | United States | 2016-May-04 | EPI_ISL_277414 | A/glaucous-winged gull/Southcentral Alaska/16MB00031/2016 | Import from public-domain | Pickett,B.; Tan,G.; Fedorova,N.; Puri,V.; Shrivastava,S.; Amedeo,P.; Isom,R.; Hu,L.; Christensen,J.; Novotny,M.; Durbin,A.; Rocchi,I.; Williams,T.; Hill,N.; Bao,Y.; Sanders,R.; Zhdanov,S.; Kiryutin,B.; Lipman,D.J.; Tatusova,T.; Hatcher,E.; Runstadler,J. |
| EPI1057612 | MP | United States | 2016-Aug-19 | EPI_ISL_277413 | A/glaucous-winged gull/Southcentral Alaska/16MB03046/2016 | Import from public-domain | Pickett,B.; Tan,G.; Fedorova,N.; Puri,V.; Shrivastava,S.; Amedeo,P.; Isom,R.; Hu,L.; Christensen,J.; Novotny,M.; Durbin,A.; Rocchi,I.; Williams,T.; Hill,N.; Bao,Y.; Sanders,R.; Zhdanov,S.; Kiryutin,B.; Lipman,D.J.; Tatusova,T.; Hatcher,E.; Runstadler,J. |
| EPI1057648 | NS | United States | 2016-Aug-19 | EPI_ISL_277413 | A/glaucous-winged gull/Southcentral Alaska/16MB03046/2016 | Import from public-domain | Pickett,B.; Tan,G.; Fedorova,N.; Puri,V.; Shrivastava,S.; Amedeo,P.; Isom,R.; Hu,L.; Christensen,J.; Novotny,M.; Durbin,A.; Rocchi,I.; Williams,T.; Hill,N.; Bao,Y.; Sanders,R.; Zhdanov,S.; Kiryutin,B.; Lipman,D.J.; Tatusova,T.; Hatcher,E.; Runstadler,J. |
| EPI1057647 | NP | United States | 2016-Aug-19 | EPI_ISL_277413 | A/glaucous-winged gull/Southcentral Alaska/16MB03046/2016 | Import from public-domain | Pickett,B.; Tan,G.; Fedorova,N.; Puri,V.; Shrivastava,S.; Amedeo,P.; Isom,R.; Hu,L.; Christensen,J.; Novotny,M.; Durbin,A.; Rocchi,I.; Williams,T.; Hill,N.; Bao,Y.; Sanders,R.; Zhdanov,S.; Kiryutin,B.; Lipman,D.J.; Tatusova,T.; Hatcher,E.; Runstadler,J. |
| EPI1057638 | MP | United States | 2016-Aug-19 | EPI_ISL_277412 | A/glaucous-winged gull/Southcentral Alaska/16MB03039/2016 | Import from public-domain | Pickett,B.; Tan,G.; Fedorova,N.; Puri,V.; Shrivastava,S.; Amedeo,P.; Isom,R.; Hu,L.; Christensen,J.; Novotny,M.; Durbin,A.; Rocchi,I.; Williams,T.; Hill,N.; Bao,Y.; Sanders,R.; Zhdanov,S.; Kiryutin,B.; Lipman,D.J.; Tatusova,T.; Hatcher,E.; Runstadler,J. |
| EPI1057640 | NP | United States | 2016-Aug-19 | EPI_ISL_277412 | A/glaucous-winged gull/Southcentral Alaska/16MB03039/2016 | Import from public-domain | Pickett,B.; Tan,G.; Fedorova,N.; Puri,V.; Shrivastava,S.; Amedeo,P.; Isom,R.; Hu,L.; Christensen,J.; Novotny,M.; Durbin,A.; Rocchi,I.; Williams,T.; Hill,N.; Bao,Y.; Sanders,R.; Zhdanov,S.; Kiryutin,B.; Lipman,D.J.; Tatusova,T.; Hatcher,E.; Runstadler,J. |
| EPI1057641 | NS | United States | 2016-Aug-19 | EPI_ISL_277412 | A/glaucous-winged gull/Southcentral Alaska/16MB03039/2016 | Import from public-domain | Pickett,B.; Tan,G.; Fedorova,N.; Puri,V.; Shrivastava,S.; Amedeo,P.; Isom,R.; Hu,L.; Christensen,J.; Novotny,M.; Durbin,A.; Rocchi,I.; Williams,T.; Hill,N.; Bao,Y.; Sanders,R.; Zhdanov,S.; Kiryutin,B.; Lipman,D.J.; Tatusova,T.; Hatcher,E.; Runstadler,J. |
| EPI1057610 | MP | United States | 2016-Aug-19 | EPI_ISL_277411 | A/glaucous-winged gull/Southcentral Alaska/16MB03027/2016 | Import from public-domain | Pickett,B.; Tan,G.; Fedorova,N.; Puri,V.; Shrivastava,S.; Amedeo,P.; Isom,R.; Hu,L.; Christensen,J.; Novotny,M.; Durbin,A.; Rocchi,I.; Williams,T.; Hill,N.; Bao,Y.; Sanders,R.; Zhdanov,S.; Kiryutin,B.; Lipman,D.J.; Tatusova,T.; Hatcher,E.; Runstadler,J. |
| EPI1057633 | NP | United States | 2016-Aug-19 | EPI_ISL_277411 | A/glaucous-winged gull/Southcentral Alaska/16MB03027/2016 | Import from public-domain | Pickett,B.; Tan,G.; Fedorova,N.; Puri,V.; Shrivastava,S.; Amedeo,P.; Isom,R.; Hu,L.; Christensen,J.; Novotny,M.; Durbin,A.; Rocchi,I.; Williams,T.; Hill,N.; Bao,Y.; Sanders,R.; Zhdanov,S.; Kiryutin,B.; Lipman,D.J.; Tatusova,T.; Hatcher,E.; Runstadler,J. |
| EPI1057634 | NS | United States | 2016-Aug-19 | EPI_ISL_277411 | A/glaucous-winged gull/Southcentral Alaska/16MB03027/2016 | Import from public-domain | Pickett,B.; Tan,G.; Fedorova,N.; Puri,V.; Shrivastava,S.; Amedeo,P.; Isom,R.; Hu,L.; Christensen,J.; Novotny,M.; Durbin,A.; Rocchi,I.; Williams,T.; Hill,N.; Bao,Y.; Sanders,R.; Zhdanov,S.; Kiryutin,B.; Lipman,D.J.; Tatusova,T.; Hatcher,E.; Runstadler,J. |
| EPI1057627 | NS | United States | 2016-Aug-16 | EPI_ISL_277410 | A/glaucous-winged gull/Southcentral Alaska/16MB02960/2016 | Import from public-domain | Pickett,B.; Tan,G.; Fedorova,N.; Puri,V.; Shrivastava,S.; Amedeo,P.; Isom,R.; Hu,L.; Christensen,J.; Novotny,M.; Durbin,A.; Rocchi,I.; Williams,T.; Hill,N.; Bao,Y.; Sanders,R.; Zhdanov,S.; Kiryutin,B.; Lipman,D.J.; Tatusova,T.; Hatcher,E.; Runstadler,J. |
| EPI1057624 | MP | United States | 2016-Aug-16 | EPI_ISL_277410 | A/glaucous-winged gull/Southcentral Alaska/16MB02960/2016 | Import from public-domain | Pickett,B.; Tan,G.; Fedorova,N.; Puri,V.; Shrivastava,S.; Amedeo,P.; Isom,R.; Hu,L.; Christensen,J.; Novotny,M.; Durbin,A.; Rocchi,I.; Williams,T.; Hill,N.; Bao,Y.; Sanders,R.; Zhdanov,S.; Kiryutin,B.; Lipman,D.J.; Tatusova,T.; Hatcher,E.; Runstadler,J. |
| EPI1057626 | NP | United States | 2016-Aug-16 | EPI_ISL_277410 | A/glaucous-winged gull/Southcentral Alaska/16MB02960/2016 | Import from public-domain | Pickett,B.; Tan,G.; Fedorova,N.; Puri,V.; Shrivastava,S.; Amedeo,P.; Isom,R.; Hu,L.; Christensen,J.; Novotny,M.; Durbin,A.; Rocchi,I.; Williams,T.; Hill,N.; Bao,Y.; Sanders,R.; Zhdanov,S.; Kiryutin,B.; Lipman,D.J.; Tatusova,T.; Hatcher,E.; Runstadler,J. |
| EPI1057617 | MP | United States | 2016-Aug-16 | EPI_ISL_277409 | A/glaucous-winged gull/Southcentral Alaska/16MB02942/2016 | Import from public-domain | Pickett,B.; Tan,G.; Fedorova,N.; Puri,V.; Shrivastava,S.; Amedeo,P.; Isom,R.; Hu,L.; Christensen,J.; Novotny,M.; Durbin,A.; Rocchi,I.; Williams,T.; Hill,N.; Bao,Y.; Sanders,R.; Zhdanov,S.; Kiryutin,B.; Lipman,D.J.; Tatusova,T.; Hatcher,E.; Runstadler,J. |

| | | | | | | | |
|---|---|---|---|---|---|---|---|
| EPI1057619 | NP | United States | 2016-Aug-16 | EPI_ISL_277409 | A/glaucous-winged gull/Southcentral Alaska/16MB02942/2016 | Import from public-domain | Pickett,B.; Tan,G.; Fedorova,N.; Puri,V.; Shrivastava,S.; Amedeo,P.; Isom,R.; Hu,L.; Christensen,J.; Novotny,M.; Durbin,A.; Rocchi,I.; Williams,T.; Hill,N.; Bao,Y.; Sanders,R.; Zhdanov,S.; Kiryutin,B.; Lipman,D.J.; Tatusova,T.; Hatcher,E.; Runstadler,J. |
| EPI1057620 | NS | United States | 2016-Aug-16 | EPI_ISL_277409 | A/glaucous-winged gull/Southcentral Alaska/16MB02942/2016 | Import from public-domain | Pickett,B.; Tan,G.; Fedorova,N.; Puri,V.; Shrivastava,S.; Amedeo,P.; Isom,R.; Hu,L.; Christensen,J.; Novotny,M.; Durbin,A.; Rocchi,I.; Williams,T.; Hill,N.; Bao,Y.; Sanders,R.; Zhdanov,S.; Kiryutin,B.; Lipman,D.J.; Tatusova,T.; Hatcher,E.; Runstadler,J. |
| EPI1057602 | MP | United States | 2016-Aug-16 | EPI_ISL_277408 | A/glaucous-winged gull/Southcentral Alaska/16MB02941/2016 | Import from public-domain | Pickett,B.; Tan,G.; Fedorova,N.; Puri,V.; Shrivastava,S.; Amedeo,P.; Isom,R.; Hu,L.; Christensen,J.; Novotny,M.; Durbin,A.; Rocchi,I.; Williams,T.; Hill,N.; Bao,Y.; Sanders,R.; Zhdanov,S.; Kiryutin,B.; Lipman,D.J.; Tatusova,T.; Hatcher,E.; Runstadler,J. |
| EPI1057603 | NP | United States | 2016-Aug-16 | EPI_ISL_277408 | A/glaucous-winged gull/Southcentral Alaska/16MB02941/2016 | Import from public-domain | Pickett,B.; Tan,G.; Fedorova,N.; Puri,V.; Shrivastava,S.; Amedeo,P.; Isom,R.; Hu,L.; Christensen,J.; Novotny,M.; Durbin,A.; Rocchi,I.; Williams,T.; Hill,N.; Bao,Y.; Sanders,R.; Zhdanov,S.; Kiryutin,B.; Lipman,D.J.; Tatusova,T.; Hatcher,E.; Runstadler,J. |
| EPI1057604 | NS | United States | 2016-Aug-16 | EPI_ISL_277408 | A/glaucous-winged gull/Southcentral Alaska/16MB02941/2016 | Import from public-domain | Pickett,B.; Tan,G.; Fedorova,N.; Puri,V.; Shrivastava,S.; Amedeo,P.; Isom,R.; Hu,L.; Christensen,J.; Novotny,M.; Durbin,A.; Rocchi,I.; Williams,T.; Hill,N.; Bao,Y.; Sanders,R.; Zhdanov,S.; Kiryutin,B.; Lipman,D.J.; Tatusova,T.; Hatcher,E.; Runstadler,J. |
| EPI1057595 | MP | United States | 2016-Aug-16 | EPI_ISL_277407 | A/glaucous-winged gull/Southcentral Alaska/16MB02936/2016 | Import from public-domain | Pickett,B.; Tan,G.; Fedorova,N.; Puri,V.; Shrivastava,S.; Amedeo,P.; Isom,R.; Hu,L.; Christensen,J.; Novotny,M.; Durbin,A.; Rocchi,I.; Williams,T.; Hill,N.; Bao,Y.; Sanders,R.; Zhdanov,S.; Kiryutin,B.; Lipman,D.J.; Tatusova,T.; Hatcher,E.; Runstadler,J. |
| EPI1057596 | NP | United States | 2016-Aug-16 | EPI_ISL_277407 | A/glaucous-winged gull/Southcentral Alaska/16MB02936/2016 | Import from public-domain | Pickett,B.; Tan,G.; Fedorova,N.; Puri,V.; Shrivastava,S.; Amedeo,P.; Isom,R.; Hu,L.; Christensen,J.; Novotny,M.; Durbin,A.; Rocchi,I.; Williams,T.; Hill,N.; Bao,Y.; Sanders,R.; Zhdanov,S.; Kiryutin,B.; Lipman,D.J.; Tatusova,T.; Hatcher,E.; Runstadler,J. |
| EPI1057597 | NS | United States | 2016-Aug-16 | EPI_ISL_277407 | A/glaucous-winged gull/Southcentral Alaska/16MB02936/2016 | Import from public-domain | Pickett,B.; Tan,G.; Fedorova,N.; Puri,V.; Shrivastava,S.; Amedeo,P.; Isom,R.; Hu,L.; Christensen,J.; Novotny,M.; Durbin,A.; Rocchi,I.; Williams,T.; Hill,N.; Bao,Y.; Sanders,R.; Zhdanov,S.; Kiryutin,B.; Lipman,D.J.; Tatusova,T.; Hatcher,E.; Runstadler,J. |
| EPI1057582 | MP | United States | 2014-Aug-20 | EPI_ISL_277380 | A/glaucous-winged gull/Southcentral Alaska/14MB01444/2014 | Import from public-domain | Pickett,B.; Tan,G.; Fedorova,N.; Puri,V.; Shrivastava,S.; Amedeo,P.; Isom,R.; Hu,L.; Christensen,J.; Novotny,M.; Durbin,A.; Rocchi,I.; Williams,T.; Hill,N.; Bao,Y.; Sanders,R.; Zhdanov,S.; Kiryutin,B.; Lipman,D.J.; Tatusova,T.; Hatcher,E.; Runstadler,J. |
| EPI1057583 | NP | United States | 2014-Aug-20 | EPI_ISL_277380 | A/glaucous-winged gull/Southcentral Alaska/14MB01444/2014 | Import from public-domain | Pickett,B.; Tan,G.; Fedorova,N.; Puri,V.; Shrivastava,S.; Amedeo,P.; Isom,R.; Hu,L.; Christensen,J.; Novotny,M.; Durbin,A.; Rocchi,I.; Williams,T.; Hill,N.; Bao,Y.; Sanders,R.; Zhdanov,S.; Kiryutin,B.; Lipman,D.J.; Tatusova,T.; Hatcher,E.; Runstadler,J. |
| EPI1057584 | NS | United States | 2014-Aug-20 | EPI_ISL_277380 | A/glaucous-winged gull/Southcentral Alaska/14MB01444/2014 | Import from public-domain | Pickett,B.; Tan,G.; Fedorova,N.; Puri,V.; Shrivastava,S.; Amedeo,P.; Isom,R.; Hu,L.; Christensen,J.; Novotny,M.; Durbin,A.; Rocchi,I.; Williams,T.; Hill,N.; Bao,Y.; Sanders,R.; Zhdanov,S.; Kiryutin,B.; Lipman,D.J.; Tatusova,T.; Hatcher,E.; Runstadler,J. |
| EPI1057576 | NP | United States | 2013-Aug-07 | EPI_ISL_277378 | A/glaucous-winged gull/Southcentral Alaska/13MB02410/2013 | Import from public-domain | Pickett,B.; Tan,G.; Fedorova,N.; Puri,V.; Shrivastava,S.; Amedeo,P.; Isom,R.; Hu,L.; Christensen,J.; Novotny,M.; Durbin,A.; Rocchi,I.; Williams,T.; Hill,N.; Bao,Y.; Sanders,R.; Zhdanov,S.; Kiryutin,B.; Lipman,D.J.; Tatusova,T.; Hatcher,E.; Runstadler,J. |
| EPI1057577 | NS | United States | 2013-Aug-07 | EPI_ISL_277378 | A/glaucous-winged gull/Southcentral Alaska/13MB02410/2013 | Import from public-domain | Pickett,B.; Tan,G.; Fedorova,N.; Puri,V.; Shrivastava,S.; Amedeo,P.; Isom,R.; Hu,L.; Christensen,J.; Novotny,M.; Durbin,A.; Rocchi,I.; Williams,T.; Hill,N.; Bao,Y.; Sanders,R.; Zhdanov,S.; Kiryutin,B.; Lipman,D.J.; Tatusova,T.; Hatcher,E.; Runstadler,J. |
| EPI1057455 | MP | United States | 2013-Aug-07 | EPI_ISL_277378 | A/glaucous-winged gull/Southcentral Alaska/13MB02410/2013 | Import from public-domain | Pickett,B.; Tan,G.; Fedorova,N.; Puri,V.; Shrivastava,S.; Amedeo,P.; Isom,R.; Hu,L.; Christensen,J.; Novotny,M.; Durbin,A.; Rocchi,I.; Williams,T.; Hill,N.; Bao,Y.; Sanders,R.; Zhdanov,S.; Kiryutin,B.; Lipman,D.J.; Tatusova,T.; Hatcher,E.; Runstadler,J. |
| EPI1057567 | MP | United States | 2013-Jul-12 | EPI_ISL_277377 | A/glaucous-winged gull/Southcentral Alaska/13MB01431/2013 | Import from public-domain | Pickett,B.; Tan,G.; Fedorova,N.; Puri,V.; Shrivastava,S.; Amedeo,P.; Isom,R.; Hu,L.; Christensen,J.; Novotny,M.; Durbin,A.; Rocchi,I.; Williams,T.; Hill,N.; Bao,Y.; Sanders,R.; Zhdanov,S.; Kiryutin,B.; Lipman,D.J.; Tatusova,T.; Hatcher,E.; Runstadler,J. |
| EPI1057569 | NP | United States | 2013-Jul-12 | EPI_ISL_277377 | A/glaucous-winged gull/Southcentral Alaska/13MB01431/2013 | Import from public-domain | Pickett,B.; Tan,G.; Fedorova,N.; Puri,V.; Shrivastava,S.; Amedeo,P.; Isom,R.; Hu,L.; Christensen,J.; Novotny,M.; Durbin,A.; Rocchi,I.; Williams,T.; Hill,N.; Bao,Y.; Sanders,R.; Zhdanov,S.; Kiryutin,B.; Lipman,D.J.; Tatusova,T.; Hatcher,E.; Runstadler,J. |
| EPI1057570 | NS | United States | 2013-Jul-12 | EPI_ISL_277377 | A/glaucous-winged gull/Southcentral Alaska/13MB01431/2013 | Import from public-domain | Pickett,B.; Tan,G.; Fedorova,N.; Puri,V.; Shrivastava,S.; Amedeo,P.; Isom,R.; Hu,L.; Christensen,J.; Novotny,M.; Durbin,A.; Rocchi,I.; Williams,T.; Hill,N.; Bao,Y.; Sanders,R.; Zhdanov,S.; Kiryutin,B.; Lipman,D.J.; Tatusova,T.; Hatcher,E.; Runstadler,J. |
| EPI1057551 | MP | United States | 2016-May-04 | EPI_ISL_277374 | A/glaucous-winged gull/Southcentral Alaska/16MB00033/2016 | Import from public-domain | Pickett,B.; Tan,G.; Fedorova,N.; Puri,V.; Shrivastava,S.; Amedeo,P.; Isom,R.; Hu,L.; Christensen,J.; Novotny,M.; Durbin,A.; Rocchi,I.; Williams,T.; Hill,N.; Bao,Y.; Sanders,R.; Zhdanov,S.; Kiryutin,B.; Lipman,D.J.; Tatusova,T.; Hatcher,E.; Runstadler,J. |
| EPI1057553 | NP | United States | 2016-May-04 | EPI_ISL_277374 | A/glaucous-winged gull/Southcentral Alaska/16MB00033/2016 | Import from public-domain | Pickett,B.; Tan,G.; Fedorova,N.; Puri,V.; Shrivastava,S.; Amedeo,P.; Isom,R.; Hu,L.; Christensen,J.; Novotny,M.; Durbin,A.; Rocchi,I.; Williams,T.; Hill,N.; Bao,Y.; Sanders,R.; Zhdanov,S.; Kiryutin,B.; Lipman,D.J.; Tatusova,T.; Hatcher,E.; Runstadler,J. |
| EPI1057554 | NS | United States | 2016-May-04 | EPI_ISL_277374 | A/glaucous-winged gull/Southcentral Alaska/16MB00033/2016 | Import from public-domain | Pickett,B.; Tan,G.; Fedorova,N.; Puri,V.; Shrivastava,S.; Amedeo,P.; Isom,R.; Hu,L.; Christensen,J.; Novotny,M.; Durbin,A.; Rocchi,I.; Williams,T.; Hill,N.; Bao,Y.; Sanders,R.; Zhdanov,S.; Kiryutin,B.; Lipman,D.J.; Tatusova,T.; Hatcher,E.; Runstadler,J. |
| EPI939915 | MP | United States | 2013-Aug-30 | EPI_ISL_252227 | A/glaucous-winged gull/Southcentral Alaska/13MB02599/2013 | Import from public-domain | Das,S.R.; Halpin,R.A.; Lin,X.; Simenauer,A.; Akopov,A.; Fedorova,N.; Puri,V.; Stockwell,T.; Amedeo,P.; Katzel,D.; Schobel,S.; Shrivastava,S.; Hill,N.; Bao,Y.; Sanders,R.; Zhdanov,S.; Kiryutin,B.; Lipman,D.J.; Tatusova,T.; Runstadler,J. |
| EPI939917 | NS | United States | 2013-Aug-30 | EPI_ISL_252227 | A/glaucous-winged gull/Southcentral Alaska/13MB02599/2013 | Import from public-domain | Das,S.R.; Halpin,R.A.; Lin,X.; Simenauer,A.; Akopov,A.; Fedorova,N.; Puri,V.; Stockwell,T.; Amedeo,P.; Katzel,D.; Schobel,S.; Shrivastava,S.; Hill,N.; Bao,Y.; Sanders,R.; Zhdanov,S.; Kiryutin,B.; Lipman,D.J.; Tatusova,T.; Runstadler,J. |
| EPI939857 | NP | United States | 2013-Aug-30 | EPI_ISL_252227 | A/glaucous-winged gull/Southcentral Alaska/13MB02599/2013 | Import from public-domain | Das,S.R.; Halpin,R.A.; Lin,X.; Simenauer,A.; Akopov,A.; Fedorova,N.; Puri,V.; Stockwell,T.; Amedeo,P.; Katzel,D.; Schobel,S.; Shrivastava,S.; Hill,N.; Bao,Y.; Sanders,R.; Zhdanov,S.; Kiryutin,B.; Lipman,D.J.; Tatusova,T.; Runstadler,J. |
| EPI939907 | MP | United States | 2013-Aug-30 | EPI_ISL_252226 | A/glaucous-winged gull/Southcentral Alaska/13MB02593/2013 | Import from public-domain | Das,S.R.; Halpin,R.A.; Lin,X.; Simenauer,A.; Akopov,A.; Fedorova,N.; Puri,V.; Stockwell,T.; Amedeo,P.; Katzel,D.; Schobel,S.; Shrivastava,S.; Hill,N.; Bao,Y.; Sanders,R.; Zhdanov,S.; Kiryutin,B.; Lipman,D.J.; Tatusova,T.; Runstadler,J. |
| EPI939909 | NP | United States | 2013-Aug-30 | EPI_ISL_252226 | A/glaucous-winged gull/Southcentral Alaska/13MB02593/2013 | Import from public-domain | Das,S.R.; Halpin,R.A.; Lin,X.; Simenauer,A.; Akopov,A.; Fedorova,N.; Puri,V.; Stockwell,T.; Amedeo,P.; Katzel,D.; Schobel,S.; Shrivastava,S.; Hill,N.; Bao,Y.; Sanders,R.; Zhdanov,S.; Kiryutin,B.; Lipman,D.J.; Tatusova,T.; Runstadler,J. |
| EPI939910 | NS | United States | 2013-Aug-30 | EPI_ISL_252226 | A/glaucous-winged gull/Southcentral Alaska/13MB02593/2013 | Import from public-domain | Das,S.R.; Halpin,R.A.; Lin,X.; Simenauer,A.; Akopov,A.; Fedorova,N.; Puri,V.; Stockwell,T.; Amedeo,P.; Katzel,D.; Schobel,S.; Shrivastava,S.; Hill,N.; Bao,Y.; Sanders,R.; Zhdanov,S.; Kiryutin,B.; Lipman,D.J.; Tatusova,T.; Runstadler,J. |
| EPI939900 | MP | United States | 2013-Aug-30 | EPI_ISL_252225 | A/glaucous-winged gull/Southcentral Alaska/13MB02582/2013 | Import from public-domain | Das,S.R.; Halpin,R.A.; Lin,X.; Simenauer,A.; Akopov,A.; Fedorova,N.; Puri,V.; Stockwell,T.; Amedeo,P.; Katzel,D.; Schobel,S.; Shrivastava,S.; Hill,N.; Bao,Y.; Sanders,R.; Zhdanov,S.; Kiryutin,B.; Lipman,D.J.; Tatusova,T.; Runstadler,J. |
| EPI939902 | NP | United States | 2013-Aug-30 | EPI_ISL_252225 | A/glaucous-winged gull/Southcentral Alaska/13MB02582/2013 | Import from public-domain | Das,S.R.; Halpin,R.A.; Lin,X.; Simenauer,A.; Akopov,A.; Fedorova,N.; Puri,V.; Stockwell,T.; Amedeo,P.; Katzel,D.; Schobel,S.; Shrivastava,S.; Hill,N.; Bao,Y.; Sanders,R.; Zhdanov,S.; Kiryutin,B.; Lipman,D.J.; Tatusova,T.; Runstadler,J. |
| EPI939903 | NS | United States | 2013-Aug-30 | EPI_ISL_252225 | A/glaucous-winged gull/Southcentral Alaska/13MB02582/2013 | Import from public-domain | Das,S.R.; Halpin,R.A.; Lin,X.; Simenauer,A.; Akopov,A.; Fedorova,N.; Puri,V.; Stockwell,T.; Amedeo,P.; Katzel,D.; Schobel,S.; Shrivastava,S.; Hill,N.; Bao,Y.; Sanders,R.; Zhdanov,S.; Kiryutin,B.; Lipman,D.J.; Tatusova,T.; Runstadler,J. |
| EPI939895 | NP | United States | 2013-Aug-30 | EPI_ISL_252224 | A/glaucous-winged gull/Southcentral Alaska/13MB02569/2013 | Import from public-domain | Das,S.R.; Halpin,R.A.; Lin,X.; Simenauer,A.; Akopov,A.; Fedorova,N.; Puri,V.; Stockwell,T.; Amedeo,P.; Katzel,D.; Schobel,S.; Shrivastava,S.; Hill,N.; Bao,Y.; Sanders,R.; Zhdanov,S.; Kiryutin,B.; Lipman,D.J.; Tatusova,T.; Runstadler,J. |
| EPI939896 | NS | United States | 2013-Aug-30 | EPI_ISL_252224 | A/glaucous-winged gull/Southcentral Alaska/13MB02569/2013 | Import from public-domain | Das,S.R.; Halpin,R.A.; Lin,X.; Simenauer,A.; Akopov,A.; Fedorova,N.; Puri,V.; Stockwell,T.; Amedeo,P.; Katzel,D.; Schobel,S.; Shrivastava,S.; Hill,N.; Bao,Y.; Sanders,R.; Zhdanov,S.; Kiryutin,B.; Lipman,D.J.; Tatusova,T.; Runstadler,J. |
| EPI939854 | MP | United States | 2013-Aug-30 | EPI_ISL_252224 | A/glaucous-winged gull/Southcentral Alaska/13MB02569/2013 | Import from public-domain | Das,S.R.; Halpin,R.A.; Lin,X.; Simenauer,A.; Akopov,A.; Fedorova,N.; Puri,V.; Stockwell,T.; Amedeo,P.; Katzel,D.; Schobel,S.; Shrivastava,S.; Hill,N.; Bao,Y.; Sanders,R.; Zhdanov,S.; Kiryutin,B.; Lipman,D.J.; Tatusova,T.; Runstadler,J. |
| EPI939888 | NP | United States | 2013-Aug-30 | EPI_ISL_252223 | A/glaucous-winged gull/Southcentral Alaska/13MB02561/2013 | Import from public-domain | Das,S.R.; Halpin,R.A.; Lin,X.; Simenauer,A.; Akopov,A.; Fedorova,N.; Puri,V.; Stockwell,T.; Amedeo,P.; Katzel,D.; Schobel,S.; Shrivastava,S.; Hill,N.; Bao,Y.; Sanders,R.; Zhdanov,S.; Kiryutin,B.; Lipman,D.J.; Tatusova,T.; Runstadler,J. |
| EPI939889 | NS | United States | 2013-Aug-30 | EPI_ISL_252223 | A/glaucous-winged gull/Southcentral Alaska/13MB02561/2013 | Import from public-domain | Das,S.R.; Halpin,R.A.; Lin,X.; Simenauer,A.; Akopov,A.; Fedorova,N.; Puri,V.; Stockwell,T.; Amedeo,P.; Katzel,D.; Schobel,S.; Shrivastava,S.; Hill,N.; Bao,Y.; Sanders,R.; Zhdanov,S.; Kiryutin,B.; Lipman,D.J.; Tatusova,T.; Runstadler,J. |
| EPI939886 | MP | United States | 2013-Aug-30 | EPI_ISL_252223 | A/glaucous-winged gull/Southcentral Alaska/13MB02561/2013 | Import from public-domain | Das,S.R.; Halpin,R.A.; Lin,X.; Simenauer,A.; Akopov,A.; Fedorova,N.; Puri,V.; Stockwell,T.; Amedeo,P.; Katzel,D.; Schobel,S.; Shrivastava,S.; Hill,N.; Bao,Y.; Sanders,R.; Zhdanov,S.; Kiryutin,B.; Lipman,D.J.; Tatusova,T.; Runstadler,J. |

| | | | | | | | |
|---|---|---|---|---|---|---|---|
| EPI939879 | MP | United States | 2013-Aug-30 | EPI_ISL_252222 | A/glaucous-winged gull/Southcentral Alaska/13MB02558/2013 | Import from public-domain | Das,S.R.; Halpin,R.A.; Lin,X.; Simenauer,A.; Akopov,A.; Fedorova,N.; Puri,V.; Stockwell,T.; Amedeo,P.; Katzel,D.; Schobel,S.; Shrivastava,S.; Hill,N.; Bao,Y.; Sanders,R.; Zhdanov,S.; Kiryutin,B.; Lipman,D.J.; Tatusova,T.; Runstadler,J. |
| EPI939881 | NP | United States | 2013-Aug-30 | EPI_ISL_252222 | A/glaucous-winged gull/Southcentral Alaska/13MB02558/2013 | Import from public-domain | Das,S.R.; Halpin,R.A.; Lin,X.; Simenauer,A.; Akopov,A.; Fedorova,N.; Puri,V.; Stockwell,T.; Amedeo,P.; Katzel,D.; Schobel,S.; Shrivastava,S.; Hill,N.; Bao,Y.; Sanders,R.; Zhdanov,S.; Kiryutin,B.; Lipman,D.J.; Tatusova,T.; Runstadler,J. |
| EPI939882 | NS | United States | 2013-Aug-30 | EPI_ISL_252222 | A/glaucous-winged gull/Southcentral Alaska/13MB02558/2013 | Import from public-domain | Das,S.R.; Halpin,R.A.; Lin,X.; Simenauer,A.; Akopov,A.; Fedorova,N.; Puri,V.; Stockwell,T.; Amedeo,P.; Katzel,D.; Schobel,S.; Shrivastava,S.; Hill,N.; Bao,Y.; Sanders,R.; Zhdanov,S.; Kiryutin,B.; Lipman,D.J.; Tatusova,T.; Runstadler,J. |
| EPI939851 | MP | United States | 2013-Aug-23 | EPI_ISL_252221 | A/glaucous-winged gull/Southcentral Alaska/13MB02527/2013 | Import from public-domain | Das,S.R.; Halpin,R.A.; Lin,X.; Simenauer,A.; Akopov,A.; Fedorova,N.; Puri,V.; Stockwell,T.; Amedeo,P.; Katzel,D.; Schobel,S.; Shrivastava,S.; Hill,N.; Bao,Y.; Sanders,R.; Zhdanov,S.; Kiryutin,B.; Lipman,D.J.; Tatusova,T.; Runstadler,J. |
| EPI939874 | NP | United States | 2013-Aug-23 | EPI_ISL_252221 | A/glaucous-winged gull/Southcentral Alaska/13MB02527/2013 | Import from public-domain | Das,S.R.; Halpin,R.A.; Lin,X.; Simenauer,A.; Akopov,A.; Fedorova,N.; Puri,V.; Stockwell,T.; Amedeo,P.; Katzel,D.; Schobel,S.; Shrivastava,S.; Hill,N.; Bao,Y.; Sanders,R.; Zhdanov,S.; Kiryutin,B.; Lipman,D.J.; Tatusova,T.; Runstadler,J. |
| EPI939875 | NS | United States | 2013-Aug-23 | EPI_ISL_252221 | A/glaucous-winged gull/Southcentral Alaska/13MB02527/2013 | Import from public-domain | Das,S.R.; Halpin,R.A.; Lin,X.; Simenauer,A.; Akopov,A.; Fedorova,N.; Puri,V.; Stockwell,T.; Amedeo,P.; Katzel,D.; Schobel,S.; Shrivastava,S.; Hill,N.; Bao,Y.; Sanders,R.; Zhdanov,S.; Kiryutin,B.; Lipman,D.J.; Tatusova,T.; Runstadler,J. |
| EPI939865 | MP | United States | 2013-Aug-23 | EPI_ISL_252220 | A/glaucous-winged gull/Southcentral Alaska/13MB02526/2013 | Import from public-domain | Das,S.R.; Halpin,R.A.; Lin,X.; Simenauer,A.; Akopov,A.; Fedorova,N.; Puri,V.; Stockwell,T.; Amedeo,P.; Katzel,D.; Schobel,S.; Shrivastava,S.; Hill,N.; Bao,Y.; Sanders,R.; Zhdanov,S.; Kiryutin,B.; Lipman,D.J.; Tatusova,T.; Runstadler,J. |
| EPI939867 | NP | United States | 2013-Aug-23 | EPI_ISL_252220 | A/glaucous-winged gull/Southcentral Alaska/13MB02526/2013 | Import from public-domain | Das,S.R.; Halpin,R.A.; Lin,X.; Simenauer,A.; Akopov,A.; Fedorova,N.; Puri,V.; Stockwell,T.; Amedeo,P.; Katzel,D.; Schobel,S.; Shrivastava,S.; Hill,N.; Bao,Y.; Sanders,R.; Zhdanov,S.; Kiryutin,B.; Lipman,D.J.; Tatusova,T.; Runstadler,J. |
| EPI939868 | NS | United States | 2013-Aug-23 | EPI_ISL_252220 | A/glaucous-winged gull/Southcentral Alaska/13MB02526/2013 | Import from public-domain | Das,S.R.; Halpin,R.A.; Lin,X.; Simenauer,A.; Akopov,A.; Fedorova,N.; Puri,V.; Stockwell,T.; Amedeo,P.; Katzel,D.; Schobel,S.; Shrivastava,S.; Hill,N.; Bao,Y.; Sanders,R.; Zhdanov,S.; Kiryutin,B.; Lipman,D.J.; Tatusova,T.; Runstadler,J. |
| EPI939858 | MP | United States | 2015-Sep-14 | EPI_ISL_252219 | A/glaucous-winged gull/Southcentral Alaska/15MB02018/2015 | Import from public-domain | Das,S.R.; Halpin,R.A.; Lin,X.; Simenauer,A.; Akopov,A.; Fedorova,N.; Puri,V.; Stockwell,T.; Amedeo,P.; Katzel,D.; Schobel,S.; Shrivastava,S.; Hill,N.; Bao,Y.; Sanders,R.; Zhdanov,S.; Kiryutin,B.; Lipman,D.J.; Tatusova,T.; Runstadler,J. |
| EPI939860 | NP | United States | 2015-Sep-14 | EPI_ISL_252219 | A/glaucous-winged gull/Southcentral Alaska/15MB02018/2015 | Import from public-domain | Das,S.R.; Halpin,R.A.; Lin,X.; Simenauer,A.; Akopov,A.; Fedorova,N.; Puri,V.; Stockwell,T.; Amedeo,P.; Katzel,D.; Schobel,S.; Shrivastava,S.; Hill,N.; Bao,Y.; Sanders,R.; Zhdanov,S.; Kiryutin,B.; Lipman,D.J.; Tatusova,T.; Runstadler,J. |
| EPI939861 | NS | United States | 2015-Sep-14 | EPI_ISL_252219 | A/glaucous-winged gull/Southcentral Alaska/15MB02018/2015 | Import from public-domain | Das,S.R.; Halpin,R.A.; Lin,X.; Simenauer,A.; Akopov,A.; Fedorova,N.; Puri,V.; Stockwell,T.; Amedeo,P.; Katzel,D.; Schobel,S.; Shrivastava,S.; Hill,N.; Bao,Y.; Sanders,R.; Zhdanov,S.; Kiryutin,B.; Lipman,D.J.; Tatusova,T.; Runstadler,J. |
| EPI939837 | MP | United States | 2015-Sep-14 | EPI_ISL_252217 | A/glaucous-winged gull/Southcentral Alaska/15MB02016/2015 | Import from public-domain | Das,S.R.; Halpin,R.A.; Lin,X.; Simenauer,A.; Akopov,A.; Fedorova,N.; Puri,V.; Stockwell,T.; Amedeo,P.; Katzel,D.; Schobel,S.; Shrivastava,S.; Hill,N.; Bao,Y.; Sanders,R.; Zhdanov,S.; Kiryutin,B.; Lipman,D.J.; Tatusova,T.; Runstadler,J. |
| EPI939838 | NP | United States | 2015-Sep-14 | EPI_ISL_252217 | A/glaucous-winged gull/Southcentral Alaska/15MB02016/2015 | Import from public-domain | Das,S.R.; Halpin,R.A.; Lin,X.; Simenauer,A.; Akopov,A.; Fedorova,N.; Puri,V.; Stockwell,T.; Amedeo,P.; Katzel,D.; Schobel,S.; Shrivastava,S.; Hill,N.; Bao,Y.; Sanders,R.; Zhdanov,S.; Kiryutin,B.; Lipman,D.J.; Tatusova,T.; Runstadler,J. |
| EPI939839 | NS | United States | 2015-Sep-14 | EPI_ISL_252217 | A/glaucous-winged gull/Southcentral Alaska/15MB02016/2015 | Import from public-domain | Das,S.R.; Halpin,R.A.; Lin,X.; Simenauer,A.; Akopov,A.; Fedorova,N.; Puri,V.; Stockwell,T.; Amedeo,P.; Katzel,D.; Schobel,S.; Shrivastava,S.; Hill,N.; Bao,Y.; Sanders,R.; Zhdanov,S.; Kiryutin,B.; Lipman,D.J.; Tatusova,T.; Runstadler,J. |
| EPI939831 | NP | United States | 2015-Sep-03 | EPI_ISL_252216 | A/glaucous-winged gull/Southcentral Alaska/15MB01776/2015 | Import from public-domain | Das,S.R.; Halpin,R.A.; Lin,X.; Simenauer,A.; Akopov,A.; Fedorova,N.; Puri,V.; Stockwell,T.; Amedeo,P.; Katzel,D.; Schobel,S.; Shrivastava,S.; Hill,N.; Bao,Y.; Sanders,R.; Zhdanov,S.; Kiryutin,B.; Lipman,D.J.; Tatusova,T.; Runstadler,J. |
| EPI939832 | NS | United States | 2015-Sep-03 | EPI_ISL_252216 | A/glaucous-winged gull/Southcentral Alaska/15MB01776/2015 | Import from public-domain | Das,S.R.; Halpin,R.A.; Lin,X.; Simenauer,A.; Akopov,A.; Fedorova,N.; Puri,V.; Stockwell,T.; Amedeo,P.; Katzel,D.; Schobel,S.; Shrivastava,S.; Hill,N.; Bao,Y.; Sanders,R.; Zhdanov,S.; Kiryutin,B.; Lipman,D.J.; Tatusova,T.; Runstadler,J. |
| EPI939799 | MP | United States | 2015-Sep-03 | EPI_ISL_252216 | A/glaucous-winged gull/Southcentral Alaska/15MB01776/2015 | Import from public-domain | Das,S.R.; Halpin,R.A.; Lin,X.; Simenauer,A.; Akopov,A.; Fedorova,N.; Puri,V.; Stockwell,T.; Amedeo,P.; Katzel,D.; Schobel,S.; Shrivastava,S.; Hill,N.; Bao,Y.; Sanders,R.; Zhdanov,S.; Kiryutin,B.; Lipman,D.J.; Tatusova,T.; Runstadler,J. |
| EPI939822 | MP | United States | 2015-Sep-01 | EPI_ISL_252215 | A/glaucous-winged gull/Southcentral Alaska/15MB01758/2015 | Import from public-domain | Das,S.R.; Halpin,R.A.; Lin,X.; Simenauer,A.; Akopov,A.; Fedorova,N.; Puri,V.; Stockwell,T.; Amedeo,P.; Katzel,D.; Schobel,S.; Shrivastava,S.; Hill,N.; Bao,Y.; Sanders,R.; Zhdanov,S.; Kiryutin,B.; Lipman,D.J.; Tatusova,T.; Runstadler,J. |
| EPI939824 | NP | United States | 2015-Sep-01 | EPI_ISL_252215 | A/glaucous-winged gull/Southcentral Alaska/15MB01758/2015 | Import from public-domain | Das,S.R.; Halpin,R.A.; Lin,X.; Simenauer,A.; Akopov,A.; Fedorova,N.; Puri,V.; Stockwell,T.; Amedeo,P.; Katzel,D.; Schobel,S.; Shrivastava,S.; Hill,N.; Bao,Y.; Sanders,R.; Zhdanov,S.; Kiryutin,B.; Lipman,D.J.; Tatusova,T.; Runstadler,J. |
| EPI939825 | NS | United States | 2015-Sep-01 | EPI_ISL_252215 | A/glaucous-winged gull/Southcentral Alaska/15MB01758/2015 | Import from public-domain | Das,S.R.; Halpin,R.A.; Lin,X.; Simenauer,A.; Akopov,A.; Fedorova,N.; Puri,V.; Stockwell,T.; Amedeo,P.; Katzel,D.; Schobel,S.; Shrivastava,S.; Hill,N.; Bao,Y.; Sanders,R.; Zhdanov,S.; Kiryutin,B.; Lipman,D.J.; Tatusova,T.; Runstadler,J. |
| EPI939818 | NS | United States | 2015-Sep-01 | EPI_ISL_252214 | A/glaucous-winged gull/Southcentral Alaska/15MB01747/2015 | Import from public-domain | Das,S.R.; Halpin,R.A.; Lin,X.; Simenauer,A.; Akopov,A.; Fedorova,N.; Puri,V.; Stockwell,T.; Amedeo,P.; Katzel,D.; Schobel,S.; Shrivastava,S.; Hill,N.; Bao,Y.; Sanders,R.; Zhdanov,S.; Kiryutin,B.; Lipman,D.J.; Tatusova,T.; Runstadler,J. |
| EPI939815 | MP | United States | 2015-Sep-01 | EPI_ISL_252214 | A/glaucous-winged gull/Southcentral Alaska/15MB01747/2015 | Import from public-domain | Das,S.R.; Halpin,R.A.; Lin,X.; Simenauer,A.; Akopov,A.; Fedorova,N.; Puri,V.; Stockwell,T.; Amedeo,P.; Katzel,D.; Schobel,S.; Shrivastava,S.; Hill,N.; Bao,Y.; Sanders,R.; Zhdanov,S.; Kiryutin,B.; Lipman,D.J.; Tatusova,T.; Runstadler,J. |
| EPI939817 | NP | United States | 2015-Sep-01 | EPI_ISL_252214 | A/glaucous-winged gull/Southcentral Alaska/15MB01747/2015 | Import from public-domain | Das,S.R.; Halpin,R.A.; Lin,X.; Simenauer,A.; Akopov,A.; Fedorova,N.; Puri,V.; Stockwell,T.; Amedeo,P.; Katzel,D.; Schobel,S.; Shrivastava,S.; Hill,N.; Bao,Y.; Sanders,R.; Zhdanov,S.; Kiryutin,B.; Lipman,D.J.; Tatusova,T.; Runstadler,J. |
| EPI939808 | MP | United States | 2015-Sep-01 | EPI_ISL_252213 | A/glaucous-winged gull/Southcentral Alaska/15MB01694/2015 | Import from public-domain | Das,S.R.; Halpin,R.A.; Lin,X.; Simenauer,A.; Akopov,A.; Fedorova,N.; Puri,V.; Stockwell,T.; Amedeo,P.; Katzel,D.; Schobel,S.; Shrivastava,S.; Hill,N.; Bao,Y.; Sanders,R.; Zhdanov,S.; Kiryutin,B.; Lipman,D.J.; Tatusova,T.; Runstadler,J. |
| EPI939810 | NP | United States | 2015-Sep-01 | EPI_ISL_252213 | A/glaucous-winged gull/Southcentral Alaska/15MB01694/2015 | Import from public-domain | Das,S.R.; Halpin,R.A.; Lin,X.; Simenauer,A.; Akopov,A.; Fedorova,N.; Puri,V.; Stockwell,T.; Amedeo,P.; Katzel,D.; Schobel,S.; Shrivastava,S.; Hill,N.; Bao,Y.; Sanders,R.; Zhdanov,S.; Kiryutin,B.; Lipman,D.J.; Tatusova,T.; Runstadler,J. |
| EPI939811 | NS | United States | 2015-Sep-01 | EPI_ISL_252213 | A/glaucous-winged gull/Southcentral Alaska/15MB01694/2015 | Import from public-domain | Das,S.R.; Halpin,R.A.; Lin,X.; Simenauer,A.; Akopov,A.; Fedorova,N.; Puri,V.; Stockwell,T.; Amedeo,P.; Katzel,D.; Schobel,S.; Shrivastava,S.; Hill,N.; Bao,Y.; Sanders,R.; Zhdanov,S.; Kiryutin,B.; Lipman,D.J.; Tatusova,T.; Runstadler,J. |
| EPI939801 | MP | United States | 2015-Aug-25 | EPI_ISL_252212 | A/glaucous-winged gull/Southcentral Alaska/15MB01680/2015 | Import from public-domain | Das,S.R.; Halpin,R.A.; Lin,X.; Simenauer,A.; Akopov,A.; Fedorova,N.; Puri,V.; Stockwell,T.; Amedeo,P.; Katzel,D.; Schobel,S.; Shrivastava,S.; Hill,N.; Bao,Y.; Sanders,R.; Zhdanov,S.; Kiryutin,B.; Lipman,D.J.; Tatusova,T.; Runstadler,J. |
| EPI939803 | NP | United States | 2015-Aug-25 | EPI_ISL_252212 | A/glaucous-winged gull/Southcentral Alaska/15MB01680/2015 | Import from public-domain | Das,S.R.; Halpin,R.A.; Lin,X.; Simenauer,A.; Akopov,A.; Fedorova,N.; Puri,V.; Stockwell,T.; Amedeo,P.; Katzel,D.; Schobel,S.; Shrivastava,S.; Hill,N.; Bao,Y.; Sanders,R.; Zhdanov,S.; Kiryutin,B.; Lipman,D.J.; Tatusova,T.; Runstadler,J. |
| EPI939804 | NS | United States | 2015-Aug-25 | EPI_ISL_252212 | A/glaucous-winged gull/Southcentral Alaska/15MB01680/2015 | Import from public-domain | Das,S.R.; Halpin,R.A.; Lin,X.; Simenauer,A.; Akopov,A.; Fedorova,N.; Puri,V.; Stockwell,T.; Amedeo,P.; Katzel,D.; Schobel,S.; Shrivastava,S.; Hill,N.; Bao,Y.; Sanders,R.; Zhdanov,S.; Kiryutin,B.; Lipman,D.J.; Tatusova,T.; Runstadler,J. |
| EPI939776 | MP | United States | 2015-Aug-25 | EPI_ISL_252208 | A/glaucous-winged gull/Southcentral Alaska/15MB01632/2015 | Import from public-domain | Das,S.R.; Halpin,R.A.; Lin,X.; Simenauer,A.; Akopov,A.; Fedorova,N.; Puri,V.; Stockwell,T.; Amedeo,P.; Katzel,D.; Schobel,S.; Shrivastava,S.; Hill,N.; Bao,Y.; Sanders,R.; Zhdanov,S.; Kiryutin,B.; Lipman,D.J.; Tatusova,T.; Runstadler,J. |
| EPI939778 | NP | United States | 2015-Aug-25 | EPI_ISL_252208 | A/glaucous-winged gull/Southcentral Alaska/15MB01632/2015 | Import from public-domain | Das,S.R.; Halpin,R.A.; Lin,X.; Simenauer,A.; Akopov,A.; Fedorova,N.; Puri,V.; Stockwell,T.; Amedeo,P.; Katzel,D.; Schobel,S.; Shrivastava,S.; Hill,N.; Bao,Y.; Sanders,R.; Zhdanov,S.; Kiryutin,B.; Lipman,D.J.; Tatusova,T.; Runstadler,J. |
| EPI939779 | NS | United States | 2015-Aug-25 | EPI_ISL_252208 | A/glaucous-winged gull/Southcentral Alaska/15MB01632/2015 | Import from public-domain | Das,S.R.; Halpin,R.A.; Lin,X.; Simenauer,A.; Akopov,A.; Fedorova,N.; Puri,V.; Stockwell,T.; Amedeo,P.; Katzel,D.; Schobel,S.; Shrivastava,S.; Hill,N.; Bao,Y.; Sanders,R.; Zhdanov,S.; Kiryutin,B.; Lipman,D.J.; Tatusova,T.; Runstadler,J. |
| EPI939768 | MP | United States | 2015-Aug-24 | EPI_ISL_252207 | A/glaucous-winged gull/Southcentral Alaska/15MB01429/2015 | Import from public-domain | Das,S.R.; Halpin,R.A.; Lin,X.; Simenauer,A.; Akopov,A.; Fedorova,N.; Puri,V.; Stockwell,T.; Amedeo,P.; Katzel,D.; Schobel,S.; Shrivastava,S.; Hill,N.; Bao,Y.; Sanders,R.; Zhdanov,S.; Kiryutin,B.; Lipman,D.J.; Tatusova,T.; Runstadler,J. |
| EPI939770 | NP | United States | 2015-Aug-24 | EPI_ISL_252207 | A/glaucous-winged gull/Southcentral Alaska/15MB01429/2015 | Import from public-domain | Das,S.R.; Halpin,R.A.; Lin,X.; Simenauer,A.; Akopov,A.; Fedorova,N.; Puri,V.; Stockwell,T.; Amedeo,P.; Katzel,D.; Schobel,S.; Shrivastava,S.; Hill,N.; Bao,Y.; Sanders,R.; Zhdanov,S.; Kiryutin,B.; Lipman,D.J.; Tatusova,T.; Runstadler,J. |

| | | | | | | | |
|---|---|---|---|---|---|---|---|
| EPI939771 | NS | United States | 2015-Aug-24 | EPI_ISL_252207 | A/glaucous-winged gull/Southcentral Alaska/15MB01429/2015 | Import from public-domain | Das,S.R.; Halpin,R.A.; Lin,X.; Simenauer,A.; Akopov,A.; Fedorova,N.; Puri,V.; Stockwell,T.; Amedeo,P.; Katzel,D.; Schobel,S.; Shrivastava,S.; Hill,N.; Bao,Y.; Sanders,R.; Zhdanov,S.; Kiryutin,B.; Lipman,D.J.; Tatusova,T.; Runstadler,J. |
| EPI939761 | MP | United States | 2015-Aug-21 | EPI_ISL_252206 | A/glaucous-winged gull/Southcentral Alaska/15MB01557/2015 | Import from public-domain | Das,S.R.; Halpin,R.A.; Lin,X.; Simenauer,A.; Akopov,A.; Fedorova,N.; Puri,V.; Stockwell,T.; Amedeo,P.; Katzel,D.; Schobel,S.; Shrivastava,S.; Hill,N.; Bao,Y.; Sanders,R.; Zhdanov,S.; Kiryutin,B.; Lipman,D.J.; Tatusova,T.; Runstadler,J. |
| EPI939763 | NP | United States | 2015-Aug-21 | EPI_ISL_252206 | A/glaucous-winged gull/Southcentral Alaska/15MB01557/2015 | Import from public-domain | Das,S.R.; Halpin,R.A.; Lin,X.; Simenauer,A.; Akopov,A.; Fedorova,N.; Puri,V.; Stockwell,T.; Amedeo,P.; Katzel,D.; Schobel,S.; Shrivastava,S.; Hill,N.; Bao,Y.; Sanders,R.; Zhdanov,S.; Kiryutin,B.; Lipman,D.J.; Tatusova,T.; Runstadler,J. |
| EPI939764 | NS | United States | 2015-Aug-21 | EPI_ISL_252206 | A/glaucous-winged gull/Southcentral Alaska/15MB01557/2015 | Import from public-domain | Das,S.R.; Halpin,R.A.; Lin,X.; Simenauer,A.; Akopov,A.; Fedorova,N.; Puri,V.; Stockwell,T.; Amedeo,P.; Katzel,D.; Schobel,S.; Shrivastava,S.; Hill,N.; Bao,Y.; Sanders,R.; Zhdanov,S.; Kiryutin,B.; Lipman,D.J.; Tatusova,T.; Runstadler,J. |
| EPI939705 | MP | United States | 2015-Sep-01 | EPI_ISL_252200 | A/glaucous-winged gull/Southcentral Alaska/15MB01735/2015 | Import from public-domain | Das,S.R.; Halpin,R.A.; Lin,X.; Simenauer,A.; Akopov,A.; Fedorova,N.; Puri,V.; Stockwell,T.; Amedeo,P.; Katzel,D.; Schobel,S.; Shrivastava,S.; Hill,N.; Bao,Y.; Sanders,R.; Zhdanov,S.; Kiryutin,B.; Lipman,D.J.; Tatusova,T.; Runstadler,J. |
| EPI939706 | NP | United States | 2015-Sep-01 | EPI_ISL_252200 | A/glaucous-winged gull/Southcentral Alaska/15MB01735/2015 | Import from public-domain | Das,S.R.; Halpin,R.A.; Lin,X.; Simenauer,A.; Akopov,A.; Fedorova,N.; Puri,V.; Stockwell,T.; Amedeo,P.; Katzel,D.; Schobel,S.; Shrivastava,S.; Hill,N.; Bao,Y.; Sanders,R.; Zhdanov,S.; Kiryutin,B.; Lipman,D.J.; Tatusova,T.; Runstadler,J. |
| EPI939707 | NS | United States | 2015-Sep-01 | EPI_ISL_252200 | A/glaucous-winged gull/Southcentral Alaska/15MB01735/2015 | Import from public-domain | Das,S.R.; Halpin,R.A.; Lin,X.; Simenauer,A.; Akopov,A.; Fedorova,N.; Puri,V.; Stockwell,T.; Amedeo,P.; Katzel,D.; Schobel,S.; Shrivastava,S.; Hill,N.; Bao,Y.; Sanders,R.; Zhdanov,S.; Kiryutin,B.; Lipman,D.J.; Tatusova,T.; Runstadler,J. |
| EPI939678 | MP | United States | 2015-Sep-01 | EPI_ISL_252199 | A/glaucous-winged gull/Southcentral Alaska/15MB01693/2015 | Import from public-domain | Das,S.R.; Halpin,R.A.; Lin,X.; Simenauer,A.; Akopov,A.; Fedorova,N.; Puri,V.; Stockwell,T.; Amedeo,P.; Katzel,D.; Schobel,S.; Shrivastava,S.; Hill,N.; Bao,Y.; Sanders,R.; Zhdanov,S.; Kiryutin,B.; Lipman,D.J.; Tatusova,T.; Runstadler,J. |
| EPI939699 | NP | United States | 2015-Sep-01 | EPI_ISL_252199 | A/glaucous-winged gull/Southcentral Alaska/15MB01693/2015 | Import from public-domain | Das,S.R.; Halpin,R.A.; Lin,X.; Simenauer,A.; Akopov,A.; Fedorova,N.; Puri,V.; Stockwell,T.; Amedeo,P.; Katzel,D.; Schobel,S.; Shrivastava,S.; Hill,N.; Bao,Y.; Sanders,R.; Zhdanov,S.; Kiryutin,B.; Lipman,D.J.; Tatusova,T.; Runstadler,J. |
| EPI939700 | NS | United States | 2015-Sep-01 | EPI_ISL_252199 | A/glaucous-winged gull/Southcentral Alaska/15MB01693/2015 | Import from public-domain | Das,S.R.; Halpin,R.A.; Lin,X.; Simenauer,A.; Akopov,A.; Fedorova,N.; Puri,V.; Stockwell,T.; Amedeo,P.; Katzel,D.; Schobel,S.; Shrivastava,S.; Hill,N.; Bao,Y.; Sanders,R.; Zhdanov,S.; Kiryutin,B.; Lipman,D.J.; Tatusova,T.; Runstadler,J. |
| EPI939690 | MP | United States | 2015-Aug-25 | EPI_ISL_252198 | A/glaucous-winged gull/Southcentral Alaska/15MB01645/2015 | Import from public-domain | Das,S.R.; Halpin,R.A.; Lin,X.; Simenauer,A.; Akopov,A.; Fedorova,N.; Puri,V.; Stockwell,T.; Amedeo,P.; Katzel,D.; Schobel,S.; Shrivastava,S.; Hill,N.; Bao,Y.; Sanders,R.; Zhdanov,S.; Kiryutin,B.; Lipman,D.J.; Tatusova,T.; Runstadler,J. |
| EPI939692 | NP | United States | 2015-Aug-25 | EPI_ISL_252198 | A/glaucous-winged gull/Southcentral Alaska/15MB01645/2015 | Import from public-domain | Das,S.R.; Halpin,R.A.; Lin,X.; Simenauer,A.; Akopov,A.; Fedorova,N.; Puri,V.; Stockwell,T.; Amedeo,P.; Katzel,D.; Schobel,S.; Shrivastava,S.; Hill,N.; Bao,Y.; Sanders,R.; Zhdanov,S.; Kiryutin,B.; Lipman,D.J.; Tatusova,T.; Runstadler,J. |
| EPI939693 | NS | United States | 2015-Aug-25 | EPI_ISL_252198 | A/glaucous-winged gull/Southcentral Alaska/15MB01645/2015 | Import from public-domain | Das,S.R.; Halpin,R.A.; Lin,X.; Simenauer,A.; Akopov,A.; Fedorova,N.; Puri,V.; Stockwell,T.; Amedeo,P.; Katzel,D.; Schobel,S.; Shrivastava,S.; Hill,N.; Bao,Y.; Sanders,R.; Zhdanov,S.; Kiryutin,B.; Lipman,D.J.; Tatusova,T.; Runstadler,J. |
| EPI939683 | MP | United States | 2015-Aug-25 | EPI_ISL_252197 | A/glaucous-winged gull/Southcentral Alaska/15MB01610/2015 | Import from public-domain | Das,S.R.; Halpin,R.A.; Lin,X.; Simenauer,A.; Akopov,A.; Fedorova,N.; Puri,V.; Stockwell,T.; Amedeo,P.; Katzel,D.; Schobel,S.; Shrivastava,S.; Hill,N.; Bao,Y.; Sanders,R.; Zhdanov,S.; Kiryutin,B.; Lipman,D.J.; Tatusova,T.; Runstadler,J. |
| EPI939685 | NP | United States | 2015-Aug-25 | EPI_ISL_252197 | A/glaucous-winged gull/Southcentral Alaska/15MB01610/2015 | Import from public-domain | Das,S.R.; Halpin,R.A.; Lin,X.; Simenauer,A.; Akopov,A.; Fedorova,N.; Puri,V.; Stockwell,T.; Amedeo,P.; Katzel,D.; Schobel,S.; Shrivastava,S.; Hill,N.; Bao,Y.; Sanders,R.; Zhdanov,S.; Kiryutin,B.; Lipman,D.J.; Tatusova,T.; Runstadler,J. |
| EPI939686 | NS | United States | 2015-Aug-25 | EPI_ISL_252197 | A/glaucous-winged gull/Southcentral Alaska/15MB01610/2015 | Import from public-domain | Das,S.R.; Halpin,R.A.; Lin,X.; Simenauer,A.; Akopov,A.; Fedorova,N.; Puri,V.; Stockwell,T.; Amedeo,P.; Katzel,D.; Schobel,S.; Shrivastava,S.; Hill,N.; Bao,Y.; Sanders,R.; Zhdanov,S.; Kiryutin,B.; Lipman,D.J.; Tatusova,T.; Runstadler,J. |
| EPI763019 | MP | United States | 2014-Sep-03 | EPI_ISL_222248 | A/glaucous-winged gull/Southcentral Alaska/14MB02081/2014 | Import from public-domain | Das,S.R.; Halpin,R.A.; Lin,X.; Simenauer,A.; Akopov,A.; Fedorova,N.; Puri,V.; Stockwell,T.; Amedeo,P.; Katzel,D.; Schobel,S.; Shrivastava,S.; Hill,N.; Bao,Y.; Sanders,R.; Zhdanov,S.; Kiryutin,B.; Lipman,D.J.; Tatusova,T.; Runstadler,J. |
| EPI763030 | NS | United States | 2014-Sep-03 | EPI_ISL_222248 | A/glaucous-winged gull/Southcentral Alaska/14MB02081/2014 | Import from public-domain | Das,S.R.; Halpin,R.A.; Lin,X.; Simenauer,A.; Akopov,A.; Fedorova,N.; Puri,V.; Stockwell,T.; Amedeo,P.; Katzel,D.; Schobel,S.; Shrivastava,S.; Hill,N.; Bao,Y.; Sanders,R.; Zhdanov,S.; Kiryutin,B.; Lipman,D.J.; Tatusova,T.; Runstadler,J. |
| EPI763029 | NP | United States | 2014-Sep-03 | EPI_ISL_222248 | A/glaucous-winged gull/Southcentral Alaska/14MB02081/2014 | Import from public-domain | Das,S.R.; Halpin,R.A.; Lin,X.; Simenauer,A.; Akopov,A.; Fedorova,N.; Puri,V.; Stockwell,T.; Amedeo,P.; Katzel,D.; Schobel,S.; Shrivastava,S.; Hill,N.; Bao,Y.; Sanders,R.; Zhdanov,S.; Kiryutin,B.; Lipman,D.J.; Tatusova,T.; Runstadler,J. |
| EPI763020 | MP | United States | 2014-Sep-03 | EPI_ISL_222247 | A/glaucous-winged gull/Southcentral Alaska/14MB02018/2014 | Import from public-domain | Das,S.R.; Halpin,R.A.; Lin,X.; Simenauer,A.; Akopov,A.; Fedorova,N.; Puri,V.; Stockwell,T.; Amedeo,P.; Katzel,D.; Schobel,S.; Shrivastava,S.; Hill,N.; Bao,Y.; Sanders,R.; Zhdanov,S.; Kiryutin,B.; Lipman,D.J.; Tatusova,T.; Runstadler,J. |
| EPI763022 | NP | United States | 2014-Sep-03 | EPI_ISL_222247 | A/glaucous-winged gull/Southcentral Alaska/14MB02018/2014 | Import from public-domain | Das,S.R.; Halpin,R.A.; Lin,X.; Simenauer,A.; Akopov,A.; Fedorova,N.; Puri,V.; Stockwell,T.; Amedeo,P.; Katzel,D.; Schobel,S.; Shrivastava,S.; Hill,N.; Bao,Y.; Sanders,R.; Zhdanov,S.; Kiryutin,B.; Lipman,D.J.; Tatusova,T.; Runstadler,J. |
| EPI763023 | NS | United States | 2014-Sep-03 | EPI_ISL_222247 | A/glaucous-winged gull/Southcentral Alaska/14MB02018/2014 | Import from public-domain | Das,S.R.; Halpin,R.A.; Lin,X.; Simenauer,A.; Akopov,A.; Fedorova,N.; Puri,V.; Stockwell,T.; Amedeo,P.; Katzel,D.; Schobel,S.; Shrivastava,S.; Hill,N.; Bao,Y.; Sanders,R.; Zhdanov,S.; Kiryutin,B.; Lipman,D.J.; Tatusova,T.; Runstadler,J. |
| EPI763007 | NP | United States | 2014-Aug-30 | EPI_ISL_222246 | A/glaucous-winged gull/Southcentral Alaska/14MB01886/2014 | Import from public-domain | Das,S.R.; Halpin,R.A.; Lin,X.; Simenauer,A.; Akopov,A.; Fedorova,N.; Puri,V.; Stockwell,T.; Amedeo,P.; Katzel,D.; Schobel,S.; Shrivastava,S.; Hill,N.; Bao,Y.; Sanders,R.; Zhdanov,S.; Kiryutin,B.; Lipman,D.J.; Tatusova,T.; Runstadler,J. |
| EPI763008 | NS | United States | 2014-Aug-30 | EPI_ISL_222246 | A/glaucous-winged gull/Southcentral Alaska/14MB01886/2014 | Import from public-domain | Das,S.R.; Halpin,R.A.; Lin,X.; Simenauer,A.; Akopov,A.; Fedorova,N.; Puri,V.; Stockwell,T.; Amedeo,P.; Katzel,D.; Schobel,S.; Shrivastava,S.; Hill,N.; Bao,Y.; Sanders,R.; Zhdanov,S.; Kiryutin,B.; Lipman,D.J.; Tatusova,T.; Runstadler,J. |
| EPI763004 | MP | United States | 2014-Aug-30 | EPI_ISL_222246 | A/glaucous-winged gull/Southcentral Alaska/14MB01886/2014 | Import from public-domain | Das,S.R.; Halpin,R.A.; Lin,X.; Simenauer,A.; Akopov,A.; Fedorova,N.; Puri,V.; Stockwell,T.; Amedeo,P.; Katzel,D.; Schobel,S.; Shrivastava,S.; Hill,N.; Bao,Y.; Sanders,R.; Zhdanov,S.; Kiryutin,B.; Lipman,D.J.; Tatusova,T.; Runstadler,J. |
| EPI762967 | MP | United States | 2014-Sep-01 | EPI_ISL_222245 | A/glaucous-winged gull/Southcentral Alaska/14MB01959/2014 | Import from public-domain | Das,S.R.; Halpin,R.A.; Lin,X.; Simenauer,A.; Akopov,A.; Fedorova,N.; Puri,V.; Stockwell,T.; Amedeo,P.; Katzel,D.; Schobel,S.; Shrivastava,S.; Hill,N.; Bao,Y.; Sanders,R.; Zhdanov,S.; Kiryutin,B.; Lipman,D.J.; Tatusova,T.; Runstadler,J. |
| EPI763012 | NP | United States | 2014-Sep-01 | EPI_ISL_222245 | A/glaucous-winged gull/Southcentral Alaska/14MB01959/2014 | Import from public-domain | Das,S.R.; Halpin,R.A.; Lin,X.; Simenauer,A.; Akopov,A.; Fedorova,N.; Puri,V.; Stockwell,T.; Amedeo,P.; Katzel,D.; Schobel,S.; Shrivastava,S.; Hill,N.; Bao,Y.; Sanders,R.; Zhdanov,S.; Kiryutin,B.; Lipman,D.J.; Tatusova,T.; Runstadler,J. |
| EPI763013 | NS | United States | 2014-Sep-01 | EPI_ISL_222245 | A/glaucous-winged gull/Southcentral Alaska/14MB01959/2014 | Import from public-domain | Das,S.R.; Halpin,R.A.; Lin,X.; Simenauer,A.; Akopov,A.; Fedorova,N.; Puri,V.; Stockwell,T.; Amedeo,P.; Katzel,D.; Schobel,S.; Shrivastava,S.; Hill,N.; Bao,Y.; Sanders,R.; Zhdanov,S.; Kiryutin,B.; Lipman,D.J.; Tatusova,T.; Runstadler,J. |
| EPI762995 | MP | United States | 2014-Sep-01 | EPI_ISL_222244 | A/glaucous-winged gull/Southcentral Alaska/14MB01926/2014 | Import from public-domain | Das,S.R.; Halpin,R.A.; Lin,X.; Simenauer,A.; Akopov,A.; Fedorova,N.; Puri,V.; Stockwell,T.; Amedeo,P.; Katzel,D.; Schobel,S.; Shrivastava,S.; Hill,N.; Bao,Y.; Sanders,R.; Zhdanov,S.; Kiryutin,B.; Lipman,D.J.; Tatusova,T.; Runstadler,J. |
| EPI762997 | NP | United States | 2014-Sep-01 | EPI_ISL_222244 | A/glaucous-winged gull/Southcentral Alaska/14MB01926/2014 | Import from public-domain | Das,S.R.; Halpin,R.A.; Lin,X.; Simenauer,A.; Akopov,A.; Fedorova,N.; Puri,V.; Stockwell,T.; Amedeo,P.; Katzel,D.; Schobel,S.; Shrivastava,S.; Hill,N.; Bao,Y.; Sanders,R.; Zhdanov,S.; Kiryutin,B.; Lipman,D.J.; Tatusova,T.; Runstadler,J. |
| EPI762998 | NS | United States | 2014-Sep-01 | EPI_ISL_222244 | A/glaucous-winged gull/Southcentral Alaska/14MB01926/2014 | Import from public-domain | Das,S.R.; Halpin,R.A.; Lin,X.; Simenauer,A.; Akopov,A.; Fedorova,N.; Puri,V.; Stockwell,T.; Amedeo,P.; Katzel,D.; Schobel,S.; Shrivastava,S.; Hill,N.; Bao,Y.; Sanders,R.; Zhdanov,S.; Kiryutin,B.; Lipman,D.J.; Tatusova,T.; Runstadler,J. |
| EPI762989 | MP | United States | 2014-Aug-30 | EPI_ISL_222243 | A/glaucous-winged gull/Southcentral Alaska/14MB01893/2014 | Import from public-domain | Das,S.R.; Halpin,R.A.; Lin,X.; Simenauer,A.; Akopov,A.; Fedorova,N.; Puri,V.; Stockwell,T.; Amedeo,P.; Katzel,D.; Schobel,S.; Shrivastava,S.; Hill,N.; Bao,Y.; Sanders,R.; Zhdanov,S.; Kiryutin,B.; Lipman,D.J.; Tatusova,T.; Runstadler,J. |
| EPI762990 | NP | United States | 2014-Aug-30 | EPI_ISL_222243 | A/glaucous-winged gull/Southcentral Alaska/14MB01893/2014 | Import from public-domain | Das,S.R.; Halpin,R.A.; Lin,X.; Simenauer,A.; Akopov,A.; Fedorova,N.; Puri,V.; Stockwell,T.; Amedeo,P.; Katzel,D.; Schobel,S.; Shrivastava,S.; Hill,N.; Bao,Y.; Sanders,R.; Zhdanov,S.; Kiryutin,B.; Lipman,D.J.; Tatusova,T.; Runstadler,J. |
| EPI762991 | NS | United States | 2014-Aug-30 | EPI_ISL_222243 | A/glaucous-winged gull/Southcentral Alaska/14MB01893/2014 | Import from public-domain | Das,S.R.; Halpin,R.A.; Lin,X.; Simenauer,A.; Akopov,A.; Fedorova,N.; Puri,V.; Stockwell,T.; Amedeo,P.; Katzel,D.; Schobel,S.; Shrivastava,S.; Hill,N.; Bao,Y.; Sanders,R.; Zhdanov,S.; Kiryutin,B.; Lipman,D.J.; Tatusova,T.; Runstadler,J. |
| EPI762964 | NP | United States | 2014-Aug-30 | EPI_ISL_222242 | A/glaucous-winged gull/Southcentral Alaska/14MB01884/2014 | Import from public-domain | Das,S.R.; Halpin,R.A.; Lin,X.; Simenauer,A.; Akopov,A.; Fedorova,N.; Puri,V.; Stockwell,T.; Amedeo,P.; Katzel,D.; Schobel,S.; Shrivastava,S.; Hill,N.; Bao,Y.; Sanders,R.; Zhdanov,S.; Kiryutin,B.; Lipman,D.J.; Tatusova,T.; Runstadler,J. |

| | | | | | | | |
|---|---|---|---|---|---|---|---|
| EPI762982 | MP | United States | 2014-Aug-30 | EPI_ISL_222242 | A/glaucous-winged gull/Southcentral Alaska/14MB01884/2014 | Import from public-domain | Das,S.R.; Halpin,R.A.; Lin,X.; Simenauer,A.; Akopov,A.; Fedorova,N.; Puri,V.; Stockwell,T.; Amedeo,P.; Katzel,D.; Schobel,S.; Shrivastava,S.; Hill,N.; Bao,Y.; Sanders,R.; Zhdanov,S.; Kiryutin,B.; Lipman,D.J.; Tatusova,T.; Runstadler,J. |
| EPI762984 | NS | United States | 2014-Aug-30 | EPI_ISL_222242 | A/glaucous-winged gull/Southcentral Alaska/14MB01884/2014 | Import from public-domain | Das,S.R.; Halpin,R.A.; Lin,X.; Simenauer,A.; Akopov,A.; Fedorova,N.; Puri,V.; Stockwell,T.; Amedeo,P.; Katzel,D.; Schobel,S.; Shrivastava,S.; Hill,N.; Bao,Y.; Sanders,R.; Zhdanov,S.; Kiryutin,B.; Lipman,D.J.; Tatusova,T.; Runstadler,J. |
| EPI762975 | MP | United States | 2014-Aug-30 | EPI_ISL_222241 | A/glaucous-winged gull/Southcentral Alaska/14MB01819/2014 | Import from public-domain | Das,S.R.; Halpin,R.A.; Lin,X.; Simenauer,A.; Akopov,A.; Fedorova,N.; Puri,V.; Stockwell,T.; Amedeo,P.; Katzel,D.; Schobel,S.; Shrivastava,S.; Hill,N.; Bao,Y.; Sanders,R.; Zhdanov,S.; Kiryutin,B.; Lipman,D.J.; Tatusova,T.; Runstadler,J. |
| EPI762976 | NP | United States | 2014-Aug-30 | EPI_ISL_222241 | A/glaucous-winged gull/Southcentral Alaska/14MB01819/2014 | Import from public-domain | Das,S.R.; Halpin,R.A.; Lin,X.; Simenauer,A.; Akopov,A.; Fedorova,N.; Puri,V.; Stockwell,T.; Amedeo,P.; Katzel,D.; Schobel,S.; Shrivastava,S.; Hill,N.; Bao,Y.; Sanders,R.; Zhdanov,S.; Kiryutin,B.; Lipman,D.J.; Tatusova,T.; Runstadler,J. |
| EPI762977 | NS | United States | 2014-Aug-30 | EPI_ISL_222241 | A/glaucous-winged gull/Southcentral Alaska/14MB01819/2014 | Import from public-domain | Das,S.R.; Halpin,R.A.; Lin,X.; Simenauer,A.; Akopov,A.; Fedorova,N.; Puri,V.; Stockwell,T.; Amedeo,P.; Katzel,D.; Schobel,S.; Shrivastava,S.; Hill,N.; Bao,Y.; Sanders,R.; Zhdanov,S.; Kiryutin,B.; Lipman,D.J.; Tatusova,T.; Runstadler,J. |
| EPI762970 | NS | United States | 2014-Aug-29 | EPI_ISL_222240 | A/glaucous-winged gull/Southcentral Alaska/14MB01770/2014 | Import from public-domain | Das,S.R.; Halpin,R.A.; Lin,X.; Simenauer,A.; Akopov,A.; Fedorova,N.; Puri,V.; Stockwell,T.; Amedeo,P.; Katzel,D.; Schobel,S.; Shrivastava,S.; Hill,N.; Bao,Y.; Sanders,R.; Zhdanov,S.; Kiryutin,B.; Lipman,D.J.; Tatusova,T.; Runstadler,J. |
| EPI762962 | MP | United States | 2014-Aug-29 | EPI_ISL_222240 | A/glaucous-winged gull/Southcentral Alaska/14MB01770/2014 | Import from public-domain | Das,S.R.; Halpin,R.A.; Lin,X.; Simenauer,A.; Akopov,A.; Fedorova,N.; Puri,V.; Stockwell,T.; Amedeo,P.; Katzel,D.; Schobel,S.; Shrivastava,S.; Hill,N.; Bao,Y.; Sanders,R.; Zhdanov,S.; Kiryutin,B.; Lipman,D.J.; Tatusova,T.; Runstadler,J. |
| EPI762969 | NP | United States | 2014-Aug-29 | EPI_ISL_222240 | A/glaucous-winged gull/Southcentral Alaska/14MB01770/2014 | Import from public-domain | Das,S.R.; Halpin,R.A.; Lin,X.; Simenauer,A.; Akopov,A.; Fedorova,N.; Puri,V.; Stockwell,T.; Amedeo,P.; Katzel,D.; Schobel,S.; Shrivastava,S.; Hill,N.; Bao,Y.; Sanders,R.; Zhdanov,S.; Kiryutin,B.; Lipman,D.J.; Tatusova,T.; Runstadler,J. |
| EPI762904 | MP | United States | 2014-Aug-29 | EPI_ISL_222219 | A/glaucous-winged gull/Southcentral Alaska/14MB01705/2014 | Import from public-domain | Das,S.R.; Halpin,R.A.; Lin,X.; Simenauer,A.; Akopov,A.; Fedorova,N.; Puri,V.; Stockwell,T.; Amedeo,P.; Katzel,D.; Schobel,S.; Shrivastava,S.; Hill,N.; Bao,Y.; Sanders,R.; Zhdanov,S.; Kiryutin,B.; Lipman,D.J.; Tatusova,T.; Runstadler,J. |
| EPI762911 | NP | United States | 2014-Aug-29 | EPI_ISL_222219 | A/glaucous-winged gull/Southcentral Alaska/14MB01705/2014 | Import from public-domain | Das,S.R.; Halpin,R.A.; Lin,X.; Simenauer,A.; Akopov,A.; Fedorova,N.; Puri,V.; Stockwell,T.; Amedeo,P.; Katzel,D.; Schobel,S.; Shrivastava,S.; Hill,N.; Bao,Y.; Sanders,R.; Zhdanov,S.; Kiryutin,B.; Lipman,D.J.; Tatusova,T.; Runstadler,J. |
| EPI762914 | NS | United States | 2014-Aug-29 | EPI_ISL_222219 | A/glaucous-winged gull/Southcentral Alaska/14MB01705/2014 | Import from public-domain | Das,S.R.; Halpin,R.A.; Lin,X.; Simenauer,A.; Akopov,A.; Fedorova,N.; Puri,V.; Stockwell,T.; Amedeo,P.; Katzel,D.; Schobel,S.; Shrivastava,S.; Hill,N.; Bao,Y.; Sanders,R.; Zhdanov,S.; Kiryutin,B.; Lipman,D.J.; Tatusova,T.; Runstadler,J. |
| EPI762880 | MP | United States | 2014-Aug-25 | EPI_ISL_222213 | A/glaucous-winged gull/Southcentral Alaska/14MB01615/2014 | Import from public-domain | Das,S.R.; Halpin,R.A.; Lin,X.; Simenauer,A.; Akopov,A.; Fedorova,N.; Puri,V.; Stockwell,T.; Amedeo,P.; Katzel,D.; Schobel,S.; Shrivastava,S.; Hill,N.; Bao,Y.; Sanders,R.; Zhdanov,S.; Kiryutin,B.; Lipman,D.J.; Tatusova,T.; Runstadler,J. |
| EPI762885 | NP | United States | 2014-Aug-25 | EPI_ISL_222213 | A/glaucous-winged gull/Southcentral Alaska/14MB01615/2014 | Import from public-domain | Das,S.R.; Halpin,R.A.; Lin,X.; Simenauer,A.; Akopov,A.; Fedorova,N.; Puri,V.; Stockwell,T.; Amedeo,P.; Katzel,D.; Schobel,S.; Shrivastava,S.; Hill,N.; Bao,Y.; Sanders,R.; Zhdanov,S.; Kiryutin,B.; Lipman,D.J.; Tatusova,T.; Runstadler,J. |
| EPI762890 | NS | United States | 2014-Aug-25 | EPI_ISL_222213 | A/glaucous-winged gull/Southcentral Alaska/14MB01615/2014 | Import from public-domain | Das,S.R.; Halpin,R.A.; Lin,X.; Simenauer,A.; Akopov,A.; Fedorova,N.; Puri,V.; Stockwell,T.; Amedeo,P.; Katzel,D.; Schobel,S.; Shrivastava,S.; Hill,N.; Bao,Y.; Sanders,R.; Zhdanov,S.; Kiryutin,B.; Lipman,D.J.; Tatusova,T.; Runstadler,J. |
| EPI762697 | NS | United States | 2014-Aug-20 | EPI_ISL_222208 | A/glaucous-winged gull/Southcentral Alaska/14MB01438/2014 | Import from public-domain | Das,S.R.; Halpin,R.A.; Lin,X.; Simenauer,A.; Akopov,A.; Fedorova,N.; Puri,V.; Stockwell,T.; Amedeo,P.; Katzel,D.; Schobel,S.; Shrivastava,S.; Hill,N.; Bao,Y.; Sanders,R.; Zhdanov,S.; Kiryutin,B.; Lipman,D.J.; Tatusova,T.; Runstadler,J. |
| EPI762856 | MP | United States | 2014-Aug-20 | EPI_ISL_222208 | A/glaucous-winged gull/Southcentral Alaska/14MB01438/2014 | Import from public-domain | Das,S.R.; Halpin,R.A.; Lin,X.; Simenauer,A.; Akopov,A.; Fedorova,N.; Puri,V.; Stockwell,T.; Amedeo,P.; Katzel,D.; Schobel,S.; Shrivastava,S.; Hill,N.; Bao,Y.; Sanders,R.; Zhdanov,S.; Kiryutin,B.; Lipman,D.J.; Tatusova,T.; Runstadler,J. |
| EPI762863 | NP | United States | 2014-Aug-20 | EPI_ISL_222208 | A/glaucous-winged gull/Southcentral Alaska/14MB01438/2014 | Import from public-domain | Das,S.R.; Halpin,R.A.; Lin,X.; Simenauer,A.; Akopov,A.; Fedorova,N.; Puri,V.; Stockwell,T.; Amedeo,P.; Katzel,D.; Schobel,S.; Shrivastava,S.; Hill,N.; Bao,Y.; Sanders,R.; Zhdanov,S.; Kiryutin,B.; Lipman,D.J.; Tatusova,T.; Runstadler,J. |
| EPI762663 | MP | United States | 2014-Aug-20 | EPI_ISL_222203 | A/glaucous-winged gull/Southcentral Alaska/14MB01422/2014 | Import from public-domain | Das,S.R.; Halpin,R.A.; Lin,X.; Simenauer,A.; Akopov,A.; Fedorova,N.; Puri,V.; Stockwell,T.; Amedeo,P.; Katzel,D.; Schobel,S.; Shrivastava,S.; Hill,N.; Bao,Y.; Sanders,R.; Zhdanov,S.; Kiryutin,B.; Lipman,D.J.; Tatusova,T.; Runstadler,J. |
| EPI762837 | NP | United States | 2014-Aug-20 | EPI_ISL_222203 | A/glaucous-winged gull/Southcentral Alaska/14MB01422/2014 | Import from public-domain | Das,S.R.; Halpin,R.A.; Lin,X.; Simenauer,A.; Akopov,A.; Fedorova,N.; Puri,V.; Stockwell,T.; Amedeo,P.; Katzel,D.; Schobel,S.; Shrivastava,S.; Hill,N.; Bao,Y.; Sanders,R.; Zhdanov,S.; Kiryutin,B.; Lipman,D.J.; Tatusova,T.; Runstadler,J. |
| EPI762841 | NS | United States | 2014-Aug-20 | EPI_ISL_222203 | A/glaucous-winged gull/Southcentral Alaska/14MB01422/2014 | Import from public-domain | Das,S.R.; Halpin,R.A.; Lin,X.; Simenauer,A.; Akopov,A.; Fedorova,N.; Puri,V.; Stockwell,T.; Amedeo,P.; Katzel,D.; Schobel,S.; Shrivastava,S.; Hill,N.; Bao,Y.; Sanders,R.; Zhdanov,S.; Kiryutin,B.; Lipman,D.J.; Tatusova,T.; Runstadler,J. |
| EPI762630 | MP | United States | 2014-Aug-20 | EPI_ISL_222198 | A/glaucous-winged gull/Southcentral Alaska/14MB01418/2014 | Import from public-domain | Das,S.R.; Halpin,R.A.; Lin,X.; Simenauer,A.; Akopov,A.; Fedorova,N.; Puri,V.; Stockwell,T.; Amedeo,P.; Katzel,D.; Schobel,S.; Shrivastava,S.; Hill,N.; Bao,Y.; Sanders,R.; Zhdanov,S.; Kiryutin,B.; Lipman,D.J.; Tatusova,T.; Runstadler,J. |
| EPI762814 | NP | United States | 2014-Aug-20 | EPI_ISL_222198 | A/glaucous-winged gull/Southcentral Alaska/14MB01418/2014 | Import from public-domain | Das,S.R.; Halpin,R.A.; Lin,X.; Simenauer,A.; Akopov,A.; Fedorova,N.; Puri,V.; Stockwell,T.; Amedeo,P.; Katzel,D.; Schobel,S.; Shrivastava,S.; Hill,N.; Bao,Y.; Sanders,R.; Zhdanov,S.; Kiryutin,B.; Lipman,D.J.; Tatusova,T.; Runstadler,J. |
| EPI762818 | NS | United States | 2014-Aug-20 | EPI_ISL_222198 | A/glaucous-winged gull/Southcentral Alaska/14MB01418/2014 | Import from public-domain | Das,S.R.; Halpin,R.A.; Lin,X.; Simenauer,A.; Akopov,A.; Fedorova,N.; Puri,V.; Stockwell,T.; Amedeo,P.; Katzel,D.; Schobel,S.; Shrivastava,S.; Hill,N.; Bao,Y.; Sanders,R.; Zhdanov,S.; Kiryutin,B.; Lipman,D.J.; Tatusova,T.; Runstadler,J. |
| EPI762786 | MP | United States | 2014-Aug-20 | EPI_ISL_222193 | A/glaucous-winged gull/Southcentral Alaska/14MB01417/2014 | Import from public-domain | Das,S.R.; Halpin,R.A.; Lin,X.; Simenauer,A.; Akopov,A.; Fedorova,N.; Puri,V.; Stockwell,T.; Amedeo,P.; Katzel,D.; Schobel,S.; Shrivastava,S.; Hill,N.; Bao,Y.; Sanders,R.; Zhdanov,S.; Kiryutin,B.; Lipman,D.J.; Tatusova,T.; Runstadler,J. |
| EPI762791 | NP | United States | 2014-Aug-20 | EPI_ISL_222193 | A/glaucous-winged gull/Southcentral Alaska/14MB01417/2014 | Import from public-domain | Das,S.R.; Halpin,R.A.; Lin,X.; Simenauer,A.; Akopov,A.; Fedorova,N.; Puri,V.; Stockwell,T.; Amedeo,P.; Katzel,D.; Schobel,S.; Shrivastava,S.; Hill,N.; Bao,Y.; Sanders,R.; Zhdanov,S.; Kiryutin,B.; Lipman,D.J.; Tatusova,T.; Runstadler,J. |
| EPI762795 | NS | United States | 2014-Aug-20 | EPI_ISL_222193 | A/glaucous-winged gull/Southcentral Alaska/14MB01417/2014 | Import from public-domain | Das,S.R.; Halpin,R.A.; Lin,X.; Simenauer,A.; Akopov,A.; Fedorova,N.; Puri,V.; Stockwell,T.; Amedeo,P.; Katzel,D.; Schobel,S.; Shrivastava,S.; Hill,N.; Bao,Y.; Sanders,R.; Zhdanov,S.; Kiryutin,B.; Lipman,D.J.; Tatusova,T.; Runstadler,J. |
| EPI762451 | NP | United States | 2014-Aug-19 | EPI_ISL_222172 | A/glaucous-winged gull/Southcentral Alaska/14MB01392/2014 | Import from public-domain | Das,S.R.; Halpin,R.A.; Lin,X.; Simenauer,A.; Akopov,A.; Fedorova,N.; Puri,V.; Stockwell,T.; Amedeo,P.; Katzel,D.; Schobel,S.; Shrivastava,S.; Hill,N.; Bao,Y.; Sanders,R.; Zhdanov,S.; Kiryutin,B.; Lipman,D.J.; Tatusova,T.; Runstadler,J. |
| EPI762452 | NS | United States | 2014-Aug-19 | EPI_ISL_222172 | A/glaucous-winged gull/Southcentral Alaska/14MB01392/2014 | Import from public-domain | Das,S.R.; Halpin,R.A.; Lin,X.; Simenauer,A.; Akopov,A.; Fedorova,N.; Puri,V.; Stockwell,T.; Amedeo,P.; Katzel,D.; Schobel,S.; Shrivastava,S.; Hill,N.; Bao,Y.; Sanders,R.; Zhdanov,S.; Kiryutin,B.; Lipman,D.J.; Tatusova,T.; Runstadler,J. |
| EPI762413 | MP | United States | 2014-Aug-19 | EPI_ISL_222172 | A/glaucous-winged gull/Southcentral Alaska/14MB01392/2014 | Import from public-domain | Das,S.R.; Halpin,R.A.; Lin,X.; Simenauer,A.; Akopov,A.; Fedorova,N.; Puri,V.; Stockwell,T.; Amedeo,P.; Katzel,D.; Schobel,S.; Shrivastava,S.; Hill,N.; Bao,Y.; Sanders,R.; Zhdanov,S.; Kiryutin,B.; Lipman,D.J.; Tatusova,T.; Runstadler,J. |
| EPI762443 | MP | United States | 2014-Aug-19 | EPI_ISL_222171 | A/glaucous-winged gull/Southcentral Alaska/14MB01383/2014 | Import from public-domain | Das,S.R.; Halpin,R.A.; Lin,X.; Simenauer,A.; Akopov,A.; Fedorova,N.; Puri,V.; Stockwell,T.; Amedeo,P.; Katzel,D.; Schobel,S.; Shrivastava,S.; Hill,N.; Bao,Y.; Sanders,R.; Zhdanov,S.; Kiryutin,B.; Lipman,D.J.; Tatusova,T.; Runstadler,J. |
| EPI762444 | NP | United States | 2014-Aug-19 | EPI_ISL_222171 | A/glaucous-winged gull/Southcentral Alaska/14MB01383/2014 | Import from public-domain | Das,S.R.; Halpin,R.A.; Lin,X.; Simenauer,A.; Akopov,A.; Fedorova,N.; Puri,V.; Stockwell,T.; Amedeo,P.; Katzel,D.; Schobel,S.; Shrivastava,S.; Hill,N.; Bao,Y.; Sanders,R.; Zhdanov,S.; Kiryutin,B.; Lipman,D.J.; Tatusova,T.; Runstadler,J. |
| EPI762445 | NS | United States | 2014-Aug-19 | EPI_ISL_222171 | A/glaucous-winged gull/Southcentral Alaska/14MB01383/2014 | Import from public-domain | Das,S.R.; Halpin,R.A.; Lin,X.; Simenauer,A.; Akopov,A.; Fedorova,N.; Puri,V.; Stockwell,T.; Amedeo,P.; Katzel,D.; Schobel,S.; Shrivastava,S.; Hill,N.; Bao,Y.; Sanders,R.; Zhdanov,S.; Kiryutin,B.; Lipman,D.J.; Tatusova,T.; Runstadler,J. |
| EPI762436 | MP | United States | 2014-Aug-19 | EPI_ISL_222170 | A/glaucous-winged gull/Southcentral Alaska/14MB01318/2014 | Import from public-domain | Das,S.R.; Halpin,R.A.; Lin,X.; Simenauer,A.; Akopov,A.; Fedorova,N.; Puri,V.; Stockwell,T.; Amedeo,P.; Katzel,D.; Schobel,S.; Shrivastava,S.; Hill,N.; Bao,Y.; Sanders,R.; Zhdanov,S.; Kiryutin,B.; Lipman,D.J.; Tatusova,T.; Runstadler,J. |
| EPI762438 | NS | United States | 2014-Aug-19 | EPI_ISL_222170 | A/glaucous-winged gull/Southcentral Alaska/14MB01318/2014 | Import from public-domain | Das,S.R.; Halpin,R.A.; Lin,X.; Simenauer,A.; Akopov,A.; Fedorova,N.; Puri,V.; Stockwell,T.; Amedeo,P.; Katzel,D.; Schobel,S.; Shrivastava,S.; Hill,N.; Bao,Y.; Sanders,R.; Zhdanov,S.; Kiryutin,B.; Lipman,D.J.; Tatusova,T.; Runstadler,J. |
| EPI762411 | NP | United States | 2014-Aug-19 | EPI_ISL_222170 | A/glaucous-winged gull/Southcentral Alaska/14MB01318/2014 | Import from public-domain | Das,S.R.; Halpin,R.A.; Lin,X.; Simenauer,A.; Akopov,A.; Fedorova,N.; Puri,V.; Stockwell,T.; Amedeo,P.; Katzel,D.; Schobel,S.; Shrivastava,S.; Hill,N.; Bao,Y.; Sanders,R.; Zhdanov,S.; Kiryutin,B.; Lipman,D.J.; Tatusova,T.; Runstadler,J. |

| | | | | | | | | |
|---|---|---|---|---|---|---|---|---|
| EPI762428 | MP | United States | 2014-Aug-19 | EPI_ISL_222169 | A/glaucous-winged gull/Southcentral Alaska/14MB01306/2014 | | Import from public-domain | Das,S.R.; Halpin,R.A.; Lin,X.; Simenauer,A.; Akopov,A.; Fedorova,N.; Puri,V.; Stockwell,T.; Amedeo,P.; Katzel,D.; Schobel,S.; Shrivastava,S.; Hill,N.; Bao,Y.; Sanders,R.; Zhdanov,S.; Kiryutin,B.; Lipman,D.J.; Tatusova,T.; Runstadler,J. |
| EPI762430 | NP | United States | 2014-Aug-19 | EPI_ISL_222169 | A/glaucous-winged gull/Southcentral Alaska/14MB01306/2014 | | Import from public-domain | Das,S.R.; Halpin,R.A.; Lin,X.; Simenauer,A.; Akopov,A.; Fedorova,N.; Puri,V.; Stockwell,T.; Amedeo,P.; Katzel,D.; Schobel,S.; Shrivastava,S.; Hill,N.; Bao,Y.; Sanders,R.; Zhdanov,S.; Kiryutin,B.; Lipman,D.J.; Tatusova,T.; Runstadler,J. |
| EPI762431 | NS | United States | 2014-Aug-19 | EPI_ISL_222169 | A/glaucous-winged gull/Southcentral Alaska/14MB01306/2014 | | Import from public-domain | Das,S.R.; Halpin,R.A.; Lin,X.; Simenauer,A.; Akopov,A.; Fedorova,N.; Puri,V.; Stockwell,T.; Amedeo,P.; Katzel,D.; Schobel,S.; Shrivastava,S.; Hill,N.; Bao,Y.; Sanders,R.; Zhdanov,S.; Kiryutin,B.; Lipman,D.J.; Tatusova,T.; Runstadler,J. |
| EPI762423 | NP | United States | 2014-Aug-12 | EPI_ISL_222168 | A/glaucous-winged gull/Southcentral Alaska/14MB00623/2014 | | Import from public-domain | Das,S.R.; Halpin,R.A.; Lin,X.; Simenauer,A.; Akopov,A.; Fedorova,N.; Puri,V.; Stockwell,T.; Amedeo,P.; Katzel,D.; Schobel,S.; Shrivastava,S.; Hill,N.; Bao,Y.; Sanders,R.; Zhdanov,S.; Kiryutin,B.; Lipman,D.J.; Tatusova,T.; Runstadler,J. |
| EPI762424 | NS | United States | 2014-Aug-12 | EPI_ISL_222168 | A/glaucous-winged gull/Southcentral Alaska/14MB00623/2014 | | Import from public-domain | Das,S.R.; Halpin,R.A.; Lin,X.; Simenauer,A.; Akopov,A.; Fedorova,N.; Puri,V.; Stockwell,T.; Amedeo,P.; Katzel,D.; Schobel,S.; Shrivastava,S.; Hill,N.; Bao,Y.; Sanders,R.; Zhdanov,S.; Kiryutin,B.; Lipman,D.J.; Tatusova,T.; Runstadler,J. |
| EPI762421 | MP | United States | 2014-Aug-12 | EPI_ISL_222168 | A/glaucous-winged gull/Southcentral Alaska/14MB00623/2014 | | Import from public-domain | Das,S.R.; Halpin,R.A.; Lin,X.; Simenauer,A.; Akopov,A.; Fedorova,N.; Puri,V.; Stockwell,T.; Amedeo,P.; Katzel,D.; Schobel,S.; Shrivastava,S.; Hill,N.; Bao,Y.; Sanders,R.; Zhdanov,S.; Kiryutin,B.; Lipman,D.J.; Tatusova,T.; Runstadler,J. |
| EPI762232 | MP | United States | 2014-Aug-19 | EPI_ISL_222145 | A/glaucous-winged gull/Southcentral Alaska/14MB01336/2014 | | Import from public-domain | Das,S.R.; Halpin,R.A.; Lin,X.; Simenauer,A.; Akopov,A.; Fedorova,N.; Puri,V.; Stockwell,T.; Amedeo,P.; Katzel,D.; Schobel,S.; Shrivastava,S.; Hill,N.; Bao,Y.; Sanders,R.; Zhdanov,S.; Kiryutin,B.; Lipman,D.J.; Tatusova,T.; Runstadler,J. |
| EPI762234 | NP | United States | 2014-Aug-19 | EPI_ISL_222145 | A/glaucous-winged gull/Southcentral Alaska/14MB01336/2014 | | Import from public-domain | Das,S.R.; Halpin,R.A.; Lin,X.; Simenauer,A.; Akopov,A.; Fedorova,N.; Puri,V.; Stockwell,T.; Amedeo,P.; Katzel,D.; Schobel,S.; Shrivastava,S.; Hill,N.; Bao,Y.; Sanders,R.; Zhdanov,S.; Kiryutin,B.; Lipman,D.J.; Tatusova,T.; Runstadler,J. |
| EPI762235 | NS | United States | 2014-Aug-19 | EPI_ISL_222145 | A/glaucous-winged gull/Southcentral Alaska/14MB01336/2014 | | Import from public-domain | Das,S.R.; Halpin,R.A.; Lin,X.; Simenauer,A.; Akopov,A.; Fedorova,N.; Puri,V.; Stockwell,T.; Amedeo,P.; Katzel,D.; Schobel,S.; Shrivastava,S.; Hill,N.; Bao,Y.; Sanders,R.; Zhdanov,S.; Kiryutin,B.; Lipman,D.J.; Tatusova,T.; Runstadler,J. |
| EPI2403215 | MP | United States | 2022-May-31 | EPI_ISL_16920547 | A/Iceland gull/Delaware/831/2022 | | Import from public-domain | Direct Submission |
| EPI2403216 | NP | United States | 2022-May-31 | EPI_ISL_16920547 | A/Iceland gull/Delaware/831/2022 | | Import from public-domain | Direct Submission |
| EPI2403213 | NS | United States | 2022-May-31 | EPI_ISL_16920547 | A/Iceland gull/Delaware/831/2022 | | Import from public-domain | Direct Submission |
| EPI2401522 | NS | United States | 2022-May-31 | EPI_ISL_16919128 | A/Iceland gull/Delaware/819/2022 | | Import from public-domain | Direct Submission |
| EPI2401524 | MP | United States | 2022-May-31 | EPI_ISL_16919128 | A/Iceland gull/Delaware/819/2022 | | Import from public-domain | Direct Submission |
| EPI2401526 | NP | United States | 2022-May-31 | EPI_ISL_16919128 | A/Iceland gull/Delaware/819/2022 | | Import from public-domain | Direct Submission |
| EPI2401345 | NP | United States | 2022-May-31 | EPI_ISL_16919094 | A/Great black-backed gull/Delaware/844/2022 | | Import from public-domain | Direct Submission |
| EPI2401348 | MP | United States | 2022-May-31 | EPI_ISL_16919094 | A/Great black-backed gull/Delaware/844/2022 | | Import from public-domain | Direct Submission |
| EPI2401346 | NS | United States | 2022-May-31 | EPI_ISL_16919094 | A/Great black-backed gull/Delaware/844/2022 | | Import from public-domain | Direct Submission |
| EPI2147949 | MP | United States | 2021-Jun-01 | EPI_ISL_14777730 | A/Great Black-Backed Gull/Delaware/610/2021 | | Import from public-domain | Direct Submission |
| EPI2147950 | NP | United States | 2021-Jun-01 | EPI_ISL_14777730 | A/Great Black-Backed Gull/Delaware/610/2021 | | Import from public-domain | Direct Submission |
| EPI2147951 | NS | United States | 2021-Jun-01 | EPI_ISL_14777730 | A/Great Black-Backed Gull/Delaware/610/2021 | | Import from public-domain | Direct Submission |
| EPI1604437 | MP | China | 2018-Oct-23 | EPI_ISL_393957 | A/great black-headed gull/Ningxia/1/2018(H16N3) | Harbin Veterinary Research Institute (CAAS) | Harbin Veterinary Research Institute (CAAS) | Yulei,Li;Minghui,Li;Jingman,Tian;Yanbing,Li |
| EPI1604435 | NP | China | 2018-Oct-23 | EPI_ISL_393957 | A/great black-headed gull/Ningxia/1/2018(H16N3) | Harbin Veterinary Research Institute (CAAS) | Harbin Veterinary Research Institute (CAAS) | Yulei,Li;Minghui,Li;Jingman,Tian;Yanbing,Li |
| EPI1604441 | NS | China | 2018-Oct-23 | EPI_ISL_393957 | A/great black-headed gull/Ningxia/1/2018(H16N3) | Harbin Veterinary Research Institute (CAAS) | Harbin Veterinary Research Institute (CAAS) | Yulei,Li;Minghui,Li;Jingman,Tian;Yanbing,Li |
| EPI1604479 | MP | China | 2018-Oct-23 | EPI_ISL_393962 | A/great black-headed gull/Ningxia/2/2018(H16N3) | Harbin Veterinary Research Institute (CAAS) | Harbin Veterinary Research Institute (CAAS) | Yulei,Li;Minghui,Li;Jingman,Tian;Yanbing,Li |
| EPI1604480 | NS | China | 2018-Oct-23 | EPI_ISL_393962 | A/great black-headed gull/Ningxia/2/2018(H16N3) | Harbin Veterinary Research Institute (CAAS) | Harbin Veterinary Research Institute (CAAS) | Yulei,Li;Minghui,Li;Jingman,Tian;Yanbing,Li |
| EPI1604477 | NP | China | 2018-Oct-23 | EPI_ISL_393962 | A/great black-headed gull/Ningxia/2/2018(H16N3) | Harbin Veterinary Research Institute (CAAS) | Harbin Veterinary Research Institute (CAAS) | Yulei,Li;Minghui,Li;Jingman,Tian;Yanbing,Li |
| EPI961526 | NS | Russian Federation | 2016-Oct-01 | EPI_ISL_256303 | A/environment/Kamchatka/117/2016 | | State Research Center of Virology and Biotechnology (VECTOR) | Ivan,Susloparov; Natalya,Goncharova; Natalya,Kolosova; Vasiliy,Marchenko; Alexander,Ryzhikov |
| EPI961508 | MP | Russian Federation | 2016-Oct-01 | EPI_ISL_256303 | A/environment/Kamchatka/117/2016 | | State Research Center of Virology and Biotechnology (VECTOR) | Ivan,Susloparov; Natalya,Goncharova; Natalya,Kolosova; Vasiliy,Marchenko; Alexander,Ryzhikov |
| EPI961491 | NP | Russian Federation | 2016-Oct-01 | EPI_ISL_256303 | A/environment/Kamchatka/117/2016 | | State Research Center of Virology and Biotechnology (VECTOR) | Ivan,Susloparov; Natalya,Goncharova; Natalya,Kolosova; Vasiliy,Marchenko; Alexander,Ryzhikov |
| EPI1298398 | NS | Chile | 2016-Apr-27 | EPI_ISL_326953 | A/dolphin gull/Chile/C10783/2016 | | Import from public-domain | Jimenez-Bluhm,P.; Hamilton,C.; van Bakel,H.; Kriti,D.; Schultz-Cherry,S. |
| EPI1298400 | NP | Chile | 2016-Apr-27 | EPI_ISL_326953 | A/dolphin gull/Chile/C10783/2016 | | Import from public-domain | Jimenez-Bluhm,P.; Hamilton,C.; van Bakel,H.; Kriti,D.; Schultz-Cherry,S. |
| EPI1298403 | MP | Chile | 2016-Apr-27 | EPI_ISL_326953 | A/dolphin gull/Chile/C10783/2016 | | Import from public-domain | Jimenez-Bluhm,P.; Hamilton,C.; van Bakel,H.; Kriti,D.; Schultz-Cherry,S. |
| EPI1301302 | NP | Chile | 2016-Dec-13 | EPI_ISL_327627 | A/black skimmer/Chile/C20077/2016 | | Import from public-domain | Bravo-Vasquez,N.; Bahl,J.; Hamilton-West,C.; Schultz-Cherry,S. |
| EPI1301284 | NS | Chile | 2016-Dec-13 | EPI_ISL_327627 | A/black skimmer/Chile/C20077/2016 | | Import from public-domain | Bravo-Vasquez,N.; Bahl,J.; Hamilton-West,C.; Schultz-Cherry,S. |

| | | | | | | | | |
|---|---|---|---|---|---|---|---|---|
| EPI1301292 | MP | Chile | 2016-Dec-13 | EPI_ISL_327627 | A/black skimmer/Chile/C20077/2016 | | Import from public-domain | Bravo-Vasquez,N.; Bahl,J.; Hamilton-West,C.; Schultz-Cherry,S. |
| EPI1301296 | MP | Chile | 2016-Dec-13 | EPI_ISL_327626 | A/black skimmer/Chile/C20084/2016 | | Import from public-domain | Bravo-Vasquez,N.; Bahl,J.; Hamilton-West,C.; Schultz-Cherry,S. |
| EPI1301303 | NS | Chile | 2016-Dec-13 | EPI_ISL_327626 | A/black skimmer/Chile/C20084/2016 | | Import from public-domain | Bravo-Vasquez,N.; Bahl,J.; Hamilton-West,C.; Schultz-Cherry,S. |
| EPI1301272 | NP | Chile | 2016-Dec-13 | EPI_ISL_327626 | A/black skimmer/Chile/C20084/2016 | | Import from public-domain | Bravo-Vasquez,N.; Bahl,J.; Hamilton-West,C.; Schultz-Cherry,S. |
| EPI1301294 | NS | Chile | 2016-Dec-13 | EPI_ISL_327625 | A/black skimmer/Chile/C20100/2016 | | Import from public-domain | Bravo-Vasquez,N.; Bahl,J.; Hamilton-West,C.; Schultz-Cherry,S. |
| EPI1301300 | MP | Chile | 2016-Dec-13 | EPI_ISL_327625 | A/black skimmer/Chile/C20100/2016 | | Import from public-domain | Bravo-Vasquez,N.; Bahl,J.; Hamilton-West,C.; Schultz-Cherry,S. |
| EPI1301270 | NP | Chile | 2016-Dec-13 | EPI_ISL_327625 | A/black skimmer/Chile/C20100/2016 | | Import from public-domain | Bravo-Vasquez,N.; Bahl,J.; Hamilton-West,C.; Schultz-Cherry,S. |
| EPI1301274 | NP | Chile | 2016-Dec-13 | EPI_ISL_327624 | A/black skimmer/Chile/C20057/2016 | | Import from public-domain | Bravo-Vasquez,N.; Bahl,J.; Hamilton-West,C.; Schultz-Cherry,S. |
| EPI1301282 | NS | Chile | 2016-Dec-13 | EPI_ISL_327624 | A/black skimmer/Chile/C20057/2016 | | Import from public-domain | Bravo-Vasquez,N.; Bahl,J.; Hamilton-West,C.; Schultz-Cherry,S. |
| EPI1301288 | MP | Chile | 2016-Dec-13 | EPI_ISL_327624 | A/black skimmer/Chile/C20057/2016 | | Import from public-domain | Bravo-Vasquez,N.; Bahl,J.; Hamilton-West,C.; Schultz-Cherry,S. |
| EPI1301264 | MP | Chile | 2016-Dec-13 | EPI_ISL_327623 | A/black skimmer/Chile/C20108/2016 | | Import from public-domain | Bravo-Vasquez,N.; Bahl,J.; Hamilton-West,C.; Schultz-Cherry,S. |
| EPI1301269 | NP | Chile | 2016-Dec-13 | EPI_ISL_327623 | A/black skimmer/Chile/C20108/2016 | | Import from public-domain | Bravo-Vasquez,N.; Bahl,J.; Hamilton-West,C.; Schultz-Cherry,S. |
| EPI1301281 | NS | Chile | 2016-Dec-13 | EPI_ISL_327623 | A/black skimmer/Chile/C20108/2016 | | Import from public-domain | Bravo-Vasquez,N.; Bahl,J.; Hamilton-West,C.; Schultz-Cherry,S. |
| EPI1301250 | NP | Chile | 2016-Dec-13 | EPI_ISL_327622 | A/black skimmer/Chile/C20140/2016 | | Import from public-domain | Bravo-Vasquez,N.; Bahl,J.; Hamilton-West,C.; Schultz-Cherry,S. |
| EPI1301256 | MP | Chile | 2016-Dec-13 | EPI_ISL_327622 | A/black skimmer/Chile/C20140/2016 | | Import from public-domain | Bravo-Vasquez,N.; Bahl,J.; Hamilton-West,C.; Schultz-Cherry,S. |
| EPI1301259 | NS | Chile | 2016-Dec-13 | EPI_ISL_327622 | A/black skimmer/Chile/C20140/2016 | | Import from public-domain | Bravo-Vasquez,N.; Bahl,J.; Hamilton-West,C.; Schultz-Cherry,S. |
| EPI1301243 | MP | Chile | 2016-Dec-13 | EPI_ISL_327621 | A/black skimmer/Chile/C20123/2016 | | Import from public-domain | Bravo-Vasquez,N.; Bahl,J.; Hamilton-West,C.; Schultz-Cherry,S. |
| EPI1301254 | NS | Chile | 2016-Dec-13 | EPI_ISL_327621 | A/black skimmer/Chile/C20123/2016 | | Import from public-domain | Bravo-Vasquez,N.; Bahl,J.; Hamilton-West,C.; Schultz-Cherry,S. |
| EPI1301236 | NP | Chile | 2016-Dec-13 | EPI_ISL_327621 | A/black skimmer/Chile/C20123/2016 | | Import from public-domain | Bravo-Vasquez,N.; Bahl,J.; Hamilton-West,C.; Schultz-Cherry,S. |
| EPI1301238 | NP | Chile | 2016-Dec-13 | EPI_ISL_327620 | A/black skimmer/Chile/C20124/2016 | | Import from public-domain | Bravo-Vasquez,N.; Bahl,J.; Hamilton-West,C.; Schultz-Cherry,S. |
| EPI1301261 | MP | Chile | 2016-Dec-13 | EPI_ISL_327620 | A/black skimmer/Chile/C20124/2016 | | Import from public-domain | Bravo-Vasquez,N.; Bahl,J.; Hamilton-West,C.; Schultz-Cherry,S. |
| EPI1301235 | NS | Chile | 2016-Dec-13 | EPI_ISL_327620 | A/black skimmer/Chile/C20124/2016 | | Import from public-domain | Bravo-Vasquez,N.; Bahl,J.; Hamilton-West,C.; Schultz-Cherry,S. |
| EPI1301242 | NP | Chile | 2016-Dec-13 | EPI_ISL_327619 | A/black skimmer/Chile/C20142/2016 | | Import from public-domain | Bravo-Vasquez,N.; Bahl,J.; Hamilton-West,C.; Schultz-Cherry,S. |
| EPI1301263 | NS | Chile | 2016-Dec-13 | EPI_ISL_327619 | A/black skimmer/Chile/C20142/2016 | | Import from public-domain | Bravo-Vasquez,N.; Bahl,J.; Hamilton-West,C.; Schultz-Cherry,S. |
| EPI1301267 | MP | Chile | 2016-Dec-13 | EPI_ISL_327619 | A/black skimmer/Chile/C20142/2016 | | Import from public-domain | Bravo-Vasquez,N.; Bahl,J.; Hamilton-West,C.; Schultz-Cherry,S. |
| EPI1301205 | MP | Chile | 2016-Dec-13 | EPI_ISL_327611 | A/elegant tern/Chile/C20093/2016 | | Import from public-domain | Bravo-Vasquez,N.; Bahl,J.; Hamilton-West,C.; Schultz-Cherry,S. |
| EPI1301206 | NS | Chile | 2016-Dec-13 | EPI_ISL_327611 | A/elegant tern/Chile/C20093/2016 | | Import from public-domain | Bravo-Vasquez,N.; Bahl,J.; Hamilton-West,C.; Schultz-Cherry,S. |
| EPI1301209 | NP | Chile | 2016-Dec-13 | EPI_ISL_327611 | A/elegant tern/Chile/C20093/2016 | | Import from public-domain | Bravo-Vasquez,N.; Bahl,J.; Hamilton-West,C.; Schultz-Cherry,S. |
| EPI1301204 | NP | Chile | 2016-Dec-13 | EPI_ISL_327610 | A/elegant tern/Chile/C20085/2016 | | Import from public-domain | Bravo-Vasquez,N.; Bahl,J.; Hamilton-West,C.; Schultz-Cherry,S. |
| EPI1301213 | MP | Chile | 2016-Dec-13 | EPI_ISL_327610 | A/elegant tern/Chile/C20085/2016 | | Import from public-domain | Bravo-Vasquez,N.; Bahl,J.; Hamilton-West,C.; Schultz-Cherry,S. |
| EPI1301215 | NS | Chile | 2016-Dec-13 | EPI_ISL_327610 | A/elegant tern/Chile/C20085/2016 | | Import from public-domain | Bravo-Vasquez,N.; Bahl,J.; Hamilton-West,C.; Schultz-Cherry,S. |
| EPI1057471 | NP | China | 2015-Dec-26 | EPI_ISL_277382 | A/swan/East China/SD588/2015(H13N8) | Institute of Poultry Science, Shandong, Academy of Agricultural Science | South China Agricultural University | Youling,Wang;Xiaoyuan,Yuan;Wu,Ai;Lihong,Qi; Yuxia,Zhang; Huaiying,Xu; Kai,Meng |
| EPI1057473 | MP | China | 2015-Dec-26 | EPI_ISL_277382 | A/swan/East China/SD588/2015(H13N8) | Institute of Poultry Science, Shandong, Academy of Agricultural Science | South China Agricultural University | Youling,Wang;Xiaoyuan,Yuan;Wu,Ai;Lihong,Qi; Yuxia,Zhang; Huaiying,Xu; Kai,Meng |
| EPI1057474 | NS | China | 2015-Dec-26 | EPI_ISL_277382 | A/swan/East China/SD588/2015(H13N8) | Institute of Poultry Science, Shandong, Academy of Agricultural Science | South China Agricultural University | Youling,Wang;Xiaoyuan,Yuan;Wu,Ai;Lihong,Qi; Yuxia,Zhang; Huaiying,Xu; Kai,Meng |
| EPI1884899 | NP | Sweden | 2021-Jul-13 | EPI_ISL_3319119 | A/Common guillemot/Sweden/SVA210729SZ0317/FB002610/I-2021 | National Veterinary Institute, SVA | National Veterinary Institute | |

| | | | | | | | | |
|---|---|---|---|---|---|---|---|---|
| EPI1884901 | MP | Sweden | 2021-Jul-13 | EPI_ISL_3319119 | A/Common guillemot/Sweden/SVA210729SZ0317/FB002610/I-2021 | National Veterinary Institute, SVA | National Veterinary Institute | |
| EPI1884902 | NS | Sweden | 2021-Jul-13 | EPI_ISL_3319119 | A/Common guillemot/Sweden/SVA210729SZ0317/FB002610/I-2021 | National Veterinary Institute, SVA | National Veterinary Institute | |
| EPI1067087 | MP | United States | 2016-May-27 | EPI_ISL_279337 | A/environment/New Jersey/UGAI16-0787/2016 | | Import from public-domain | Pickett,B.; Fedorova,N.; Puri,V.; Shrivastava,S.; Amedeo,P.; Isom,R.; Hu,L.; Durbin,A.; Rocchi,I.; Williams,T.; Poulson,R.L.; Bao,Y.; Sanders,R.; Zhdanov,S.; Kiryutin,B.; Lipman,D.J.; Tatusova,T.; Stallknecht,D.E. |
| EPI1067089 | NP | United States | 2016-May-27 | EPI_ISL_279337 | A/environment/New Jersey/UGAI16-0787/2016 | | Import from public-domain | Pickett,B.; Fedorova,N.; Puri,V.; Shrivastava,S.; Amedeo,P.; Isom,R.; Hu,L.; Durbin,A.; Rocchi,I.; Williams,T.; Poulson,R.L.; Bao,Y.; Sanders,R.; Zhdanov,S.; Kiryutin,B.; Lipman,D.J.; Tatusova,T.; Stallknecht,D.E. |
| EPI1067090 | NS | United States | 2016-May-27 | EPI_ISL_279337 | A/environment/New Jersey/UGAI16-0787/2016 | | Import from public-domain | Pickett,B.; Fedorova,N.; Puri,V.; Shrivastava,S.; Amedeo,P.; Isom,R.; Hu,L.; Durbin,A.; Rocchi,I.; Williams,T.; Poulson,R.L.; Bao,Y.; Sanders,R.; Zhdanov,S.; Kiryutin,B.; Lipman,D.J.; Tatusova,T.; Stallknecht,D.E. |
| EPI1066960 | MP | United States | 2016-May-16 | EPI_ISL_279322 | A/environment/New Jersey/UGAI16-1048/2016 | | Import from public-domain | Pickett,B.; Fedorova,N.; Puri,V.; Shrivastava,S.; Amedeo,P.; Isom,R.; Hu,L.; Durbin,A.; Rocchi,I.; Williams,T.; Poulson,R.L.; Bao,Y.; Sanders,R.; Zhdanov,S.; Kiryutin,B.; Lipman,D.J.; Tatusova,T.; Stallknecht,D.E. |
| EPI1066962 | NP | United States | 2016-May-16 | EPI_ISL_279322 | A/environment/New Jersey/UGAI16-1048/2016 | | Import from public-domain | Pickett,B.; Fedorova,N.; Puri,V.; Shrivastava,S.; Amedeo,P.; Isom,R.; Hu,L.; Durbin,A.; Rocchi,I.; Williams,T.; Poulson,R.L.; Bao,Y.; Sanders,R.; Zhdanov,S.; Kiryutin,B.; Lipman,D.J.; Tatusova,T.; Stallknecht,D.E. |
| EPI1066963 | NS | United States | 2016-May-16 | EPI_ISL_279322 | A/environment/New Jersey/UGAI16-1048/2016 | | Import from public-domain | Pickett,B.; Fedorova,N.; Puri,V.; Shrivastava,S.; Amedeo,P.; Isom,R.; Hu,L.; Durbin,A.; Rocchi,I.; Williams,T.; Poulson,R.L.; Bao,Y.; Sanders,R.; Zhdanov,S.; Kiryutin,B.; Lipman,D.J.; Tatusova,T.; Stallknecht,D.E. |
| EPI1066268 | MP | United States | 2016-May-25 | EPI_ISL_279224 | A/environment/New Jersey/UGAI16-1713/2016 | | Import from public-domain | Pickett,B.; Fedorova,N.; Puri,V.; Shrivastava,S.; Amedeo,P.; Isom,R.; Hu,L.; Durbin,A.; Rocchi,I.; Williams,T.; Poulson,R.L.; Bao,Y.; Sanders,R.; Zhdanov,S.; Kiryutin,B.; Lipman,D.J.; Tatusova,T.; Stallknecht,D.E. |
| EPI1066270 | NP | United States | 2016-May-25 | EPI_ISL_279224 | A/environment/New Jersey/UGAI16-1713/2016 | | Import from public-domain | Pickett,B.; Fedorova,N.; Puri,V.; Shrivastava,S.; Amedeo,P.; Isom,R.; Hu,L.; Durbin,A.; Rocchi,I.; Williams,T.; Poulson,R.L.; Bao,Y.; Sanders,R.; Zhdanov,S.; Kiryutin,B.; Lipman,D.J.; Tatusova,T.; Stallknecht,D.E. |
| EPI1066271 | NS | United States | 2016-May-25 | EPI_ISL_279224 | A/environment/New Jersey/UGAI16-1713/2016 | | Import from public-domain | Pickett,B.; Fedorova,N.; Puri,V.; Shrivastava,S.; Amedeo,P.; Isom,R.; Hu,L.; Durbin,A.; Rocchi,I.; Williams,T.; Poulson,R.L.; Bao,Y.; Sanders,R.; Zhdanov,S.; Kiryutin,B.; Lipman,D.J.; Tatusova,T.; Stallknecht,D.E. |
| EPI823782 | NP | Russian Federation | 2016-May-01 | EPI_ISL_231687 | A/environment/Kamchatka/24/2016 | State Research Center of Virology and Biotechnology (VECTOR) | WHO National Influenza Centre Russian Federation | Fadeev,Artem; Komissarov,Andrey; Egorova,Anna; Sintsova,Ksenia; Musaeva,Tamila; Susloparov,Ivan; Marchenko,Vasiliy; Ryzhikov,Aleksandr |
| EPI823787 | MP | Russian Federation | 2016-May-01 | EPI_ISL_231687 | A/environment/Kamchatka/24/2016 | State Research Center of Virology and Biotechnology (VECTOR) | WHO National Influenza Centre Russian Federation | Fadeev,Artem; Komissarov,Andrey; Egorova,Anna; Sintsova,Ksenia; Musaeva,Tamila; Susloparov,Ivan; Marchenko,Vasiliy; Ryzhikov,Aleksandr |
| EPI823792 | NS | Russian Federation | 2016-May-01 | EPI_ISL_231687 | A/environment/Kamchatka/24/2016 | State Research Center of Virology and Biotechnology (VECTOR) | WHO National Influenza Centre Russian Federation | Fadeev,Artem; Komissarov,Andrey; Egorova,Anna; Sintsova,Ksenia; Musaeva,Tamila; Susloparov,Ivan; Marchenko,Vasiliy; Ryzhikov,Aleksandr |
| EPI823768 | MP | Russian Federation | 2016-May-01 | EPI_ISL_231686 | A/environment/Kamchatka/23/2016 | State Research Center of Virology and Biotechnology (VECTOR) | WHO National Influenza Centre Russian Federation | Fadeev,Artem; Komissarov,Andrey; Egorova,Anna; Sintsova,Ksenia; Musaeva,Tamila; Susloparov,Ivan; Marchenko,Vasiliy; Ryzhikov,Aleksandr |
| EPI823766 | NP | Russian Federation | 2016-May-01 | EPI_ISL_231686 | A/environment/Kamchatka/23/2016 | State Research Center of Virology and Biotechnology (VECTOR) | WHO National Influenza Centre Russian Federation | Fadeev,Artem; Komissarov,Andrey; Egorova,Anna; Sintsova,Ksenia; Musaeva,Tamila; Susloparov,Ivan; Marchenko,Vasiliy; Ryzhikov,Aleksandr |
| EPI823773 | NS | Russian Federation | 2016-May-01 | EPI_ISL_231686 | A/environment/Kamchatka/23/2016 | State Research Center of Virology and Biotechnology (VECTOR) | WHO National Influenza Centre Russian Federation | Fadeev,Artem; Komissarov,Andrey; Egorova,Anna; Sintsova,Ksenia; Musaeva,Tamila; Susloparov,Ivan; Marchenko,Vasiliy; Ryzhikov,Aleksandr |
| EPI2145460 | NP | Australia | 2019-Oct-30 | EPI_ISL_14769829 | A/wild waterbird/Victoria/19-04759-011/2019 | | Import from public-domain | Wille,M.; Grillo,V.; Ban de Gouvea Pedroso,S.; Burgess,G.W.; Crawley,A.; Dickason,C.; Hansbro,P.M.; Hoque,M.A.; Horwood,P.F.; Kirkland,P.D.; Kung,N.Y.; Lynch,S.E.; Martin,S.; McArthur,M.; O'Riley,K.; Read,A.J.; Warner,S.; Hoye,B.J.; Lisovski,S.; Leen,T.; Hurt,A.; Butler,J.; Broz,I.; Davies,K.R.; Mileto,P.; Neave,M.J.; Ste |
| EPI2145462 | MP | Australia | 2019-Oct-30 | EPI_ISL_14769829 | A/wild waterbird/Victoria/19-04759-011/2019 | | Import from public-domain | Wille,M.; Grillo,V.; Ban de Gouvea Pedroso,S.; Burgess,G.W.; Crawley,A.; Dickason,C.; Hansbro,P.M.; Hoque,M.A.; Horwood,P.F.; Kirkland,P.D.; Kung,N.Y.; Lynch,S.E.; Martin,S.; McArthur,M.; O'Riley,K.; Read,A.J.; Warner,S.; Hoye,B.J.; Lisovski,S.; Leen,T.; Hurt,A.; Butler,J.; Broz,I.; Davies,K.R.; Mileto,P.; Neave,M.J.; Ste |
| EPI2145463 | NS | Australia | 2019-Oct-30 | EPI_ISL_14769829 | A/wild waterbird/Victoria/19-04759-011/2019 | | Import from public-domain | Wille,M.; Grillo,V.; Ban de Gouvea Pedroso,S.; Burgess,G.W.; Crawley,A.; Dickason,C.; Hansbro,P.M.; Hoque,M.A.; Horwood,P.F.; Kirkland,P.D.; Kung,N.Y.; Lynch,S.E.; Martin,S.; McArthur,M.; O'Riley,K.; Read,A.J.; Warner,S.; Hoye,B.J.; Lisovski,S.; Leen,T.; Hurt,A.; Butler,J.; Broz,I.; Davies,K.R.; Mileto,P.; Neave,M.J.; Ste |
| EPI1932185 | NP | Peru | 2019-Oct-17 | EPI_ISL_6795367 | A/Kelp gull/Peru/M8/2019 | | Import from public-domain | Simas,P.V.M.; Castro-Sanguinetti,G.; Apaza-Chiara,A.P.; Callupe-Leyva,J.A.; Rondon-Espinoza,J.A.; Ramirez-Velasquez,M.G.; Carranza,C.; Flores,C.; Rivera-Geronimo,H.; Calle-Espinoza,S.Y.; Gavidia-Chucan,C.M.; Fernandez,M.; More-Bayona,J.A.; Gonzalez-Veliz,R.I.; Icochea-D'Arrigo,M.E.; Vakharia,V.N. |
| EPI1932186 | MP | Peru | 2019-Oct-17 | EPI_ISL_6795367 | A/Kelp gull/Peru/M8/2019 | | Import from public-domain | Simas,P.V.M.; Castro-Sanguinetti,G.; Apaza-Chiara,A.P.; Callupe-Leyva,J.A.; Rondon-Espinoza,J.A.; Ramirez-Velasquez,M.G.; Carranza,C.; Flores,C.; Rivera-Geronimo,H.; Calle-Espinoza,S.Y.; Gavidia-Chucan,C.M.; Fernandez,M.; More-Bayona,J.A.; Gonzalez-Veliz,R.I.; Icochea-D'Arrigo,M.E.; Vakharia,V.N. |
| EPI1933026 | NS | Peru | 2019-Oct-17 | EPI_ISL_6795367 | A/Kelp gull/Peru/M8/2019 | | Import from public-domain | Simas,P.V.M.; Castro-Sanguinetti,G.; Apaza-Chiara,A.P.; Callupe-Leyva,J.A.; Rondon-Espinoza,J.A.; Ramirez-Velasquez,M.G.; Carranza,C.; Flores,C.; Rivera-Geronimo,H.; Calle-Espinoza,S.Y.; Gavidia-Chucan,C.M.; Fernandez,M.; More-Bayona,J.A.; Gonzalez-Veliz,R.I.; Icochea-D'Arrigo,M.E.; Vakharia,V.N. |
| EPI1565259 | NP | China | 2017-May-01 | EPI_ISL_381339 | A/Duck/China/Weihai/2017 | | Import from public-domain | Yu,Z.; Cheng,K. |
| EPI1565261 | MP | China | 2017-May-01 | EPI_ISL_381339 | A/Duck/China/Weihai/2017 | | Import from public-domain | Yu,Z.; Cheng,K. |
| EPI1565262 | NS | China | 2017-May-01 | EPI_ISL_381339 | A/Duck/China/Weihai/2017 | | Import from public-domain | Yu,Z.; Cheng,K. |
| EPI729743 | NP | United States | 2014-Sep-29 | EPI_ISL_215572 | A/northern pintail/Alaska/886/2014 | | Import from public-domain | Ramey,A.M.; Reeves,A.B.; TeSlaa,J.L.; Nashold,S.; Donnelly,T.; Bahl,J.; Hall,J.S.; Donnelly,T.F. |
| EPI729745 | MP | United States | 2014-Sep-29 | EPI_ISL_215572 | A/northern pintail/Alaska/886/2014 | | Import from public-domain | Ramey,A.M.; Reeves,A.B.; TeSlaa,J.L.; Nashold,S.; Donnelly,T.; Bahl,J.; Hall,J.S.; Donnelly,T.F. |
| EPI729746 | NS | United States | 2014-Sep-29 | EPI_ISL_215572 | A/northern pintail/Alaska/886/2014 | | Import from public-domain | Ramey,A.M.; Reeves,A.B.; TeSlaa,J.L.; Nashold,S.; Donnelly,T.; Bahl,J.; Hall,J.S.; Donnelly,T.F. |
| EPI729759 | NP | United States | 2014-Sep-22 | EPI_ISL_215574 | A/mallard/Alaska/903/2014 | | Import from public-domain | Ramey,A.M.; Reeves,A.B.; TeSlaa,J.L.; Nashold,S.; Donnelly,T.; Bahl,J.; Hall,J.S.; Donnelly,T.F. |
| EPI729761 | MP | United States | 2014-Sep-22 | EPI_ISL_215574 | A/mallard/Alaska/903/2014 | | Import from public-domain | Ramey,A.M.; Reeves,A.B.; TeSlaa,J.L.; Nashold,S.; Donnelly,T.; Bahl,J.; Hall,J.S.; Donnelly,T.F. |
| EPI729762 | NS | United States | 2014-Sep-22 | EPI_ISL_215574 | A/mallard/Alaska/903/2014 | | Import from public-domain | Ramey,A.M.; Reeves,A.B.; TeSlaa,J.L.; Nashold,S.; Donnelly,T.; Bahl,J.; Hall,J.S.; Donnelly,T.F. |
| EPI2403130 | NS | United States | 2022-May-31 | EPI_ISL_16920536 | A/Laughing gull/Delaware/809/2022 | | Import from public-domain | Direct Submission |
| EPI2403131 | MP | United States | 2022-May-31 | EPI_ISL_16920536 | A/Laughing gull/Delaware/809/2022 | | Import from public-domain | Direct Submission |
| EPI2403133 | NP | United States | 2022-May-31 | EPI_ISL_16920536 | A/Laughing gull/Delaware/809/2022 | | Import from public-domain | Direct Submission |

| | | | | | | | |
|---|---|---|---|---|---|---|---|
| EPI2401472 | NS | United States | 2022-May-31 | EPI_ISL_16919121 | A/Laughing gull/Delaware/827/2022 | Import from public-domain | Direct Submission |
| EPI2401474 | MP | United States | 2022-May-31 | EPI_ISL_16919121 | A/Laughing gull/Delaware/827/2022 | Import from public-domain | Direct Submission |
| EPI2401476 | NP | United States | 2022-May-31 | EPI_ISL_16919121 | A/Laughing gull/Delaware/827/2022 | Import from public-domain | Direct Submission |
| EPI2401396 | NS | United States | 2022-May-31 | EPI_ISL_16919101 | A/Laughing Gull/Delaware/801/2022 | Import from public-domain | Direct Submission |
| EPI2401397 | MP | United States | 2022-May-31 | EPI_ISL_16919101 | A/Laughing Gull/Delaware/801/2022 | Import from public-domain | Direct Submission |
| EPI2401399 | NP | United States | 2022-May-31 | EPI_ISL_16919101 | A/Laughing Gull/Delaware/801/2022 | Import from public-domain | Direct Submission |
| EPI2147993 | MP | United States | 2021-Jun-01 | EPI_ISL_14777737 | A/Laughing Gull/Delaware/591/2021 | Import from public-domain | Direct Submission |
| EPI2147994 | NP | United States | 2021-Jun-01 | EPI_ISL_14777737 | A/Laughing Gull/Delaware/591/2021 | Import from public-domain | Direct Submission |
| EPI2147995 | NS | United States | 2021-Jun-01 | EPI_ISL_14777737 | A/Laughing Gull/Delaware/591/2021 | Import from public-domain | Direct Submission |
| EPI2147985 | MP | United States | 2021-Jun-01 | EPI_ISL_14777736 | A/Laughing Gull/Delaware/294/2021 | Import from public-domain | Direct Submission |
| EPI2147986 | NP | United States | 2021-Jun-01 | EPI_ISL_14777736 | A/Laughing Gull/Delaware/294/2021 | Import from public-domain | Direct Submission |
| EPI2147987 | NS | United States | 2021-Jun-01 | EPI_ISL_14777736 | A/Laughing Gull/Delaware/294/2021 | Import from public-domain | Direct Submission |
| EPI2147854 | MP | United States | 2021-Jun-01 | EPI_ISL_14777715 | A/Laughing Gull/Delaware/583/2021 | Import from public-domain | Direct Submission |
| EPI2147855 | NP | United States | 2021-Jun-01 | EPI_ISL_14777715 | A/Laughing Gull/Delaware/583/2021 | Import from public-domain | Direct Submission |
| EPI2147856 | NS | United States | 2021-Jun-01 | EPI_ISL_14777715 | A/Laughing Gull/Delaware/583/2021 | Import from public-domain | Direct Submission |
| EPI1789453 | NP | United States | 2019-May-20 | EPI_ISL_505801 | A/Laughing Gull/Delaware/589/2019 | Import from public-domain | Woodard,K.; Kercher,L.; Mukherjee,N.; Webby,R. |
| EPI1789457 | NS | United States | 2019-May-20 | EPI_ISL_505801 | A/Laughing Gull/Delaware/589/2019 | Import from public-domain | Woodard,K.; Kercher,L.; Mukherjee,N.; Webby,R. |
| EPI1789459 | MP | United States | 2019-May-20 | EPI_ISL_505801 | A/Laughing Gull/Delaware/589/2019 | Import from public-domain | Woodard,K.; Kercher,L.; Mukherjee,N.; Webby,R. |
| EPI1525707 | NP | United States | 2014-May-20 | EPI_ISL_372160 | A/laughing gull/Delaware Bay/358/2014 | Import from public-domain | Hicks,J.T.; Friedman,K.; Qiu,X.; Kim,D.-K.; Hixson,J.; Krauss,S.; Bahl,J. |
| EPI1525713 | MP | United States | 2014-May-20 | EPI_ISL_372160 | A/laughing gull/Delaware Bay/358/2014 | Import from public-domain | Hicks,J.T.; Friedman,K.; Qiu,X.; Kim,D.-K.; Hixson,J.; Krauss,S.; Bahl,J. |
| EPI1525716 | NS | United States | 2014-May-20 | EPI_ISL_372160 | A/laughing gull/Delaware Bay/358/2014 | Import from public-domain | Hicks,J.T.; Friedman,K.; Qiu,X.; Kim,D.-K.; Hixson,J.; Krauss,S.; Bahl,J. |
| EPI1525676 | NS | United States | 2014-May-20 | EPI_ISL_372153 | A/laughing gull/Delaware Bay/328/2014 | Import from public-domain | Hicks,J.T.; Friedman,K.; Qiu,X.; Kim,D.-K.; Hixson,J.; Krauss,S.; Bahl,J. |
| EPI1525650 | MP | United States | 2014-May-20 | EPI_ISL_372153 | A/laughing gull/Delaware Bay/328/2014 | Import from public-domain | Hicks,J.T.; Friedman,K.; Qiu,X.; Kim,D.-K.; Hixson,J.; Krauss,S.; Bahl,J. |
| EPI1525669 | NP | United States | 2014-May-20 | EPI_ISL_372153 | A/laughing gull/Delaware Bay/328/2014 | Import from public-domain | Hicks,J.T.; Friedman,K.; Qiu,X.; Kim,D.-K.; Hixson,J.; Krauss,S.; Bahl,J. |
| EPI1525633 | MP | United States | 2014-May-20 | EPI_ISL_372149 | A/laughing gull/Delaware Bay/351/2014 | Import from public-domain | Hicks,J.T.; Friedman,K.; Qiu,X.; Kim,D.-K.; Hixson,J.; Krauss,S.; Bahl,J. |
| EPI1525637 | NP | United States | 2014-May-20 | EPI_ISL_372149 | A/laughing gull/Delaware Bay/351/2014 | Import from public-domain | Hicks,J.T.; Friedman,K.; Qiu,X.; Kim,D.-K.; Hixson,J.; Krauss,S.; Bahl,J. |
| EPI1525640 | NS | United States | 2014-May-20 | EPI_ISL_372149 | A/laughing gull/Delaware Bay/351/2014 | Import from public-domain | Hicks,J.T.; Friedman,K.; Qiu,X.; Kim,D.-K.; Hixson,J.; Krauss,S.; Bahl,J. |
| EPI1525572 | MP | United States | 2014-May-20 | EPI_ISL_372142 | A/laughing gull/Delaware Bay/340/2014 | Import from public-domain | Hicks,J.T.; Friedman,K.; Qiu,X.; Kim,D.-K.; Hixson,J.; Krauss,S.; Bahl,J. |
| EPI1525574 | NS | United States | 2014-May-20 | EPI_ISL_372142 | A/laughing gull/Delaware Bay/340/2014 | Import from public-domain | Hicks,J.T.; Friedman,K.; Qiu,X.; Kim,D.-K.; Hixson,J.; Krauss,S.; Bahl,J. |
| EPI1525581 | NP | United States | 2014-May-20 | EPI_ISL_372142 | A/laughing gull/Delaware Bay/340/2014 | Import from public-domain | Hicks,J.T.; Friedman,K.; Qiu,X.; Kim,D.-K.; Hixson,J.; Krauss,S.; Bahl,J. |
| EPI1525366 | NS | United States | 2014-May-20 | EPI_ISL_372117 | A/laughing gull/Delaware Bay/327/2014 | Import from public-domain | Hicks,J.T.; Friedman,K.; Qiu,X.; Kim,D.-K.; Hixson,J.; Krauss,S.; Bahl,J. |
| EPI1525368 | NP | United States | 2014-May-20 | EPI_ISL_372117 | A/laughing gull/Delaware Bay/327/2014 | Import from public-domain | Hicks,J.T.; Friedman,K.; Qiu,X.; Kim,D.-K.; Hixson,J.; Krauss,S.; Bahl,J. |
| EPI1525370 | MP | United States | 2014-May-20 | EPI_ISL_372117 | A/laughing gull/Delaware Bay/327/2014 | Import from public-domain | Hicks,J.T.; Friedman,K.; Qiu,X.; Kim,D.-K.; Hixson,J.; Krauss,S.; Bahl,J. |
| EPI1525295 | NS | United States | 2014-May-19 | EPI_ISL_372108 | A/laughing gull/Delaware Bay/30/2014 | Import from public-domain | Hicks,J.T.; Friedman,K.; Qiu,X.; Kim,D.-K.; Hixson,J.; Krauss,S.; Bahl,J. |
| EPI1525298 | NP | United States | 2014-May-19 | EPI_ISL_372108 | A/laughing gull/Delaware Bay/30/2014 | Import from public-domain | Hicks,J.T.; Friedman,K.; Qiu,X.; Kim,D.-K.; Hixson,J.; Krauss,S.; Bahl,J. |

| | | | | | | | |
|---|---|---|---|---|---|---|---|
| EPI1525301 | MP | United States | 2014-May-19 | EPI_ISL_372108 | A/laughing gull/Delaware Bay/30/2014 | Import from public-domain | Hicks,J.T.; Friedman,K.; Qiu,X.; Kim,D.-K.; Hixson,J.; Krauss,S.; Bahl,J. |
| EPI1525190 | NP | United States | 2014-May-19 | EPI_ISL_372092 | A/laughing gull/Delaware Bay/39/2014 | Import from public-domain | Hicks,J.T.; Friedman,K.; Qiu,X.; Kim,D.-K.; Hixson,J.; Krauss,S.; Bahl,J. |
| EPI1525192 | NS | United States | 2014-May-19 | EPI_ISL_372092 | A/laughing gull/Delaware Bay/39/2014 | Import from public-domain | Hicks,J.T.; Friedman,K.; Qiu,X.; Kim,D.-K.; Hixson,J.; Krauss,S.; Bahl,J. |
| EPI1525195 | MP | United States | 2014-May-19 | EPI_ISL_372092 | A/laughing gull/Delaware Bay/39/2014 | Import from public-domain | Hicks,J.T.; Friedman,K.; Qiu,X.; Kim,D.-K.; Hixson,J.; Krauss,S.; Bahl,J. |
| EPI1525150 | NS | United States | 2014-May-20 | EPI_ISL_372087 | A/laughing gull/Delaware Bay/350/2014 | Import from public-domain | Hicks,J.T.; Friedman,K.; Qiu,X.; Kim,D.-K.; Hixson,J.; Krauss,S.; Bahl,J. |
| EPI1525151 | MP | United States | 2014-May-20 | EPI_ISL_372087 | A/laughing gull/Delaware Bay/350/2014 | Import from public-domain | Hicks,J.T.; Friedman,K.; Qiu,X.; Kim,D.-K.; Hixson,J.; Krauss,S.; Bahl,J. |
| EPI1525154 | NP | United States | 2014-May-20 | EPI_ISL_372087 | A/laughing gull/Delaware Bay/350/2014 | Import from public-domain | Hicks,J.T.; Friedman,K.; Qiu,X.; Kim,D.-K.; Hixson,J.; Krauss,S.; Bahl,J. |
| EPI1530536 | MP | United States | 2018-May-22 | EPI_ISL_373079 | A/laughing gull/Delaware Bay/458/2018 | Import from public-domain | Woodard,K.; Turner,J.C.; Franks,J.; Friedman,K.; Kercher,L.; Webby,R.J. |
| EPI1530538 | NS | United States | 2018-May-22 | EPI_ISL_373079 | A/laughing gull/Delaware Bay/458/2018 | Import from public-domain | Woodard,K.; Turner,J.C.; Franks,J.; Friedman,K.; Kercher,L.; Webby,R.J. |
| EPI1530541 | NP | United States | 2018-May-22 | EPI_ISL_373079 | A/laughing gull/Delaware Bay/458/2018 | Import from public-domain | Woodard,K.; Turner,J.C.; Franks,J.; Friedman,K.; Kercher,L.; Webby,R.J. |
| EPI1530523 | NS | United States | 2018-May-22 | EPI_ISL_373077 | A/laughing gull/Delaware Bay/433/2018 | Import from public-domain | Woodard,K.; Turner,J.C.; Franks,J.; Friedman,K.; Kercher,L.; Webby,R.J. |
| EPI1530527 | NP | United States | 2018-May-22 | EPI_ISL_373077 | A/laughing gull/Delaware Bay/433/2018 | Import from public-domain | Woodard,K.; Turner,J.C.; Franks,J.; Friedman,K.; Kercher,L.; Webby,R.J. |
| EPI1530528 | MP | United States | 2018-May-22 | EPI_ISL_373077 | A/laughing gull/Delaware Bay/433/2018 | Import from public-domain | Woodard,K.; Turner,J.C.; Franks,J.; Friedman,K.; Kercher,L.; Webby,R.J. |
| EPI1530490 | NS | United States | 2018-May-22 | EPI_ISL_373073 | A/laughing gull/Delaware Bay/436/2018 | Import from public-domain | Woodard,K.; Turner,J.C.; Franks,J.; Friedman,K.; Kercher,L.; Webby,R.J. |
| EPI1530493 | NP | United States | 2018-May-22 | EPI_ISL_373073 | A/laughing gull/Delaware Bay/436/2018 | Import from public-domain | Woodard,K.; Turner,J.C.; Franks,J.; Friedman,K.; Kercher,L.; Webby,R.J. |
| EPI1530496 | MP | United States | 2018-May-22 | EPI_ISL_373073 | A/laughing gull/Delaware Bay/436/2018 | Import from public-domain | Woodard,K.; Turner,J.C.; Franks,J.; Friedman,K.; Kercher,L.; Webby,R.J. |
| EPI1530428 | MP | United States | 2018-May-22 | EPI_ISL_373065 | A/laughing gull/Delaware Bay/453/2018 | Import from public-domain | Woodard,K.; Turner,J.C.; Franks,J.; Friedman,K.; Kercher,L.; Webby,R.J. |
| EPI1530431 | NS | United States | 2018-May-22 | EPI_ISL_373065 | A/laughing gull/Delaware Bay/453/2018 | Import from public-domain | Woodard,K.; Turner,J.C.; Franks,J.; Friedman,K.; Kercher,L.; Webby,R.J. |
| EPI1530433 | NP | United States | 2018-May-22 | EPI_ISL_373065 | A/laughing gull/Delaware Bay/453/2018 | Import from public-domain | Woodard,K.; Turner,J.C.; Franks,J.; Friedman,K.; Kercher,L.; Webby,R.J. |
| EPI1530382 | NP | United States | 2018-May-23 | EPI_ISL_373059 | A/laughing gull/Delaware Bay/464/2018 | Import from public-domain | Woodard,K.; Turner,J.C.; Franks,J.; Friedman,K.; Kercher,L.; Webby,R.J. |
| EPI1530383 | MP | United States | 2018-May-23 | EPI_ISL_373059 | A/laughing gull/Delaware Bay/464/2018 | Import from public-domain | Woodard,K.; Turner,J.C.; Franks,J.; Friedman,K.; Kercher,L.; Webby,R.J. |
| EPI1530384 | NS | United States | 2018-May-23 | EPI_ISL_373059 | A/laughing gull/Delaware Bay/464/2018 | Import from public-domain | Woodard,K.; Turner,J.C.; Franks,J.; Friedman,K.; Kercher,L.; Webby,R.J. |
| EPI1530365 | MP | United States | 2018-May-22 | EPI_ISL_373057 | A/laughing gull/Delaware Bay/451/2018 | Import from public-domain | Woodard,K.; Turner,J.C.; Franks,J.; Friedman,K.; Kercher,L.; Webby,R.J. |
| EPI1530368 | NS | United States | 2018-May-22 | EPI_ISL_373057 | A/laughing gull/Delaware Bay/451/2018 | Import from public-domain | Woodard,K.; Turner,J.C.; Franks,J.; Friedman,K.; Kercher,L.; Webby,R.J. |
| EPI1530370 | NP | United States | 2018-May-22 | EPI_ISL_373057 | A/laughing gull/Delaware Bay/451/2018 | Import from public-domain | Woodard,K.; Turner,J.C.; Franks,J.; Friedman,K.; Kercher,L.; Webby,R.J. |
| EPI1228279 | NS | United States | 2017-May-23 | EPI_ISL_309577 | A/laughing gull/New Jersey/UGAI17-2856/2017 | Import from public-domain | Tan,G.; Pickett,B.; Fedorova,N.; Amedeo,P.; Isom,R.; Hu,L.; Christensen,J.; Durbin,A.; Williams,T.; Arumemi,F.; Poulson,R.; Bao,Y.; Sanders,R.; Zhdanov,S.; Kiryutin,B.; Lipman,D.J.; Tatusova,T.; Hatcher,E.; Stallknecht,D. |
| EPI1228276 | MP | United States | 2017-May-23 | EPI_ISL_309577 | A/laughing gull/New Jersey/UGAI17-2856/2017 | Import from public-domain | Tan,G.; Pickett,B.; Fedorova,N.; Amedeo,P.; Isom,R.; Hu,L.; Christensen,J.; Durbin,A.; Williams,T.; Arumemi,F.; Poulson,R.; Bao,Y.; Sanders,R.; Zhdanov,S.; Kiryutin,B.; Lipman,D.J.; Tatusova,T.; Hatcher,E.; Stallknecht,D. |
| EPI1228278 | NP | United States | 2017-May-23 | EPI_ISL_309577 | A/laughing gull/New Jersey/UGAI17-2856/2017 | Import from public-domain | Tan,G.; Pickett,B.; Fedorova,N.; Amedeo,P.; Isom,R.; Hu,L.; Christensen,J.; Durbin,A.; Williams,T.; Arumemi,F.; Poulson,R.; Bao,Y.; Sanders,R.; Zhdanov,S.; Kiryutin,B.; Lipman,D.J.; Tatusova,T.; Hatcher,E.; Stallknecht,D. |
| EPI1228272 | NS | United States | 2017-May-23 | EPI_ISL_309576 | A/laughing gull/New Jersey/UGAI17-2850/2017 | Import from public-domain | Tan,G.; Pickett,B.; Fedorova,N.; Amedeo,P.; Isom,R.; Hu,L.; Christensen,J.; Durbin,A.; Williams,T.; Arumemi,F.; Poulson,R.; Bao,Y.; Sanders,R.; Zhdanov,S.; Kiryutin,B.; Lipman,D.J.; Tatusova,T.; Hatcher,E.; Stallknecht,D. |
| EPI1228269 | MP | United States | 2017-May-23 | EPI_ISL_309576 | A/laughing gull/New Jersey/UGAI17-2850/2017 | Import from public-domain | Tan,G.; Pickett,B.; Fedorova,N.; Amedeo,P.; Isom,R.; Hu,L.; Christensen,J.; Durbin,A.; Williams,T.; Arumemi,F.; Poulson,R.; Bao,Y.; Sanders,R.; Zhdanov,S.; Kiryutin,B.; Lipman,D.J.; Tatusova,T.; Hatcher,E.; Stallknecht,D. |
| EPI1228271 | NP | United States | 2017-May-23 | EPI_ISL_309576 | A/laughing gull/New Jersey/UGAI17-2850/2017 | Import from public-domain | Tan,G.; Pickett,B.; Fedorova,N.; Amedeo,P.; Isom,R.; Hu,L.; Christensen,J.; Durbin,A.; Williams,T.; Arumemi,F.; Poulson,R.; Bao,Y.; Sanders,R.; Zhdanov,S.; Kiryutin,B.; Lipman,D.J.; Tatusova,T.; Hatcher,E.; Stallknecht,D. |
| EPI1227232 | MP | United States | 2017-May-23 | EPI_ISL_309426 | A/laughing gull/New Jersey/UGAI17-2843/2017 | Import from public-domain | Tan,G.; Pickett,B.; Fedorova,N.; Amedeo,P.; Isom,R.; Hu,L.; Christensen,J.; Durbin,A.; Williams,T.; Arumemi,F.; Poulson,R.; Bao,Y.; Sanders,R.; Zhdanov,S.; Kiryutin,B.; Lipman,D.J.; Tatusova,T.; Hatcher,E.; Stallknecht,D. |
| EPI1227234 | NP | United States | 2017-May-23 | EPI_ISL_309426 | A/laughing gull/New Jersey/UGAI17-2843/2017 | Import from public-domain | Tan,G.; Pickett,B.; Fedorova,N.; Amedeo,P.; Isom,R.; Hu,L.; Christensen,J.; Durbin,A.; Williams,T.; Arumemi,F.; Poulson,R.; Bao,Y.; Sanders,R.; Zhdanov,S.; Kiryutin,B.; Lipman,D.J.; Tatusova,T.; Hatcher,E.; Stallknecht,D. |
| EPI1227235 | NS | United States | 2017-May-23 | EPI_ISL_309426 | A/laughing gull/New Jersey/UGAI17-2843/2017 | Import from public-domain | Tan,G.; Pickett,B.; Fedorova,N.; Amedeo,P.; Isom,R.; Hu,L.; Christensen,J.; Durbin,A.; Williams,T.; Arumemi,F.; Poulson,R.; Bao,Y.; Sanders,R.; Zhdanov,S.; Kiryutin,B.; Lipman,D.J.; Tatusova,T.; Hatcher,E.; Stallknecht,D. |
| EPI1227224 | MP | United States | 2017-May-23 | EPI_ISL_309425 | A/laughing gull/New Jersey/UGAI17-2839/2017 | Import from public-domain | Tan,G.; Pickett,B.; Fedorova,N.; Amedeo,P.; Isom,R.; Hu,L.; Christensen,J.; Durbin,A.; Williams,T.; Arumemi,F.; Poulson,R.; Bao,Y.; Sanders,R.; Zhdanov,S.; Kiryutin,B.; Lipman,D.J.; Tatusova,T.; Hatcher,E.; Stallknecht,D. |

| | | | | | | | |
|---|---|---|---|---|---|---|---|
| EPI1227227 | NP | United States | 2017-May-23 | EPI_ISL_309425 | A/laughing gull/New Jersey/UGAI17-2839/2017 | Import from public-domain | Tan,G.; Pickett,B.; Fedorova,N.; Amedeo,P.; Isom,R.; Hu,L.; Christensen,J.; Durbin,A.; Williams,T.; Arumemi,F.; Poulson,R.; Bao,Y.; Sanders,R.; Zhdanov,S.; Kiryutin,B.; Lipman,D.J.; Tatusova,T.; Hatcher,E.; Stallknecht,D. |
| EPI1227228 | NS | United States | 2017-May-23 | EPI_ISL_309425 | A/laughing gull/New Jersey/UGAI17-2839/2017 | Import from public-domain | Tan,G.; Pickett,B.; Fedorova,N.; Amedeo,P.; Isom,R.; Hu,L.; Christensen,J.; Durbin,A.; Williams,T.; Arumemi,F.; Poulson,R.; Bao,Y.; Sanders,R.; Zhdanov,S.; Kiryutin,B.; Lipman,D.J.; Tatusova,T.; Hatcher,E.; Stallknecht,D. |
| EPI1789445 | MP | United States | 2019-May-20 | EPI_ISL_505800 | A/Herring Gull/Delaware/597/2019 | Import from public-domain | Woodard,K.; Kercher,L.; Mukherjee,N.; Webby,R. |
| EPI1789448 | NP | United States | 2019-May-20 | EPI_ISL_505800 | A/Herring Gull/Delaware/597/2019 | Import from public-domain | Woodard,K.; Kercher,L.; Mukherjee,N.; Webby,R. |
| EPI1789451 | NS | United States | 2019-May-20 | EPI_ISL_505800 | A/Herring Gull/Delaware/597/2019 | Import from public-domain | Woodard,K.; Kercher,L.; Mukherjee,N.; Webby,R. |
| EPI1789437 | MP | United States | 2019-May-20 | EPI_ISL_505799 | A/Herring Gull/Delaware/608/2019 | Import from public-domain | Woodard,K.; Kercher,L.; Mukherjee,N.; Webby,R. |
| EPI1789439 | NS | United States | 2019-May-20 | EPI_ISL_505799 | A/Herring Gull/Delaware/608/2019 | Import from public-domain | Woodard,K.; Kercher,L.; Mukherjee,N.; Webby,R. |
| EPI1789443 | NP | United States | 2019-May-20 | EPI_ISL_505799 | A/Herring Gull/Delaware/608/2019 | Import from public-domain | Woodard,K.; Kercher,L.; Mukherjee,N.; Webby,R. |
| EPI1549759 | NS | United States | 2014-May-19 | EPI_ISL_378352 | A/herring gull/Delaware Bay/22/2014 | Import from public-domain | Hicks,J.T.; Friedman,K.; Qiu,X.; Kim,D.-K.; Hixson,J.; Krauss,S.; Bahl,J. |
| EPI1549763 | NP | United States | 2014-May-19 | EPI_ISL_378352 | A/herring gull/Delaware Bay/22/2014 | Import from public-domain | Hicks,J.T.; Friedman,K.; Qiu,X.; Kim,D.-K.; Hixson,J.; Krauss,S.; Bahl,J. |
| EPI1549765 | MP | United States | 2014-May-19 | EPI_ISL_378352 | A/herring gull/Delaware Bay/22/2014 | Import from public-domain | Hicks,J.T.; Friedman,K.; Qiu,X.; Kim,D.-K.; Hixson,J.; Krauss,S.; Bahl,J. |
| EPI1549514 | NP | China | 2013-Oct-01 | EPI_ISL_378286 | A/mallard/Dalian/DZ-137/2013 | Import from public-domain | Fan,S.; Gao,X.; Wang,T.; Sun,W.; Ying,Y.; Li,X.; Zhao,Y.; Yang,S.; Gao,Y.; Xia,X. |
| EPI1549516 | MP | China | 2013-Oct-01 | EPI_ISL_378286 | A/mallard/Dalian/DZ-137/2013 | Import from public-domain | Fan,S.; Gao,X.; Wang,T.; Sun,W.; Ying,Y.; Li,X.; Zhao,Y.; Yang,S.; Gao,Y.; Xia,X. |
| EPI1549517 | NS | China | 2013-Oct-01 | EPI_ISL_378286 | A/mallard/Dalian/DZ-137/2013 | Import from public-domain | Fan,S.; Gao,X.; Wang,T.; Sun,W.; Ying,Y.; Li,X.; Zhao,Y.; Yang,S.; Gao,Y.; Xia,X. |
| EPI1619665 | NS | China | 2014-Oct-18 | EPI_ISL_397343 | A/Eurasian curlew/Liaoning/ZH-385/2014 | Import from public-domain | Gao,X. |
| EPI1619662 | NP | China | 2014-Oct-18 | EPI_ISL_397343 | A/Eurasian curlew/Liaoning/ZH-385/2014 | Import from public-domain | Gao,X. |
| EPI1619664 | MP | China | 2014-Oct-18 | EPI_ISL_397343 | A/Eurasian curlew/Liaoning/ZH-385/2014 | Import from public-domain | Gao,X. |
| EPI1619657 | NS | China | 2014-Oct-18 | EPI_ISL_397342 | A/Eurasian curlew/Liaoning/ZH-186/2014 | Import from public-domain | Gao,X. |
| EPI1619654 | NP | China | 2014-Oct-18 | EPI_ISL_397342 | A/Eurasian curlew/Liaoning/ZH-186/2014 | Import from public-domain | Gao,X. |
| EPI1619656 | MP | China | 2014-Oct-18 | EPI_ISL_397342 | A/Eurasian curlew/Liaoning/ZH-186/2014 | Import from public-domain | Gao,X. |
| EPI2142813 | NP | United States | 2019-Sep-24 | EPI_ISL_14767366 | A/glaucous-winged gull/Alaska/AK19-396/2019 | Import from public-domain | Reeves,A.B.; Ramey,A.M.; Poulson,R.L.; Stallknecht,D.E. |
| EPI2142815 | MP | United States | 2019-Sep-24 | EPI_ISL_14767366 | A/glaucous-winged gull/Alaska/AK19-396/2019 | Import from public-domain | Reeves,A.B.; Ramey,A.M.; Poulson,R.L.; Stallknecht,D.E. |
| EPI2142816 | NS | United States | 2019-Sep-24 | EPI_ISL_14767366 | A/glaucous-winged gull/Alaska/AK19-396/2019 | Import from public-domain | Reeves,A.B.; Ramey,A.M.; Poulson,R.L.; Stallknecht,D.E. |
| EPI2142805 | NP | United States | 2019-Sep-24 | EPI_ISL_14767365 | A/glaucous-winged gull/Alaska/AK19-391/2019 | Import from public-domain | Reeves,A.B.; Ramey,A.M.; Poulson,R.L.; Stallknecht,D.E. |
| EPI2142808 | NS | United States | 2019-Sep-24 | EPI_ISL_14767365 | A/glaucous-winged gull/Alaska/AK19-391/2019 | Import from public-domain | Reeves,A.B.; Ramey,A.M.; Poulson,R.L.; Stallknecht,D.E. |
| EPI2142807 | MP | United States | 2019-Sep-24 | EPI_ISL_14767365 | A/glaucous-winged gull/Alaska/AK19-391/2019 | Import from public-domain | Reeves,A.B.; Ramey,A.M.; Poulson,R.L.; Stallknecht,D.E. |
| EPI2142800 | NS | United States | 2019-Sep-24 | EPI_ISL_14767322 | A/glaucous-winged gull/Alaska/AK19-381/2019 | Import from public-domain | Reeves,A.B.; Ramey,A.M.; Poulson,R.L.; Stallknecht,D.E. |
| EPI2142797 | NP | United States | 2019-Sep-24 | EPI_ISL_14767322 | A/glaucous-winged gull/Alaska/AK19-381/2019 | Import from public-domain | Reeves,A.B.; Ramey,A.M.; Poulson,R.L.; Stallknecht,D.E. |
| EPI2142799 | MP | United States | 2019-Sep-24 | EPI_ISL_14767322 | A/glaucous-winged gull/Alaska/AK19-381/2019 | Import from public-domain | Reeves,A.B.; Ramey,A.M.; Poulson,R.L.; Stallknecht,D.E. |
| EPI2142789 | NP | United States | 2018-Sep-25 | EPI_ISL_14767321 | A/glaucous-winged gull/Alaska/AK18-288/2018 | Import from public-domain | Reeves,A.B.; Ramey,A.M.; Poulson,R.L.; Stallknecht,D.E. |
| EPI2142791 | MP | United States | 2018-Sep-25 | EPI_ISL_14767321 | A/glaucous-winged gull/Alaska/AK18-288/2018 | Import from public-domain | Reeves,A.B.; Ramey,A.M.; Poulson,R.L.; Stallknecht,D.E. |
| EPI2142792 | NS | United States | 2018-Sep-25 | EPI_ISL_14767321 | A/glaucous-winged gull/Alaska/AK18-288/2018 | Import from public-domain | Reeves,A.B.; Ramey,A.M.; Poulson,R.L.; Stallknecht,D.E. |
| EPI2403229 | MP | United States | 2022-May-31 | EPI_ISL_16920549 | A/Red knot/Delaware/1087/2022 | Import from public-domain | Direct Submission |
| EPI2403227 | NS | United States | 2022-May-31 | EPI_ISL_16920549 | A/Red knot/Delaware/1087/2022 | Import from public-domain | Direct Submission |
| EPI2403230 | NP | United States | 2022-May-31 | EPI_ISL_16920549 | A/Red knot/Delaware/1087/2022 | Import from public-domain | Direct Submission |

| | | | | | | | |
|---|---|---|---|---|---|---|---|
| EPI2401457 | NS | United States | 2022-May-31 | EPI_ISL_16919118 | A/Red knot/Delaware/1058/2022 | Import from public-domain | Direct Submission |
| EPI2401459 | MP | United States | 2022-May-31 | EPI_ISL_16919118 | A/Red knot/Delaware/1058/2022 | Import from public-domain | Direct Submission |
| EPI2401460 | NP | United States | 2022-May-31 | EPI_ISL_16919118 | A/Red knot/Delaware/1058/2022 | Import from public-domain | Direct Submission |
| EPI2147876 | MP | United States | 2021-Jun-01 | EPI_ISL_14777719 | A/Red Knot/Delaware/224/2021 | Import from public-domain | Direct Submission |
| EPI2147877 | NP | United States | 2021-Jun-01 | EPI_ISL_14777719 | A/Red Knot/Delaware/224/2021 | Import from public-domain | Direct Submission |
| EPI2147878 | NS | United States | 2021-Jun-01 | EPI_ISL_14777719 | A/Red Knot/Delaware/224/2021 | Import from public-domain | Direct Submission |
| EPI1895236 | MP | United States | 2020-May-29 | EPI_ISL_4061163 | A/Red Knot/Delaware Bay/420/2020 | Import from public-domain | Direct Submission |
| EPI1895235 | NP | United States | 2020-May-29 | EPI_ISL_4061163 | A/Red Knot/Delaware Bay/420/2020 | Import from public-domain | Direct Submission |
| EPI1895234 | NS | United States | 2020-May-29 | EPI_ISL_4061163 | A/Red Knot/Delaware Bay/420/2020 | Import from public-domain | Direct Submission |
| EPI1894735 | MP | United States | 2020-May-29 | EPI_ISL_4056657 | A/Red Knot/Delaware Bay/605/2020 | Import from public-domain | Direct Submission |
| EPI1894738 | NS | United States | 2020-May-29 | EPI_ISL_4056657 | A/Red Knot/Delaware Bay/605/2020 | Import from public-domain | Direct Submission |
| EPI1894739 | NP | United States | 2020-May-29 | EPI_ISL_4056657 | A/Red Knot/Delaware Bay/605/2020 | Import from public-domain | Direct Submission |
| EPI2403144 | MP | United States | 2022-May-31 | EPI_ISL_16920538 | A/Sanderling/Delaware/192/2022 | Import from public-domain | Direct Submission |
| EPI2403146 | NP | United States | 2022-May-31 | EPI_ISL_16920538 | A/Sanderling/Delaware/192/2022 | Import from public-domain | Direct Submission |
| EPI2403147 | NS | United States | 2022-May-31 | EPI_ISL_16920538 | A/Sanderling/Delaware/192/2022 | Import from public-domain | Direct Submission |
| EPI1301114 | NS | Chile | 2016-Dec-13 | EPI_ISL_327600 | A/whimbrel/Chile/C20073/2016 | Import from public-domain | Bravo-Vasquez,N.; Bahl,J.; Hamilton-West,C.; Schultz-Cherry,S. |
| EPI1301122 | NP | Chile | 2016-Dec-13 | EPI_ISL_327600 | A/whimbrel/Chile/C20073/2016 | Import from public-domain | Bravo-Vasquez,N.; Bahl,J.; Hamilton-West,C.; Schultz-Cherry,S. |
| EPI1301139 | MP | Chile | 2016-Dec-13 | EPI_ISL_327600 | A/whimbrel/Chile/C20073/2016 | Import from public-domain | Bravo-Vasquez,N.; Bahl,J.; Hamilton-West,C.; Schultz-Cherry,S. |
| EPI1301125 | NP | Chile | 2016-Dec-13 | EPI_ISL_327599 | A/whimbrel/Chile/C20075/2016 | Import from public-domain | Bravo-Vasquez,N.; Bahl,J.; Hamilton-West,C.; Schultz-Cherry,S. |
| EPI1301136 | NS | Chile | 2016-Dec-13 | EPI_ISL_327599 | A/whimbrel/Chile/C20075/2016 | Import from public-domain | Bravo-Vasquez,N.; Bahl,J.; Hamilton-West,C.; Schultz-Cherry,S. |
| EPI1301145 | MP | Chile | 2016-Dec-13 | EPI_ISL_327599 | A/whimbrel/Chile/C20075/2016 | Import from public-domain | Bravo-Vasquez,N.; Bahl,J.; Hamilton-West,C.; Schultz-Cherry,S. |
| EPI1301106 | MP | Chile | 2016-Dec-13 | EPI_ISL_327598 | A/whimbrel/Chile/C20106/2016 | Import from public-domain | Bravo-Vasquez,N.; Bahl,J.; Hamilton-West,C.; Schultz-Cherry,S. |
| EPI1301107 | NS | Chile | 2016-Dec-13 | EPI_ISL_327598 | A/whimbrel/Chile/C20106/2016 | Import from public-domain | Bravo-Vasquez,N.; Bahl,J.; Hamilton-West,C.; Schultz-Cherry,S. |
| EPI1301115 | NP | Chile | 2016-Dec-13 | EPI_ISL_327598 | A/whimbrel/Chile/C20106/2016 | Import from public-domain | Bravo-Vasquez,N.; Bahl,J.; Hamilton-West,C.; Schultz-Cherry,S. |
| EPI1301084 | NP | Chile | 2016-Dec-13 | EPI_ISL_327597 | A/whimbrel/Chile/C20144/2016 | Import from public-domain | Bravo-Vasquez,N.; Bahl,J.; Hamilton-West,C.; Schultz-Cherry,S. |
| EPI1301113 | MP | Chile | 2016-Dec-13 | EPI_ISL_327597 | A/whimbrel/Chile/C20144/2016 | Import from public-domain | Bravo-Vasquez,N.; Bahl,J.; Hamilton-West,C.; Schultz-Cherry,S. |
| EPI1301121 | NS | Chile | 2016-Dec-13 | EPI_ISL_327597 | A/whimbrel/Chile/C20144/2016 | Import from public-domain | Bravo-Vasquez,N.; Bahl,J.; Hamilton-West,C.; Schultz-Cherry,S. |
| EPI1301091 | MP | Chile | 2016-Dec-13 | EPI_ISL_327595 | A/whimbrel/Chile/C20147/2016 | Import from public-domain | Bravo-Vasquez,N.; Bahl,J.; Hamilton-West,C.; Schultz-Cherry,S. |
| EPI1301100 | NS | Chile | 2016-Dec-13 | EPI_ISL_327595 | A/whimbrel/Chile/C20147/2016 | Import from public-domain | Bravo-Vasquez,N.; Bahl,J.; Hamilton-West,C.; Schultz-Cherry,S. |
| EPI1301065 | NP | Chile | 2016-Dec-13 | EPI_ISL_327595 | A/whimbrel/Chile/C20147/2016 | Import from public-domain | Bravo-Vasquez,N.; Bahl,J.; Hamilton-West,C.; Schultz-Cherry,S. |
| EPI1298543 | NS | Chile | 2016-Apr-27 | EPI_ISL_326975 | A/whimbrel/Chile/C10834/2016 | Import from public-domain | Jimenez-Bluhm,P.; Hamilton,C.; van Bakel,H.; Kriti,D.; Schultz-Cherry,S. |
| EPI1298525 | MP | Chile | 2016-Apr-27 | EPI_ISL_326975 | A/whimbrel/Chile/C10834/2016 | Import from public-domain | Jimenez-Bluhm,P.; Hamilton,C.; van Bakel,H.; Kriti,D.; Schultz-Cherry,S. |
| EPI1298479 | NP | Chile | 2016-Apr-27 | EPI_ISL_326975 | A/whimbrel/Chile/C10834/2016 | Import from public-domain | Jimenez-Bluhm,P.; Hamilton,C.; van Bakel,H.; Kriti,D.; Schultz-Cherry,S. |
| EPI1298530 | NP | Chile | 2016-May-31 | EPI_ISL_326973 | A/whimbrel/Chile/C12523/2016 | Import from public-domain | Jimenez-Bluhm,P.; Hamilton,C.; van Bakel,H.; Kriti,D.; Schultz-Cherry,S. |
| EPI1298551 | MP | Chile | 2016-May-31 | EPI_ISL_326973 | A/whimbrel/Chile/C12523/2016 | Import from public-domain | Jimenez-Bluhm,P.; Hamilton,C.; van Bakel,H.; Kriti,D.; Schultz-Cherry,S. |

| | | | | | | | | |
|---|---|---|---|---|---|---|---|---|
| EPI1298495 | NS | Chile | 2016-May-31 | EPI_ISL_326973 | A/whimbrel/Chile/C12523/2016 | | Import from public-domain | Jimenez-Bluhm,P.; Hamilton,C.; van Bakel,H.; Kriti,D.; Schultz-Cherry,S. |
| EPI1942891 | NP | Belgium | 2021-Dec-09 | EPI_ISL_7622539 | A/Chroicocephalus_ridibundus/Belgium/13464/2020 | Sciensano - Animal Infectious Diseases | Sciensano, Department of Animal Infectious Diseases | Van Borm, Steven; Vandenbussche, Frank; Roupie, Virginie; Lambrecht, Benedicte; Steensels, Mieke |
| EPI1942893 | MP | Belgium | 2021-Dec-09 | EPI_ISL_7622539 | A/Chroicocephalus_ridibundus/Belgium/13464/2020 | Sciensano - Animal Infectious Diseases | Sciensano, Department of Animal Infectious Diseases | Van Borm, Steven; Vandenbussche, Frank; Roupie, Virginie; Lambrecht, Benedicte; Steensels, Mieke |
| EPI1942894 | NS | Belgium | 2021-Dec-09 | EPI_ISL_7622539 | A/Chroicocephalus_ridibundus/Belgium/13464/2020 | Sciensano - Animal Infectious Diseases | Sciensano, Department of Animal Infectious Diseases | Van Borm, Steven; Vandenbussche, Frank; Roupie, Virginie; Lambrecht, Benedicte; Steensels, Mieke |
| EPI1307697 | NS | Netherlands | 2014-Jul-11 | EPI_ISL_329049 | A/black-headed gull/Netherlands/31/2014 | | Import from public-domain | Poen,M.J.; Verhagen,J.H.; Vuong,O.; Scheuer,R.D.; Pas,S.D.; Fouchier,R.A.M. |
| EPI1307662 | NP | Netherlands | 2014-Jul-11 | EPI_ISL_329049 | A/black-headed gull/Netherlands/31/2014 | | Import from public-domain | Poen,M.J.; Verhagen,J.H.; Vuong,O.; Scheuer,R.D.; Pas,S.D.; Fouchier,R.A.M. |
| EPI1307665 | MP | Netherlands | 2014-Jul-11 | EPI_ISL_329049 | A/black-headed gull/Netherlands/31/2014 | | Import from public-domain | Poen,M.J.; Verhagen,J.H.; Vuong,O.; Scheuer,R.D.; Pas,S.D.; Fouchier,R.A.M. |
| EPI1307681 | NS | Netherlands | 2014-Jul-24 | EPI_ISL_329047 | A/black-headed gull/Netherlands/36/2014 | | Import from public-domain | Poen,M.J.; Verhagen,J.H.; Vuong,O.; Scheuer,R.D.; Pas,S.D.; Fouchier,R.A.M. |
| EPI1307690 | NP | Netherlands | 2014-Jul-24 | EPI_ISL_329047 | A/black-headed gull/Netherlands/36/2014 | | Import from public-domain | Poen,M.J.; Verhagen,J.H.; Vuong,O.; Scheuer,R.D.; Pas,S.D.; Fouchier,R.A.M. |
| EPI1307652 | MP | Netherlands | 2014-Jul-24 | EPI_ISL_329047 | A/black-headed gull/Netherlands/36/2014 | | Import from public-domain | Poen,M.J.; Verhagen,J.H.; Vuong,O.; Scheuer,R.D.; Pas,S.D.; Fouchier,R.A.M. |
| EPI1307655 | NP | Netherlands | 2014-Jul-11 | EPI_ISL_329046 | A/black-headed gull/Netherlands/34/2014 | | Import from public-domain | Poen,M.J.; Verhagen,J.H.; Vuong,O.; Scheuer,R.D.; Pas,S.D.; Fouchier,R.A.M. |
| EPI1307666 | NS | Netherlands | 2014-Jul-11 | EPI_ISL_329046 | A/black-headed gull/Netherlands/34/2014 | | Import from public-domain | Poen,M.J.; Verhagen,J.H.; Vuong,O.; Scheuer,R.D.; Pas,S.D.; Fouchier,R.A.M. |
| EPI1307667 | MP | Netherlands | 2014-Jul-11 | EPI_ISL_329046 | A/black-headed gull/Netherlands/34/2014 | | Import from public-domain | Poen,M.J.; Verhagen,J.H.; Vuong,O.; Scheuer,R.D.; Pas,S.D.; Fouchier,R.A.M. |
| EPI1307678 | MP | Netherlands | 2014-Jul-11 | EPI_ISL_329045 | A/black-headed gull/Netherlands/33/2014 | | Import from public-domain | Poen,M.J.; Verhagen,J.H.; Vuong,O.; Scheuer,R.D.; Pas,S.D.; Fouchier,R.A.M. |
| EPI1307684 | NP | Netherlands | 2014-Jul-11 | EPI_ISL_329045 | A/black-headed gull/Netherlands/33/2014 | | Import from public-domain | Poen,M.J.; Verhagen,J.H.; Vuong,O.; Scheuer,R.D.; Pas,S.D.; Fouchier,R.A.M. |
| EPI1307649 | NS | Netherlands | 2014-Jul-11 | EPI_ISL_329045 | A/black-headed gull/Netherlands/33/2014 | | Import from public-domain | Poen,M.J.; Verhagen,J.H.; Vuong,O.; Scheuer,R.D.; Pas,S.D.; Fouchier,R.A.M. |
| EPI1307672 | NP | Netherlands | 2014-Jul-09 | EPI_ISL_329044 | A/black-headed gull/Netherlands/4/2014 | | Import from public-domain | Poen,M.J.; Verhagen,J.H.; Vuong,O.; Scheuer,R.D.; Pas,S.D.; Fouchier,R.A.M. |
| EPI1307687 | NS | Netherlands | 2014-Jul-09 | EPI_ISL_329044 | A/black-headed gull/Netherlands/4/2014 | | Import from public-domain | Poen,M.J.; Verhagen,J.H.; Vuong,O.; Scheuer,R.D.; Pas,S.D.; Fouchier,R.A.M. |
| EPI1307651 | MP | Netherlands | 2014-Jul-09 | EPI_ISL_329044 | A/black-headed gull/Netherlands/4/2014 | | Import from public-domain | Poen,M.J.; Verhagen,J.H.; Vuong,O.; Scheuer,R.D.; Pas,S.D.; Fouchier,R.A.M. |
| EPI1307174 | NP | Netherlands | 2016-Jul-15 | EPI_ISL_328991 | A/black-headed gull/Netherlands/1/2016 | | Import from public-domain | Poen,M.J.; Verhagen,J.H.; Vuong,O.; Scheuer,R.D.; Pas,S.D.; Fouchier,R.A.M. |
| EPI1307182 | MP | Netherlands | 2016-Jul-15 | EPI_ISL_328991 | A/black-headed gull/Netherlands/1/2016 | | Import from public-domain | Poen,M.J.; Verhagen,J.H.; Vuong,O.; Scheuer,R.D.; Pas,S.D.; Fouchier,R.A.M. |
| EPI1307189 | NS | Netherlands | 2016-Jul-15 | EPI_ISL_328991 | A/black-headed gull/Netherlands/1/2016 | | Import from public-domain | Poen,M.J.; Verhagen,J.H.; Vuong,O.; Scheuer,R.D.; Pas,S.D.; Fouchier,R.A.M. |
| EPI1307166 | MP | Netherlands | 2016-Aug-06 | EPI_ISL_328990 | A/black-headed gull/Netherlands/10/2016 | | Import from public-domain | Poen,M.J.; Verhagen,J.H.; Vuong,O.; Scheuer,R.D.; Pas,S.D.; Fouchier,R.A.M. |
| EPI1307181 | NP | Netherlands | 2016-Aug-06 | EPI_ISL_328990 | A/black-headed gull/Netherlands/10/2016 | | Import from public-domain | Poen,M.J.; Verhagen,J.H.; Vuong,O.; Scheuer,R.D.; Pas,S.D.; Fouchier,R.A.M. |
| EPI1307188 | NS | Netherlands | 2016-Aug-06 | EPI_ISL_328990 | A/black-headed gull/Netherlands/10/2016 | | Import from public-domain | Poen,M.J.; Verhagen,J.H.; Vuong,O.; Scheuer,R.D.; Pas,S.D.; Fouchier,R.A.M. |
| EPI1307195 | NS | Netherlands | 2016-Jul-15 | EPI_ISL_328989 | A/black-headed gull/Netherlands/2/2016 | | Import from public-domain | Poen,M.J.; Verhagen,J.H.; Vuong,O.; Scheuer,R.D.; Pas,S.D.; Fouchier,R.A.M. |
| EPI1307163 | MP | Netherlands | 2016-Jul-15 | EPI_ISL_328989 | A/black-headed gull/Netherlands/2/2016 | | Import from public-domain | Poen,M.J.; Verhagen,J.H.; Vuong,O.; Scheuer,R.D.; Pas,S.D.; Fouchier,R.A.M. |
| EPI1307172 | NP | Netherlands | 2016-Jul-15 | EPI_ISL_328989 | A/black-headed gull/Netherlands/2/2016 | | Import from public-domain | Poen,M.J.; Verhagen,J.H.; Vuong,O.; Scheuer,R.D.; Pas,S.D.; Fouchier,R.A.M. |
| EPI1307161 | MP | Netherlands | 2016-Aug-06 | EPI_ISL_328988 | A/black-headed gull/Netherlands/11/2016 | | Import from public-domain | Poen,M.J.; Verhagen,J.H.; Vuong,O.; Scheuer,R.D.; Pas,S.D.; Fouchier,R.A.M. |
| EPI1307175 | NP | Netherlands | 2016-Aug-06 | EPI_ISL_328988 | A/black-headed gull/Netherlands/11/2016 | | Import from public-domain | Poen,M.J.; Verhagen,J.H.; Vuong,O.; Scheuer,R.D.; Pas,S.D.; Fouchier,R.A.M. |
| EPI1307187 | NS | Netherlands | 2016-Aug-06 | EPI_ISL_328988 | A/black-headed gull/Netherlands/11/2016 | | Import from public-domain | Poen,M.J.; Verhagen,J.H.; Vuong,O.; Scheuer,R.D.; Pas,S.D.; Fouchier,R.A.M. |
| EPI1307133 | MP | Netherlands | 2016-Aug-06 | EPI_ISL_328984 | A/black-headed gull/Netherlands/8/2016 | | Import from public-domain | Poen,M.J.; Verhagen,J.H.; Vuong,O.; Scheuer,R.D.; Pas,S.D.; Fouchier,R.A.M. |
| EPI1307139 | NS | Netherlands | 2016-Aug-06 | EPI_ISL_328984 | A/black-headed gull/Netherlands/8/2016 | | Import from public-domain | Poen,M.J.; Verhagen,J.H.; Vuong,O.; Scheuer,R.D.; Pas,S.D.; Fouchier,R.A.M. |
| EPI1307111 | NP | Netherlands | 2016-Aug-06 | EPI_ISL_328984 | A/black-headed gull/Netherlands/8/2016 | | Import from public-domain | Poen,M.J.; Verhagen,J.H.; Vuong,O.; Scheuer,R.D.; Pas,S.D.; Fouchier,R.A.M. |
| EPI1307126 | MP | Netherlands | 2016-Jul-15 | EPI_ISL_328983 | A/black-headed gull/Netherlands/6/2016 | | Import from public-domain | Poen,M.J.; Verhagen,J.H.; Vuong,O.; Scheuer,R.D.; Pas,S.D.; Fouchier,R.A.M. |

| | | | | | | | |
|---|---|---|---|---|---|---|---|
| EPI1307131 | NS | Netherlands | 2016-Jul-15 | EPI_ISL_328983 | A/black-headed gull/Netherlands/6/2016 | Import from public-domain | Poen,M.J.; Verhagen,J.H.; Vuong,O.; Scheuer,R.D.; Pas,S.D.; Fouchier,R.A.M. |
| EPI1307140 | NP | Netherlands | 2016-Jul-15 | EPI_ISL_328983 | A/black-headed gull/Netherlands/6/2016 | Import from public-domain | Poen,M.J.; Verhagen,J.H.; Vuong,O.; Scheuer,R.D.; Pas,S.D.; Fouchier,R.A.M. |
| EPI1307130 | NP | Netherlands | 2016-Jul-15 | EPI_ISL_328981 | A/black-headed gull/Netherlands/3/2016 | Import from public-domain | Poen,M.J.; Verhagen,J.H.; Vuong,O.; Scheuer,R.D.; Pas,S.D.; Fouchier,R.A.M. |
| EPI1307102 | MP | Netherlands | 2016-Jul-15 | EPI_ISL_328981 | A/black-headed gull/Netherlands/3/2016 | Import from public-domain | Poen,M.J.; Verhagen,J.H.; Vuong,O.; Scheuer,R.D.; Pas,S.D.; Fouchier,R.A.M. |
| EPI1307159 | NS | Netherlands | 2016-Jul-15 | EPI_ISL_328981 | A/black-headed gull/Netherlands/3/2016 | Import from public-domain | Poen,M.J.; Verhagen,J.H.; Vuong,O.; Scheuer,R.D.; Pas,S.D.; Fouchier,R.A.M. |
| EPI1307124 | NS | Netherlands | 2016-Jun-29 | EPI_ISL_328980 | A/black-headed gull/Netherlands/7/2016 | Import from public-domain | Poen,M.J.; Verhagen,J.H.; Vuong,O.; Scheuer,R.D.; Pas,S.D.; Fouchier,R.A.M. |
| EPI1307112 | MP | Netherlands | 2016-Jun-29 | EPI_ISL_328980 | A/black-headed gull/Netherlands/7/2016 | Import from public-domain | Poen,M.J.; Verhagen,J.H.; Vuong,O.; Scheuer,R.D.; Pas,S.D.; Fouchier,R.A.M. |
| EPI1307114 | NP | Netherlands | 2016-Jun-29 | EPI_ISL_328980 | A/black-headed gull/Netherlands/7/2016 | Import from public-domain | Poen,M.J.; Verhagen,J.H.; Vuong,O.; Scheuer,R.D.; Pas,S.D.; Fouchier,R.A.M. |
| EPI1307093 | NP | Netherlands | 2016-Aug-06 | EPI_ISL_328978 | A/black-headed gull/Netherlands/9/2016 | Import from public-domain | Poen,M.J.; Verhagen,J.H.; Vuong,O.; Scheuer,R.D.; Pas,S.D.; Fouchier,R.A.M. |
| EPI1307096 | NS | Netherlands | 2016-Aug-06 | EPI_ISL_328978 | A/black-headed gull/Netherlands/9/2016 | Import from public-domain | Poen,M.J.; Verhagen,J.H.; Vuong,O.; Scheuer,R.D.; Pas,S.D.; Fouchier,R.A.M. |
| EPI1307098 | MP | Netherlands | 2016-Aug-06 | EPI_ISL_328978 | A/black-headed gull/Netherlands/9/2016 | Import from public-domain | Poen,M.J.; Verhagen,J.H.; Vuong,O.; Scheuer,R.D.; Pas,S.D.; Fouchier,R.A.M. |
| EPI1014017 | NS | Netherlands | 2014-Jun-17 | EPI_ISL_267793 | A/black-headed gull/Netherlands/37/2014 | Import from public-domain | |
| EPI1014138 | NP | Netherlands | 2014-Jun-17 | EPI_ISL_267793 | A/black-headed gull/Netherlands/37/2014 | Import from public-domain | |
| EPI1014702 | MP | Netherlands | 2014-Jun-17 | EPI_ISL_267793 | A/black-headed gull/Netherlands/37/2014 | Import from public-domain | |
| EPI1013490 | NP | Netherlands | 2013-Jul-17 | EPI_ISL_267789 | A/black-headed gull/Netherlands/8/2013 | Import from public-domain | |
| EPI1014802 | NS | Netherlands | 2013-Jul-17 | EPI_ISL_267789 | A/black-headed gull/Netherlands/8/2013 | Import from public-domain | |
| EPI1014896 | MP | Netherlands | 2013-Jul-17 | EPI_ISL_267789 | A/black-headed gull/Netherlands/8/2013 | Import from public-domain | |
| EPI1014046 | NS | Netherlands | 2014-Dec-24 | EPI_ISL_267770 | A/black-headed gull/Netherlands/18/2014 | Import from public-domain | |
| EPI1013701 | NP | Netherlands | 2014-Dec-24 | EPI_ISL_267770 | A/black-headed gull/Netherlands/18/2014 | Import from public-domain | |
| EPI1013880 | MP | Netherlands | 2014-Dec-24 | EPI_ISL_267770 | A/black-headed gull/Netherlands/18/2014 | Import from public-domain | |
| EPI1013375 | NS | Netherlands | 2014-Jun-23 | EPI_ISL_267768 | A/black-headed gull/Netherlands/7/2014 | Import from public-domain | |
| EPI1013740 | NP | Netherlands | 2014-Jun-23 | EPI_ISL_267768 | A/black-headed gull/Netherlands/7/2014 | Import from public-domain | |
| EPI1013566 | MP | Netherlands | 2014-Jun-23 | EPI_ISL_267768 | A/black-headed gull/Netherlands/7/2014 | Import from public-domain | |
| EPI1011079 | NS | Netherlands | 2014-Jul-09 | EPI_ISL_267413 | A/black-headed gull/Netherlands/9/2014 | Import from public-domain | |
| EPI1014599 | MP | Netherlands | 2014-Jul-09 | EPI_ISL_267413 | A/black-headed gull/Netherlands/9/2014 | Import from public-domain | |
| EPI1014396 | NP | Netherlands | 2014-Jul-09 | EPI_ISL_267413 | A/black-headed gull/Netherlands/9/2014 | Import from public-domain | |
| EPI1011073 | MP | Netherlands | 2013-Jul-06 | EPI_ISL_267411 | A/black-headed gull/Netherlands/6/2013 | Import from public-domain | |
| EPI1014063 | NP | Netherlands | 2013-Jul-06 | EPI_ISL_267411 | A/black-headed gull/Netherlands/6/2013 | Import from public-domain | |
| EPI1013788 | NS | Netherlands | 2013-Jul-06 | EPI_ISL_267411 | A/black-headed gull/Netherlands/6/2013 | Import from public-domain | |
| EPI1010822 | NS | Netherlands | 2014-Jul-11 | EPI_ISL_267343 | A/black-headed gull/Netherlands/14/2014 | Import from public-domain | |
| EPI1010921 | MP | Netherlands | 2014-Jul-11 | EPI_ISL_267343 | A/black-headed gull/Netherlands/14/2014 | Import from public-domain | |
| EPI1013341 | NP | Netherlands | 2014-Jul-11 | EPI_ISL_267343 | A/black-headed gull/Netherlands/14/2014 | Import from public-domain | |
| EPI1010743 | MP | Netherlands | 2013-Jul-17 | EPI_ISL_267305 | A/black-headed gull/Netherlands/9/2013 | Import from public-domain | |
| EPI1013333 | NS | Netherlands | 2013-Jul-17 | EPI_ISL_267305 | A/black-headed gull/Netherlands/9/2013 | Import from public-domain | |
| EPI1014170 | NP | Netherlands | 2013-Jul-17 | EPI_ISL_267305 | A/black-headed gull/Netherlands/9/2013 | Import from public-domain | |

| | | | | | | | |
|---|---|---|---|---|---|---|---|
| EPI1010838 | MP | Netherlands | 2014-Jul-09 | EPI_ISL_267230 | A/black-headed gull/Netherlands/13/2014 | Import from public-domain | |
| EPI1010978 | NS | Netherlands | 2014-Jul-09 | EPI_ISL_267230 | A/black-headed gull/Netherlands/13/2014 | Import from public-domain | |
| EPI1014858 | NP | Netherlands | 2014-Jul-09 | EPI_ISL_267230 | A/black-headed gull/Netherlands/13/2014 | Import from public-domain | |
| EPI1011334 | MP | Netherlands | 2014-Jul-11 | EPI_ISL_267220 | A/black-headed gull/Netherlands/16/2014 | Import from public-domain | |
| EPI1014300 | NP | Netherlands | 2014-Jul-11 | EPI_ISL_267220 | A/black-headed gull/Netherlands/16/2014 | Import from public-domain | |
| EPI1014949 | NS | Netherlands | 2014-Jul-11 | EPI_ISL_267220 | A/black-headed gull/Netherlands/16/2014 | Import from public-domain | |
| EPI1010603 | NS | Netherlands | 2014-Jun-17 | EPI_ISL_267215 | A/black-headed gull/Netherlands/24/2014 | Import from public-domain | |
| EPI1014646 | NP | Netherlands | 2014-Jun-17 | EPI_ISL_267215 | A/black-headed gull/Netherlands/24/2014 | Import from public-domain | |
| EPI1013848 | MP | Netherlands | 2014-Jun-17 | EPI_ISL_267215 | A/black-headed gull/Netherlands/24/2014 | Import from public-domain | |
| EPI1013330 | NP | Netherlands | 2015-Jan-27 | EPI_ISL_267201 | A/black-headed gull/Netherlands/1/2015 | Import from public-domain | |
| EPI1014532 | NS | Netherlands | 2015-Jan-27 | EPI_ISL_267201 | A/black-headed gull/Netherlands/1/2015 | Import from public-domain | |
| EPI1013697 | MP | Netherlands | 2015-Jan-27 | EPI_ISL_267201 | A/black-headed gull/Netherlands/1/2015 | Import from public-domain | |
| EPI1010944 | NS | Netherlands | 2014-Jul-09 | EPI_ISL_267179 | A/black-headed gull/Netherlands/2/2014 | Import from public-domain | |
| EPI1011291 | MP | Netherlands | 2014-Jul-09 | EPI_ISL_267179 | A/black-headed gull/Netherlands/2/2014 | Import from public-domain | |
| EPI1014082 | NP | Netherlands | 2014-Jul-09 | EPI_ISL_267179 | A/black-headed gull/Netherlands/2/2014 | Import from public-domain | |
| EPI890104 | NS | Netherlands | 2014-Jul-11 | EPI_ISL_243662 | A/black-headed gull/Netherlands/29/2014 | Import from public-domain | Poen,M.J.; Verhagen,J.H.; Vuong,O.; Scheuer,R.D.; Pas,S.D.; Fouchier,R.A.M. |
| EPI890910 | MP | Netherlands | 2014-Jul-11 | EPI_ISL_243662 | A/black-headed gull/Netherlands/29/2014 | Import from public-domain | Poen,M.J.; Verhagen,J.H.; Vuong,O.; Scheuer,R.D.; Pas,S.D.; Fouchier,R.A.M. |
| EPI890544 | NP | Netherlands | 2014-Jul-11 | EPI_ISL_243662 | A/black-headed gull/Netherlands/29/2014 | Import from public-domain | Poen,M.J.; Verhagen,J.H.; Vuong,O.; Scheuer,R.D.; Pas,S.D.; Fouchier,R.A.M. |
| EPI890172 | NS | Netherlands | 2014-Jun-27 | EPI_ISL_243645 | A/black-headed gull/Netherlands/5/2014 | Import from public-domain | Poen,M.J.; Verhagen,J.H.; Vuong,O.; Scheuer,R.D.; Pas,S.D.; Fouchier,R.A.M. |
| EPI890985 | MP | Netherlands | 2014-Jun-27 | EPI_ISL_243645 | A/black-headed gull/Netherlands/5/2014 | Import from public-domain | Poen,M.J.; Verhagen,J.H.; Vuong,O.; Scheuer,R.D.; Pas,S.D.; Fouchier,R.A.M. |
| EPI889894 | NP | Netherlands | 2014-Jun-27 | EPI_ISL_243645 | A/black-headed gull/Netherlands/5/2014 | Import from public-domain | Poen,M.J.; Verhagen,J.H.; Vuong,O.; Scheuer,R.D.; Pas,S.D.; Fouchier,R.A.M. |
| EPI889737 | NP | Netherlands | 2014-Jul-11 | EPI_ISL_243631 | A/black-headed gull/Netherlands/35/2014 | Import from public-domain | Poen,M.J.; Verhagen,J.H.; Vuong,O.; Scheuer,R.D.; Pas,S.D.; Fouchier,R.A.M. |
| EPI890722 | NS | Netherlands | 2014-Jul-11 | EPI_ISL_243631 | A/black-headed gull/Netherlands/35/2014 | Import from public-domain | Poen,M.J.; Verhagen,J.H.; Vuong,O.; Scheuer,R.D.; Pas,S.D.; Fouchier,R.A.M. |
| EPI890896 | MP | Netherlands | 2014-Jul-11 | EPI_ISL_243631 | A/black-headed gull/Netherlands/35/2014 | Import from public-domain | Poen,M.J.; Verhagen,J.H.; Vuong,O.; Scheuer,R.D.; Pas,S.D.; Fouchier,R.A.M. |
| EPI890070 | MP | Netherlands | 2014-Jun-23 | EPI_ISL_243628 | A/black-headed gull/Netherlands/8/2014 | Import from public-domain | Poen,M.J.; Verhagen,J.H.; Vuong,O.; Scheuer,R.D.; Pas,S.D.; Fouchier,R.A.M. |
| EPI890110 | NS | Netherlands | 2014-Jun-23 | EPI_ISL_243628 | A/black-headed gull/Netherlands/8/2014 | Import from public-domain | Poen,M.J.; Verhagen,J.H.; Vuong,O.; Scheuer,R.D.; Pas,S.D.; Fouchier,R.A.M. |
| EPI889724 | NP | Netherlands | 2014-Jun-23 | EPI_ISL_243628 | A/black-headed gull/Netherlands/8/2014 | Import from public-domain | Poen,M.J.; Verhagen,J.H.; Vuong,O.; Scheuer,R.D.; Pas,S.D.; Fouchier,R.A.M. |
| EPI890498 | NP | Netherlands | 2014-Jun-17 | EPI_ISL_243616 | A/black-headed gull/Netherlands/25/2014 | Import from public-domain | Poen,M.J.; Verhagen,J.H.; Vuong,O.; Scheuer,R.D.; Pas,S.D.; Fouchier,R.A.M. |
| EPI890909 | NS | Netherlands | 2014-Jun-17 | EPI_ISL_243616 | A/black-headed gull/Netherlands/25/2014 | Import from public-domain | Poen,M.J.; Verhagen,J.H.; Vuong,O.; Scheuer,R.D.; Pas,S.D.; Fouchier,R.A.M. |
| EPI890050 | MP | Netherlands | 2014-Jun-17 | EPI_ISL_243616 | A/black-headed gull/Netherlands/25/2014 | Import from public-domain | Poen,M.J.; Verhagen,J.H.; Vuong,O.; Scheuer,R.D.; Pas,S.D.; Fouchier,R.A.M. |
| EPI889646 | MP | Netherlands | 2014-Jul-11 | EPI_ISL_243587 | A/black-headed gull/Netherlands/15/2014 | Import from public-domain | Poen,M.J.; Verhagen,J.H.; Vuong,O.; Scheuer,R.D.; Pas,S.D.; Fouchier,R.A.M. |
| EPI890729 | NS | Netherlands | 2014-Jul-11 | EPI_ISL_243587 | A/black-headed gull/Netherlands/15/2014 | Import from public-domain | Poen,M.J.; Verhagen,J.H.; Vuong,O.; Scheuer,R.D.; Pas,S.D.; Fouchier,R.A.M. |
| EPI890002 | NP | Netherlands | 2014-Jul-11 | EPI_ISL_243587 | A/black-headed gull/Netherlands/15/2014 | Import from public-domain | Poen,M.J.; Verhagen,J.H.; Vuong,O.; Scheuer,R.D.; Pas,S.D.; Fouchier,R.A.M. |
| EPI890068 | MP | Netherlands | 2013-Jul-06 | EPI_ISL_243564 | A/black-headed gull/Netherlands/4/2013 | Import from public-domain | Poen,M.J.; Verhagen,J.H.; Vuong,O.; Scheuer,R.D.; Pas,S.D.; Fouchier,R.A.M. |
| EPI889513 | NP | Netherlands | 2013-Jul-06 | EPI_ISL_243564 | A/black-headed gull/Netherlands/4/2013 | Import from public-domain | Poen,M.J.; Verhagen,J.H.; Vuong,O.; Scheuer,R.D.; Pas,S.D.; Fouchier,R.A.M. |

| | | | | | | | |
|---|---|---|---|---|---|---|---|
| EPI891056 | NS | Netherlands | 2013-Jul-06 | EPI_ISL_243564 | A/black-headed gull/Netherlands/4/2013 | Import from public-domain | Poen,M.J.; Verhagen,J.H.; Vuong,O.; Scheuer,R.D.; Pas,S.D.; Fouchier,R.A.M. |
| EPI891190 | NS | Netherlands | 2014-Jul-09 | EPI_ISL_243562 | A/black-headed gull/Netherlands/1/2014 | Import from public-domain | Poen,M.J.; Verhagen,J.H.; Vuong,O.; Scheuer,R.D.; Pas,S.D.; Fouchier,R.A.M. |
| EPI890901 | NP | Netherlands | 2014-Jul-09 | EPI_ISL_243562 | A/black-headed gull/Netherlands/1/2014 | Import from public-domain | Poen,M.J.; Verhagen,J.H.; Vuong,O.; Scheuer,R.D.; Pas,S.D.; Fouchier,R.A.M. |
| EPI889558 | MP | Netherlands | 2014-Jul-09 | EPI_ISL_243562 | A/black-headed gull/Netherlands/1/2014 | Import from public-domain | Poen,M.J.; Verhagen,J.H.; Vuong,O.; Scheuer,R.D.; Pas,S.D.; Fouchier,R.A.M. |
| EPI890894 | NS | Netherlands | 2015-Jun-30 | EPI_ISL_243560 | A/black-headed gull/Netherlands/2/2015 | Import from public-domain | Poen,M.J.; Verhagen,J.H.; Vuong,O.; Scheuer,R.D.; Pas,S.D.; Fouchier,R.A.M. |
| EPI890118 | MP | Netherlands | 2015-Jun-30 | EPI_ISL_243560 | A/black-headed gull/Netherlands/2/2015 | Import from public-domain | Poen,M.J.; Verhagen,J.H.; Vuong,O.; Scheuer,R.D.; Pas,S.D.; Fouchier,R.A.M. |
| EPI889505 | NP | Netherlands | 2015-Jun-30 | EPI_ISL_243560 | A/black-headed gull/Netherlands/2/2015 | Import from public-domain | Poen,M.J.; Verhagen,J.H.; Vuong,O.; Scheuer,R.D.; Pas,S.D.; Fouchier,R.A.M. |
| EPI899583 | NS | Netherlands | 2014-Jun-17 | EPI_ISL_243556 | A/black-headed gull/Netherlands/23/2014 | Import from public-domain | Poen,M.J.; Verhagen,J.H.; Vuong,O.; Scheuer,R.D.; Pas,S.D.; Fouchier,R.A.M. |
| EPI889496 | NP | Netherlands | 2014-Jun-17 | EPI_ISL_243556 | A/black-headed gull/Netherlands/23/2014 | Import from public-domain | Poen,M.J.; Verhagen,J.H.; Vuong,O.; Scheuer,R.D.; Pas,S.D.; Fouchier,R.A.M. |
| EPI890983 | MP | Netherlands | 2014-Jun-17 | EPI_ISL_243556 | A/black-headed gull/Netherlands/23/2014 | Import from public-domain | Poen,M.J.; Verhagen,J.H.; Vuong,O.; Scheuer,R.D.; Pas,S.D.; Fouchier,R.A.M. |
| EPI890221 | NP | Netherlands | 2013-Jul-06 | EPI_ISL_243555 | A/black-headed gull/Netherlands/5/2013 | Import from public-domain | Poen,M.J.; Verhagen,J.H.; Vuong,O.; Scheuer,R.D.; Pas,S.D.; Fouchier,R.A.M. |
| EPI889493 | MP | Netherlands | 2013-Jul-06 | EPI_ISL_243555 | A/black-headed gull/Netherlands/5/2013 | Import from public-domain | Poen,M.J.; Verhagen,J.H.; Vuong,O.; Scheuer,R.D.; Pas,S.D.; Fouchier,R.A.M. |
| EPI889804 | NS | Netherlands | 2013-Jul-06 | EPI_ISL_243555 | A/black-headed gull/Netherlands/5/2013 | Import from public-domain | Poen,M.J.; Verhagen,J.H.; Vuong,O.; Scheuer,R.D.; Pas,S.D.; Fouchier,R.A.M. |
| EPI890825 | MP | Netherlands | 2014-Jul-04 | EPI_ISL_243554 | A/black-headed gull/Netherlands/28/2014 | Import from public-domain | Poen,M.J.; Verhagen,J.H.; Vuong,O.; Scheuer,R.D.; Pas,S.D.; Fouchier,R.A.M. |
| EPI889489 | NS | Netherlands | 2014-Jul-04 | EPI_ISL_243554 | A/black-headed gull/Netherlands/28/2014 | Import from public-domain | Poen,M.J.; Verhagen,J.H.; Vuong,O.; Scheuer,R.D.; Pas,S.D.; Fouchier,R.A.M. |
| EPI890412 | NP | Netherlands | 2014-Jul-04 | EPI_ISL_243554 | A/black-headed gull/Netherlands/28/2014 | Import from public-domain | Poen,M.J.; Verhagen,J.H.; Vuong,O.; Scheuer,R.D.; Pas,S.D.; Fouchier,R.A.M. |
| EPI890770 | MP | Netherlands | 2014-Jun-30 | EPI_ISL_243552 | A/black-headed gull/Netherlands/27/2014 | Import from public-domain | Poen,M.J.; Verhagen,J.H.; Vuong,O.; Scheuer,R.D.; Pas,S.D.; Fouchier,R.A.M. |
| EPI890291 | NP | Netherlands | 2014-Jun-30 | EPI_ISL_243552 | A/black-headed gull/Netherlands/27/2014 | Import from public-domain | Poen,M.J.; Verhagen,J.H.; Vuong,O.; Scheuer,R.D.; Pas,S.D.; Fouchier,R.A.M. |
| EPI891167 | NS | Netherlands | 2014-Jun-30 | EPI_ISL_243552 | A/black-headed gull/Netherlands/27/2014 | Import from public-domain | Poen,M.J.; Verhagen,J.H.; Vuong,O.; Scheuer,R.D.; Pas,S.D.; Fouchier,R.A.M. |
| EPI889840 | NS | Netherlands | 2015-Jun-30 | EPI_ISL_243545 | A/black-headed gull/Netherlands/4/2015 | Import from public-domain | Poen,M.J.; Verhagen,J.H.; Vuong,O.; Scheuer,R.D.; Pas,S.D.; Fouchier,R.A.M. |
| EPI889652 | MP | Netherlands | 2015-Jun-30 | EPI_ISL_243545 | A/black-headed gull/Netherlands/4/2015 | Import from public-domain | Poen,M.J.; Verhagen,J.H.; Vuong,O.; Scheuer,R.D.; Pas,S.D.; Fouchier,R.A.M. |
| EPI891074 | NP | Netherlands | 2015-Jun-30 | EPI_ISL_243545 | A/black-headed gull/Netherlands/4/2015 | Import from public-domain | Poen,M.J.; Verhagen,J.H.; Vuong,O.; Scheuer,R.D.; Pas,S.D.; Fouchier,R.A.M. |
| EPI889754 | NS | Netherlands | 2014-Jun-23 | EPI_ISL_243511 | A/black-headed gull/Netherlands/26/2014 | Import from public-domain | Poen,M.J.; Verhagen,J.H.; Vuong,O.; Scheuer,R.D.; Pas,S.D.; Fouchier,R.A.M. |
| EPI889755 | MP | Netherlands | 2014-Jun-23 | EPI_ISL_243511 | A/black-headed gull/Netherlands/26/2014 | Import from public-domain | Poen,M.J.; Verhagen,J.H.; Vuong,O.; Scheuer,R.D.; Pas,S.D.; Fouchier,R.A.M. |
| EPI891114 | NP | Netherlands | 2014-Jun-23 | EPI_ISL_243511 | A/black-headed gull/Netherlands/26/2014 | Import from public-domain | Poen,M.J.; Verhagen,J.H.; Vuong,O.; Scheuer,R.D.; Pas,S.D.; Fouchier,R.A.M. |
| EPI889686 | NS | Netherlands | 2013-Jul-19 | EPI_ISL_243493 | A/black-headed gull/Netherlands/10/2013 | Import from public-domain | Poen,M.J.; Verhagen,J.H.; Vuong,O.; Scheuer,R.D.; Pas,S.D.; Fouchier,R.A.M. |
| EPI890795 | MP | Netherlands | 2013-Jul-19 | EPI_ISL_243493 | A/black-headed gull/Netherlands/10/2013 | Import from public-domain | Poen,M.J.; Verhagen,J.H.; Vuong,O.; Scheuer,R.D.; Pas,S.D.; Fouchier,R.A.M. |
| EPI890171 | NP | Netherlands | 2013-Jul-19 | EPI_ISL_243493 | A/black-headed gull/Netherlands/10/2013 | Import from public-domain | Poen,M.J.; Verhagen,J.H.; Vuong,O.; Scheuer,R.D.; Pas,S.D.; Fouchier,R.A.M. |
| EPI890081 | NP | Netherlands | 2015-Jun-30 | EPI_ISL_243463 | A/black-headed gull/Netherlands/5/2015 | Import from public-domain | Poen,M.J.; Verhagen,J.H.; Vuong,O.; Scheuer,R.D.; Pas,S.D.; Fouchier,R.A.M. |
| EPI889619 | NS | Netherlands | 2015-Jun-30 | EPI_ISL_243463 | A/black-headed gull/Netherlands/5/2015 | Import from public-domain | Poen,M.J.; Verhagen,J.H.; Vuong,O.; Scheuer,R.D.; Pas,S.D.; Fouchier,R.A.M. |
| EPI890690 | MP | Netherlands | 2015-Jun-30 | EPI_ISL_243463 | A/black-headed gull/Netherlands/5/2015 | Import from public-domain | Poen,M.J.; Verhagen,J.H.; Vuong,O.; Scheuer,R.D.; Pas,S.D.; Fouchier,R.A.M. |
| EPI889813 | NP | Netherlands | 2014-Jul-09 | EPI_ISL_243458 | A/black-headed gull/Netherlands/12/2014 | Import from public-domain | Poen,M.J.; Verhagen,J.H.; Vuong,O.; Scheuer,R.D.; Pas,S.D.; Fouchier,R.A.M. |
| EPI889830 | MP | Netherlands | 2014-Jul-09 | EPI_ISL_243458 | A/black-headed gull/Netherlands/12/2014 | Import from public-domain | Poen,M.J.; Verhagen,J.H.; Vuong,O.; Scheuer,R.D.; Pas,S.D.; Fouchier,R.A.M. |
| EPI889334 | NS | Netherlands | 2014-Jul-09 | EPI_ISL_243458 | A/black-headed gull/Netherlands/12/2014 | Import from public-domain | Poen,M.J.; Verhagen,J.H.; Vuong,O.; Scheuer,R.D.; Pas,S.D.; Fouchier,R.A.M. |
| EPI890712 | NS | Netherlands | 2013-Jul-06 | EPI_ISL_243447 | A/black-headed gull/Netherlands/7/2013 | Import from public-domain | Poen,M.J.; Verhagen,J.H.; Vuong,O.; Scheuer,R.D.; Pas,S.D.; Fouchier,R.A.M. |

| | | | | | | | | |
|---|---|---|---|---|---|---|---|---|
| EPI890928 | NP | Netherlands | 2013-Jul-06 | EPI_ISL_243447 | A/black-headed gull/Netherlands/7/2013 | | Import from public-domain | Poen,M.J.; Verhagen,J.H.; Vuong,O.; Scheuer,R.D.; Pas,S.D.; Fouchier,R.A.M. |
| EPI889865 | MP | Netherlands | 2013-Jul-06 | EPI_ISL_243447 | A/black-headed gull/Netherlands/7/2013 | | Import from public-domain | Poen,M.J.; Verhagen,J.H.; Vuong,O.; Scheuer,R.D.; Pas,S.D.; Fouchier,R.A.M. |
| EPI889393 | NP | Netherlands | 2014-Jun-23 | EPI_ISL_243443 | A/black-headed gull/Netherlands/11/2014 | | Import from public-domain | Poen,M.J.; Verhagen,J.H.; Vuong,O.; Scheuer,R.D.; Pas,S.D.; Fouchier,R.A.M. |
| EPI891025 | NS | Netherlands | 2014-Jun-23 | EPI_ISL_243443 | A/black-headed gull/Netherlands/11/2014 | | Import from public-domain | Poen,M.J.; Verhagen,J.H.; Vuong,O.; Scheuer,R.D.; Pas,S.D.; Fouchier,R.A.M. |
| EPI890028 | MP | Netherlands | 2014-Jun-23 | EPI_ISL_243443 | A/black-headed gull/Netherlands/11/2014 | | Import from public-domain | Poen,M.J.; Verhagen,J.H.; Vuong,O.; Scheuer,R.D.; Pas,S.D.; Fouchier,R.A.M. |
| EPI890616 | NP | Netherlands | 2014-Jul-11 | EPI_ISL_243437 | A/black-headed gull/Netherlands/32/2014 | | Import from public-domain | Poen,M.J.; Verhagen,J.H.; Vuong,O.; Scheuer,R.D.; Pas,S.D.; Fouchier,R.A.M. |
| EPI890332 | MP | Netherlands | 2014-Jul-11 | EPI_ISL_243437 | A/black-headed gull/Netherlands/32/2014 | | Import from public-domain | Poen,M.J.; Verhagen,J.H.; Vuong,O.; Scheuer,R.D.; Pas,S.D.; Fouchier,R.A.M. |
| EPI891137 | NS | Netherlands | 2014-Jul-11 | EPI_ISL_243437 | A/black-headed gull/Netherlands/32/2014 | | Import from public-domain | Poen,M.J.; Verhagen,J.H.; Vuong,O.; Scheuer,R.D.; Pas,S.D.; Fouchier,R.A.M. |
| EPI889953 | NS | Netherlands | 2014-Jun-23 | EPI_ISL_243419 | A/black-headed gull/Netherlands/3/2014 | | Import from public-domain | Poen,M.J.; Verhagen,J.H.; Vuong,O.; Scheuer,R.D.; Pas,S.D.; Fouchier,R.A.M. |
| EPI890961 | MP | Netherlands | 2014-Jun-23 | EPI_ISL_243419 | A/black-headed gull/Netherlands/3/2014 | | Import from public-domain | Poen,M.J.; Verhagen,J.H.; Vuong,O.; Scheuer,R.D.; Pas,S.D.; Fouchier,R.A.M. |
| EPI889980 | NP | Netherlands | 2014-Jun-23 | EPI_ISL_243419 | A/black-headed gull/Netherlands/3/2014 | | Import from public-domain | Poen,M.J.; Verhagen,J.H.; Vuong,O.; Scheuer,R.D.; Pas,S.D.; Fouchier,R.A.M. |
| EPI1537037 | MP | Netherlands | 2017-Jul-03 | EPI_ISL_376203 | A/black-headed Gull/Netherlands/5/2017 | | Import from public-domain | Poen,M.J.; Vuong,O.; Scheuer,R.D.; Pas,S.D.; Fouchier,R.A.M. |
| EPI1537008 | NP | Netherlands | 2017-Jul-03 | EPI_ISL_376203 | A/black-headed Gull/Netherlands/5/2017 | | Import from public-domain | Poen,M.J.; Vuong,O.; Scheuer,R.D.; Pas,S.D.; Fouchier,R.A.M. |
| EPI1537069 | NS | Netherlands | 2017-Jul-03 | EPI_ISL_376203 | A/black-headed Gull/Netherlands/5/2017 | | Import from public-domain | Poen,M.J.; Vuong,O.; Scheuer,R.D.; Pas,S.D.; Fouchier,R.A.M. |
| EPI1013920 | NS | Netherlands | 2014-Jul-11 | EPI_ISL_267361 | A/black-headed gull/Netherlands/30/2014 | | Import from public-domain | |
| EPI1013748 | NP | Netherlands | 2014-Jul-11 | EPI_ISL_267361 | A/black-headed gull/Netherlands/30/2014 | | Import from public-domain | |
| EPI1013699 | MP | Netherlands | 2014-Jul-11 | EPI_ISL_267361 | A/black-headed gull/Netherlands/30/2014 | | Import from public-domain | |
| EPI1525321 | NP | United States | 2014-May-21 | EPI_ISL_372111 | A/ruddy turnstone/Delaware Bay/486/2014 | | Import from public-domain | Hicks,J.T.; Friedman,K.; Qiu,X.; Kim,D.-K.; Hixson,J.; Krauss,S.; Bahl,J. |
| EPI1525323 | NS | United States | 2014-May-21 | EPI_ISL_372111 | A/ruddy turnstone/Delaware Bay/486/2014 | | Import from public-domain | Hicks,J.T.; Friedman,K.; Qiu,X.; Kim,D.-K.; Hixson,J.; Krauss,S.; Bahl,J. |
| EPI1525325 | MP | United States | 2014-May-21 | EPI_ISL_372111 | A/ruddy turnstone/Delaware Bay/486/2014 | | Import from public-domain | Hicks,J.T.; Friedman,K.; Qiu,X.; Kim,D.-K.; Hixson,J.; Krauss,S.; Bahl,J. |
| EPI1525173 | NS | United States | 2014-May-19 | EPI_ISL_372090 | A/ruddy turnstone/Delaware Bay/48/2014 | | Import from public-domain | Hicks,J.T.; Friedman,K.; Qiu,X.; Kim,D.-K.; Hixson,J.; Krauss,S.; Bahl,J. |
| EPI1525178 | MP | United States | 2014-May-19 | EPI_ISL_372090 | A/ruddy turnstone/Delaware Bay/48/2014 | | Import from public-domain | Hicks,J.T.; Friedman,K.; Qiu,X.; Kim,D.-K.; Hixson,J.; Krauss,S.; Bahl,J. |
| EPI1525180 | NP | United States | 2014-May-19 | EPI_ISL_372090 | A/ruddy turnstone/Delaware Bay/48/2014 | | Import from public-domain | Hicks,J.T.; Friedman,K.; Qiu,X.; Kim,D.-K.; Hixson,J.; Krauss,S.; Bahl,J. |
| EPI1622928 | NS | United States | 2014-May-21 | EPI_ISL_397706 | A/ruddy turnstone/New Jersey/UGAI14-1436/2014 | | Import from public-domain | Tan,G.; Pickett,B.; Fedorova,N.; Amedeo,P.; Hu,L.; Christensen,J.; Miller,J.; Durbin,A.; Williams,T.; Poulson,R.; Stallknecht,D. |
| EPI1622925 | MP | United States | 2014-May-21 | EPI_ISL_397706 | A/ruddy turnstone/New Jersey/UGAI14-1436/2014 | | Import from public-domain | Tan,G.; Pickett,B.; Fedorova,N.; Amedeo,P.; Hu,L.; Christensen,J.; Miller,J.; Durbin,A.; Williams,T.; Poulson,R.; Stallknecht,D. |
| EPI1622927 | NP | United States | 2014-May-21 | EPI_ISL_397706 | A/ruddy turnstone/New Jersey/UGAI14-1436/2014 | | Import from public-domain | Tan,G.; Pickett,B.; Fedorova,N.; Amedeo,P.; Hu,L.; Christensen,J.; Miller,J.; Durbin,A.; Williams,T.; Poulson,R.; Stallknecht,D. |
| EPI1581795 | NS | United States | 2018-Sep-26 | EPI_ISL_388040 | A/Glaucous Gull/Northern Alaska/18AR00289/2018 | | Import from public-domain | Tan,G.; Fedorova,N.; Amedeo,P.; Christensen,J.; Miller,J.; Durbin,A.; Williams,T.; Cadiz,C.; Bishop,M.; Hill,N.; Runstadler,J. |
| EPI1581792 | MP | United States | 2018-Sep-26 | EPI_ISL_388040 | A/Glaucous Gull/Northern Alaska/18AR00289/2018 | | Import from public-domain | Tan,G.; Fedorova,N.; Amedeo,P.; Christensen,J.; Miller,J.; Durbin,A.; Williams,T.; Cadiz,C.; Bishop,M.; Hill,N.; Runstadler,J. |
| EPI1581794 | NP | United States | 2018-Sep-26 | EPI_ISL_388040 | A/Glaucous Gull/Northern Alaska/18AR00289/2018 | | Import from public-domain | Tan,G.; Fedorova,N.; Amedeo,P.; Christensen,J.; Miller,J.; Durbin,A.; Williams,T.; Cadiz,C.; Bishop,M.; Hill,N.; Runstadler,J. |
| EPI1621155 | NS | Chile | 2016-Dec-13 | EPI_ISL_397478 | A/white-backed stilt/Chile/C20090/2016 | | Import from public-domain | Bravo-Vasquez,N.; Bahl,J.; Hamilton-West,C.; Schultz-Cherry,S. |
| EPI1621040 | MP | Chile | 2016-Dec-13 | EPI_ISL_397478 | A/white-backed stilt/Chile/C20090/2016 | | Import from public-domain | Bravo-Vasquez,N.; Bahl,J.; Hamilton-West,C.; Schultz-Cherry,S. |
| EPI1621220 | NP | Chile | 2016-Dec-13 | EPI_ISL_397478 | A/white-backed stilt/Chile/C20090/2016 | | Import from public-domain | Bravo-Vasquez,N.; Bahl,J.; Hamilton-West,C.; Schultz-Cherry,S. |
| EPI1956633 | NP | Netherlands | 2022-Jan-03 | EPI_ISL_8650947 | A/red fox/Netherlands/22000097-004/2022 | Wageningen Bioveterinary Research | Wageningen Bioveterinary Research | Beerens, Nancy; Harders, Frank; Pritz-Verschuren, Sylvia; Roose, Marit; Venema, Sandra; Germeraad, Evelien; Engelsma, Marc; Heutink, Rene |
| EPI1956634 | NS | Netherlands | 2022-Jan-03 | EPI_ISL_8650947 | A/red fox/Netherlands/22000097-004/2022 | Wageningen Bioveterinary Research | Wageningen Bioveterinary Research | Beerens, Nancy; Harders, Frank; Pritz-Verschuren, Sylvia; Roose, Marit; Venema, Sandra; Germeraad, Evelien; Engelsma, Marc; Heutink, Rene |
| EPI1956635 | MP | Netherlands | 2022-Jan-03 | EPI_ISL_8650947 | A/red fox/Netherlands/22000097-004/2022 | Wageningen Bioveterinary Research | Wageningen Bioveterinary Research | Beerens, Nancy; Harders, Frank; Pritz-Verschuren, Sylvia; Roose, Marit; Venema, Sandra; Germeraad, Evelien; Engelsma, Marc; Heutink, Rene |

| | | | | | | | | |
|---|---|---|---|---|---|---|---|---|
| EPI1958451 | NS | Netherlands | 2022-Jan-08 | EPI_ISL_8799320 | A/polecat/Netherlands/22000516-003/2022 | Wageningen Bioveterinary Research | Wageningen Bioveterinary Research | Beerens, Nancy; Harders, Frank; Pritz-Verschuren, Sylvia; Roose, Marit; Venema, Sandra; Germeraad, Evelien; Engelsma, Marc; Heutink, Rene |
| EPI1958452 | MP | Netherlands | 2022-Jan-08 | EPI_ISL_8799320 | A/polecat/Netherlands/22000516-003/2022 | Wageningen Bioveterinary Research | Wageningen Bioveterinary Research | Beerens, Nancy; Harders, Frank; Pritz-Verschuren, Sylvia; Roose, Marit; Venema, Sandra; Germeraad, Evelien; Engelsma, Marc; Heutink, Rene |
| EPI1958450 | NP | Netherlands | 2022-Jan-08 | EPI_ISL_8799320 | A/polecat/Netherlands/22000516-003/2022 | Wageningen Bioveterinary Research | Wageningen Bioveterinary Research | Beerens, Nancy; Harders, Frank; Pritz-Verschuren, Sylvia; Roose, Marit; Venema, Sandra; Germeraad, Evelien; Engelsma, Marc; Heutink, Rene |
| EPI1965206 | NP | Netherlands | 2022-Jan-06 | EPI_ISL_9111095 | A/goose/Netherlands/22000487-001/2022 | Wageningen Bioveterinary Research | Wageningen Bioveterinary Research | Beerens, Nancy; Harders, Frank; Pritz-Verschuren, Sylvia; Roose, Marit; Venema, Sandra; Germeraad, Evelien; Engelsma, Marc; Heutink, Rene |
| EPI1965207 | NS | Netherlands | 2022-Jan-06 | EPI_ISL_9111095 | A/goose/Netherlands/22000487-001/2022 | Wageningen Bioveterinary Research | Wageningen Bioveterinary Research | Beerens, Nancy; Harders, Frank; Pritz-Verschuren, Sylvia; Roose, Marit; Venema, Sandra; Germeraad, Evelien; Engelsma, Marc; Heutink, Rene |
| EPI1965208 | MP | Netherlands | 2022-Jan-06 | EPI_ISL_9111095 | A/goose/Netherlands/22000487-001/2022 | Wageningen Bioveterinary Research | Wageningen Bioveterinary Research | Beerens, Nancy; Harders, Frank; Pritz-Verschuren, Sylvia; Roose, Marit; Venema, Sandra; Germeraad, Evelien; Engelsma, Marc; Heutink, Rene |
| EPI1965222 | NP | Netherlands | 2022-Jan-17 | EPI_ISL_9111104 | A/otter/Netherlands/22001014-005/2022 | Wageningen Bioveterinary Research | Wageningen Bioveterinary Research | Beerens, Nancy; Harders, Frank; Pritz-Verschuren, Sylvia; Roose, Marit; Venema, Sandra; Germeraad, Evelien; Engelsma, Marc; Heutink, Rene |
| EPI1965223 | NS | Netherlands | 2022-Jan-17 | EPI_ISL_9111104 | A/otter/Netherlands/22001014-005/2022 | Wageningen Bioveterinary Research | Wageningen Bioveterinary Research | Beerens, Nancy; Harders, Frank; Pritz-Verschuren, Sylvia; Roose, Marit; Venema, Sandra; Germeraad, Evelien; Engelsma, Marc; Heutink, Rene |
| EPI1965224 | MP | Netherlands | 2022-Jan-17 | EPI_ISL_9111104 | A/otter/Netherlands/22001014-005/2022 | Wageningen Bioveterinary Research | Wageningen Bioveterinary Research | Beerens, Nancy; Harders, Frank; Pritz-Verschuren, Sylvia; Roose, Marit; Venema, Sandra; Germeraad, Evelien; Engelsma, Marc; Heutink, Rene |
| EPI1965230 | NP | Netherlands | 2022-Jan-22 | EPI_ISL_9111105 | A/chicken/Netherlands/22001401-001005/2022 | Wageningen Bioveterinary Research | Wageningen Bioveterinary Research | Beerens, Nancy; Harders, Frank; Pritz-Verschuren, Sylvia; Roose, Marit; Venema, Sandra; Germeraad, Evelien; Engelsma, Marc; Heutink, Rene |
| EPI1965231 | NS | Netherlands | 2022-Jan-22 | EPI_ISL_9111105 | A/chicken/Netherlands/22001401-001005/2022 | Wageningen Bioveterinary Research | Wageningen Bioveterinary Research | Beerens, Nancy; Harders, Frank; Pritz-Verschuren, Sylvia; Roose, Marit; Venema, Sandra; Germeraad, Evelien; Engelsma, Marc; Heutink, Rene |
| EPI1965232 | MP | Netherlands | 2022-Jan-22 | EPI_ISL_9111105 | A/chicken/Netherlands/22001401-001005/2022 | Wageningen Bioveterinary Research | Wageningen Bioveterinary Research | Beerens, Nancy; Harders, Frank; Pritz-Verschuren, Sylvia; Roose, Marit; Venema, Sandra; Germeraad, Evelien; Engelsma, Marc; Heutink, Rene |
| EPI1965254 | NP | Netherlands | 2022-Jan-04 | EPI_ISL_9117228 | A/northern goshawk/Netherlands/22000305-002/2022 | Wageningen Bioveterinary Research | Wageningen Bioveterinary Research | Beerens, Nancy; Harders, Frank; Pritz-Verschuren, Sylvia; Roose, Marit; Venema, Sandra; Germeraad, Evelien; Engelsma, Marc; Heutink, Rene |
| EPI1965255 | NS | Netherlands | 2022-Jan-04 | EPI_ISL_9117228 | A/northern goshawk/Netherlands/22000305-002/2022 | Wageningen Bioveterinary Research | Wageningen Bioveterinary Research | Beerens, Nancy; Harders, Frank; Pritz-Verschuren, Sylvia; Roose, Marit; Venema, Sandra; Germeraad, Evelien; Engelsma, Marc; Heutink, Rene |
| EPI1965256 | MP | Netherlands | 2022-Jan-04 | EPI_ISL_9117228 | A/northern goshawk/Netherlands/22000305-002/2022 | Wageningen Bioveterinary Research | Wageningen Bioveterinary Research | Beerens, Nancy; Harders, Frank; Pritz-Verschuren, Sylvia; Roose, Marit; Venema, Sandra; Germeraad, Evelien; Engelsma, Marc; Heutink, Rene |
| EPI1966010 | NP | Belgium | 2022-Jan-04 | EPI_ISL_9162331 | A/Phasianus_colchicus/Belgium/294/2022 | Sciensano - Animal Infectious Diseases | Sciensano, Department of Animal Infectious Diseases | Van Borm, Steven; Roupie, Virginie; Lambrecht, Benedicte; Mathijs, Elisabeth; Steensels, Mieke |
| EPI1966014 | MP | Belgium | 2022-Jan-04 | EPI_ISL_9162331 | A/Phasianus_colchicus/Belgium/294/2022 | Sciensano - Animal Infectious Diseases | Sciensano, Department of Animal Infectious Diseases | Van Borm, Steven; Roupie, Virginie; Lambrecht, Benedicte; Mathijs, Elisabeth; Steensels, Mieke |
| EPI1966015 | NS | Belgium | 2022-Jan-04 | EPI_ISL_9162331 | A/Phasianus_colchicus/Belgium/294/2022 | Sciensano - Animal Infectious Diseases | Sciensano, Department of Animal Infectious Diseases | Van Borm, Steven; Roupie, Virginie; Lambrecht, Benedicte; Mathijs, Elisabeth; Steensels, Mieke |
| EPI1966254 | NP | Netherlands | 2022-Jan-06 | EPI_ISL_9221519 | A/barnacle goose/Netherlands/22000419-002/2022 | Wageningen Bioveterinary Research | Wageningen Bioveterinary Research | Beerens, Nancy; Harders, Frank; Pritz-Verschuren, Sylvia; Roose, Marit; Venema, Sandra; Germeraad, Evelien; Engelsma, Marc; Heutink, Rene |
| EPI1966255 | NS | Netherlands | 2022-Jan-06 | EPI_ISL_9221519 | A/barnacle goose/Netherlands/22000419-002/2022 | Wageningen Bioveterinary Research | Wageningen Bioveterinary Research | Beerens, Nancy; Harders, Frank; Pritz-Verschuren, Sylvia; Roose, Marit; Venema, Sandra; Germeraad, Evelien; Engelsma, Marc; Heutink, Rene |
| EPI1966256 | MP | Netherlands | 2022-Jan-06 | EPI_ISL_9221519 | A/barnacle goose/Netherlands/22000419-002/2022 | Wageningen Bioveterinary Research | Wageningen Bioveterinary Research | Beerens, Nancy; Harders, Frank; Pritz-Verschuren, Sylvia; Roose, Marit; Venema, Sandra; Germeraad, Evelien; Engelsma, Marc; Heutink, Rene |
| EPI1966947 | NP | Netherlands | 2022-Jan-04 | EPI_ISL_9261741 | A/Barnacle goose/Netherlands/1/2022 | Erasmus Medical Center | Erasmus Medical Center | |
| EPI1966948 | NS | Netherlands | 2022-Jan-04 | EPI_ISL_9261741 | A/Barnacle goose/Netherlands/1/2022 | Erasmus Medical Center | Erasmus Medical Center | |
| EPI1966949 | MP | Netherlands | 2022-Jan-04 | EPI_ISL_9261741 | A/Barnacle goose/Netherlands/1/2022 | Erasmus Medical Center | Erasmus Medical Center | |
| EPI1966955 | NP | Netherlands | 2022-Jan-06 | EPI_ISL_9261742 | A/Great black-backed gull/1/2022 | Erasmus Medical Center | Erasmus Medical Center | |
| EPI1966956 | NS | Netherlands | 2022-Jan-06 | EPI_ISL_9261742 | A/Great black-backed gull/1/2022 | Erasmus Medical Center | Erasmus Medical Center | |
| EPI1966957 | MP | Netherlands | 2022-Jan-06 | EPI_ISL_9261742 | A/Great black-backed gull/1/2022 | Erasmus Medical Center | Erasmus Medical Center | |
| EPI1966963 | NP | Netherlands | 2022-Jan-11 | EPI_ISL_9261743 | A/Oystercatcher/Netherlands/1/2022 | Erasmus Medical Center | Erasmus Medical Center | |
| EPI1966964 | NS | Netherlands | 2022-Jan-11 | EPI_ISL_9261743 | A/Oystercatcher/Netherlands/1/2022 | Erasmus Medical Center | Erasmus Medical Center | |
| EPI1966965 | MP | Netherlands | 2022-Jan-11 | EPI_ISL_9261743 | A/Oystercatcher/Netherlands/1/2022 | Erasmus Medical Center | Erasmus Medical Center | |
| EPI1966971 | NP | Netherlands | 2022-Jan-11 | EPI_ISL_9261744 | A/Barnacle goose/Netherlands/2/2022 | Erasmus Medical Center | Erasmus Medical Center | |
| EPI1966972 | NS | Netherlands | 2022-Jan-11 | EPI_ISL_9261744 | A/Barnacle goose/Netherlands/2/2022 | Erasmus Medical Center | Erasmus Medical Center | |
| EPI1966973 | MP | Netherlands | 2022-Jan-11 | EPI_ISL_9261744 | A/Barnacle goose/Netherlands/2/2022 | Erasmus Medical Center | Erasmus Medical Center | |
| EPI1966979 | NP | Netherlands | 2022-Jan-11 | EPI_ISL_9261745 | A/Barnacle goose/Netherlands/3/2022 | Erasmus Medical Center | Erasmus Medical Center | |
| EPI1966980 | NS | Netherlands | 2022-Jan-11 | EPI_ISL_9261745 | A/Barnacle goose/Netherlands/3/2022 | Erasmus Medical Center | Erasmus Medical Center | |

| | | | | | | | | |
|---|---|---|---|---|---|---|---|---|
| EPI1966981 | MP | Netherlands | 2022-Jan-11 | EPI_ISL_9261745 | A/Barnacle goose/Netherlands/3/2022 | Erasmus Medical Center | Erasmus Medical Center | |
| EPI1966987 | NP | Netherlands | 2022-Jan-11 | EPI_ISL_9261746 | A/Barnacle goose/Netherlands/4/2022 | Erasmus Medical Center | Erasmus Medical Center | |
| EPI1966988 | NS | Netherlands | 2022-Jan-11 | EPI_ISL_9261746 | A/Barnacle goose/Netherlands/4/2022 | Erasmus Medical Center | Erasmus Medical Center | |
| EPI1966989 | MP | Netherlands | 2022-Jan-11 | EPI_ISL_9261746 | A/Barnacle goose/Netherlands/4/2022 | Erasmus Medical Center | Erasmus Medical Center | |
| EPI1966994 | NP | Netherlands | 2022-Jan-14 | EPI_ISL_9261747 | A/Barnacle goose/Netherlands/5/2022 | Erasmus Medical Center | Erasmus Medical Center | |
| EPI1966995 | NS | Netherlands | 2022-Jan-14 | EPI_ISL_9261747 | A/Barnacle goose/Netherlands/5/2022 | Erasmus Medical Center | Erasmus Medical Center | |
| EPI1966996 | MP | Netherlands | 2022-Jan-14 | EPI_ISL_9261747 | A/Barnacle goose/Netherlands/5/2022 | Erasmus Medical Center | Erasmus Medical Center | |
| EPI1967002 | NP | Netherlands | 2022-Jan-14 | EPI_ISL_9261748 | A/Grey heron/Netherlands/1/2022 | Erasmus Medical Center | Erasmus Medical Center | |
| EPI1967003 | NS | Netherlands | 2022-Jan-14 | EPI_ISL_9261748 | A/Grey heron/Netherlands/1/2022 | Erasmus Medical Center | Erasmus Medical Center | |
| EPI1967004 | MP | Netherlands | 2022-Jan-14 | EPI_ISL_9261748 | A/Grey heron/Netherlands/1/2022 | Erasmus Medical Center | Erasmus Medical Center | |
| EPI1967010 | NP | Netherlands | 2022-Jan-14 | EPI_ISL_9261749 | A/Caspian gull/Netherlands/1/2022 | Erasmus Medical Center | Erasmus Medical Center | |
| EPI1967011 | NS | Netherlands | 2022-Jan-14 | EPI_ISL_9261749 | A/Caspian gull/Netherlands/1/2022 | Erasmus Medical Center | Erasmus Medical Center | |
| EPI1967012 | MP | Netherlands | 2022-Jan-14 | EPI_ISL_9261749 | A/Caspian gull/Netherlands/1/2022 | Erasmus Medical Center | Erasmus Medical Center | |
| EPI1967018 | NP | Netherlands | 2022-Jan-14 | EPI_ISL_9261750 | A/Black-headed gull/Netherlands/1/2022 | Erasmus Medical Center | Erasmus Medical Center | |
| EPI1967019 | NS | Netherlands | 2022-Jan-14 | EPI_ISL_9261750 | A/Black-headed gull/Netherlands/1/2022 | Erasmus Medical Center | Erasmus Medical Center | |
| EPI1967020 | MP | Netherlands | 2022-Jan-14 | EPI_ISL_9261750 | A/Black-headed gull/Netherlands/1/2022 | Erasmus Medical Center | Erasmus Medical Center | |
| EPI1967026 | NP | Netherlands | 2022-Jan-14 | EPI_ISL_9261751 | A/Barnacle goose/Netherlands/6/2022 | Erasmus Medical Center | Erasmus Medical Center | |
| EPI1967027 | NS | Netherlands | 2022-Jan-14 | EPI_ISL_9261751 | A/Barnacle goose/Netherlands/6/2022 | Erasmus Medical Center | Erasmus Medical Center | |
| EPI1967028 | MP | Netherlands | 2022-Jan-14 | EPI_ISL_9261751 | A/Barnacle goose/Netherlands/6/2022 | Erasmus Medical Center | Erasmus Medical Center | |
| EPI1980925 | NP | Netherlands | 2022-Jan-04 | EPI_ISL_9616210 | A/gadwall/Netherlands/22000307-001/2022 | Wageningen Bioveterinary Research | Wageningen Bioveterinary Research | Beerens, Nancy; Harders, Frank; Pritz-Verschuren, Sylvia; Roose, Marit; Venema, Sandra; Germeraad, Evelien; Engelsma, Marc; Heutink, Rene |
| EPI1980926 | NS | Netherlands | 2022-Jan-04 | EPI_ISL_9616210 | A/gadwall/Netherlands/22000307-001/2022 | Wageningen Bioveterinary Research | Wageningen Bioveterinary Research | Beerens, Nancy; Harders, Frank; Pritz-Verschuren, Sylvia; Roose, Marit; Venema, Sandra; Germeraad, Evelien; Engelsma, Marc; Heutink, Rene |
| EPI1980927 | MP | Netherlands | 2022-Jan-04 | EPI_ISL_9616210 | A/gadwall/Netherlands/22000307-001/2022 | Wageningen Bioveterinary Research | Wageningen Bioveterinary Research | Beerens, Nancy; Harders, Frank; Pritz-Verschuren, Sylvia; Roose, Marit; Venema, Sandra; Germeraad, Evelien; Engelsma, Marc; Heutink, Rene |
| EPI1980933 | NP | Netherlands | 2022-Jan-06 | EPI_ISL_9616212 | A/red knot/Netherlands/22000409-002/2022 | Wageningen Bioveterinary Research | Wageningen Bioveterinary Research | Beerens, Nancy; Harders, Frank; Pritz-Verschuren, Sylvia; Roose, Marit; Venema, Sandra; Germeraad, Evelien; Engelsma, Marc; Heutink, Rene |
| EPI1980934 | NS | Netherlands | 2022-Jan-06 | EPI_ISL_9616212 | A/red knot/Netherlands/22000409-002/2022 | Wageningen Bioveterinary Research | Wageningen Bioveterinary Research | Beerens, Nancy; Harders, Frank; Pritz-Verschuren, Sylvia; Roose, Marit; Venema, Sandra; Germeraad, Evelien; Engelsma, Marc; Heutink, Rene |
| EPI1980935 | MP | Netherlands | 2022-Jan-06 | EPI_ISL_9616212 | A/red knot/Netherlands/22000409-002/2022 | Wageningen Bioveterinary Research | Wageningen Bioveterinary Research | Beerens, Nancy; Harders, Frank; Pritz-Verschuren, Sylvia; Roose, Marit; Venema, Sandra; Germeraad, Evelien; Engelsma, Marc; Heutink, Rene |
| EPI1980973 | NP | Netherlands | 2022-Jan-11 | EPI_ISL_9616263 | A/brant goose/Netherlands/22000648-002/2022 | Wageningen Bioveterinary Research | Wageningen Bioveterinary Research | Beerens, Nancy; Harders, Frank; Pritz-Verschuren, Sylvia; Roose, Marit; Venema, Sandra; Germeraad, Evelien; Engelsma, Marc; Heutink, Rene |
| EPI1980974 | NS | Netherlands | 2022-Jan-11 | EPI_ISL_9616263 | A/brant goose/Netherlands/22000648-002/2022 | Wageningen Bioveterinary Research | Wageningen Bioveterinary Research | Beerens, Nancy; Harders, Frank; Pritz-Verschuren, Sylvia; Roose, Marit; Venema, Sandra; Germeraad, Evelien; Engelsma, Marc; Heutink, Rene |
| EPI1980975 | MP | Netherlands | 2022-Jan-11 | EPI_ISL_9616263 | A/brant goose/Netherlands/22000648-002/2022 | Wageningen Bioveterinary Research | Wageningen Bioveterinary Research | Beerens, Nancy; Harders, Frank; Pritz-Verschuren, Sylvia; Roose, Marit; Venema, Sandra; Germeraad, Evelien; Engelsma, Marc; Heutink, Rene |
| EPI1980989 | NP | Netherlands | 2022-Jan-27 | EPI_ISL_9616265 | A/red fox/Netherlands/22001901-005/2022 | Wageningen Bioveterinary Research | Wageningen Bioveterinary Research | Beerens, Nancy; Harders, Frank; Pritz-Verschuren, Sylvia; Roose, Marit; Venema, Sandra; Germeraad, Evelien; Engelsma, Marc; Heutink, Rene |
| EPI1980990 | NS | Netherlands | 2022-Jan-27 | EPI_ISL_9616265 | A/red fox/Netherlands/22001901-005/2022 | Wageningen Bioveterinary Research | Wageningen Bioveterinary Research | Beerens, Nancy; Harders, Frank; Pritz-Verschuren, Sylvia; Roose, Marit; Venema, Sandra; Germeraad, Evelien; Engelsma, Marc; Heutink, Rene |
| EPI1980991 | MP | Netherlands | 2022-Jan-27 | EPI_ISL_9616265 | A/red fox/Netherlands/22001901-005/2022 | Wageningen Bioveterinary Research | Wageningen Bioveterinary Research | Beerens, Nancy; Harders, Frank; Pritz-Verschuren, Sylvia; Roose, Marit; Venema, Sandra; Germeraad, Evelien; Engelsma, Marc; Heutink, Rene |
| EPI1980997 | NP | Netherlands | 2022-Jan-24 | EPI_ISL_9616266 | A/polecat/Netherlands/22002047-001/2022 | Wageningen Bioveterinary Research | Wageningen Bioveterinary Research | Beerens, Nancy; Harders, Frank; Pritz-Verschuren, Sylvia; Roose, Marit; Venema, Sandra; Germeraad, Evelien; Engelsma, Marc; Heutink, Rene |
| EPI1980998 | NS | Netherlands | 2022-Jan-24 | EPI_ISL_9616266 | A/polecat/Netherlands/22002047-001/2022 | Wageningen Bioveterinary Research | Wageningen Bioveterinary Research | Beerens, Nancy; Harders, Frank; Pritz-Verschuren, Sylvia; Roose, Marit; Venema, Sandra; Germeraad, Evelien; Engelsma, Marc; Heutink, Rene |
| EPI1980999 | MP | Netherlands | 2022-Jan-24 | EPI_ISL_9616266 | A/polecat/Netherlands/22002047-001/2022 | Wageningen Bioveterinary Research | Wageningen Bioveterinary Research | Beerens, Nancy; Harders, Frank; Pritz-Verschuren, Sylvia; Roose, Marit; Venema, Sandra; Germeraad, Evelien; Engelsma, Marc; Heutink, Rene |
| EPI1981005 | NP | Netherlands | 2022-Jan-31 | EPI_ISL_9616271 | A/red fox/Netherlands/22002190-003/2022 | Wageningen Bioveterinary Research | Wageningen Bioveterinary Research | Beerens, Nancy; Harders, Frank; Pritz-Verschuren, Sylvia; Roose, Marit; Venema, Sandra; Germeraad, Evelien; Engelsma, Marc; Heutink, Rene |

| | | | | | | | | |
|---|---|---|---|---|---|---|---|---|
| EPI1981006 | NS | Netherlands | 2022-Jan-31 | EPI_ISL_9616271 | A/red fox/Netherlands/22002190-003/2022 | Wageningen Bioveterinary Research | Wageningen Bioveterinary Research | Beerens, Nancy; Harders, Frank; Pritz-Verschuren, Sylvia; Roose, Marit; Venema, Sandra; Germeraad, Evelien; Engelsma, Marc; Heutink, Rene |
| EPI1981007 | MP | Netherlands | 2022-Jan-31 | EPI_ISL_9616271 | A/red fox/Netherlands/22002190-003/2022 | Wageningen Bioveterinary Research | Wageningen Bioveterinary Research | Beerens, Nancy; Harders, Frank; Pritz-Verschuren, Sylvia; Roose, Marit; Venema, Sandra; Germeraad, Evelien; Engelsma, Marc; Heutink, Rene |
| EPI1981013 | NP | Netherlands | 2022-Jan-31 | EPI_ISL_9616272 | A/red fox/Netherlands/22002191-003/2022 | Wageningen Bioveterinary Research | Wageningen Bioveterinary Research | Beerens, Nancy; Harders, Frank; Pritz-Verschuren, Sylvia; Roose, Marit; Venema, Sandra; Germeraad, Evelien; Engelsma, Marc; Heutink, Rene |
| EPI1981014 | NS | Netherlands | 2022-Jan-31 | EPI_ISL_9616272 | A/red fox/Netherlands/22002191-003/2022 | Wageningen Bioveterinary Research | Wageningen Bioveterinary Research | Beerens, Nancy; Harders, Frank; Pritz-Verschuren, Sylvia; Roose, Marit; Venema, Sandra; Germeraad, Evelien; Engelsma, Marc; Heutink, Rene |
| EPI1981015 | MP | Netherlands | 2022-Jan-31 | EPI_ISL_9616272 | A/red fox/Netherlands/22002191-003/2022 | Wageningen Bioveterinary Research | Wageningen Bioveterinary Research | Beerens, Nancy; Harders, Frank; Pritz-Verschuren, Sylvia; Roose, Marit; Venema, Sandra; Germeraad, Evelien; Engelsma, Marc; Heutink, Rene |
| EPI1983202 | NP | Netherlands | 2022-Jan-03 | EPI_ISL_9690841 | A/great black-backed gull/Netherlands/22000090-002/2022 | Wageningen Bioveterinary Research | Wageningen Bioveterinary Research | Beerens, Nancy; Harders, Frank; Pritz-Verschuren, Sylvia; Roose, Marit; Venema, Sandra; Germeraad, Evelien; Engelsma, Marc; Heutink, Rene |
| EPI1983203 | NS | Netherlands | 2022-Jan-03 | EPI_ISL_9690841 | A/great black-backed gull/Netherlands/22000090-002/2022 | Wageningen Bioveterinary Research | Wageningen Bioveterinary Research | Beerens, Nancy; Harders, Frank; Pritz-Verschuren, Sylvia; Roose, Marit; Venema, Sandra; Germeraad, Evelien; Engelsma, Marc; Heutink, Rene |
| EPI1983204 | MP | Netherlands | 2022-Jan-03 | EPI_ISL_9690841 | A/great black-backed gull/Netherlands/22000090-002/2022 | Wageningen Bioveterinary Research | Wageningen Bioveterinary Research | Beerens, Nancy; Harders, Frank; Pritz-Verschuren, Sylvia; Roose, Marit; Venema, Sandra; Germeraad, Evelien; Engelsma, Marc; Heutink, Rene |
| EPI1983210 | NP | Netherlands | 2022-Jan-03 | EPI_ISL_9690956 | A/common buzzard/Netherlands/22000192-001/2022 | Wageningen Bioveterinary Research | Wageningen Bioveterinary Research | Beerens, Nancy; Harders, Frank; Pritz-Verschuren, Sylvia; Roose, Marit; Venema, Sandra; Germeraad, Evelien; Engelsma, Marc; Heutink, Rene |
| EPI1983211 | NS | Netherlands | 2022-Jan-03 | EPI_ISL_9690956 | A/common buzzard/Netherlands/22000192-001/2022 | Wageningen Bioveterinary Research | Wageningen Bioveterinary Research | Beerens, Nancy; Harders, Frank; Pritz-Verschuren, Sylvia; Roose, Marit; Venema, Sandra; Germeraad, Evelien; Engelsma, Marc; Heutink, Rene |
| EPI1983212 | MP | Netherlands | 2022-Jan-03 | EPI_ISL_9690956 | A/common buzzard/Netherlands/22000192-001/2022 | Wageningen Bioveterinary Research | Wageningen Bioveterinary Research | Beerens, Nancy; Harders, Frank; Pritz-Verschuren, Sylvia; Roose, Marit; Venema, Sandra; Germeraad, Evelien; Engelsma, Marc; Heutink, Rene |
| EPI1987211 | NP | Netherlands | 2022-Feb-07 | EPI_ISL_10220427 | A/red fox/Netherlands/22002636-005/2022 | Wageningen Bioveterinary Research | Wageningen Bioveterinary Research | Beerens, Nancy; Harders, Frank; Pritz-Verschuren, Sylvia; Roose, Marit; Venema, Sandra; Germeraad, Evelien; Engelsma, Marc; Heutink, Rene |
| EPI1987212 | NS | Netherlands | 2022-Feb-07 | EPI_ISL_10220427 | A/red fox/Netherlands/22002636-005/2022 | Wageningen Bioveterinary Research | Wageningen Bioveterinary Research | Beerens, Nancy; Harders, Frank; Pritz-Verschuren, Sylvia; Roose, Marit; Venema, Sandra; Germeraad, Evelien; Engelsma, Marc; Heutink, Rene |
| EPI1987213 | MP | Netherlands | 2022-Feb-07 | EPI_ISL_10220427 | A/red fox/Netherlands/22002636-005/2022 | Wageningen Bioveterinary Research | Wageningen Bioveterinary Research | Beerens, Nancy; Harders, Frank; Pritz-Verschuren, Sylvia; Roose, Marit; Venema, Sandra; Germeraad, Evelien; Engelsma, Marc; Heutink, Rene |
| EPI1987219 | NP | Netherlands | 2022-Feb-08 | EPI_ISL_10220428 | A/red fox/Netherlands/22002722-004/2022 | Wageningen Bioveterinary Research | Wageningen Bioveterinary Research | Beerens, Nancy; Harders, Frank; Pritz-Verschuren, Sylvia; Roose, Marit; Venema, Sandra; Germeraad, Evelien; Engelsma, Marc; Heutink, Rene |
| EPI1987220 | NS | Netherlands | 2022-Feb-08 | EPI_ISL_10220428 | A/red fox/Netherlands/22002722-004/2022 | Wageningen Bioveterinary Research | Wageningen Bioveterinary Research | Beerens, Nancy; Harders, Frank; Pritz-Verschuren, Sylvia; Roose, Marit; Venema, Sandra; Germeraad, Evelien; Engelsma, Marc; Heutink, Rene |
| EPI1987221 | MP | Netherlands | 2022-Feb-08 | EPI_ISL_10220428 | A/red fox/Netherlands/22002722-004/2022 | Wageningen Bioveterinary Research | Wageningen Bioveterinary Research | Beerens, Nancy; Harders, Frank; Pritz-Verschuren, Sylvia; Roose, Marit; Venema, Sandra; Germeraad, Evelien; Engelsma, Marc; Heutink, Rene |
| EPI1987227 | NP | Netherlands | 2022-Feb-09 | EPI_ISL_10220429 | A/polecat/Netherlands/22002792-005/2022 | Wageningen Bioveterinary Research | Wageningen Bioveterinary Research | Beerens, Nancy; Harders, Frank; Pritz-Verschuren, Sylvia; Roose, Marit; Venema, Sandra; Germeraad, Evelien; Engelsma, Marc; Heutink, Rene |
| EPI1987228 | NS | Netherlands | 2022-Feb-09 | EPI_ISL_10220429 | A/polecat/Netherlands/22002792-005/2022 | Wageningen Bioveterinary Research | Wageningen Bioveterinary Research | Beerens, Nancy; Harders, Frank; Pritz-Verschuren, Sylvia; Roose, Marit; Venema, Sandra; Germeraad, Evelien; Engelsma, Marc; Heutink, Rene |
| EPI1987229 | MP | Netherlands | 2022-Feb-09 | EPI_ISL_10220429 | A/polecat/Netherlands/22002792-005/2022 | Wageningen Bioveterinary Research | Wageningen Bioveterinary Research | Beerens, Nancy; Harders, Frank; Pritz-Verschuren, Sylvia; Roose, Marit; Venema, Sandra; Germeraad, Evelien; Engelsma, Marc; Heutink, Rene |
| EPI1987251 | NP | Netherlands | 2022-Feb-13 | EPI_ISL_10220432 | A/badger/Netherlands/22003078-004/2022 | Wageningen Bioveterinary Research | Wageningen Bioveterinary Research | Beerens, Nancy; Harders, Frank; Pritz-Verschuren, Sylvia; Roose, Marit; Venema, Sandra; Germeraad, Evelien; Engelsma, Marc; Heutink, Rene |
| EPI1987252 | NS | Netherlands | 2022-Feb-13 | EPI_ISL_10220432 | A/badger/Netherlands/22003078-004/2022 | Wageningen Bioveterinary Research | Wageningen Bioveterinary Research | Beerens, Nancy; Harders, Frank; Pritz-Verschuren, Sylvia; Roose, Marit; Venema, Sandra; Germeraad, Evelien; Engelsma, Marc; Heutink, Rene |
| EPI1987253 | MP | Netherlands | 2022-Feb-13 | EPI_ISL_10220432 | A/badger/Netherlands/22003078-004/2022 | Wageningen Bioveterinary Research | Wageningen Bioveterinary Research | Beerens, Nancy; Harders, Frank; Pritz-Verschuren, Sylvia; Roose, Marit; Venema, Sandra; Germeraad, Evelien; Engelsma, Marc; Heutink, Rene |
| EPI1987341 | NP | Netherlands | 2022-Feb-04 | EPI_ISL_10255580 | A/wild bird/Netherlands/22002833-002/2022 | Wageningen Bioveterinary Research | Wageningen Bioveterinary Research | Beerens, Nancy; Harders, Frank; Pritz-Verschuren, Sylvia; Roose, Marit; Venema, Sandra; Germeraad, Evelien; Engelsma, Marc; Heutink, Rene |
| EPI1987342 | NS | Netherlands | 2022-Feb-04 | EPI_ISL_10255580 | A/wild bird/Netherlands/22002833-002/2022 | Wageningen Bioveterinary Research | Wageningen Bioveterinary Research | Beerens, Nancy; Harders, Frank; Pritz-Verschuren, Sylvia; Roose, Marit; Venema, Sandra; Germeraad, Evelien; Engelsma, Marc; Heutink, Rene |
| EPI1987343 | MP | Netherlands | 2022-Feb-04 | EPI_ISL_10255580 | A/wild bird/Netherlands/22002833-002/2022 | Wageningen Bioveterinary Research | Wageningen Bioveterinary Research | Beerens, Nancy; Harders, Frank; Pritz-Verschuren, Sylvia; Roose, Marit; Venema, Sandra; Germeraad, Evelien; Engelsma, Marc; Heutink, Rene |
| EPI1987349 | NP | Netherlands | 2022-Feb-11 | EPI_ISL_10255581 | A/red fox/Netherlands/22002992-003/2022 | Wageningen Bioveterinary Research | Wageningen Bioveterinary Research | Beerens, Nancy; Harders, Frank; Pritz-Verschuren, Sylvia; Roose, Marit; Venema, Sandra; Germeraad, Evelien; Engelsma, Marc; Heutink, Rene |
| EPI1987350 | NS | Netherlands | 2022-Feb-11 | EPI_ISL_10255581 | A/red fox/Netherlands/22002992-003/2022 | Wageningen Bioveterinary Research | Wageningen Bioveterinary Research | Beerens, Nancy; Harders, Frank; Pritz-Verschuren, Sylvia; Roose, Marit; Venema, Sandra; Germeraad, Evelien; Engelsma, Marc; Heutink, Rene |
| EPI1987351 | MP | Netherlands | 2022-Feb-11 | EPI_ISL_10255581 | A/red fox/Netherlands/22002992-003/2022 | Wageningen Bioveterinary Research | Wageningen Bioveterinary Research | Beerens, Nancy; Harders, Frank; Pritz-Verschuren, Sylvia; Roose, Marit; Venema, Sandra; Germeraad, Evelien; Engelsma, Marc; Heutink, Rene |
| EPI1987357 | NP | Netherlands | 2022-Feb-11 | EPI_ISL_10255587 | A/red fox/Netherlands/22002993-003 /2022 | Wageningen Bioveterinary Research | Wageningen Bioveterinary Research | Beerens, Nancy; Harders, Frank; Pritz-Verschuren, Sylvia; Roose, Marit; Venema, Sandra; Germeraad, Evelien; Engelsma, Marc; Heutink, Rene |
| EPI1987358 | NS | Netherlands | 2022-Feb-11 | EPI_ISL_10255587 | A/red fox/Netherlands/22002993-003 /2022 | Wageningen Bioveterinary Research | Wageningen Bioveterinary Research | Beerens, Nancy; Harders, Frank; Pritz-Verschuren, Sylvia; Roose, Marit; Venema, Sandra; Germeraad, Evelien; Engelsma, Marc; Heutink, Rene |
| EPI1987359 | MP | Netherlands | 2022-Feb-11 | EPI_ISL_10255587 | A/red fox/Netherlands/22002993-003 /2022 | Wageningen Bioveterinary Research | Wageningen Bioveterinary Research | Beerens, Nancy; Harders, Frank; Pritz-Verschuren, Sylvia; Roose, Marit; Venema, Sandra; Germeraad, Evelien; Engelsma, Marc; Heutink, Rene |
| EPI1987376 | NP | Germany | 2022-Jan-24 | EPI_ISL_10261376 | A/pigeon/Germany-NW/AI00951/2022 | Chemisches- und Veterinäruntersuchungsamt Westfalen | Friedrich-Loeffler-Institut | |
| EPI1987374 | MP | Germany | 2022-Jan-24 | EPI_ISL_10261376 | A/pigeon/Germany-NW/AI00951/2022 | Chemisches- und Veterinäruntersuchungsamt Westfalen | Friedrich-Loeffler-Institut | |
| EPI1987377 | NS | Germany | 2022-Jan-24 | EPI_ISL_10261376 | A/pigeon/Germany-NW/AI00951/2022 | Chemisches- und Veterinäruntersuchungsamt Westfalen | Friedrich-Loeffler-Institut | |

| | | | | | | | | |
|---|---|---|---|---|---|---|---|---|
| EPI1987667 | NP | Netherlands | 2022-Jan-28 | EPI_ISL_10347219 | A/Barnacle Goose/Netherlands/8/2022 | Erasmus Medical Center | Erasmus Medical Center | |
| EPI1987668 | NS | Netherlands | 2022-Jan-28 | EPI_ISL_10347219 | A/Barnacle Goose/Netherlands/8/2022 | Erasmus Medical Center | Erasmus Medical Center | |
| EPI1987669 | MP | Netherlands | 2022-Jan-28 | EPI_ISL_10347219 | A/Barnacle Goose/Netherlands/8/2022 | Erasmus Medical Center | Erasmus Medical Center | |
| EPI1988030 | NP | Netherlands | 2022-Feb-02 | EPI_ISL_10454874 | A/barnacle goose/Netherlands/22002507-002/2022 | Wageningen Bioveterinary Research | Wageningen Bioveterinary Research | Beerens, Nancy; Harders, Frank; Pritz-Verschuren, Sylvia; Roose, Marit; Venema, Sandra; Germeraad, Evelien; Engelsma, Marc; Heutink, Rene |
| EPI1988031 | NS | Netherlands | 2022-Feb-02 | EPI_ISL_10454874 | A/barnacle goose/Netherlands/22002507-002/2022 | Wageningen Bioveterinary Research | Wageningen Bioveterinary Research | Beerens, Nancy; Harders, Frank; Pritz-Verschuren, Sylvia; Roose, Marit; Venema, Sandra; Germeraad, Evelien; Engelsma, Marc; Heutink, Rene |
| EPI1988032 | MP | Netherlands | 2022-Feb-02 | EPI_ISL_10454874 | A/barnacle goose/Netherlands/22002507-002/2022 | Wageningen Bioveterinary Research | Wageningen Bioveterinary Research | Beerens, Nancy; Harders, Frank; Pritz-Verschuren, Sylvia; Roose, Marit; Venema, Sandra; Germeraad, Evelien; Engelsma, Marc; Heutink, Rene |
| EPI1988038 | NP | Netherlands | 2022-Feb-03 | EPI_ISL_10454886 | A/barnacle goose/Netherlands/22002511-001/2022 | Wageningen Bioveterinary Research | Wageningen Bioveterinary Research | Beerens, Nancy; Harders, Frank; Pritz-Verschuren, Sylvia; Roose, Marit; Venema, Sandra; Germeraad, Evelien; Engelsma, Marc; Heutink, Rene |
| EPI1988039 | NS | Netherlands | 2022-Feb-03 | EPI_ISL_10454886 | A/barnacle goose/Netherlands/22002511-001/2022 | Wageningen Bioveterinary Research | Wageningen Bioveterinary Research | Beerens, Nancy; Harders, Frank; Pritz-Verschuren, Sylvia; Roose, Marit; Venema, Sandra; Germeraad, Evelien; Engelsma, Marc; Heutink, Rene |
| EPI1988040 | MP | Netherlands | 2022-Feb-03 | EPI_ISL_10454886 | A/barnacle goose/Netherlands/22002511-001/2022 | Wageningen Bioveterinary Research | Wageningen Bioveterinary Research | Beerens, Nancy; Harders, Frank; Pritz-Verschuren, Sylvia; Roose, Marit; Venema, Sandra; Germeraad, Evelien; Engelsma, Marc; Heutink, Rene |
| EPI1988046 | NP | Netherlands | 2022-Feb-18 | EPI_ISL_10454887 | A/red fox/Netherlands/22003495-003/2022 | Wageningen Bioveterinary Research | Wageningen Bioveterinary Research | Beerens, Nancy; Harders, Frank; Pritz-Verschuren, Sylvia; Roose, Marit; Venema, Sandra; Germeraad, Evelien; Engelsma, Marc; Heutink, Rene |
| EPI1988047 | NS | Netherlands | 2022-Feb-18 | EPI_ISL_10454887 | A/red fox/Netherlands/22003495-003/2022 | Wageningen Bioveterinary Research | Wageningen Bioveterinary Research | Beerens, Nancy; Harders, Frank; Pritz-Verschuren, Sylvia; Roose, Marit; Venema, Sandra; Germeraad, Evelien; Engelsma, Marc; Heutink, Rene |
| EPI1988048 | MP | Netherlands | 2022-Feb-18 | EPI_ISL_10454887 | A/red fox/Netherlands/22003495-003/2022 | Wageningen Bioveterinary Research | Wageningen Bioveterinary Research | Beerens, Nancy; Harders, Frank; Pritz-Verschuren, Sylvia; Roose, Marit; Venema, Sandra; Germeraad, Evelien; Engelsma, Marc; Heutink, Rene |
| EPI1988854 | MP | Germany | 2022-Jan-25 | EPI_ISL_10497305 | A/red knot/Germany-SH/AI01010/2022 | Landeslabor Schleswig-Holstein | Friedrich-Loeffler-Institut | |
| EPI1988856 | NP | Germany | 2022-Jan-25 | EPI_ISL_10497305 | A/red knot/Germany-SH/AI01010/2022 | Landeslabor Schleswig-Holstein | Friedrich-Loeffler-Institut | |
| EPI1988857 | NS | Germany | 2022-Jan-25 | EPI_ISL_10497305 | A/red knot/Germany-SH/AI01010/2022 | Landeslabor Schleswig-Holstein | Friedrich-Loeffler-Institut | |
| EPI1988865 | NS | Germany | 2022-Feb-11 | EPI_ISL_10497306 | A/buzzard/Germany-BB/AI01212/2022 | Landeslabor Berlin-Brandenburg | Friedrich-Loeffler-Institut | |
| EPI1988862 | MP | Germany | 2022-Feb-11 | EPI_ISL_10497306 | A/buzzard/Germany-BB/AI01212/2022 | Landeslabor Berlin-Brandenburg | Friedrich-Loeffler-Institut | |
| EPI1988864 | NP | Germany | 2022-Feb-11 | EPI_ISL_10497306 | A/buzzard/Germany-BB/AI01212/2022 | Landeslabor Berlin-Brandenburg | Friedrich-Loeffler-Institut | |
| EPI1988978 | NP | Belgium | 2022-Jan-23 | EPI_ISL_10576444 | A/Anser_anser/Belgium/1809_0002/2022 | Sciensano - Animal Infectious Diseases | Sciensano, Department of Animal Infectious Diseases | Van Borm, Steven; Roupie, Virginie; Lambrecht, Benedicte; Mathijs, Elisabeth; Steensels, Mieke |
| EPI1988981 | NS | Belgium | 2022-Jan-23 | EPI_ISL_10576444 | A/Anser_anser/Belgium/1809_0002/2022 | Sciensano - Animal Infectious Diseases | Sciensano, Department of Animal Infectious Diseases | Van Borm, Steven; Roupie, Virginie; Lambrecht, Benedicte; Mathijs, Elisabeth; Steensels, Mieke |
| EPI1988980 | MP | Belgium | 2022-Jan-23 | EPI_ISL_10576444 | A/Anser_anser/Belgium/1809_0002/2022 | Sciensano - Animal Infectious Diseases | Sciensano, Department of Animal Infectious Diseases | Van Borm, Steven; Roupie, Virginie; Lambrecht, Benedicte; Mathijs, Elisabeth; Steensels, Mieke |
| EPI1990441 | NP | Netherlands | 2022-Feb-01 | EPI_ISL_10724259 | A/seagull/Netherlands/22002274-002/2022 | Wageningen Bioveterinary Research | Wageningen Bioveterinary Research | Beerens, Nancy; Harders, Frank; Pritz-Verschuren, Sylvia; Roose, Marit; Venema, Sandra; Germeraad, Evelien; Engelsma, Marc; Heutink, Rene |
| EPI1990442 | NS | Netherlands | 2022-Feb-01 | EPI_ISL_10724259 | A/seagull/Netherlands/22002274-002/2022 | Wageningen Bioveterinary Research | Wageningen Bioveterinary Research | Beerens, Nancy; Harders, Frank; Pritz-Verschuren, Sylvia; Roose, Marit; Venema, Sandra; Germeraad, Evelien; Engelsma, Marc; Heutink, Rene |
| EPI1990443 | MP | Netherlands | 2022-Feb-01 | EPI_ISL_10724259 | A/seagull/Netherlands/22002274-002/2022 | Wageningen Bioveterinary Research | Wageningen Bioveterinary Research | Beerens, Nancy; Harders, Frank; Pritz-Verschuren, Sylvia; Roose, Marit; Venema, Sandra; Germeraad, Evelien; Engelsma, Marc; Heutink, Rene |
| EPI1994439 | NP | Netherlands | 2022-Mar-01 | EPI_ISL_10915976 | A/red fox/Netherlands/22004172-003/2022 | Wageningen Bioveterinary Research | Wageningen Bioveterinary Research | Beerens, Nancy; Harders, Frank; Pritz-Verschuren, Sylvia; Roose, Marit; Venema, Sandra; Germeraad, Evelien; Engelsma, Marc; Heutink, Rene |
| EPI1994440 | NS | Netherlands | 2022-Mar-01 | EPI_ISL_10915976 | A/red fox/Netherlands/22004172-003/2022 | Wageningen Bioveterinary Research | Wageningen Bioveterinary Research | Beerens, Nancy; Harders, Frank; Pritz-Verschuren, Sylvia; Roose, Marit; Venema, Sandra; Germeraad, Evelien; Engelsma, Marc; Heutink, Rene |
| EPI1994441 | MP | Netherlands | 2022-Mar-01 | EPI_ISL_10915976 | A/red fox/Netherlands/22004172-003/2022 | Wageningen Bioveterinary Research | Wageningen Bioveterinary Research | Beerens, Nancy; Harders, Frank; Pritz-Verschuren, Sylvia; Roose, Marit; Venema, Sandra; Germeraad, Evelien; Engelsma, Marc; Heutink, Rene |
| EPI1994516 | NP | Sweden | 2022-Jan-25 | EPI_ISL_10943657 | A/northern goshawk/Sweden/SVA220210SZ0305/FP000317/O-2022 | National Veterinary Institute, SVA | National Veterinary Institute | |
| EPI1994518 | MP | Sweden | 2022-Jan-25 | EPI_ISL_10943657 | A/northern goshawk/Sweden/SVA220210SZ0305/FP000317/O-2022 | National Veterinary Institute, SVA | National Veterinary Institute | |
| EPI1994519 | NS | Sweden | 2022-Jan-25 | EPI_ISL_10943657 | A/northern goshawk/Sweden/SVA220210SZ0305/FP000317/O-2022 | National Veterinary Institute, SVA | National Veterinary Institute | |
| EPI1994982 | NP | Netherlands | 2022-Feb-12 | EPI_ISL_10992813 | A/Mallard/Netherlands/1/2022 | Erasmus Medical Center | Erasmus Medical Center | |
| EPI1994983 | NS | Netherlands | 2022-Feb-12 | EPI_ISL_10992813 | A/Mallard/Netherlands/1/2022 | Erasmus Medical Center | Erasmus Medical Center | |
| EPI1994984 | MP | Netherlands | 2022-Feb-12 | EPI_ISL_10992813 | A/Mallard/Netherlands/1/2022 | Erasmus Medical Center | Erasmus Medical Center | |
| EPI1994990 | NP | Netherlands | 2022-Feb-14 | EPI_ISL_10993074 | A/Mallard/Netherlands/2/2022 | Erasmus Medical Center | Erasmus Medical Center | |
| EPI1994991 | NS | Netherlands | 2022-Feb-14 | EPI_ISL_10993074 | A/Mallard/Netherlands/2/2022 | Erasmus Medical Center | Erasmus Medical Center | |

| | | | | | | | | |
|---|---|---|---|---|---|---|---|---|
| EPI1994992 | MP | Netherlands | 2022-Feb-14 | EPI_ISL_10993074 | A/Mallard/Netherlands/2/2022 | Erasmus Medical Center | Erasmus Medical Center | |
| EPI1995038 | NP | Netherlands | 2022-Jan-29 | EPI_ISL_10993196 | A/Sanderling/Netherlands/1/2022 | Erasmus Medical Center | Erasmus Medical Center | |
| EPI1995039 | NS | Netherlands | 2022-Jan-29 | EPI_ISL_10993196 | A/Sanderling/Netherlands/1/2022 | Erasmus Medical Center | Erasmus Medical Center | |
| EPI1995040 | MP | Netherlands | 2022-Jan-29 | EPI_ISL_10993196 | A/Sanderling/Netherlands/1/2022 | Erasmus Medical Center | Erasmus Medical Center | |
| EPI1995266 | NP | Netherlands | 2022-Jan-03 | EPI_ISL_11019784 | A/red fox/Netherlands/22000097-005/2022 | Wageningen Bioveterinary Research | Wageningen Bioveterinary Research | Beerens, Nancy; Harders, Frank; Pritz-Verschuren, Sylvia; Roose, Marit; Venema, Sandra; Germeraad, Evelien; Engelsma, Marc; Heutink, Rene |
| EPI1995267 | NS | Netherlands | 2022-Jan-03 | EPI_ISL_11019784 | A/red fox/Netherlands/22000097-005/2022 | Wageningen Bioveterinary Research | Wageningen Bioveterinary Research | Beerens, Nancy; Harders, Frank; Pritz-Verschuren, Sylvia; Roose, Marit; Venema, Sandra; Germeraad, Evelien; Engelsma, Marc; Heutink, Rene |
| EPI1995268 | MP | Netherlands | 2022-Jan-03 | EPI_ISL_11019784 | A/red fox/Netherlands/22000097-005/2022 | Wageningen Bioveterinary Research | Wageningen Bioveterinary Research | Beerens, Nancy; Harders, Frank; Pritz-Verschuren, Sylvia; Roose, Marit; Venema, Sandra; Germeraad, Evelien; Engelsma, Marc; Heutink, Rene |
| EPI1995275 | NP | Netherlands | 2022-Feb-07 | EPI_ISL_11019882 | A/red fox/Netherlands/22002636-002/2022 | Wageningen Bioveterinary Research | Wageningen Bioveterinary Research | Beerens, Nancy; Harders, Frank; Pritz-Verschuren, Sylvia; Roose, Marit; Venema, Sandra; Germeraad, Evelien; Engelsma, Marc; Heutink, Rene |
| EPI1995276 | NS | Netherlands | 2022-Feb-07 | EPI_ISL_11019882 | A/red fox/Netherlands/22002636-002/2022 | Wageningen Bioveterinary Research | Wageningen Bioveterinary Research | Beerens, Nancy; Harders, Frank; Pritz-Verschuren, Sylvia; Roose, Marit; Venema, Sandra; Germeraad, Evelien; Engelsma, Marc; Heutink, Rene |
| EPI1995277 | MP | Netherlands | 2022-Feb-07 | EPI_ISL_11019882 | A/red fox/Netherlands/22002636-002/2022 | Wageningen Bioveterinary Research | Wageningen Bioveterinary Research | Beerens, Nancy; Harders, Frank; Pritz-Verschuren, Sylvia; Roose, Marit; Venema, Sandra; Germeraad, Evelien; Engelsma, Marc; Heutink, Rene |
| EPI1995555 | NP | Netherlands | 2022-Feb-07 | EPI_ISL_11112340 | A/Barnacle Goose/Netherlands/10/2022 | Erasmus Medical Center | Erasmus Medical Center | |
| EPI1995557 | NS | Netherlands | 2022-Feb-07 | EPI_ISL_11112340 | A/Barnacle Goose/Netherlands/10/2022 | Erasmus Medical Center | Erasmus Medical Center | |
| EPI1995559 | MP | Netherlands | 2022-Feb-07 | EPI_ISL_11112340 | A/Barnacle Goose/Netherlands/10/2022 | Erasmus Medical Center | Erasmus Medical Center | |
| EPI1995571 | NP | Netherlands | 2022-Feb-09 | EPI_ISL_11112342 | A/Barnacle Goose/Netherlands/11/2022 | Erasmus Medical Center | Erasmus Medical Center | |
| EPI1995573 | NS | Netherlands | 2022-Feb-09 | EPI_ISL_11112342 | A/Barnacle Goose/Netherlands/11/2022 | Erasmus Medical Center | Erasmus Medical Center | |
| EPI1995575 | MP | Netherlands | 2022-Feb-09 | EPI_ISL_11112342 | A/Barnacle Goose/Netherlands/11/2022 | Erasmus Medical Center | Erasmus Medical Center | |
| EPI1995588 | NP | Netherlands | 2022-Feb-09 | EPI_ISL_11112344 | A/Barnacle Goose/Netherlands/12/2022 | Erasmus Medical Center | Erasmus Medical Center | |
| EPI1995590 | NS | Netherlands | 2022-Feb-09 | EPI_ISL_11112344 | A/Barnacle Goose/Netherlands/12/2022 | Erasmus Medical Center | Erasmus Medical Center | |
| EPI1995591 | MP | Netherlands | 2022-Feb-09 | EPI_ISL_11112344 | A/Barnacle Goose/Netherlands/12/2022 | Erasmus Medical Center | Erasmus Medical Center | |
| EPI1995604 | NP | Netherlands | 2022-Feb-23 | EPI_ISL_11112346 | A/Barnacle Goose/Netherlands/13/2022 | Erasmus Medical Center | Erasmus Medical Center | |
| EPI1995606 | NS | Netherlands | 2022-Feb-23 | EPI_ISL_11112346 | A/Barnacle Goose/Netherlands/13/2022 | Erasmus Medical Center | Erasmus Medical Center | |
| EPI1995607 | MP | Netherlands | 2022-Feb-23 | EPI_ISL_11112346 | A/Barnacle Goose/Netherlands/13/2022 | Erasmus Medical Center | Erasmus Medical Center | |
| EPI1995620 | NP | Netherlands | 2022-Feb-23 | EPI_ISL_11112348 | A/Barnacle Goose/Netherlands/14/2022 | Erasmus Medical Center | Erasmus Medical Center | |
| EPI1995622 | NS | Netherlands | 2022-Feb-23 | EPI_ISL_11112348 | A/Barnacle Goose/Netherlands/14/2022 | Erasmus Medical Center | Erasmus Medical Center | |
| EPI1995623 | MP | Netherlands | 2022-Feb-23 | EPI_ISL_11112348 | A/Barnacle Goose/Netherlands/14/2022 | Erasmus Medical Center | Erasmus Medical Center | |
| EPI1995631 | NP | Netherlands | 2022-Feb-26 | EPI_ISL_11112350 | A/Barnacle Goose/Netherlands/15/2022 | Erasmus Medical Center | Erasmus Medical Center | |
| EPI1995633 | NS | Netherlands | 2022-Feb-26 | EPI_ISL_11112350 | A/Barnacle Goose/Netherlands/15/2022 | Erasmus Medical Center | Erasmus Medical Center | |
| EPI1995635 | MP | Netherlands | 2022-Feb-26 | EPI_ISL_11112350 | A/Barnacle Goose/Netherlands/15/2022 | Erasmus Medical Center | Erasmus Medical Center | |
| EPI1995647 | NP | Netherlands | 2022-Feb-07 | EPI_ISL_11112352 | A/Barnacle Goose/Netherlands/9/2022 | Erasmus Medical Center | Erasmus Medical Center | |
| EPI1995649 | NS | Netherlands | 2022-Feb-07 | EPI_ISL_11112352 | A/Barnacle Goose/Netherlands/9/2022 | Erasmus Medical Center | Erasmus Medical Center | |
| EPI1995651 | MP | Netherlands | 2022-Feb-07 | EPI_ISL_11112352 | A/Barnacle Goose/Netherlands/9/2022 | Erasmus Medical Center | Erasmus Medical Center | |
| EPI1995663 | NP | Netherlands | 2022-Feb-01 | EPI_ISL_11112354 | A/Common Gull/Netherlands/1/2022 | Erasmus Medical Center | Erasmus Medical Center | |
| EPI1995665 | NS | Netherlands | 2022-Feb-01 | EPI_ISL_11112354 | A/Common Gull/Netherlands/1/2022 | Erasmus Medical Center | Erasmus Medical Center | |
| EPI1995667 | MP | Netherlands | 2022-Feb-01 | EPI_ISL_11112354 | A/Common Gull/Netherlands/1/2022 | Erasmus Medical Center | Erasmus Medical Center | |
| EPI1995679 | NP | Netherlands | 2022-Feb-09 | EPI_ISL_11112356 | A/Eurasian Curlew/Netherlands/1/2022 | Erasmus Medical Center | Erasmus Medical Center | |

| | | | | | | | | |
|---|---|---|---|---|---|---|---|---|
| EPI1995681 | NS | Netherlands | 2022-Feb-09 | EPI_ISL_11112356 | A/Eurasian Curlew/Netherlands/1/2022 | Erasmus Medical Center | Erasmus Medical Center | |
| EPI1995683 | MP | Netherlands | 2022-Feb-09 | EPI_ISL_11112356 | A/Eurasian Curlew/Netherlands/1/2022 | Erasmus Medical Center | Erasmus Medical Center | |
| EPI1995695 | NP | Netherlands | 2022-Feb-26 | EPI_ISL_11112358 | A/European Herring Gull/Netherlands/2/2022 | Erasmus Medical Center | Erasmus Medical Center | |
| EPI1995697 | NS | Netherlands | 2022-Feb-26 | EPI_ISL_11112358 | A/European Herring Gull/Netherlands/2/2022 | Erasmus Medical Center | Erasmus Medical Center | |
| EPI1995699 | MP | Netherlands | 2022-Feb-26 | EPI_ISL_11112358 | A/European Herring Gull/Netherlands/2/2022 | Erasmus Medical Center | Erasmus Medical Center | |
| EPI1995711 | NP | Netherlands | 2022-Feb-23 | EPI_ISL_11112360 | A/Great black-backed Gull/Netherlands/2/2022 | Erasmus Medical Center | Erasmus Medical Center | |
| EPI1995713 | NS | Netherlands | 2022-Feb-23 | EPI_ISL_11112360 | A/Great black-backed Gull/Netherlands/2/2022 | Erasmus Medical Center | Erasmus Medical Center | |
| EPI1995715 | MP | Netherlands | 2022-Feb-23 | EPI_ISL_11112360 | A/Great black-backed Gull/Netherlands/2/2022 | Erasmus Medical Center | Erasmus Medical Center | |
| EPI1995727 | NP | Netherlands | 2022-Feb-02 | EPI_ISL_11112362 | A/Greylag Goose/Netherlands/1/2022 | Erasmus Medical Center | Erasmus Medical Center | |
| EPI1995729 | NS | Netherlands | 2022-Feb-02 | EPI_ISL_11112362 | A/Greylag Goose/Netherlands/1/2022 | Erasmus Medical Center | Erasmus Medical Center | |
| EPI1995731 | MP | Netherlands | 2022-Feb-02 | EPI_ISL_11112362 | A/Greylag Goose/Netherlands/1/2022 | Erasmus Medical Center | Erasmus Medical Center | |
| EPI1995840 | NP | Netherlands | 2022-Feb-23 | EPI_ISL_11112484 | A/Caspian Gull/Netherlands/2/2022 | Erasmus Medical Center | Erasmus Medical Center | |
| EPI1995841 | NS | Netherlands | 2022-Feb-23 | EPI_ISL_11112484 | A/Caspian Gull/Netherlands/2/2022 | Erasmus Medical Center | Erasmus Medical Center | |
| EPI1995842 | MP | Netherlands | 2022-Feb-23 | EPI_ISL_11112484 | A/Caspian Gull/Netherlands/2/2022 | Erasmus Medical Center | Erasmus Medical Center | |
| EPI1998131 | NP | Ireland | 2022-Feb-14 | EPI_ISL_11259283 | A/fox/Ireland/3866_22VIR2064-1/2022 | Central Veterinary Research Laboratory | Istituto Zooprofilattico Sperimentale delle Venezie | Byrne, C.; Garcia, K.; Cuartero, L.G.; Barbierato, G.; Zecchin, B.; Fusaro, A.; Schivo, A.; Salviato, A.; Palumbo, E.; Giussani, E.; Monne, I.; Terregino, C. |
| EPI1998132 | NS | Ireland | 2022-Feb-14 | EPI_ISL_11259283 | A/fox/Ireland/3866_22VIR2064-1/2022 | Central Veterinary Research Laboratory | Istituto Zooprofilattico Sperimentale delle Venezie | Byrne, C.; Garcia, K.; Cuartero, L.G.; Barbierato, G.; Zecchin, B.; Fusaro, A.; Schivo, A.; Salviato, A.; Palumbo, E.; Giussani, E.; Monne, I.; Terregino, C. |
| EPI1998133 | MP | Ireland | 2022-Feb-14 | EPI_ISL_11259283 | A/fox/Ireland/3866_22VIR2064-1/2022 | Central Veterinary Research Laboratory | Istituto Zooprofilattico Sperimentale delle Venezie | Byrne, C.; Garcia, K.; Cuartero, L.G.; Barbierato, G.; Zecchin, B.; Fusaro, A.; Schivo, A.; Salviato, A.; Palumbo, E.; Giussani, E.; Monne, I.; Terregino, C. |
| EPI1998139 | NP | Ireland | 2022-Feb-14 | EPI_ISL_11259284 | A/goose/Ireland/3869_22VIR2064-2/2022 | Central Veterinary Research Laboratory | Istituto Zooprofilattico Sperimentale delle Venezie | Byrne, C.; Garcia, K.; Cuartero, L.G.; Barbierato, G.; Zecchin, B.; Fusaro, A.; Schivo, A.; Salviato, A.; Palumbo, E.; Giussani, E.; Monne, I.; Terregino, C. |
| EPI1998140 | NS | Ireland | 2022-Feb-14 | EPI_ISL_11259284 | A/goose/Ireland/3869_22VIR2064-2/2022 | Central Veterinary Research Laboratory | Istituto Zooprofilattico Sperimentale delle Venezie | Byrne, C.; Garcia, K.; Cuartero, L.G.; Barbierato, G.; Zecchin, B.; Fusaro, A.; Schivo, A.; Salviato, A.; Palumbo, E.; Giussani, E.; Monne, I.; Terregino, C. |
| EPI1998141 | MP | Ireland | 2022-Feb-14 | EPI_ISL_11259284 | A/goose/Ireland/3869_22VIR2064-2/2022 | Central Veterinary Research Laboratory | Istituto Zooprofilattico Sperimentale delle Venezie | Byrne, C.; Garcia, K.; Cuartero, L.G.; Barbierato, G.; Zecchin, B.; Fusaro, A.; Schivo, A.; Salviato, A.; Palumbo, E.; Giussani, E.; Monne, I.; Terregino, C. |
| EPI2001589 | NP | United Kingdom | 2022-Jan-06 | EPI_ISL_11406398 | A/chicken/England/002070/2022 | Animal and Plant Health Agency (APHA) | Animal and Plant Health Agency (APHA) | |
| EPI2001590 | NS | United Kingdom | 2022-Jan-06 | EPI_ISL_11406398 | A/chicken/England/002070/2022 | Animal and Plant Health Agency (APHA) | Animal and Plant Health Agency (APHA) | |
| EPI2001591 | MP | United Kingdom | 2022-Jan-06 | EPI_ISL_11406398 | A/chicken/England/002070/2022 | Animal and Plant Health Agency (APHA) | Animal and Plant Health Agency (APHA) | |
| EPI2001597 | NP | United Kingdom | 2022-Jan-12 | EPI_ISL_11406399 | A/turkey/England/004737/2022 | Animal and Plant Health Agency (APHA) | Animal and Plant Health Agency (APHA) | |
| EPI2001598 | NS | United Kingdom | 2022-Jan-12 | EPI_ISL_11406399 | A/turkey/England/004737/2022 | Animal and Plant Health Agency (APHA) | Animal and Plant Health Agency (APHA) | |
| EPI2001599 | MP | United Kingdom | 2022-Jan-12 | EPI_ISL_11406399 | A/turkey/England/004737/2022 | Animal and Plant Health Agency (APHA) | Animal and Plant Health Agency (APHA) | |
| EPI2001613 | NP | United Kingdom | 2022-Jan-03 | EPI_ISL_11406401 | A/chicken/England/000187/2022 | Animal and Plant Health Agency (APHA) | Animal and Plant Health Agency (APHA) | |
| EPI2001614 | NS | United Kingdom | 2022-Jan-03 | EPI_ISL_11406401 | A/chicken/England/000187/2022 | Animal and Plant Health Agency (APHA) | Animal and Plant Health Agency (APHA) | |
| EPI2001615 | MP | United Kingdom | 2022-Jan-03 | EPI_ISL_11406401 | A/chicken/England/000187/2022 | Animal and Plant Health Agency (APHA) | Animal and Plant Health Agency (APHA) | |
| EPI2001621 | NP | United Kingdom | 2022-Jan-23 | EPI_ISL_11406402 | A/domestic_duck/England/007588/2022 | Animal and Plant Health Agency (APHA) | Animal and Plant Health Agency (APHA) | |
| EPI2001622 | NS | United Kingdom | 2022-Jan-23 | EPI_ISL_11406402 | A/domestic_duck/England/007588/2022 | Animal and Plant Health Agency (APHA) | Animal and Plant Health Agency (APHA) | |
| EPI2001623 | MP | United Kingdom | 2022-Jan-23 | EPI_ISL_11406402 | A/domestic_duck/England/007588/2022 | Animal and Plant Health Agency (APHA) | Animal and Plant Health Agency (APHA) | |
| EPI2001629 | NP | United Kingdom | 2022-Jan-31 | EPI_ISL_11406403 | A/black-headed_gull/England/306270/2022 | Animal and Plant Health Agency (APHA) | Animal and Plant Health Agency (APHA) | |
| EPI2001630 | NS | United Kingdom | 2022-Jan-31 | EPI_ISL_11406403 | A/black-headed_gull/England/306270/2022 | Animal and Plant Health Agency (APHA) | Animal and Plant Health Agency (APHA) | |
| EPI2001631 | MP | United Kingdom | 2022-Jan-31 | EPI_ISL_11406403 | A/black-headed_gull/England/306270/2022 | Animal and Plant Health Agency (APHA) | Animal and Plant Health Agency (APHA) | |

| | | | | | | | | |
|---|---|---|---|---|---|---|---|---|
| EPI2002653 | NS | Belgium | 2022-Mar-10 | EPI_ISL_11449674 | A/Gallus_gallus/Belgium/3194_0001/2022 | Sciensano - Animal Infectious Diseases | Sciensano, Department of Animal Infectious Diseases | Van Borm, Steven; Roupie, Virginie; Lambrecht, Benedicte; Mathijs, Elisabeth; Steensels, Mieke |
| EPI2002650 | NP | Belgium | 2022-Mar-10 | EPI_ISL_11449674 | A/Gallus_gallus/Belgium/3194_0001/2022 | Sciensano - Animal Infectious Diseases | Sciensano, Department of Animal Infectious Diseases | Van Borm, Steven; Roupie, Virginie; Lambrecht, Benedicte; Mathijs, Elisabeth; Steensels, Mieke |
| EPI2002652 | MP | Belgium | 2022-Mar-10 | EPI_ISL_11449674 | A/Gallus_gallus/Belgium/3194_0001/2022 | Sciensano - Animal Infectious Diseases | Sciensano, Department of Animal Infectious Diseases | Van Borm, Steven; Roupie, Virginie; Lambrecht, Benedicte; Mathijs, Elisabeth; Steensels, Mieke |
| EPI2008046 | NP | Netherlands | 2022-Mar-03 | EPI_ISL_11560233 | A/Brent goose/Netherlands/1/2022 | Erasmus Medical Center | Erasmus Medical Center | |
| EPI2008047 | NS | Netherlands | 2022-Mar-03 | EPI_ISL_11560233 | A/Brent goose/Netherlands/1/2022 | Erasmus Medical Center | Erasmus Medical Center | |
| EPI2008048 | MP | Netherlands | 2022-Mar-03 | EPI_ISL_11560233 | A/Brent goose/Netherlands/1/2022 | Erasmus Medical Center | Erasmus Medical Center | |
| EPI2008054 | NP | Netherlands | 2022-Mar-03 | EPI_ISL_11560235 | A/Barnacle Goose/Netherlands/16/2022 | Erasmus Medical Center | Erasmus Medical Center | |
| EPI2008055 | NS | Netherlands | 2022-Mar-03 | EPI_ISL_11560235 | A/Barnacle Goose/Netherlands/16/2022 | Erasmus Medical Center | Erasmus Medical Center | |
| EPI2008056 | MP | Netherlands | 2022-Mar-03 | EPI_ISL_11560235 | A/Barnacle Goose/Netherlands/16/2022 | Erasmus Medical Center | Erasmus Medical Center | |
| EPI2008062 | NP | Netherlands | 2022-Mar-03 | EPI_ISL_11560236 | A/Barnacle Goose/Netherlands/17/2022 | Erasmus Medical Center | Erasmus Medical Center | |
| EPI2008063 | NS | Netherlands | 2022-Mar-03 | EPI_ISL_11560236 | A/Barnacle Goose/Netherlands/17/2022 | Erasmus Medical Center | Erasmus Medical Center | |
| EPI2008064 | MP | Netherlands | 2022-Mar-03 | EPI_ISL_11560236 | A/Barnacle Goose/Netherlands/17/2022 | Erasmus Medical Center | Erasmus Medical Center | |
| EPI2008070 | NP | Netherlands | 2022-Mar-04 | EPI_ISL_11560323 | A/Barnacle Goose/Netherlands/18/2022 | Erasmus Medical Center | Erasmus Medical Center | |
| EPI2008071 | NS | Netherlands | 2022-Mar-04 | EPI_ISL_11560323 | A/Barnacle Goose/Netherlands/18/2022 | Erasmus Medical Center | Erasmus Medical Center | |
| EPI2008072 | MP | Netherlands | 2022-Mar-04 | EPI_ISL_11560323 | A/Barnacle Goose/Netherlands/18/2022 | Erasmus Medical Center | Erasmus Medical Center | |
| EPI2008078 | NP | Netherlands | 2022-Mar-06 | EPI_ISL_11560324 | A/Graylag goose/Netherlands/2/20222 | Erasmus Medical Center | Erasmus Medical Center | |
| EPI2008079 | NS | Netherlands | 2022-Mar-06 | EPI_ISL_11560324 | A/Graylag goose/Netherlands/2/20222 | Erasmus Medical Center | Erasmus Medical Center | |
| EPI2008080 | MP | Netherlands | 2022-Mar-06 | EPI_ISL_11560324 | A/Graylag goose/Netherlands/2/20222 | Erasmus Medical Center | Erasmus Medical Center | |
| EPI2008086 | NP | Netherlands | 2022-Mar-15 | EPI_ISL_11560325 | A/Great black-backed Gull/Netherlands/3/2022 | Erasmus Medical Center | Erasmus Medical Center | |
| EPI2008087 | NS | Netherlands | 2022-Mar-15 | EPI_ISL_11560325 | A/Great black-backed Gull/Netherlands/3/2022 | Erasmus Medical Center | Erasmus Medical Center | |
| EPI2008088 | MP | Netherlands | 2022-Mar-15 | EPI_ISL_11560325 | A/Great black-backed Gull/Netherlands/3/2022 | Erasmus Medical Center | Erasmus Medical Center | |
| EPI2008094 | NP | United Kingdom | 2022-Feb-02 | EPI_ISL_11561589 | A/chicken/England/011981/2022 | Animal and Plant Health Agency (APHA) | Animal and Plant Health Agency (APHA) | |
| EPI2008095 | NS | United Kingdom | 2022-Feb-02 | EPI_ISL_11561589 | A/chicken/England/011981/2022 | Animal and Plant Health Agency (APHA) | Animal and Plant Health Agency (APHA) | |
| EPI2008096 | MP | United Kingdom | 2022-Feb-02 | EPI_ISL_11561589 | A/chicken/England/011981/2022 | Animal and Plant Health Agency (APHA) | Animal and Plant Health Agency (APHA) | |
| EPI2008102 | NP | United Kingdom | 2022-Feb-08 | EPI_ISL_11561590 | A/chicken/England/012967/2022 | Animal and Plant Health Agency (APHA) | Animal and Plant Health Agency (APHA) | |
| EPI2008103 | NS | United Kingdom | 2022-Feb-08 | EPI_ISL_11561590 | A/chicken/England/012967/2022 | Animal and Plant Health Agency (APHA) | Animal and Plant Health Agency (APHA) | |
| EPI2008104 | MP | United Kingdom | 2022-Feb-08 | EPI_ISL_11561590 | A/chicken/England/012967/2022 | Animal and Plant Health Agency (APHA) | Animal and Plant Health Agency (APHA) | |
| EPI2008110 | NP | United Kingdom | 2022-Feb-08 | EPI_ISL_11561591 | A/domestic_duck/England/012973/2022 | Animal and Plant Health Agency (APHA) | Animal and Plant Health Agency (APHA) | |
| EPI2008111 | NS | United Kingdom | 2022-Feb-08 | EPI_ISL_11561591 | A/domestic_duck/England/012973/2022 | Animal and Plant Health Agency (APHA) | Animal and Plant Health Agency (APHA) | |
| EPI2008112 | MP | United Kingdom | 2022-Feb-08 | EPI_ISL_11561591 | A/domestic_duck/England/012973/2022 | Animal and Plant Health Agency (APHA) | Animal and Plant Health Agency (APHA) | |
| EPI2008118 | NP | United Kingdom | 2022-Feb-20 | EPI_ISL_11561592 | A/turkey/England/016515/2022 | Animal and Plant Health Agency (APHA) | Animal and Plant Health Agency (APHA) | |
| EPI2008119 | NS | United Kingdom | 2022-Feb-20 | EPI_ISL_11561592 | A/turkey/England/016515/2022 | Animal and Plant Health Agency (APHA) | Animal and Plant Health Agency (APHA) | |
| EPI2008120 | MP | United Kingdom | 2022-Feb-20 | EPI_ISL_11561592 | A/turkey/England/016515/2022 | Animal and Plant Health Agency (APHA) | Animal and Plant Health Agency (APHA) | |
| EPI2008126 | NP | United Kingdom | 2022-Feb-03 | EPI_ISL_11561593 | A/black-headed_gull/England/388256/2022 | Animal and Plant Health Agency (APHA) | Animal and Plant Health Agency (APHA) | |
| EPI2008127 | NS | United Kingdom | 2022-Feb-03 | EPI_ISL_11561593 | A/black-headed_gull/England/388256/2022 | Animal and Plant Health Agency (APHA) | Animal and Plant Health Agency (APHA) | |

| | | | | | | | | |
|---|---|---|---|---|---|---|---|---|
| EPI2008128 | MP | United Kingdom | 2022-Feb-03 | EPI_ISL_11561593 | A/black-headed_gull/England/388256/2022 | Animal and Plant Health Agency (APHA) | Animal and Plant Health Agency (APHA) | |
| EPI2008525 | NS | Sweden | 2022-Jan-21 | EPI_ISL_11580255 | A/Greylag goose/Sweden/SVA220308SZ0382/FB000559/M-2022 | National Veterinary Institute, SVA | National Veterinary Institute | |
| EPI2008522 | NP | Sweden | 2022-Jan-21 | EPI_ISL_11580255 | A/Greylag goose/Sweden/SVA220308SZ0382/FB000559/M-2022 | National Veterinary Institute, SVA | National Veterinary Institute | |
| EPI2008524 | MP | Sweden | 2022-Jan-21 | EPI_ISL_11580255 | A/Greylag goose/Sweden/SVA220308SZ0382/FB000559/M-2022 | National Veterinary Institute, SVA | National Veterinary Institute | |
| EPI2013084 | NS | Belgium | 2022-Mar-28 | EPI_ISL_11880371 | A/Gallus_gallus/Belgium/4190_0002/2022 | Sciensano - Animal Infectious Diseases | Sciensano, Department of Animal Infectious Diseases | Van Borm, Steven; Roupie, Virginie; Lambrecht, Benedicte; Mathijs, Elisabeth; Steensels, Mieke |
| EPI2013083 | NP | Belgium | 2022-Mar-28 | EPI_ISL_11880371 | A/Gallus_gallus/Belgium/4190_0002/2022 | Sciensano - Animal Infectious Diseases | Sciensano, Department of Animal Infectious Diseases | Van Borm, Steven; Roupie, Virginie; Lambrecht, Benedicte; Mathijs, Elisabeth; Steensels, Mieke |
| EPI2013085 | MP | Belgium | 2022-Mar-28 | EPI_ISL_11880371 | A/Gallus_gallus/Belgium/4190_0002/2022 | Sciensano - Animal Infectious Diseases | Sciensano, Department of Animal Infectious Diseases | Van Borm, Steven; Roupie, Virginie; Lambrecht, Benedicte; Mathijs, Elisabeth; Steensels, Mieke |
| EPI2014986 | NP | Romania | 2022-Feb-01 | EPI_ISL_11922816 | A/swan/Romania/10324_22VIR2749-2/2022 | Institute for Diagnosis & Animal Health (IDAH) | Istituto Zooprofilattico Sperimentale delle Venezie | Barbuceanu, F.; Onita, I.; Neicut, A.; Motiu, R.; Burlacu, R.; Barbierato, G.; Zecchin, B.; Fusaro, A.; Schivo, A.; Salviato, A.; Palumbo, E.; Giussani, E.; Monne, I.; Terregino, C. |
| EPI2014987 | NS | Romania | 2022-Feb-01 | EPI_ISL_11922816 | A/swan/Romania/10324_22VIR2749-2/2022 | Institute for Diagnosis & Animal Health (IDAH) | Istituto Zooprofilattico Sperimentale delle Venezie | Barbuceanu, F.; Onita, I.; Neicut, A.; Motiu, R.; Burlacu, R.; Barbierato, G.; Zecchin, B.; Fusaro, A.; Schivo, A.; Salviato, A.; Palumbo, E.; Giussani, E.; Monne, I.; Terregino, C. |
| EPI2014988 | MP | Romania | 2022-Feb-01 | EPI_ISL_11922816 | A/swan/Romania/10324_22VIR2749-2/2022 | Institute for Diagnosis & Animal Health (IDAH) | Istituto Zooprofilattico Sperimentale delle Venezie | Barbuceanu, F.; Onita, I.; Neicut, A.; Motiu, R.; Burlacu, R.; Barbierato, G.; Zecchin, B.; Fusaro, A.; Schivo, A.; Salviato, A.; Palumbo, E.; Giussani, E.; Monne, I.; Terregino, C. |
| EPI2014994 | NP | Romania | 2022-Feb-03 | EPI_ISL_11922817 | A/swan/Romania/10394_22VIR2749-3/2022 | Institute for Diagnosis & Animal Health (IDAH) | Istituto Zooprofilattico Sperimentale delle Venezie | Barbuceanu, F.; Onita, I.; Neicut, A.; Motiu, R.; Burlacu, R.; Barbierato, G.; Zecchin, B.; Fusaro, A.; Schivo, A.; Salviato, A.; Palumbo, E.; Giussani, E.; Monne, I.; Terregino, C. |
| EPI2014995 | NS | Romania | 2022-Feb-03 | EPI_ISL_11922817 | A/swan/Romania/10394_22VIR2749-3/2022 | Institute for Diagnosis & Animal Health (IDAH) | Istituto Zooprofilattico Sperimentale delle Venezie | Barbuceanu, F.; Onita, I.; Neicut, A.; Motiu, R.; Burlacu, R.; Barbierato, G.; Zecchin, B.; Fusaro, A.; Schivo, A.; Salviato, A.; Palumbo, E.; Giussani, E.; Monne, I.; Terregino, C. |
| EPI2014996 | MP | Romania | 2022-Feb-03 | EPI_ISL_11922817 | A/swan/Romania/10394_22VIR2749-3/2022 | Institute for Diagnosis & Animal Health (IDAH) | Istituto Zooprofilattico Sperimentale delle Venezie | Barbuceanu, F.; Onita, I.; Neicut, A.; Motiu, R.; Burlacu, R.; Barbierato, G.; Zecchin, B.; Fusaro, A.; Schivo, A.; Salviato, A.; Palumbo, E.; Giussani, E.; Monne, I.; Terregino, C. |
| EPI2015002 | NP | Romania | 2022-Feb-09 | EPI_ISL_11922818 | A/swan/Romania/10455_22VIR2749-4/2022 | Institute for Diagnosis & Animal Health (IDAH) | Istituto Zooprofilattico Sperimentale delle Venezie | Barbuceanu, F.; Onita, I.; Neicut, A.; Motiu, R.; Burlacu, R.; Barbierato, G.; Zecchin, B.; Fusaro, A.; Schivo, A.; Salviato, A.; Palumbo, E.; Giussani, E.; Monne, I.; Terregino, C. |
| EPI2015003 | NS | Romania | 2022-Feb-09 | EPI_ISL_11922818 | A/swan/Romania/10455_22VIR2749-4/2022 | Institute for Diagnosis & Animal Health (IDAH) | Istituto Zooprofilattico Sperimentale delle Venezie | Barbuceanu, F.; Onita, I.; Neicut, A.; Motiu, R.; Burlacu, R.; Barbierato, G.; Zecchin, B.; Fusaro, A.; Schivo, A.; Salviato, A.; Palumbo, E.; Giussani, E.; Monne, I.; Terregino, C. |
| EPI2015004 | MP | Romania | 2022-Feb-09 | EPI_ISL_11922818 | A/swan/Romania/10455_22VIR2749-4/2022 | Institute for Diagnosis & Animal Health (IDAH) | Istituto Zooprofilattico Sperimentale delle Venezie | Barbuceanu, F.; Onita, I.; Neicut, A.; Motiu, R.; Burlacu, R.; Barbierato, G.; Zecchin, B.; Fusaro, A.; Schivo, A.; Salviato, A.; Palumbo, E.; Giussani, E.; Monne, I.; Terregino, C. |
| EPI2015010 | NP | Romania | 2022-Feb-10 | EPI_ISL_11922819 | A/laying_hen/Romania/10470_22VIR2749-5/2022 | Institute for Diagnosis & Animal Health (IDAH) | Istituto Zooprofilattico Sperimentale delle Venezie | Barbuceanu, F.; Onita, I.; Neicut, A.; Motiu, R.; Burlacu, R.; Barbierato, G.; Zecchin, B.; Fusaro, A.; Schivo, A.; Salviato, A.; Palumbo, E.; Giussani, E.; Monne, I.; Terregino, C. |
| EPI2015011 | NS | Romania | 2022-Feb-10 | EPI_ISL_11922819 | A/laying_hen/Romania/10470_22VIR2749-5/2022 | Institute for Diagnosis & Animal Health (IDAH) | Istituto Zooprofilattico Sperimentale delle Venezie | Barbuceanu, F.; Onita, I.; Neicut, A.; Motiu, R.; Burlacu, R.; Barbierato, G.; Zecchin, B.; Fusaro, A.; Schivo, A.; Salviato, A.; Palumbo, E.; Giussani, E.; Monne, I.; Terregino, C. |
| EPI2015012 | MP | Romania | 2022-Feb-10 | EPI_ISL_11922819 | A/laying_hen/Romania/10470_22VIR2749-5/2022 | Institute for Diagnosis & Animal Health (IDAH) | Istituto Zooprofilattico Sperimentale delle Venezie | Barbuceanu, F.; Onita, I.; Neicut, A.; Motiu, R.; Burlacu, R.; Barbierato, G.; Zecchin, B.; Fusaro, A.; Schivo, A.; Salviato, A.; Palumbo, E.; Giussani, E.; Monne, I.; Terregino, C. |
| EPI2015018 | NP | Romania | 2022-Feb-22 | EPI_ISL_11922820 | A/swan/Romania/10656_22VIR2749-6/2022 | Institute for Diagnosis & Animal Health (IDAH) | Istituto Zooprofilattico Sperimentale delle Venezie | Barbuceanu, F.; Onita, I.; Neicut, A.; Motiu, R.; Burlacu, R.; Barbierato, G.; Zecchin, B.; Fusaro, A.; Schivo, A.; Salviato, A.; Palumbo, E.; Giussani, E.; Monne, I.; Terregino, C. |
| EPI2015019 | NS | Romania | 2022-Feb-22 | EPI_ISL_11922820 | A/swan/Romania/10656_22VIR2749-6/2022 | Institute for Diagnosis & Animal Health (IDAH) | Istituto Zooprofilattico Sperimentale delle Venezie | Barbuceanu, F.; Onita, I.; Neicut, A.; Motiu, R.; Burlacu, R.; Barbierato, G.; Zecchin, B.; Fusaro, A.; Schivo, A.; Salviato, A.; Palumbo, E.; Giussani, E.; Monne, I.; Terregino, C. |
| EPI2015020 | MP | Romania | 2022-Feb-22 | EPI_ISL_11922820 | A/swan/Romania/10656_22VIR2749-6/2022 | Institute for Diagnosis & Animal Health (IDAH) | Istituto Zooprofilattico Sperimentale delle Venezie | Barbuceanu, F.; Onita, I.; Neicut, A.; Motiu, R.; Burlacu, R.; Barbierato, G.; Zecchin, B.; Fusaro, A.; Schivo, A.; Salviato, A.; Palumbo, E.; Giussani, E.; Monne, I.; Terregino, C. |
| EPI2015027 | NS | Romania | 2022-Feb-23 | EPI_ISL_11922821 | A/swan/Romania/10678_22VIR2749-7/2022 | Institute for Diagnosis & Animal Health (IDAH) | Istituto Zooprofilattico Sperimentale delle Venezie | Barbuceanu, F.; Onita, I.; Neicut, A.; Motiu, R.; Burlacu, R.; Barbierato, G.; Zecchin, B.; Fusaro, A.; Schivo, A.; Salviato, A.; Palumbo, E.; Giussani, E.; Monne, I.; Terregino, C. |
| EPI2015028 | MP | Romania | 2022-Feb-23 | EPI_ISL_11922821 | A/swan/Romania/10678_22VIR2749-7/2022 | Institute for Diagnosis & Animal Health (IDAH) | Istituto Zooprofilattico Sperimentale delle Venezie | Barbuceanu, F.; Onita, I.; Neicut, A.; Motiu, R.; Burlacu, R.; Barbierato, G.; Zecchin, B.; Fusaro, A.; Schivo, A.; Salviato, A.; Palumbo, E.; Giussani, E.; Monne, I.; Terregino, C. |
| EPI2015026 | NP | Romania | 2022-Feb-23 | EPI_ISL_11922821 | A/swan/Romania/10678_22VIR2749-7/2022 | Institute for Diagnosis & Animal Health (IDAH) | Istituto Zooprofilattico Sperimentale delle Venezie | Barbuceanu, F.; Onita, I.; Neicut, A.; Motiu, R.; Burlacu, R.; Barbierato, G.; Zecchin, B.; Fusaro, A.; Schivo, A.; Salviato, A.; Palumbo, E.; Giussani, E.; Monne, I.; Terregino, C. |
| EPI2015034 | NP | Romania | 2022-Mar-07 | EPI_ISL_11922822 | A/swan/Romania/10986_22VIR2749-8/2022 | Institute for Diagnosis & Animal Health (IDAH) | Istituto Zooprofilattico Sperimentale delle Venezie | Barbuceanu, F.; Onita, I.; Neicut, A.; Motiu, R.; Burlacu, R.; Barbierato, G.; Zecchin, B.; Fusaro, A.; Schivo, A.; Salviato, A.; Palumbo, E.; Giussani, E.; Monne, I.; Terregino, C. |
| EPI2015035 | NS | Romania | 2022-Mar-07 | EPI_ISL_11922822 | A/swan/Romania/10986_22VIR2749-8/2022 | Institute for Diagnosis & Animal Health (IDAH) | Istituto Zooprofilattico Sperimentale delle Venezie | Barbuceanu, F.; Onita, I.; Neicut, A.; Motiu, R.; Burlacu, R.; Barbierato, G.; Zecchin, B.; Fusaro, A.; Schivo, A.; Salviato, A.; Palumbo, E.; Giussani, E.; Monne, I.; Terregino, C. |
| EPI2015036 | MP | Romania | 2022-Mar-07 | EPI_ISL_11922822 | A/swan/Romania/10986_22VIR2749-8/2022 | Institute for Diagnosis & Animal Health (IDAH) | Istituto Zooprofilattico Sperimentale delle Venezie | Barbuceanu, F.; Onita, I.; Neicut, A.; Motiu, R.; Burlacu, R.; Barbierato, G.; Zecchin, B.; Fusaro, A.; Schivo, A.; Salviato, A.; Palumbo, E.; Giussani, E.; Monne, I.; Terregino, C. |
| EPI2018061 | NP | Netherlands | 2022-Jan-26 | EPI_ISL_12066188 | A/Fox/Netherlands/EMC1/2022 | Erasmus Medical Center | Erasmus Medical Center | R.A.M., Fouchier; S., Therwessen; O., Vuong; I., Chestakova; R., Sikkema; P., Lexmond; M., Pronk |
| EPI2018062 | NS | Netherlands | 2022-Jan-26 | EPI_ISL_12066188 | A/Fox/Netherlands/EMC1/2022 | Erasmus Medical Center | Erasmus Medical Center | R.A.M., Fouchier; S., Therwessen; O., Vuong; I., Chestakova; R., Sikkema; P., Lexmond; M., Pronk |
| EPI2018063 | MP | Netherlands | 2022-Jan-26 | EPI_ISL_12066188 | A/Fox/Netherlands/EMC1/2022 | Erasmus Medical Center | Erasmus Medical Center | R.A.M., Fouchier; S., Therwessen; O., Vuong; I., Chestakova; R., Sikkema; P., Lexmond; M., Pronk |
| EPI2018068 | NP | Netherlands | 2022-Jan-10 | EPI_ISL_12069288 | A/Fox/Netherlands/EMC2/2022 | Erasmus Medical Center | Erasmus Medical Center | R.A.M., Fouchier; S., Thewessen; O., Vuong; I., Chestakova; R., Sikkema; P., Lexmond; M., Pronk |
| EPI2018069 | NS | Netherlands | 2022-Jan-10 | EPI_ISL_12069288 | A/Fox/Netherlands/EMC2/2022 | Erasmus Medical Center | Erasmus Medical Center | R.A.M., Fouchier; S., Thewessen; O., Vuong; I., Chestakova; R., Sikkema; P., Lexmond; M., Pronk |
| EPI2018070 | MP | Netherlands | 2022-Jan-10 | EPI_ISL_12069288 | A/Fox/Netherlands/EMC2/2022 | Erasmus Medical Center | Erasmus Medical Center | R.A.M., Fouchier; S., Thewessen; O., Vuong; I., Chestakova; R., Sikkema; P., Lexmond; M., Pronk |
| EPI2018076 | NP | Netherlands | 2022-Jan-22 | EPI_ISL_12069289 | A/Fox/Netherlands/EMC3/2022 | Erasmus Medical Center | Erasmus Medical Center | R.A.M., Fouchier; S., Thewessen; O., Vuong; I., Chestakova; R., Sikkema; P., Lexmond; M., Pronk |

| | | | | | | | | |
|---|---|---|---|---|---|---|---|---|
| EPI2018077 | NS | Netherlands | 2022-Jan-22 | EPI_ISL_12069289 | A/Fox/Netherlands/EMC3/2022 | Erasmus Medical Center | Erasmus Medical Center | R.A.M., Fouchier; S., Thewessen; O., Vuong; I., Chestakova; R., Sikkema; P., Lexmond; M., Pronk |
| EPI2018078 | MP | Netherlands | 2022-Jan-22 | EPI_ISL_12069289 | A/Fox/Netherlands/EMC3/2022 | Erasmus Medical Center | Erasmus Medical Center | R.A.M., Fouchier; S., Thewessen; O., Vuong; I., Chestakova; R., Sikkema; P., Lexmond; M., Pronk |
| EPI2018092 | NP | Netherlands | 2022-Mar-28 | EPI_ISL_12069978 | A/greylag goose/Netherlands/22006190-002/2022 | Wageningen Bioveterinary Research | Wageningen Bioveterinary Research | Beerens, Nancy; Harders, Frank; Pritz-Verschuren, Sylvia; Roose, Marit; Venema, Sandra; Germeraad, Evelien; Engelsma, Marc; Heutink, Rene |
| EPI2018093 | NS | Netherlands | 2022-Mar-28 | EPI_ISL_12069978 | A/greylag goose/Netherlands/22006190-002/2022 | Wageningen Bioveterinary Research | Wageningen Bioveterinary Research | Beerens, Nancy; Harders, Frank; Pritz-Verschuren, Sylvia; Roose, Marit; Venema, Sandra; Germeraad, Evelien; Engelsma, Marc; Heutink, Rene |
| EPI2018094 | MP | Netherlands | 2022-Mar-28 | EPI_ISL_12069978 | A/greylag goose/Netherlands/22006190-002/2022 | Wageningen Bioveterinary Research | Wageningen Bioveterinary Research | Beerens, Nancy; Harders, Frank; Pritz-Verschuren, Sylvia; Roose, Marit; Venema, Sandra; Germeraad, Evelien; Engelsma, Marc; Heutink, Rene |
| EPI2018100 | NP | Netherlands | 2022-Mar-27 | EPI_ISL_12069979 | A/black-backed gull/Netherlands/22006192-001/2022 | Wageningen Bioveterinary Research | Wageningen Bioveterinary Research | Beerens, Nancy; Harders, Frank; Pritz-Verschuren, Sylvia; Roose, Marit; Venema, Sandra; Germeraad, Evelien; Engelsma, Marc; Heutink, Rene |
| EPI2018101 | NS | Netherlands | 2022-Mar-27 | EPI_ISL_12069979 | A/black-backed gull/Netherlands/22006192-001/2022 | Wageningen Bioveterinary Research | Wageningen Bioveterinary Research | Beerens, Nancy; Harders, Frank; Pritz-Verschuren, Sylvia; Roose, Marit; Venema, Sandra; Germeraad, Evelien; Engelsma, Marc; Heutink, Rene |
| EPI2018102 | MP | Netherlands | 2022-Mar-27 | EPI_ISL_12069979 | A/black-backed gull/Netherlands/22006192-001/2022 | Wageningen Bioveterinary Research | Wageningen Bioveterinary Research | Beerens, Nancy; Harders, Frank; Pritz-Verschuren, Sylvia; Roose, Marit; Venema, Sandra; Germeraad, Evelien; Engelsma, Marc; Heutink, Rene |
| EPI2022141 | NP | Netherlands | 2022-Apr-05 | EPI_ISL_12177629 | A/black-backed gull/Netherlands/22006711-001/2022 | Wageningen Bioveterinary Research | Wageningen Bioveterinary Research | Beerens, Nancy; Harders, Frank; Pritz-Verschuren, Sylvia; Roose, Marit; Venema, Sandra; Germeraad, Evelien; Engelsma, Marc; Heutink, Rene |
| EPI2022142 | NS | Netherlands | 2022-Apr-05 | EPI_ISL_12177629 | A/black-backed gull/Netherlands/22006711-001/2022 | Wageningen Bioveterinary Research | Wageningen Bioveterinary Research | Beerens, Nancy; Harders, Frank; Pritz-Verschuren, Sylvia; Roose, Marit; Venema, Sandra; Germeraad, Evelien; Engelsma, Marc; Heutink, Rene |
| EPI2022143 | MP | Netherlands | 2022-Apr-05 | EPI_ISL_12177629 | A/black-backed gull/Netherlands/22006711-001/2022 | Wageningen Bioveterinary Research | Wageningen Bioveterinary Research | Beerens, Nancy; Harders, Frank; Pritz-Verschuren, Sylvia; Roose, Marit; Venema, Sandra; Germeraad, Evelien; Engelsma, Marc; Heutink, Rene |
| EPI2022938 | NP | Belgium | 2022-Mar-19 | EPI_ISL_12210335 | A/Falco_peregrinus/Belgium/4055_0002/2022 | Sciensano - Animal Infectious Diseases | Sciensano, Department of Animal Infectious Diseases | Van Borm, Steven; Roupie, Virginie; Lambrecht, Benedicte; Mathijs, Elisabeth; Steensels, Mieke |
| EPI2022941 | NS | Belgium | 2022-Mar-19 | EPI_ISL_12210335 | A/Falco_peregrinus/Belgium/4055_0002/2022 | Sciensano - Animal Infectious Diseases | Sciensano, Department of Animal Infectious Diseases | Van Borm, Steven; Roupie, Virginie; Lambrecht, Benedicte; Mathijs, Elisabeth; Steensels, Mieke |
| EPI2022940 | MP | Belgium | 2022-Mar-19 | EPI_ISL_12210335 | A/Falco_peregrinus/Belgium/4055_0002/2022 | Sciensano - Animal Infectious Diseases | Sciensano, Department of Animal Infectious Diseases | Van Borm, Steven; Roupie, Virginie; Lambrecht, Benedicte; Mathijs, Elisabeth; Steensels, Mieke |
| EPI2023595 | NP | Netherlands | 2022-Mar-23 | EPI_ISL_12215416 | A/greylag goose/Netherlands/22005844-002/2022 | Wageningen Bioveterinary Research | Wageningen Bioveterinary Research | Beerens, Nancy; Harders, Frank; Pritz-Verschuren, Sylvia; Roose, Marit; Venema, Sandra; Germeraad, Evelien; Engelsma, Marc; Heutink, Rene |
| EPI2023596 | NS | Netherlands | 2022-Mar-23 | EPI_ISL_12215416 | A/greylag goose/Netherlands/22005844-002/2022 | Wageningen Bioveterinary Research | Wageningen Bioveterinary Research | Beerens, Nancy; Harders, Frank; Pritz-Verschuren, Sylvia; Roose, Marit; Venema, Sandra; Germeraad, Evelien; Engelsma, Marc; Heutink, Rene |
| EPI2023597 | MP | Netherlands | 2022-Mar-23 | EPI_ISL_12215416 | A/greylag goose/Netherlands/22005844-002/2022 | Wageningen Bioveterinary Research | Wageningen Bioveterinary Research | Beerens, Nancy; Harders, Frank; Pritz-Verschuren, Sylvia; Roose, Marit; Venema, Sandra; Germeraad, Evelien; Engelsma, Marc; Heutink, Rene |
| EPI2028762 | NP | Netherlands | 2022-Apr-17 | EPI_ISL_12474793 | A/barnacle goose/Netherlands/22007405-004/2022 | Wageningen Bioveterinary Research | Wageningen Bioveterinary Research | Beerens, Nancy; Harders, Frank; Pritz-Verschuren, Sylvia; Roose, Marit; Venema, Sandra; Germeraad, Evelien; Engelsma, Marc; Heutink, Rene |
| EPI2028763 | NS | Netherlands | 2022-Apr-17 | EPI_ISL_12474793 | A/barnacle goose/Netherlands/22007405-004/2022 | Wageningen Bioveterinary Research | Wageningen Bioveterinary Research | Beerens, Nancy; Harders, Frank; Pritz-Verschuren, Sylvia; Roose, Marit; Venema, Sandra; Germeraad, Evelien; Engelsma, Marc; Heutink, Rene |
| EPI2028764 | MP | Netherlands | 2022-Apr-17 | EPI_ISL_12474793 | A/barnacle goose/Netherlands/22007405-004/2022 | Wageningen Bioveterinary Research | Wageningen Bioveterinary Research | Beerens, Nancy; Harders, Frank; Pritz-Verschuren, Sylvia; Roose, Marit; Venema, Sandra; Germeraad, Evelien; Engelsma, Marc; Heutink, Rene |
| EPI2028770 | NP | Netherlands | 2022-Apr-11 | EPI_ISL_12474794 | A/greylag goose /Netherlands/22006859-001/2022 | Wageningen Bioveterinary Research | Wageningen Bioveterinary Research | Beerens, Nancy; Harders, Frank; Pritz-Verschuren, Sylvia; Roose, Marit; Venema, Sandra; Germeraad, Evelien; Engelsma, Marc; Heutink, Rene |
| EPI2028771 | NS | Netherlands | 2022-Apr-11 | EPI_ISL_12474794 | A/greylag goose /Netherlands/22006859-001/2022 | Wageningen Bioveterinary Research | Wageningen Bioveterinary Research | Beerens, Nancy; Harders, Frank; Pritz-Verschuren, Sylvia; Roose, Marit; Venema, Sandra; Germeraad, Evelien; Engelsma, Marc; Heutink, Rene |
| EPI2028772 | MP | Netherlands | 2022-Apr-11 | EPI_ISL_12474794 | A/greylag goose /Netherlands/22006859-001/2022 | Wageningen Bioveterinary Research | Wageningen Bioveterinary Research | Beerens, Nancy; Harders, Frank; Pritz-Verschuren, Sylvia; Roose, Marit; Venema, Sandra; Germeraad, Evelien; Engelsma, Marc; Heutink, Rene |
| EPI2028987 | NS | Netherlands | 2022-Apr-06 | EPI_ISL_12512879 | A/Black-headed gull/Netherlands/2/2022 | Erasmus Medical Center | Erasmus Medical Center | |
| EPI2028984 | MP | Netherlands | 2022-Apr-06 | EPI_ISL_12512879 | A/Black-headed gull/Netherlands/2/2022 | Erasmus Medical Center | Erasmus Medical Center | |
| EPI2028986 | NP | Netherlands | 2022-Apr-06 | EPI_ISL_12512879 | A/Black-headed gull/Netherlands/2/2022 | Erasmus Medical Center | Erasmus Medical Center | |
| EPI2029044 | NP | Netherlands | 2022-Apr-14 | EPI_ISL_12514425 | A/Black-headed gull/Netherlands/3/2022 | Erasmus Medical Center | Erasmus Medical Center | |
| EPI2029045 | NS | Netherlands | 2022-Apr-14 | EPI_ISL_12514425 | A/Black-headed gull/Netherlands/3/2022 | Erasmus Medical Center | Erasmus Medical Center | |
| EPI2029046 | MP | Netherlands | 2022-Apr-14 | EPI_ISL_12514425 | A/Black-headed gull/Netherlands/3/2022 | Erasmus Medical Center | Erasmus Medical Center | |
| EPI2029052 | NP | Netherlands | 2022-Apr-14 | EPI_ISL_12514442 | A/Black-headed gull/Netherlands/4/2022 | Erasmus Medical Center | Erasmus Medical Center | |
| EPI2029053 | NS | Netherlands | 2022-Apr-14 | EPI_ISL_12514442 | A/Black-headed gull/Netherlands/4/2022 | Erasmus Medical Center | Erasmus Medical Center | |
| EPI2029054 | MP | Netherlands | 2022-Apr-14 | EPI_ISL_12514442 | A/Black-headed gull/Netherlands/4/2022 | Erasmus Medical Center | Erasmus Medical Center | |
| EPI2029060 | NP | Netherlands | 2022-Apr-14 | EPI_ISL_12514483 | A/Caspian Gull/Netherlands/3/2022 | Erasmus Medical Center | Erasmus Medical Center | |
| EPI2029061 | NS | Netherlands | 2022-Apr-14 | EPI_ISL_12514483 | A/Caspian Gull/Netherlands/3/2022 | Erasmus Medical Center | Erasmus Medical Center | |
| EPI2029062 | MP | Netherlands | 2022-Apr-14 | EPI_ISL_12514483 | A/Caspian Gull/Netherlands/3/2022 | Erasmus Medical Center | Erasmus Medical Center | |

| | | | | | | | | |
|---|---|---|---|---|---|---|---|---|
| EPI2029068 | NP | Netherlands | 2022-Apr-11 | EPI_ISL_12514532 | A/Common raven/Netherlands/1/2022 | Erasmus Medical Center | Erasmus Medical Center | |
| EPI2029069 | NS | Netherlands | 2022-Apr-11 | EPI_ISL_12514532 | A/Common raven/Netherlands/1/2022 | Erasmus Medical Center | Erasmus Medical Center | |
| EPI2029070 | MP | Netherlands | 2022-Apr-11 | EPI_ISL_12514532 | A/Common raven/Netherlands/1/2022 | Erasmus Medical Center | Erasmus Medical Center | |
| EPI2029076 | NP | Netherlands | 2022-Apr-13 | EPI_ISL_12514573 | A/Song Thrush/Netherlands/1/2022 | Erasmus Medical Center | Erasmus Medical Center | |
| EPI2029077 | NS | Netherlands | 2022-Apr-13 | EPI_ISL_12514573 | A/Song Thrush/Netherlands/1/2022 | Erasmus Medical Center | Erasmus Medical Center | |
| EPI2029078 | MP | Netherlands | 2022-Apr-13 | EPI_ISL_12514573 | A/Song Thrush/Netherlands/1/2022 | Erasmus Medical Center | Erasmus Medical Center | |
| EPI2029084 | NP | Netherlands | 2022-Apr-14 | EPI_ISL_12514622 | A/Common raven/Netherlands/2/2022 | Erasmus Medical Center | Erasmus Medical Center | |
| EPI2029085 | NS | Netherlands | 2022-Apr-14 | EPI_ISL_12514622 | A/Common raven/Netherlands/2/2022 | Erasmus Medical Center | Erasmus Medical Center | |
| EPI2029086 | MP | Netherlands | 2022-Apr-14 | EPI_ISL_12514622 | A/Common raven/Netherlands/2/2022 | Erasmus Medical Center | Erasmus Medical Center | |
| EPI2029092 | NP | Netherlands | 2022-Apr-14 | EPI_ISL_12514681 | A/European Herring Gull/Netherlands/3/2022 | Erasmus Medical Center | Erasmus Medical Center | |
| EPI2029093 | NS | Netherlands | 2022-Apr-14 | EPI_ISL_12514681 | A/European Herring Gull/Netherlands/3/2022 | Erasmus Medical Center | Erasmus Medical Center | |
| EPI2029094 | MP | Netherlands | 2022-Apr-14 | EPI_ISL_12514681 | A/European Herring Gull/Netherlands/3/2022 | Erasmus Medical Center | Erasmus Medical Center | |
| EPI2050816 | MP | Luxembourg | 2022-Jan-17 | EPI_ISL_13026245 | A/Canada goose/Luxembourg/22012198/2022 | Laboratoire de Médecine vétérinaire de l'Etat | Luxembourg Institute of Health | Aurélie Sausy, Chantal Snoeck |
| EPI2050817 | NS | Luxembourg | 2022-Jan-17 | EPI_ISL_13026245 | A/Canada goose/Luxembourg/22012198/2022 | Laboratoire de Médecine vétérinaire de l'Etat | Luxembourg Institute of Health | Aurélie Sausy, Chantal Snoeck |
| EPI2050820 | NP | Luxembourg | 2022-Jan-17 | EPI_ISL_13026245 | A/Canada goose/Luxembourg/22012198/2022 | Laboratoire de Médecine vétérinaire de l'Etat | Luxembourg Institute of Health | Aurélie Sausy, Chantal Snoeck |
| EPI2050824 | MP | Luxembourg | 2022-Feb-04 | EPI_ISL_13026246 | A/Barnacle goose/Luxembourg/22033922/2022 | Laboratoire de Médecine vétérinaire de l'Etat | Luxembourg Institute of Health | Aurélie Sausy, Chantal Snoeck |
| EPI2050828 | NP | Luxembourg | 2022-Feb-04 | EPI_ISL_13026246 | A/Barnacle goose/Luxembourg/22033922/2022 | Laboratoire de Médecine vétérinaire de l'Etat | Luxembourg Institute of Health | Aurélie Sausy, Chantal Snoeck |
| EPI2050825 | NS | Luxembourg | 2022-Feb-04 | EPI_ISL_13026246 | A/Barnacle goose/Luxembourg/22033922/2022 | Laboratoire de Médecine vétérinaire de l'Etat | Luxembourg Institute of Health | Aurélie Sausy, Chantal Snoeck |
| EPI2061036 | NP | Netherlands | 2022-Apr-29 | EPI_ISL_13201050 | A/Lesser Black-backed Gull/Netherlands/1/2022 | Erasmus Medical Center | Erasmus Medical Center | |
| EPI2061037 | NS | Netherlands | 2022-Apr-29 | EPI_ISL_13201050 | A/Lesser Black-backed Gull/Netherlands/1/2022 | Erasmus Medical Center | Erasmus Medical Center | |
| EPI2061038 | MP | Netherlands | 2022-Apr-29 | EPI_ISL_13201050 | A/Lesser Black-backed Gull/Netherlands/1/2022 | Erasmus Medical Center | Erasmus Medical Center | |
| EPI2061044 | NP | Netherlands | 2022-Mar-05 | EPI_ISL_13201074 | A/European polecat/Netherlands/1/2022 | Erasmus Medical Center | Erasmus Medical Center | |
| EPI2061045 | NS | Netherlands | 2022-Mar-05 | EPI_ISL_13201074 | A/European polecat/Netherlands/1/2022 | Erasmus Medical Center | Erasmus Medical Center | |
| EPI2061046 | MP | Netherlands | 2022-Mar-05 | EPI_ISL_13201074 | A/European polecat/Netherlands/1/2022 | Erasmus Medical Center | Erasmus Medical Center | |
| EPI2062302 | MP | Iceland | 2022-Apr-14 | EPI_ISL_13246267 | A/chicken/Iceland/2022AI02564/2022 | University of Iceland, Keldur | Friedrich-Loeffler-Institut | |
| EPI2062304 | NP | Iceland | 2022-Apr-14 | EPI_ISL_13246267 | A/chicken/Iceland/2022AI02564/2022 | University of Iceland, Keldur | Friedrich-Loeffler-Institut | |
| EPI2062305 | NS | Iceland | 2022-Apr-14 | EPI_ISL_13246267 | A/chicken/Iceland/2022AI02564/2022 | University of Iceland, Keldur | Friedrich-Loeffler-Institut | |
| EPI2062333 | NP | Iceland | 2022-Apr-14 | EPI_ISL_13246657 | A/chicken/Iceland/2022AI02565/2022 | University of Iceland, Keldur | Friedrich-Loeffler-Institut | |
| EPI2062331 | MP | Iceland | 2022-Apr-14 | EPI_ISL_13246657 | A/chicken/Iceland/2022AI02565/2022 | University of Iceland, Keldur | Friedrich-Loeffler-Institut | |
| EPI2062334 | NS | Iceland | 2022-Apr-14 | EPI_ISL_13246657 | A/chicken/Iceland/2022AI02565/2022 | University of Iceland, Keldur | Friedrich-Loeffler-Institut | |
| EPI2068304 | NP | Netherlands | 2022-May-26 | EPI_ISL_13300319 | A/brant goose/Netherlands/22009881-002/2022 | Wageningen Bioveterinary Research | Wageningen Bioveterinary Research | Beerens, Nancy; Harders, Frank; Pritz-Verschuren, Sylvia; Roose, Marit; Venema, Sandra; Germeraad, Evelien; Engelsma, Marc; Heutink, Rene |
| EPI2068305 | NS | Netherlands | 2022-May-26 | EPI_ISL_13300319 | A/brant goose/Netherlands/22009881-002/2022 | Wageningen Bioveterinary Research | Wageningen Bioveterinary Research | Beerens, Nancy; Harders, Frank; Pritz-Verschuren, Sylvia; Roose, Marit; Venema, Sandra; Germeraad, Evelien; Engelsma, Marc; Heutink, Rene |
| EPI2068306 | MP | Netherlands | 2022-May-26 | EPI_ISL_13300319 | A/brant goose/Netherlands/22009881-002/2022 | Wageningen Bioveterinary Research | Wageningen Bioveterinary Research | Beerens, Nancy; Harders, Frank; Pritz-Verschuren, Sylvia; Roose, Marit; Venema, Sandra; Germeraad, Evelien; Engelsma, Marc; Heutink, Rene |
| EPI2068336 | NP | Netherlands | 2022-May-27 | EPI_ISL_13300324 | A/white stork/Netherlands/22009973-002/2022 | Wageningen Bioveterinary Research | Wageningen Bioveterinary Research | Beerens, Nancy; Harders, Frank; Pritz-Verschuren, Sylvia; Roose, Marit; Venema, Sandra; Germeraad, Evelien; Engelsma, Marc; Heutink, Rene |
| EPI2068337 | NS | Netherlands | 2022-May-27 | EPI_ISL_13300324 | A/white stork/Netherlands/22009973-002/2022 | Wageningen Bioveterinary Research | Wageningen Bioveterinary Research | Beerens, Nancy; Harders, Frank; Pritz-Verschuren, Sylvia; Roose, Marit; Venema, Sandra; Germeraad, Evelien; Engelsma, Marc; Heutink, Rene |

| | | | | | | | | |
|---|---|---|---|---|---|---|---|---|
| EPI2068338 | MP | Netherlands | 2022-May-27 | EPI_ISL_13300324 | A/white stork/Netherlands/22009973-002/2022 | Wageningen Bioveterinary Research | Wageningen Bioveterinary Research | Beerens, Nancy; Harders, Frank; Pritz-Verschuren, Sylvia; Roose, Marit; Venema, Sandra; Germeraad, Evelien; Engelsma, Marc; Heutink, Rene |
| EPI2069017 | MP | Sweden | 2022-May-25 | EPI_ISL_13338081 | A/Northern gannet/Sweden/SVA220525SZ0402/FB001671/O-2022 | National Veterinary Institute, SVA | National Veterinary Institute | |
| EPI2069015 | NP | Sweden | 2022-May-25 | EPI_ISL_13338081 | A/Northern gannet/Sweden/SVA220525SZ0402/FB001671/O-2022 | National Veterinary Institute, SVA | National Veterinary Institute | |
| EPI2069018 | NS | Sweden | 2022-May-25 | EPI_ISL_13338081 | A/Northern gannet/Sweden/SVA220525SZ0402/FB001671/O-2022 | National Veterinary Institute, SVA | National Veterinary Institute | |
| EPI2071969 | NP | United Kingdom | 2022-Jan-02 | EPI_ISL_13370631 | A/domestic_duck/England/000357/2022 | Animal and Plant Health Agency (APHA) | Animal and Plant Health Agency (APHA) | |
| EPI2071970 | NS | United Kingdom | 2022-Jan-02 | EPI_ISL_13370631 | A/domestic_duck/England/000357/2022 | Animal and Plant Health Agency (APHA) | Animal and Plant Health Agency (APHA) | |
| EPI2071971 | MP | United Kingdom | 2022-Jan-02 | EPI_ISL_13370631 | A/domestic_duck/England/000357/2022 | Animal and Plant Health Agency (APHA) | Animal and Plant Health Agency (APHA) | |
| EPI2071977 | NP | United Kingdom | 2022-Jan-02 | EPI_ISL_13370632 | A/chicken/England/000095/2022 | Animal and Plant Health Agency (APHA) | Animal and Plant Health Agency (APHA) | |
| EPI2071978 | NS | United Kingdom | 2022-Jan-02 | EPI_ISL_13370632 | A/chicken/England/000095/2022 | Animal and Plant Health Agency (APHA) | Animal and Plant Health Agency (APHA) | |
| EPI2071979 | MP | United Kingdom | 2022-Jan-02 | EPI_ISL_13370632 | A/chicken/England/000095/2022 | Animal and Plant Health Agency (APHA) | Animal and Plant Health Agency (APHA) | |
| EPI2071985 | NP | United Kingdom | 2022-Jan-04 | EPI_ISL_13370633 | A/turkey/England/000733/2022 | Animal and Plant Health Agency (APHA) | Animal and Plant Health Agency (APHA) | |
| EPI2071986 | NS | United Kingdom | 2022-Jan-04 | EPI_ISL_13370633 | A/turkey/England/000733/2022 | Animal and Plant Health Agency (APHA) | Animal and Plant Health Agency (APHA) | |
| EPI2071987 | MP | United Kingdom | 2022-Jan-04 | EPI_ISL_13370633 | A/turkey/England/000733/2022 | Animal and Plant Health Agency (APHA) | Animal and Plant Health Agency (APHA) | |
| EPI2071993 | NP | United Kingdom | 2022-Jan-08 | EPI_ISL_13370634 | A/chicken/England/002376/2022 | Animal and Plant Health Agency (APHA) | Animal and Plant Health Agency (APHA) | |
| EPI2071994 | NS | United Kingdom | 2022-Jan-08 | EPI_ISL_13370634 | A/chicken/England/002376/2022 | Animal and Plant Health Agency (APHA) | Animal and Plant Health Agency (APHA) | |
| EPI2071995 | MP | United Kingdom | 2022-Jan-08 | EPI_ISL_13370634 | A/chicken/England/002376/2022 | Animal and Plant Health Agency (APHA) | Animal and Plant Health Agency (APHA) | |
| EPI2072001 | NP | United Kingdom | 2022-Jan-08 | EPI_ISL_13370679 | A/common_gull/England/245481/2022 | Animal and Plant Health Agency (APHA) | Animal and Plant Health Agency (APHA) | |
| EPI2072002 | NS | United Kingdom | 2022-Jan-08 | EPI_ISL_13370679 | A/common_gull/England/245481/2022 | Animal and Plant Health Agency (APHA) | Animal and Plant Health Agency (APHA) | |
| EPI2072003 | MP | United Kingdom | 2022-Jan-08 | EPI_ISL_13370679 | A/common_gull/England/245481/2022 | Animal and Plant Health Agency (APHA) | Animal and Plant Health Agency (APHA) | |
| EPI2072009 | NP | United Kingdom | 2022-Jan-12 | EPI_ISL_13370692 | A/turkey/England/004667/2022 | Animal and Plant Health Agency (APHA) | Animal and Plant Health Agency (APHA) | |
| EPI2072010 | NS | United Kingdom | 2022-Jan-12 | EPI_ISL_13370692 | A/turkey/England/004667/2022 | Animal and Plant Health Agency (APHA) | Animal and Plant Health Agency (APHA) | |
| EPI2072011 | MP | United Kingdom | 2022-Jan-12 | EPI_ISL_13370692 | A/turkey/England/004667/2022 | Animal and Plant Health Agency (APHA) | Animal and Plant Health Agency (APHA) | |
| EPI2072017 | NP | Isle of Man | 2022-Jan-14 | EPI_ISL_13370693 | A/Greylag_goose/Isle_of_Man/010298/2022 | Animal and Plant Health Agency (APHA) | Animal and Plant Health Agency (APHA) | |
| EPI2072018 | NS | Isle of Man | 2022-Jan-14 | EPI_ISL_13370693 | A/Greylag_goose/Isle_of_Man/010298/2022 | Animal and Plant Health Agency (APHA) | Animal and Plant Health Agency (APHA) | |
| EPI2072019 | MP | Isle of Man | 2022-Jan-14 | EPI_ISL_13370693 | A/Greylag_goose/Isle_of_Man/010298/2022 | Animal and Plant Health Agency (APHA) | Animal and Plant Health Agency (APHA) | |
| EPI2072025 | NP | United Kingdom | 2022-Jan-19 | EPI_ISL_13370694 | A/Domestic_goose/England/006847/2022 | Animal and Plant Health Agency (APHA) | Animal and Plant Health Agency (APHA) | |
| EPI2072026 | NS | United Kingdom | 2022-Jan-19 | EPI_ISL_13370694 | A/Domestic_goose/England/006847/2022 | Animal and Plant Health Agency (APHA) | Animal and Plant Health Agency (APHA) | |
| EPI2072027 | MP | United Kingdom | 2022-Jan-19 | EPI_ISL_13370694 | A/Domestic_goose/England/006847/2022 | Animal and Plant Health Agency (APHA) | Animal and Plant Health Agency (APHA) | |
| EPI2072033 | NP | United Kingdom | 2022-Jan-20 | EPI_ISL_13370695 | A/turkey/England/006963/2022 | Animal and Plant Health Agency (APHA) | Animal and Plant Health Agency (APHA) | |
| EPI2072034 | NS | United Kingdom | 2022-Jan-20 | EPI_ISL_13370695 | A/turkey/England/006963/2022 | Animal and Plant Health Agency (APHA) | Animal and Plant Health Agency (APHA) | |
| EPI2072035 | MP | United Kingdom | 2022-Jan-20 | EPI_ISL_13370695 | A/turkey/England/006963/2022 | Animal and Plant Health Agency (APHA) | Animal and Plant Health Agency (APHA) | |
| EPI2072041 | NP | United Kingdom | 2022-Jan-23 | EPI_ISL_13370696 | A/turkey/England/007586/2022 | Animal and Plant Health Agency (APHA) | Animal and Plant Health Agency (APHA) | |
| EPI2072042 | NS | United Kingdom | 2022-Jan-23 | EPI_ISL_13370696 | A/turkey/England/007586/2022 | Animal and Plant Health Agency (APHA) | Animal and Plant Health Agency (APHA) | |
| EPI2072043 | MP | United Kingdom | 2022-Jan-23 | EPI_ISL_13370696 | A/turkey/England/007586/2022 | Animal and Plant Health Agency (APHA) | Animal and Plant Health Agency (APHA) | |
| EPI2072049 | NP | United Kingdom | 2022-Jan-23 | EPI_ISL_13370697 | A/chicken/England/007584/2022 | Animal and Plant Health Agency (APHA) | Animal and Plant Health Agency (APHA) | |

| | | | | | | | |
|---|---|---|---|---|---|---|---|
| EPI2072050 | NS | United Kingdom | 2022-Jan-23 | EPI_ISL_13370697 | A/chicken/England/007584/2022 | Animal and Plant Health Agency (APHA) | Animal and Plant Health Agency (APHA) |
| EPI2072051 | MP | United Kingdom | 2022-Jan-23 | EPI_ISL_13370697 | A/chicken/England/007584/2022 | Animal and Plant Health Agency (APHA) | Animal and Plant Health Agency (APHA) |
| EPI2072058 | NS | United Kingdom | 2022-Jan-26 | EPI_ISL_13370698 | A/domestic_goose/England/008781/2022 | Animal and Plant Health Agency (APHA) | Animal and Plant Health Agency (APHA) |
| EPI2072059 | MP | United Kingdom | 2022-Jan-26 | EPI_ISL_13370698 | A/domestic_goose/England/008781/2022 | Animal and Plant Health Agency (APHA) | Animal and Plant Health Agency (APHA) |
| EPI2072057 | NP | United Kingdom | 2022-Jan-26 | EPI_ISL_13370698 | A/domestic_goose/England/008781/2022 | Animal and Plant Health Agency (APHA) | Animal and Plant Health Agency (APHA) |
| EPI2072065 | NP | United Kingdom | 2022-Jan-26 | EPI_ISL_13370699 | A/chicken/England/008775/2022 | Animal and Plant Health Agency (APHA) | Animal and Plant Health Agency (APHA) |
| EPI2072066 | NS | United Kingdom | 2022-Jan-26 | EPI_ISL_13370699 | A/chicken/England/008775/2022 | Animal and Plant Health Agency (APHA) | Animal and Plant Health Agency (APHA) |
| EPI2072067 | MP | United Kingdom | 2022-Jan-26 | EPI_ISL_13370699 | A/chicken/England/008775/2022 | Animal and Plant Health Agency (APHA) | Animal and Plant Health Agency (APHA) |
| EPI2072073 | NP | United Kingdom | 2022-Jan-27 | EPI_ISL_13370700 | A/turkey/England/009687/2022 | Animal and Plant Health Agency (APHA) | Animal and Plant Health Agency (APHA) |
| EPI2072074 | NS | United Kingdom | 2022-Jan-27 | EPI_ISL_13370700 | A/turkey/England/009687/2022 | Animal and Plant Health Agency (APHA) | Animal and Plant Health Agency (APHA) |
| EPI2072075 | MP | United Kingdom | 2022-Jan-27 | EPI_ISL_13370700 | A/turkey/England/009687/2022 | Animal and Plant Health Agency (APHA) | Animal and Plant Health Agency (APHA) |
| EPI2072081 | NP | United Kingdom | 2022-Jan-27 | EPI_ISL_13370701 | A/chicken/England/009811/2022 | Animal and Plant Health Agency (APHA) | Animal and Plant Health Agency (APHA) |
| EPI2072082 | NS | United Kingdom | 2022-Jan-27 | EPI_ISL_13370701 | A/chicken/England/009811/2022 | Animal and Plant Health Agency (APHA) | Animal and Plant Health Agency (APHA) |
| EPI2072083 | MP | United Kingdom | 2022-Jan-27 | EPI_ISL_13370701 | A/chicken/England/009811/2022 | Animal and Plant Health Agency (APHA) | Animal and Plant Health Agency (APHA) |
| EPI2072089 | NP | United Kingdom | 2022-Jan-27 | EPI_ISL_13370702 | A/mallard_duck/England/388009/2022 | Animal and Plant Health Agency (APHA) | Animal and Plant Health Agency (APHA) |
| EPI2072090 | NS | United Kingdom | 2022-Jan-27 | EPI_ISL_13370702 | A/mallard_duck/England/388009/2022 | Animal and Plant Health Agency (APHA) | Animal and Plant Health Agency (APHA) |
| EPI2072091 | MP | United Kingdom | 2022-Jan-27 | EPI_ISL_13370702 | A/mallard_duck/England/388009/2022 | Animal and Plant Health Agency (APHA) | Animal and Plant Health Agency (APHA) |
| EPI2072097 | NP | United Kingdom | 2022-Feb-03 | EPI_ISL_13370703 | A/domestic_duck/England/012247/2022 | Animal and Plant Health Agency (APHA) | Animal and Plant Health Agency (APHA) |
| EPI2072098 | NS | United Kingdom | 2022-Feb-03 | EPI_ISL_13370703 | A/domestic_duck/England/012247/2022 | Animal and Plant Health Agency (APHA) | Animal and Plant Health Agency (APHA) |
| EPI2072099 | MP | United Kingdom | 2022-Feb-03 | EPI_ISL_13370703 | A/domestic_duck/England/012247/2022 | Animal and Plant Health Agency (APHA) | Animal and Plant Health Agency (APHA) |
| EPI2072105 | NP | United Kingdom | 2022-Feb-12 | EPI_ISL_13370704 | A/chicken/England/014330/2022 | Animal and Plant Health Agency (APHA) | Animal and Plant Health Agency (APHA) |
| EPI2072106 | NS | United Kingdom | 2022-Feb-12 | EPI_ISL_13370704 | A/chicken/England/014330/2022 | Animal and Plant Health Agency (APHA) | Animal and Plant Health Agency (APHA) |
| EPI2072107 | MP | United Kingdom | 2022-Feb-12 | EPI_ISL_13370704 | A/chicken/England/014330/2022 | Animal and Plant Health Agency (APHA) | Animal and Plant Health Agency (APHA) |
| EPI2072113 | NP | United Kingdom | 2022-Feb-20 | EPI_ISL_13370705 | A/pheasant/Wales/016441/2022 | Animal and Plant Health Agency (APHA) | Animal and Plant Health Agency (APHA) |
| EPI2072114 | NS | United Kingdom | 2022-Feb-20 | EPI_ISL_13370705 | A/pheasant/Wales/016441/2022 | Animal and Plant Health Agency (APHA) | Animal and Plant Health Agency (APHA) |
| EPI2072115 | MP | United Kingdom | 2022-Feb-20 | EPI_ISL_13370705 | A/pheasant/Wales/016441/2022 | Animal and Plant Health Agency (APHA) | Animal and Plant Health Agency (APHA) |
| EPI2072129 | NP | United Kingdom | 2022-Feb-21 | EPI_ISL_13370707 | A/domestic_duck/England/017166/2022 | Animal and Plant Health Agency (APHA) | Animal and Plant Health Agency (APHA) |
| EPI2072130 | NS | United Kingdom | 2022-Feb-21 | EPI_ISL_13370707 | A/domestic_duck/England/017166/2022 | Animal and Plant Health Agency (APHA) | Animal and Plant Health Agency (APHA) |
| EPI2072131 | MP | United Kingdom | 2022-Feb-21 | EPI_ISL_13370707 | A/domestic_duck/England/017166/2022 | Animal and Plant Health Agency (APHA) | Animal and Plant Health Agency (APHA) |
| EPI2072137 | NP | United Kingdom | 2022-Feb-25 | EPI_ISL_13370708 | A/domestic_goose/England/019349/2022 | Animal and Plant Health Agency (APHA) | Animal and Plant Health Agency (APHA) |
| EPI2072138 | NS | United Kingdom | 2022-Feb-25 | EPI_ISL_13370708 | A/domestic_goose/England/019349/2022 | Animal and Plant Health Agency (APHA) | Animal and Plant Health Agency (APHA) |
| EPI2072139 | MP | United Kingdom | 2022-Feb-25 | EPI_ISL_13370708 | A/domestic_goose/England/019349/2022 | Animal and Plant Health Agency (APHA) | Animal and Plant Health Agency (APHA) |
| EPI2072145 | NP | United Kingdom | 2022-Feb-26 | EPI_ISL_13370709 | A/pheasant/England/019746/2022 | Animal and Plant Health Agency (APHA) | Animal and Plant Health Agency (APHA) |
| EPI2072146 | NS | United Kingdom | 2022-Feb-26 | EPI_ISL_13370709 | A/pheasant/England/019746/2022 | Animal and Plant Health Agency (APHA) | Animal and Plant Health Agency (APHA) |
| EPI2072147 | MP | United Kingdom | 2022-Feb-26 | EPI_ISL_13370709 | A/pheasant/England/019746/2022 | Animal and Plant Health Agency (APHA) | Animal and Plant Health Agency (APHA) |

| | | | | | | | |
|---|---|---|---|---|---|---|---|
| EPI2072153 | NP | United Kingdom | 2022-Feb-28 | EPI_ISL_13370710 | A/domestic_duck/England/019894/2022 | Animal and Plant Health Agency (APHA) | Animal and Plant Health Agency (APHA) |
| EPI2072154 | NS | United Kingdom | 2022-Feb-28 | EPI_ISL_13370710 | A/domestic_duck/England/019894/2022 | Animal and Plant Health Agency (APHA) | Animal and Plant Health Agency (APHA) |
| EPI2072155 | MP | United Kingdom | 2022-Feb-28 | EPI_ISL_13370710 | A/domestic_duck/England/019894/2022 | Animal and Plant Health Agency (APHA) | Animal and Plant Health Agency (APHA) |
| EPI2072177 | NP | United Kingdom | 2022-Jan-28 | EPI_ISL_13370894 | A/coot/England/388108/2022 | Animal and Plant Health Agency (APHA) | Animal and Plant Health Agency (APHA) |
| EPI2072178 | NS | United Kingdom | 2022-Jan-28 | EPI_ISL_13370894 | A/coot/England/388108/2022 | Animal and Plant Health Agency (APHA) | Animal and Plant Health Agency (APHA) |
| EPI2072179 | MP | United Kingdom | 2022-Jan-28 | EPI_ISL_13370894 | A/coot/England/388108/2022 | Animal and Plant Health Agency (APHA) | Animal and Plant Health Agency (APHA) |
| EPI2072185 | NP | United Kingdom | 2022-Feb-05 | EPI_ISL_13370895 | A/magpie/Scotland/012805/2022 | Animal and Plant Health Agency (APHA) | Animal and Plant Health Agency (APHA) |
| EPI2072186 | NS | United Kingdom | 2022-Feb-05 | EPI_ISL_13370895 | A/magpie/Scotland/012805/2022 | Animal and Plant Health Agency (APHA) | Animal and Plant Health Agency (APHA) |
| EPI2072187 | MP | United Kingdom | 2022-Feb-05 | EPI_ISL_13370895 | A/magpie/Scotland/012805/2022 | Animal and Plant Health Agency (APHA) | Animal and Plant Health Agency (APHA) |
| EPI2072193 | NP | Jersey | 2022-Feb-22 | EPI_ISL_13370896 | A/Red-breasted_goose/Jersey/018413/2022 | Animal and Plant Health Agency (APHA) | Animal and Plant Health Agency (APHA) |
| EPI2072194 | NS | Jersey | 2022-Feb-22 | EPI_ISL_13370896 | A/Red-breasted_goose/Jersey/018413/2022 | Animal and Plant Health Agency (APHA) | Animal and Plant Health Agency (APHA) |
| EPI2072195 | MP | Jersey | 2022-Feb-22 | EPI_ISL_13370896 | A/Red-breasted_goose/Jersey/018413/2022 | Animal and Plant Health Agency (APHA) | Animal and Plant Health Agency (APHA) |
| EPI2072201 | NP | Jersey | 2022-Feb-22 | EPI_ISL_13370897 | A/Red-breasted_goose/Jersey/018414/2022 | Animal and Plant Health Agency (APHA) | Animal and Plant Health Agency (APHA) |
| EPI2072202 | NS | Jersey | 2022-Feb-22 | EPI_ISL_13370897 | A/Red-breasted_goose/Jersey/018414/2022 | Animal and Plant Health Agency (APHA) | Animal and Plant Health Agency (APHA) |
| EPI2072203 | MP | Jersey | 2022-Feb-22 | EPI_ISL_13370897 | A/Red-breasted_goose/Jersey/018414/2022 | Animal and Plant Health Agency (APHA) | Animal and Plant Health Agency (APHA) |
| EPI2072209 | NP | United Kingdom | 2022-Mar-07 | EPI_ISL_13370907 | A/mute_swan/Scotland/029918/2022 | Animal and Plant Health Agency (APHA) | Animal and Plant Health Agency (APHA) |
| EPI2072210 | NS | United Kingdom | 2022-Mar-07 | EPI_ISL_13370907 | A/mute_swan/Scotland/029918/2022 | Animal and Plant Health Agency (APHA) | Animal and Plant Health Agency (APHA) |
| EPI2072211 | MP | United Kingdom | 2022-Mar-07 | EPI_ISL_13370907 | A/mute_swan/Scotland/029918/2022 | Animal and Plant Health Agency (APHA) | Animal and Plant Health Agency (APHA) |
| EPI2072217 | NP | United Kingdom | 2022-Mar-07 | EPI_ISL_13370908 | A/mute_swan/Scotland/029919/2022 | Animal and Plant Health Agency (APHA) | Animal and Plant Health Agency (APHA) |
| EPI2072218 | NS | United Kingdom | 2022-Mar-07 | EPI_ISL_13370908 | A/mute_swan/Scotland/029919/2022 | Animal and Plant Health Agency (APHA) | Animal and Plant Health Agency (APHA) |
| EPI2072219 | MP | United Kingdom | 2022-Mar-07 | EPI_ISL_13370908 | A/mute_swan/Scotland/029919/2022 | Animal and Plant Health Agency (APHA) | Animal and Plant Health Agency (APHA) |
| EPI2072225 | NP | United Kingdom | 2022-Mar-09 | EPI_ISL_13370909 | A/chicken/Scotland/024309/2022 | Animal and Plant Health Agency (APHA) | Animal and Plant Health Agency (APHA) |
| EPI2072226 | NS | United Kingdom | 2022-Mar-09 | EPI_ISL_13370909 | A/chicken/Scotland/024309/2022 | Animal and Plant Health Agency (APHA) | Animal and Plant Health Agency (APHA) |
| EPI2072227 | MP | United Kingdom | 2022-Mar-09 | EPI_ISL_13370909 | A/chicken/Scotland/024309/2022 | Animal and Plant Health Agency (APHA) | Animal and Plant Health Agency (APHA) |
| EPI2072233 | NP | United Kingdom | 2022-Mar-09 | EPI_ISL_13370910 | A/domestic_duck/Scotland/024305/2022 | Animal and Plant Health Agency (APHA) | Animal and Plant Health Agency (APHA) |
| EPI2072234 | NS | United Kingdom | 2022-Mar-09 | EPI_ISL_13370910 | A/domestic_duck/Scotland/024305/2022 | Animal and Plant Health Agency (APHA) | Animal and Plant Health Agency (APHA) |
| EPI2072235 | MP | United Kingdom | 2022-Mar-09 | EPI_ISL_13370910 | A/domestic_duck/Scotland/024305/2022 | Animal and Plant Health Agency (APHA) | Animal and Plant Health Agency (APHA) |
| EPI2072242 | NS | United Kingdom | 2022-Mar-10 | EPI_ISL_13370911 | A/domestic_duck/England/025226/2022 | Animal and Plant Health Agency (APHA) | Animal and Plant Health Agency (APHA) |
| EPI2072243 | MP | United Kingdom | 2022-Mar-10 | EPI_ISL_13370911 | A/domestic_duck/England/025226/2022 | Animal and Plant Health Agency (APHA) | Animal and Plant Health Agency (APHA) |
| EPI2072241 | NP | United Kingdom | 2022-Mar-10 | EPI_ISL_13370911 | A/domestic_duck/England/025226/2022 | Animal and Plant Health Agency (APHA) | Animal and Plant Health Agency (APHA) |
| EPI2072249 | NP | United Kingdom | 2022-Mar-11 | EPI_ISL_13370912 | A/domestic_duck/England/025639/2022 | Animal and Plant Health Agency (APHA) | Animal and Plant Health Agency (APHA) |
| EPI2072250 | NS | United Kingdom | 2022-Mar-11 | EPI_ISL_13370912 | A/domestic_duck/England/025639/2022 | Animal and Plant Health Agency (APHA) | Animal and Plant Health Agency (APHA) |
| EPI2072251 | MP | United Kingdom | 2022-Mar-11 | EPI_ISL_13370912 | A/domestic_duck/England/025639/2022 | Animal and Plant Health Agency (APHA) | Animal and Plant Health Agency (APHA) |
| EPI2072257 | NP | United Kingdom | 2022-Mar-18 | EPI_ISL_13370913 | A/chicken/Scotland/029509/2022 | Animal and Plant Health Agency (APHA) | Animal and Plant Health Agency (APHA) |
| EPI2072258 | NS | United Kingdom | 2022-Mar-18 | EPI_ISL_13370913 | A/chicken/Scotland/029509/2022 | Animal and Plant Health Agency (APHA) | Animal and Plant Health Agency (APHA) |

| | | | | | | | |
|---|---|---|---|---|---|---|---|
| EPI2072259 | MP | United Kingdom | 2022-Mar-18 | EPI_ISL_13370913 | A/chicken/Scotland/029509/2022 | Animal and Plant Health Agency (APHA) | Animal and Plant Health Agency (APHA) |
| EPI2072265 | NP | United Kingdom | 2022-Mar-20 | EPI_ISL_13370914 | A/domestic_duck/England/029617/2022 | Animal and Plant Health Agency (APHA) | Animal and Plant Health Agency (APHA) |
| EPI2072266 | NS | United Kingdom | 2022-Mar-20 | EPI_ISL_13370914 | A/domestic_duck/England/029617/2022 | Animal and Plant Health Agency (APHA) | Animal and Plant Health Agency (APHA) |
| EPI2072267 | MP | United Kingdom | 2022-Mar-20 | EPI_ISL_13370914 | A/domestic_duck/England/029617/2022 | Animal and Plant Health Agency (APHA) | Animal and Plant Health Agency (APHA) |
| EPI2072273 | NP | United Kingdom | 2022-Mar-26 | EPI_ISL_13370915 | A/domestic_duck/England/032919/2022 | Animal and Plant Health Agency (APHA) | Animal and Plant Health Agency (APHA) |
| EPI2072274 | NS | United Kingdom | 2022-Mar-26 | EPI_ISL_13370915 | A/domestic_duck/England/032919/2022 | Animal and Plant Health Agency (APHA) | Animal and Plant Health Agency (APHA) |
| EPI2072275 | MP | United Kingdom | 2022-Mar-26 | EPI_ISL_13370915 | A/domestic_duck/England/032919/2022 | Animal and Plant Health Agency (APHA) | Animal and Plant Health Agency (APHA) |
| EPI2072281 | NP | United Kingdom | 2022-Mar-27 | EPI_ISL_13370916 | A/chicken/England/033318/2022 | Animal and Plant Health Agency (APHA) | Animal and Plant Health Agency (APHA) |
| EPI2072282 | NS | United Kingdom | 2022-Mar-27 | EPI_ISL_13370916 | A/chicken/England/033318/2022 | Animal and Plant Health Agency (APHA) | Animal and Plant Health Agency (APHA) |
| EPI2072283 | MP | United Kingdom | 2022-Mar-27 | EPI_ISL_13370916 | A/chicken/England/033318/2022 | Animal and Plant Health Agency (APHA) | Animal and Plant Health Agency (APHA) |
| EPI2072289 | NP | United Kingdom | 2022-Mar-28 | EPI_ISL_13370917 | A/chicken/England/034820/2022 | Animal and Plant Health Agency (APHA) | Animal and Plant Health Agency (APHA) |
| EPI2072290 | NS | United Kingdom | 2022-Mar-28 | EPI_ISL_13370917 | A/chicken/England/034820/2022 | Animal and Plant Health Agency (APHA) | Animal and Plant Health Agency (APHA) |
| EPI2072291 | MP | United Kingdom | 2022-Mar-28 | EPI_ISL_13370917 | A/chicken/England/034820/2022 | Animal and Plant Health Agency (APHA) | Animal and Plant Health Agency (APHA) |
| EPI2072299 | MP | United Kingdom | 2022-Apr-04 | EPI_ISL_13370918 | A/domestic_duck/England/041295/2022 | Animal and Plant Health Agency (APHA) | Animal and Plant Health Agency (APHA) |
| EPI2072297 | NP | United Kingdom | 2022-Apr-04 | EPI_ISL_13370918 | A/domestic_duck/England/041295/2022 | Animal and Plant Health Agency (APHA) | Animal and Plant Health Agency (APHA) |
| EPI2072298 | NS | United Kingdom | 2022-Apr-04 | EPI_ISL_13370918 | A/domestic_duck/England/041295/2022 | Animal and Plant Health Agency (APHA) | Animal and Plant Health Agency (APHA) |
| EPI2072305 | NP | United Kingdom | 2022-Apr-04 | EPI_ISL_13370919 | A/domestic_goose/England/041268/2022 | Animal and Plant Health Agency (APHA) | Animal and Plant Health Agency (APHA) |
| EPI2072306 | NS | United Kingdom | 2022-Apr-04 | EPI_ISL_13370919 | A/domestic_goose/England/041268/2022 | Animal and Plant Health Agency (APHA) | Animal and Plant Health Agency (APHA) |
| EPI2072307 | MP | United Kingdom | 2022-Apr-04 | EPI_ISL_13370919 | A/domestic_goose/England/041268/2022 | Animal and Plant Health Agency (APHA) | Animal and Plant Health Agency (APHA) |
| EPI2072313 | NP | United Kingdom | 2022-Apr-04 | EPI_ISL_13370920 | A/domestic_duck/England/040831/2022 | Animal and Plant Health Agency (APHA) | Animal and Plant Health Agency (APHA) |
| EPI2072314 | NS | United Kingdom | 2022-Apr-04 | EPI_ISL_13370920 | A/domestic_duck/England/040831/2022 | Animal and Plant Health Agency (APHA) | Animal and Plant Health Agency (APHA) |
| EPI2072315 | MP | United Kingdom | 2022-Apr-04 | EPI_ISL_13370920 | A/domestic_duck/England/040831/2022 | Animal and Plant Health Agency (APHA) | Animal and Plant Health Agency (APHA) |
| EPI2072321 | NP | United Kingdom | 2022-Apr-06 | EPI_ISL_13370921 | A/domestic_duck/England/042266/2022 | Animal and Plant Health Agency (APHA) | Animal and Plant Health Agency (APHA) |
| EPI2072322 | NS | United Kingdom | 2022-Apr-06 | EPI_ISL_13370921 | A/domestic_duck/England/042266/2022 | Animal and Plant Health Agency (APHA) | Animal and Plant Health Agency (APHA) |
| EPI2072323 | MP | United Kingdom | 2022-Apr-06 | EPI_ISL_13370921 | A/domestic_duck/England/042266/2022 | Animal and Plant Health Agency (APHA) | Animal and Plant Health Agency (APHA) |
| EPI2072329 | NP | United Kingdom | 2022-Apr-06 | EPI_ISL_13370922 | A/chicken/England/042391/2022 | Animal and Plant Health Agency (APHA) | Animal and Plant Health Agency (APHA) |
| EPI2072330 | NS | United Kingdom | 2022-Apr-06 | EPI_ISL_13370922 | A/chicken/England/042391/2022 | Animal and Plant Health Agency (APHA) | Animal and Plant Health Agency (APHA) |
| EPI2072331 | MP | United Kingdom | 2022-Apr-06 | EPI_ISL_13370922 | A/chicken/England/042391/2022 | Animal and Plant Health Agency (APHA) | Animal and Plant Health Agency (APHA) |
| EPI2072337 | NP | United Kingdom | 2022-Apr-12 | EPI_ISL_13370923 | A/chicken/England/046274/2022 | Animal and Plant Health Agency (APHA) | Animal and Plant Health Agency (APHA) |
| EPI2072338 | NS | United Kingdom | 2022-Apr-12 | EPI_ISL_13370923 | A/chicken/England/046274/2022 | Animal and Plant Health Agency (APHA) | Animal and Plant Health Agency (APHA) |
| EPI2072339 | MP | United Kingdom | 2022-Apr-12 | EPI_ISL_13370923 | A/chicken/England/046274/2022 | Animal and Plant Health Agency (APHA) | Animal and Plant Health Agency (APHA) |
| EPI2072345 | NP | United Kingdom | 2022-Apr-21 | EPI_ISL_13370924 | A/chicken/England/053826/2022 | Animal and Plant Health Agency (APHA) | Animal and Plant Health Agency (APHA) |
| EPI2072346 | NS | United Kingdom | 2022-Apr-21 | EPI_ISL_13370924 | A/chicken/England/053826/2022 | Animal and Plant Health Agency (APHA) | Animal and Plant Health Agency (APHA) |
| EPI2072347 | MP | United Kingdom | 2022-Apr-21 | EPI_ISL_13370924 | A/chicken/England/053826/2022 | Animal and Plant Health Agency (APHA) | Animal and Plant Health Agency (APHA) |
| EPI2073617 | NP | Netherlands | 2022-Jun-03 | EPI_ISL_13429290 | A/Caspian Gull/Netherlands/4/2022 | Erasmus Medical Center | Erasmus Medical Center |

| | | | | | | | |
|---|---|---|---|---|---|---|---|
| EPI2073618 | NS | Netherlands | 2022-Jun-03 | EPI_ISL_13429290 | A/Caspian Gull/Netherlands/4/2022 | Erasmus Medical Center | Erasmus Medical Center |
| EPI2073619 | MP | Netherlands | 2022-Jun-03 | EPI_ISL_13429290 | A/Caspian Gull/Netherlands/4/2022 | Erasmus Medical Center | Erasmus Medical Center |
| EPI2073625 | NP | Netherlands | 2022-Jun-06 | EPI_ISL_13429291 | A/Sandwich Tern/Netherlands/8/2022 | Erasmus Medical Center | Erasmus Medical Center |
| EPI2073626 | NS | Netherlands | 2022-Jun-06 | EPI_ISL_13429291 | A/Sandwich Tern/Netherlands/8/2022 | Erasmus Medical Center | Erasmus Medical Center |
| EPI2073627 | MP | Netherlands | 2022-Jun-06 | EPI_ISL_13429291 | A/Sandwich Tern/Netherlands/8/2022 | Erasmus Medical Center | Erasmus Medical Center |
| EPI2073633 | NP | Netherlands | 2022-Jun-06 | EPI_ISL_13429292 | A/Sandwich Tern/Netherlands/5/2022 | Erasmus Medical Center | Erasmus Medical Center |
| EPI2073634 | NS | Netherlands | 2022-Jun-06 | EPI_ISL_13429292 | A/Sandwich Tern/Netherlands/5/2022 | Erasmus Medical Center | Erasmus Medical Center |
| EPI2073635 | MP | Netherlands | 2022-Jun-06 | EPI_ISL_13429292 | A/Sandwich Tern/Netherlands/5/2022 | Erasmus Medical Center | Erasmus Medical Center |
| EPI2073641 | NP | Netherlands | 2022-Jun-03 | EPI_ISL_13429293 | A/Black-headed gull/Netherlands/5/2022 | Erasmus Medical Center | Erasmus Medical Center |
| EPI2073642 | NS | Netherlands | 2022-Jun-03 | EPI_ISL_13429293 | A/Black-headed gull/Netherlands/5/2022 | Erasmus Medical Center | Erasmus Medical Center |
| EPI2073643 | MP | Netherlands | 2022-Jun-03 | EPI_ISL_13429293 | A/Black-headed gull/Netherlands/5/2022 | Erasmus Medical Center | Erasmus Medical Center |
| EPI2073649 | NP | Netherlands | 2022-Jun-03 | EPI_ISL_13429294 | A/Black-headed gull/Netherlands/6/2022 | Erasmus Medical Center | Erasmus Medical Center |
| EPI2073650 | NS | Netherlands | 2022-Jun-03 | EPI_ISL_13429294 | A/Black-headed gull/Netherlands/6/2022 | Erasmus Medical Center | Erasmus Medical Center |
| EPI2073651 | MP | Netherlands | 2022-Jun-03 | EPI_ISL_13429294 | A/Black-headed gull/Netherlands/6/2022 | Erasmus Medical Center | Erasmus Medical Center |
| EPI2073657 | NP | Netherlands | 2022-Jun-04 | EPI_ISL_13429295 | A/Sandwich Tern/Netherlands/1/2022 | Erasmus Medical Center | Erasmus Medical Center |
| EPI2073658 | NS | Netherlands | 2022-Jun-04 | EPI_ISL_13429295 | A/Sandwich Tern/Netherlands/1/2022 | Erasmus Medical Center | Erasmus Medical Center |
| EPI2073659 | MP | Netherlands | 2022-Jun-04 | EPI_ISL_13429295 | A/Sandwich Tern/Netherlands/1/2022 | Erasmus Medical Center | Erasmus Medical Center |
| EPI2073665 | NP | Netherlands | 2022-Jun-03 | EPI_ISL_13429296 | A/Sandwich Tern/Netherlands/2/2022 | Erasmus Medical Center | Erasmus Medical Center |
| EPI2073666 | NS | Netherlands | 2022-Jun-03 | EPI_ISL_13429296 | A/Sandwich Tern/Netherlands/2/2022 | Erasmus Medical Center | Erasmus Medical Center |
| EPI2073667 | MP | Netherlands | 2022-Jun-03 | EPI_ISL_13429296 | A/Sandwich Tern/Netherlands/2/2022 | Erasmus Medical Center | Erasmus Medical Center |
| EPI2073673 | NP | Netherlands | 2022-Jun-03 | EPI_ISL_13429297 | A/Sandwich Tern/Netherlands/3/2022 | Erasmus Medical Center | Erasmus Medical Center |
| EPI2073674 | NS | Netherlands | 2022-Jun-03 | EPI_ISL_13429297 | A/Sandwich Tern/Netherlands/3/2022 | Erasmus Medical Center | Erasmus Medical Center |
| EPI2073675 | MP | Netherlands | 2022-Jun-03 | EPI_ISL_13429297 | A/Sandwich Tern/Netherlands/3/2022 | Erasmus Medical Center | Erasmus Medical Center |
| EPI2073681 | NP | Netherlands | 2022-Jun-03 | EPI_ISL_13429298 | A/Common Tern/Netherlands/1/2022 | Erasmus Medical Center | Erasmus Medical Center |
| EPI2073682 | NS | Netherlands | 2022-Jun-03 | EPI_ISL_13429298 | A/Common Tern/Netherlands/1/2022 | Erasmus Medical Center | Erasmus Medical Center |
| EPI2073683 | MP | Netherlands | 2022-Jun-03 | EPI_ISL_13429298 | A/Common Tern/Netherlands/1/2022 | Erasmus Medical Center | Erasmus Medical Center |
| EPI2073689 | NP | Netherlands | 2022-Jun-06 | EPI_ISL_13429299 | A/Sandwich Tern/Netherlands/6/2022 | Erasmus Medical Center | Erasmus Medical Center |
| EPI2073690 | NS | Netherlands | 2022-Jun-06 | EPI_ISL_13429299 | A/Sandwich Tern/Netherlands/6/2022 | Erasmus Medical Center | Erasmus Medical Center |
| EPI2073691 | MP | Netherlands | 2022-Jun-06 | EPI_ISL_13429299 | A/Sandwich Tern/Netherlands/6/2022 | Erasmus Medical Center | Erasmus Medical Center |
| EPI2073697 | NP | Netherlands | 2022-Jun-06 | EPI_ISL_13429300 | A/Sandwich Tern/Netherlands/7/2022 | Erasmus Medical Center | Erasmus Medical Center |
| EPI2073698 | NS | Netherlands | 2022-Jun-06 | EPI_ISL_13429300 | A/Sandwich Tern/Netherlands/7/2022 | Erasmus Medical Center | Erasmus Medical Center |
| EPI2073699 | MP | Netherlands | 2022-Jun-06 | EPI_ISL_13429300 | A/Sandwich Tern/Netherlands/7/2022 | Erasmus Medical Center | Erasmus Medical Center |
| EPI2075593 | NP | United Kingdom | 2022-May-18 | EPI_ISL_13453559 | A/domestic_duck/England/073865/2022 | Animal and Plant Health Agency (APHA) | Animal and Plant Health Agency (APHA) |
| EPI2075595 | NS | United Kingdom | 2022-May-18 | EPI_ISL_13453559 | A/domestic_duck/England/073865/2022 | Animal and Plant Health Agency (APHA) | Animal and Plant Health Agency (APHA) |
| EPI2075596 | MP | United Kingdom | 2022-May-18 | EPI_ISL_13453559 | A/domestic_duck/England/073865/2022 | Animal and Plant Health Agency (APHA) | Animal and Plant Health Agency (APHA) |

| | | | | | | | |
|---|---|---|---|---|---|---|---|
| EPI2075602 | NP | United Kingdom | 2022-May-29 | EPI_ISL_13453560 | A/chicken/Scotland/078873/2022 | Animal and Plant Health Agency (APHA) | Animal and Plant Health Agency (APHA) |
| EPI2075603 | NS | United Kingdom | 2022-May-29 | EPI_ISL_13453560 | A/chicken/Scotland/078873/2022 | Animal and Plant Health Agency (APHA) | Animal and Plant Health Agency (APHA) |
| EPI2075604 | MP | United Kingdom | 2022-May-29 | EPI_ISL_13453560 | A/chicken/Scotland/078873/2022 | Animal and Plant Health Agency (APHA) | Animal and Plant Health Agency (APHA) |
| EPI2075635 | NP | United Kingdom | 2022-Mar-18 | EPI_ISL_13453564 | A/goose/Scotland/031099/2022 | Animal and Plant Health Agency (APHA) | Animal and Plant Health Agency (APHA) |
| EPI2075636 | NS | United Kingdom | 2022-Mar-18 | EPI_ISL_13453564 | A/goose/Scotland/031099/2022 | Animal and Plant Health Agency (APHA) | Animal and Plant Health Agency (APHA) |
| EPI2075637 | MP | United Kingdom | 2022-Mar-18 | EPI_ISL_13453564 | A/goose/Scotland/031099/2022 | Animal and Plant Health Agency (APHA) | Animal and Plant Health Agency (APHA) |
| EPI2075651 | NP | United Kingdom | 2022-May-31 | EPI_ISL_13453567 | A/turkey/England/081395/2022 | Animal and Plant Health Agency (APHA) | Animal and Plant Health Agency (APHA) |
| EPI2075652 | NS | United Kingdom | 2022-May-31 | EPI_ISL_13453567 | A/turkey/England/081395/2022 | Animal and Plant Health Agency (APHA) | Animal and Plant Health Agency (APHA) |
| EPI2075653 | MP | United Kingdom | 2022-May-31 | EPI_ISL_13453567 | A/turkey/England/081395/2022 | Animal and Plant Health Agency (APHA) | Animal and Plant Health Agency (APHA) |
| EPI2075659 | NP | United Kingdom | 2022-Jun-05 | EPI_ISL_13453568 | A/turkey/England/081490/2022 | Animal and Plant Health Agency (APHA) | Animal and Plant Health Agency (APHA) |
| EPI2075660 | NS | United Kingdom | 2022-Jun-05 | EPI_ISL_13453568 | A/turkey/England/081490/2022 | Animal and Plant Health Agency (APHA) | Animal and Plant Health Agency (APHA) |
| EPI2075661 | MP | United Kingdom | 2022-Jun-05 | EPI_ISL_13453568 | A/turkey/England/081490/2022 | Animal and Plant Health Agency (APHA) | Animal and Plant Health Agency (APHA) |
| EPI2075667 | NP | United Kingdom | 2022-Apr-26 | EPI_ISL_13453569 | A/tawny_owl/England/113433/2022 | Animal and Plant Health Agency (APHA) | Animal and Plant Health Agency (APHA) |
| EPI2075668 | NS | United Kingdom | 2022-Apr-26 | EPI_ISL_13453569 | A/tawny_owl/England/113433/2022 | Animal and Plant Health Agency (APHA) | Animal and Plant Health Agency (APHA) |
| EPI2075669 | MP | United Kingdom | 2022-Apr-26 | EPI_ISL_13453569 | A/tawny_owl/England/113433/2022 | Animal and Plant Health Agency (APHA) | Animal and Plant Health Agency (APHA) |
| EPI2075675 | NP | United Kingdom | 2022-Apr-29 | EPI_ISL_13453570 | A/great_skua/Scotland/5/2022 | Animal and Plant Health Agency (APHA) | Animal and Plant Health Agency (APHA) |
| EPI2075676 | NS | United Kingdom | 2022-Apr-29 | EPI_ISL_13453570 | A/great_skua/Scotland/5/2022 | Animal and Plant Health Agency (APHA) | Animal and Plant Health Agency (APHA) |
| EPI2075677 | MP | United Kingdom | 2022-Apr-29 | EPI_ISL_13453570 | A/great_skua/Scotland/5/2022 | Animal and Plant Health Agency (APHA) | Animal and Plant Health Agency (APHA) |
| EPI2075683 | NP | United Kingdom | 2022-May-10 | EPI_ISL_13453571 | A/gannet/Scotland/3/2022 | Animal and Plant Health Agency (APHA) | Animal and Plant Health Agency (APHA) |
| EPI2075684 | NS | United Kingdom | 2022-May-10 | EPI_ISL_13453571 | A/gannet/Scotland/3/2022 | Animal and Plant Health Agency (APHA) | Animal and Plant Health Agency (APHA) |
| EPI2075685 | MP | United Kingdom | 2022-May-10 | EPI_ISL_13453571 | A/gannet/Scotland/3/2022 | Animal and Plant Health Agency (APHA) | Animal and Plant Health Agency (APHA) |
| EPI2076254 | NP | United Kingdom | 2022-Jan-02 | EPI_ISL_13486700 | A/Barnacle_goose/Scotland/003579/2022 | Animal and Plant Health Agency (APHA) | Animal and Plant Health Agency (APHA) |
| EPI2076255 | NS | United Kingdom | 2022-Jan-02 | EPI_ISL_13486700 | A/Barnacle_goose/Scotland/003579/2022 | Animal and Plant Health Agency (APHA) | Animal and Plant Health Agency (APHA) |
| EPI2076256 | MP | United Kingdom | 2022-Jan-02 | EPI_ISL_13486700 | A/Barnacle_goose/Scotland/003579/2022 | Animal and Plant Health Agency (APHA) | Animal and Plant Health Agency (APHA) |
| EPI2076262 | NP | United Kingdom | 2022-Jan-02 | EPI_ISL_13486713 | A/Barnacle_goose/Scotland/003588/2022 | Animal and Plant Health Agency (APHA) | Animal and Plant Health Agency (APHA) |
| EPI2076263 | NS | United Kingdom | 2022-Jan-02 | EPI_ISL_13486713 | A/Barnacle_goose/Scotland/003588/2022 | Animal and Plant Health Agency (APHA) | Animal and Plant Health Agency (APHA) |
| EPI2076264 | MP | United Kingdom | 2022-Jan-02 | EPI_ISL_13486713 | A/Barnacle_goose/Scotland/003588/2022 | Animal and Plant Health Agency (APHA) | Animal and Plant Health Agency (APHA) |
| EPI2076326 | NP | United Kingdom | 2022-Mar-01 | EPI_ISL_13486807 | A/Common_buzzard/Scotland/020817/2022 | Animal and Plant Health Agency (APHA) | Animal and Plant Health Agency (APHA) |
| EPI2076327 | NS | United Kingdom | 2022-Mar-01 | EPI_ISL_13486807 | A/Common_buzzard/Scotland/020817/2022 | Animal and Plant Health Agency (APHA) | Animal and Plant Health Agency (APHA) |
| EPI2076328 | MP | United Kingdom | 2022-Mar-01 | EPI_ISL_13486807 | A/Common_buzzard/Scotland/020817/2022 | Animal and Plant Health Agency (APHA) | Animal and Plant Health Agency (APHA) |
| EPI2076342 | NP | United Kingdom | 2022-Mar-03 | EPI_ISL_13486831 | A/Common_eider/England/023549/2022 | Animal and Plant Health Agency (APHA) | Animal and Plant Health Agency (APHA) |
| EPI2076343 | NS | United Kingdom | 2022-Mar-03 | EPI_ISL_13486831 | A/Common_eider/England/023549/2022 | Animal and Plant Health Agency (APHA) | Animal and Plant Health Agency (APHA) |
| EPI2076344 | MP | United Kingdom | 2022-Mar-03 | EPI_ISL_13486831 | A/Common_eider/England/023549/2022 | Animal and Plant Health Agency (APHA) | Animal and Plant Health Agency (APHA) |
| EPI2076350 | NP | United Kingdom | 2022-Mar-02 | EPI_ISL_13486844 | A/goose/Scotland/022633/2022 | Animal and Plant Health Agency (APHA) | Animal and Plant Health Agency (APHA) |
| EPI2076351 | NS | United Kingdom | 2022-Mar-02 | EPI_ISL_13486844 | A/goose/Scotland/022633/2022 | Animal and Plant Health Agency (APHA) | Animal and Plant Health Agency (APHA) |

| | | | | | | | | |
|---|---|---|---|---|---|---|---|---|
| EPI2076352 | MP | United Kingdom | 2022-Mar-02 | EPI_ISL_13486844 | A/goose/Scotland/022633/2022 | Animal and Plant Health Agency (APHA) | Animal and Plant Health Agency (APHA) | |
| EPI2076360 | MP | United Kingdom | 2022-Mar-02 | EPI_ISL_13486857 | A/goose/Scotland/022634/2022 | Animal and Plant Health Agency (APHA) | Animal and Plant Health Agency (APHA) | |
| EPI2076358 | NP | United Kingdom | 2022-Mar-02 | EPI_ISL_13486857 | A/goose/Scotland/022634/2022 | Animal and Plant Health Agency (APHA) | Animal and Plant Health Agency (APHA) | |
| EPI2076359 | NS | United Kingdom | 2022-Mar-02 | EPI_ISL_13486857 | A/goose/Scotland/022634/2022 | Animal and Plant Health Agency (APHA) | Animal and Plant Health Agency (APHA) | |
| EPI2076366 | NP | United Kingdom | 2022-Mar-07 | EPI_ISL_13486869 | A/Greylag_goose/Scotland/024915/2022 | Animal and Plant Health Agency (APHA) | Animal and Plant Health Agency (APHA) | |
| EPI2076367 | NS | United Kingdom | 2022-Mar-07 | EPI_ISL_13486869 | A/Greylag_goose/Scotland/024915/2022 | Animal and Plant Health Agency (APHA) | Animal and Plant Health Agency (APHA) | |
| EPI2076368 | MP | United Kingdom | 2022-Mar-07 | EPI_ISL_13486869 | A/Greylag_goose/Scotland/024915/2022 | Animal and Plant Health Agency (APHA) | Animal and Plant Health Agency (APHA) | |
| EPI2076382 | NP | United Kingdom | 2022-Mar-04 | EPI_ISL_13486890 | A/Mute_Swan/Wales/057697/2022 | Animal and Plant Health Agency (APHA) | Animal and Plant Health Agency (APHA) | |
| EPI2076383 | NS | United Kingdom | 2022-Mar-04 | EPI_ISL_13486890 | A/Mute_Swan/Wales/057697/2022 | Animal and Plant Health Agency (APHA) | Animal and Plant Health Agency (APHA) | |
| EPI2076384 | MP | United Kingdom | 2022-Mar-04 | EPI_ISL_13486890 | A/Mute_Swan/Wales/057697/2022 | Animal and Plant Health Agency (APHA) | Animal and Plant Health Agency (APHA) | |
| EPI2076390 | NP | United Kingdom | 2022-Feb-25 | EPI_ISL_13486962 | A/Sparrowhawk/Scotland/022623/2022 | Animal and Plant Health Agency (APHA) | Animal and Plant Health Agency (APHA) | |
| EPI2076391 | NS | United Kingdom | 2022-Feb-25 | EPI_ISL_13486962 | A/Sparrowhawk/Scotland/022623/2022 | Animal and Plant Health Agency (APHA) | Animal and Plant Health Agency (APHA) | |
| EPI2076392 | MP | United Kingdom | 2022-Feb-25 | EPI_ISL_13486962 | A/Sparrowhawk/Scotland/022623/2022 | Animal and Plant Health Agency (APHA) | Animal and Plant Health Agency (APHA) | |
| EPI2077986 | MP | Belgium | 2022-Jun-04 | EPI_ISL_13514370 | A/Thalasseus sandvicensis/Belgium/7473_0009/2022 | Sciensano - Animal Infectious Diseases | Sciensano, Department of Animal Infectious Diseases | Van Borm, Steven; Roupie, Virginie; Lambrecht, Benedicte; Mathijs, Elisabeth; Steensels, Mieke |
| EPI2077984 | NP | Belgium | 2022-Jun-04 | EPI_ISL_13514370 | A/Thalasseus sandvicensis/Belgium/7473_0009/2022 | Sciensano - Animal Infectious Diseases | Sciensano, Department of Animal Infectious Diseases | Van Borm, Steven; Roupie, Virginie; Lambrecht, Benedicte; Mathijs, Elisabeth; Steensels, Mieke |
| EPI2077987 | NS | Belgium | 2022-Jun-04 | EPI_ISL_13514370 | A/Thalasseus sandvicensis/Belgium/7473_0009/2022 | Sciensano - Animal Infectious Diseases | Sciensano, Department of Animal Infectious Diseases | Van Borm, Steven; Roupie, Virginie; Lambrecht, Benedicte; Mathijs, Elisabeth; Steensels, Mieke |
| EPI2077994 | MP | Belgium | 2022-Jun-20 | EPI_ISL_13514375 | A/Gallus gallus/Belgium/8092_0002/2022 | Sciensano - Animal Infectious Diseases | Sciensano, Department of Animal Infectious Diseases | Van Borm, Steven; Roupie, Virginie; Lambrecht, Benedicte; Mathijs, Elisabeth; Steensels, Mieke |
| EPI2077995 | NS | Belgium | 2022-Jun-20 | EPI_ISL_13514375 | A/Gallus gallus/Belgium/8092_0002/2022 | Sciensano - Animal Infectious Diseases | Sciensano, Department of Animal Infectious Diseases | Van Borm, Steven; Roupie, Virginie; Lambrecht, Benedicte; Mathijs, Elisabeth; Steensels, Mieke |
| EPI2077992 | NP | Belgium | 2022-Jun-20 | EPI_ISL_13514375 | A/Gallus gallus/Belgium/8092_0002/2022 | Sciensano - Animal Infectious Diseases | Sciensano, Department of Animal Infectious Diseases | Van Borm, Steven; Roupie, Virginie; Lambrecht, Benedicte; Mathijs, Elisabeth; Steensels, Mieke |
| EPI2078357 | MP | France | 2022-May-11 | EPI_ISL_13519451 | A/gull/France/22P015977/2022 | Anses (Ploufragan-Plouzané) | ANSES Agence Nationale De Securite Sanitaire De L'alimentation | |
| EPI2078355 | NP | France | 2022-May-11 | EPI_ISL_13519451 | A/gull/France/22P015977/2022 | Anses (Ploufragan-Plouzané) | ANSES Agence Nationale De Securite Sanitaire De L'alimentation | |
| EPI2078358 | NS | France | 2022-May-11 | EPI_ISL_13519451 | A/gull/France/22P015977/2022 | Anses (Ploufragan-Plouzané) | ANSES Agence Nationale De Securite Sanitaire De L'alimentation | |
| EPI2082704 | MP | Netherlands | 2022-Jun-06 | EPI_ISL_13613219 | A/Greylag Goose/Netherlands/4/2022 | Erasmus Medical Center | Erasmus Medical Center | |
| EPI2082702 | NP | Netherlands | 2022-Jun-06 | EPI_ISL_13613219 | A/Greylag Goose/Netherlands/4/2022 | Erasmus Medical Center | Erasmus Medical Center | |
| EPI2082703 | NS | Netherlands | 2022-Jun-06 | EPI_ISL_13613219 | A/Greylag Goose/Netherlands/4/2022 | Erasmus Medical Center | Erasmus Medical Center | |
| EPI2082710 | NP | Netherlands | 2022-Jun-06 | EPI_ISL_13613220 | A/Greylag Goose/Netherlands/5/2022 | Erasmus Medical Center | Erasmus Medical Center | |
| EPI2082711 | NS | Netherlands | 2022-Jun-06 | EPI_ISL_13613220 | A/Greylag Goose/Netherlands/5/2022 | Erasmus Medical Center | Erasmus Medical Center | |
| EPI2082712 | MP | Netherlands | 2022-Jun-06 | EPI_ISL_13613220 | A/Greylag Goose/Netherlands/5/2022 | Erasmus Medical Center | Erasmus Medical Center | |
| EPI2082718 | NP | Netherlands | 2022-Jun-06 | EPI_ISL_13613221 | A/Greylag Goose/Netherlands/6/2022 | Erasmus Medical Center | Erasmus Medical Center | |
| EPI2082719 | NS | Netherlands | 2022-Jun-06 | EPI_ISL_13613221 | A/Greylag Goose/Netherlands/6/2022 | Erasmus Medical Center | Erasmus Medical Center | |
| EPI2082720 | MP | Netherlands | 2022-Jun-06 | EPI_ISL_13613221 | A/Greylag Goose/Netherlands/6/2022 | Erasmus Medical Center | Erasmus Medical Center | |
| EPI2082726 | NP | Netherlands | 2022-Jun-06 | EPI_ISL_13613222 | A/Greylag Goose/Netherlands/7/2022 | Erasmus Medical Center | Erasmus Medical Center | |
| EPI2082727 | NS | Netherlands | 2022-Jun-06 | EPI_ISL_13613222 | A/Greylag Goose/Netherlands/7/2022 | Erasmus Medical Center | Erasmus Medical Center | |
| EPI2082728 | MP | Netherlands | 2022-Jun-06 | EPI_ISL_13613222 | A/Greylag Goose/Netherlands/7/2022 | Erasmus Medical Center | Erasmus Medical Center | |
| EPI2082734 | NP | Netherlands | 2022-Jun-06 | EPI_ISL_13613223 | A/Greylag Goose/Netherlands/8/2022 | Erasmus Medical Center | Erasmus Medical Center | |

| | | | | | | | |
|---|---|---|---|---|---|---|---|
| EPI2082735 | NS | Netherlands | 2022-Jun-06 | EPI_ISL_13613223 | A/Greylag Goose/Netherlands/8/2022 | Erasmus Medical Center | Erasmus Medical Center |
| EPI2082736 | MP | Netherlands | 2022-Jun-06 | EPI_ISL_13613223 | A/Greylag Goose/Netherlands/8/2022 | Erasmus Medical Center | Erasmus Medical Center |
| EPI2082742 | NP | Netherlands | 2022-Jun-10 | EPI_ISL_13613224 | A/Common Tern/Netherlands/2/2022 | Erasmus Medical Center | Erasmus Medical Center |
| EPI2082743 | NS | Netherlands | 2022-Jun-10 | EPI_ISL_13613224 | A/Common Tern/Netherlands/2/2022 | Erasmus Medical Center | Erasmus Medical Center |
| EPI2082744 | MP | Netherlands | 2022-Jun-10 | EPI_ISL_13613224 | A/Common Tern/Netherlands/2/2022 | Erasmus Medical Center | Erasmus Medical Center |
| EPI2082750 | NP | Netherlands | 2022-Jun-10 | EPI_ISL_13613225 | A/Black-headed gull/Netherlands/7/2022 | Erasmus Medical Center | Erasmus Medical Center |
| EPI2082751 | NS | Netherlands | 2022-Jun-10 | EPI_ISL_13613225 | A/Black-headed gull/Netherlands/7/2022 | Erasmus Medical Center | Erasmus Medical Center |
| EPI2082752 | MP | Netherlands | 2022-Jun-10 | EPI_ISL_13613225 | A/Black-headed gull/Netherlands/7/2022 | Erasmus Medical Center | Erasmus Medical Center |
| EPI2082758 | NP | Netherlands | 2022-Jun-10 | EPI_ISL_13613226 | A/Black-headed gull/Netherlands/8/2022 | Erasmus Medical Center | Erasmus Medical Center |
| EPI2082759 | NS | Netherlands | 2022-Jun-10 | EPI_ISL_13613226 | A/Black-headed gull/Netherlands/8/2022 | Erasmus Medical Center | Erasmus Medical Center |
| EPI2082760 | MP | Netherlands | 2022-Jun-10 | EPI_ISL_13613226 | A/Black-headed gull/Netherlands/8/2022 | Erasmus Medical Center | Erasmus Medical Center |
| EPI2082766 | NP | Netherlands | 2022-Jun-10 | EPI_ISL_13613227 | A/Black-headed gull/Netherlands/9/2022 | Erasmus Medical Center | Erasmus Medical Center |
| EPI2082767 | NS | Netherlands | 2022-Jun-10 | EPI_ISL_13613227 | A/Black-headed gull/Netherlands/9/2022 | Erasmus Medical Center | Erasmus Medical Center |
| EPI2082768 | MP | Netherlands | 2022-Jun-10 | EPI_ISL_13613227 | A/Black-headed gull/Netherlands/9/2022 | Erasmus Medical Center | Erasmus Medical Center |
| EPI2082774 | NP | Netherlands | 2022-Jun-12 | EPI_ISL_13613228 | A/Sandwich Tern/Netherlands/9/2022 | Erasmus Medical Center | Erasmus Medical Center |
| EPI2082775 | NS | Netherlands | 2022-Jun-12 | EPI_ISL_13613228 | A/Sandwich Tern/Netherlands/9/2022 | Erasmus Medical Center | Erasmus Medical Center |
| EPI2082776 | MP | Netherlands | 2022-Jun-12 | EPI_ISL_13613228 | A/Sandwich Tern/Netherlands/9/2022 | Erasmus Medical Center | Erasmus Medical Center |
| EPI2082782 | NP | Netherlands | 2022-Jun-12 | EPI_ISL_13613229 | A/Sandwich Tern/Netherlands/10/2022 | Erasmus Medical Center | Erasmus Medical Center |
| EPI2082783 | NS | Netherlands | 2022-Jun-12 | EPI_ISL_13613229 | A/Sandwich Tern/Netherlands/10/2022 | Erasmus Medical Center | Erasmus Medical Center |
| EPI2082784 | MP | Netherlands | 2022-Jun-12 | EPI_ISL_13613229 | A/Sandwich Tern/Netherlands/10/2022 | Erasmus Medical Center | Erasmus Medical Center |
| EPI2082790 | NP | Netherlands | 2022-Jun-13 | EPI_ISL_13613230 | A/Common Tern/Netherlands/3/2022 | Erasmus Medical Center | Erasmus Medical Center |
| EPI2082791 | NS | Netherlands | 2022-Jun-13 | EPI_ISL_13613230 | A/Common Tern/Netherlands/3/2022 | Erasmus Medical Center | Erasmus Medical Center |
| EPI2082792 | MP | Netherlands | 2022-Jun-13 | EPI_ISL_13613230 | A/Common Tern/Netherlands/3/2022 | Erasmus Medical Center | Erasmus Medical Center |
| EPI2088880 | NP | Netherlands | 2022-Jun-17 | EPI_ISL_13778462 | A/Lesser Black-backed Gull/Netherlands/2/2022 | Erasmus Medical Center | Erasmus Medical Center |
| EPI2088881 | NS | Netherlands | 2022-Jun-17 | EPI_ISL_13778462 | A/Lesser Black-backed Gull/Netherlands/2/2022 | Erasmus Medical Center | Erasmus Medical Center |
| EPI2088882 | MP | Netherlands | 2022-Jun-17 | EPI_ISL_13778462 | A/Lesser Black-backed Gull/Netherlands/2/2022 | Erasmus Medical Center | Erasmus Medical Center |
| EPI2088896 | NP | Netherlands | 2022-Jun-19 | EPI_ISL_13778466 | A/Oystercatcher/Netherlands/2/2022 | Erasmus Medical Center | Erasmus Medical Center |
| EPI2088897 | NS | Netherlands | 2022-Jun-19 | EPI_ISL_13778466 | A/Oystercatcher/Netherlands/2/2022 | Erasmus Medical Center | Erasmus Medical Center |
| EPI2088898 | MP | Netherlands | 2022-Jun-19 | EPI_ISL_13778466 | A/Oystercatcher/Netherlands/2/2022 | Erasmus Medical Center | Erasmus Medical Center |
| EPI2088903 | NP | Netherlands | 2022-Jun-19 | EPI_ISL_13778467 | A/Sandwich Tern/Netherlands/11/2022 | Erasmus Medical Center | Erasmus Medical Center |
| EPI2088904 | NS | Netherlands | 2022-Jun-19 | EPI_ISL_13778467 | A/Sandwich Tern/Netherlands/11/2022 | Erasmus Medical Center | Erasmus Medical Center |
| EPI2088905 | MP | Netherlands | 2022-Jun-19 | EPI_ISL_13778467 | A/Sandwich Tern/Netherlands/11/2022 | Erasmus Medical Center | Erasmus Medical Center |
| EPI2088911 | NP | Netherlands | 2022-Jun-07 | EPI_ISL_13778468 | A/European Herring Gull/Netherlands/5/2022 | Erasmus Medical Center | Erasmus Medical Center |
| EPI2088912 | NS | Netherlands | 2022-Jun-07 | EPI_ISL_13778468 | A/European Herring Gull/Netherlands/5/2022 | Erasmus Medical Center | Erasmus Medical Center |
| EPI2088913 | MP | Netherlands | 2022-Jun-07 | EPI_ISL_13778468 | A/European Herring Gull/Netherlands/5/2022 | Erasmus Medical Center | Erasmus Medical Center |

| | | | | | | | |
|---|---|---|---|---|---|---|---|
| EPI2088919 | NP | Netherlands | 2022-Jun-07 | EPI_ISL_13778469 | A/European Herring Gull/Netherlands/6/2022 | Erasmus Medical Center | Erasmus Medical Center |
| EPI2088920 | NS | Netherlands | 2022-Jun-07 | EPI_ISL_13778469 | A/European Herring Gull/Netherlands/6/2022 | Erasmus Medical Center | Erasmus Medical Center |
| EPI2088921 | MP | Netherlands | 2022-Jun-07 | EPI_ISL_13778469 | A/European Herring Gull/Netherlands/6/2022 | Erasmus Medical Center | Erasmus Medical Center |
| EPI2088927 | NP | Netherlands | 2022-Jun-07 | EPI_ISL_13778474 | A/European Herring Gull/Netherlands/7/2022 | Erasmus Medical Center | Erasmus Medical Center |
| EPI2088928 | NS | Netherlands | 2022-Jun-07 | EPI_ISL_13778474 | A/European Herring Gull/Netherlands/7/2022 | Erasmus Medical Center | Erasmus Medical Center |
| EPI2088929 | MP | Netherlands | 2022-Jun-07 | EPI_ISL_13778474 | A/European Herring Gull/Netherlands/7/2022 | Erasmus Medical Center | Erasmus Medical Center |
| EPI2088935 | NP | Netherlands | 2022-Jun-07 | EPI_ISL_13778475 | A/Sandwich Tern/Netherlands/12/2022 | Erasmus Medical Center | Erasmus Medical Center |
| EPI2088936 | NS | Netherlands | 2022-Jun-07 | EPI_ISL_13778475 | A/Sandwich Tern/Netherlands/12/2022 | Erasmus Medical Center | Erasmus Medical Center |
| EPI2088937 | MP | Netherlands | 2022-Jun-07 | EPI_ISL_13778475 | A/Sandwich Tern/Netherlands/12/2022 | Erasmus Medical Center | Erasmus Medical Center |
| EPI2088943 | NP | Netherlands | 2022-Jun-07 | EPI_ISL_13778476 | A/Sandwich Tern/Netherlands/13/2022 | Erasmus Medical Center | Erasmus Medical Center |
| EPI2088944 | NS | Netherlands | 2022-Jun-07 | EPI_ISL_13778476 | A/Sandwich Tern/Netherlands/13/2022 | Erasmus Medical Center | Erasmus Medical Center |
| EPI2088945 | MP | Netherlands | 2022-Jun-07 | EPI_ISL_13778476 | A/Sandwich Tern/Netherlands/13/2022 | Erasmus Medical Center | Erasmus Medical Center |
| EPI2088951 | NP | Netherlands | 2022-Jun-16 | EPI_ISL_13778477 | A/Common Tern/Netherlands/8/2022 | Erasmus Medical Center | Erasmus Medical Center |
| EPI2088952 | NS | Netherlands | 2022-Jun-16 | EPI_ISL_13778477 | A/Common Tern/Netherlands/8/2022 | Erasmus Medical Center | Erasmus Medical Center |
| EPI2088953 | MP | Netherlands | 2022-Jun-16 | EPI_ISL_13778477 | A/Common Tern/Netherlands/8/2022 | Erasmus Medical Center | Erasmus Medical Center |
| EPI2088959 | NP | Netherlands | 2022-Jun-24 | EPI_ISL_13778478 | A/Sandwich Tern/Netherlands/14/2022 | Erasmus Medical Center | Erasmus Medical Center |
| EPI2088960 | NS | Netherlands | 2022-Jun-24 | EPI_ISL_13778478 | A/Sandwich Tern/Netherlands/14/2022 | Erasmus Medical Center | Erasmus Medical Center |
| EPI2088961 | MP | Netherlands | 2022-Jun-24 | EPI_ISL_13778478 | A/Sandwich Tern/Netherlands/14/2022 | Erasmus Medical Center | Erasmus Medical Center |
| EPI2088967 | NP | Netherlands | 2022-Jun-24 | EPI_ISL_13778479 | A/Common Tern/Netherlands/9/2022 | Erasmus Medical Center | Erasmus Medical Center |
| EPI2088968 | NS | Netherlands | 2022-Jun-24 | EPI_ISL_13778479 | A/Common Tern/Netherlands/9/2022 | Erasmus Medical Center | Erasmus Medical Center |
| EPI2088969 | MP | Netherlands | 2022-Jun-24 | EPI_ISL_13778479 | A/Common Tern/Netherlands/9/2022 | Erasmus Medical Center | Erasmus Medical Center |
| EPI2088975 | NP | Netherlands | 2022-Jun-24 | EPI_ISL_13778522 | A/Sandwich Tern/Netherlands/15/2022 | Erasmus Medical Center | Erasmus Medical Center |
| EPI2088976 | NS | Netherlands | 2022-Jun-24 | EPI_ISL_13778522 | A/Sandwich Tern/Netherlands/15/2022 | Erasmus Medical Center | Erasmus Medical Center |
| EPI2088977 | MP | Netherlands | 2022-Jun-24 | EPI_ISL_13778522 | A/Sandwich Tern/Netherlands/15/2022 | Erasmus Medical Center | Erasmus Medical Center |
| EPI2088983 | NP | Netherlands | 2022-Jun-20 | EPI_ISL_13778523 | A/Common Tern/Netherlands/4/2022 | Erasmus Medical Center | Erasmus Medical Center |
| EPI2088984 | NS | Netherlands | 2022-Jun-20 | EPI_ISL_13778523 | A/Common Tern/Netherlands/4/2022 | Erasmus Medical Center | Erasmus Medical Center |
| EPI2088985 | MP | Netherlands | 2022-Jun-20 | EPI_ISL_13778523 | A/Common Tern/Netherlands/4/2022 | Erasmus Medical Center | Erasmus Medical Center |
| EPI2088991 | NP | Netherlands | 2022-Jun-20 | EPI_ISL_13778524 | A/Common Tern/Netherlands/5/2022 | Erasmus Medical Center | Erasmus Medical Center |
| EPI2088992 | NS | Netherlands | 2022-Jun-20 | EPI_ISL_13778524 | A/Common Tern/Netherlands/5/2022 | Erasmus Medical Center | Erasmus Medical Center |
| EPI2088993 | MP | Netherlands | 2022-Jun-20 | EPI_ISL_13778524 | A/Common Tern/Netherlands/5/2022 | Erasmus Medical Center | Erasmus Medical Center |
| EPI2088999 | NP | Netherlands | 2022-Jun-20 | EPI_ISL_13778525 | A/Common Tern/Netherlands/6/2022 | Erasmus Medical Center | Erasmus Medical Center |
| EPI2089000 | NS | Netherlands | 2022-Jun-20 | EPI_ISL_13778525 | A/Common Tern/Netherlands/6/2022 | Erasmus Medical Center | Erasmus Medical Center |
| EPI2089001 | MP | Netherlands | 2022-Jun-20 | EPI_ISL_13778525 | A/Common Tern/Netherlands/6/2022 | Erasmus Medical Center | Erasmus Medical Center |
| EPI2089007 | NP | Netherlands | 2022-Jun-20 | EPI_ISL_13778526 | A/Common Tern/Netherlands/7/2022 | Erasmus Medical Center | Erasmus Medical Center |
| EPI2089008 | NS | Netherlands | 2022-Jun-20 | EPI_ISL_13778526 | A/Common Tern/Netherlands/7/2022 | Erasmus Medical Center | Erasmus Medical Center |

| | | | | | | | |
|---|---|---|---|---|---|---|---|
| EPI2089009 | MP | Netherlands | 2022-Jun-20 | EPI_ISL_13778526 | A/Common Tern/Netherlands/7/2022 | Erasmus Medical Center | Erasmus Medical Center |
| EPI2089015 | NP | United Kingdom | 2022-Jun-15 | EPI_ISL_13782459 | A/chicken/England/085598/2022 | Animal and Plant Health Agency (APHA) | Animal and Plant Health Agency (APHA) |
| EPI2089016 | NS | United Kingdom | 2022-Jun-15 | EPI_ISL_13782459 | A/chicken/England/085598/2022 | Animal and Plant Health Agency (APHA) | Animal and Plant Health Agency (APHA) |
| EPI2089017 | MP | United Kingdom | 2022-Jun-15 | EPI_ISL_13782459 | A/chicken/England/085598/2022 | Animal and Plant Health Agency (APHA) | Animal and Plant Health Agency (APHA) |
| EPI2089031 | NP | United Kingdom | 2022-Jan-08 | EPI_ISL_13782461 | A/pink_footed_goose/Scotland/003617/2022 | Animal and Plant Health Agency (APHA) | Animal and Plant Health Agency (APHA) |
| EPI2089032 | NS | United Kingdom | 2022-Jan-08 | EPI_ISL_13782461 | A/pink_footed_goose/Scotland/003617/2022 | Animal and Plant Health Agency (APHA) | Animal and Plant Health Agency (APHA) |
| EPI2089033 | MP | United Kingdom | 2022-Jan-08 | EPI_ISL_13782461 | A/pink_footed_goose/Scotland/003617/2022 | Animal and Plant Health Agency (APHA) | Animal and Plant Health Agency (APHA) |
| EPI2097271 | NP | Netherlands | 2022-Jul-03 | EPI_ISL_13902858 | A/Eurasian Curlew/Netherlands/2/2022 | Erasmus Medical Center | Erasmus Medical Center |
| EPI2097272 | NS | Netherlands | 2022-Jul-03 | EPI_ISL_13902858 | A/Eurasian Curlew/Netherlands/2/2022 | Erasmus Medical Center | Erasmus Medical Center |
| EPI2097273 | MP | Netherlands | 2022-Jul-03 | EPI_ISL_13902858 | A/Eurasian Curlew/Netherlands/2/2022 | Erasmus Medical Center | Erasmus Medical Center |
| EPI2100192 | NP | United Kingdom | 2022-Mar-26 | EPI_ISL_13969421 | A/Pink-footed_Goose/Scotland/036871/2022 | Animal and Plant Health Agency (APHA) | Animal and Plant Health Agency (APHA) |
| EPI2100193 | NS | United Kingdom | 2022-Mar-26 | EPI_ISL_13969421 | A/Pink-footed_Goose/Scotland/036871/2022 | Animal and Plant Health Agency (APHA) | Animal and Plant Health Agency (APHA) |
| EPI2100194 | MP | United Kingdom | 2022-Mar-26 | EPI_ISL_13969421 | A/Pink-footed_Goose/Scotland/036871/2022 | Animal and Plant Health Agency (APHA) | Animal and Plant Health Agency (APHA) |
| EPI2100216 | NP | United Kingdom | 2022-Jun-06 | EPI_ISL_13969424 | A/Gannet/Scotland/084490/2022 | Animal and Plant Health Agency (APHA) | Animal and Plant Health Agency (APHA) |
| EPI2100217 | NS | United Kingdom | 2022-Jun-06 | EPI_ISL_13969424 | A/Gannet/Scotland/084490/2022 | Animal and Plant Health Agency (APHA) | Animal and Plant Health Agency (APHA) |
| EPI2100218 | MP | United Kingdom | 2022-Jun-06 | EPI_ISL_13969424 | A/Gannet/Scotland/084490/2022 | Animal and Plant Health Agency (APHA) | Animal and Plant Health Agency (APHA) |
| EPI2100224 | NP | United Kingdom | 2022-Jun-07 | EPI_ISL_13969425 | A/Gannet/Scotland/084624/2022 | Animal and Plant Health Agency (APHA) | Animal and Plant Health Agency (APHA) |
| EPI2100225 | NS | United Kingdom | 2022-Jun-07 | EPI_ISL_13969425 | A/Gannet/Scotland/084624/2022 | Animal and Plant Health Agency (APHA) | Animal and Plant Health Agency (APHA) |
| EPI2100226 | MP | United Kingdom | 2022-Jun-07 | EPI_ISL_13969425 | A/Gannet/Scotland/084624/2022 | Animal and Plant Health Agency (APHA) | Animal and Plant Health Agency (APHA) |
| EPI2100240 | NP | United Kingdom | 2022-Jun-05 | EPI_ISL_13969427 | A/Common_tern/England/248697/2022 | Animal and Plant Health Agency (APHA) | Animal and Plant Health Agency (APHA) |
| EPI2100241 | NS | United Kingdom | 2022-Jun-05 | EPI_ISL_13969427 | A/Common_tern/England/248697/2022 | Animal and Plant Health Agency (APHA) | Animal and Plant Health Agency (APHA) |
| EPI2100242 | MP | United Kingdom | 2022-Jun-05 | EPI_ISL_13969427 | A/Common_tern/England/248697/2022 | Animal and Plant Health Agency (APHA) | Animal and Plant Health Agency (APHA) |
| EPI2100256 | NP | United Kingdom | 2022-Mar-24 | EPI_ISL_13969429 | A/White-fronted_goose/England/311038/2022 | Animal and Plant Health Agency (APHA) | Animal and Plant Health Agency (APHA) |
| EPI2100257 | NS | United Kingdom | 2022-Mar-24 | EPI_ISL_13969429 | A/White-fronted_goose/England/311038/2022 | Animal and Plant Health Agency (APHA) | Animal and Plant Health Agency (APHA) |
| EPI2100258 | MP | United Kingdom | 2022-Mar-24 | EPI_ISL_13969429 | A/White-fronted_goose/England/311038/2022 | Animal and Plant Health Agency (APHA) | Animal and Plant Health Agency (APHA) |
| EPI2100264 | NP | United Kingdom | 2022-Apr-10 | EPI_ISL_13969430 | A/Mute_swan/Wales/058560/2022 | Animal and Plant Health Agency (APHA) | Animal and Plant Health Agency (APHA) |
| EPI2100265 | NS | United Kingdom | 2022-Apr-10 | EPI_ISL_13969430 | A/Mute_swan/Wales/058560/2022 | Animal and Plant Health Agency (APHA) | Animal and Plant Health Agency (APHA) |
| EPI2100266 | MP | United Kingdom | 2022-Apr-10 | EPI_ISL_13969430 | A/Mute_swan/Wales/058560/2022 | Animal and Plant Health Agency (APHA) | Animal and Plant Health Agency (APHA) |
| EPI2109880 | NP | United Kingdom | 2022-Jul-04 | EPI_ISL_14064983 | A/chicken/Scotland/093091/2022 | Animal and Plant Health Agency (APHA) | Animal and Plant Health Agency (APHA) |
| EPI2109881 | NS | United Kingdom | 2022-Jul-04 | EPI_ISL_14064983 | A/chicken/Scotland/093091/2022 | Animal and Plant Health Agency (APHA) | Animal and Plant Health Agency (APHA) |
| EPI2109882 | MP | United Kingdom | 2022-Jul-04 | EPI_ISL_14064983 | A/chicken/Scotland/093091/2022 | Animal and Plant Health Agency (APHA) | Animal and Plant Health Agency (APHA) |
| EPI2109888 | NP | United Kingdom | 2022-Jul-06 | EPI_ISL_14064984 | A/chicken/England/093459/2022 | Animal and Plant Health Agency (APHA) | Animal and Plant Health Agency (APHA) |
| EPI2109889 | NS | United Kingdom | 2022-Jul-06 | EPI_ISL_14064984 | A/chicken/England/093459/2022 | Animal and Plant Health Agency (APHA) | Animal and Plant Health Agency (APHA) |
| EPI2109890 | MP | United Kingdom | 2022-Jul-06 | EPI_ISL_14064984 | A/chicken/England/093459/2022 | Animal and Plant Health Agency (APHA) | Animal and Plant Health Agency (APHA) |
| EPI2109896 | NP | United Kingdom | 2022-Jul-06 | EPI_ISL_14064985 | A/Domestic_goose/England/093469/2022 | Animal and Plant Health Agency (APHA) | Animal and Plant Health Agency (APHA) |

| | | | | | | | |
|---|---|---|---|---|---|---|---|
| EPI2109897 | NS | United Kingdom | 2022-Jul-06 | EPI_ISL_14064985 | A/Domestic_goose/England/093469/2022 | Animal and Plant Health Agency (APHA) | Animal and Plant Health Agency (APHA) |
| EPI2109898 | MP | United Kingdom | 2022-Jul-06 | EPI_ISL_14064985 | A/Domestic_goose/England/093469/2022 | Animal and Plant Health Agency (APHA) | Animal and Plant Health Agency (APHA) |
| EPI2113995 | NP | Netherlands | 2022-Jul-07 | EPI_ISL_14163711 | A/Common Tern/Netherlands/10/2022 | Erasmus Medical Center | Erasmus Medical Center |
| EPI2113996 | NS | Netherlands | 2022-Jul-07 | EPI_ISL_14163711 | A/Common Tern/Netherlands/10/2022 | Erasmus Medical Center | Erasmus Medical Center |
| EPI2113997 | MP | Netherlands | 2022-Jul-07 | EPI_ISL_14163711 | A/Common Tern/Netherlands/10/2022 | Erasmus Medical Center | Erasmus Medical Center |
| EPI2114003 | NP | Netherlands | 2022-Jul-07 | EPI_ISL_14163712 | A/European Herring Gull/Netherlands/8/2022 | Erasmus Medical Center | Erasmus Medical Center |
| EPI2114004 | NS | Netherlands | 2022-Jul-07 | EPI_ISL_14163712 | A/European Herring Gull/Netherlands/8/2022 | Erasmus Medical Center | Erasmus Medical Center |
| EPI2114005 | MP | Netherlands | 2022-Jul-07 | EPI_ISL_14163712 | A/European Herring Gull/Netherlands/8/2022 | Erasmus Medical Center | Erasmus Medical Center |
| EPI2114011 | NP | Netherlands | 2022-Jul-08 | EPI_ISL_14163713 | A/Caspian Gull/Netherlands/5/2022 | Erasmus Medical Center | Erasmus Medical Center |
| EPI2114012 | NS | Netherlands | 2022-Jul-08 | EPI_ISL_14163713 | A/Caspian Gull/Netherlands/5/2022 | Erasmus Medical Center | Erasmus Medical Center |
| EPI2114013 | MP | Netherlands | 2022-Jul-08 | EPI_ISL_14163713 | A/Caspian Gull/Netherlands/5/2022 | Erasmus Medical Center | Erasmus Medical Center |
| EPI2114019 | NP | Netherlands | 2022-Jul-04 | EPI_ISL_14163714 | A/European Herring Gull/Netherlands/9/2022 | Erasmus Medical Center | Erasmus Medical Center |
| EPI2114020 | NS | Netherlands | 2022-Jul-04 | EPI_ISL_14163714 | A/European Herring Gull/Netherlands/9/2022 | Erasmus Medical Center | Erasmus Medical Center |
| EPI2114021 | MP | Netherlands | 2022-Jul-04 | EPI_ISL_14163714 | A/European Herring Gull/Netherlands/9/2022 | Erasmus Medical Center | Erasmus Medical Center |
| EPI2114027 | NP | Netherlands | 2022-Jul-12 | EPI_ISL_14163715 | A/Greylag Goose/Netherlands/9/2022 | Erasmus Medical Center | Erasmus Medical Center |
| EPI2114028 | NS | Netherlands | 2022-Jul-12 | EPI_ISL_14163715 | A/Greylag Goose/Netherlands/9/2022 | Erasmus Medical Center | Erasmus Medical Center |
| EPI2114029 | MP | Netherlands | 2022-Jul-12 | EPI_ISL_14163715 | A/Greylag Goose/Netherlands/9/2022 | Erasmus Medical Center | Erasmus Medical Center |
| EPI2114051 | NP | Netherlands | 2022-Jul-11 | EPI_ISL_14171729 | A/Eurasian Spoonbill/Netherlands/1A/2022 | Erasmus Medical Center | Erasmus Medical Center |
| EPI2114052 | NS | Netherlands | 2022-Jul-11 | EPI_ISL_14171729 | A/Eurasian Spoonbill/Netherlands/1A/2022 | Erasmus Medical Center | Erasmus Medical Center |
| EPI2114053 | MP | Netherlands | 2022-Jul-11 | EPI_ISL_14171729 | A/Eurasian Spoonbill/Netherlands/1A/2022 | Erasmus Medical Center | Erasmus Medical Center |
| EPI2114059 | NP | Netherlands | 2022-Jul-11 | EPI_ISL_14171741 | A/Eurasian Spoonbill/Netherlands/1B/2022 | Erasmus Medical Center | Erasmus Medical Center |
| EPI2114060 | NS | Netherlands | 2022-Jul-11 | EPI_ISL_14171741 | A/Eurasian Spoonbill/Netherlands/1B/2022 | Erasmus Medical Center | Erasmus Medical Center |
| EPI2114061 | MP | Netherlands | 2022-Jul-11 | EPI_ISL_14171741 | A/Eurasian Spoonbill/Netherlands/1B/2022 | Erasmus Medical Center | Erasmus Medical Center |
| EPI2119165 | MP | Netherlands | 2022-Jul-24 | EPI_ISL_14233919 | A/Eurasian Spoonbill/Netherlands/3/2022 | Erasmus Medical Center | Erasmus Medical Center |
| EPI2119163 | NP | Netherlands | 2022-Jul-24 | EPI_ISL_14233919 | A/Eurasian Spoonbill/Netherlands/3/2022 | Erasmus Medical Center | Erasmus Medical Center |
| EPI2119164 | NS | Netherlands | 2022-Jul-24 | EPI_ISL_14233919 | A/Eurasian Spoonbill/Netherlands/3/2022 | Erasmus Medical Center | Erasmus Medical Center |
| EPI2119173 | MP | Netherlands | 2022-Jul-15 | EPI_ISL_14233920 | A/Common Tern/Netherlands/17/2022 | Erasmus Medical Center | Erasmus Medical Center |
| EPI2119171 | NP | Netherlands | 2022-Jul-15 | EPI_ISL_14233920 | A/Common Tern/Netherlands/17/2022 | Erasmus Medical Center | Erasmus Medical Center |
| EPI2119172 | NS | Netherlands | 2022-Jul-15 | EPI_ISL_14233920 | A/Common Tern/Netherlands/17/2022 | Erasmus Medical Center | Erasmus Medical Center |
| EPI2119181 | MP | Netherlands | 2022-Jul-15 | EPI_ISL_14233921 | A/Common Tern/Netherlands/18/2022 | Erasmus Medical Center | Erasmus Medical Center |
| EPI2119179 | NP | Netherlands | 2022-Jul-15 | EPI_ISL_14233921 | A/Common Tern/Netherlands/18/2022 | Erasmus Medical Center | Erasmus Medical Center |
| EPI2119180 | NS | Netherlands | 2022-Jul-15 | EPI_ISL_14233921 | A/Common Tern/Netherlands/18/2022 | Erasmus Medical Center | Erasmus Medical Center |
| EPI2119189 | MP | Netherlands | 2022-Jul-19 | EPI_ISL_14233922 | A/European Herring Gull/Netherlands/11/2022 | Erasmus Medical Center | Erasmus Medical Center |
| EPI2119187 | NP | Netherlands | 2022-Jul-19 | EPI_ISL_14233922 | A/European Herring Gull/Netherlands/11/2022 | Erasmus Medical Center | Erasmus Medical Center |
| EPI2119188 | NS | Netherlands | 2022-Jul-19 | EPI_ISL_14233922 | A/European Herring Gull/Netherlands/11/2022 | Erasmus Medical Center | Erasmus Medical Center |

| | | | | | | | | |
|---|---|---|---|---|---|---|---|---|
| EPI2119197 | MP | Netherlands | 2022-Jul-15 | EPI_ISL_14233923 | A/Caspian Gull/Netherlands/7/2022 | Erasmus Medical Center | Erasmus Medical Center | |
| EPI2119195 | NP | Netherlands | 2022-Jul-15 | EPI_ISL_14233923 | A/Caspian Gull/Netherlands/7/2022 | Erasmus Medical Center | Erasmus Medical Center | |
| EPI2119196 | NS | Netherlands | 2022-Jul-15 | EPI_ISL_14233923 | A/Caspian Gull/Netherlands/7/2022 | Erasmus Medical Center | Erasmus Medical Center | |
| EPI2119203 | MP | Netherlands | 2022-Jul-22 | EPI_ISL_14233924 | A/Common Tern/Netherlands/19/2022 | Erasmus Medical Center | Erasmus Medical Center | |
| EPI2119201 | NP | Netherlands | 2022-Jul-22 | EPI_ISL_14233924 | A/Common Tern/Netherlands/19/2022 | Erasmus Medical Center | Erasmus Medical Center | |
| EPI2119202 | NS | Netherlands | 2022-Jul-22 | EPI_ISL_14233924 | A/Common Tern/Netherlands/19/2022 | Erasmus Medical Center | Erasmus Medical Center | |
| EPI2119211 | MP | Netherlands | 2022-Jul-22 | EPI_ISL_14233925 | A/Common Tern/Netherlands/20/2022 | Erasmus Medical Center | Erasmus Medical Center | |
| EPI2119209 | NP | Netherlands | 2022-Jul-22 | EPI_ISL_14233925 | A/Common Tern/Netherlands/20/2022 | Erasmus Medical Center | Erasmus Medical Center | |
| EPI2119210 | NS | Netherlands | 2022-Jul-22 | EPI_ISL_14233925 | A/Common Tern/Netherlands/20/2022 | Erasmus Medical Center | Erasmus Medical Center | |
| EPI2119219 | MP | Netherlands | 2022-Jul-14 | EPI_ISL_14233926 | A/European Herring Gull/Netherlands/10/2022 | Erasmus Medical Center | Erasmus Medical Center | |
| EPI2119217 | NP | Netherlands | 2022-Jul-14 | EPI_ISL_14233926 | A/European Herring Gull/Netherlands/10/2022 | Erasmus Medical Center | Erasmus Medical Center | |
| EPI2119218 | NS | Netherlands | 2022-Jul-14 | EPI_ISL_14233926 | A/European Herring Gull/Netherlands/10/2022 | Erasmus Medical Center | Erasmus Medical Center | |
| EPI2119225 | MP | Netherlands | 2022-Jul-14 | EPI_ISL_14233927 | A/Common Tern/Netherlands/11/2022 | Erasmus Medical Center | Erasmus Medical Center | |
| EPI2119223 | NP | Netherlands | 2022-Jul-14 | EPI_ISL_14233927 | A/Common Tern/Netherlands/11/2022 | Erasmus Medical Center | Erasmus Medical Center | |
| EPI2119224 | NS | Netherlands | 2022-Jul-14 | EPI_ISL_14233927 | A/Common Tern/Netherlands/11/2022 | Erasmus Medical Center | Erasmus Medical Center | |
| EPI2119239 | MP | Netherlands | 2022-Jul-14 | EPI_ISL_14233929 | A/Common Tern/Netherlands/12/2022 | Erasmus Medical Center | Erasmus Medical Center | |
| EPI2119237 | NP | Netherlands | 2022-Jul-14 | EPI_ISL_14233929 | A/Common Tern/Netherlands/12/2022 | Erasmus Medical Center | Erasmus Medical Center | |
| EPI2119238 | NS | Netherlands | 2022-Jul-14 | EPI_ISL_14233929 | A/Common Tern/Netherlands/12/2022 | Erasmus Medical Center | Erasmus Medical Center | |
| EPI2119244 | NP | Netherlands | 2022-Jul-14 | EPI_ISL_14233930 | A/Common Tern/Netherlands/14/2022 | Erasmus Medical Center | Erasmus Medical Center | |
| EPI2119245 | NS | Netherlands | 2022-Jul-14 | EPI_ISL_14233930 | A/Common Tern/Netherlands/14/2022 | Erasmus Medical Center | Erasmus Medical Center | |
| EPI2119246 | MP | Netherlands | 2022-Jul-14 | EPI_ISL_14233930 | A/Common Tern/Netherlands/14/2022 | Erasmus Medical Center | Erasmus Medical Center | |
| EPI2119254 | MP | Netherlands | 2022-Jul-14 | EPI_ISL_14233931 | A/Common Tern/Netherlands/15/2022 | Erasmus Medical Center | Erasmus Medical Center | |
| EPI2119252 | NP | Netherlands | 2022-Jul-14 | EPI_ISL_14233931 | A/Common Tern/Netherlands/15/2022 | Erasmus Medical Center | Erasmus Medical Center | |
| EPI2119253 | NS | Netherlands | 2022-Jul-14 | EPI_ISL_14233931 | A/Common Tern/Netherlands/15/2022 | Erasmus Medical Center | Erasmus Medical Center | |
| EPI2119262 | MP | Netherlands | 2022-Jul-14 | EPI_ISL_14233932 | A/Common Tern/Netherlands/16/2022 | Erasmus Medical Center | Erasmus Medical Center | |
| EPI2119260 | NP | Netherlands | 2022-Jul-14 | EPI_ISL_14233932 | A/Common Tern/Netherlands/16/2022 | Erasmus Medical Center | Erasmus Medical Center | |
| EPI2119261 | NS | Netherlands | 2022-Jul-14 | EPI_ISL_14233932 | A/Common Tern/Netherlands/16/2022 | Erasmus Medical Center | Erasmus Medical Center | |
| EPI2119270 | MP | Netherlands | 2022-Jul-15 | EPI_ISL_14233944 | A/Caspian Gull/Netherlands/6/2022 | Erasmus Medical Center | Erasmus Medical Center | |
| EPI2119268 | NP | Netherlands | 2022-Jul-15 | EPI_ISL_14233944 | A/Caspian Gull/Netherlands/6/2022 | Erasmus Medical Center | Erasmus Medical Center | |
| EPI2119269 | NS | Netherlands | 2022-Jul-15 | EPI_ISL_14233944 | A/Caspian Gull/Netherlands/6/2022 | Erasmus Medical Center | Erasmus Medical Center | |
| EPI2122828 | MP | Belgium | 2022-Jan-06 | EPI_ISL_14389133 | A/Larus_argentatus/Belgium/595_0008/2022 | Sciensano - Animal Infectious Diseases | Sciensano, Department of Animal Infectious Diseases | Van Borm, Steven; Vandenbussche, Frank; Roupie, Virginie; Lambrecht, Benedicte; Steensels, Mieke |
| EPI2122829 | NS | Belgium | 2022-Jan-06 | EPI_ISL_14389133 | A/Larus_argentatus/Belgium/595_0008/2022 | Sciensano - Animal Infectious Diseases | Sciensano, Department of Animal Infectious Diseases | Van Borm, Steven; Vandenbussche, Frank; Roupie, Virginie; Lambrecht, Benedicte; Steensels, Mieke |
| EPI2122826 | NP | Belgium | 2022-Jan-06 | EPI_ISL_14389133 | A/Larus_argentatus/Belgium/595_0008/2022 | Sciensano - Animal Infectious Diseases | Sciensano, Department of Animal Infectious Diseases | Van Borm, Steven; Vandenbussche, Frank; Roupie, Virginie; Lambrecht, Benedicte; Steensels, Mieke |
| EPI2122836 | MP | Belgium | 2022-Jan-24 | EPI_ISL_14389148 | A/Tachybaptus_ruficollis/Belgium/1234_0008/2022 | Sciensano - Animal Infectious Diseases | Sciensano, Department of Animal Infectious Diseases | Van Borm, Steven; Vandenbussche, Frank; Roupie, Virginie; Lambrecht, Benedicte; Steensels, Mieke |
| EPI2122837 | NS | Belgium | 2022-Jan-24 | EPI_ISL_14389148 | A/Tachybaptus_ruficollis/Belgium/1234_0008/2022 | Sciensano - Animal Infectious Diseases | Sciensano, Department of Animal Infectious Diseases | Van Borm, Steven; Vandenbussche, Frank; Roupie, Virginie; Lambrecht, Benedicte; Steensels, Mieke |

| | | | | | | | | |
|---|---|---|---|---|---|---|---|---|
| EPI2122834 | NP | Belgium | 2022-Jan-24 | EPI_ISL_14389148 | A/Tachybaptus_ruficollis/Belgium/1234_0008/2022 | Sciensano - Animal Infectious Diseases | Sciensano, Department of Animal Infectious Diseases | Van Borm, Steven; Vandenbussche, Frank; Roupie, Virginie; Lambrecht, Benedicte; Steensels, Mieke |
| EPI2122853 | NP | Belgium | 2022-Jan-26 | EPI_ISL_14389524 | A/Anser_anser_domesticus/Belgium/1668_0016/2022 | Sciensano - Animal Infectious Diseases | Sciensano, Department of Animal Infectious Diseases | Van Borm, Steven; Vandenbussche, Frank; Roupie, Virginie; Lambrecht, Benedicte; Steensels, Mieke |
| EPI2122856 | NS | Belgium | 2022-Jan-26 | EPI_ISL_14389524 | A/Anser_anser_domesticus/Belgium/1668_0016/2022 | Sciensano - Animal Infectious Diseases | Sciensano, Department of Animal Infectious Diseases | Van Borm, Steven; Vandenbussche, Frank; Roupie, Virginie; Lambrecht, Benedicte; Steensels, Mieke |
| EPI2122855 | MP | Belgium | 2022-Jan-26 | EPI_ISL_14389524 | A/Anser_anser_domesticus/Belgium/1668_0016/2022 | Sciensano - Animal Infectious Diseases | Sciensano, Department of Animal Infectious Diseases | Van Borm, Steven; Vandenbussche, Frank; Roupie, Virginie; Lambrecht, Benedicte; Steensels, Mieke |
| EPI2122864 | NS | Belgium | 2022-Jan-21 | EPI_ISL_14390100 | A/Larus_canus/Belgium/1668_0019/2022 | Sciensano - Animal Infectious Diseases | Sciensano, Department of Animal Infectious Diseases | Van Borm, Steven; Vandenbussche, Frank; Roupie, Virginie; Lambrecht, Benedicte; Steensels, Mieke |
| EPI2122863 | MP | Belgium | 2022-Jan-21 | EPI_ISL_14390100 | A/Larus_canus/Belgium/1668_0019/2022 | Sciensano - Animal Infectious Diseases | Sciensano, Department of Animal Infectious Diseases | Van Borm, Steven; Vandenbussche, Frank; Roupie, Virginie; Lambrecht, Benedicte; Steensels, Mieke |
| EPI2122861 | NP | Belgium | 2022-Jan-21 | EPI_ISL_14390100 | A/Larus_canus/Belgium/1668_0019/2022 | Sciensano - Animal Infectious Diseases | Sciensano, Department of Animal Infectious Diseases | Van Borm, Steven; Vandenbussche, Frank; Roupie, Virginie; Lambrecht, Benedicte; Steensels, Mieke |
| EPI2122869 | NP | Belgium | 2022-Feb-02 | EPI_ISL_14390385 | A/Phalacrocorax_carbo/Belgium/1734_0002/2022 | Sciensano - Animal Infectious Diseases | Sciensano, Department of Animal Infectious Diseases | Van Borm, Steven; Vandenbussche, Frank; Roupie, Virginie; Lambrecht, Benedicte; Steensels, Mieke |
| EPI2122871 | MP | Belgium | 2022-Feb-02 | EPI_ISL_14390385 | A/Phalacrocorax_carbo/Belgium/1734_0002/2022 | Sciensano - Animal Infectious Diseases | Sciensano, Department of Animal Infectious Diseases | Van Borm, Steven; Vandenbussche, Frank; Roupie, Virginie; Lambrecht, Benedicte; Steensels, Mieke |
| EPI2122872 | NS | Belgium | 2022-Feb-02 | EPI_ISL_14390385 | A/Phalacrocorax_carbo/Belgium/1734_0002/2022 | Sciensano - Animal Infectious Diseases | Sciensano, Department of Animal Infectious Diseases | Van Borm, Steven; Vandenbussche, Frank; Roupie, Virginie; Lambrecht, Benedicte; Steensels, Mieke |
| EPI2122910 | NP | Belgium | 2022-Feb-20 | EPI_ISL_14391865 | A/Buteo_buteo/Belgium/2606_0006/2022 | Sciensano - Animal Infectious Diseases | Sciensano, Department of Animal Infectious Diseases | Van Borm, Steven; Vandenbussche, Frank; Roupie, Virginie; Lambrecht, Benedicte; Steensels, Mieke |
| EPI2122913 | NS | Belgium | 2022-Feb-20 | EPI_ISL_14391865 | A/Buteo_buteo/Belgium/2606_0006/2022 | Sciensano - Animal Infectious Diseases | Sciensano, Department of Animal Infectious Diseases | Van Borm, Steven; Vandenbussche, Frank; Roupie, Virginie; Lambrecht, Benedicte; Steensels, Mieke |
| EPI2122912 | MP | Belgium | 2022-Feb-20 | EPI_ISL_14391865 | A/Buteo_buteo/Belgium/2606_0006/2022 | Sciensano - Animal Infectious Diseases | Sciensano, Department of Animal Infectious Diseases | Van Borm, Steven; Vandenbussche, Frank; Roupie, Virginie; Lambrecht, Benedicte; Steensels, Mieke |
| EPI2122918 | NP | Belgium | 2022-Feb-17 | EPI_ISL_14392127 | A/Branta_leucopsis/Belgium/2606_0009/2022 | Sciensano - Animal Infectious Diseases | Sciensano, Department of Animal Infectious Diseases | Van Borm, Steven; Vandenbussche, Frank; Roupie, Virginie; Lambrecht, Benedicte; Steensels, Mieke |
| EPI2122920 | MP | Belgium | 2022-Feb-17 | EPI_ISL_14392127 | A/Branta_leucopsis/Belgium/2606_0009/2022 | Sciensano - Animal Infectious Diseases | Sciensano, Department of Animal Infectious Diseases | Van Borm, Steven; Vandenbussche, Frank; Roupie, Virginie; Lambrecht, Benedicte; Steensels, Mieke |
| EPI2122921 | NS | Belgium | 2022-Feb-17 | EPI_ISL_14392127 | A/Branta_leucopsis/Belgium/2606_0009/2022 | Sciensano - Animal Infectious Diseases | Sciensano, Department of Animal Infectious Diseases | Van Borm, Steven; Vandenbussche, Frank; Roupie, Virginie; Lambrecht, Benedicte; Steensels, Mieke |
| EPI2122938 | NP | Netherlands | 2022-Apr-02 | EPI_ISL_14393097 | A/Fox/Netherlands/EMC4/2022 | Erasmus Medical Center | Erasmus Medical Center | |
| EPI2122939 | NS | Netherlands | 2022-Apr-02 | EPI_ISL_14393097 | A/Fox/Netherlands/EMC4/2022 | Erasmus Medical Center | Erasmus Medical Center | |
| EPI2122940 | MP | Netherlands | 2022-Apr-02 | EPI_ISL_14393097 | A/Fox/Netherlands/EMC4/2022 | Erasmus Medical Center | Erasmus Medical Center | |
| EPI2122946 | NP | Netherlands | 2022-Feb-09 | EPI_ISL_14393115 | A/Fox/Netherlands/EMC5/2022 | Erasmus Medical Center | Erasmus Medical Center | |
| EPI2122947 | NS | Netherlands | 2022-Feb-09 | EPI_ISL_14393115 | A/Fox/Netherlands/EMC5/2022 | Erasmus Medical Center | Erasmus Medical Center | |
| EPI2122948 | MP | Netherlands | 2022-Feb-09 | EPI_ISL_14393115 | A/Fox/Netherlands/EMC5/2022 | Erasmus Medical Center | Erasmus Medical Center | |
| EPI2122954 | NS | Netherlands | 2022-Feb-15 | EPI_ISL_14393126 | A/Fox/Netherlands/EMC6/2022 | Erasmus Medical Center | Erasmus Medical Center | |
| EPI2122955 | MP | Netherlands | 2022-Feb-15 | EPI_ISL_14393126 | A/Fox/Netherlands/EMC6/2022 | Erasmus Medical Center | Erasmus Medical Center | |
| EPI2122953 | NP | Netherlands | 2022-Feb-15 | EPI_ISL_14393126 | A/Fox/Netherlands/EMC6/2022 | Erasmus Medical Center | Erasmus Medical Center | |
| EPI2122973 | NP | Belgium | 2022-Apr-07 | EPI_ISL_14393465 | A/Branta_canadensis/Belgium/4821_0001/2022 | Sciensano - Animal Infectious Diseases | Sciensano, Department of Animal Infectious Diseases | Van Borm, Steven; Vandenbussche, Frank; Roupie, Virginie; Lambrecht, Benedicte; Steensels, Mieke |
| EPI2122976 | NS | Belgium | 2022-Apr-07 | EPI_ISL_14393465 | A/Branta_canadensis/Belgium/4821_0001/2022 | Sciensano - Animal Infectious Diseases | Sciensano, Department of Animal Infectious Diseases | Van Borm, Steven; Vandenbussche, Frank; Roupie, Virginie; Lambrecht, Benedicte; Steensels, Mieke |
| EPI2122975 | MP | Belgium | 2022-Apr-07 | EPI_ISL_14393465 | A/Branta_canadensis/Belgium/4821_0001/2022 | Sciensano - Animal Infectious Diseases | Sciensano, Department of Animal Infectious Diseases | Van Borm, Steven; Vandenbussche, Frank; Roupie, Virginie; Lambrecht, Benedicte; Steensels, Mieke |
| EPI2122981 | NP | Belgium | 2022-Apr-13 | EPI_ISL_14393671 | A/Branta_canadensis/Belgium/5177_0003/2022 | Sciensano - Animal Infectious Diseases | Sciensano, Department of Animal Infectious Diseases | Van Borm, Steven; Vandenbussche, Frank; Roupie, Virginie; Lambrecht, Benedicte; Steensels, Mieke |
| EPI2122983 | MP | Belgium | 2022-Apr-13 | EPI_ISL_14393671 | A/Branta_canadensis/Belgium/5177_0003/2022 | Sciensano - Animal Infectious Diseases | Sciensano, Department of Animal Infectious Diseases | Van Borm, Steven; Vandenbussche, Frank; Roupie, Virginie; Lambrecht, Benedicte; Steensels, Mieke |
| EPI2122984 | NS | Belgium | 2022-Apr-13 | EPI_ISL_14393671 | A/Branta_canadensis/Belgium/5177_0003/2022 | Sciensano - Animal Infectious Diseases | Sciensano, Department of Animal Infectious Diseases | Van Borm, Steven; Vandenbussche, Frank; Roupie, Virginie; Lambrecht, Benedicte; Steensels, Mieke |
| EPI2128847 | NP | Netherlands | 2022-Jul-29 | EPI_ISL_14469689 | A/Common Tern/Netherlands/21/2022 | Erasmus Medical Center | Erasmus Medical Center | |
| EPI2128848 | NS | Netherlands | 2022-Jul-29 | EPI_ISL_14469689 | A/Common Tern/Netherlands/21/2022 | Erasmus Medical Center | Erasmus Medical Center | |
| EPI2128849 | MP | Netherlands | 2022-Jul-29 | EPI_ISL_14469689 | A/Common Tern/Netherlands/21/2022 | Erasmus Medical Center | Erasmus Medical Center | |
| EPI2128855 | NP | Netherlands | 2022-Jul-29 | EPI_ISL_14469690 | A/Common Tern/Netherlands/22/2022 | Erasmus Medical Center | Erasmus Medical Center | |

| | | | | | | | | |
|---|---|---|---|---|---|---|---|---|
| EPI2128856 | NS | Netherlands | 2022-Jul-29 | EPI_ISL_14469690 | A/Common Tern/Netherlands/22/2022 | Erasmus Medical Center | Erasmus Medical Center | |
| EPI2128857 | MP | Netherlands | 2022-Jul-29 | EPI_ISL_14469690 | A/Common Tern/Netherlands/22/2022 | Erasmus Medical Center | Erasmus Medical Center | |
| EPI2128863 | NP | Netherlands | 2022-Jul-29 | EPI_ISL_14469691 | A/Common Tern/Netherlands/23/2022 | Erasmus Medical Center | Erasmus Medical Center | |
| EPI2128864 | NS | Netherlands | 2022-Jul-29 | EPI_ISL_14469691 | A/Common Tern/Netherlands/23/2022 | Erasmus Medical Center | Erasmus Medical Center | |
| EPI2128865 | MP | Netherlands | 2022-Jul-29 | EPI_ISL_14469691 | A/Common Tern/Netherlands/23/2022 | Erasmus Medical Center | Erasmus Medical Center | |
| EPI2128871 | NP | Netherlands | 2022-Jul-29 | EPI_ISL_14469692 | A/Common Tern/Netherlands/24/2022 | Erasmus Medical Center | Erasmus Medical Center | |
| EPI2128872 | NS | Netherlands | 2022-Jul-29 | EPI_ISL_14469692 | A/Common Tern/Netherlands/24/2022 | Erasmus Medical Center | Erasmus Medical Center | |
| EPI2128873 | MP | Netherlands | 2022-Jul-29 | EPI_ISL_14469692 | A/Common Tern/Netherlands/24/2022 | Erasmus Medical Center | Erasmus Medical Center | |
| EPI2128879 | NP | Netherlands | 2022-Jul-30 | EPI_ISL_14469693 | A/Common Tern/Netherlands/25/2022 | Erasmus Medical Center | Erasmus Medical Center | |
| EPI2128880 | NS | Netherlands | 2022-Jul-30 | EPI_ISL_14469693 | A/Common Tern/Netherlands/25/2022 | Erasmus Medical Center | Erasmus Medical Center | |
| EPI2128881 | MP | Netherlands | 2022-Jul-30 | EPI_ISL_14469693 | A/Common Tern/Netherlands/25/2022 | Erasmus Medical Center | Erasmus Medical Center | |
| EPI2129741 | NP | Netherlands | 2022-Jun-03 | EPI_ISL_14493135 | A/white stork/Netherlands/22012595-002/2022 | Wageningen Bioveterinary Research | Wageningen Bioveterinary Research | Beerens, Nancy; Harders, Frank; Pritz-Verschuren, Sylvia; Roose, Marit; Venema, Sandra; Germeraad, Evelien; Engelsma, Marc; Heutink, Rene |
| EPI2129742 | NS | Netherlands | 2022-Jun-03 | EPI_ISL_14493135 | A/white stork/Netherlands/22012595-002/2022 | Wageningen Bioveterinary Research | Wageningen Bioveterinary Research | Beerens, Nancy; Harders, Frank; Pritz-Verschuren, Sylvia; Roose, Marit; Venema, Sandra; Germeraad, Evelien; Engelsma, Marc; Heutink, Rene |
| EPI2129743 | MP | Netherlands | 2022-Jun-03 | EPI_ISL_14493135 | A/white stork/Netherlands/22012595-002/2022 | Wageningen Bioveterinary Research | Wageningen Bioveterinary Research | Beerens, Nancy; Harders, Frank; Pritz-Verschuren, Sylvia; Roose, Marit; Venema, Sandra; Germeraad, Evelien; Engelsma, Marc; Heutink, Rene |
| EPI2129817 | NP | Belgium | 2022-Apr-16 | EPI_ISL_14493899 | A/Vulpes_vulpes/Belgium/8660_0016/2022 | Sciensano - Animal Infectious Diseases | Sciensano, Department of Animal Infectious Diseases | Van Borm, Steven; Mathijs, Elisabeth; Roupie, Virginie; Vervaeke, Muriel; Lambrecht, Benedicte; Steensels, Mieke |
| EPI2129819 | MP | Belgium | 2022-Apr-16 | EPI_ISL_14493899 | A/Vulpes_vulpes/Belgium/8660_0016/2022 | Sciensano - Animal Infectious Diseases | Sciensano, Department of Animal Infectious Diseases | Van Borm, Steven; Mathijs, Elisabeth; Roupie, Virginie; Vervaeke, Muriel; Lambrecht, Benedicte; Steensels, Mieke |
| EPI2129820 | NS | Belgium | 2022-Apr-16 | EPI_ISL_14493899 | A/Vulpes_vulpes/Belgium/8660_0016/2022 | Sciensano - Animal Infectious Diseases | Sciensano, Department of Animal Infectious Diseases | Van Borm, Steven; Mathijs, Elisabeth; Roupie, Virginie; Vervaeke, Muriel; Lambrecht, Benedicte; Steensels, Mieke |
| EPI2129828 | NS | Belgium | 2022-Jul-08 | EPI_ISL_14494351 | A/Larus_argentatus/Belgium/9013_0001/2022 | Sciensano - Animal Infectious Diseases | Sciensano, Department of Animal Infectious Diseases | Van Borm, Steven; Mathijs, Elisabeth; Roupie, Virginie; Lambrecht, Benedicte; Steensels, Mieke |
| EPI2129825 | NP | Belgium | 2022-Jul-08 | EPI_ISL_14494351 | A/Larus_argentatus/Belgium/9013_0001/2022 | Sciensano - Animal Infectious Diseases | Sciensano, Department of Animal Infectious Diseases | Van Borm, Steven; Mathijs, Elisabeth; Roupie, Virginie; Lambrecht, Benedicte; Steensels, Mieke |
| EPI2129827 | MP | Belgium | 2022-Jul-08 | EPI_ISL_14494351 | A/Larus_argentatus/Belgium/9013_0001/2022 | Sciensano - Animal Infectious Diseases | Sciensano, Department of Animal Infectious Diseases | Van Borm, Steven; Mathijs, Elisabeth; Roupie, Virginie; Lambrecht, Benedicte; Steensels, Mieke |
| EPI2129835 | MP | Belgium | 2022-Jun-16 | EPI_ISL_14494796 | A/Vulpes_vulpes/Belgium/9031_0008/2022 | Sciensano - Animal Infectious Diseases | Sciensano, Department of Animal Infectious Diseases | Van Borm, Steven; Mathijs, Elisabeth; Roupie, Virginie; Vervaeke, Muriel; Lambrecht, Benedicte; Steensels, Mieke |
| EPI2129836 | NS | Belgium | 2022-Jun-16 | EPI_ISL_14494796 | A/Vulpes_vulpes/Belgium/9031_0008/2022 | Sciensano - Animal Infectious Diseases | Sciensano, Department of Animal Infectious Diseases | Van Borm, Steven; Mathijs, Elisabeth; Roupie, Virginie; Vervaeke, Muriel; Lambrecht, Benedicte; Steensels, Mieke |
| EPI2129833 | NP | Belgium | 2022-Jun-16 | EPI_ISL_14494796 | A/Vulpes_vulpes/Belgium/9031_0008/2022 | Sciensano - Animal Infectious Diseases | Sciensano, Department of Animal Infectious Diseases | Van Borm, Steven; Mathijs, Elisabeth; Roupie, Virginie; Vervaeke, Muriel; Lambrecht, Benedicte; Steensels, Mieke |
| EPI2129843 | MP | Belgium | 2022-Aug-03 | EPI_ISL_14494933 | A/Gallus_gallus/Belgium/9548_0001/2022 | Sciensano - Animal Infectious Diseases | Sciensano, Department of Animal Infectious Diseases | Van Borm, Steven; Mathijs, Elisabeth; Roupie, Virginie; Lambrecht, Benedicte; Steensels, Mieke |
| EPI2129844 | NS | Belgium | 2022-Aug-03 | EPI_ISL_14494933 | A/Gallus_gallus/Belgium/9548_0001/2022 | Sciensano - Animal Infectious Diseases | Sciensano, Department of Animal Infectious Diseases | Van Borm, Steven; Mathijs, Elisabeth; Roupie, Virginie; Lambrecht, Benedicte; Steensels, Mieke |
| EPI2129841 | NP | Belgium | 2022-Aug-03 | EPI_ISL_14494933 | A/Gallus_gallus/Belgium/9548_0001/2022 | Sciensano - Animal Infectious Diseases | Sciensano, Department of Animal Infectious Diseases | Van Borm, Steven; Mathijs, Elisabeth; Roupie, Virginie; Lambrecht, Benedicte; Steensels, Mieke |
| EPI2129856 | NP | Netherlands | 2022-Aug-10 | EPI_ISL_14497321 | A/Eurasian Spoonbill/Netherlands/4/2022 | Erasmus Medical Center | Erasmus Medical Center | |
| EPI2129857 | NS | Netherlands | 2022-Aug-10 | EPI_ISL_14497321 | A/Eurasian Spoonbill/Netherlands/4/2022 | Erasmus Medical Center | Erasmus Medical Center | |
| EPI2129858 | MP | Netherlands | 2022-Aug-10 | EPI_ISL_14497321 | A/Eurasian Spoonbill/Netherlands/4/2022 | Erasmus Medical Center | Erasmus Medical Center | |
| EPI2129879 | NP | United Kingdom | 2022-Jul-26 | EPI_ISL_14497837 | A/domestic_duck/England/100990/2022 | Animal and Plant Health Agency (APHA) | Animal and Plant Health Agency (APHA) | |
| EPI2129880 | NS | United Kingdom | 2022-Jul-26 | EPI_ISL_14497837 | A/domestic_duck/England/100990/2022 | Animal and Plant Health Agency (APHA) | Animal and Plant Health Agency (APHA) | |
| EPI2129881 | MP | United Kingdom | 2022-Jul-26 | EPI_ISL_14497837 | A/domestic_duck/England/100990/2022 | Animal and Plant Health Agency (APHA) | Animal and Plant Health Agency (APHA) | |
| EPI2129896 | NP | United Kingdom | 2022-Aug-05 | EPI_ISL_14497855 | A/domestic_duck/England/104859/2022 | Animal and Plant Health Agency (APHA) | Animal and Plant Health Agency (APHA) | |
| EPI2129897 | NS | United Kingdom | 2022-Aug-05 | EPI_ISL_14497855 | A/domestic_duck/England/104859/2022 | Animal and Plant Health Agency (APHA) | Animal and Plant Health Agency (APHA) | |
| EPI2129898 | MP | United Kingdom | 2022-Aug-05 | EPI_ISL_14497855 | A/domestic_duck/England/104859/2022 | Animal and Plant Health Agency (APHA) | Animal and Plant Health Agency (APHA) | |

| | | | | | | | |
|---|---|---|---|---|---|---|---|
| EPI2129904 | NP | United Kingdom | 2022-Aug-07 | EPI_ISL_14497870 | A/domestic_duck/England/105414/2022 | Animal and Plant Health Agency (APHA) | Animal and Plant Health Agency (APHA) |
| EPI2129905 | NS | United Kingdom | 2022-Aug-07 | EPI_ISL_14497870 | A/domestic_duck/England/105414/2022 | Animal and Plant Health Agency (APHA) | Animal and Plant Health Agency (APHA) |
| EPI2129906 | MP | United Kingdom | 2022-Aug-07 | EPI_ISL_14497870 | A/domestic_duck/England/105414/2022 | Animal and Plant Health Agency (APHA) | Animal and Plant Health Agency (APHA) |
| EPI2129912 | NP | United Kingdom | 2022-Aug-09 | EPI_ISL_14497885 | A/domestic_goose/England/105863/2022 | Animal and Plant Health Agency (APHA) | Animal and Plant Health Agency (APHA) |
| EPI2129913 | NS | United Kingdom | 2022-Aug-09 | EPI_ISL_14497885 | A/domestic_goose/England/105863/2022 | Animal and Plant Health Agency (APHA) | Animal and Plant Health Agency (APHA) |
| EPI2129914 | MP | United Kingdom | 2022-Aug-09 | EPI_ISL_14497885 | A/domestic_goose/England/105863/2022 | Animal and Plant Health Agency (APHA) | Animal and Plant Health Agency (APHA) |
| EPI2131804 | NP | Germany | 2022-Jun-07 | EPI_ISL_14571430 | A/common tern/Germany-NI/AI03185/2022 | Lebensmittel- und Veterinärinstitut Oldenburg - Standort Veterinärinstitut | Friedrich-Loeffler-Institut |
| EPI2131805 | NS | Germany | 2022-Jun-07 | EPI_ISL_14571430 | A/common tern/Germany-NI/AI03185/2022 | Lebensmittel- und Veterinärinstitut Oldenburg - Standort Veterinärinstitut | Friedrich-Loeffler-Institut |
| EPI2131802 | MP | Germany | 2022-Jun-07 | EPI_ISL_14571430 | A/common tern/Germany-NI/AI03185/2022 | Lebensmittel- und Veterinärinstitut Oldenburg - Standort Veterinärinstitut | Friedrich-Loeffler-Institut |
| EPI2131811 | MP | Germany | 2022-Jun-07 | EPI_ISL_14571432 | A/common tern/Germany-NI/AI03186/2022 | Lebensmittel- und Veterinärinstitut Oldenburg - Standort Veterinärinstitut | Friedrich-Loeffler-Institut |
| EPI2131813 | NP | Germany | 2022-Jun-07 | EPI_ISL_14571432 | A/common tern/Germany-NI/AI03186/2022 | Lebensmittel- und Veterinärinstitut Oldenburg - Standort Veterinärinstitut | Friedrich-Loeffler-Institut |
| EPI2131815 | NS | Germany | 2022-Jun-07 | EPI_ISL_14571432 | A/common tern/Germany-NI/AI03186/2022 | Lebensmittel- und Veterinärinstitut Oldenburg - Standort Veterinärinstitut | Friedrich-Loeffler-Institut |
| EPI2131825 | MP | Germany | 2022-Jun-16 | EPI_ISL_14571434 | A/common tern/Germany-NI/AI03512/2022 | Lebensmittel- und Veterinärinstitut Oldenburg - Standort Veterinärinstitut | Friedrich-Loeffler-Institut |
| EPI2131828 | NP | Germany | 2022-Jun-16 | EPI_ISL_14571434 | A/common tern/Germany-NI/AI03512/2022 | Lebensmittel- und Veterinärinstitut Oldenburg - Standort Veterinärinstitut | Friedrich-Loeffler-Institut |
| EPI2131829 | NS | Germany | 2022-Jun-16 | EPI_ISL_14571434 | A/common tern/Germany-NI/AI03512/2022 | Lebensmittel- und Veterinärinstitut Oldenburg - Standort Veterinärinstitut | Friedrich-Loeffler-Institut |
| EPI2131834 | MP | Germany | 2022-Jun-14 | EPI_ISL_14571436 | A/common tern/Germany-NI/AI03520/2022 | Lebensmittel- und Veterinärinstitut Oldenburg - Standort Veterinärinstitut | Friedrich-Loeffler-Institut |
| EPI2131836 | NP | Germany | 2022-Jun-14 | EPI_ISL_14571436 | A/common tern/Germany-NI/AI03520/2022 | Lebensmittel- und Veterinärinstitut Oldenburg - Standort Veterinärinstitut | Friedrich-Loeffler-Institut |
| EPI2131837 | NS | Germany | 2022-Jun-14 | EPI_ISL_14571436 | A/common tern/Germany-NI/AI03520/2022 | Lebensmittel- und Veterinärinstitut Oldenburg - Standort Veterinärinstitut | Friedrich-Loeffler-Institut |
| EPI2131849 | MP | Germany | 2022-Jun-14 | EPI_ISL_14571438 | A/common tern/Germany-NI/AI03523/2022 | Lebensmittel- und Veterinärinstitut Oldenburg - Standort Veterinärinstitut | Friedrich-Loeffler-Institut |
| EPI2131852 | NP | Germany | 2022-Jun-14 | EPI_ISL_14571438 | A/common tern/Germany-NI/AI03523/2022 | Lebensmittel- und Veterinärinstitut Oldenburg - Standort Veterinärinstitut | Friedrich-Loeffler-Institut |
| EPI2131853 | NS | Germany | 2022-Jun-14 | EPI_ISL_14571438 | A/common tern/Germany-NI/AI03523/2022 | Lebensmittel- und Veterinärinstitut Oldenburg - Standort Veterinärinstitut | Friedrich-Loeffler-Institut |
| EPI2131898 | MP | Germany | 2022-May-03 | EPI_ISL_14571448 | A/great cormorant/Germany-MV/AI02666/2022 | Landesamt für Landwirtschaft, Lebensmittelsicherheit und Fischerei (LALLF) | Friedrich-Loeffler-Institut |
| EPI2131900 | NP | Germany | 2022-May-03 | EPI_ISL_14571448 | A/great cormorant/Germany-MV/AI02666/2022 | Landesamt für Landwirtschaft, Lebensmittelsicherheit und Fischerei (LALLF) | Friedrich-Loeffler-Institut |
| EPI2131901 | NS | Germany | 2022-May-03 | EPI_ISL_14571448 | A/great cormorant/Germany-MV/AI02666/2022 | Landesamt für Landwirtschaft, Lebensmittelsicherheit und Fischerei (LALLF) | Friedrich-Loeffler-Institut |
| EPI2131908 | NP | Germany | 2022-May-06 | EPI_ISL_14571449 | A/great cormorant/Germany-MV/AI02774/2022 | Landesamt für Landwirtschaft, Lebensmittelsicherheit und Fischerei (LALLF) | Friedrich-Loeffler-Institut |
| EPI2131906 | MP | Germany | 2022-May-06 | EPI_ISL_14571449 | A/great cormorant/Germany-MV/AI02774/2022 | Landesamt für Landwirtschaft, Lebensmittelsicherheit und Fischerei (LALLF) | Friedrich-Loeffler-Institut |
| EPI2131909 | NS | Germany | 2022-May-06 | EPI_ISL_14571449 | A/great cormorant/Germany-MV/AI02774/2022 | Landesamt für Landwirtschaft, Lebensmittelsicherheit und Fischerei (LALLF) | Friedrich-Loeffler-Institut |
| EPI2132555 | NS | Germany | 2022-Jun-16 | EPI_ISL_14573001 | A/northern gannet/Germany-SH/AI03496/2022 | Landeslabor Schleswig-Holstein | Friedrich-Loeffler-Institut |
| EPI2132554 | NP | Germany | 2022-Jun-16 | EPI_ISL_14573001 | A/northern gannet/Germany-SH/AI03496/2022 | Landeslabor Schleswig-Holstein | Friedrich-Loeffler-Institut |
| EPI2132551 | MP | Germany | 2022-Jun-16 | EPI_ISL_14573001 | A/northern gannet/Germany-SH/AI03496/2022 | Landeslabor Schleswig-Holstein | Friedrich-Loeffler-Institut |
| EPI2132569 | NS | Germany | 2022-Jun-15 | EPI_ISL_14573031 | A/northern gannet/Germany-SH/AI03498/2022 | Landeslabor Schleswig-Holstein | Friedrich-Loeffler-Institut |
| EPI2132565 | MP | Germany | 2022-Jun-15 | EPI_ISL_14573031 | A/northern gannet/Germany-SH/AI03498/2022 | Landeslabor Schleswig-Holstein | Friedrich-Loeffler-Institut |
| EPI2132568 | NP | Germany | 2022-Jun-15 | EPI_ISL_14573031 | A/northern gannet/Germany-SH/AI03498/2022 | Landeslabor Schleswig-Holstein | Friedrich-Loeffler-Institut |
| EPI2132610 | NP | Germany | 2022-May-31 | EPI_ISL_14573037 | A/sandwich tern/Germany-MV/AI03131/2022 | Landesamt für Landwirtschaft, Lebensmittelsicherheit und Fischerei (LALLF) | Friedrich-Loeffler-Institut |
| EPI2132611 | NS | Germany | 2022-May-31 | EPI_ISL_14573037 | A/sandwich tern/Germany-MV/AI03131/2022 | Landesamt für Landwirtschaft, Lebensmittelsicherheit und Fischerei (LALLF) | Friedrich-Loeffler-Institut |

| | | | | | | | |
|---|---|---|---|---|---|---|---|
| EPI2132608 | MP | Germany | 2022-May-31 | EPI_ISL_14573037 | A/sandwich tern/Germany-MV/AI03131/2022 | Landesamt für Landwirtschaft, Lebensmittelsicherheit und Fischerei (LALLF) | Friedrich-Loeffler-Institut |
| EPI2132621 | NS | Germany | 2022-Jun-02 | EPI_ISL_14573043 | A/sandwich tern/Germany-NI/AI03180/2022 | Lebensmittel- und Veterinärinstitut Oldenburg - Standort Veterinärinstitut | Friedrich-Loeffler-Institut |
| EPI2132617 | MP | Germany | 2022-Jun-02 | EPI_ISL_14573043 | A/sandwich tern/Germany-NI/AI03180/2022 | Lebensmittel- und Veterinärinstitut Oldenburg - Standort Veterinärinstitut | Friedrich-Loeffler-Institut |
| EPI2132619 | NP | Germany | 2022-Jun-02 | EPI_ISL_14573043 | A/sandwich tern/Germany-NI/AI03180/2022 | Lebensmittel- und Veterinärinstitut Oldenburg - Standort Veterinärinstitut | Friedrich-Loeffler-Institut |
| EPI2132633 | NS | Germany | 2022-Jun-02 | EPI_ISL_14573044 | A/sandwich tern/Germany-NI/AI03181/2022 | Lebensmittel- und Veterinärinstitut Oldenburg - Standort Veterinärinstitut | Friedrich-Loeffler-Institut |
| EPI2132629 | MP | Germany | 2022-Jun-02 | EPI_ISL_14573044 | A/sandwich tern/Germany-NI/AI03181/2022 | Lebensmittel- und Veterinärinstitut Oldenburg - Standort Veterinärinstitut | Friedrich-Loeffler-Institut |
| EPI2132632 | NP | Germany | 2022-Jun-02 | EPI_ISL_14573044 | A/sandwich tern/Germany-NI/AI03181/2022 | Lebensmittel- und Veterinärinstitut Oldenburg - Standort Veterinärinstitut | Friedrich-Loeffler-Institut |
| EPI2132640 | MP | Germany | 2022-Jun-07 | EPI_ISL_14573045 | A/sandwich tern/Germany-SH/AI03333/2022 | Landeslabor Schleswig-Holstein | Friedrich-Loeffler-Institut |
| EPI2132642 | NP | Germany | 2022-Jun-07 | EPI_ISL_14573045 | A/sandwich tern/Germany-SH/AI03333/2022 | Landeslabor Schleswig-Holstein | Friedrich-Loeffler-Institut |
| EPI2132643 | NS | Germany | 2022-Jun-07 | EPI_ISL_14573045 | A/sandwich tern/Germany-SH/AI03333/2022 | Landeslabor Schleswig-Holstein | Friedrich-Loeffler-Institut |
| EPI2132648 | MP | Germany | 2022-Jun-07 | EPI_ISL_14573052 | A/sandwich tern/Germany-SH/AI03335/2022 | Landeslabor Schleswig-Holstein | Friedrich-Loeffler-Institut |
| EPI2132651 | NP | Germany | 2022-Jun-07 | EPI_ISL_14573052 | A/sandwich tern/Germany-SH/AI03335/2022 | Landeslabor Schleswig-Holstein | Friedrich-Loeffler-Institut |
| EPI2132652 | NS | Germany | 2022-Jun-07 | EPI_ISL_14573052 | A/sandwich tern/Germany-SH/AI03335/2022 | Landeslabor Schleswig-Holstein | Friedrich-Loeffler-Institut |
| EPI2132662 | MP | Germany | 2022-Jun-09 | EPI_ISL_14573054 | A/sandwich tern/Germany-SH/AI03497/2022 | Landeslabor Schleswig-Holstein | Friedrich-Loeffler-Institut |
| EPI2132665 | NP | Germany | 2022-Jun-09 | EPI_ISL_14573054 | A/sandwich tern/Germany-SH/AI03497/2022 | Landeslabor Schleswig-Holstein | Friedrich-Loeffler-Institut |
| EPI2132666 | NS | Germany | 2022-Jun-09 | EPI_ISL_14573054 | A/sandwich tern/Germany-SH/AI03497/2022 | Landeslabor Schleswig-Holstein | Friedrich-Loeffler-Institut |
| EPI2134873 | NS | France | 2022-Jul-13 | EPI_ISL_14702899 | A/nothern_gannet/France/22P019331/2022 | Anses (Ploufragan-Plouzané) | ANSES Agence Nationale De Securite Sanitaire De L'alimentation |
| EPI2134872 | MP | France | 2022-Jul-13 | EPI_ISL_14702899 | A/nothern_gannet/France/22P019331/2022 | Anses (Ploufragan-Plouzané) | ANSES Agence Nationale De Securite Sanitaire De L'alimentation |
| EPI2134870 | NP | France | 2022-Jul-13 | EPI_ISL_14702899 | A/nothern_gannet/France/22P019331/2022 | Anses (Ploufragan-Plouzané) | ANSES Agence Nationale De Securite Sanitaire De L'alimentation |
| EPI2136048 | NP | France | 2022-May-09 | EPI_ISL_14722960 | A/vulture/France/22P018210/2022 | Anses (Ploufragan-Plouzané) | ANSES Agence Nationale De Securite Sanitaire De L'alimentation |
| EPI2136050 | MP | France | 2022-May-09 | EPI_ISL_14722960 | A/vulture/France/22P018210/2022 | Anses (Ploufragan-Plouzané) | ANSES Agence Nationale De Securite Sanitaire De L'alimentation |
| EPI2136051 | NS | France | 2022-May-09 | EPI_ISL_14722960 | A/vulture/France/22P018210/2022 | Anses (Ploufragan-Plouzané) | ANSES Agence Nationale De Securite Sanitaire De L'alimentation |
| EPI2150628 | NS | Sweden | 2022-Jun-29 | EPI_ISL_14810369 | A/porpoise /Sweden/SVA220712SZ0367/FB002184/O-2022 | National Veterinary Institute, SVA | National Veterinary Institute |
| EPI2150625 | NP | Sweden | 2022-Jun-29 | EPI_ISL_14810369 | A/porpoise /Sweden/SVA220712SZ0367/FB002184/O-2022 | National Veterinary Institute, SVA | National Veterinary Institute |
| EPI2150627 | MP | Sweden | 2022-Jun-29 | EPI_ISL_14810369 | A/porpoise /Sweden/SVA220712SZ0367/FB002184/O-2022 | National Veterinary Institute, SVA | National Veterinary Institute |
| EPI2150865 | MP | Sweden | 2022-Jun-20 | EPI_ISL_14811896 | A/northern gannet/Sweden/SVA220623SZ0232/FB001982/O-2022 | National Veterinary Institute, SVA | National Veterinary Institute |
| EPI2150866 | NS | Sweden | 2022-Jun-20 | EPI_ISL_14811896 | A/northern gannet/Sweden/SVA220623SZ0232/FB001982/O-2022 | National Veterinary Institute, SVA | National Veterinary Institute |
| EPI2150863 | NP | Sweden | 2022-Jun-20 | EPI_ISL_14811896 | A/northern gannet/Sweden/SVA220623SZ0232/FB001982/O-2022 | National Veterinary Institute, SVA | National Veterinary Institute |
| EPI2150871 | NP | Sweden | 2022-Jun-15 | EPI_ISL_14811929 | A/northern gannet/Sweden/SVA220623SZ0232/FB001984/N-2022 | National Veterinary Institute, SVA | National Veterinary Institute |
| EPI2150873 | MP | Sweden | 2022-Jun-15 | EPI_ISL_14811929 | A/northern gannet/Sweden/SVA220623SZ0232/FB001984/N-2022 | National Veterinary Institute, SVA | National Veterinary Institute |
| EPI2150874 | NS | Sweden | 2022-Jun-15 | EPI_ISL_14811929 | A/northern gannet/Sweden/SVA220623SZ0232/FB001984/N-2022 | National Veterinary Institute, SVA | National Veterinary Institute |
| EPI2154997 | MP | France | 2022-Jul-06 | EPI_ISL_14841913 | A/mallard/22P019377/France/2022 | Anses (Ploufragan-Plouzané) | ANSES Agence Nationale De Securite Sanitaire De L'alimentation |
| EPI2154998 | NS | France | 2022-Jul-06 | EPI_ISL_14841913 | A/mallard/22P019377/France/2022 | Anses (Ploufragan-Plouzané) | ANSES Agence Nationale De Securite Sanitaire De L'alimentation |
| EPI2154995 | NP | France | 2022-Jul-06 | EPI_ISL_14841913 | A/mallard/22P019377/France/2022 | Anses (Ploufragan-Plouzané) | ANSES Agence Nationale De Securite Sanitaire De L'alimentation |
| EPI2158609 | NP | United Kingdom | 2022-Mar-09 | EPI_ISL_14864638 | A/fox/England/M0083-03-22/2022 | Animal and Plant Health Agency (APHA) | Animal and Plant Health Agency (APHA) |

| | | | | | | | | |
|---|---|---|---|---|---|---|---|---|
| EPI2158610 | NS | United Kingdom | 2022-Mar-09 | EPI_ISL_14864638 | A/fox/England/M0083-03-22/2022 | Animal and Plant Health Agency (APHA) | Animal and Plant Health Agency (APHA) | |
| EPI2158611 | MP | United Kingdom | 2022-Mar-09 | EPI_ISL_14864638 | A/fox/England/M0083-03-22/2022 | Animal and Plant Health Agency (APHA) | Animal and Plant Health Agency (APHA) | |
| EPI2158633 | NP | United Kingdom | 2022-Jan-01 | EPI_ISL_14864641 | A/otter/Scotland/M100839/2022 | Animal and Plant Health Agency (APHA) | Animal and Plant Health Agency (APHA) | |
| EPI2158634 | NS | United Kingdom | 2022-Jan-01 | EPI_ISL_14864641 | A/otter/Scotland/M100839/2022 | Animal and Plant Health Agency (APHA) | Animal and Plant Health Agency (APHA) | |
| EPI2158635 | MP | United Kingdom | 2022-Jan-01 | EPI_ISL_14864641 | A/otter/Scotland/M100839/2022 | Animal and Plant Health Agency (APHA) | Animal and Plant Health Agency (APHA) | |
| EPI2158641 | NP | United Kingdom | 2022-Jan-01 | EPI_ISL_14864642 | A/otter/Scotland/M100366/2022 | Animal and Plant Health Agency (APHA) | Animal and Plant Health Agency (APHA) | |
| EPI2158642 | NS | United Kingdom | 2022-Jan-01 | EPI_ISL_14864642 | A/otter/Scotland/M100366/2022 | Animal and Plant Health Agency (APHA) | Animal and Plant Health Agency (APHA) | |
| EPI2158643 | MP | United Kingdom | 2022-Jan-01 | EPI_ISL_14864642 | A/otter/Scotland/M100366/2022 | Animal and Plant Health Agency (APHA) | Animal and Plant Health Agency (APHA) | |
| EPI2158705 | NP | United Kingdom | 2022-Aug-26 | EPI_ISL_14864650 | A/turkey/England/111923/2022 | Animal and Plant Health Agency (APHA) | Animal and Plant Health Agency (APHA) | |
| EPI2158706 | NS | United Kingdom | 2022-Aug-26 | EPI_ISL_14864650 | A/turkey/England/111923/2022 | Animal and Plant Health Agency (APHA) | Animal and Plant Health Agency (APHA) | |
| EPI2158707 | MP | United Kingdom | 2022-Aug-26 | EPI_ISL_14864650 | A/turkey/England/111923/2022 | Animal and Plant Health Agency (APHA) | Animal and Plant Health Agency (APHA) | |
| EPI2158721 | NP | United Kingdom | 2022-Feb-01 | EPI_ISL_14864652 | A/long-tailed_skua/Scotland/22/2022 | Animal and Plant Health Agency (APHA) | Animal and Plant Health Agency (APHA) | |
| EPI2158722 | NS | United Kingdom | 2022-Feb-01 | EPI_ISL_14864652 | A/long-tailed_skua/Scotland/22/2022 | Animal and Plant Health Agency (APHA) | Animal and Plant Health Agency (APHA) | |
| EPI2158723 | MP | United Kingdom | 2022-Feb-01 | EPI_ISL_14864652 | A/long-tailed_skua/Scotland/22/2022 | Animal and Plant Health Agency (APHA) | Animal and Plant Health Agency (APHA) | |
| EPI2167212 | NP | Poland | 2022-Jul-13 | EPI_ISL_14917968 | A/common_murre/Poland/MB151/2022 | National Veterinary Research Institut Poland, PIWet-PIB | National Veterinary Research Institut Poland, PIWet-PIB | Swieton E., Smietanka K. |
| EPI2167217 | NS | Poland | 2022-Jul-13 | EPI_ISL_14917968 | A/common_murre/Poland/MB151/2022 | National Veterinary Research Institut Poland, PIWet-PIB | National Veterinary Research Institut Poland, PIWet-PIB | Swieton E., Smietanka K. |
| EPI2167215 | MP | Poland | 2022-Jul-13 | EPI_ISL_14917968 | A/common_murre/Poland/MB151/2022 | National Veterinary Research Institut Poland, PIWet-PIB | National Veterinary Research Institut Poland, PIWet-PIB | Swieton E., Smietanka K. |
| EPI2167225 | NS | Poland | 2022-May-30 | EPI_ISL_14917979 | A/herring_gull/Poland/MB138/2022 | National Veterinary Research Institut Poland, PIWet-PIB | National Veterinary Research Institut Poland, PIWet-PIB | Swieton E., Smietanka K. |
| EPI2167220 | NP | Poland | 2022-May-30 | EPI_ISL_14917979 | A/herring_gull/Poland/MB138/2022 | National Veterinary Research Institut Poland, PIWet-PIB | National Veterinary Research Institut Poland, PIWet-PIB | Swieton E., Smietanka K. |
| EPI2167223 | MP | Poland | 2022-May-30 | EPI_ISL_14917979 | A/herring_gull/Poland/MB138/2022 | National Veterinary Research Institut Poland, PIWet-PIB | National Veterinary Research Institut Poland, PIWet-PIB | Swieton E., Smietanka K. |
| EPI2167233 | NS | Poland | 2022-May-30 | EPI_ISL_14917999 | A/black-headed_gull/Poland/MB139/2022 | National Veterinary Research Institut Poland, PIWet-PIB | National Veterinary Research Institut Poland, PIWet-PIB | Swieton E., Smietanka K. |
| EPI2167228 | NP | Poland | 2022-May-30 | EPI_ISL_14917999 | A/black-headed_gull/Poland/MB139/2022 | National Veterinary Research Institut Poland, PIWet-PIB | National Veterinary Research Institut Poland, PIWet-PIB | Swieton E., Smietanka K. |
| EPI2167231 | MP | Poland | 2022-May-30 | EPI_ISL_14917999 | A/black-headed_gull/Poland/MB139/2022 | National Veterinary Research Institut Poland, PIWet-PIB | National Veterinary Research Institut Poland, PIWet-PIB | Swieton E., Smietanka K. |
| EPI2176011 | MP | France | 2022-Aug-24 | EPI_ISL_14933724 | A/duck/France/22P020165/2022 | Anses (Ploufragan-Plouzané) | ANSES Agence Nationale De Securite Sanitaire De L'alimentation | |
| EPI2176013 | NS | France | 2022-Aug-24 | EPI_ISL_14933724 | A/duck/France/22P020165/2022 | Anses (Ploufragan-Plouzané) | ANSES Agence Nationale De Securite Sanitaire De L'alimentation | |
| EPI2176008 | NP | France | 2022-Aug-24 | EPI_ISL_14933724 | A/duck/France/22P020165/2022 | Anses (Ploufragan-Plouzané) | ANSES Agence Nationale De Securite Sanitaire De L'alimentation | |
| EPI2181299 | MP | United Kingdom | 2022-Sep-05 | EPI_ISL_15038818 | A/pheasant/England/115311/2022 | Animal and Plant Health Agency (APHA) | Animal and Plant Health Agency (APHA) | |
| EPI2181297 | NP | United Kingdom | 2022-Sep-05 | EPI_ISL_15038818 | A/pheasant/England/115311/2022 | Animal and Plant Health Agency (APHA) | Animal and Plant Health Agency (APHA) | |
| EPI2181298 | NS | United Kingdom | 2022-Sep-05 | EPI_ISL_15038818 | A/pheasant/England/115311/2022 | Animal and Plant Health Agency (APHA) | Animal and Plant Health Agency (APHA) | |
| EPI2181305 | NP | United Kingdom | 2022-Sep-05 | EPI_ISL_15038819 | A/partridge/England/115171/2022 | Animal and Plant Health Agency (APHA) | Animal and Plant Health Agency (APHA) | |
| EPI2181307 | MP | United Kingdom | 2022-Sep-05 | EPI_ISL_15038819 | A/partridge/England/115171/2022 | Animal and Plant Health Agency (APHA) | Animal and Plant Health Agency (APHA) | |
| EPI2181306 | NS | United Kingdom | 2022-Sep-05 | EPI_ISL_15038819 | A/partridge/England/115171/2022 | Animal and Plant Health Agency (APHA) | Animal and Plant Health Agency (APHA) | |
| EPI2181313 | NP | United Kingdom | 2022-Sep-05 | EPI_ISL_15038820 | A/chicken/England/115126/2022 | Animal and Plant Health Agency (APHA) | Animal and Plant Health Agency (APHA) | |
| EPI2181314 | NS | United Kingdom | 2022-Sep-05 | EPI_ISL_15038820 | A/chicken/England/115126/2022 | Animal and Plant Health Agency (APHA) | Animal and Plant Health Agency (APHA) | |
| EPI2181315 | MP | United Kingdom | 2022-Sep-05 | EPI_ISL_15038820 | A/chicken/England/115126/2022 | Animal and Plant Health Agency (APHA) | Animal and Plant Health Agency (APHA) | |

| | | | | | | | |
|---|---|---|---|---|---|---|---|
| EPI2181330 | NS | United Kingdom | 2022-Sep-02 | EPI_ISL_15038822 | A/chicken/England/114595/2022 | Animal and Plant Health Agency (APHA) | Animal and Plant Health Agency (APHA) |
| EPI2181331 | MP | United Kingdom | 2022-Sep-02 | EPI_ISL_15038822 | A/chicken/England/114595/2022 | Animal and Plant Health Agency (APHA) | Animal and Plant Health Agency (APHA) |
| EPI2181329 | NP | United Kingdom | 2022-Sep-02 | EPI_ISL_15038822 | A/chicken/England/114595/2022 | Animal and Plant Health Agency (APHA) | Animal and Plant Health Agency (APHA) |
| EPI2181757 | NP | Netherlands | 2022-Aug-10 | EPI_ISL_15069398 | A/European Herring Gull/Netherlands/12/2022 | Erasmus Medical Center | Erasmus Medical Center |
| EPI2181758 | NS | Netherlands | 2022-Aug-10 | EPI_ISL_15069398 | A/European Herring Gull/Netherlands/12/2022 | Erasmus Medical Center | Erasmus Medical Center |
| EPI2181759 | MP | Netherlands | 2022-Aug-10 | EPI_ISL_15069398 | A/European Herring Gull/Netherlands/12/2022 | Erasmus Medical Center | Erasmus Medical Center |
| EPI2181765 | NP | Netherlands | 2022-Aug-10 | EPI_ISL_15069399 | A/Black-headed gull/Netherlands/10/2022 | Erasmus Medical Center | Erasmus Medical Center |
| EPI2181766 | NS | Netherlands | 2022-Aug-10 | EPI_ISL_15069399 | A/Black-headed gull/Netherlands/10/2022 | Erasmus Medical Center | Erasmus Medical Center |
| EPI2181767 | MP | Netherlands | 2022-Aug-10 | EPI_ISL_15069399 | A/Black-headed gull/Netherlands/10/2022 | Erasmus Medical Center | Erasmus Medical Center |
| EPI2181773 | NP | Netherlands | 2022-Aug-10 | EPI_ISL_15069400 | A/Mute Swan/Netherlands/1/2022 | Erasmus Medical Center | Erasmus Medical Center |
| EPI2181774 | NS | Netherlands | 2022-Aug-10 | EPI_ISL_15069400 | A/Mute Swan/Netherlands/1/2022 | Erasmus Medical Center | Erasmus Medical Center |
| EPI2181775 | MP | Netherlands | 2022-Aug-10 | EPI_ISL_15069400 | A/Mute Swan/Netherlands/1/2022 | Erasmus Medical Center | Erasmus Medical Center |
| EPI2181781 | NP | Netherlands | 2022-Aug-12 | EPI_ISL_15069401 | A/Common Tern/Netherlands/26/2022 | Erasmus Medical Center | Erasmus Medical Center |
| EPI2181782 | NS | Netherlands | 2022-Aug-12 | EPI_ISL_15069401 | A/Common Tern/Netherlands/26/2022 | Erasmus Medical Center | Erasmus Medical Center |
| EPI2181783 | MP | Netherlands | 2022-Aug-12 | EPI_ISL_15069401 | A/Common Tern/Netherlands/26/2022 | Erasmus Medical Center | Erasmus Medical Center |
| EPI2185345 | NP | Netherlands | 2022-Aug-17 | EPI_ISL_15088295 | A/Common Buzzard/Netherlands/1/2022 | Erasmus Medical Center | Erasmus Medical Center |
| EPI2185346 | NS | Netherlands | 2022-Aug-17 | EPI_ISL_15088295 | A/Common Buzzard/Netherlands/1/2022 | Erasmus Medical Center | Erasmus Medical Center |
| EPI2185347 | MP | Netherlands | 2022-Aug-17 | EPI_ISL_15088295 | A/Common Buzzard/Netherlands/1/2022 | Erasmus Medical Center | Erasmus Medical Center |
| EPI2185353 | NP | Netherlands | 2022-Aug-19 | EPI_ISL_15088296 | A/European Herring Gull/Netherlands/13/2022 | Erasmus Medical Center | Erasmus Medical Center |
| EPI2185354 | NS | Netherlands | 2022-Aug-19 | EPI_ISL_15088296 | A/European Herring Gull/Netherlands/13/2022 | Erasmus Medical Center | Erasmus Medical Center |
| EPI2185355 | MP | Netherlands | 2022-Aug-19 | EPI_ISL_15088296 | A/European Herring Gull/Netherlands/13/2022 | Erasmus Medical Center | Erasmus Medical Center |
| EPI2185361 | NP | Netherlands | 2022-Aug-23 | EPI_ISL_15088297 | A/European Herring Gull/Netherlands/14/2022 | Erasmus Medical Center | Erasmus Medical Center |
| EPI2185362 | NS | Netherlands | 2022-Aug-23 | EPI_ISL_15088297 | A/European Herring Gull/Netherlands/14/2022 | Erasmus Medical Center | Erasmus Medical Center |
| EPI2185363 | MP | Netherlands | 2022-Aug-23 | EPI_ISL_15088297 | A/European Herring Gull/Netherlands/14/2022 | Erasmus Medical Center | Erasmus Medical Center |
| EPI2185369 | NP | Netherlands | 2022-Aug-23 | EPI_ISL_15088298 | A/Northern Gannet/Netherlands/1/2022 | Erasmus Medical Center | Erasmus Medical Center |
| EPI2185370 | NS | Netherlands | 2022-Aug-23 | EPI_ISL_15088298 | A/Northern Gannet/Netherlands/1/2022 | Erasmus Medical Center | Erasmus Medical Center |
| EPI2185371 | MP | Netherlands | 2022-Aug-23 | EPI_ISL_15088298 | A/Northern Gannet/Netherlands/1/2022 | Erasmus Medical Center | Erasmus Medical Center |
| EPI2185377 | NP | Netherlands | 2022-Aug-26 | EPI_ISL_15088299 | A/Greylag Goose/Netherlands/10/2022 | Erasmus Medical Center | Erasmus Medical Center |
| EPI2185378 | NS | Netherlands | 2022-Aug-26 | EPI_ISL_15088299 | A/Greylag Goose/Netherlands/10/2022 | Erasmus Medical Center | Erasmus Medical Center |
| EPI2185379 | MP | Netherlands | 2022-Aug-26 | EPI_ISL_15088299 | A/Greylag Goose/Netherlands/10/2022 | Erasmus Medical Center | Erasmus Medical Center |
| EPI2185385 | NP | Netherlands | 2022-Aug-26 | EPI_ISL_15088300 | A/Greylag Goose/Netherlands/11/2022 | Erasmus Medical Center | Erasmus Medical Center |
| EPI2185386 | NS | Netherlands | 2022-Aug-26 | EPI_ISL_15088300 | A/Greylag Goose/Netherlands/11/2022 | Erasmus Medical Center | Erasmus Medical Center |
| EPI2185387 | MP | Netherlands | 2022-Aug-26 | EPI_ISL_15088300 | A/Greylag Goose/Netherlands/11/2022 | Erasmus Medical Center | Erasmus Medical Center |
| EPI2185393 | NP | Netherlands | 2022-Aug-26 | EPI_ISL_15088301 | A/Greylag Goose/Netherlands/12/2022 | Erasmus Medical Center | Erasmus Medical Center |
| EPI2185394 | NS | Netherlands | 2022-Aug-26 | EPI_ISL_15088301 | A/Greylag Goose/Netherlands/12/2022 | Erasmus Medical Center | Erasmus Medical Center |

| | | | | | | | |
|---|---|---|---|---|---|---|---|
| EPI2185395 | MP | Netherlands | 2022-Aug-26 | EPI_ISL_15088301 | A/Greylag Goose/Netherlands/12/2022 | Erasmus Medical Center | Erasmus Medical Center |
| EPI2185400 | NP | Netherlands | 2022-Aug-27 | EPI_ISL_15088302 | A/Greylag Goose/Netherlands/13/2022 | Erasmus Medical Center | Erasmus Medical Center |
| EPI2185401 | NS | Netherlands | 2022-Aug-27 | EPI_ISL_15088302 | A/Greylag Goose/Netherlands/13/2022 | Erasmus Medical Center | Erasmus Medical Center |
| EPI2185402 | MP | Netherlands | 2022-Aug-27 | EPI_ISL_15088302 | A/Greylag Goose/Netherlands/13/2022 | Erasmus Medical Center | Erasmus Medical Center |
| EPI2185408 | NP | Netherlands | 2022-Aug-29 | EPI_ISL_15088303 | A/Lesser Black-backed Gull/Netherlands/3/2022 | Erasmus Medical Center | Erasmus Medical Center |
| EPI2185409 | NS | Netherlands | 2022-Aug-29 | EPI_ISL_15088303 | A/Lesser Black-backed Gull/Netherlands/3/2022 | Erasmus Medical Center | Erasmus Medical Center |
| EPI2185410 | MP | Netherlands | 2022-Aug-29 | EPI_ISL_15088303 | A/Lesser Black-backed Gull/Netherlands/3/2022 | Erasmus Medical Center | Erasmus Medical Center |
| EPI2185416 | NP | Netherlands | 2022-Aug-29 | EPI_ISL_15088304 | A/European Herring Gull/Netherlands/15/2022 | Erasmus Medical Center | Erasmus Medical Center |
| EPI2185417 | NS | Netherlands | 2022-Aug-29 | EPI_ISL_15088304 | A/European Herring Gull/Netherlands/15/2022 | Erasmus Medical Center | Erasmus Medical Center |
| EPI2185418 | MP | Netherlands | 2022-Aug-29 | EPI_ISL_15088304 | A/European Herring Gull/Netherlands/15/2022 | Erasmus Medical Center | Erasmus Medical Center |
| EPI2185424 | NP | Netherlands | 2022-Aug-29 | EPI_ISL_15088305 | A/Common Teal/Netherlands/1/2022 | Erasmus Medical Center | Erasmus Medical Center |
| EPI2185425 | NS | Netherlands | 2022-Aug-29 | EPI_ISL_15088305 | A/Common Teal/Netherlands/1/2022 | Erasmus Medical Center | Erasmus Medical Center |
| EPI2185426 | MP | Netherlands | 2022-Aug-29 | EPI_ISL_15088305 | A/Common Teal/Netherlands/1/2022 | Erasmus Medical Center | Erasmus Medical Center |
| EPI2185432 | NP | Netherlands | 2022-Aug-18 | EPI_ISL_15088306 | A/Mallard/Netherlands/3/2022 | Erasmus Medical Center | Erasmus Medical Center |
| EPI2185433 | NS | Netherlands | 2022-Aug-18 | EPI_ISL_15088306 | A/Mallard/Netherlands/3/2022 | Erasmus Medical Center | Erasmus Medical Center |
| EPI2185434 | MP | Netherlands | 2022-Aug-18 | EPI_ISL_15088306 | A/Mallard/Netherlands/3/2022 | Erasmus Medical Center | Erasmus Medical Center |
| EPI2185446 | NP | Netherlands | 2022-Aug-21 | EPI_ISL_15088308 | A/Mallard/Netherlands/5/2022 | Erasmus Medical Center | Erasmus Medical Center |
| EPI2185447 | NS | Netherlands | 2022-Aug-21 | EPI_ISL_15088308 | A/Mallard/Netherlands/5/2022 | Erasmus Medical Center | Erasmus Medical Center |
| EPI2185448 | MP | Netherlands | 2022-Aug-21 | EPI_ISL_15088308 | A/Mallard/Netherlands/5/2022 | Erasmus Medical Center | Erasmus Medical Center |
| EPI2185453 | NP | Netherlands | 2022-Aug-24 | EPI_ISL_15088309 | A/Mallard/Netherlands/6/2022 | Erasmus Medical Center | Erasmus Medical Center |
| EPI2185454 | NS | Netherlands | 2022-Aug-24 | EPI_ISL_15088309 | A/Mallard/Netherlands/6/2022 | Erasmus Medical Center | Erasmus Medical Center |
| EPI2185455 | MP | Netherlands | 2022-Aug-24 | EPI_ISL_15088309 | A/Mallard/Netherlands/6/2022 | Erasmus Medical Center | Erasmus Medical Center |
| EPI2185484 | MP | United Kingdom | 2022-Aug-26 | EPI_ISL_15115556 | A/turkey/England/112259/2022 | Animal and Plant Health Agency (APHA) | Animal and Plant Health Agency (APHA) |
| EPI2185482 | NP | United Kingdom | 2022-Aug-26 | EPI_ISL_15115556 | A/turkey/England/112259/2022 | Animal and Plant Health Agency (APHA) | Animal and Plant Health Agency (APHA) |
| EPI2185486 | NS | United Kingdom | 2022-Aug-26 | EPI_ISL_15115556 | A/turkey/England/112259/2022 | Animal and Plant Health Agency (APHA) | Animal and Plant Health Agency (APHA) |
| EPI2191362 | NP | Netherlands | 2022-Sep-04 | EPI_ISL_15267012 | A/Lesser Black-backed Gull/Netherlands/4/2022 | Erasmus Medical Center | Erasmus Medical Center |
| EPI2191363 | NS | Netherlands | 2022-Sep-04 | EPI_ISL_15267012 | A/Lesser Black-backed Gull/Netherlands/4/2022 | Erasmus Medical Center | Erasmus Medical Center |
| EPI2191364 | MP | Netherlands | 2022-Sep-04 | EPI_ISL_15267012 | A/Lesser Black-backed Gull/Netherlands/4/2022 | Erasmus Medical Center | Erasmus Medical Center |
| EPI2191370 | NP | Netherlands | 2022-Jun-16 | EPI_ISL_15267013 | A/Eurasian Spoonbill/Netherlands/5/2022 | Erasmus Medical Center | Erasmus Medical Center |
| EPI2191371 | NS | Netherlands | 2022-Jun-16 | EPI_ISL_15267013 | A/Eurasian Spoonbill/Netherlands/5/2022 | Erasmus Medical Center | Erasmus Medical Center |
| EPI2191372 | MP | Netherlands | 2022-Jun-16 | EPI_ISL_15267013 | A/Eurasian Spoonbill/Netherlands/5/2022 | Erasmus Medical Center | Erasmus Medical Center |
| EPI2191378 | NP | Netherlands | 2022-Jun-16 | EPI_ISL_15267014 | A/Eurasian Spoonbill/Netherlands/6/2022 | Erasmus Medical Center | Erasmus Medical Center |
| EPI2191379 | NS | Netherlands | 2022-Jun-16 | EPI_ISL_15267014 | A/Eurasian Spoonbill/Netherlands/6/2022 | Erasmus Medical Center | Erasmus Medical Center |
| EPI2191380 | MP | Netherlands | 2022-Jun-16 | EPI_ISL_15267014 | A/Eurasian Spoonbill/Netherlands/6/2022 | Erasmus Medical Center | Erasmus Medical Center |
| EPI2191386 | NP | Netherlands | 2022-Jun-16 | EPI_ISL_15267016 | A/Eurasian Spoonbill/Netherlands/7/2022 | Erasmus Medical Center | Erasmus Medical Center |

| | | | | | | | |
|---|---|---|---|---|---|---|---|
| EPI2191387 | NS | Netherlands | 2022-Jun-16 | EPI_ISL_15267016 | A/Eurasian Spoonbill/Netherlands/7/2022 | Erasmus Medical Center | Erasmus Medical Center |
| EPI2191388 | MP | Netherlands | 2022-Jun-16 | EPI_ISL_15267016 | A/Eurasian Spoonbill/Netherlands/7/2022 | Erasmus Medical Center | Erasmus Medical Center |
| EPI2191394 | NP | Netherlands | 2022-Aug-31 | EPI_ISL_15267017 | A/Eurasian Spoonbill/Netherlands/8/2022 | Erasmus Medical Center | Erasmus Medical Center |
| EPI2191395 | NS | Netherlands | 2022-Aug-31 | EPI_ISL_15267017 | A/Eurasian Spoonbill/Netherlands/8/2022 | Erasmus Medical Center | Erasmus Medical Center |
| EPI2191396 | MP | Netherlands | 2022-Aug-31 | EPI_ISL_15267017 | A/Eurasian Spoonbill/Netherlands/8/2022 | Erasmus Medical Center | Erasmus Medical Center |
| EPI2191402 | NP | Netherlands | 2022-Aug-31 | EPI_ISL_15267018 | A/Greylag Goose/Netherlands/14/2022 | Erasmus Medical Center | Erasmus Medical Center |
| EPI2191403 | NS | Netherlands | 2022-Aug-31 | EPI_ISL_15267018 | A/Greylag Goose/Netherlands/14/2022 | Erasmus Medical Center | Erasmus Medical Center |
| EPI2191404 | MP | Netherlands | 2022-Aug-31 | EPI_ISL_15267018 | A/Greylag Goose/Netherlands/14/2022 | Erasmus Medical Center | Erasmus Medical Center |
| EPI2191410 | NP | Netherlands | 2022-Sep-02 | EPI_ISL_15267019 | A/Greylag Goose/Netherlands/15/2022 | Erasmus Medical Center | Erasmus Medical Center |
| EPI2191411 | NS | Netherlands | 2022-Sep-02 | EPI_ISL_15267019 | A/Greylag Goose/Netherlands/15/2022 | Erasmus Medical Center | Erasmus Medical Center |
| EPI2191412 | MP | Netherlands | 2022-Sep-02 | EPI_ISL_15267019 | A/Greylag Goose/Netherlands/15/2022 | Erasmus Medical Center | Erasmus Medical Center |
| EPI2191418 | NP | Netherlands | 2022-Sep-02 | EPI_ISL_15267020 | A/Greylag Goose/Netherlands/16/2022 | Erasmus Medical Center | Erasmus Medical Center |
| EPI2191419 | NS | Netherlands | 2022-Sep-02 | EPI_ISL_15267020 | A/Greylag Goose/Netherlands/16/2022 | Erasmus Medical Center | Erasmus Medical Center |
| EPI2191420 | MP | Netherlands | 2022-Sep-02 | EPI_ISL_15267020 | A/Greylag Goose/Netherlands/16/2022 | Erasmus Medical Center | Erasmus Medical Center |
| EPI2191426 | NP | Netherlands | 2022-Sep-02 | EPI_ISL_15267021 | A/Common Tern/Netherlands/27/2022 | Erasmus Medical Center | Erasmus Medical Center |
| EPI2191427 | NS | Netherlands | 2022-Sep-02 | EPI_ISL_15267021 | A/Common Tern/Netherlands/27/2022 | Erasmus Medical Center | Erasmus Medical Center |
| EPI2191428 | MP | Netherlands | 2022-Sep-02 | EPI_ISL_15267021 | A/Common Tern/Netherlands/27/2022 | Erasmus Medical Center | Erasmus Medical Center |
| EPI2191434 | NP | Netherlands | 2022-Sep-03 | EPI_ISL_15267022 | A/European Herring Gull/Netherlands/16/2022 | Erasmus Medical Center | Erasmus Medical Center |
| EPI2191435 | NS | Netherlands | 2022-Sep-03 | EPI_ISL_15267022 | A/European Herring Gull/Netherlands/16/2022 | Erasmus Medical Center | Erasmus Medical Center |
| EPI2191436 | MP | Netherlands | 2022-Sep-03 | EPI_ISL_15267022 | A/European Herring Gull/Netherlands/16/2022 | Erasmus Medical Center | Erasmus Medical Center |
| EPI2191442 | NP | Netherlands | 2022-Jul-13 | EPI_ISL_15267023 | A/Northern Gannet/Netherlands/2/2022 | Erasmus Medical Center | Erasmus Medical Center |
| EPI2191443 | NS | Netherlands | 2022-Jul-13 | EPI_ISL_15267023 | A/Northern Gannet/Netherlands/2/2022 | Erasmus Medical Center | Erasmus Medical Center |
| EPI2191444 | MP | Netherlands | 2022-Jul-13 | EPI_ISL_15267023 | A/Northern Gannet/Netherlands/2/2022 | Erasmus Medical Center | Erasmus Medical Center |
| EPI2191448 | NP | Netherlands | 2022-Jul-10 | EPI_ISL_15267024 | A/Northern Gannet/Netherlands/3/2022 | Erasmus Medical Center | Erasmus Medical Center |
| EPI2191449 | NS | Netherlands | 2022-Jul-10 | EPI_ISL_15267024 | A/Northern Gannet/Netherlands/3/2022 | Erasmus Medical Center | Erasmus Medical Center |
| EPI2191450 | MP | Netherlands | 2022-Jul-10 | EPI_ISL_15267024 | A/Northern Gannet/Netherlands/3/2022 | Erasmus Medical Center | Erasmus Medical Center |
| EPI2191455 | NP | Netherlands | 2022-Jul-09 | EPI_ISL_15267025 | A/Northern Gannet/Netherlands/4/2022 | Erasmus Medical Center | Erasmus Medical Center |
| EPI2191456 | NS | Netherlands | 2022-Jul-09 | EPI_ISL_15267025 | A/Northern Gannet/Netherlands/4/2022 | Erasmus Medical Center | Erasmus Medical Center |
| EPI2191457 | MP | Netherlands | 2022-Jul-09 | EPI_ISL_15267025 | A/Northern Gannet/Netherlands/4/2022 | Erasmus Medical Center | Erasmus Medical Center |
| EPI2191462 | NP | Netherlands | 2022-Jun-26 | EPI_ISL_15267026 | A/Northern Gannet/Netherlands/5/2022 | Erasmus Medical Center | Erasmus Medical Center |
| EPI2191463 | NS | Netherlands | 2022-Jun-26 | EPI_ISL_15267026 | A/Northern Gannet/Netherlands/5/2022 | Erasmus Medical Center | Erasmus Medical Center |
| EPI2191464 | MP | Netherlands | 2022-Jun-26 | EPI_ISL_15267026 | A/Northern Gannet/Netherlands/5/2022 | Erasmus Medical Center | Erasmus Medical Center |
| EPI2191470 | NP | Netherlands | 2022-Jun-21 | EPI_ISL_15267027 | A/Northern Gannet/Netherlands/6/2022 | Erasmus Medical Center | Erasmus Medical Center |
| EPI2191471 | NS | Netherlands | 2022-Jun-21 | EPI_ISL_15267027 | A/Northern Gannet/Netherlands/6/2022 | Erasmus Medical Center | Erasmus Medical Center |
| EPI2191472 | MP | Netherlands | 2022-Jun-21 | EPI_ISL_15267027 | A/Northern Gannet/Netherlands/6/2022 | Erasmus Medical Center | Erasmus Medical Center |

| | | | | | | | | |
|---|---|---|---|---|---|---|---|---|
| EPI2193352 | NP | Netherlands | 2022-Sep-10 | EPI_ISL_15364788 | A/European Herring Gull/Netherlands/17/2022 | Erasmus Medical Center | Erasmus Medical Center | |
| EPI2193353 | NS | Netherlands | 2022-Sep-10 | EPI_ISL_15364788 | A/European Herring Gull/Netherlands/17/2022 | Erasmus Medical Center | Erasmus Medical Center | |
| EPI2193354 | MP | Netherlands | 2022-Sep-10 | EPI_ISL_15364788 | A/European Herring Gull/Netherlands/17/2022 | Erasmus Medical Center | Erasmus Medical Center | |
| EPI2193359 | NP | Netherlands | 2022-Sep-10 | EPI_ISL_15364789 | A/European Herring Gull/Netherlands/18/2022 | Erasmus Medical Center | Erasmus Medical Center | |
| EPI2193360 | NS | Netherlands | 2022-Sep-10 | EPI_ISL_15364789 | A/European Herring Gull/Netherlands/18/2022 | Erasmus Medical Center | Erasmus Medical Center | |
| EPI2193361 | MP | Netherlands | 2022-Sep-10 | EPI_ISL_15364789 | A/European Herring Gull/Netherlands/18/2022 | Erasmus Medical Center | Erasmus Medical Center | |
| EPI2193367 | NP | Netherlands | 2022-Sep-16 | EPI_ISL_15364790 | A/Greylag Goose/Netherlands/17/2022 | Erasmus Medical Center | Erasmus Medical Center | |
| EPI2193368 | NS | Netherlands | 2022-Sep-16 | EPI_ISL_15364790 | A/Greylag Goose/Netherlands/17/2022 | Erasmus Medical Center | Erasmus Medical Center | |
| EPI2193369 | MP | Netherlands | 2022-Sep-16 | EPI_ISL_15364790 | A/Greylag Goose/Netherlands/17/2022 | Erasmus Medical Center | Erasmus Medical Center | |
| EPI2193381 | NP | Netherlands | 2022-Sep-14 | EPI_ISL_15364792 | A/Mallard/Netherlands/11/2022 | Erasmus Medical Center | Erasmus Medical Center | |
| EPI2193382 | NS | Netherlands | 2022-Sep-14 | EPI_ISL_15364792 | A/Mallard/Netherlands/11/2022 | Erasmus Medical Center | Erasmus Medical Center | |
| EPI2193383 | MP | Netherlands | 2022-Sep-14 | EPI_ISL_15364792 | A/Mallard/Netherlands/11/2022 | Erasmus Medical Center | Erasmus Medical Center | |
| EPI2193389 | NP | Netherlands | 2022-Sep-19 | EPI_ISL_15364793 | A/Mallard/Netherlands/12/2022 | Erasmus Medical Center | Erasmus Medical Center | |
| EPI2193390 | NS | Netherlands | 2022-Sep-19 | EPI_ISL_15364793 | A/Mallard/Netherlands/12/2022 | Erasmus Medical Center | Erasmus Medical Center | |
| EPI2193391 | MP | Netherlands | 2022-Sep-19 | EPI_ISL_15364793 | A/Mallard/Netherlands/12/2022 | Erasmus Medical Center | Erasmus Medical Center | |
| EPI2193396 | NP | Netherlands | 2022-Sep-26 | EPI_ISL_15364794 | A/Mallard/Netherlands/13/2022 | Erasmus Medical Center | Erasmus Medical Center | |
| EPI2193397 | NS | Netherlands | 2022-Sep-26 | EPI_ISL_15364794 | A/Mallard/Netherlands/13/2022 | Erasmus Medical Center | Erasmus Medical Center | |
| EPI2193398 | MP | Netherlands | 2022-Sep-26 | EPI_ISL_15364794 | A/Mallard/Netherlands/13/2022 | Erasmus Medical Center | Erasmus Medical Center | |
| EPI2193404 | NP | Netherlands | 2022-Sep-08 | EPI_ISL_15364795 | A/Mallard/Netherlands/8/2022 | Erasmus Medical Center | Erasmus Medical Center | |
| EPI2193405 | NS | Netherlands | 2022-Sep-08 | EPI_ISL_15364795 | A/Mallard/Netherlands/8/2022 | Erasmus Medical Center | Erasmus Medical Center | |
| EPI2193406 | MP | Netherlands | 2022-Sep-08 | EPI_ISL_15364795 | A/Mallard/Netherlands/8/2022 | Erasmus Medical Center | Erasmus Medical Center | |
| EPI2193412 | NP | Netherlands | 2022-Sep-11 | EPI_ISL_15364796 | A/Mallard/Netherlands/9/2022 | Erasmus Medical Center | Erasmus Medical Center | |
| EPI2193413 | NS | Netherlands | 2022-Sep-11 | EPI_ISL_15364796 | A/Mallard/Netherlands/9/2022 | Erasmus Medical Center | Erasmus Medical Center | |
| EPI2193414 | MP | Netherlands | 2022-Sep-11 | EPI_ISL_15364796 | A/Mallard/Netherlands/9/2022 | Erasmus Medical Center | Erasmus Medical Center | |
| EPI2193420 | NP | Netherlands | 2022-Sep-19 | EPI_ISL_15364797 | A/Mute Swan/Netherlands/2/2022 | Erasmus Medical Center | Erasmus Medical Center | |
| EPI2193421 | NS | Netherlands | 2022-Sep-19 | EPI_ISL_15364797 | A/Mute Swan/Netherlands/2/2022 | Erasmus Medical Center | Erasmus Medical Center | |
| EPI2193422 | MP | Netherlands | 2022-Sep-19 | EPI_ISL_15364797 | A/Mute Swan/Netherlands/2/2022 | Erasmus Medical Center | Erasmus Medical Center | |
| EPI2197217 | NP | Finland | 2022-Feb-01 | EPI_ISL_15518269 | A/lynx/Finland/Vi209-22_22VIR3127-2/2022 | Finnish Food Authority | Istituto Zooprofilattico Sperimentale Delle Venezie | Tammiranta, N.;Kantala, T.;Laamanen, I.;Gadd, T.;Zecchin, B.;Fusaro, A.;Schivo, A.;Salviato, A.;Palumbo, E.;Milani, A.;Giussani, E.;Pastori, A.;Monne, I.;Terregino, C. |
| EPI2197218 | NS | Finland | 2022-Feb-01 | EPI_ISL_15518269 | A/lynx/Finland/Vi209-22_22VIR3127-2/2022 | Finnish Food Authority | Istituto Zooprofilattico Sperimentale Delle Venezie | Tammiranta, N.;Kantala, T.;Laamanen, I.;Gadd, T.;Zecchin, B.;Fusaro, A.;Schivo, A.;Salviato, A.;Palumbo, E.;Milani, A.;Giussani, E.;Pastori, A.;Monne, I.;Terregino, C. |
| EPI2197219 | MP | Finland | 2022-Feb-01 | EPI_ISL_15518269 | A/lynx/Finland/Vi209-22_22VIR3127-2/2022 | Finnish Food Authority | Istituto Zooprofilattico Sperimentale Delle Venezie | Tammiranta, N.;Kantala, T.;Laamanen, I.;Gadd, T.;Zecchin, B.;Fusaro, A.;Schivo, A.;Salviato, A.;Palumbo, E.;Milani, A.;Giussani, E.;Pastori, A.;Monne, I.;Terregino, C. |
| EPI2197374 | NS | Poland | 2022-Sep-20 | EPI_ISL_15535272 | A/domestic_goose/Poland/H397-N/2022 | National Veterinary Research Institut Poland, PIWet-PIB | National Veterinary Research Institut Poland, PIWet-PIB | Świętoń E., Śmietanka K. |
| EPI2197369 | NP | Poland | 2022-Sep-20 | EPI_ISL_15535272 | A/domestic_goose/Poland/H397-N/2022 | National Veterinary Research Institut Poland, PIWet-PIB | National Veterinary Research Institut Poland, PIWet-PIB | Świętoń E., Śmietanka K. |
| EPI2197372 | MP | Poland | 2022-Sep-20 | EPI_ISL_15535272 | A/domestic_goose/Poland/H397-N/2022 | National Veterinary Research Institut Poland, PIWet-PIB | National Veterinary Research Institut Poland, PIWet-PIB | Świętoń E., Śmietanka K. |
| EPI2197455 | NP | Spain | 2022-Sep-23 | EPI_ISL_15542438 | A/CastillaLaMancha/3739/2022 | Instituto de Salud Carlos III | Instituto de Salud Carlos III | Pozo,F; Iglesias-Caballero, M.; Molinero,M.; Camarero,S.; Mart?n,V.; Reyes,N.; V?zquez-Mor?n,S.; Casas,I. |
| EPI2197456 | NS | Spain | 2022-Sep-23 | EPI_ISL_15542438 | A/CastillaLaMancha/3739/2022 | Instituto de Salud Carlos III | Instituto de Salud Carlos III | Pozo,F; Iglesias-Caballero, M.; Molinero,M.; Camarero,S.; Mart?n,V.; Reyes,N.; V?zquez-Mor?n,S.; Casas,I. |

| | | | | | | | | |
|---|---|---|---|---|---|---|---|---|
| EPI2352712 | MP | Spain | 2022-Sep-23 | EPI_ISL_15542438 | A/CastillaLaMancha/3739/2022 | Instituto de Salud Carlos III | Instituto de Salud Carlos III | Pozo,F; Iglesias-Caballero, M.; Molinero,M.; Camarero,S.; Mart?n,V.; Reyes,N.; V?zquez-Mor?n,S.; Casas,I. |
| EPI2197787 | NP | Netherlands | 2022-Sep-21 | EPI_ISL_15579535 | A/European Herring Gull/Netherlands/19/2022 | Erasmus Medical Center | Erasmus Medical Center | |
| EPI2197788 | NS | Netherlands | 2022-Sep-21 | EPI_ISL_15579535 | A/European Herring Gull/Netherlands/19/2022 | Erasmus Medical Center | Erasmus Medical Center | |
| EPI2197789 | MP | Netherlands | 2022-Sep-21 | EPI_ISL_15579535 | A/European Herring Gull/Netherlands/19/2022 | Erasmus Medical Center | Erasmus Medical Center | |
| EPI2197795 | NP | Netherlands | 2022-Sep-25 | EPI_ISL_15579536 | A/Greylag Goose/Netherlands/18/2022 | Erasmus Medical Center | Erasmus Medical Center | |
| EPI2197796 | NS | Netherlands | 2022-Sep-25 | EPI_ISL_15579536 | A/Greylag Goose/Netherlands/18/2022 | Erasmus Medical Center | Erasmus Medical Center | |
| EPI2197797 | MP | Netherlands | 2022-Sep-25 | EPI_ISL_15579536 | A/Greylag Goose/Netherlands/18/2022 | Erasmus Medical Center | Erasmus Medical Center | |
| EPI2197802 | NP | Netherlands | 2022-Sep-26 | EPI_ISL_15579537 | A/Barnacle Goose/Netherlands/19/2022 | Erasmus Medical Center | Erasmus Medical Center | |
| EPI2197803 | NS | Netherlands | 2022-Sep-26 | EPI_ISL_15579537 | A/Barnacle Goose/Netherlands/19/2022 | Erasmus Medical Center | Erasmus Medical Center | |
| EPI2197804 | MP | Netherlands | 2022-Sep-26 | EPI_ISL_15579537 | A/Barnacle Goose/Netherlands/19/2022 | Erasmus Medical Center | Erasmus Medical Center | |
| EPI2197810 | NP | Netherlands | 2022-Sep-28 | EPI_ISL_15579538 | A/Eurasian Curlew/Netherlands/4/2022 | Erasmus Medical Center | Erasmus Medical Center | |
| EPI2197811 | NS | Netherlands | 2022-Sep-28 | EPI_ISL_15579538 | A/Eurasian Curlew/Netherlands/4/2022 | Erasmus Medical Center | Erasmus Medical Center | |
| EPI2197812 | MP | Netherlands | 2022-Sep-28 | EPI_ISL_15579538 | A/Eurasian Curlew/Netherlands/4/2022 | Erasmus Medical Center | Erasmus Medical Center | |
| EPI2197818 | NP | Netherlands | 2022-Oct-02 | EPI_ISL_15579539 | A/European Herring Gull/Netherlands/20/2022 | Erasmus Medical Center | Erasmus Medical Center | |
| EPI2197819 | NS | Netherlands | 2022-Oct-02 | EPI_ISL_15579539 | A/European Herring Gull/Netherlands/20/2022 | Erasmus Medical Center | Erasmus Medical Center | |
| EPI2197820 | MP | Netherlands | 2022-Oct-02 | EPI_ISL_15579539 | A/European Herring Gull/Netherlands/20/2022 | Erasmus Medical Center | Erasmus Medical Center | |
| EPI2197826 | NP | Netherlands | 2022-Sep-21 | EPI_ISL_15579540 | A/Mallard/Netherlands/15/2022 | Erasmus Medical Center | Erasmus Medical Center | |
| EPI2197827 | NS | Netherlands | 2022-Sep-21 | EPI_ISL_15579540 | A/Mallard/Netherlands/15/2022 | Erasmus Medical Center | Erasmus Medical Center | |
| EPI2197828 | MP | Netherlands | 2022-Sep-21 | EPI_ISL_15579540 | A/Mallard/Netherlands/15/2022 | Erasmus Medical Center | Erasmus Medical Center | |
| EPI2197850 | NP | Netherlands | 2022-Oct-01 | EPI_ISL_15579543 | A/Mallard/Netherlands/18/2022 | Erasmus Medical Center | Erasmus Medical Center | |
| EPI2197851 | NS | Netherlands | 2022-Oct-01 | EPI_ISL_15579543 | A/Mallard/Netherlands/18/2022 | Erasmus Medical Center | Erasmus Medical Center | |
| EPI2197852 | MP | Netherlands | 2022-Oct-01 | EPI_ISL_15579543 | A/Mallard/Netherlands/18/2022 | Erasmus Medical Center | Erasmus Medical Center | |
| EPI2197858 | NP | Netherlands | 2022-Oct-09 | EPI_ISL_15579544 | A/Great Black-backed Gull/4/2022 | Erasmus Medical Center | Erasmus Medical Center | |
| EPI2197859 | NS | Netherlands | 2022-Oct-09 | EPI_ISL_15579544 | A/Great Black-backed Gull/4/2022 | Erasmus Medical Center | Erasmus Medical Center | |
| EPI2197860 | MP | Netherlands | 2022-Oct-09 | EPI_ISL_15579544 | A/Great Black-backed Gull/4/2022 | Erasmus Medical Center | Erasmus Medical Center | |
| EPI2197866 | NP | Netherlands | 2022-Oct-09 | EPI_ISL_15579545 | A/Sanderling/Netherlands/2/2022 | Erasmus Medical Center | Erasmus Medical Center | |
| EPI2197867 | NS | Netherlands | 2022-Oct-09 | EPI_ISL_15579545 | A/Sanderling/Netherlands/2/2022 | Erasmus Medical Center | Erasmus Medical Center | |
| EPI2197868 | MP | Netherlands | 2022-Oct-09 | EPI_ISL_15579545 | A/Sanderling/Netherlands/2/2022 | Erasmus Medical Center | Erasmus Medical Center | |
| EPI2198049 | NP | Netherlands | 2022-Oct-12 | EPI_ISL_15581789 | A/Mallard/Netherlands/19/2022 | Erasmus Medical Center | Erasmus Medical Center | |
| EPI2198050 | NS | Netherlands | 2022-Oct-12 | EPI_ISL_15581789 | A/Mallard/Netherlands/19/2022 | Erasmus Medical Center | Erasmus Medical Center | |
| EPI2198051 | MP | Netherlands | 2022-Oct-12 | EPI_ISL_15581789 | A/Mallard/Netherlands/19/2022 | Erasmus Medical Center | Erasmus Medical Center | |
| EPI2198064 | NP | Netherlands | 2022-Oct-11 | EPI_ISL_15581791 | A/Mallard/Netherlands/21/2022 | Erasmus Medical Center | Erasmus Medical Center | |
| EPI2198065 | NS | Netherlands | 2022-Oct-11 | EPI_ISL_15581791 | A/Mallard/Netherlands/21/2022 | Erasmus Medical Center | Erasmus Medical Center | |
| EPI2198066 | MP | Netherlands | 2022-Oct-11 | EPI_ISL_15581791 | A/Mallard/Netherlands/21/2022 | Erasmus Medical Center | Erasmus Medical Center | |
| EPI2198071 | NP | Netherlands | 2022-Oct-13 | EPI_ISL_15581792 | A/Mallard/Netherlands/22/2022 | Erasmus Medical Center | Erasmus Medical Center | |

| | | | | | | | |
|---|---|---|---|---|---|---|---|
| EPI2198072 | NS | Netherlands | 2022-Oct-13 | EPI_ISL_15581792 | A/Mallard/Netherlands/22/2022 | Erasmus Medical Center | Erasmus Medical Center |
| EPI2198073 | MP | Netherlands | 2022-Oct-13 | EPI_ISL_15581792 | A/Mallard/Netherlands/22/2022 | Erasmus Medical Center | Erasmus Medical Center |
| EPI2198093 | NP | United Kingdom | 2022-Oct-04 | EPI_ISL_15585748 | A/Chicken/England/126673/2022 | Animal and Plant Health Agency (APHA) | Animal and Plant Health Agency (APHA) |
| EPI2198094 | NS | United Kingdom | 2022-Oct-04 | EPI_ISL_15585748 | A/Chicken/England/126673/2022 | Animal and Plant Health Agency (APHA) | Animal and Plant Health Agency (APHA) |
| EPI2198095 | MP | United Kingdom | 2022-Oct-04 | EPI_ISL_15585748 | A/Chicken/England/126673/2022 | Animal and Plant Health Agency (APHA) | Animal and Plant Health Agency (APHA) |
| EPI2198101 | NP | United Kingdom | 2022-Oct-06 | EPI_ISL_15585749 | A/Chicken/England/127733/2022 | Animal and Plant Health Agency (APHA) | Animal and Plant Health Agency (APHA) |
| EPI2198102 | NS | United Kingdom | 2022-Oct-06 | EPI_ISL_15585749 | A/Chicken/England/127733/2022 | Animal and Plant Health Agency (APHA) | Animal and Plant Health Agency (APHA) |
| EPI2198103 | MP | United Kingdom | 2022-Oct-06 | EPI_ISL_15585749 | A/Chicken/England/127733/2022 | Animal and Plant Health Agency (APHA) | Animal and Plant Health Agency (APHA) |
| EPI2198109 | NP | United Kingdom | 2022-Oct-06 | EPI_ISL_15585750 | A/Chicken/England/127861/2022 | Animal and Plant Health Agency (APHA) | Animal and Plant Health Agency (APHA) |
| EPI2198110 | NS | United Kingdom | 2022-Oct-06 | EPI_ISL_15585750 | A/Chicken/England/127861/2022 | Animal and Plant Health Agency (APHA) | Animal and Plant Health Agency (APHA) |
| EPI2198111 | MP | United Kingdom | 2022-Oct-06 | EPI_ISL_15585750 | A/Chicken/England/127861/2022 | Animal and Plant Health Agency (APHA) | Animal and Plant Health Agency (APHA) |
| EPI2198117 | NP | United Kingdom | 2022-Oct-07 | EPI_ISL_15585751 | A/Chicken/England/128321/2022 | Animal and Plant Health Agency (APHA) | Animal and Plant Health Agency (APHA) |
| EPI2198118 | NS | United Kingdom | 2022-Oct-07 | EPI_ISL_15585751 | A/Chicken/England/128321/2022 | Animal and Plant Health Agency (APHA) | Animal and Plant Health Agency (APHA) |
| EPI2198119 | MP | United Kingdom | 2022-Oct-07 | EPI_ISL_15585751 | A/Chicken/England/128321/2022 | Animal and Plant Health Agency (APHA) | Animal and Plant Health Agency (APHA) |
| EPI2198125 | NP | United Kingdom | 2022-Oct-08 | EPI_ISL_15585752 | A/Chicken/England/128527/2022 | Animal and Plant Health Agency (APHA) | Animal and Plant Health Agency (APHA) |
| EPI2198126 | NS | United Kingdom | 2022-Oct-08 | EPI_ISL_15585752 | A/Chicken/England/128527/2022 | Animal and Plant Health Agency (APHA) | Animal and Plant Health Agency (APHA) |
| EPI2198127 | MP | United Kingdom | 2022-Oct-08 | EPI_ISL_15585752 | A/Chicken/England/128527/2022 | Animal and Plant Health Agency (APHA) | Animal and Plant Health Agency (APHA) |
| EPI2198133 | NP | United Kingdom | 2022-Oct-10 | EPI_ISL_15585753 | A/Chicken/England/129627/2022 | Animal and Plant Health Agency (APHA) | Animal and Plant Health Agency (APHA) |
| EPI2198134 | NS | United Kingdom | 2022-Oct-10 | EPI_ISL_15585753 | A/Chicken/England/129627/2022 | Animal and Plant Health Agency (APHA) | Animal and Plant Health Agency (APHA) |
| EPI2198135 | MP | United Kingdom | 2022-Oct-10 | EPI_ISL_15585753 | A/Chicken/England/129627/2022 | Animal and Plant Health Agency (APHA) | Animal and Plant Health Agency (APHA) |
| EPI2198141 | NP | United Kingdom | 2022-Oct-10 | EPI_ISL_15585754 | A/Chicken/England/129733/2022 | Animal and Plant Health Agency (APHA) | Animal and Plant Health Agency (APHA) |
| EPI2198142 | NS | United Kingdom | 2022-Oct-10 | EPI_ISL_15585754 | A/Chicken/England/129733/2022 | Animal and Plant Health Agency (APHA) | Animal and Plant Health Agency (APHA) |
| EPI2198143 | MP | United Kingdom | 2022-Oct-10 | EPI_ISL_15585754 | A/Chicken/England/129733/2022 | Animal and Plant Health Agency (APHA) | Animal and Plant Health Agency (APHA) |
| EPI2198149 | NP | United Kingdom | 2022-Oct-10 | EPI_ISL_15585755 | A/Chicken/England/129801/2022 | Animal and Plant Health Agency (APHA) | Animal and Plant Health Agency (APHA) |
| EPI2198150 | NS | United Kingdom | 2022-Oct-10 | EPI_ISL_15585755 | A/Chicken/England/129801/2022 | Animal and Plant Health Agency (APHA) | Animal and Plant Health Agency (APHA) |
| EPI2198151 | MP | United Kingdom | 2022-Oct-10 | EPI_ISL_15585755 | A/Chicken/England/129801/2022 | Animal and Plant Health Agency (APHA) | Animal and Plant Health Agency (APHA) |
| EPI2198165 | NP | United Kingdom | 2022-Oct-18 | EPI_ISL_15585757 | A/Chicken/England/134205/2022 | Animal and Plant Health Agency (APHA) | Animal and Plant Health Agency (APHA) |
| EPI2198166 | NS | United Kingdom | 2022-Oct-18 | EPI_ISL_15585757 | A/Chicken/England/134205/2022 | Animal and Plant Health Agency (APHA) | Animal and Plant Health Agency (APHA) |
| EPI2198167 | MP | United Kingdom | 2022-Oct-18 | EPI_ISL_15585757 | A/Chicken/England/134205/2022 | Animal and Plant Health Agency (APHA) | Animal and Plant Health Agency (APHA) |
| EPI2198189 | NP | United Kingdom | 2022-Oct-05 | EPI_ISL_15585760 | A/Domestic_Goose/England/127100/2022 | Animal and Plant Health Agency (APHA) | Animal and Plant Health Agency (APHA) |
| EPI2198190 | NS | United Kingdom | 2022-Oct-05 | EPI_ISL_15585760 | A/Domestic_Goose/England/127100/2022 | Animal and Plant Health Agency (APHA) | Animal and Plant Health Agency (APHA) |
| EPI2198191 | MP | United Kingdom | 2022-Oct-05 | EPI_ISL_15585760 | A/Domestic_Goose/England/127100/2022 | Animal and Plant Health Agency (APHA) | Animal and Plant Health Agency (APHA) |
| EPI2198197 | NP | United Kingdom | 2022-Oct-11 | EPI_ISL_15585761 | A/Domestic_Goose/England/130370/2022 | Animal and Plant Health Agency (APHA) | Animal and Plant Health Agency (APHA) |
| EPI2198198 | NS | United Kingdom | 2022-Oct-11 | EPI_ISL_15585761 | A/Domestic_Goose/England/130370/2022 | Animal and Plant Health Agency (APHA) | Animal and Plant Health Agency (APHA) |
| EPI2198199 | MP | United Kingdom | 2022-Oct-11 | EPI_ISL_15585761 | A/Domestic_Goose/England/130370/2022 | Animal and Plant Health Agency (APHA) | Animal and Plant Health Agency (APHA) |

| | | | | | | | |
|---|---|---|---|---|---|---|---|
| EPI2198213 | NP | United Kingdom | 2022-Oct-06 | EPI_ISL_15585763 | A/Turkey/England/127741/2022 | Animal and Plant Health Agency (APHA) | Animal and Plant Health Agency (APHA) |
| EPI2198214 | NS | United Kingdom | 2022-Oct-06 | EPI_ISL_15585763 | A/Turkey/England/127741/2022 | Animal and Plant Health Agency (APHA) | Animal and Plant Health Agency (APHA) |
| EPI2198215 | MP | United Kingdom | 2022-Oct-06 | EPI_ISL_15585763 | A/Turkey/England/127741/2022 | Animal and Plant Health Agency (APHA) | Animal and Plant Health Agency (APHA) |
| EPI2198221 | NP | United Kingdom | 2022-Oct-07 | EPI_ISL_15585764 | A/Turkey/England/128362/2022 | Animal and Plant Health Agency (APHA) | Animal and Plant Health Agency (APHA) |
| EPI2198222 | NS | United Kingdom | 2022-Oct-07 | EPI_ISL_15585764 | A/Turkey/England/128362/2022 | Animal and Plant Health Agency (APHA) | Animal and Plant Health Agency (APHA) |
| EPI2198223 | MP | United Kingdom | 2022-Oct-07 | EPI_ISL_15585764 | A/Turkey/England/128362/2022 | Animal and Plant Health Agency (APHA) | Animal and Plant Health Agency (APHA) |
| EPI2198229 | NP | United Kingdom | 2022-Oct-09 | EPI_ISL_15585765 | A/Turkey/England/128999/2022 | Animal and Plant Health Agency (APHA) | Animal and Plant Health Agency (APHA) |
| EPI2198230 | NS | United Kingdom | 2022-Oct-09 | EPI_ISL_15585765 | A/Turkey/England/128999/2022 | Animal and Plant Health Agency (APHA) | Animal and Plant Health Agency (APHA) |
| EPI2198231 | MP | United Kingdom | 2022-Oct-09 | EPI_ISL_15585765 | A/Turkey/England/128999/2022 | Animal and Plant Health Agency (APHA) | Animal and Plant Health Agency (APHA) |
| EPI2198237 | NP | United Kingdom | 2022-Oct-09 | EPI_ISL_15585839 | A/Turkey/England/129035/2022 | Animal and Plant Health Agency (APHA) | Animal and Plant Health Agency (APHA) |
| EPI2198238 | NS | United Kingdom | 2022-Oct-09 | EPI_ISL_15585839 | A/Turkey/England/129035/2022 | Animal and Plant Health Agency (APHA) | Animal and Plant Health Agency (APHA) |
| EPI2198239 | MP | United Kingdom | 2022-Oct-09 | EPI_ISL_15585839 | A/Turkey/England/129035/2022 | Animal and Plant Health Agency (APHA) | Animal and Plant Health Agency (APHA) |
| EPI2198245 | NP | United Kingdom | 2022-Oct-10 | EPI_ISL_15585840 | A/Turkey/England/129898/2022 | Animal and Plant Health Agency (APHA) | Animal and Plant Health Agency (APHA) |
| EPI2198246 | NS | United Kingdom | 2022-Oct-10 | EPI_ISL_15585840 | A/Turkey/England/129898/2022 | Animal and Plant Health Agency (APHA) | Animal and Plant Health Agency (APHA) |
| EPI2198247 | MP | United Kingdom | 2022-Oct-10 | EPI_ISL_15585840 | A/Turkey/England/129898/2022 | Animal and Plant Health Agency (APHA) | Animal and Plant Health Agency (APHA) |
| EPI2198253 | NP | United Kingdom | 2022-Oct-10 | EPI_ISL_15585841 | A/Turkey/England/129968/2022 | Animal and Plant Health Agency (APHA) | Animal and Plant Health Agency (APHA) |
| EPI2198254 | NS | United Kingdom | 2022-Oct-10 | EPI_ISL_15585841 | A/Turkey/England/129968/2022 | Animal and Plant Health Agency (APHA) | Animal and Plant Health Agency (APHA) |
| EPI2198255 | MP | United Kingdom | 2022-Oct-10 | EPI_ISL_15585841 | A/Turkey/England/129968/2022 | Animal and Plant Health Agency (APHA) | Animal and Plant Health Agency (APHA) |
| EPI2198263 | MP | United Kingdom | 2022-Oct-10 | EPI_ISL_15585842 | A/Turkey/England/130038/2022 | Animal and Plant Health Agency (APHA) | Animal and Plant Health Agency (APHA) |
| EPI2198261 | NP | United Kingdom | 2022-Oct-10 | EPI_ISL_15585842 | A/Turkey/England/130038/2022 | Animal and Plant Health Agency (APHA) | Animal and Plant Health Agency (APHA) |
| EPI2198262 | NS | United Kingdom | 2022-Oct-10 | EPI_ISL_15585842 | A/Turkey/England/130038/2022 | Animal and Plant Health Agency (APHA) | Animal and Plant Health Agency (APHA) |
| EPI2198269 | NP | United Kingdom | 2022-Oct-10 | EPI_ISL_15585843 | A/Turkey/England/130119/2022 | Animal and Plant Health Agency (APHA) | Animal and Plant Health Agency (APHA) |
| EPI2198270 | NS | United Kingdom | 2022-Oct-10 | EPI_ISL_15585843 | A/Turkey/England/130119/2022 | Animal and Plant Health Agency (APHA) | Animal and Plant Health Agency (APHA) |
| EPI2198271 | MP | United Kingdom | 2022-Oct-10 | EPI_ISL_15585843 | A/Turkey/England/130119/2022 | Animal and Plant Health Agency (APHA) | Animal and Plant Health Agency (APHA) |
| EPI2198277 | NP | United Kingdom | 2022-Oct-11 | EPI_ISL_15585844 | A/Turkey/England/130299/2022 | Animal and Plant Health Agency (APHA) | Animal and Plant Health Agency (APHA) |
| EPI2198278 | NS | United Kingdom | 2022-Oct-11 | EPI_ISL_15585844 | A/Turkey/England/130299/2022 | Animal and Plant Health Agency (APHA) | Animal and Plant Health Agency (APHA) |
| EPI2198279 | MP | United Kingdom | 2022-Oct-11 | EPI_ISL_15585844 | A/Turkey/England/130299/2022 | Animal and Plant Health Agency (APHA) | Animal and Plant Health Agency (APHA) |
| EPI2198317 | NP | United Kingdom | 2022-Sep-17 | EPI_ISL_15585856 | A/chicken/England/118828/2022 | Animal and Plant Health Agency (APHA) | Animal and Plant Health Agency (APHA) |
| EPI2198318 | NS | United Kingdom | 2022-Sep-17 | EPI_ISL_15585856 | A/chicken/England/118828/2022 | Animal and Plant Health Agency (APHA) | Animal and Plant Health Agency (APHA) |
| EPI2198319 | MP | United Kingdom | 2022-Sep-17 | EPI_ISL_15585856 | A/chicken/England/118828/2022 | Animal and Plant Health Agency (APHA) | Animal and Plant Health Agency (APHA) |
| EPI2198325 | NP | United Kingdom | 2022-Sep-18 | EPI_ISL_15585857 | A/chicken/England/118935/2022 | Animal and Plant Health Agency (APHA) | Animal and Plant Health Agency (APHA) |
| EPI2198326 | NS | United Kingdom | 2022-Sep-18 | EPI_ISL_15585857 | A/chicken/England/118935/2022 | Animal and Plant Health Agency (APHA) | Animal and Plant Health Agency (APHA) |
| EPI2198327 | MP | United Kingdom | 2022-Sep-18 | EPI_ISL_15585857 | A/chicken/England/118935/2022 | Animal and Plant Health Agency (APHA) | Animal and Plant Health Agency (APHA) |
| EPI2198333 | NP | United Kingdom | 2022-Sep-19 | EPI_ISL_15585858 | A/chicken/England/119017/2022 | Animal and Plant Health Agency (APHA) | Animal and Plant Health Agency (APHA) |
| EPI2198334 | NS | United Kingdom | 2022-Sep-19 | EPI_ISL_15585858 | A/chicken/England/119017/2022 | Animal and Plant Health Agency (APHA) | Animal and Plant Health Agency (APHA) |

| | | | | | | | |
|---|---|---|---|---|---|---|---|
| EPI2198335 | MP | United Kingdom | 2022-Sep-19 | EPI_ISL_15585858 | A/chicken/England/119017/2022 | Animal and Plant Health Agency (APHA) | Animal and Plant Health Agency (APHA) |
| EPI2198357 | NP | United Kingdom | 2022-Sep-23 | EPI_ISL_15585861 | A/chicken/England/121695/2022 | Animal and Plant Health Agency (APHA) | Animal and Plant Health Agency (APHA) |
| EPI2198358 | NS | United Kingdom | 2022-Sep-23 | EPI_ISL_15585861 | A/chicken/England/121695/2022 | Animal and Plant Health Agency (APHA) | Animal and Plant Health Agency (APHA) |
| EPI2198359 | MP | United Kingdom | 2022-Sep-23 | EPI_ISL_15585861 | A/chicken/England/121695/2022 | Animal and Plant Health Agency (APHA) | Animal and Plant Health Agency (APHA) |
| EPI2198366 | NS | United Kingdom | 2022-Sep-23 | EPI_ISL_15585862 | A/chicken/England/121771/2022 | Animal and Plant Health Agency (APHA) | Animal and Plant Health Agency (APHA) |
| EPI2198367 | MP | United Kingdom | 2022-Sep-23 | EPI_ISL_15585862 | A/chicken/England/121771/2022 | Animal and Plant Health Agency (APHA) | Animal and Plant Health Agency (APHA) |
| EPI2198365 | NP | United Kingdom | 2022-Sep-23 | EPI_ISL_15585862 | A/chicken/England/121771/2022 | Animal and Plant Health Agency (APHA) | Animal and Plant Health Agency (APHA) |
| EPI2198373 | NP | United Kingdom | 2022-Sep-27 | EPI_ISL_15585863 | A/chicken/England/123195/2022 | Animal and Plant Health Agency (APHA) | Animal and Plant Health Agency (APHA) |
| EPI2198374 | NS | United Kingdom | 2022-Sep-27 | EPI_ISL_15585863 | A/chicken/England/123195/2022 | Animal and Plant Health Agency (APHA) | Animal and Plant Health Agency (APHA) |
| EPI2198375 | MP | United Kingdom | 2022-Sep-27 | EPI_ISL_15585863 | A/chicken/England/123195/2022 | Animal and Plant Health Agency (APHA) | Animal and Plant Health Agency (APHA) |
| EPI2198381 | NP | United Kingdom | 2022-Sep-28 | EPI_ISL_15585864 | A/chicken/England/123975/2022 | Animal and Plant Health Agency (APHA) | Animal and Plant Health Agency (APHA) |
| EPI2198382 | NS | United Kingdom | 2022-Sep-28 | EPI_ISL_15585864 | A/chicken/England/123975/2022 | Animal and Plant Health Agency (APHA) | Animal and Plant Health Agency (APHA) |
| EPI2198383 | MP | United Kingdom | 2022-Sep-28 | EPI_ISL_15585864 | A/chicken/England/123975/2022 | Animal and Plant Health Agency (APHA) | Animal and Plant Health Agency (APHA) |
| EPI2198389 | NP | United Kingdom | 2022-Sep-30 | EPI_ISL_15585865 | A/chicken/England/124798/2022 | Animal and Plant Health Agency (APHA) | Animal and Plant Health Agency (APHA) |
| EPI2198390 | NS | United Kingdom | 2022-Sep-30 | EPI_ISL_15585865 | A/chicken/England/124798/2022 | Animal and Plant Health Agency (APHA) | Animal and Plant Health Agency (APHA) |
| EPI2198391 | MP | United Kingdom | 2022-Sep-30 | EPI_ISL_15585865 | A/chicken/England/124798/2022 | Animal and Plant Health Agency (APHA) | Animal and Plant Health Agency (APHA) |
| EPI2198397 | NP | United Kingdom | 2022-Oct-03 | EPI_ISL_15585880 | A/chicken/England/125420/2022 | Animal and Plant Health Agency (APHA) | Animal and Plant Health Agency (APHA) |
| EPI2198398 | NS | United Kingdom | 2022-Oct-03 | EPI_ISL_15585880 | A/chicken/England/125420/2022 | Animal and Plant Health Agency (APHA) | Animal and Plant Health Agency (APHA) |
| EPI2198399 | MP | United Kingdom | 2022-Oct-03 | EPI_ISL_15585880 | A/chicken/England/125420/2022 | Animal and Plant Health Agency (APHA) | Animal and Plant Health Agency (APHA) |
| EPI2198405 | NP | United Kingdom | 2022-Oct-04 | EPI_ISL_15585881 | A/chicken/England/130482/2022 | Animal and Plant Health Agency (APHA) | Animal and Plant Health Agency (APHA) |
| EPI2198406 | NS | United Kingdom | 2022-Oct-04 | EPI_ISL_15585881 | A/chicken/England/130482/2022 | Animal and Plant Health Agency (APHA) | Animal and Plant Health Agency (APHA) |
| EPI2198407 | MP | United Kingdom | 2022-Oct-04 | EPI_ISL_15585881 | A/chicken/England/130482/2022 | Animal and Plant Health Agency (APHA) | Animal and Plant Health Agency (APHA) |
| EPI2198413 | NP | United Kingdom | 2022-Oct-12 | EPI_ISL_15585882 | A/chicken/England/131142/2022 | Animal and Plant Health Agency (APHA) | Animal and Plant Health Agency (APHA) |
| EPI2198414 | NS | United Kingdom | 2022-Oct-12 | EPI_ISL_15585882 | A/chicken/England/131142/2022 | Animal and Plant Health Agency (APHA) | Animal and Plant Health Agency (APHA) |
| EPI2198415 | MP | United Kingdom | 2022-Oct-12 | EPI_ISL_15585882 | A/chicken/England/131142/2022 | Animal and Plant Health Agency (APHA) | Animal and Plant Health Agency (APHA) |
| EPI2198429 | NP | United Kingdom | 2022-Oct-13 | EPI_ISL_15585884 | A/chicken/England/131916/2022 | Animal and Plant Health Agency (APHA) | Animal and Plant Health Agency (APHA) |
| EPI2198430 | NS | United Kingdom | 2022-Oct-13 | EPI_ISL_15585884 | A/chicken/England/131916/2022 | Animal and Plant Health Agency (APHA) | Animal and Plant Health Agency (APHA) |
| EPI2198431 | MP | United Kingdom | 2022-Oct-13 | EPI_ISL_15585884 | A/chicken/England/131916/2022 | Animal and Plant Health Agency (APHA) | Animal and Plant Health Agency (APHA) |
| EPI2198437 | NP | United Kingdom | 2022-Oct-14 | EPI_ISL_15585885 | A/chicken/England/132529/2022 | Animal and Plant Health Agency (APHA) | Animal and Plant Health Agency (APHA) |
| EPI2198438 | NS | United Kingdom | 2022-Oct-14 | EPI_ISL_15585885 | A/chicken/England/132529/2022 | Animal and Plant Health Agency (APHA) | Animal and Plant Health Agency (APHA) |
| EPI2198439 | MP | United Kingdom | 2022-Oct-14 | EPI_ISL_15585885 | A/chicken/England/132529/2022 | Animal and Plant Health Agency (APHA) | Animal and Plant Health Agency (APHA) |
| EPI2198445 | NP | United Kingdom | 2022-Oct-16 | EPI_ISL_15585886 | A/chicken/England/133051/2022 | Animal and Plant Health Agency (APHA) | Animal and Plant Health Agency (APHA) |
| EPI2198446 | NS | United Kingdom | 2022-Oct-16 | EPI_ISL_15585886 | A/chicken/England/133051/2022 | Animal and Plant Health Agency (APHA) | Animal and Plant Health Agency (APHA) |
| EPI2198447 | MP | United Kingdom | 2022-Oct-16 | EPI_ISL_15585886 | A/chicken/England/133051/2022 | Animal and Plant Health Agency (APHA) | Animal and Plant Health Agency (APHA) |
| EPI2198461 | NP | United Kingdom | 2022-Aug-30 | EPI_ISL_15585888 | A/chicken/Scotland/130966/2022 | Animal and Plant Health Agency (APHA) | Animal and Plant Health Agency (APHA) |

| | | | | | | | |
|---|---|---|---|---|---|---|---|
| EPI2198462 | NS | United Kingdom | 2022-Aug-30 | EPI_ISL_15585888 | A/chicken/Scotland/130966/2022 | Animal and Plant Health Agency (APHA) | Animal and Plant Health Agency (APHA) |
| EPI2198463 | MP | United Kingdom | 2022-Aug-30 | EPI_ISL_15585888 | A/chicken/Scotland/130966/2022 | Animal and Plant Health Agency (APHA) | Animal and Plant Health Agency (APHA) |
| EPI2198477 | NP | United Kingdom | 2022-Sep-04 | EPI_ISL_15585890 | A/chicken/Wales/116603/2022 | Animal and Plant Health Agency (APHA) | Animal and Plant Health Agency (APHA) |
| EPI2198478 | NS | United Kingdom | 2022-Sep-04 | EPI_ISL_15585890 | A/chicken/Wales/116603/2022 | Animal and Plant Health Agency (APHA) | Animal and Plant Health Agency (APHA) |
| EPI2198479 | MP | United Kingdom | 2022-Sep-04 | EPI_ISL_15585890 | A/chicken/Wales/116603/2022 | Animal and Plant Health Agency (APHA) | Animal and Plant Health Agency (APHA) |
| EPI2198485 | NP | United Kingdom | 2022-Oct-17 | EPI_ISL_15585891 | A/domestic_goose/England/120019/2022 | Animal and Plant Health Agency (APHA) | Animal and Plant Health Agency (APHA) |
| EPI2198486 | NS | United Kingdom | 2022-Oct-17 | EPI_ISL_15585891 | A/domestic_goose/England/120019/2022 | Animal and Plant Health Agency (APHA) | Animal and Plant Health Agency (APHA) |
| EPI2198487 | MP | United Kingdom | 2022-Oct-17 | EPI_ISL_15585891 | A/domestic_goose/England/120019/2022 | Animal and Plant Health Agency (APHA) | Animal and Plant Health Agency (APHA) |
| EPI2198493 | NP | United Kingdom | 2022-Sep-21 | EPI_ISL_15585892 | A/domestic_goose/England/125128/2022 | Animal and Plant Health Agency (APHA) | Animal and Plant Health Agency (APHA) |
| EPI2198494 | NS | United Kingdom | 2022-Sep-21 | EPI_ISL_15585892 | A/domestic_goose/England/125128/2022 | Animal and Plant Health Agency (APHA) | Animal and Plant Health Agency (APHA) |
| EPI2198495 | MP | United Kingdom | 2022-Sep-21 | EPI_ISL_15585892 | A/domestic_goose/England/125128/2022 | Animal and Plant Health Agency (APHA) | Animal and Plant Health Agency (APHA) |
| EPI2198501 | NP | United Kingdom | 2022-Oct-03 | EPI_ISL_15585893 | A/domestic_goose/England/125496/2022 | Animal and Plant Health Agency (APHA) | Animal and Plant Health Agency (APHA) |
| EPI2198502 | NS | United Kingdom | 2022-Oct-03 | EPI_ISL_15585893 | A/domestic_goose/England/125496/2022 | Animal and Plant Health Agency (APHA) | Animal and Plant Health Agency (APHA) |
| EPI2198503 | MP | United Kingdom | 2022-Oct-03 | EPI_ISL_15585893 | A/domestic_goose/England/125496/2022 | Animal and Plant Health Agency (APHA) | Animal and Plant Health Agency (APHA) |
| EPI2198517 | NP | United Kingdom | 2022-Oct-14 | EPI_ISL_15585895 | A/domestic_goose/England/132400/2022 | Animal and Plant Health Agency (APHA) | Animal and Plant Health Agency (APHA) |
| EPI2198518 | NS | United Kingdom | 2022-Oct-14 | EPI_ISL_15585895 | A/domestic_goose/England/132400/2022 | Animal and Plant Health Agency (APHA) | Animal and Plant Health Agency (APHA) |
| EPI2198519 | MP | United Kingdom | 2022-Oct-14 | EPI_ISL_15585895 | A/domestic_goose/England/132400/2022 | Animal and Plant Health Agency (APHA) | Animal and Plant Health Agency (APHA) |
| EPI2198525 | NP | United Kingdom | 2022-Sep-12 | EPI_ISL_15585896 | A/greylag_goose/England/125886/2022 | Animal and Plant Health Agency (APHA) | Animal and Plant Health Agency (APHA) |
| EPI2198526 | NS | United Kingdom | 2022-Sep-12 | EPI_ISL_15585896 | A/greylag_goose/England/125886/2022 | Animal and Plant Health Agency (APHA) | Animal and Plant Health Agency (APHA) |
| EPI2198527 | MP | United Kingdom | 2022-Sep-12 | EPI_ISL_15585896 | A/greylag_goose/England/125886/2022 | Animal and Plant Health Agency (APHA) | Animal and Plant Health Agency (APHA) |
| EPI2198533 | NP | United Kingdom | 2022-Oct-03 | EPI_ISL_15585897 | A/greylag_goose/England/312865/2022 | Animal and Plant Health Agency (APHA) | Animal and Plant Health Agency (APHA) |
| EPI2198534 | NS | United Kingdom | 2022-Oct-03 | EPI_ISL_15585897 | A/greylag_goose/England/312865/2022 | Animal and Plant Health Agency (APHA) | Animal and Plant Health Agency (APHA) |
| EPI2198535 | MP | United Kingdom | 2022-Oct-03 | EPI_ISL_15585897 | A/greylag_goose/England/312865/2022 | Animal and Plant Health Agency (APHA) | Animal and Plant Health Agency (APHA) |
| EPI2198557 | NP | United Kingdom | 2022-Sep-29 | EPI_ISL_15585900 | A/herring_gull/England/324803/2022 | Animal and Plant Health Agency (APHA) | Animal and Plant Health Agency (APHA) |
| EPI2198558 | NS | United Kingdom | 2022-Sep-29 | EPI_ISL_15585900 | A/herring_gull/England/324803/2022 | Animal and Plant Health Agency (APHA) | Animal and Plant Health Agency (APHA) |
| EPI2198559 | MP | United Kingdom | 2022-Sep-29 | EPI_ISL_15585900 | A/herring_gull/England/324803/2022 | Animal and Plant Health Agency (APHA) | Animal and Plant Health Agency (APHA) |
| EPI2198589 | NP | United Kingdom | 2022-Sep-05 | EPI_ISL_15585904 | A/pheasant/England/132162/2022 | Animal and Plant Health Agency (APHA) | Animal and Plant Health Agency (APHA) |
| EPI2198590 | NS | United Kingdom | 2022-Sep-05 | EPI_ISL_15585904 | A/pheasant/England/132162/2022 | Animal and Plant Health Agency (APHA) | Animal and Plant Health Agency (APHA) |
| EPI2198591 | MP | United Kingdom | 2022-Sep-05 | EPI_ISL_15585904 | A/pheasant/England/132162/2022 | Animal and Plant Health Agency (APHA) | Animal and Plant Health Agency (APHA) |
| EPI2198597 | NP | United Kingdom | 2022-Oct-15 | EPI_ISL_15585905 | A/pheasant/England/251536/2022 | Animal and Plant Health Agency (APHA) | Animal and Plant Health Agency (APHA) |
| EPI2198598 | NS | United Kingdom | 2022-Oct-15 | EPI_ISL_15585905 | A/pheasant/England/251536/2022 | Animal and Plant Health Agency (APHA) | Animal and Plant Health Agency (APHA) |
| EPI2198599 | MP | United Kingdom | 2022-Oct-15 | EPI_ISL_15585905 | A/pheasant/England/251536/2022 | Animal and Plant Health Agency (APHA) | Animal and Plant Health Agency (APHA) |
| EPI2198605 | NP | United Kingdom | 2022-Aug-26 | EPI_ISL_15585906 | A/pheasant/England/393437/2022 | Animal and Plant Health Agency (APHA) | Animal and Plant Health Agency (APHA) |
| EPI2198606 | NS | United Kingdom | 2022-Aug-26 | EPI_ISL_15585906 | A/pheasant/England/393437/2022 | Animal and Plant Health Agency (APHA) | Animal and Plant Health Agency (APHA) |
| EPI2198607 | MP | United Kingdom | 2022-Aug-26 | EPI_ISL_15585906 | A/pheasant/England/393437/2022 | Animal and Plant Health Agency (APHA) | Animal and Plant Health Agency (APHA) |

| | | | | | | | |
|---|---|---|---|---|---|---|---|
| EPI2198621 | NP | United Kingdom | 2022-Sep-26 | EPI_ISL_15585908 | A/turkey/England/125196/2022 | Animal and Plant Health Agency (APHA) | Animal and Plant Health Agency (APHA) |
| EPI2198622 | NS | United Kingdom | 2022-Sep-26 | EPI_ISL_15585908 | A/turkey/England/125196/2022 | Animal and Plant Health Agency (APHA) | Animal and Plant Health Agency (APHA) |
| EPI2198623 | MP | United Kingdom | 2022-Sep-26 | EPI_ISL_15585908 | A/turkey/England/125196/2022 | Animal and Plant Health Agency (APHA) | Animal and Plant Health Agency (APHA) |
| EPI2198629 | NP | United Kingdom | 2022-Oct-15 | EPI_ISL_15585909 | A/turkey/England/132310/2022 | Animal and Plant Health Agency (APHA) | Animal and Plant Health Agency (APHA) |
| EPI2198630 | NS | United Kingdom | 2022-Oct-15 | EPI_ISL_15585909 | A/turkey/England/132310/2022 | Animal and Plant Health Agency (APHA) | Animal and Plant Health Agency (APHA) |
| EPI2198631 | MP | United Kingdom | 2022-Oct-15 | EPI_ISL_15585909 | A/turkey/England/132310/2022 | Animal and Plant Health Agency (APHA) | Animal and Plant Health Agency (APHA) |
| EPI2198637 | NP | United Kingdom | 2022-Oct-15 | EPI_ISL_15585910 | A/turkey/England/132460/2022 | Animal and Plant Health Agency (APHA) | Animal and Plant Health Agency (APHA) |
| EPI2198638 | NS | United Kingdom | 2022-Oct-15 | EPI_ISL_15585910 | A/turkey/England/132460/2022 | Animal and Plant Health Agency (APHA) | Animal and Plant Health Agency (APHA) |
| EPI2198639 | MP | United Kingdom | 2022-Oct-15 | EPI_ISL_15585910 | A/turkey/England/132460/2022 | Animal and Plant Health Agency (APHA) | Animal and Plant Health Agency (APHA) |
| EPI2198645 | NP | United Kingdom | 2022-Oct-15 | EPI_ISL_15585911 | A/turkey/England/132948/2022 | Animal and Plant Health Agency (APHA) | Animal and Plant Health Agency (APHA) |
| EPI2198646 | NS | United Kingdom | 2022-Oct-15 | EPI_ISL_15585911 | A/turkey/England/132948/2022 | Animal and Plant Health Agency (APHA) | Animal and Plant Health Agency (APHA) |
| EPI2198647 | MP | United Kingdom | 2022-Oct-15 | EPI_ISL_15585911 | A/turkey/England/132948/2022 | Animal and Plant Health Agency (APHA) | Animal and Plant Health Agency (APHA) |
| EPI2198653 | NP | United Kingdom | 2022-Oct-17 | EPI_ISL_15585919 | A/turkey/England/133112/2022 | Animal and Plant Health Agency (APHA) | Animal and Plant Health Agency (APHA) |
| EPI2198654 | NS | United Kingdom | 2022-Oct-17 | EPI_ISL_15585919 | A/turkey/England/133112/2022 | Animal and Plant Health Agency (APHA) | Animal and Plant Health Agency (APHA) |
| EPI2198655 | MP | United Kingdom | 2022-Oct-17 | EPI_ISL_15585919 | A/turkey/England/133112/2022 | Animal and Plant Health Agency (APHA) | Animal and Plant Health Agency (APHA) |
| EPI2198661 | NP | United Kingdom | 2022-Oct-18 | EPI_ISL_15585920 | A/turkey/Wales/132240/2022 | Animal and Plant Health Agency (APHA) | Animal and Plant Health Agency (APHA) |
| EPI2198662 | NS | United Kingdom | 2022-Oct-18 | EPI_ISL_15585920 | A/turkey/Wales/132240/2022 | Animal and Plant Health Agency (APHA) | Animal and Plant Health Agency (APHA) |
| EPI2198663 | MP | United Kingdom | 2022-Oct-18 | EPI_ISL_15585920 | A/turkey/Wales/132240/2022 | Animal and Plant Health Agency (APHA) | Animal and Plant Health Agency (APHA) |
| EPI2198671 | MP | United Kingdom | 2022-Oct-10 | EPI_ISL_15586107 | A/Bufflehead_Duck/Northern_Ireland/012321/2022 | Animal and Plant Health Agency (APHA) | Animal and Plant Health Agency (APHA) |
| EPI2198669 | NP | United Kingdom | 2022-Oct-10 | EPI_ISL_15586107 | A/Bufflehead_Duck/Northern_Ireland/012321/2022 | Animal and Plant Health Agency (APHA) | Animal and Plant Health Agency (APHA) |
| EPI2198670 | NS | United Kingdom | 2022-Oct-10 | EPI_ISL_15586107 | A/Bufflehead_Duck/Northern_Ireland/012321/2022 | Animal and Plant Health Agency (APHA) | Animal and Plant Health Agency (APHA) |
| EPI2198677 | NP | United Kingdom | 2022-Oct-07 | EPI_ISL_15586108 | A/Carolina_Duck/England/128445/2022 | Animal and Plant Health Agency (APHA) | Animal and Plant Health Agency (APHA) |
| EPI2198678 | NS | United Kingdom | 2022-Oct-07 | EPI_ISL_15586108 | A/Carolina_Duck/England/128445/2022 | Animal and Plant Health Agency (APHA) | Animal and Plant Health Agency (APHA) |
| EPI2198679 | MP | United Kingdom | 2022-Oct-07 | EPI_ISL_15586108 | A/Carolina_Duck/England/128445/2022 | Animal and Plant Health Agency (APHA) | Animal and Plant Health Agency (APHA) |
| EPI2198685 | NP | United Kingdom | 2022-Oct-06 | EPI_ISL_15586109 | A/Domestic_Duck/England/128223/2022 | Animal and Plant Health Agency (APHA) | Animal and Plant Health Agency (APHA) |
| EPI2198686 | NS | United Kingdom | 2022-Oct-06 | EPI_ISL_15586109 | A/Domestic_Duck/England/128223/2022 | Animal and Plant Health Agency (APHA) | Animal and Plant Health Agency (APHA) |
| EPI2198687 | MP | United Kingdom | 2022-Oct-06 | EPI_ISL_15586109 | A/Domestic_Duck/England/128223/2022 | Animal and Plant Health Agency (APHA) | Animal and Plant Health Agency (APHA) |
| EPI2198693 | NP | United Kingdom | 2022-Oct-08 | EPI_ISL_15586110 | A/Domestic_Duck/England/128591/2022 | Animal and Plant Health Agency (APHA) | Animal and Plant Health Agency (APHA) |
| EPI2198694 | NS | United Kingdom | 2022-Oct-08 | EPI_ISL_15586110 | A/Domestic_Duck/England/128591/2022 | Animal and Plant Health Agency (APHA) | Animal and Plant Health Agency (APHA) |
| EPI2198695 | MP | United Kingdom | 2022-Oct-08 | EPI_ISL_15586110 | A/Domestic_Duck/England/128591/2022 | Animal and Plant Health Agency (APHA) | Animal and Plant Health Agency (APHA) |
| EPI2198701 | NP | United Kingdom | 2022-Oct-08 | EPI_ISL_15586111 | A/Domestic_Duck/England/128721/2022 | Animal and Plant Health Agency (APHA) | Animal and Plant Health Agency (APHA) |
| EPI2198702 | NS | United Kingdom | 2022-Oct-08 | EPI_ISL_15586111 | A/Domestic_Duck/England/128721/2022 | Animal and Plant Health Agency (APHA) | Animal and Plant Health Agency (APHA) |
| EPI2198703 | MP | United Kingdom | 2022-Oct-08 | EPI_ISL_15586111 | A/Domestic_Duck/England/128721/2022 | Animal and Plant Health Agency (APHA) | Animal and Plant Health Agency (APHA) |
| EPI2198709 | NP | United Kingdom | 2022-Oct-09 | EPI_ISL_15586112 | A/Domestic_Duck/England/128822/2022 | Animal and Plant Health Agency (APHA) | Animal and Plant Health Agency (APHA) |
| EPI2198710 | NS | United Kingdom | 2022-Oct-09 | EPI_ISL_15586112 | A/Domestic_Duck/England/128822/2022 | Animal and Plant Health Agency (APHA) | Animal and Plant Health Agency (APHA) |

| | | | | | | | |
|---|---|---|---|---|---|---|---|
| EPI2198711 | MP | United Kingdom | 2022-Oct-09 | EPI_ISL_15586112 | A/Domestic_Duck/England/128822/2022 | Animal and Plant Health Agency (APHA) | Animal and Plant Health Agency (APHA) |
| EPI2198717 | NP | United Kingdom | 2022-Oct-09 | EPI_ISL_15586113 | A/Domestic_Duck/England/128927/2022 | Animal and Plant Health Agency (APHA) | Animal and Plant Health Agency (APHA) |
| EPI2198718 | NS | United Kingdom | 2022-Oct-09 | EPI_ISL_15586113 | A/Domestic_Duck/England/128927/2022 | Animal and Plant Health Agency (APHA) | Animal and Plant Health Agency (APHA) |
| EPI2198719 | MP | United Kingdom | 2022-Oct-09 | EPI_ISL_15586113 | A/Domestic_Duck/England/128927/2022 | Animal and Plant Health Agency (APHA) | Animal and Plant Health Agency (APHA) |
| EPI2198725 | NP | United Kingdom | 2022-Oct-18 | EPI_ISL_15586114 | A/Domestic_Duck/England/134267/2022 | Animal and Plant Health Agency (APHA) | Animal and Plant Health Agency (APHA) |
| EPI2198726 | NS | United Kingdom | 2022-Oct-18 | EPI_ISL_15586114 | A/Domestic_Duck/England/134267/2022 | Animal and Plant Health Agency (APHA) | Animal and Plant Health Agency (APHA) |
| EPI2198727 | MP | United Kingdom | 2022-Oct-18 | EPI_ISL_15586114 | A/Domestic_Duck/England/134267/2022 | Animal and Plant Health Agency (APHA) | Animal and Plant Health Agency (APHA) |
| EPI2198733 | NP | United Kingdom | 2022-Oct-19 | EPI_ISL_15586115 | A/Domestic_Duck/England/134956/2022 | Animal and Plant Health Agency (APHA) | Animal and Plant Health Agency (APHA) |
| EPI2198734 | NS | United Kingdom | 2022-Oct-19 | EPI_ISL_15586115 | A/Domestic_Duck/England/134956/2022 | Animal and Plant Health Agency (APHA) | Animal and Plant Health Agency (APHA) |
| EPI2198735 | MP | United Kingdom | 2022-Oct-19 | EPI_ISL_15586115 | A/Domestic_Duck/England/134956/2022 | Animal and Plant Health Agency (APHA) | Animal and Plant Health Agency (APHA) |
| EPI2198741 | NP | United Kingdom | 2022-Oct-10 | EPI_ISL_15586116 | A/Nene_Goose/Northern_Ireland/013322/2022 | Animal and Plant Health Agency (APHA) | Animal and Plant Health Agency (APHA) |
| EPI2198742 | NS | United Kingdom | 2022-Oct-10 | EPI_ISL_15586116 | A/Nene_Goose/Northern_Ireland/013322/2022 | Animal and Plant Health Agency (APHA) | Animal and Plant Health Agency (APHA) |
| EPI2198743 | MP | United Kingdom | 2022-Oct-10 | EPI_ISL_15586116 | A/Nene_Goose/Northern_Ireland/013322/2022 | Animal and Plant Health Agency (APHA) | Animal and Plant Health Agency (APHA) |
| EPI2198749 | NP | United Kingdom | 2022-Oct-07 | EPI_ISL_15586117 | A/Whistling_Duck/England/128441/2022 | Animal and Plant Health Agency (APHA) | Animal and Plant Health Agency (APHA) |
| EPI2198750 | NS | United Kingdom | 2022-Oct-07 | EPI_ISL_15586117 | A/Whistling_Duck/England/128441/2022 | Animal and Plant Health Agency (APHA) | Animal and Plant Health Agency (APHA) |
| EPI2198751 | MP | United Kingdom | 2022-Oct-07 | EPI_ISL_15586117 | A/Whistling_Duck/England/128441/2022 | Animal and Plant Health Agency (APHA) | Animal and Plant Health Agency (APHA) |
| EPI2198789 | NP | United Kingdom | 2022-Sep-09 | EPI_ISL_15586122 | A/domestic_duck/England/117887/2022 | Animal and Plant Health Agency (APHA) | Animal and Plant Health Agency (APHA) |
| EPI2198790 | NS | United Kingdom | 2022-Sep-09 | EPI_ISL_15586122 | A/domestic_duck/England/117887/2022 | Animal and Plant Health Agency (APHA) | Animal and Plant Health Agency (APHA) |
| EPI2198791 | MP | United Kingdom | 2022-Sep-09 | EPI_ISL_15586122 | A/domestic_duck/England/117887/2022 | Animal and Plant Health Agency (APHA) | Animal and Plant Health Agency (APHA) |
| EPI2198805 | NP | United Kingdom | 2022-Sep-20 | EPI_ISL_15586124 | A/domestic_duck/England/125290/2022 | Animal and Plant Health Agency (APHA) | Animal and Plant Health Agency (APHA) |
| EPI2198806 | NS | United Kingdom | 2022-Sep-20 | EPI_ISL_15586124 | A/domestic_duck/England/125290/2022 | Animal and Plant Health Agency (APHA) | Animal and Plant Health Agency (APHA) |
| EPI2198807 | MP | United Kingdom | 2022-Sep-20 | EPI_ISL_15586124 | A/domestic_duck/England/125290/2022 | Animal and Plant Health Agency (APHA) | Animal and Plant Health Agency (APHA) |
| EPI2198813 | NP | United Kingdom | 2022-Oct-02 | EPI_ISL_15586125 | A/domestic_duck/England/129575/2022 | Animal and Plant Health Agency (APHA) | Animal and Plant Health Agency (APHA) |
| EPI2198814 | NS | United Kingdom | 2022-Oct-02 | EPI_ISL_15586125 | A/domestic_duck/England/129575/2022 | Animal and Plant Health Agency (APHA) | Animal and Plant Health Agency (APHA) |
| EPI2198815 | MP | United Kingdom | 2022-Oct-02 | EPI_ISL_15586125 | A/domestic_duck/England/129575/2022 | Animal and Plant Health Agency (APHA) | Animal and Plant Health Agency (APHA) |
| EPI2198823 | MP | United Kingdom | 2022-Oct-12 | EPI_ISL_15586126 | A/domestic_duck/England/130575/2022 | Animal and Plant Health Agency (APHA) | Animal and Plant Health Agency (APHA) |
| EPI2198821 | NP | United Kingdom | 2022-Oct-12 | EPI_ISL_15586126 | A/domestic_duck/England/130575/2022 | Animal and Plant Health Agency (APHA) | Animal and Plant Health Agency (APHA) |
| EPI2198822 | NS | United Kingdom | 2022-Oct-12 | EPI_ISL_15586126 | A/domestic_duck/England/130575/2022 | Animal and Plant Health Agency (APHA) | Animal and Plant Health Agency (APHA) |
| EPI2198829 | NP | United Kingdom | 2022-Oct-12 | EPI_ISL_15586127 | A/domestic_duck/England/131065/2022 | Animal and Plant Health Agency (APHA) | Animal and Plant Health Agency (APHA) |
| EPI2198830 | NS | United Kingdom | 2022-Oct-12 | EPI_ISL_15586127 | A/domestic_duck/England/131065/2022 | Animal and Plant Health Agency (APHA) | Animal and Plant Health Agency (APHA) |
| EPI2198831 | MP | United Kingdom | 2022-Oct-12 | EPI_ISL_15586127 | A/domestic_duck/England/131065/2022 | Animal and Plant Health Agency (APHA) | Animal and Plant Health Agency (APHA) |
| EPI2198837 | NP | United Kingdom | 2022-Oct-13 | EPI_ISL_15586128 | A/domestic_duck/England/132948/2022 | Animal and Plant Health Agency (APHA) | Animal and Plant Health Agency (APHA) |
| EPI2198838 | NS | United Kingdom | 2022-Oct-13 | EPI_ISL_15586128 | A/domestic_duck/England/132948/2022 | Animal and Plant Health Agency (APHA) | Animal and Plant Health Agency (APHA) |
| EPI2198839 | MP | United Kingdom | 2022-Oct-13 | EPI_ISL_15586128 | A/domestic_duck/England/132948/2022 | Animal and Plant Health Agency (APHA) | Animal and Plant Health Agency (APHA) |
| EPI2198869 | NP | United Kingdom | 2022-Sep-02 | EPI_ISL_15586132 | A/mute_Swan/Scotland/126261/2022 | Animal and Plant Health Agency (APHA) | Animal and Plant Health Agency (APHA) |

| | | | | | | | |
|---|---|---|---|---|---|---|---|
| EPI2198870 | NS | United Kingdom | 2022-Sep-02 | EPI_ISL_15586132 | A/mute_Swan/Scotland/126261/2022 | Animal and Plant Health Agency (APHA) | Animal and Plant Health Agency (APHA) |
| EPI2198871 | MP | United Kingdom | 2022-Sep-02 | EPI_ISL_15586132 | A/mute_Swan/Scotland/126261/2022 | Animal and Plant Health Agency (APHA) | Animal and Plant Health Agency (APHA) |
| EPI2198878 | NS | United Kingdom | 2022-Sep-18 | EPI_ISL_15586133 | A/mute_swan/England/117298/2022 | Animal and Plant Health Agency (APHA) | Animal and Plant Health Agency (APHA) |
| EPI2198879 | MP | United Kingdom | 2022-Sep-18 | EPI_ISL_15586133 | A/mute_swan/England/117298/2022 | Animal and Plant Health Agency (APHA) | Animal and Plant Health Agency (APHA) |
| EPI2198877 | NP | United Kingdom | 2022-Sep-18 | EPI_ISL_15586133 | A/mute_swan/England/117298/2022 | Animal and Plant Health Agency (APHA) | Animal and Plant Health Agency (APHA) |
| EPI2198885 | NP | United Kingdom | 2022-Sep-18 | EPI_ISL_15586134 | A/mute_swan/England/125329/2022 | Animal and Plant Health Agency (APHA) | Animal and Plant Health Agency (APHA) |
| EPI2198886 | NS | United Kingdom | 2022-Sep-18 | EPI_ISL_15586134 | A/mute_swan/England/125329/2022 | Animal and Plant Health Agency (APHA) | Animal and Plant Health Agency (APHA) |
| EPI2198887 | MP | United Kingdom | 2022-Sep-18 | EPI_ISL_15586134 | A/mute_swan/England/125329/2022 | Animal and Plant Health Agency (APHA) | Animal and Plant Health Agency (APHA) |
| EPI2198893 | NP | United Kingdom | 2022-Sep-27 | EPI_ISL_15586135 | A/mute_swan/England/253063/2022 | Animal and Plant Health Agency (APHA) | Animal and Plant Health Agency (APHA) |
| EPI2198894 | NS | United Kingdom | 2022-Sep-27 | EPI_ISL_15586135 | A/mute_swan/England/253063/2022 | Animal and Plant Health Agency (APHA) | Animal and Plant Health Agency (APHA) |
| EPI2198895 | MP | United Kingdom | 2022-Sep-27 | EPI_ISL_15586135 | A/mute_swan/England/253063/2022 | Animal and Plant Health Agency (APHA) | Animal and Plant Health Agency (APHA) |
| EPI2198901 | NP | United Kingdom | 2022-Oct-03 | EPI_ISL_15586136 | A/mute_swan/England/328136/2022 | Animal and Plant Health Agency (APHA) | Animal and Plant Health Agency (APHA) |
| EPI2198902 | NS | United Kingdom | 2022-Oct-03 | EPI_ISL_15586136 | A/mute_swan/England/328136/2022 | Animal and Plant Health Agency (APHA) | Animal and Plant Health Agency (APHA) |
| EPI2198903 | MP | United Kingdom | 2022-Oct-03 | EPI_ISL_15586136 | A/mute_swan/England/328136/2022 | Animal and Plant Health Agency (APHA) | Animal and Plant Health Agency (APHA) |
| EPI2208349 | MP | Belgium | 2022-Sep-18 | EPI_ISL_15697406 | A/Pavo/Belgium/10906_0006/2022 | Sciensano - Animal Infectious Diseases | Sciensano, Department of Animal Infectious Diseases |
| EPI2208350 | NS | Belgium | 2022-Sep-18 | EPI_ISL_15697406 | A/Pavo/Belgium/10906_0006/2022 | Sciensano - Animal Infectious Diseases | Sciensano, Department of Animal Infectious Diseases |
| EPI2208347 | NP | Belgium | 2022-Sep-18 | EPI_ISL_15697406 | A/Pavo/Belgium/10906_0006/2022 | Sciensano - Animal Infectious Diseases | Sciensano, Department of Animal Infectious Diseases |
| EPI2208444 | NP | Belgium | 2022-Sep-19 | EPI_ISL_15697407 | A/Gallus_gallus/Belgium/10968_0005/2022 | Sciensano - Animal Infectious Diseases | Sciensano, Department of Animal Infectious Diseases |
| EPI2208494 | MP | Belgium | 2022-Sep-19 | EPI_ISL_15697407 | A/Gallus_gallus/Belgium/10968_0005/2022 | Sciensano - Animal Infectious Diseases | Sciensano, Department of Animal Infectious Diseases |
| EPI2208532 | NS | Belgium | 2022-Sep-19 | EPI_ISL_15697407 | A/Gallus_gallus/Belgium/10968_0005/2022 | Sciensano - Animal Infectious Diseases | Sciensano, Department of Animal Infectious Diseases |
| EPI2208896 | NP | Belgium | 2022-Sep-26 | EPI_ISL_15698209 | A/Gallus_gallus/Belgium/11248_0001/2022 | Sciensano - Animal Infectious Diseases | Sciensano, Department of Animal Infectious Diseases |
| EPI2208898 | MP | Belgium | 2022-Sep-26 | EPI_ISL_15698209 | A/Gallus_gallus/Belgium/11248_0001/2022 | Sciensano - Animal Infectious Diseases | Sciensano, Department of Animal Infectious Diseases |
| EPI2208899 | NS | Belgium | 2022-Sep-26 | EPI_ISL_15698209 | A/Gallus_gallus/Belgium/11248_0001/2022 | Sciensano - Animal Infectious Diseases | Sciensano, Department of Animal Infectious Diseases |
| EPI2208906 | MP | Belgium | 2022-Sep-23 | EPI_ISL_15698370 | A/Ardea_cinerea/Belgium/11304_0003/2022 | Sciensano - Animal Infectious Diseases | Sciensano, Department of Animal Infectious Diseases |
| EPI2208904 | NP | Belgium | 2022-Sep-23 | EPI_ISL_15698370 | A/Ardea_cinerea/Belgium/11304_0003/2022 | Sciensano - Animal Infectious Diseases | Sciensano, Department of Animal Infectious Diseases |
| EPI2208907 | NS | Belgium | 2022-Sep-23 | EPI_ISL_15698370 | A/Ardea_cinerea/Belgium/11304_0003/2022 | Sciensano - Animal Infectious Diseases | Sciensano, Department of Animal Infectious Diseases |
| EPI2208912 | NP | Belgium | 2022-Sep-21 | EPI_ISL_15699041 | A/Cygnus_olor/Belgium/11304_0009/2022 | Sciensano - Animal Infectious Diseases | Sciensano, Department of Animal Infectious Diseases |
| EPI2208914 | MP | Belgium | 2022-Sep-21 | EPI_ISL_15699041 | A/Cygnus_olor/Belgium/11304_0009/2022 | Sciensano - Animal Infectious Diseases | Sciensano, Department of Animal Infectious Diseases |
| EPI2208915 | NS | Belgium | 2022-Sep-21 | EPI_ISL_15699041 | A/Cygnus_olor/Belgium/11304_0009/2022 | Sciensano - Animal Infectious Diseases | Sciensano, Department of Animal Infectious Diseases |
| EPI2208922 | MP | Belgium | 2022-Sep-29 | EPI_ISL_15699790 | A/Gallus_gallus/Belgium/11387_0002/2022 | Sciensano - Animal Infectious Diseases | Sciensano, Department of Animal Infectious Diseases |
| EPI2208920 | NP | Belgium | 2022-Sep-29 | EPI_ISL_15699790 | A/Gallus_gallus/Belgium/11387_0002/2022 | Sciensano - Animal Infectious Diseases | Sciensano, Department of Animal Infectious Diseases |
| EPI2208923 | NS | Belgium | 2022-Sep-29 | EPI_ISL_15699790 | A/Gallus_gallus/Belgium/11387_0002/2022 | Sciensano - Animal Infectious Diseases | Sciensano, Department of Animal Infectious Diseases |
| EPI2208930 | MP | Belgium | 2022-Oct-01 | EPI_ISL_15699799 | A/Gallus_gallus/Belgium/11445_0002/2022 | Sciensano - Animal Infectious Diseases | Sciensano, Department of Animal Infectious Diseases |
| EPI2208931 | NS | Belgium | 2022-Oct-01 | EPI_ISL_15699799 | A/Gallus_gallus/Belgium/11445_0002/2022 | Sciensano - Animal Infectious Diseases | Sciensano, Department of Animal Infectious Diseases |
| EPI2208928 | NP | Belgium | 2022-Oct-01 | EPI_ISL_15699799 | A/Gallus_gallus/Belgium/11445_0002/2022 | Sciensano - Animal Infectious Diseases | Sciensano, Department of Animal Infectious Diseases |

| | | | | | | | | |
|---|---|---|---|---|---|---|---|---|
| EPI2208936 | NP | Belgium | 2022-Oct-10 | EPI_ISL_15699929 | A/Gallus_gallus/Belgium/11943_0003/2022 | Sciensano - Animal Infectious Diseases | Sciensano, Department of Animal Infectious Diseases | |
| EPI2208938 | MP | Belgium | 2022-Oct-10 | EPI_ISL_15699929 | A/Gallus_gallus/Belgium/11943_0003/2022 | Sciensano - Animal Infectious Diseases | Sciensano, Department of Animal Infectious Diseases | |
| EPI2208939 | NS | Belgium | 2022-Oct-10 | EPI_ISL_15699929 | A/Gallus_gallus/Belgium/11943_0003/2022 | Sciensano - Animal Infectious Diseases | Sciensano, Department of Animal Infectious Diseases | |
| EPI2214465 | NP | Netherlands | 2022-Oct-22 | EPI_ISL_15759949 | A/Common Teal/Netherlands/3/2022 | Erasmus Medical Center | Erasmus Medical Center | |
| EPI2214466 | NS | Netherlands | 2022-Oct-22 | EPI_ISL_15759949 | A/Common Teal/Netherlands/3/2022 | Erasmus Medical Center | Erasmus Medical Center | |
| EPI2214467 | MP | Netherlands | 2022-Oct-22 | EPI_ISL_15759949 | A/Common Teal/Netherlands/3/2022 | Erasmus Medical Center | Erasmus Medical Center | |
| EPI2214472 | NP | Netherlands | 2022-Oct-31 | EPI_ISL_15759950 | A/Common Buzzard/Netherlands/2/2022 | Erasmus Medical Center | Erasmus Medical Center | |
| EPI2214473 | NS | Netherlands | 2022-Oct-31 | EPI_ISL_15759950 | A/Common Buzzard/Netherlands/2/2022 | Erasmus Medical Center | Erasmus Medical Center | |
| EPI2214474 | MP | Netherlands | 2022-Oct-31 | EPI_ISL_15759950 | A/Common Buzzard/Netherlands/2/2022 | Erasmus Medical Center | Erasmus Medical Center | |
| EPI2214480 | NP | Netherlands | 2022-Oct-18 | EPI_ISL_15759951 | A/Mallard/Netherlands/24/2022 | Erasmus Medical Center | Erasmus Medical Center | |
| EPI2214481 | NS | Netherlands | 2022-Oct-18 | EPI_ISL_15759951 | A/Mallard/Netherlands/24/2022 | Erasmus Medical Center | Erasmus Medical Center | |
| EPI2214482 | MP | Netherlands | 2022-Oct-18 | EPI_ISL_15759951 | A/Mallard/Netherlands/24/2022 | Erasmus Medical Center | Erasmus Medical Center | |
| EPI2214493 | NP | Netherlands | 2022-Oct-25 | EPI_ISL_15759953 | A/Mallard/Netherlands/26/2022 | Erasmus Medical Center | Erasmus Medical Center | |
| EPI2214494 | NS | Netherlands | 2022-Oct-25 | EPI_ISL_15759953 | A/Mallard/Netherlands/26/2022 | Erasmus Medical Center | Erasmus Medical Center | |
| EPI2214495 | MP | Netherlands | 2022-Oct-25 | EPI_ISL_15759953 | A/Mallard/Netherlands/26/2022 | Erasmus Medical Center | Erasmus Medical Center | |
| EPI2218142 | NP | Belgium | 2022-Sep-16 | EPI_ISL_15812579 | A/Anser_anser_domesticus/Belgium/10918_0002/2022 | Sciensano - Animal Infectious Diseases | Sciensano, Department of Animal Infectious Diseases | Van Borm, Steven; Roupie, Virginie; Lambrecht, Benedicte; Steensels, Mieke |
| EPI2218144 | MP | Belgium | 2022-Sep-16 | EPI_ISL_15812579 | A/Anser_anser_domesticus/Belgium/10918_0002/2022 | Sciensano - Animal Infectious Diseases | Sciensano, Department of Animal Infectious Diseases | Van Borm, Steven; Roupie, Virginie; Lambrecht, Benedicte; Steensels, Mieke |
| EPI2218145 | NS | Belgium | 2022-Sep-16 | EPI_ISL_15812579 | A/Anser_anser_domesticus/Belgium/10918_0002/2022 | Sciensano - Animal Infectious Diseases | Sciensano, Department of Animal Infectious Diseases | Van Borm, Steven; Roupie, Virginie; Lambrecht, Benedicte; Steensels, Mieke |
| EPI2218155 | NS | Belgium | 2022-Oct-04 | EPI_ISL_15812678 | A/Anas_platyrhynchos_domesticus/Belgium/11599_0005/2022 | Sciensano - Animal Infectious Diseases | Sciensano, Department of Animal Infectious Diseases | Van Borm, Steven; Mathijs, Elisabeth; Roupie, Virginie; Lambrecht, Benedicte; Steensels, Mieke |
| EPI2218154 | MP | Belgium | 2022-Oct-04 | EPI_ISL_15812678 | A/Anas_platyrhynchos_domesticus/Belgium/11599_0005/2022 | Sciensano - Animal Infectious Diseases | Sciensano, Department of Animal Infectious Diseases | Van Borm, Steven; Mathijs, Elisabeth; Roupie, Virginie; Lambrecht, Benedicte; Steensels, Mieke |
| EPI2218152 | NP | Belgium | 2022-Oct-04 | EPI_ISL_15812678 | A/Anas_platyrhynchos_domesticus/Belgium/11599_0005/2022 | Sciensano - Animal Infectious Diseases | Sciensano, Department of Animal Infectious Diseases | Van Borm, Steven; Mathijs, Elisabeth; Roupie, Virginie; Lambrecht, Benedicte; Steensels, Mieke |
| EPI2220393 | MP | France | 2022-Nov-08 | EPI_ISL_15857992 | A/chicken/France/22P023203/2022 | Anses (Ploufragan-Plouzané) | ANSES Agence Nationale De Securite Sanitaire De L'alimentation | |
| EPI2220391 | NP | France | 2022-Nov-08 | EPI_ISL_15857992 | A/chicken/France/22P023203/2022 | Anses (Ploufragan-Plouzané) | ANSES Agence Nationale De Securite Sanitaire De L'alimentation | |
| EPI2220394 | NS | France | 2022-Nov-08 | EPI_ISL_15857992 | A/chicken/France/22P023203/2022 | Anses (Ploufragan-Plouzané) | ANSES Agence Nationale De Securite Sanitaire De L'alimentation | |
| EPI2222969 | NP | Netherlands | 2022-Oct-31 | EPI_ISL_15909904 | A/Barnacle Goose/Netherlands/22019555-001/2022 | Wageningen Bioveterinary Research | Wageningen Bioveterinary Research | Beerens, Nancy; Harders, Frank; Pritz-Verschuren, Sylvia; Roose, Marit; Venema, Sandra; Germeraad, Evelien; Engelsma, Marc; Heutink, Rene |
| EPI2222970 | NS | Netherlands | 2022-Oct-31 | EPI_ISL_15909904 | A/Barnacle Goose/Netherlands/22019555-001/2022 | Wageningen Bioveterinary Research | Wageningen Bioveterinary Research | Beerens, Nancy; Harders, Frank; Pritz-Verschuren, Sylvia; Roose, Marit; Venema, Sandra; Germeraad, Evelien; Engelsma, Marc; Heutink, Rene |
| EPI2222971 | MP | Netherlands | 2022-Oct-31 | EPI_ISL_15909904 | A/Barnacle Goose/Netherlands/22019555-001/2022 | Wageningen Bioveterinary Research | Wageningen Bioveterinary Research | Beerens, Nancy; Harders, Frank; Pritz-Verschuren, Sylvia; Roose, Marit; Venema, Sandra; Germeraad, Evelien; Engelsma, Marc; Heutink, Rene |
| EPI2223234 | NP | Netherlands | 2022-Oct-29 | EPI_ISL_15925875 | A/Eurasian Wigeon/Netherlands/1/2022 | Erasmus Medical Center | Erasmus Medical Center | |
| EPI2223235 | NS | Netherlands | 2022-Oct-29 | EPI_ISL_15925875 | A/Eurasian Wigeon/Netherlands/1/2022 | Erasmus Medical Center | Erasmus Medical Center | |
| EPI2223236 | MP | Netherlands | 2022-Oct-29 | EPI_ISL_15925875 | A/Eurasian Wigeon/Netherlands/1/2022 | Erasmus Medical Center | Erasmus Medical Center | |
| EPI2223242 | NP | Netherlands | 2022-Oct-29 | EPI_ISL_15925876 | A/Eurasian Wigeon/Netherlands/2/2022 | Erasmus Medical Center | Erasmus Medical Center | |
| EPI2223243 | NS | Netherlands | 2022-Oct-29 | EPI_ISL_15925876 | A/Eurasian Wigeon/Netherlands/2/2022 | Erasmus Medical Center | Erasmus Medical Center | |
| EPI2223244 | MP | Netherlands | 2022-Oct-29 | EPI_ISL_15925876 | A/Eurasian Wigeon/Netherlands/2/2022 | Erasmus Medical Center | Erasmus Medical Center | |
| EPI2223257 | NP | Netherlands | 2022-Nov-01 | EPI_ISL_15925878 | A/Common Teal/Netherlands/5/2022 | Erasmus Medical Center | Erasmus Medical Center | |
| EPI2223258 | NS | Netherlands | 2022-Nov-01 | EPI_ISL_15925878 | A/Common Teal/Netherlands/5/2022 | Erasmus Medical Center | Erasmus Medical Center | |

| | | | | | | | | |
|---|---|---|---|---|---|---|---|---|
| EPI2223259 | MP | Netherlands | 2022-Nov-01 | EPI_ISL_15925878 | A/Common Teal/Netherlands/5/2022 | Erasmus Medical Center | Erasmus Medical Center | |
| EPI2223272 | NP | Netherlands | 2022-Nov-09 | EPI_ISL_15925880 | A/Gadwall/Netherlands/1/2022 | Erasmus Medical Center | Erasmus Medical Center | |
| EPI2223273 | NS | Netherlands | 2022-Nov-09 | EPI_ISL_15925880 | A/Gadwall/Netherlands/1/2022 | Erasmus Medical Center | Erasmus Medical Center | |
| EPI2223274 | MP | Netherlands | 2022-Nov-09 | EPI_ISL_15925880 | A/Gadwall/Netherlands/1/2022 | Erasmus Medical Center | Erasmus Medical Center | |
| EPI2223280 | NP | Netherlands | 2022-Nov-09 | EPI_ISL_15925881 | A/Gadwall/Netherlands/2/2022 | Erasmus Medical Center | Erasmus Medical Center | |
| EPI2223281 | NS | Netherlands | 2022-Nov-09 | EPI_ISL_15925881 | A/Gadwall/Netherlands/2/2022 | Erasmus Medical Center | Erasmus Medical Center | |
| EPI2223282 | MP | Netherlands | 2022-Nov-09 | EPI_ISL_15925881 | A/Gadwall/Netherlands/2/2022 | Erasmus Medical Center | Erasmus Medical Center | |
| EPI2223288 | NP | Netherlands | 2022-Oct-21 | EPI_ISL_15925882 | A/Eurasian Wigeon/Netherlands/3/2022 | Erasmus Medical Center | Erasmus Medical Center | |
| EPI2223289 | NS | Netherlands | 2022-Oct-21 | EPI_ISL_15925882 | A/Eurasian Wigeon/Netherlands/3/2022 | Erasmus Medical Center | Erasmus Medical Center | |
| EPI2223290 | MP | Netherlands | 2022-Oct-21 | EPI_ISL_15925882 | A/Eurasian Wigeon/Netherlands/3/2022 | Erasmus Medical Center | Erasmus Medical Center | |
| EPI2223296 | NP | Netherlands | 2022-Nov-01 | EPI_ISL_15925883 | A/Eurasian Wigeon/Netherlands/4/2022 | Erasmus Medical Center | Erasmus Medical Center | |
| EPI2223297 | NS | Netherlands | 2022-Nov-01 | EPI_ISL_15925883 | A/Eurasian Wigeon/Netherlands/4/2022 | Erasmus Medical Center | Erasmus Medical Center | |
| EPI2223298 | MP | Netherlands | 2022-Nov-01 | EPI_ISL_15925883 | A/Eurasian Wigeon/Netherlands/4/2022 | Erasmus Medical Center | Erasmus Medical Center | |
| EPI2223304 | NP | Netherlands | 2022-Nov-11 | EPI_ISL_15925884 | A/Great Cormorant/Netherlands/1/2022 | Erasmus Medical Center | Erasmus Medical Center | |
| EPI2223305 | NS | Netherlands | 2022-Nov-11 | EPI_ISL_15925884 | A/Great Cormorant/Netherlands/1/2022 | Erasmus Medical Center | Erasmus Medical Center | |
| EPI2223306 | MP | Netherlands | 2022-Nov-11 | EPI_ISL_15925884 | A/Great Cormorant/Netherlands/1/2022 | Erasmus Medical Center | Erasmus Medical Center | |
| EPI2223312 | NP | Netherlands | 2022-Nov-11 | EPI_ISL_15925885 | A/Black-headed Gull/Netherlands/11/2022 | Erasmus Medical Center | Erasmus Medical Center | |
| EPI2223313 | NS | Netherlands | 2022-Nov-11 | EPI_ISL_15925885 | A/Black-headed Gull/Netherlands/11/2022 | Erasmus Medical Center | Erasmus Medical Center | |
| EPI2223314 | MP | Netherlands | 2022-Nov-11 | EPI_ISL_15925885 | A/Black-headed Gull/Netherlands/11/2022 | Erasmus Medical Center | Erasmus Medical Center | |
| EPI2231267 | NS | United Kingdom | 2022-Nov-15 | EPI_ISL_15942297 | A/domestic_duck/England/152473/2022 | Animal and Plant Health Agency (APHA) | Animal and Plant Health Agency (APHA) | Maskell,Daniel |
| EPI2231264 | NP | United Kingdom | 2022-Nov-15 | EPI_ISL_15942297 | A/domestic_duck/England/152473/2022 | Animal and Plant Health Agency (APHA) | Animal and Plant Health Agency (APHA) | Maskell,Daniel |
| EPI2231266 | MP | United Kingdom | 2022-Nov-15 | EPI_ISL_15942297 | A/domestic_duck/England/152473/2022 | Animal and Plant Health Agency (APHA) | Animal and Plant Health Agency (APHA) | Maskell,Daniel |
| EPI2232643 | NP | United Kingdom | 2022-Oct-19 | EPI_ISL_15957618 | A/turkey/England/137790/2022 | Animal and Plant Health Agency (APHA) | Animal and Plant Health Agency (APHA) | Byrne, A.; Howton, N.; Maskell, D.; Mollett, B.; Peers-Dent, J. |
| EPI2232644 | NS | United Kingdom | 2022-Oct-19 | EPI_ISL_15957618 | A/turkey/England/137790/2022 | Animal and Plant Health Agency (APHA) | Animal and Plant Health Agency (APHA) | Byrne, A.; Howton, N.; Maskell, D.; Mollett, B.; Peers-Dent, J. |
| EPI2232645 | MP | United Kingdom | 2022-Oct-19 | EPI_ISL_15957618 | A/turkey/England/137790/2022 | Animal and Plant Health Agency (APHA) | Animal and Plant Health Agency (APHA) | Byrne, A.; Howton, N.; Maskell, D.; Mollett, B.; Peers-Dent, J. |
| EPI2232651 | NP | United Kingdom | 2022-Oct-18 | EPI_ISL_15957624 | A/turkey/England/137861/2022 | Animal and Plant Health Agency (APHA) | Animal and Plant Health Agency (APHA) | Byrne, A.; Howton, N.; Maskell, D.; Mollett, B.; Peers-Dent, J. |
| EPI2232652 | NS | United Kingdom | 2022-Oct-18 | EPI_ISL_15957624 | A/turkey/England/137861/2022 | Animal and Plant Health Agency (APHA) | Animal and Plant Health Agency (APHA) | Byrne, A.; Howton, N.; Maskell, D.; Mollett, B.; Peers-Dent, J. |
| EPI2232653 | MP | United Kingdom | 2022-Oct-18 | EPI_ISL_15957624 | A/turkey/England/137861/2022 | Animal and Plant Health Agency (APHA) | Animal and Plant Health Agency (APHA) | Byrne, A.; Howton, N.; Maskell, D.; Mollett, B.; Peers-Dent, J. |
| EPI2232667 | NP | United Kingdom | 2022-Oct-20 | EPI_ISL_15957635 | A/turkey/England/137992/2022 | Animal and Plant Health Agency (APHA) | Animal and Plant Health Agency (APHA) | Byrne, A.; Howton, N.; Maskell, D.; Mollett, B.; Peers-Dent, J. |
| EPI2232668 | NS | United Kingdom | 2022-Oct-20 | EPI_ISL_15957635 | A/turkey/England/137992/2022 | Animal and Plant Health Agency (APHA) | Animal and Plant Health Agency (APHA) | Byrne, A.; Howton, N.; Maskell, D.; Mollett, B.; Peers-Dent, J. |
| EPI2232669 | MP | United Kingdom | 2022-Oct-20 | EPI_ISL_15957635 | A/turkey/England/137992/2022 | Animal and Plant Health Agency (APHA) | Animal and Plant Health Agency (APHA) | Byrne, A.; Howton, N.; Maskell, D.; Mollett, B.; Peers-Dent, J. |
| EPI2232675 | NP | United Kingdom | 2022-Oct-20 | EPI_ISL_15957641 | A/chicken/England/138092/2022 | Animal and Plant Health Agency (APHA) | Animal and Plant Health Agency (APHA) | Byrne, A.; Howton, N.; Maskell, D.; Mollett, B.; Peers-Dent, J. |
| EPI2232676 | NS | United Kingdom | 2022-Oct-20 | EPI_ISL_15957641 | A/chicken/England/138092/2022 | Animal and Plant Health Agency (APHA) | Animal and Plant Health Agency (APHA) | Byrne, A.; Howton, N.; Maskell, D.; Mollett, B.; Peers-Dent, J. |
| EPI2232677 | MP | United Kingdom | 2022-Oct-20 | EPI_ISL_15957641 | A/chicken/England/138092/2022 | Animal and Plant Health Agency (APHA) | Animal and Plant Health Agency (APHA) | Byrne, A.; Howton, N.; Maskell, D.; Mollett, B.; Peers-Dent, J. |
| EPI2232683 | NP | United Kingdom | 2022-Oct-20 | EPI_ISL_15957648 | A/chicken/England/138155/2022 | Animal and Plant Health Agency (APHA) | Animal and Plant Health Agency (APHA) | Byrne, A.; Howton, N.; Maskell, D.; Mollett, B.; Peers-Dent, J. |

| | | | | | | | | |
|---|---|---|---|---|---|---|---|---|
| EPI2232684 | NS | United Kingdom | 2022-Oct-20 | EPI_ISL_15957648 | A/chicken/England/138155/2022 | Animal and Plant Health Agency (APHA) | Animal and Plant Health Agency (APHA) | Byrne, A.; Howton, N.; Maskell, D.; Mollett, B.; Peers-Dent, J. |
| EPI2232685 | MP | United Kingdom | 2022-Oct-20 | EPI_ISL_15957648 | A/chicken/England/138155/2022 | Animal and Plant Health Agency (APHA) | Animal and Plant Health Agency (APHA) | Byrne, A.; Howton, N.; Maskell, D.; Mollett, B.; Peers-Dent, J. |
| EPI2232691 | NP | United Kingdom | 2022-Oct-20 | EPI_ISL_15957654 | A/chicken/England/138249/2022 | Animal and Plant Health Agency (APHA) | Animal and Plant Health Agency (APHA) | Byrne, A.; Howton, N.; Maskell, D.; Mollett, B.; Peers-Dent, J. |
| EPI2232692 | NS | United Kingdom | 2022-Oct-20 | EPI_ISL_15957654 | A/chicken/England/138249/2022 | Animal and Plant Health Agency (APHA) | Animal and Plant Health Agency (APHA) | Byrne, A.; Howton, N.; Maskell, D.; Mollett, B.; Peers-Dent, J. |
| EPI2232693 | MP | United Kingdom | 2022-Oct-20 | EPI_ISL_15957654 | A/chicken/England/138249/2022 | Animal and Plant Health Agency (APHA) | Animal and Plant Health Agency (APHA) | Byrne, A.; Howton, N.; Maskell, D.; Mollett, B.; Peers-Dent, J. |
| EPI2232699 | NP | United Kingdom | 2022-Oct-21 | EPI_ISL_15957660 | A/chicken/Wales/138294/2022 | Animal and Plant Health Agency (APHA) | Animal and Plant Health Agency (APHA) | Byrne, A.; Howton, N.; Maskell, D.; Mollett, B.; Peers-Dent, J. |
| EPI2232700 | NS | United Kingdom | 2022-Oct-21 | EPI_ISL_15957660 | A/chicken/Wales/138294/2022 | Animal and Plant Health Agency (APHA) | Animal and Plant Health Agency (APHA) | Byrne, A.; Howton, N.; Maskell, D.; Mollett, B.; Peers-Dent, J. |
| EPI2232701 | MP | United Kingdom | 2022-Oct-21 | EPI_ISL_15957660 | A/chicken/Wales/138294/2022 | Animal and Plant Health Agency (APHA) | Animal and Plant Health Agency (APHA) | Byrne, A.; Howton, N.; Maskell, D.; Mollett, B.; Peers-Dent, J. |
| EPI2232707 | NP | United Kingdom | 2022-Oct-20 | EPI_ISL_15957666 | A/turkey/England/138429/2022 | Animal and Plant Health Agency (APHA) | Animal and Plant Health Agency (APHA) | Byrne, A.; Howton, N.; Maskell, D.; Mollett, B.; Peers-Dent, J. |
| EPI2232708 | NS | United Kingdom | 2022-Oct-20 | EPI_ISL_15957666 | A/turkey/England/138429/2022 | Animal and Plant Health Agency (APHA) | Animal and Plant Health Agency (APHA) | Byrne, A.; Howton, N.; Maskell, D.; Mollett, B.; Peers-Dent, J. |
| EPI2232709 | MP | United Kingdom | 2022-Oct-20 | EPI_ISL_15957666 | A/turkey/England/138429/2022 | Animal and Plant Health Agency (APHA) | Animal and Plant Health Agency (APHA) | Byrne, A.; Howton, N.; Maskell, D.; Mollett, B.; Peers-Dent, J. |
| EPI2232715 | NP | United Kingdom | 2022-Oct-21 | EPI_ISL_15957672 | A/chicken/England/138486/2022 | Animal and Plant Health Agency (APHA) | Animal and Plant Health Agency (APHA) | Byrne, A.; Howton, N.; Maskell, D.; Mollett, B.; Peers-Dent, J. |
| EPI2232716 | NS | United Kingdom | 2022-Oct-21 | EPI_ISL_15957672 | A/chicken/England/138486/2022 | Animal and Plant Health Agency (APHA) | Animal and Plant Health Agency (APHA) | Byrne, A.; Howton, N.; Maskell, D.; Mollett, B.; Peers-Dent, J. |
| EPI2232717 | MP | United Kingdom | 2022-Oct-21 | EPI_ISL_15957672 | A/chicken/England/138486/2022 | Animal and Plant Health Agency (APHA) | Animal and Plant Health Agency (APHA) | Byrne, A.; Howton, N.; Maskell, D.; Mollett, B.; Peers-Dent, J. |
| EPI2232724 | NS | United Kingdom | 2022-Oct-21 | EPI_ISL_15957678 | A/chicken/England/138597/2022 | Animal and Plant Health Agency (APHA) | Animal and Plant Health Agency (APHA) | Byrne, A.; Howton, N.; Maskell, D.; Mollett, B.; Peers-Dent, J. |
| EPI2232725 | MP | United Kingdom | 2022-Oct-21 | EPI_ISL_15957678 | A/chicken/England/138597/2022 | Animal and Plant Health Agency (APHA) | Animal and Plant Health Agency (APHA) | Byrne, A.; Howton, N.; Maskell, D.; Mollett, B.; Peers-Dent, J. |
| EPI2232723 | NP | United Kingdom | 2022-Oct-21 | EPI_ISL_15957678 | A/chicken/England/138597/2022 | Animal and Plant Health Agency (APHA) | Animal and Plant Health Agency (APHA) | Byrne, A.; Howton, N.; Maskell, D.; Mollett, B.; Peers-Dent, J. |
| EPI2232731 | NP | United Kingdom | 2022-Oct-22 | EPI_ISL_15957684 | A/domestic_goose/England/138821/2022 | Animal and Plant Health Agency (APHA) | Animal and Plant Health Agency (APHA) | Byrne, A.; Howton, N.; Maskell, D.; Mollett, B.; Peers-Dent, J. |
| EPI2232732 | NS | United Kingdom | 2022-Oct-22 | EPI_ISL_15957684 | A/domestic_goose/England/138821/2022 | Animal and Plant Health Agency (APHA) | Animal and Plant Health Agency (APHA) | Byrne, A.; Howton, N.; Maskell, D.; Mollett, B.; Peers-Dent, J. |
| EPI2232733 | MP | United Kingdom | 2022-Oct-22 | EPI_ISL_15957684 | A/domestic_goose/England/138821/2022 | Animal and Plant Health Agency (APHA) | Animal and Plant Health Agency (APHA) | Byrne, A.; Howton, N.; Maskell, D.; Mollett, B.; Peers-Dent, J. |
| EPI2232739 | NP | United Kingdom | 2022-Oct-22 | EPI_ISL_15957690 | A/turkey/England/140408/2022 | Animal and Plant Health Agency (APHA) | Animal and Plant Health Agency (APHA) | Byrne, A.; Howton, N.; Maskell, D.; Mollett, B.; Peers-Dent, J. |
| EPI2232740 | NS | United Kingdom | 2022-Oct-22 | EPI_ISL_15957690 | A/turkey/England/140408/2022 | Animal and Plant Health Agency (APHA) | Animal and Plant Health Agency (APHA) | Byrne, A.; Howton, N.; Maskell, D.; Mollett, B.; Peers-Dent, J. |
| EPI2232741 | MP | United Kingdom | 2022-Oct-22 | EPI_ISL_15957690 | A/turkey/England/140408/2022 | Animal and Plant Health Agency (APHA) | Animal and Plant Health Agency (APHA) | Byrne, A.; Howton, N.; Maskell, D.; Mollett, B.; Peers-Dent, J. |
| EPI2232747 | NP | United Kingdom | 2022-Oct-23 | EPI_ISL_15957696 | A/chicken/England/140572/2022 | Animal and Plant Health Agency (APHA) | Animal and Plant Health Agency (APHA) | Byrne, A.; Howton, N.; Maskell, D.; Mollett, B.; Peers-Dent, J. |
| EPI2232748 | NS | United Kingdom | 2022-Oct-23 | EPI_ISL_15957696 | A/chicken/England/140572/2022 | Animal and Plant Health Agency (APHA) | Animal and Plant Health Agency (APHA) | Byrne, A.; Howton, N.; Maskell, D.; Mollett, B.; Peers-Dent, J. |
| EPI2232749 | MP | United Kingdom | 2022-Oct-23 | EPI_ISL_15957696 | A/chicken/England/140572/2022 | Animal and Plant Health Agency (APHA) | Animal and Plant Health Agency (APHA) | Byrne, A.; Howton, N.; Maskell, D.; Mollett, B.; Peers-Dent, J. |
| EPI2232755 | NP | United Kingdom | 2022-Oct-23 | EPI_ISL_15957702 | A/turkey/England/140663/2022 | Animal and Plant Health Agency (APHA) | Animal and Plant Health Agency (APHA) | Byrne, A.; Howton, N.; Maskell, D.; Mollett, B.; Peers-Dent, J. |
| EPI2232756 | NS | United Kingdom | 2022-Oct-23 | EPI_ISL_15957702 | A/turkey/England/140663/2022 | Animal and Plant Health Agency (APHA) | Animal and Plant Health Agency (APHA) | Byrne, A.; Howton, N.; Maskell, D.; Mollett, B.; Peers-Dent, J. |
| EPI2232757 | MP | United Kingdom | 2022-Oct-23 | EPI_ISL_15957702 | A/turkey/England/140663/2022 | Animal and Plant Health Agency (APHA) | Animal and Plant Health Agency (APHA) | Byrne, A.; Howton, N.; Maskell, D.; Mollett, B.; Peers-Dent, J. |
| EPI2232763 | NP | United Kingdom | 2022-Oct-24 | EPI_ISL_15957708 | A/turkey/England/140733/2022 | Animal and Plant Health Agency (APHA) | Animal and Plant Health Agency (APHA) | Byrne, A.; Howton, N.; Maskell, D.; Mollett, B.; Peers-Dent, J. |
| EPI2232764 | NS | United Kingdom | 2022-Oct-24 | EPI_ISL_15957708 | A/turkey/England/140733/2022 | Animal and Plant Health Agency (APHA) | Animal and Plant Health Agency (APHA) | Byrne, A.; Howton, N.; Maskell, D.; Mollett, B.; Peers-Dent, J. |
| EPI2232765 | MP | United Kingdom | 2022-Oct-24 | EPI_ISL_15957708 | A/turkey/England/140733/2022 | Animal and Plant Health Agency (APHA) | Animal and Plant Health Agency (APHA) | Byrne, A.; Howton, N.; Maskell, D.; Mollett, B.; Peers-Dent, J. |
| EPI2232779 | NP | United Kingdom | 2022-Oct-26 | EPI_ISL_15957722 | A/domestic_duck/England/141358/2022 | Animal and Plant Health Agency (APHA) | Animal and Plant Health Agency (APHA) | Byrne, A.; Howton, N.; Maskell, D.; Mollett, B.; Peers-Dent, J. |
| EPI2232780 | NS | United Kingdom | 2022-Oct-26 | EPI_ISL_15957722 | A/domestic_duck/England/141358/2022 | Animal and Plant Health Agency (APHA) | Animal and Plant Health Agency (APHA) | Byrne, A.; Howton, N.; Maskell, D.; Mollett, B.; Peers-Dent, J. |
| EPI2232781 | MP | United Kingdom | 2022-Oct-26 | EPI_ISL_15957722 | A/domestic_duck/England/141358/2022 | Animal and Plant Health Agency (APHA) | Animal and Plant Health Agency (APHA) | Byrne, A.; Howton, N.; Maskell, D.; Mollett, B.; Peers-Dent, J. |

| | | | | | | | | |
|---|---|---|---|---|---|---|---|---|
| EPI2232787 | NP | United Kingdom | 2022-Oct-28 | EPI_ISL_15957728 | A/Chicken/England/141539/2022 | Animal and Plant Health Agency (APHA) | Animal and Plant Health Agency (APHA) | Byrne, A.; Howton, N.; Maskell, D.; Mollett, B.; Peers-Dent, J. |
| EPI2232788 | NS | United Kingdom | 2022-Oct-28 | EPI_ISL_15957728 | A/Chicken/England/141539/2022 | Animal and Plant Health Agency (APHA) | Animal and Plant Health Agency (APHA) | Byrne, A.; Howton, N.; Maskell, D.; Mollett, B.; Peers-Dent, J. |
| EPI2232789 | MP | United Kingdom | 2022-Oct-28 | EPI_ISL_15957728 | A/Chicken/England/141539/2022 | Animal and Plant Health Agency (APHA) | Animal and Plant Health Agency (APHA) | Byrne, A.; Howton, N.; Maskell, D.; Mollett, B.; Peers-Dent, J. |
| EPI2232795 | NP | United Kingdom | 2022-Oct-29 | EPI_ISL_15957730 | A/Turkey/England/141755/2022 | Animal and Plant Health Agency (APHA) | Animal and Plant Health Agency (APHA) | Byrne, A.; Howton, N.; Maskell, D.; Mollett, B.; Peers-Dent, J. |
| EPI2232796 | NS | United Kingdom | 2022-Oct-29 | EPI_ISL_15957730 | A/Turkey/England/141755/2022 | Animal and Plant Health Agency (APHA) | Animal and Plant Health Agency (APHA) | Byrne, A.; Howton, N.; Maskell, D.; Mollett, B.; Peers-Dent, J. |
| EPI2232797 | MP | United Kingdom | 2022-Oct-29 | EPI_ISL_15957730 | A/Turkey/England/141755/2022 | Animal and Plant Health Agency (APHA) | Animal and Plant Health Agency (APHA) | Byrne, A.; Howton, N.; Maskell, D.; Mollett, B.; Peers-Dent, J. |
| EPI2232803 | NP | United Kingdom | 2022-Oct-29 | EPI_ISL_15957731 | A/Turkey/England/141822/2022 | Animal and Plant Health Agency (APHA) | Animal and Plant Health Agency (APHA) | Byrne, A.; Howton, N.; Maskell, D.; Mollett, B.; Peers-Dent, J. |
| EPI2232804 | NS | United Kingdom | 2022-Oct-29 | EPI_ISL_15957731 | A/Turkey/England/141822/2022 | Animal and Plant Health Agency (APHA) | Animal and Plant Health Agency (APHA) | Byrne, A.; Howton, N.; Maskell, D.; Mollett, B.; Peers-Dent, J. |
| EPI2232805 | MP | United Kingdom | 2022-Oct-29 | EPI_ISL_15957731 | A/Turkey/England/141822/2022 | Animal and Plant Health Agency (APHA) | Animal and Plant Health Agency (APHA) | Byrne, A.; Howton, N.; Maskell, D.; Mollett, B.; Peers-Dent, J. |
| EPI2232811 | NP | United Kingdom | 2022-Oct-30 | EPI_ISL_15957744 | A/Chicken/Scotland/141941/2022 | Animal and Plant Health Agency (APHA) | Animal and Plant Health Agency (APHA) | Byrne, A.; Howton, N.; Maskell, D.; Mollett, B.; Peers-Dent, J. |
| EPI2232812 | NS | United Kingdom | 2022-Oct-30 | EPI_ISL_15957744 | A/Chicken/Scotland/141941/2022 | Animal and Plant Health Agency (APHA) | Animal and Plant Health Agency (APHA) | Byrne, A.; Howton, N.; Maskell, D.; Mollett, B.; Peers-Dent, J. |
| EPI2232813 | MP | United Kingdom | 2022-Oct-30 | EPI_ISL_15957744 | A/Chicken/Scotland/141941/2022 | Animal and Plant Health Agency (APHA) | Animal and Plant Health Agency (APHA) | Byrne, A.; Howton, N.; Maskell, D.; Mollett, B.; Peers-Dent, J. |
| EPI2232819 | NP | United Kingdom | 2022-Oct-29 | EPI_ISL_15957766 | A/Domestic_Duck/England/141951/2022 | Animal and Plant Health Agency (APHA) | Animal and Plant Health Agency (APHA) | Byrne, A.; Howton, N.; Maskell, D.; Mollett, B.; Peers-Dent, J. |
| EPI2232820 | NS | United Kingdom | 2022-Oct-29 | EPI_ISL_15957766 | A/Domestic_Duck/England/141951/2022 | Animal and Plant Health Agency (APHA) | Animal and Plant Health Agency (APHA) | Byrne, A.; Howton, N.; Maskell, D.; Mollett, B.; Peers-Dent, J. |
| EPI2232821 | MP | United Kingdom | 2022-Oct-29 | EPI_ISL_15957766 | A/Domestic_Duck/England/141951/2022 | Animal and Plant Health Agency (APHA) | Animal and Plant Health Agency (APHA) | Byrne, A.; Howton, N.; Maskell, D.; Mollett, B.; Peers-Dent, J. |
| EPI2232827 | NP | United Kingdom | 2022-Oct-29 | EPI_ISL_15957774 | A/Chicken/England/141990/2022 | Animal and Plant Health Agency (APHA) | Animal and Plant Health Agency (APHA) | Byrne, A.; Howton, N.; Maskell, D.; Mollett, B.; Peers-Dent, J. |
| EPI2232828 | NS | United Kingdom | 2022-Oct-29 | EPI_ISL_15957774 | A/Chicken/England/141990/2022 | Animal and Plant Health Agency (APHA) | Animal and Plant Health Agency (APHA) | Byrne, A.; Howton, N.; Maskell, D.; Mollett, B.; Peers-Dent, J. |
| EPI2232829 | MP | United Kingdom | 2022-Oct-29 | EPI_ISL_15957774 | A/Chicken/England/141990/2022 | Animal and Plant Health Agency (APHA) | Animal and Plant Health Agency (APHA) | Byrne, A.; Howton, N.; Maskell, D.; Mollett, B.; Peers-Dent, J. |
| EPI2232835 | NP | United Kingdom | 2022-Oct-29 | EPI_ISL_15957779 | A/Chicken/Scotland/142099/2022 | Animal and Plant Health Agency (APHA) | Animal and Plant Health Agency (APHA) | Byrne, A.; Howton, N.; Maskell, D.; Mollett, B.; Peers-Dent, J. |
| EPI2232836 | NS | United Kingdom | 2022-Oct-29 | EPI_ISL_15957779 | A/Chicken/Scotland/142099/2022 | Animal and Plant Health Agency (APHA) | Animal and Plant Health Agency (APHA) | Byrne, A.; Howton, N.; Maskell, D.; Mollett, B.; Peers-Dent, J. |
| EPI2232837 | MP | United Kingdom | 2022-Oct-29 | EPI_ISL_15957779 | A/Chicken/Scotland/142099/2022 | Animal and Plant Health Agency (APHA) | Animal and Plant Health Agency (APHA) | Byrne, A.; Howton, N.; Maskell, D.; Mollett, B.; Peers-Dent, J. |
| EPI2232843 | NP | United Kingdom | 2022-Oct-31 | EPI_ISL_15957785 | A/Chicken/England/142196/2022 | Animal and Plant Health Agency (APHA) | Animal and Plant Health Agency (APHA) | Byrne, A.; Howton, N.; Maskell, D.; Mollett, B.; Peers-Dent, J. |
| EPI2232844 | NS | United Kingdom | 2022-Oct-31 | EPI_ISL_15957785 | A/Chicken/England/142196/2022 | Animal and Plant Health Agency (APHA) | Animal and Plant Health Agency (APHA) | Byrne, A.; Howton, N.; Maskell, D.; Mollett, B.; Peers-Dent, J. |
| EPI2232845 | MP | United Kingdom | 2022-Oct-31 | EPI_ISL_15957785 | A/Chicken/England/142196/2022 | Animal and Plant Health Agency (APHA) | Animal and Plant Health Agency (APHA) | Byrne, A.; Howton, N.; Maskell, D.; Mollett, B.; Peers-Dent, J. |
| EPI2232851 | NP | United Kingdom | 2022-Oct-31 | EPI_ISL_15957792 | A/Chicken/England/142361/2022 | Animal and Plant Health Agency (APHA) | Animal and Plant Health Agency (APHA) | Byrne, A.; Howton, N.; Maskell, D.; Mollett, B.; Peers-Dent, J. |
| EPI2232852 | NS | United Kingdom | 2022-Oct-31 | EPI_ISL_15957792 | A/Chicken/England/142361/2022 | Animal and Plant Health Agency (APHA) | Animal and Plant Health Agency (APHA) | Byrne, A.; Howton, N.; Maskell, D.; Mollett, B.; Peers-Dent, J. |
| EPI2232853 | MP | United Kingdom | 2022-Oct-31 | EPI_ISL_15957792 | A/Chicken/England/142361/2022 | Animal and Plant Health Agency (APHA) | Animal and Plant Health Agency (APHA) | Byrne, A.; Howton, N.; Maskell, D.; Mollett, B.; Peers-Dent, J. |
| EPI2232859 | NP | United Kingdom | 2022-Nov-01 | EPI_ISL_15957798 | A/Turkey/England/143029/2022 | Animal and Plant Health Agency (APHA) | Animal and Plant Health Agency (APHA) | Byrne, A.; Howton, N.; Maskell, D.; Mollett, B.; Peers-Dent, J. |
| EPI2232860 | NS | United Kingdom | 2022-Nov-01 | EPI_ISL_15957798 | A/Turkey/England/143029/2022 | Animal and Plant Health Agency (APHA) | Animal and Plant Health Agency (APHA) | Byrne, A.; Howton, N.; Maskell, D.; Mollett, B.; Peers-Dent, J. |
| EPI2232861 | MP | United Kingdom | 2022-Nov-01 | EPI_ISL_15957798 | A/Turkey/England/143029/2022 | Animal and Plant Health Agency (APHA) | Animal and Plant Health Agency (APHA) | Byrne, A.; Howton, N.; Maskell, D.; Mollett, B.; Peers-Dent, J. |
| EPI2232867 | NP | United Kingdom | 2022-Nov-01 | EPI_ISL_15957804 | A/Turkey/England/143099/2022 | Animal and Plant Health Agency (APHA) | Animal and Plant Health Agency (APHA) | Byrne, A.; Howton, N.; Maskell, D.; Mollett, B.; Peers-Dent, J. |
| EPI2232868 | NS | United Kingdom | 2022-Nov-01 | EPI_ISL_15957804 | A/Turkey/England/143099/2022 | Animal and Plant Health Agency (APHA) | Animal and Plant Health Agency (APHA) | Byrne, A.; Howton, N.; Maskell, D.; Mollett, B.; Peers-Dent, J. |
| EPI2232869 | MP | United Kingdom | 2022-Nov-01 | EPI_ISL_15957804 | A/Turkey/England/143099/2022 | Animal and Plant Health Agency (APHA) | Animal and Plant Health Agency (APHA) | Byrne, A.; Howton, N.; Maskell, D.; Mollett, B.; Peers-Dent, J. |
| EPI2232877 | MP | United Kingdom | 2022-Nov-01 | EPI_ISL_15957810 | A/Turkey/England/143169/2022 | Animal and Plant Health Agency (APHA) | Animal and Plant Health Agency (APHA) | Byrne, A.; Howton, N.; Maskell, D.; Mollett, B.; Peers-Dent, J. |
| EPI2232875 | NP | United Kingdom | 2022-Nov-01 | EPI_ISL_15957810 | A/Turkey/England/143169/2022 | Animal and Plant Health Agency (APHA) | Animal and Plant Health Agency (APHA) | Byrne, A.; Howton, N.; Maskell, D.; Mollett, B.; Peers-Dent, J. |

| | | | | | | | | |
|---|---|---|---|---|---|---|---|---|
| EPI2232876 | NS | United Kingdom | 2022-Nov-01 | EPI_ISL_15957810 | A/Turkey/England/143169/2022 | Animal and Plant Health Agency (APHA) | Animal and Plant Health Agency (APHA) | Byrne, A.; Howton, N.; Maskell, D.; Mollett, B.; Peers-Dent, J. |
| EPI2232883 | NP | United Kingdom | 2022-Nov-02 | EPI_ISL_15957817 | A/Turkey/England/143570/2022 | Animal and Plant Health Agency (APHA) | Animal and Plant Health Agency (APHA) | Byrne, A.; Howton, N.; Maskell, D.; Mollett, B.; Peers-Dent, J. |
| EPI2232884 | NS | United Kingdom | 2022-Nov-02 | EPI_ISL_15957817 | A/Turkey/England/143570/2022 | Animal and Plant Health Agency (APHA) | Animal and Plant Health Agency (APHA) | Byrne, A.; Howton, N.; Maskell, D.; Mollett, B.; Peers-Dent, J. |
| EPI2232885 | MP | United Kingdom | 2022-Nov-02 | EPI_ISL_15957817 | A/Turkey/England/143570/2022 | Animal and Plant Health Agency (APHA) | Animal and Plant Health Agency (APHA) | Byrne, A.; Howton, N.; Maskell, D.; Mollett, B.; Peers-Dent, J. |
| EPI2232891 | NP | United Kingdom | 2022-Nov-02 | EPI_ISL_15957824 | A/Chicken/England/143642/2022 | Animal and Plant Health Agency (APHA) | Animal and Plant Health Agency (APHA) | Byrne, A.; Howton, N.; Maskell, D.; Mollett, B.; Peers-Dent, J. |
| EPI2232892 | NS | United Kingdom | 2022-Nov-02 | EPI_ISL_15957824 | A/Chicken/England/143642/2022 | Animal and Plant Health Agency (APHA) | Animal and Plant Health Agency (APHA) | Byrne, A.; Howton, N.; Maskell, D.; Mollett, B.; Peers-Dent, J. |
| EPI2232893 | MP | United Kingdom | 2022-Nov-02 | EPI_ISL_15957824 | A/Chicken/England/143642/2022 | Animal and Plant Health Agency (APHA) | Animal and Plant Health Agency (APHA) | Byrne, A.; Howton, N.; Maskell, D.; Mollett, B.; Peers-Dent, J. |
| EPI2232899 | NP | United Kingdom | 2022-Nov-02 | EPI_ISL_15957830 | A/Turkey/England/143717/2022 | Animal and Plant Health Agency (APHA) | Animal and Plant Health Agency (APHA) | Byrne, A.; Howton, N.; Maskell, D.; Mollett, B.; Peers-Dent, J. |
| EPI2232900 | NS | United Kingdom | 2022-Nov-02 | EPI_ISL_15957830 | A/Turkey/England/143717/2022 | Animal and Plant Health Agency (APHA) | Animal and Plant Health Agency (APHA) | Byrne, A.; Howton, N.; Maskell, D.; Mollett, B.; Peers-Dent, J. |
| EPI2232901 | MP | United Kingdom | 2022-Nov-02 | EPI_ISL_15957830 | A/Turkey/England/143717/2022 | Animal and Plant Health Agency (APHA) | Animal and Plant Health Agency (APHA) | Byrne, A.; Howton, N.; Maskell, D.; Mollett, B.; Peers-Dent, J. |
| EPI2232907 | NP | United Kingdom | 2022-Nov-02 | EPI_ISL_15957837 | A/Turkey/England/144040/2022 | Animal and Plant Health Agency (APHA) | Animal and Plant Health Agency (APHA) | Byrne, A.; Howton, N.; Maskell, D.; Mollett, B.; Peers-Dent, J. |
| EPI2232908 | NS | United Kingdom | 2022-Nov-02 | EPI_ISL_15957837 | A/Turkey/England/144040/2022 | Animal and Plant Health Agency (APHA) | Animal and Plant Health Agency (APHA) | Byrne, A.; Howton, N.; Maskell, D.; Mollett, B.; Peers-Dent, J. |
| EPI2232909 | MP | United Kingdom | 2022-Nov-02 | EPI_ISL_15957837 | A/Turkey/England/144040/2022 | Animal and Plant Health Agency (APHA) | Animal and Plant Health Agency (APHA) | Byrne, A.; Howton, N.; Maskell, D.; Mollett, B.; Peers-Dent, J. |
| EPI2232915 | NP | United Kingdom | 2022-Nov-03 | EPI_ISL_15957844 | A/Domestic_Goose/England/145097/2022 | Animal and Plant Health Agency (APHA) | Animal and Plant Health Agency (APHA) | Byrne, A.; Howton, N.; Maskell, D.; Mollett, B.; Peers-Dent, J. |
| EPI2232916 | NS | United Kingdom | 2022-Nov-03 | EPI_ISL_15957844 | A/Domestic_Goose/England/145097/2022 | Animal and Plant Health Agency (APHA) | Animal and Plant Health Agency (APHA) | Byrne, A.; Howton, N.; Maskell, D.; Mollett, B.; Peers-Dent, J. |
| EPI2232917 | MP | United Kingdom | 2022-Nov-03 | EPI_ISL_15957844 | A/Domestic_Goose/England/145097/2022 | Animal and Plant Health Agency (APHA) | Animal and Plant Health Agency (APHA) | Byrne, A.; Howton, N.; Maskell, D.; Mollett, B.; Peers-Dent, J. |
| EPI2232923 | NP | United Kingdom | 2022-Nov-02 | EPI_ISL_15957850 | A/chicken/Scotland/145272/2022 | Animal and Plant Health Agency (APHA) | Animal and Plant Health Agency (APHA) | Byrne, A.; Howton, N.; Maskell, D.; Mollett, B.; Peers-Dent, J. |
| EPI2232924 | NS | United Kingdom | 2022-Nov-02 | EPI_ISL_15957850 | A/chicken/Scotland/145272/2022 | Animal and Plant Health Agency (APHA) | Animal and Plant Health Agency (APHA) | Byrne, A.; Howton, N.; Maskell, D.; Mollett, B.; Peers-Dent, J. |
| EPI2232925 | MP | United Kingdom | 2022-Nov-02 | EPI_ISL_15957850 | A/chicken/Scotland/145272/2022 | Animal and Plant Health Agency (APHA) | Animal and Plant Health Agency (APHA) | Byrne, A.; Howton, N.; Maskell, D.; Mollett, B.; Peers-Dent, J. |
| EPI2232931 | NP | United Kingdom | 2022-Nov-03 | EPI_ISL_15957857 | A/turkey/England/145357/2022 | Animal and Plant Health Agency (APHA) | Animal and Plant Health Agency (APHA) | Byrne, A.; Howton, N.; Maskell, D.; Mollett, B.; Peers-Dent, J. |
| EPI2232932 | NS | United Kingdom | 2022-Nov-03 | EPI_ISL_15957857 | A/turkey/England/145357/2022 | Animal and Plant Health Agency (APHA) | Animal and Plant Health Agency (APHA) | Byrne, A.; Howton, N.; Maskell, D.; Mollett, B.; Peers-Dent, J. |
| EPI2232933 | MP | United Kingdom | 2022-Nov-03 | EPI_ISL_15957857 | A/turkey/England/145357/2022 | Animal and Plant Health Agency (APHA) | Animal and Plant Health Agency (APHA) | Byrne, A.; Howton, N.; Maskell, D.; Mollett, B.; Peers-Dent, J. |
| EPI2232939 | NP | United Kingdom | 2022-Nov-03 | EPI_ISL_15957865 | A/turkey/England/145657/2022 | Animal and Plant Health Agency (APHA) | Animal and Plant Health Agency (APHA) | Byrne, A.; Howton, N.; Maskell, D.; Mollett, B.; Peers-Dent, J. |
| EPI2232940 | NS | United Kingdom | 2022-Nov-03 | EPI_ISL_15957865 | A/turkey/England/145657/2022 | Animal and Plant Health Agency (APHA) | Animal and Plant Health Agency (APHA) | Byrne, A.; Howton, N.; Maskell, D.; Mollett, B.; Peers-Dent, J. |
| EPI2232941 | MP | United Kingdom | 2022-Nov-03 | EPI_ISL_15957865 | A/turkey/England/145657/2022 | Animal and Plant Health Agency (APHA) | Animal and Plant Health Agency (APHA) | Byrne, A.; Howton, N.; Maskell, D.; Mollett, B.; Peers-Dent, J. |
| EPI2232947 | NP | United Kingdom | 2022-Nov-04 | EPI_ISL_15957872 | A/chicken/Scotland/146221/2022 | Animal and Plant Health Agency (APHA) | Animal and Plant Health Agency (APHA) | Byrne, A.; Howton, N.; Maskell, D.; Mollett, B.; Peers-Dent, J. |
| EPI2232948 | NS | United Kingdom | 2022-Nov-04 | EPI_ISL_15957872 | A/chicken/Scotland/146221/2022 | Animal and Plant Health Agency (APHA) | Animal and Plant Health Agency (APHA) | Byrne, A.; Howton, N.; Maskell, D.; Mollett, B.; Peers-Dent, J. |
| EPI2232949 | MP | United Kingdom | 2022-Nov-04 | EPI_ISL_15957872 | A/chicken/Scotland/146221/2022 | Animal and Plant Health Agency (APHA) | Animal and Plant Health Agency (APHA) | Byrne, A.; Howton, N.; Maskell, D.; Mollett, B.; Peers-Dent, J. |
| EPI2232955 | NP | United Kingdom | 2022-Nov-05 | EPI_ISL_15957878 | A/domestic_duck/England/146319/2022 | Animal and Plant Health Agency (APHA) | Animal and Plant Health Agency (APHA) | Byrne, A.; Howton, N.; Maskell, D.; Mollett, B.; Peers-Dent, J. |
| EPI2232956 | NS | United Kingdom | 2022-Nov-05 | EPI_ISL_15957878 | A/domestic_duck/England/146319/2022 | Animal and Plant Health Agency (APHA) | Animal and Plant Health Agency (APHA) | Byrne, A.; Howton, N.; Maskell, D.; Mollett, B.; Peers-Dent, J. |
| EPI2232957 | MP | United Kingdom | 2022-Nov-05 | EPI_ISL_15957878 | A/domestic_duck/England/146319/2022 | Animal and Plant Health Agency (APHA) | Animal and Plant Health Agency (APHA) | Byrne, A.; Howton, N.; Maskell, D.; Mollett, B.; Peers-Dent, J. |
| EPI2232963 | NP | United Kingdom | 2022-Nov-05 | EPI_ISL_15957885 | A/domestic_goose/England/146384/2022 | Animal and Plant Health Agency (APHA) | Animal and Plant Health Agency (APHA) | Byrne, A.; Howton, N.; Maskell, D.; Mollett, B.; Peers-Dent, J. |
| EPI2232964 | NS | United Kingdom | 2022-Nov-05 | EPI_ISL_15957885 | A/domestic_goose/England/146384/2022 | Animal and Plant Health Agency (APHA) | Animal and Plant Health Agency (APHA) | Byrne, A.; Howton, N.; Maskell, D.; Mollett, B.; Peers-Dent, J. |
| EPI2232965 | MP | United Kingdom | 2022-Nov-05 | EPI_ISL_15957885 | A/domestic_goose/England/146384/2022 | Animal and Plant Health Agency (APHA) | Animal and Plant Health Agency (APHA) | Byrne, A.; Howton, N.; Maskell, D.; Mollett, B.; Peers-Dent, J. |
| EPI2232980 | NS | United Kingdom | 2022-Nov-06 | EPI_ISL_15957899 | A/turkey/England/146512/2022 | Animal and Plant Health Agency (APHA) | Animal and Plant Health Agency (APHA) | Byrne, A.; Howton, N.; Maskell, D.; Mollett, B.; Peers-Dent, J. |

| | | | | | | | | |
|---|---|---|---|---|---|---|---|---|
| EPI2232981 | MP | United Kingdom | 2022-Nov-06 | EPI_ISL_15957899 | A/turkey/England/146512/2022 | Animal and Plant Health Agency (APHA) | Animal and Plant Health Agency (APHA) | Byrne, A.; Howton, N.; Maskell, D.; Mollett, B.; Peers-Dent, J. |
| EPI2232979 | NP | United Kingdom | 2022-Nov-06 | EPI_ISL_15957899 | A/turkey/England/146512/2022 | Animal and Plant Health Agency (APHA) | Animal and Plant Health Agency (APHA) | Byrne, A.; Howton, N.; Maskell, D.; Mollett, B.; Peers-Dent, J. |
| EPI2232987 | NP | United Kingdom | 2022-Nov-06 | EPI_ISL_15957906 | A/domestic_goose/Wales/146569/2022 | Animal and Plant Health Agency (APHA) | Animal and Plant Health Agency (APHA) | Byrne, A.; Howton, N.; Maskell, D.; Mollett, B.; Peers-Dent, J. |
| EPI2232988 | NS | United Kingdom | 2022-Nov-06 | EPI_ISL_15957906 | A/domestic_goose/Wales/146569/2022 | Animal and Plant Health Agency (APHA) | Animal and Plant Health Agency (APHA) | Byrne, A.; Howton, N.; Maskell, D.; Mollett, B.; Peers-Dent, J. |
| EPI2232989 | MP | United Kingdom | 2022-Nov-06 | EPI_ISL_15957906 | A/domestic_goose/Wales/146569/2022 | Animal and Plant Health Agency (APHA) | Animal and Plant Health Agency (APHA) | Byrne, A.; Howton, N.; Maskell, D.; Mollett, B.; Peers-Dent, J. |
| EPI2232995 | NP | United Kingdom | 2022-Nov-06 | EPI_ISL_15957912 | A/guinea_fowl/Wales/146571/2022 | Animal and Plant Health Agency (APHA) | Animal and Plant Health Agency (APHA) | Byrne, A.; Howton, N.; Maskell, D.; Mollett, B.; Peers-Dent, J. |
| EPI2232996 | NS | United Kingdom | 2022-Nov-06 | EPI_ISL_15957912 | A/guinea_fowl/Wales/146571/2022 | Animal and Plant Health Agency (APHA) | Animal and Plant Health Agency (APHA) | Byrne, A.; Howton, N.; Maskell, D.; Mollett, B.; Peers-Dent, J. |
| EPI2232997 | MP | United Kingdom | 2022-Nov-06 | EPI_ISL_15957912 | A/guinea_fowl/Wales/146571/2022 | Animal and Plant Health Agency (APHA) | Animal and Plant Health Agency (APHA) | Byrne, A.; Howton, N.; Maskell, D.; Mollett, B.; Peers-Dent, J. |
| EPI2233003 | NP | United Kingdom | 2022-Nov-07 | EPI_ISL_15957919 | A/turkey/England/147354/2022 | Animal and Plant Health Agency (APHA) | Animal and Plant Health Agency (APHA) | Byrne, A.; Howton, N.; Maskell, D.; Mollett, B.; Peers-Dent, J. |
| EPI2233004 | NS | United Kingdom | 2022-Nov-07 | EPI_ISL_15957919 | A/turkey/England/147354/2022 | Animal and Plant Health Agency (APHA) | Animal and Plant Health Agency (APHA) | Byrne, A.; Howton, N.; Maskell, D.; Mollett, B.; Peers-Dent, J. |
| EPI2233005 | MP | United Kingdom | 2022-Nov-07 | EPI_ISL_15957919 | A/turkey/England/147354/2022 | Animal and Plant Health Agency (APHA) | Animal and Plant Health Agency (APHA) | Byrne, A.; Howton, N.; Maskell, D.; Mollett, B.; Peers-Dent, J. |
| EPI2233011 | NP | United Kingdom | 2022-Nov-08 | EPI_ISL_15957925 | A/turkey/England/147806/2022 | Animal and Plant Health Agency (APHA) | Animal and Plant Health Agency (APHA) | Byrne, A.; Howton, N.; Maskell, D.; Mollett, B.; Peers-Dent, J. |
| EPI2233012 | NS | United Kingdom | 2022-Nov-08 | EPI_ISL_15957925 | A/turkey/England/147806/2022 | Animal and Plant Health Agency (APHA) | Animal and Plant Health Agency (APHA) | Byrne, A.; Howton, N.; Maskell, D.; Mollett, B.; Peers-Dent, J. |
| EPI2233013 | MP | United Kingdom | 2022-Nov-08 | EPI_ISL_15957925 | A/turkey/England/147806/2022 | Animal and Plant Health Agency (APHA) | Animal and Plant Health Agency (APHA) | Byrne, A.; Howton, N.; Maskell, D.; Mollett, B.; Peers-Dent, J. |
| EPI2233019 | NP | United Kingdom | 2022-Nov-08 | EPI_ISL_15957933 | A/turkey/England/147878/2022 | Animal and Plant Health Agency (APHA) | Animal and Plant Health Agency (APHA) | Byrne, A.; Howton, N.; Maskell, D.; Mollett, B.; Peers-Dent, J. |
| EPI2233020 | NS | United Kingdom | 2022-Nov-08 | EPI_ISL_15957933 | A/turkey/England/147878/2022 | Animal and Plant Health Agency (APHA) | Animal and Plant Health Agency (APHA) | Byrne, A.; Howton, N.; Maskell, D.; Mollett, B.; Peers-Dent, J. |
| EPI2233021 | MP | United Kingdom | 2022-Nov-08 | EPI_ISL_15957933 | A/turkey/England/147878/2022 | Animal and Plant Health Agency (APHA) | Animal and Plant Health Agency (APHA) | Byrne, A.; Howton, N.; Maskell, D.; Mollett, B.; Peers-Dent, J. |
| EPI2233035 | NP | United Kingdom | 2022-Nov-09 | EPI_ISL_15957946 | A/chicken/England/149505/2022 | Animal and Plant Health Agency (APHA) | Animal and Plant Health Agency (APHA) | Byrne, A.; Howton, N.; Maskell, D.; Mollett, B.; Peers-Dent, J. |
| EPI2233036 | NS | United Kingdom | 2022-Nov-09 | EPI_ISL_15957946 | A/chicken/England/149505/2022 | Animal and Plant Health Agency (APHA) | Animal and Plant Health Agency (APHA) | Byrne, A.; Howton, N.; Maskell, D.; Mollett, B.; Peers-Dent, J. |
| EPI2233037 | MP | United Kingdom | 2022-Nov-09 | EPI_ISL_15957946 | A/chicken/England/149505/2022 | Animal and Plant Health Agency (APHA) | Animal and Plant Health Agency (APHA) | Byrne, A.; Howton, N.; Maskell, D.; Mollett, B.; Peers-Dent, J. |
| EPI2233043 | NP | United Kingdom | 2022-Nov-09 | EPI_ISL_15957953 | A/chicken/England/149805/2022 | Animal and Plant Health Agency (APHA) | Animal and Plant Health Agency (APHA) | Byrne, A.; Howton, N.; Maskell, D.; Mollett, B.; Peers-Dent, J. |
| EPI2233044 | NS | United Kingdom | 2022-Nov-09 | EPI_ISL_15957953 | A/chicken/England/149805/2022 | Animal and Plant Health Agency (APHA) | Animal and Plant Health Agency (APHA) | Byrne, A.; Howton, N.; Maskell, D.; Mollett, B.; Peers-Dent, J. |
| EPI2233045 | MP | United Kingdom | 2022-Nov-09 | EPI_ISL_15957953 | A/chicken/England/149805/2022 | Animal and Plant Health Agency (APHA) | Animal and Plant Health Agency (APHA) | Byrne, A.; Howton, N.; Maskell, D.; Mollett, B.; Peers-Dent, J. |
| EPI2233051 | NP | United Kingdom | 2022-Nov-11 | EPI_ISL_15957960 | A/chicken/England/150281/2022 | Animal and Plant Health Agency (APHA) | Animal and Plant Health Agency (APHA) | Byrne, A.; Howton, N.; Maskell, D.; Mollett, B.; Peers-Dent, J. |
| EPI2233052 | NS | United Kingdom | 2022-Nov-11 | EPI_ISL_15957960 | A/chicken/England/150281/2022 | Animal and Plant Health Agency (APHA) | Animal and Plant Health Agency (APHA) | Byrne, A.; Howton, N.; Maskell, D.; Mollett, B.; Peers-Dent, J. |
| EPI2233053 | MP | United Kingdom | 2022-Nov-11 | EPI_ISL_15957960 | A/chicken/England/150281/2022 | Animal and Plant Health Agency (APHA) | Animal and Plant Health Agency (APHA) | Byrne, A.; Howton, N.; Maskell, D.; Mollett, B.; Peers-Dent, J. |
| EPI2233059 | NP | United Kingdom | 2022-Nov-13 | EPI_ISL_15957966 | A/chicken/England/150351/2022 | Animal and Plant Health Agency (APHA) | Animal and Plant Health Agency (APHA) | Byrne, A.; Howton, N.; Maskell, D.; Mollett, B.; Peers-Dent, J. |
| EPI2233060 | NS | United Kingdom | 2022-Nov-13 | EPI_ISL_15957966 | A/chicken/England/150351/2022 | Animal and Plant Health Agency (APHA) | Animal and Plant Health Agency (APHA) | Byrne, A.; Howton, N.; Maskell, D.; Mollett, B.; Peers-Dent, J. |
| EPI2233061 | MP | United Kingdom | 2022-Nov-13 | EPI_ISL_15957966 | A/chicken/England/150351/2022 | Animal and Plant Health Agency (APHA) | Animal and Plant Health Agency (APHA) | Byrne, A.; Howton, N.; Maskell, D.; Mollett, B.; Peers-Dent, J. |
| EPI2233067 | NP | United Kingdom | 2022-Nov-13 | EPI_ISL_15957972 | A/turkey/England/150421/2022 | Animal and Plant Health Agency (APHA) | Animal and Plant Health Agency (APHA) | Byrne, A.; Howton, N.; Maskell, D.; Mollett, B.; Peers-Dent, J. |
| EPI2233068 | NS | United Kingdom | 2022-Nov-13 | EPI_ISL_15957972 | A/turkey/England/150421/2022 | Animal and Plant Health Agency (APHA) | Animal and Plant Health Agency (APHA) | Byrne, A.; Howton, N.; Maskell, D.; Mollett, B.; Peers-Dent, J. |
| EPI2233069 | MP | United Kingdom | 2022-Nov-13 | EPI_ISL_15957972 | A/turkey/England/150421/2022 | Animal and Plant Health Agency (APHA) | Animal and Plant Health Agency (APHA) | Byrne, A.; Howton, N.; Maskell, D.; Mollett, B.; Peers-Dent, J. |
| EPI2233075 | NP | United Kingdom | 2022-Nov-13 | EPI_ISL_15957978 | A/chicken/England/151023/2022 | Animal and Plant Health Agency (APHA) | Animal and Plant Health Agency (APHA) | Byrne, A.; Howton, N.; Maskell, D.; Mollett, B.; Peers-Dent, J. |
| EPI2233076 | NS | United Kingdom | 2022-Nov-13 | EPI_ISL_15957978 | A/chicken/England/151023/2022 | Animal and Plant Health Agency (APHA) | Animal and Plant Health Agency (APHA) | Byrne, A.; Howton, N.; Maskell, D.; Mollett, B.; Peers-Dent, J. |
| EPI2233077 | MP | United Kingdom | 2022-Nov-13 | EPI_ISL_15957978 | A/chicken/England/151023/2022 | Animal and Plant Health Agency (APHA) | Animal and Plant Health Agency (APHA) | Byrne, A.; Howton, N.; Maskell, D.; Mollett, B.; Peers-Dent, J. |

| | | | | | | | | |
|---|---|---|---|---|---|---|---|---|
| EPI2233099 | NP | United Kingdom | 2022-Nov-15 | EPI_ISL_15957998 | A/turkey/England/152353/2022 | Animal and Plant Health Agency (APHA) | Animal and Plant Health Agency (APHA) | Byrne, A.; Howton, N.; Maskell, D.; Mollett, B.; Peers-Dent, J. |
| EPI2233100 | NS | United Kingdom | 2022-Nov-15 | EPI_ISL_15957998 | A/turkey/England/152353/2022 | Animal and Plant Health Agency (APHA) | Animal and Plant Health Agency (APHA) | Byrne, A.; Howton, N.; Maskell, D.; Mollett, B.; Peers-Dent, J. |
| EPI2233101 | MP | United Kingdom | 2022-Nov-15 | EPI_ISL_15957998 | A/turkey/England/152353/2022 | Animal and Plant Health Agency (APHA) | Animal and Plant Health Agency (APHA) | Byrne, A.; Howton, N.; Maskell, D.; Mollett, B.; Peers-Dent, J. |
| EPI2233107 | NP | United Kingdom | 2022-Nov-15 | EPI_ISL_15958004 | A/domestic_duck/England/152473/2022 | Animal and Plant Health Agency (APHA) | Animal and Plant Health Agency (APHA) | Byrne, A.; Howton, N.; Maskell, D.; Mollett, B.; Peers-Dent, J. |
| EPI2233108 | NS | United Kingdom | 2022-Nov-15 | EPI_ISL_15958004 | A/domestic_duck/England/152473/2022 | Animal and Plant Health Agency (APHA) | Animal and Plant Health Agency (APHA) | Byrne, A.; Howton, N.; Maskell, D.; Mollett, B.; Peers-Dent, J. |
| EPI2233109 | MP | United Kingdom | 2022-Nov-15 | EPI_ISL_15958004 | A/domestic_duck/England/152473/2022 | Animal and Plant Health Agency (APHA) | Animal and Plant Health Agency (APHA) | Byrne, A.; Howton, N.; Maskell, D.; Mollett, B.; Peers-Dent, J. |
| EPI2233115 | NP | United Kingdom | 2022-Nov-03 | EPI_ISL_15958011 | A/Domestic_Duck/England/144945/2022 | Animal and Plant Health Agency (APHA) | Animal and Plant Health Agency (APHA) | Byrne, A.; Howton, N.; Maskell, D.; Mollett, B.; Peers-Dent, J. |
| EPI2233116 | NS | United Kingdom | 2022-Nov-03 | EPI_ISL_15958011 | A/Domestic_Duck/England/144945/2022 | Animal and Plant Health Agency (APHA) | Animal and Plant Health Agency (APHA) | Byrne, A.; Howton, N.; Maskell, D.; Mollett, B.; Peers-Dent, J. |
| EPI2233117 | MP | United Kingdom | 2022-Nov-03 | EPI_ISL_15958011 | A/Domestic_Duck/England/144945/2022 | Animal and Plant Health Agency (APHA) | Animal and Plant Health Agency (APHA) | Byrne, A.; Howton, N.; Maskell, D.; Mollett, B.; Peers-Dent, J. |
| EPI2233139 | NP | United Kingdom | 2022-Oct-21 | EPI_ISL_15958027 | A/Pheasant/Wales/394561/2022 | Animal and Plant Health Agency (APHA) | Animal and Plant Health Agency (APHA) | Byrne, A.; Howton, N.; Maskell, D.; Mollett, B.; Peers-Dent, J. |
| EPI2233140 | NS | United Kingdom | 2022-Oct-21 | EPI_ISL_15958027 | A/Pheasant/Wales/394561/2022 | Animal and Plant Health Agency (APHA) | Animal and Plant Health Agency (APHA) | Byrne, A.; Howton, N.; Maskell, D.; Mollett, B.; Peers-Dent, J. |
| EPI2233141 | MP | United Kingdom | 2022-Oct-21 | EPI_ISL_15958027 | A/Pheasant/Wales/394561/2022 | Animal and Plant Health Agency (APHA) | Animal and Plant Health Agency (APHA) | Byrne, A.; Howton, N.; Maskell, D.; Mollett, B.; Peers-Dent, J. |
| EPI2238844 | NP | Netherlands | 2022-Nov-21 | EPI_ISL_16040744 | A/Goose/Netherlands/22020788-001/2022 | Wageningen Bioveterinary Research | Wageningen Bioveterinary Research | Beerens, Nancy; Harders, Frank; Pritz-Verschuren, Sylvia; Roose, Marit; Venema, Sandra; Germeraad, Evelien; Engelsma, Marc; Heutink, Rene |
| EPI2238845 | NS | Netherlands | 2022-Nov-21 | EPI_ISL_16040744 | A/Goose/Netherlands/22020788-001/2022 | Wageningen Bioveterinary Research | Wageningen Bioveterinary Research | Beerens, Nancy; Harders, Frank; Pritz-Verschuren, Sylvia; Roose, Marit; Venema, Sandra; Germeraad, Evelien; Engelsma, Marc; Heutink, Rene |
| EPI2238846 | MP | Netherlands | 2022-Nov-21 | EPI_ISL_16040744 | A/Goose/Netherlands/22020788-001/2022 | Wageningen Bioveterinary Research | Wageningen Bioveterinary Research | Beerens, Nancy; Harders, Frank; Pritz-Verschuren, Sylvia; Roose, Marit; Venema, Sandra; Germeraad, Evelien; Engelsma, Marc; Heutink, Rene |
| EPI2240703 | NP | Netherlands | 2022-Nov-04 | EPI_ISL_16056568 | A/Eurasian Wigeon/Netherlands/5/2022 | Erasmus Medical Center | Erasmus Medical Center | |
| EPI2240704 | NS | Netherlands | 2022-Nov-04 | EPI_ISL_16056568 | A/Eurasian Wigeon/Netherlands/5/2022 | Erasmus Medical Center | Erasmus Medical Center | |
| EPI2240705 | MP | Netherlands | 2022-Nov-04 | EPI_ISL_16056568 | A/Eurasian Wigeon/Netherlands/5/2022 | Erasmus Medical Center | Erasmus Medical Center | |
| EPI2240711 | NP | Netherlands | 2022-Nov-15 | EPI_ISL_16056569 | A/Gadwall/Netherlands/3/2022 | Erasmus Medical Center | Erasmus Medical Center | |
| EPI2240712 | NS | Netherlands | 2022-Nov-15 | EPI_ISL_16056569 | A/Gadwall/Netherlands/3/2022 | Erasmus Medical Center | Erasmus Medical Center | |
| EPI2240713 | MP | Netherlands | 2022-Nov-15 | EPI_ISL_16056569 | A/Gadwall/Netherlands/3/2022 | Erasmus Medical Center | Erasmus Medical Center | |
| EPI2240719 | NP | Netherlands | 2022-Nov-08 | EPI_ISL_16056570 | A/Mallard Duck/Netherlands/27/2022 | Erasmus Medical Center | Erasmus Medical Center | |
| EPI2240720 | NS | Netherlands | 2022-Nov-08 | EPI_ISL_16056570 | A/Mallard Duck/Netherlands/27/2022 | Erasmus Medical Center | Erasmus Medical Center | |
| EPI2240721 | MP | Netherlands | 2022-Nov-08 | EPI_ISL_16056570 | A/Mallard Duck/Netherlands/27/2022 | Erasmus Medical Center | Erasmus Medical Center | |
| EPI2240727 | NP | Netherlands | 2022-Nov-09 | EPI_ISL_16056571 | A/Mallard Duck/Netherlands/28/2022 | Erasmus Medical Center | Erasmus Medical Center | |
| EPI2240728 | NS | Netherlands | 2022-Nov-09 | EPI_ISL_16056571 | A/Mallard Duck/Netherlands/28/2022 | Erasmus Medical Center | Erasmus Medical Center | |
| EPI2240729 | MP | Netherlands | 2022-Nov-09 | EPI_ISL_16056571 | A/Mallard Duck/Netherlands/28/2022 | Erasmus Medical Center | Erasmus Medical Center | |
| EPI2240735 | NP | Netherlands | 2022-Nov-17 | EPI_ISL_16056572 | A/Mallard Duck/Netherlands/29/2022 | Erasmus Medical Center | Erasmus Medical Center | |
| EPI2240736 | NS | Netherlands | 2022-Nov-17 | EPI_ISL_16056572 | A/Mallard Duck/Netherlands/29/2022 | Erasmus Medical Center | Erasmus Medical Center | |
| EPI2240737 | MP | Netherlands | 2022-Nov-17 | EPI_ISL_16056572 | A/Mallard Duck/Netherlands/29/2022 | Erasmus Medical Center | Erasmus Medical Center | |
| EPI2242295 | NS | Germany | 2022-Jun-30 | EPI_ISL_16096040 | A/artic tern/Germany-SH/AI03798/2022 | Landeslabor Schleswig-Holstein | Friedrich-Loeffler-Institut | |
| EPI2242294 | NP | Germany | 2022-Jun-30 | EPI_ISL_16096040 | A/artic tern/Germany-SH/AI03798/2022 | Landeslabor Schleswig-Holstein | Friedrich-Loeffler-Institut | |
| EPI2242292 | MP | Germany | 2022-Jun-30 | EPI_ISL_16096040 | A/artic tern/Germany-SH/AI03798/2022 | Landeslabor Schleswig-Holstein | Friedrich-Loeffler-Institut | |
| EPI2242300 | MP | Germany | 2022-Feb-07 | EPI_ISL_16096042 | A/black-headed gull/Germany-BY/AI01062/2022 | Bayrisches Landesamt für Gesundheit und Lebensmittelsicherheit (LGL) | Friedrich-Loeffler-Institut | |
| EPI2242302 | NP | Germany | 2022-Feb-07 | EPI_ISL_16096042 | A/black-headed gull/Germany-BY/AI01062/2022 | Bayrisches Landesamt für Gesundheit und Lebensmittelsicherheit (LGL) | Friedrich-Loeffler-Institut | |

| | | | | | | | | |
|---|---|---|---|---|---|---|---|---|
| EPI2242303 | NS | Germany | 2022-Feb-07 | EPI_ISL_16096042 | A/black-headed gull/Germany-BY/AI01062/2022 | Bayrisches Landesamt für Gesundheit und Lebensmittelsicherheit (LGL) | Friedrich-Loeffler-Institut | |
| EPI2242309 | MP | Germany | 2022-Jul-02 | EPI_ISL_16096044 | A/black-headed gull/Germany-HH/AI03795/2022 | Institut für Hygiene und Umwelt | Friedrich-Loeffler-Institut | |
| EPI2242311 | NP | Germany | 2022-Jul-02 | EPI_ISL_16096044 | A/black-headed gull/Germany-HH/AI03795/2022 | Institut für Hygiene und Umwelt | Friedrich-Loeffler-Institut | |
| EPI2242312 | NS | Germany | 2022-Jul-02 | EPI_ISL_16096044 | A/black-headed gull/Germany-HH/AI03795/2022 | Institut für Hygiene und Umwelt | Friedrich-Loeffler-Institut | |
| EPI2242318 | MP | Germany | 2022-Jul-04 | EPI_ISL_16096047 | A/black-headed gull/Germany-NI/AI04164/2022 | Lebensmittel- und Veterinärinstitut Oldenburg - Standort Veterinärinstitut | Friedrich-Loeffler-Institut | |
| EPI2242320 | NP | Germany | 2022-Jul-04 | EPI_ISL_16096047 | A/black-headed gull/Germany-NI/AI04164/2022 | Lebensmittel- und Veterinärinstitut Oldenburg - Standort Veterinärinstitut | Friedrich-Loeffler-Institut | |
| EPI2242321 | NS | Germany | 2022-Jul-04 | EPI_ISL_16096047 | A/black-headed gull/Germany-NI/AI04164/2022 | Lebensmittel- und Veterinärinstitut Oldenburg - Standort Veterinärinstitut | Friedrich-Loeffler-Institut | |
| EPI2242457 | MP | Germany | 2022-Jul-14 | EPI_ISL_16098026 | A/common tern/Germany-NI/AI04284/2022 | Lebensmittel- und Veterinärinstitut Oldenburg - Standort Veterinärinstitut | Friedrich-Loeffler-Institut | |
| EPI2242459 | NP | Germany | 2022-Jul-14 | EPI_ISL_16098026 | A/common tern/Germany-NI/AI04284/2022 | Lebensmittel- und Veterinärinstitut Oldenburg - Standort Veterinärinstitut | Friedrich-Loeffler-Institut | |
| EPI2242460 | NS | Germany | 2022-Jul-14 | EPI_ISL_16098026 | A/common tern/Germany-NI/AI04284/2022 | Lebensmittel- und Veterinärinstitut Oldenburg - Standort Veterinärinstitut | Friedrich-Loeffler-Institut | |
| EPI2244092 | NP | Denmark | 2022-Jun-19 | EPI_ISL_16112044 | A/Arctic_tern/Denmark/05803-1.01/2022 | Statens Serumm Institute | Statens Serum Institute | Hjulsager,Charlotte; Liang,Yuan |
| EPI2244094 | MP | Denmark | 2022-Jun-19 | EPI_ISL_16112044 | A/Arctic_tern/Denmark/05803-1.01/2022 | Statens Serumm Institute | Statens Serum Institute | Hjulsager,Charlotte; Liang,Yuan |
| EPI2244095 | NS | Denmark | 2022-Jun-19 | EPI_ISL_16112044 | A/Arctic_tern/Denmark/05803-1.01/2022 | Statens Serumm Institute | Statens Serum Institute | Hjulsager,Charlotte; Liang,Yuan |
| EPI2244157 | MP | Denmark | 2022-Apr-20 | EPI_ISL_16113089 | A/barnacle_goose/Denmark/03332-1.02/2022 | Statens Serum Institute | Statens Serum Institute | Hjulsager,Charlotte; Liang,Yuan |
| EPI2244158 | NS | Denmark | 2022-Apr-20 | EPI_ISL_16113089 | A/barnacle_goose/Denmark/03332-1.02/2022 | Statens Serum Institute | Statens Serum Institute | Hjulsager,Charlotte; Liang,Yuan |
| EPI2244155 | NP | Denmark | 2022-Apr-20 | EPI_ISL_16113089 | A/barnacle_goose/Denmark/03332-1.02/2022 | Statens Serum Institute | Statens Serum Institute | Hjulsager,Charlotte; Liang,Yuan |
| EPI2244211 | NP | Denmark | 2022-Jul-17 | EPI_ISL_16113090 | A/peregrine_falcon/Denmark/06880-1.02/2022 | Statens Serumm Institute | Statens Serum Institute | Hjulsager,Charlotte; Liang,Yuan |
| EPI2244213 | MP | Denmark | 2022-Jul-17 | EPI_ISL_16113090 | A/peregrine_falcon/Denmark/06880-1.02/2022 | Statens Serumm Institute | Statens Serum Institute | Hjulsager,Charlotte; Liang,Yuan |
| EPI2244214 | NS | Denmark | 2022-Jul-17 | EPI_ISL_16113090 | A/peregrine_falcon/Denmark/06880-1.02/2022 | Statens Serumm Institute | Statens Serum Institute | Hjulsager,Charlotte; Liang,Yuan |
| EPI2244205 | MP | Denmark | 2022-Jun-14 | EPI_ISL_16113225 | A/sandwich_tern/Denmark/05712-1.02/2022 | Statens Serum Institute | Statens Serum Institute | Hjulsager, Charlotte; Liang,Yuan |
| EPI2244206 | NS | Denmark | 2022-Jun-14 | EPI_ISL_16113225 | A/sandwich_tern/Denmark/05712-1.02/2022 | Statens Serum Institute | Statens Serum Institute | Hjulsager, Charlotte; Liang,Yuan |
| EPI2244203 | NP | Denmark | 2022-Jun-14 | EPI_ISL_16113225 | A/sandwich_tern/Denmark/05712-1.02/2022 | Statens Serum Institute | Statens Serum Institute | Hjulsager, Charlotte; Liang,Yuan |
| EPI2244197 | MP | Denmark | 2022-Jun-17 | EPI_ISL_16113229 | A/sandwich_tern/Denmark/05802-1.01_2022 | Statens Serum Institute | Statens Serum Institute | Hjulsager,Charlotte; Liang,Yuan |
| EPI2244195 | NP | Denmark | 2022-Jun-17 | EPI_ISL_16113229 | A/sandwich_tern/Denmark/05802-1.01_2022 | Statens Serum Institute | Statens Serum Institute | Hjulsager,Charlotte; Liang,Yuan |
| EPI2244198 | NS | Denmark | 2022-Jun-17 | EPI_ISL_16113229 | A/sandwich_tern/Denmark/05802-1.01_2022 | Statens Serum Institute | Statens Serum Institute | Hjulsager,Charlotte; Liang,Yuan |
| EPI2244187 | NP | Denmark | 2022-Jun-19 | EPI_ISL_16113230 | A/sandwich_tern/Denmark/05806-1.01/2022 | Statens Serumm Institute | Statens Serum Institute | Hjulsager,Charlotte; Liang,Yuan |
| EPI2244189 | MP | Denmark | 2022-Jun-19 | EPI_ISL_16113230 | A/sandwich_tern/Denmark/05806-1.01/2022 | Statens Serumm Institute | Statens Serum Institute | Hjulsager,Charlotte; Liang,Yuan |
| EPI2244190 | NS | Denmark | 2022-Jun-19 | EPI_ISL_16113230 | A/sandwich_tern/Denmark/05806-1.01/2022 | Statens Serumm Institute | Statens Serum Institute | Hjulsager,Charlotte; Liang,Yuan |
| EPI2244182 | NS | Denmark | 2022-Jun-08 | EPI_ISL_16113231 | A/tawny_owl/Denmark/05372-1.01/2022 | Statens Serumm Institute | Statens Serum Institute | Hjulsager,Charlotte; Liang,Yuan |
| EPI2244179 | NP | Denmark | 2022-Jun-08 | EPI_ISL_16113231 | A/tawny_owl/Denmark/05372-1.01/2022 | Statens Serumm Institute | Statens Serum Institute | Hjulsager,Charlotte; Liang,Yuan |
| EPI2244181 | MP | Denmark | 2022-Jun-08 | EPI_ISL_16113231 | A/tawny_owl/Denmark/05372-1.01/2022 | Statens Serumm Institute | Statens Serum Institute | Hjulsager,Charlotte; Liang,Yuan |
| EPI2244171 | NP | Denmark | 2022-Jun-08 | EPI_ISL_16113242 | A/tawny_owl/Denmark/05373-1.01/2022 | Statens Serum Institute | Statens Serum Institute | Hjulsager,Charlotte; Liang,Yuan |
| EPI2244174 | NS | Denmark | 2022-Jun-08 | EPI_ISL_16113242 | A/tawny_owl/Denmark/05373-1.01/2022 | Statens Serum Institute | Statens Serum Institute | Hjulsager,Charlotte; Liang,Yuan |
| EPI2244173 | MP | Denmark | 2022-Jun-08 | EPI_ISL_16113242 | A/tawny_owl/Denmark/05373-1.01/2022 | Statens Serum Institute | Statens Serum Institute | Hjulsager,Charlotte; Liang,Yuan |
| EPI2244166 | NS | Denmark | 2022-Jun-11 | EPI_ISL_16113366 | A/white-tailed_eagle/Denmark/05716-1.04/2022 | Statens Serum Institute | Statens Serum Institute | Hjulsager,Charlotte; Liang,Yuan |

| | | | | | | | | |
|---|---|---|---|---|---|---|---|---|
| EPI2244165 | MP | Denmark | 2022-Jun-11 | EPI_ISL_16113366 | A/white-tailed_eagle/Denmark/05716-1.04/2022 | Statens Serum Institute | Statens Serum Institute | Hjulsager,Charlotte; Liang,Yuan |
| EPI2244163 | NP | Denmark | 2022-Jun-11 | EPI_ISL_16113366 | A/white-tailed_eagle/Denmark/05716-1.04/2022 | Statens Serum Institute | Statens Serum Institute | Hjulsager,Charlotte; Liang,Yuan |
| EPI2244219 | NP | Denmark | 2022-Aug-30 | EPI_ISL_16115054 | A/common_buzzard/Denmark/08287-1.01/2022 | Statens Serum Institute | Statens Serum Institute | Hjulsager,Charlotte; Liang,Yuan |
| EPI2244222 | NS | Denmark | 2022-Aug-30 | EPI_ISL_16115054 | A/common_buzzard/Denmark/08287-1.01/2022 | Statens Serum Institute | Statens Serum Institute | Hjulsager,Charlotte; Liang,Yuan |
| EPI2244221 | MP | Denmark | 2022-Aug-30 | EPI_ISL_16115054 | A/common_buzzard/Denmark/08287-1.01/2022 | Statens Serum Institute | Statens Serum Institute | Hjulsager,Charlotte; Liang,Yuan |
| EPI2244299 | NS | Germany | 2022-Jul-25 | EPI_ISL_16122075 | A/domestic goose/Germany-SH/AI04267/2022 | Landeslabor Schleswig-Holstein | Friedrich-Loeffler-Institut | |
| EPI2244296 | MP | Germany | 2022-Jul-25 | EPI_ISL_16122075 | A/domestic goose/Germany-SH/AI04267/2022 | Landeslabor Schleswig-Holstein | Friedrich-Loeffler-Institut | |
| EPI2244298 | NP | Germany | 2022-Jul-25 | EPI_ISL_16122075 | A/domestic goose/Germany-SH/AI04267/2022 | Landeslabor Schleswig-Holstein | Friedrich-Loeffler-Institut | |
| EPI2244370 | NP | Germany | 2022-Jul-08 | EPI_ISL_16122154 | A/northern gannet/Germany-NI/AI04234/2022 | Lebensmittel- und Veterinärinstitut Oldenburg - Standort Veterinärinstitut | Friedrich-Loeffler-Institut | |
| EPI2244368 | MP | Germany | 2022-Jul-08 | EPI_ISL_16122154 | A/northern gannet/Germany-NI/AI04234/2022 | Lebensmittel- und Veterinärinstitut Oldenburg - Standort Veterinärinstitut | Friedrich-Loeffler-Institut | |
| EPI2244371 | NS | Germany | 2022-Jul-08 | EPI_ISL_16122154 | A/northern gannet/Germany-NI/AI04234/2022 | Lebensmittel- und Veterinärinstitut Oldenburg - Standort Veterinärinstitut | Friedrich-Loeffler-Institut | |
| EPI2254385 | MP | Luxembourg | 2022-Dec-11 | EPI_ISL_16206845 | A/goose/Luxembourg/22253283/2022 | Laboratoire de médecine vétérinaire de l'Etat | Luxembourg Institute of Health | Chantal Snoeck, Aurélie Sausy |
| EPI2254384 | NS | Luxembourg | 2022-Dec-11 | EPI_ISL_16206845 | A/goose/Luxembourg/22253283/2022 | Laboratoire de médecine vétérinaire de l'Etat | Luxembourg Institute of Health | Chantal Snoeck, Aurélie Sausy |
| EPI2254386 | NP | Luxembourg | 2022-Dec-11 | EPI_ISL_16206845 | A/goose/Luxembourg/22253283/2022 | Laboratoire de médecine vétérinaire de l'Etat | Luxembourg Institute of Health | Chantal Snoeck, Aurélie Sausy |
| EPI2259803 | NP | Netherlands | 2022-Dec-07 | EPI_ISL_16253341 | A/Mallard/Netherlands/31/2022 | Erasmus Medical Center | Erasmus Medical Center | |
| EPI2259804 | NS | Netherlands | 2022-Dec-07 | EPI_ISL_16253341 | A/Mallard/Netherlands/31/2022 | Erasmus Medical Center | Erasmus Medical Center | |
| EPI2259805 | MP | Netherlands | 2022-Dec-07 | EPI_ISL_16253341 | A/Mallard/Netherlands/31/2022 | Erasmus Medical Center | Erasmus Medical Center | |
| EPI2277802 | NP | United Kingdom | 2022-Oct-31 | EPI_ISL_16384125 | A/pheasant/England/331075/2022 | Animal and Plant Health Agency (APHA) | Animal and Plant Health Agency (APHA) | |
| EPI2277803 | NS | United Kingdom | 2022-Oct-31 | EPI_ISL_16384125 | A/pheasant/England/331075/2022 | Animal and Plant Health Agency (APHA) | Animal and Plant Health Agency (APHA) | |
| EPI2277804 | MP | United Kingdom | 2022-Oct-31 | EPI_ISL_16384125 | A/pheasant/England/331075/2022 | Animal and Plant Health Agency (APHA) | Animal and Plant Health Agency (APHA) | |
| EPI2277818 | NP | United Kingdom | 2022-Oct-28 | EPI_ISL_16384127 | A/grey_heron/England/142125/2022 | Animal and Plant Health Agency (APHA) | Animal and Plant Health Agency (APHA) | |
| EPI2277819 | NS | United Kingdom | 2022-Oct-28 | EPI_ISL_16384127 | A/grey_heron/England/142125/2022 | Animal and Plant Health Agency (APHA) | Animal and Plant Health Agency (APHA) | |
| EPI2277820 | MP | United Kingdom | 2022-Oct-28 | EPI_ISL_16384127 | A/grey_heron/England/142125/2022 | Animal and Plant Health Agency (APHA) | Animal and Plant Health Agency (APHA) | |
| EPI2277826 | NP | United Kingdom | 2022-Oct-27 | EPI_ISL_16384128 | A/turkey/England/141610/2022 | Animal and Plant Health Agency (APHA) | Animal and Plant Health Agency (APHA) | |
| EPI2277827 | NS | United Kingdom | 2022-Oct-27 | EPI_ISL_16384128 | A/turkey/England/141610/2022 | Animal and Plant Health Agency (APHA) | Animal and Plant Health Agency (APHA) | |
| EPI2277828 | MP | United Kingdom | 2022-Oct-27 | EPI_ISL_16384128 | A/turkey/England/141610/2022 | Animal and Plant Health Agency (APHA) | Animal and Plant Health Agency (APHA) | |
| EPI2277834 | NP | United Kingdom | 2022-Oct-21 | EPI_ISL_16384129 | A/turkey/England/138376/2022 | Animal and Plant Health Agency (APHA) | Animal and Plant Health Agency (APHA) | |
| EPI2277835 | NS | United Kingdom | 2022-Oct-21 | EPI_ISL_16384129 | A/turkey/England/138376/2022 | Animal and Plant Health Agency (APHA) | Animal and Plant Health Agency (APHA) | |
| EPI2277836 | MP | United Kingdom | 2022-Oct-21 | EPI_ISL_16384129 | A/turkey/England/138376/2022 | Animal and Plant Health Agency (APHA) | Animal and Plant Health Agency (APHA) | |
| EPI2277842 | NP | United Kingdom | 2022-Oct-20 | EPI_ISL_16384137 | A/domestic_duck/England/137953/2022 | Animal and Plant Health Agency (APHA) | Animal and Plant Health Agency (APHA) | |
| EPI2277843 | NS | United Kingdom | 2022-Oct-20 | EPI_ISL_16384137 | A/domestic_duck/England/137953/2022 | Animal and Plant Health Agency (APHA) | Animal and Plant Health Agency (APHA) | |
| EPI2277844 | MP | United Kingdom | 2022-Oct-20 | EPI_ISL_16384137 | A/domestic_duck/England/137953/2022 | Animal and Plant Health Agency (APHA) | Animal and Plant Health Agency (APHA) | |
| EPI2277850 | NP | United Kingdom | 2022-Oct-14 | EPI_ISL_16384138 | A/domestic_goose/England/132125/2022 | Animal and Plant Health Agency (APHA) | Animal and Plant Health Agency (APHA) | |
| EPI2277851 | NS | United Kingdom | 2022-Oct-14 | EPI_ISL_16384138 | A/domestic_goose/England/132125/2022 | Animal and Plant Health Agency (APHA) | Animal and Plant Health Agency (APHA) | |
| EPI2277852 | MP | United Kingdom | 2022-Oct-14 | EPI_ISL_16384138 | A/domestic_goose/England/132125/2022 | Animal and Plant Health Agency (APHA) | Animal and Plant Health Agency (APHA) | |

| | | | | | | | |
|---|---|---|---|---|---|---|---|
| EPI2277858 | NP | United Kingdom | 2022-Sep-22 | EPI_ISL_16384139 | A/chicken/England/121576/2022 | Animal and Plant Health Agency (APHA) | Animal and Plant Health Agency (APHA) |
| EPI2277859 | NS | United Kingdom | 2022-Sep-22 | EPI_ISL_16384139 | A/chicken/England/121576/2022 | Animal and Plant Health Agency (APHA) | Animal and Plant Health Agency (APHA) |
| EPI2277860 | MP | United Kingdom | 2022-Sep-22 | EPI_ISL_16384139 | A/chicken/England/121576/2022 | Animal and Plant Health Agency (APHA) | Animal and Plant Health Agency (APHA) |
| EPI2277866 | NP | United Kingdom | 2022-Nov-16 | EPI_ISL_16384140 | A/Domestic_goose/England/153783/2022 | Animal and Plant Health Agency (APHA) | Animal and Plant Health Agency (APHA) |
| EPI2277867 | NS | United Kingdom | 2022-Nov-16 | EPI_ISL_16384140 | A/Domestic_goose/England/153783/2022 | Animal and Plant Health Agency (APHA) | Animal and Plant Health Agency (APHA) |
| EPI2277868 | MP | United Kingdom | 2022-Nov-16 | EPI_ISL_16384140 | A/Domestic_goose/England/153783/2022 | Animal and Plant Health Agency (APHA) | Animal and Plant Health Agency (APHA) |
| EPI2277874 | NP | United Kingdom | 2022-Nov-16 | EPI_ISL_16384141 | A/Chicken/Scotland/153852/2022 | Animal and Plant Health Agency (APHA) | Animal and Plant Health Agency (APHA) |
| EPI2277875 | NS | United Kingdom | 2022-Nov-16 | EPI_ISL_16384141 | A/Chicken/Scotland/153852/2022 | Animal and Plant Health Agency (APHA) | Animal and Plant Health Agency (APHA) |
| EPI2277876 | MP | United Kingdom | 2022-Nov-16 | EPI_ISL_16384141 | A/Chicken/Scotland/153852/2022 | Animal and Plant Health Agency (APHA) | Animal and Plant Health Agency (APHA) |
| EPI2277882 | NP | United Kingdom | 2022-Nov-19 | EPI_ISL_16384142 | A/Chicken/England/154150/2022 | Animal and Plant Health Agency (APHA) | Animal and Plant Health Agency (APHA) |
| EPI2277883 | NS | United Kingdom | 2022-Nov-19 | EPI_ISL_16384142 | A/Chicken/England/154150/2022 | Animal and Plant Health Agency (APHA) | Animal and Plant Health Agency (APHA) |
| EPI2277884 | MP | United Kingdom | 2022-Nov-19 | EPI_ISL_16384142 | A/Chicken/England/154150/2022 | Animal and Plant Health Agency (APHA) | Animal and Plant Health Agency (APHA) |
| EPI2277890 | NP | United Kingdom | 2022-Nov-19 | EPI_ISL_16384143 | A/Turkey/England/154210/2022 | Animal and Plant Health Agency (APHA) | Animal and Plant Health Agency (APHA) |
| EPI2277891 | NS | United Kingdom | 2022-Nov-19 | EPI_ISL_16384143 | A/Turkey/England/154210/2022 | Animal and Plant Health Agency (APHA) | Animal and Plant Health Agency (APHA) |
| EPI2277892 | MP | United Kingdom | 2022-Nov-19 | EPI_ISL_16384143 | A/Turkey/England/154210/2022 | Animal and Plant Health Agency (APHA) | Animal and Plant Health Agency (APHA) |
| EPI2277898 | NP | United Kingdom | 2022-Nov-19 | EPI_ISL_16384144 | A/Domestic_duck/England/154301/2022 | Animal and Plant Health Agency (APHA) | Animal and Plant Health Agency (APHA) |
| EPI2277899 | NS | United Kingdom | 2022-Nov-19 | EPI_ISL_16384144 | A/Domestic_duck/England/154301/2022 | Animal and Plant Health Agency (APHA) | Animal and Plant Health Agency (APHA) |
| EPI2277900 | MP | United Kingdom | 2022-Nov-19 | EPI_ISL_16384144 | A/Domestic_duck/England/154301/2022 | Animal and Plant Health Agency (APHA) | Animal and Plant Health Agency (APHA) |
| EPI2277906 | NP | United Kingdom | 2022-Nov-19 | EPI_ISL_16384145 | A/Chicken/Scotland/153380/2022 | Animal and Plant Health Agency (APHA) | Animal and Plant Health Agency (APHA) |
| EPI2277907 | NS | United Kingdom | 2022-Nov-19 | EPI_ISL_16384145 | A/Chicken/Scotland/153380/2022 | Animal and Plant Health Agency (APHA) | Animal and Plant Health Agency (APHA) |
| EPI2277908 | MP | United Kingdom | 2022-Nov-19 | EPI_ISL_16384145 | A/Chicken/Scotland/153380/2022 | Animal and Plant Health Agency (APHA) | Animal and Plant Health Agency (APHA) |
| EPI2277915 | NS | United Kingdom | 2022-Nov-20 | EPI_ISL_16384146 | A/Chicken/England/154451/2022 | Animal and Plant Health Agency (APHA) | Animal and Plant Health Agency (APHA) |
| EPI2277916 | MP | United Kingdom | 2022-Nov-20 | EPI_ISL_16384146 | A/Chicken/England/154451/2022 | Animal and Plant Health Agency (APHA) | Animal and Plant Health Agency (APHA) |
| EPI2277914 | NP | United Kingdom | 2022-Nov-20 | EPI_ISL_16384146 | A/Chicken/England/154451/2022 | Animal and Plant Health Agency (APHA) | Animal and Plant Health Agency (APHA) |
| EPI2277922 | NP | United Kingdom | 2022-Nov-20 | EPI_ISL_16384147 | A/Chicken/England/155942/2022 | Animal and Plant Health Agency (APHA) | Animal and Plant Health Agency (APHA) |
| EPI2277923 | NS | United Kingdom | 2022-Nov-20 | EPI_ISL_16384147 | A/Chicken/England/155942/2022 | Animal and Plant Health Agency (APHA) | Animal and Plant Health Agency (APHA) |
| EPI2277924 | MP | United Kingdom | 2022-Nov-20 | EPI_ISL_16384147 | A/Chicken/England/155942/2022 | Animal and Plant Health Agency (APHA) | Animal and Plant Health Agency (APHA) |
| EPI2277930 | NP | United Kingdom | 2022-Nov-21 | EPI_ISL_16384148 | A/Turkey/England/156090/2022 | Animal and Plant Health Agency (APHA) | Animal and Plant Health Agency (APHA) |
| EPI2277931 | NS | United Kingdom | 2022-Nov-21 | EPI_ISL_16384148 | A/Turkey/England/156090/2022 | Animal and Plant Health Agency (APHA) | Animal and Plant Health Agency (APHA) |
| EPI2277932 | MP | United Kingdom | 2022-Nov-21 | EPI_ISL_16384148 | A/Turkey/England/156090/2022 | Animal and Plant Health Agency (APHA) | Animal and Plant Health Agency (APHA) |
| EPI2277962 | NP | United Kingdom | 2022-Nov-17 | EPI_ISL_16384152 | A/Barnacle_goose/Northern_Ireland/159014/2022 | Animal and Plant Health Agency (APHA) | Animal and Plant Health Agency (APHA) |
| EPI2277963 | NS | United Kingdom | 2022-Nov-17 | EPI_ISL_16384152 | A/Barnacle_goose/Northern_Ireland/159014/2022 | Animal and Plant Health Agency (APHA) | Animal and Plant Health Agency (APHA) |
| EPI2277964 | MP | United Kingdom | 2022-Nov-17 | EPI_ISL_16384152 | A/Barnacle_goose/Northern_Ireland/159014/2022 | Animal and Plant Health Agency (APHA) | Animal and Plant Health Agency (APHA) |
| EPI2277970 | NP | United Kingdom | 2022-Nov-25 | EPI_ISL_16384153 | A/Chicken/England/159386/2022 | Animal and Plant Health Agency (APHA) | Animal and Plant Health Agency (APHA) |
| EPI2277971 | NS | United Kingdom | 2022-Nov-25 | EPI_ISL_16384153 | A/Chicken/England/159386/2022 | Animal and Plant Health Agency (APHA) | Animal and Plant Health Agency (APHA) |

| | | | | | | | |
|---|---|---|---|---|---|---|---|
| EPI2277972 | MP | United Kingdom | 2022-Nov-25 | EPI_ISL_16384153 | A/Chicken/England/159386/2022 | Animal and Plant Health Agency (APHA) | Animal and Plant Health Agency (APHA) |
| EPI2277978 | NP | United Kingdom | 2022-Nov-26 | EPI_ISL_16384154 | A/Chicken/Scotland/159456/2022 | Animal and Plant Health Agency (APHA) | Animal and Plant Health Agency (APHA) |
| EPI2277979 | NS | United Kingdom | 2022-Nov-26 | EPI_ISL_16384154 | A/Chicken/Scotland/159456/2022 | Animal and Plant Health Agency (APHA) | Animal and Plant Health Agency (APHA) |
| EPI2277980 | MP | United Kingdom | 2022-Nov-26 | EPI_ISL_16384154 | A/Chicken/Scotland/159456/2022 | Animal and Plant Health Agency (APHA) | Animal and Plant Health Agency (APHA) |
| EPI2277986 | NP | United Kingdom | 2022-Nov-26 | EPI_ISL_16384155 | A/Chicken/England/159529/2022 | Animal and Plant Health Agency (APHA) | Animal and Plant Health Agency (APHA) |
| EPI2277987 | NS | United Kingdom | 2022-Nov-26 | EPI_ISL_16384155 | A/Chicken/England/159529/2022 | Animal and Plant Health Agency (APHA) | Animal and Plant Health Agency (APHA) |
| EPI2277988 | MP | United Kingdom | 2022-Nov-26 | EPI_ISL_16384155 | A/Chicken/England/159529/2022 | Animal and Plant Health Agency (APHA) | Animal and Plant Health Agency (APHA) |
| EPI2277994 | NP | United Kingdom | 2022-Nov-28 | EPI_ISL_16384156 | A/Domestic_duck/England/159770/2022 | Animal and Plant Health Agency (APHA) | Animal and Plant Health Agency (APHA) |
| EPI2277995 | NS | United Kingdom | 2022-Nov-28 | EPI_ISL_16384156 | A/Domestic_duck/England/159770/2022 | Animal and Plant Health Agency (APHA) | Animal and Plant Health Agency (APHA) |
| EPI2277996 | MP | United Kingdom | 2022-Nov-28 | EPI_ISL_16384156 | A/Domestic_duck/England/159770/2022 | Animal and Plant Health Agency (APHA) | Animal and Plant Health Agency (APHA) |
| EPI2278002 | NP | United Kingdom | 2022-Nov-29 | EPI_ISL_16384157 | A/Peregrine_falcon/England/161113/2022 | Animal and Plant Health Agency (APHA) | Animal and Plant Health Agency (APHA) |
| EPI2278003 | NS | United Kingdom | 2022-Nov-29 | EPI_ISL_16384157 | A/Peregrine_falcon/England/161113/2022 | Animal and Plant Health Agency (APHA) | Animal and Plant Health Agency (APHA) |
| EPI2278004 | MP | United Kingdom | 2022-Nov-29 | EPI_ISL_16384157 | A/Peregrine_falcon/England/161113/2022 | Animal and Plant Health Agency (APHA) | Animal and Plant Health Agency (APHA) |
| EPI2278010 | NP | United Kingdom | 2022-Nov-29 | EPI_ISL_16384158 | A/Domestic_Goose/England/161197/2022 | Animal and Plant Health Agency (APHA) | Animal and Plant Health Agency (APHA) |
| EPI2278011 | NS | United Kingdom | 2022-Nov-29 | EPI_ISL_16384158 | A/Domestic_Goose/England/161197/2022 | Animal and Plant Health Agency (APHA) | Animal and Plant Health Agency (APHA) |
| EPI2278012 | MP | United Kingdom | 2022-Nov-29 | EPI_ISL_16384158 | A/Domestic_Goose/England/161197/2022 | Animal and Plant Health Agency (APHA) | Animal and Plant Health Agency (APHA) |
| EPI2278020 | MP | United Kingdom | 2022-Dec-04 | EPI_ISL_16384159 | A/Humboldt_penguin/England/161651/2022 | Animal and Plant Health Agency (APHA) | Animal and Plant Health Agency (APHA) |
| EPI2278018 | NP | United Kingdom | 2022-Dec-04 | EPI_ISL_16384159 | A/Humboldt_penguin/England/161651/2022 | Animal and Plant Health Agency (APHA) | Animal and Plant Health Agency (APHA) |
| EPI2278019 | NS | United Kingdom | 2022-Dec-04 | EPI_ISL_16384159 | A/Humboldt_penguin/England/161651/2022 | Animal and Plant Health Agency (APHA) | Animal and Plant Health Agency (APHA) |
| EPI2278026 | NP | United Kingdom | 2022-Dec-06 | EPI_ISL_16384160 | A/chicken/Scotland/163935/2022 | Animal and Plant Health Agency (APHA) | Animal and Plant Health Agency (APHA) |
| EPI2278027 | NS | United Kingdom | 2022-Dec-06 | EPI_ISL_16384160 | A/chicken/Scotland/163935/2022 | Animal and Plant Health Agency (APHA) | Animal and Plant Health Agency (APHA) |
| EPI2278028 | MP | United Kingdom | 2022-Dec-06 | EPI_ISL_16384160 | A/chicken/Scotland/163935/2022 | Animal and Plant Health Agency (APHA) | Animal and Plant Health Agency (APHA) |
| EPI2278034 | NP | United Kingdom | 2022-Dec-06 | EPI_ISL_16384161 | A/domestic_duck/England/164793/2022 | Animal and Plant Health Agency (APHA) | Animal and Plant Health Agency (APHA) |
| EPI2278035 | NS | United Kingdom | 2022-Dec-06 | EPI_ISL_16384161 | A/domestic_duck/England/164793/2022 | Animal and Plant Health Agency (APHA) | Animal and Plant Health Agency (APHA) |
| EPI2278036 | MP | United Kingdom | 2022-Dec-06 | EPI_ISL_16384161 | A/domestic_duck/England/164793/2022 | Animal and Plant Health Agency (APHA) | Animal and Plant Health Agency (APHA) |
| EPI2278042 | NP | United Kingdom | 2022-Dec-14 | EPI_ISL_16384162 | A/chicken/England/169222/2022 | Animal and Plant Health Agency (APHA) | Animal and Plant Health Agency (APHA) |
| EPI2278043 | NS | United Kingdom | 2022-Dec-14 | EPI_ISL_16384162 | A/chicken/England/169222/2022 | Animal and Plant Health Agency (APHA) | Animal and Plant Health Agency (APHA) |
| EPI2278044 | MP | United Kingdom | 2022-Dec-14 | EPI_ISL_16384162 | A/chicken/England/169222/2022 | Animal and Plant Health Agency (APHA) | Animal and Plant Health Agency (APHA) |
| EPI2278050 | NP | United Kingdom | 2022-Dec-14 | EPI_ISL_16384163 | A/domestic_goose/England/169312/2022 | Animal and Plant Health Agency (APHA) | Animal and Plant Health Agency (APHA) |
| EPI2278051 | NS | United Kingdom | 2022-Dec-14 | EPI_ISL_16384163 | A/domestic_goose/England/169312/2022 | Animal and Plant Health Agency (APHA) | Animal and Plant Health Agency (APHA) |
| EPI2278052 | MP | United Kingdom | 2022-Dec-14 | EPI_ISL_16384163 | A/domestic_goose/England/169312/2022 | Animal and Plant Health Agency (APHA) | Animal and Plant Health Agency (APHA) |
| EPI2278074 | NP | United Kingdom | 2022-Nov-22 | EPI_ISL_16384166 | A/mute_swan/Scotland/164387/2022 | Animal and Plant Health Agency (APHA) | Animal and Plant Health Agency (APHA) |
| EPI2278075 | NS | United Kingdom | 2022-Nov-22 | EPI_ISL_16384166 | A/mute_swan/Scotland/164387/2022 | Animal and Plant Health Agency (APHA) | Animal and Plant Health Agency (APHA) |
| EPI2278076 | MP | United Kingdom | 2022-Nov-22 | EPI_ISL_16384166 | A/mute_swan/Scotland/164387/2022 | Animal and Plant Health Agency (APHA) | Animal and Plant Health Agency (APHA) |
| EPI2278090 | NP | United Kingdom | 2022-Nov-30 | EPI_ISL_16384168 | A/greylag_goose/England/255248/2022 | Animal and Plant Health Agency (APHA) | Animal and Plant Health Agency (APHA) |

| | | | | | | | |
|---|---|---|---|---|---|---|---|
| EPI2278091 | NS | United Kingdom | 2022-Nov-30 | EPI_ISL_16384168 | A/greylag_goose/England/255248/2022 | Animal and Plant Health Agency (APHA) | Animal and Plant Health Agency (APHA) |
| EPI2278092 | MP | United Kingdom | 2022-Nov-30 | EPI_ISL_16384168 | A/greylag_goose/England/255248/2022 | Animal and Plant Health Agency (APHA) | Animal and Plant Health Agency (APHA) |
| EPI2278146 | NP | United Kingdom | 2022-Dec-01 | EPI_ISL_16384175 | A/mute_swan/England/119719/2022 | Animal and Plant Health Agency (APHA) | Animal and Plant Health Agency (APHA) |
| EPI2278147 | NS | United Kingdom | 2022-Dec-01 | EPI_ISL_16384175 | A/mute_swan/England/119719/2022 | Animal and Plant Health Agency (APHA) | Animal and Plant Health Agency (APHA) |
| EPI2278148 | MP | United Kingdom | 2022-Dec-01 | EPI_ISL_16384175 | A/mute_swan/England/119719/2022 | Animal and Plant Health Agency (APHA) | Animal and Plant Health Agency (APHA) |
| EPI2278162 | NP | United Kingdom | 2022-Dec-01 | EPI_ISL_16384177 | A/canada_goose/England/119759/2022 | Animal and Plant Health Agency (APHA) | Animal and Plant Health Agency (APHA) |
| EPI2278163 | NS | United Kingdom | 2022-Dec-01 | EPI_ISL_16384177 | A/canada_goose/England/119759/2022 | Animal and Plant Health Agency (APHA) | Animal and Plant Health Agency (APHA) |
| EPI2278164 | MP | United Kingdom | 2022-Dec-01 | EPI_ISL_16384177 | A/canada_goose/England/119759/2022 | Animal and Plant Health Agency (APHA) | Animal and Plant Health Agency (APHA) |
| EPI2278178 | NP | United Kingdom | 2022-Dec-05 | EPI_ISL_16384179 | A/mute_swan/England/119898/2022 | Animal and Plant Health Agency (APHA) | Animal and Plant Health Agency (APHA) |
| EPI2278179 | NS | United Kingdom | 2022-Dec-05 | EPI_ISL_16384179 | A/mute_swan/England/119898/2022 | Animal and Plant Health Agency (APHA) | Animal and Plant Health Agency (APHA) |
| EPI2278180 | MP | United Kingdom | 2022-Dec-05 | EPI_ISL_16384179 | A/mute_swan/England/119898/2022 | Animal and Plant Health Agency (APHA) | Animal and Plant Health Agency (APHA) |
| EPI2278186 | NP | United Kingdom | 2022-Dec-06 | EPI_ISL_16384180 | A/greylag_goose/England/119911/2022 | Animal and Plant Health Agency (APHA) | Animal and Plant Health Agency (APHA) |
| EPI2278187 | NS | United Kingdom | 2022-Dec-06 | EPI_ISL_16384180 | A/greylag_goose/England/119911/2022 | Animal and Plant Health Agency (APHA) | Animal and Plant Health Agency (APHA) |
| EPI2278188 | MP | United Kingdom | 2022-Dec-06 | EPI_ISL_16384180 | A/greylag_goose/England/119911/2022 | Animal and Plant Health Agency (APHA) | Animal and Plant Health Agency (APHA) |
| EPI2278194 | NP | United Kingdom | 2022-Dec-06 | EPI_ISL_16384181 | A/mute_swan/England/119920/2022 | Animal and Plant Health Agency (APHA) | Animal and Plant Health Agency (APHA) |
| EPI2278195 | NS | United Kingdom | 2022-Dec-06 | EPI_ISL_16384181 | A/mute_swan/England/119920/2022 | Animal and Plant Health Agency (APHA) | Animal and Plant Health Agency (APHA) |
| EPI2278196 | MP | United Kingdom | 2022-Dec-06 | EPI_ISL_16384181 | A/mute_swan/England/119920/2022 | Animal and Plant Health Agency (APHA) | Animal and Plant Health Agency (APHA) |
| EPI2278202 | NP | United Kingdom | 2022-Dec-06 | EPI_ISL_16384182 | A/greylag_goose/England/119927/2022 | Animal and Plant Health Agency (APHA) | Animal and Plant Health Agency (APHA) |
| EPI2278203 | NS | United Kingdom | 2022-Dec-06 | EPI_ISL_16384182 | A/greylag_goose/England/119927/2022 | Animal and Plant Health Agency (APHA) | Animal and Plant Health Agency (APHA) |
| EPI2278204 | MP | United Kingdom | 2022-Dec-06 | EPI_ISL_16384182 | A/greylag_goose/England/119927/2022 | Animal and Plant Health Agency (APHA) | Animal and Plant Health Agency (APHA) |
| EPI2278282 | NP | United Kingdom | 2022-Dec-05 | EPI_ISL_16384192 | A/greylag_goose/England/334954/2022 | Animal and Plant Health Agency (APHA) | Animal and Plant Health Agency (APHA) |
| EPI2278283 | NS | United Kingdom | 2022-Dec-05 | EPI_ISL_16384192 | A/greylag_goose/England/334954/2022 | Animal and Plant Health Agency (APHA) | Animal and Plant Health Agency (APHA) |
| EPI2278284 | MP | United Kingdom | 2022-Dec-05 | EPI_ISL_16384192 | A/greylag_goose/England/334954/2022 | Animal and Plant Health Agency (APHA) | Animal and Plant Health Agency (APHA) |
| EPI2278306 | NP | United Kingdom | 2022-Nov-30 | EPI_ISL_16384195 | A/mute_swan/England/395870/2022 | Animal and Plant Health Agency (APHA) | Animal and Plant Health Agency (APHA) |
| EPI2278307 | NS | United Kingdom | 2022-Nov-30 | EPI_ISL_16384195 | A/mute_swan/England/395870/2022 | Animal and Plant Health Agency (APHA) | Animal and Plant Health Agency (APHA) |
| EPI2278308 | MP | United Kingdom | 2022-Nov-30 | EPI_ISL_16384195 | A/mute_swan/England/395870/2022 | Animal and Plant Health Agency (APHA) | Animal and Plant Health Agency (APHA) |
| EPI2278314 | NP | United Kingdom | 2022-Nov-30 | EPI_ISL_16384196 | A/canada_goose/England/395873/2022 | Animal and Plant Health Agency (APHA) | Animal and Plant Health Agency (APHA) |
| EPI2278315 | NS | United Kingdom | 2022-Nov-30 | EPI_ISL_16384196 | A/canada_goose/England/395873/2022 | Animal and Plant Health Agency (APHA) | Animal and Plant Health Agency (APHA) |
| EPI2278316 | MP | United Kingdom | 2022-Nov-30 | EPI_ISL_16384196 | A/canada_goose/England/395873/2022 | Animal and Plant Health Agency (APHA) | Animal and Plant Health Agency (APHA) |
| EPI2278330 | NP | United Kingdom | 2022-Nov-13 | EPI_ISL_16384198 | A/Chicken/England/167913/2022 | Animal and Plant Health Agency (APHA) | Animal and Plant Health Agency (APHA) |
| EPI2278331 | NS | United Kingdom | 2022-Nov-13 | EPI_ISL_16384198 | A/Chicken/England/167913/2022 | Animal and Plant Health Agency (APHA) | Animal and Plant Health Agency (APHA) |
| EPI2278332 | MP | United Kingdom | 2022-Nov-13 | EPI_ISL_16384198 | A/Chicken/England/167913/2022 | Animal and Plant Health Agency (APHA) | Animal and Plant Health Agency (APHA) |
| EPI2278338 | NP | United Kingdom | 2022-Dec-19 | EPI_ISL_16384199 | A/domestic_duck/England/170578/2022 | Animal and Plant Health Agency (APHA) | Animal and Plant Health Agency (APHA) |
| EPI2278339 | NS | United Kingdom | 2022-Dec-19 | EPI_ISL_16384199 | A/domestic_duck/England/170578/2022 | Animal and Plant Health Agency (APHA) | Animal and Plant Health Agency (APHA) |
| EPI2278340 | MP | United Kingdom | 2022-Dec-19 | EPI_ISL_16384199 | A/domestic_duck/England/170578/2022 | Animal and Plant Health Agency (APHA) | Animal and Plant Health Agency (APHA) |

| | | | | | | | | |
|---|---|---|---|---|---|---|---|---|
| EPI2281682 | NS | France | 2022-Dec-24 | EPI_ISL_16395206 | A/cat/France/22P026544/2022 | Anses (Ploufragan-Plouzané) | ANSES Agence Nationale De Securite Sanitaire De L'alimentation | |
| EPI2281681 | MP | France | 2022-Dec-24 | EPI_ISL_16395206 | A/cat/France/22P026544/2022 | Anses (Ploufragan-Plouzané) | ANSES Agence Nationale De Securite Sanitaire De L'alimentation | |
| EPI2281679 | NP | France | 2022-Dec-24 | EPI_ISL_16395206 | A/cat/France/22P026544/2022 | Anses (Ploufragan-Plouzané) | ANSES Agence Nationale De Securite Sanitaire De L'alimentation | |
| EPI2281683 | NP | United Kingdom | 2022-Oct-18 | EPI_ISL_16398192 | A/chicken/England/137719/2022 | Animal and Plant Health Agency (APHA) | Animal and Plant Health Agency (APHA) | |
| EPI2281684 | NS | United Kingdom | 2022-Oct-18 | EPI_ISL_16398192 | A/chicken/England/137719/2022 | Animal and Plant Health Agency (APHA) | Animal and Plant Health Agency (APHA) | |
| EPI2281685 | MP | United Kingdom | 2022-Oct-18 | EPI_ISL_16398192 | A/chicken/England/137719/2022 | Animal and Plant Health Agency (APHA) | Animal and Plant Health Agency (APHA) | |
| EPI2283154 | NP | United Kingdom | 2022-Dec-12 | EPI_ISL_16443924 | A/domestic_duck/England/167913/2022 | Animal and Plant Health Agency (APHA) | Animal and Plant Health Agency (APHA) | |
| EPI2283155 | NS | United Kingdom | 2022-Dec-12 | EPI_ISL_16443924 | A/domestic_duck/England/167913/2022 | Animal and Plant Health Agency (APHA) | Animal and Plant Health Agency (APHA) | |
| EPI2283156 | MP | United Kingdom | 2022-Dec-12 | EPI_ISL_16443924 | A/domestic_duck/England/167913/2022 | Animal and Plant Health Agency (APHA) | Animal and Plant Health Agency (APHA) | |
| EPI2283162 | NS | United Kingdom | 2022-Dec-10 | EPI_ISL_16444163 | A/domestic_duck/England/165186/2022 | Animal and Plant Health Agency (APHA) | Animal and Plant Health Agency (APHA) | |
| EPI2283163 | MP | United Kingdom | 2022-Dec-10 | EPI_ISL_16444163 | A/domestic_duck/England/165186/2022 | Animal and Plant Health Agency (APHA) | Animal and Plant Health Agency (APHA) | |
| EPI2300848 | NP | United Kingdom | 2022-Dec-10 | EPI_ISL_16444163 | A/domestic_duck/England/165186/2022 | Animal and Plant Health Agency (APHA) | Animal and Plant Health Agency (APHA) | |
| EPI2291495 | NP | Spain | 2022-Aug-21 | EPI_ISL_16507094 | A/northern_gannet/Spain/22VIR12774-1_3004-12/2022 | Istituto Zooprofilattico Sperimentale delle Venezie, EU/OIE/Reference Laboratory and FAO Reference Centre for AI and ND | Istituto Zooprofilattico Sperimentale Delle Venezie | Ruano,M.J.; Rocha,A.; Sanchez,A.; Aguero,M.; Pastori, A.; Zecchin, B.; Fusaro, A.; Schivo, A.; Salviato, A.; Palumbo, E.; Giussani, E.; Monne, I.; Terregino, C. |
| EPI2291496 | NS | Spain | 2022-Aug-21 | EPI_ISL_16507094 | A/northern_gannet/Spain/22VIR12774-1_3004-12/2022 | Istituto Zooprofilattico Sperimentale delle Venezie, EU/OIE/Reference Laboratory and FAO Reference Centre for AI and ND | Istituto Zooprofilattico Sperimentale Delle Venezie | Ruano,M.J.; Rocha,A.; Sanchez,A.; Aguero,M.; Pastori, A.; Zecchin, B.; Fusaro, A.; Schivo, A.; Salviato, A.; Palumbo, E.; Giussani, E.; Monne, I.; Terregino, C. |
| EPI2291497 | MP | Spain | 2022-Aug-21 | EPI_ISL_16507094 | A/northern_gannet/Spain/22VIR12774-1_3004-12/2022 | Istituto Zooprofilattico Sperimentale delle Venezie, EU/OIE/Reference Laboratory and FAO Reference Centre for AI and ND | Istituto Zooprofilattico Sperimentale Delle Venezie | Ruano,M.J.; Rocha,A.; Sanchez,A.; Aguero,M.; Pastori, A.; Zecchin, B.; Fusaro, A.; Schivo, A.; Salviato, A.; Palumbo, E.; Giussani, E.; Monne, I.; Terregino, C. |
| EPI2291503 | NP | Spain | 2022-Oct-26 | EPI_ISL_16507095 | A/mink/Spain/22VIR12774-13_3869-2/2022 | Istituto Zooprofilattico Sperimentale delle Venezie, EU/OIE/Reference Laboratory and FAO Reference Centre for AI and ND | Istituto Zooprofilattico Sperimentale Delle Venezie | Ruano,M.J.; Rocha,A.; Sanchez,A.; Aguero,M.; Pastori, A.; Zecchin, B.; Fusaro, A.; Schivo, A.; Salviato, A.; Palumbo, E.; Giussani, E.; Monne, I.; Terregino, C. |
| EPI2291504 | NS | Spain | 2022-Oct-26 | EPI_ISL_16507095 | A/mink/Spain/22VIR12774-13_3869-2/2022 | Istituto Zooprofilattico Sperimentale delle Venezie, EU/OIE/Reference Laboratory and FAO Reference Centre for AI and ND | Istituto Zooprofilattico Sperimentale Delle Venezie | Ruano,M.J.; Rocha,A.; Sanchez,A.; Aguero,M.; Pastori, A.; Zecchin, B.; Fusaro, A.; Schivo, A.; Salviato, A.; Palumbo, E.; Giussani, E.; Monne, I.; Terregino, C. |
| EPI2291505 | MP | Spain | 2022-Oct-26 | EPI_ISL_16507095 | A/mink/Spain/22VIR12774-13_3869-2/2022 | Istituto Zooprofilattico Sperimentale delle Venezie, EU/OIE/Reference Laboratory and FAO Reference Centre for AI and ND | Istituto Zooprofilattico Sperimentale Delle Venezie | Ruano,M.J.; Rocha,A.; Sanchez,A.; Aguero,M.; Pastori, A.; Zecchin, B.; Fusaro, A.; Schivo, A.; Salviato, A.; Palumbo, E.; Giussani, E.; Monne, I.; Terregino, C. |
| EPI2291513 | MP | Spain | 2022-Oct-26 | EPI_ISL_16507096 | A/mink/Spain/22VIR12774-14_3869-3/2022 | Istituto Zooprofilattico Sperimentale delle Venezie, EU/OIE/Reference Laboratory and FAO Reference Centre for AI and ND | Istituto Zooprofilattico Sperimentale Delle Venezie | Ruano,M.J.; Rocha,A.; Sanchez,A.; Aguero,M.; Pastori, A.; Zecchin, B.; Fusaro, A.; Schivo, A.; Salviato, A.; Palumbo, E.; Giussani, E.; Monne, I.; Terregino, C. |
| EPI2291511 | NP | Spain | 2022-Oct-26 | EPI_ISL_16507096 | A/mink/Spain/22VIR12774-14_3869-3/2022 | Istituto Zooprofilattico Sperimentale delle Venezie, EU/OIE/Reference Laboratory and FAO Reference Centre for AI and ND | Istituto Zooprofilattico Sperimentale Delle Venezie | Ruano,M.J.; Rocha,A.; Sanchez,A.; Aguero,M.; Pastori, A.; Zecchin, B.; Fusaro, A.; Schivo, A.; Salviato, A.; Palumbo, E.; Giussani, E.; Monne, I.; Terregino, C. |
| EPI2291512 | NS | Spain | 2022-Oct-26 | EPI_ISL_16507096 | A/mink/Spain/22VIR12774-14_3869-3/2022 | Istituto Zooprofilattico Sperimentale delle Venezie, EU/OIE/Reference Laboratory and FAO Reference Centre for AI and ND | Istituto Zooprofilattico Sperimentale Delle Venezie | Ruano,M.J.; Rocha,A.; Sanchez,A.; Aguero,M.; Pastori, A.; Zecchin, B.; Fusaro, A.; Schivo, A.; Salviato, A.; Palumbo, E.; Giussani, E.; Monne, I.; Terregino, C. |
| EPI2291519 | NP | Spain | 2022-Nov-17 | EPI_ISL_16507097 | A/seagull/Spain/22VIR12774-15_4347-15/2022 | Istituto Zooprofilattico Sperimentale delle Venezie, EU/OIE/Reference Laboratory and FAO Reference Centre for AI and ND | Istituto Zooprofilattico Sperimentale Delle Venezie | Ruano,M.J.; Rocha,A.; Sanchez,A.; Aguero,M.; Pastori, A.; Zecchin, B.; Fusaro, A.; Schivo, A.; Salviato, A.; Palumbo, E.; Giussani, E.; Monne, I.; Terregino, C. |
| EPI2291520 | NS | Spain | 2022-Nov-17 | EPI_ISL_16507097 | A/seagull/Spain/22VIR12774-15_4347-15/2022 | Istituto Zooprofilattico Sperimentale delle Venezie, EU/OIE/Reference Laboratory and FAO Reference Centre for AI and ND | Istituto Zooprofilattico Sperimentale Delle Venezie | Ruano,M.J.; Rocha,A.; Sanchez,A.; Aguero,M.; Pastori, A.; Zecchin, B.; Fusaro, A.; Schivo, A.; Salviato, A.; Palumbo, E.; Giussani, E.; Monne, I.; Terregino, C. |
| EPI2291521 | MP | Spain | 2022-Nov-17 | EPI_ISL_16507097 | A/seagull/Spain/22VIR12774-15_4347-15/2022 | Istituto Zooprofilattico Sperimentale delle Venezie, EU/OIE/Reference Laboratory and FAO Reference Centre for AI and ND | Istituto Zooprofilattico Sperimentale Delle Venezie | Ruano,M.J.; Rocha,A.; Sanchez,A.; Aguero,M.; Pastori, A.; Zecchin, B.; Fusaro, A.; Schivo, A.; Salviato, A.; Palumbo, E.; Giussani, E.; Monne, I.; Terregino, C. |
| EPI2291527 | NP | Spain | 2022-Aug-29 | EPI_ISL_16507098 | A/wild_goose/Spain/22VIR12774-16_3024-6/2022 | Istituto Zooprofilattico Sperimentale delle Venezie, EU/OIE/Reference Laboratory and FAO Reference Centre for AI and ND | Istituto Zooprofilattico Sperimentale Delle Venezie | Ruano,M.J.; Rocha,A.; Sanchez,A.; Aguero,M.; Pastori, A.; Zecchin, B.; Fusaro, A.; Schivo, A.; Salviato, A.; Palumbo, E.; Giussani, E.; Monne, I.; Terregino, C. |
| EPI2291528 | NS | Spain | 2022-Aug-29 | EPI_ISL_16507098 | A/wild_goose/Spain/22VIR12774-16_3024-6/2022 | Istituto Zooprofilattico Sperimentale delle Venezie, EU/OIE/Reference Laboratory and FAO Reference Centre for AI and ND | Istituto Zooprofilattico Sperimentale Delle Venezie | Ruano,M.J.; Rocha,A.; Sanchez,A.; Aguero,M.; Pastori, A.; Zecchin, B.; Fusaro, A.; Schivo, A.; Salviato, A.; Palumbo, E.; Giussani, E.; Monne, I.; Terregino, C. |
| EPI2291529 | MP | Spain | 2022-Aug-29 | EPI_ISL_16507098 | A/wild_goose/Spain/22VIR12774-16_3024-6/2022 | Istituto Zooprofilattico Sperimentale delle Venezie, EU/OIE/Reference Laboratory and FAO Reference Centre for AI and ND | Istituto Zooprofilattico Sperimentale Delle Venezie | Ruano,M.J.; Rocha,A.; Sanchez,A.; Aguero,M.; Pastori, A.; Zecchin, B.; Fusaro, A.; Schivo, A.; Salviato, A.; Palumbo, E.; Giussani, E.; Monne, I.; Terregino, C. |
| EPI2291535 | NP | Spain | 2022-Sep-27 | EPI_ISL_16507100 | A/northern_gannet/Spain/22VIR12774-17_3389-5/2022 | Istituto Zooprofilattico Sperimentale delle Venezie, EU/OIE/Reference Laboratory and FAO Reference Centre for AI and ND | Istituto Zooprofilattico Sperimentale Delle Venezie | Ruano,M.J.; Rocha,A.; Sanchez,A.; Aguero,M.; Pastori, A.; Zecchin, B.; Fusaro, A.; Schivo, A.; Salviato, A.; Palumbo, E.; Giussani, E.; Monne, I.; Terregino, C. |
| EPI2291536 | NS | Spain | 2022-Sep-27 | EPI_ISL_16507100 | A/northern_gannet/Spain/22VIR12774-17_3389-5/2022 | Istituto Zooprofilattico Sperimentale delle Venezie, EU/OIE/Reference Laboratory and FAO Reference Centre for AI and ND | Istituto Zooprofilattico Sperimentale Delle Venezie | Ruano,M.J.; Rocha,A.; Sanchez,A.; Aguero,M.; Pastori, A.; Zecchin, B.; Fusaro, A.; Schivo, A.; Salviato, A.; Palumbo, E.; Giussani, E.; Monne, I.; Terregino, C. |
| EPI2291537 | MP | Spain | 2022-Sep-27 | EPI_ISL_16507100 | A/northern_gannet/Spain/22VIR12774-17_3389-5/2022 | Istituto Zooprofilattico Sperimentale delle Venezie, EU/OIE/Reference Laboratory and FAO Reference Centre for AI and ND | Istituto Zooprofilattico Sperimentale Delle Venezie | Ruano,M.J.; Rocha,A.; Sanchez,A.; Aguero,M.; Pastori, A.; Zecchin, B.; Fusaro, A.; Schivo, A.; Salviato, A.; Palumbo, E.; Giussani, E.; Monne, I.; Terregino, C. |
| EPI2291543 | NP | Spain | 2022-Sep-24 | EPI_ISL_16507101 | A/northern_gannet/Spain/22VIR12774-18_3454-2/2022 | Istituto Zooprofilattico Sperimentale delle Venezie, EU/OIE/Reference Laboratory and FAO Reference Centre for AI and ND | Istituto Zooprofilattico Sperimentale Delle Venezie | Ruano,M.J.; Rocha,A.; Sanchez,A.; Aguero,M.; Pastori, A.; Zecchin, B.; Fusaro, A.; Schivo, A.; Salviato, A.; Palumbo, E.; Giussani, E.; Monne, I.; Terregino, C. |
| EPI2291544 | NS | Spain | 2022-Sep-24 | EPI_ISL_16507101 | A/northern_gannet/Spain/22VIR12774-18_3454-2/2022 | Istituto Zooprofilattico Sperimentale delle Venezie, EU/OIE/Reference Laboratory and FAO Reference Centre for AI and ND | Istituto Zooprofilattico Sperimentale Delle Venezie | Ruano,M.J.; Rocha,A.; Sanchez,A.; Aguero,M.; Pastori, A.; Zecchin, B.; Fusaro, A.; Schivo, A.; Salviato, A.; Palumbo, E.; Giussani, E.; Monne, I.; Terregino, C. |
| EPI2291545 | MP | Spain | 2022-Sep-24 | EPI_ISL_16507101 | A/northern_gannet/Spain/22VIR12774-18_3454-2/2022 | Istituto Zooprofilattico Sperimentale delle Venezie, EU/OIE/Reference Laboratory and FAO Reference Centre for AI and ND | Istituto Zooprofilattico Sperimentale Delle Venezie | Ruano,M.J.; Rocha,A.; Sanchez,A.; Aguero,M.; Pastori, A.; Zecchin, B.; Fusaro, A.; Schivo, A.; Salviato, A.; Palumbo, E.; Giussani, E.; Monne, I.; Terregino, C. |
| EPI2291552 | MP | Spain | 2022-Sep-29 | EPI_ISL_16507102 | A/Egretta_garzetta/Spain/22VIR12774-19_3519-2/2022 | Istituto Zooprofilattico Sperimentale delle Venezie, EU/OIE/Reference Laboratory and FAO Reference Centre for AI and ND | Istituto Zooprofilattico Sperimentale Delle Venezie | Ruano,M.J.; Rocha,A.; Sanchez,A.; Aguero,M.; Pastori, A.; Zecchin, B.; Fusaro, A.; Schivo, A.; Salviato, A.; Palumbo, E.; Giussani, E.; Monne, I.; Terregino, C. |
| EPI2291550 | NP | Spain | 2022-Sep-29 | EPI_ISL_16507102 | A/Egretta_garzetta/Spain/22VIR12774-19_3519-2/2022 | Istituto Zooprofilattico Sperimentale delle Venezie, EU/OIE/Reference Laboratory and FAO Reference Centre for AI and ND | Istituto Zooprofilattico Sperimentale Delle Venezie | Ruano,M.J.; Rocha,A.; Sanchez,A.; Aguero,M.; Pastori, A.; Zecchin, B.; Fusaro, A.; Schivo, A.; Salviato, A.; Palumbo, E.; Giussani, E.; Monne, I.; Terregino, C. |

| | | | | | | | | |
|---|---|---|---|---|---|---|---|---|
| EPI2291551 | NS | Spain | 2022-Sep-29 | EPI_ISL_16507102 | A/Egretta_garzetta/Spain/22VIR12774-19_3519-2/2022 | Istituto Zooprofilattico Sperimentale delle Venezie, EU/OIE/Reference Laboratory and FAO Reference Centre for AI and ND | Istituto Zooprofilattico Sperimentale Delle Venezie | Ruano,M.J.; Rocha,A.; Sanchez,A.; Aguero,M.; Pastori, A.; Zecchin, B.; Fusaro, A.; Schivo, A.; Salviato, A.; Palumbo, E.; Giussani, E.; Monne, I.; Terregino, C. |
| EPI2291558 | NP | Spain | 2022-Sep-21 | EPI_ISL_16507103 | A/northern_gannet/Spain/22VIR12774-20_3614-2/2022 | Istituto Zooprofilattico Sperimentale delle Venezie, EU/OIE/Reference Laboratory and FAO Reference Centre for AI and ND | Istituto Zooprofilattico Sperimentale Delle Venezie | Ruano,M.J.; Rocha,A.; Sanchez,A.; Aguero,M.; Pastori, A.; Zecchin, B.; Fusaro, A.; Schivo, A.; Salviato, A.; Palumbo, E.; Giussani, E.; Monne, I.; Terregino, C. |
| EPI2291559 | NS | Spain | 2022-Sep-21 | EPI_ISL_16507103 | A/northern_gannet/Spain/22VIR12774-20_3614-2/2022 | Istituto Zooprofilattico Sperimentale delle Venezie, EU/OIE/Reference Laboratory and FAO Reference Centre for AI and ND | Istituto Zooprofilattico Sperimentale Delle Venezie | Ruano,M.J.; Rocha,A.; Sanchez,A.; Aguero,M.; Pastori, A.; Zecchin, B.; Fusaro, A.; Schivo, A.; Salviato, A.; Palumbo, E.; Giussani, E.; Monne, I.; Terregino, C. |
| EPI2291560 | MP | Spain | 2022-Sep-21 | EPI_ISL_16507103 | A/northern_gannet/Spain/22VIR12774-20_3614-2/2022 | Istituto Zooprofilattico Sperimentale delle Venezie, EU/OIE/Reference Laboratory and FAO Reference Centre for AI and ND | Istituto Zooprofilattico Sperimentale Delle Venezie | Ruano,M.J.; Rocha,A.; Sanchez,A.; Aguero,M.; Pastori, A.; Zecchin, B.; Fusaro, A.; Schivo, A.; Salviato, A.; Palumbo, E.; Giussani, E.; Monne, I.; Terregino, C. |
| EPI2291565 | NP | Spain | 2022-Oct-17 | EPI_ISL_16507104 | A/wild_goose/Spain/22VIR12774-21_3814-3/2022 | Istituto Zooprofilattico Sperimentale delle Venezie, EU/OIE/Reference Laboratory and FAO Reference Centre for AI and ND | Istituto Zooprofilattico Sperimentale Delle Venezie | Ruano,M.J.; Rocha,A.; Sanchez,A.; Aguero,M.; Pastori, A.; Zecchin, B.; Fusaro, A.; Schivo, A.; Salviato, A.; Palumbo, E.; Giussani, E.; Monne, I.; Terregino, C. |
| EPI2291566 | NS | Spain | 2022-Oct-17 | EPI_ISL_16507104 | A/wild_goose/Spain/22VIR12774-21_3814-3/2022 | Istituto Zooprofilattico Sperimentale delle Venezie, EU/OIE/Reference Laboratory and FAO Reference Centre for AI and ND | Istituto Zooprofilattico Sperimentale Delle Venezie | Ruano,M.J.; Rocha,A.; Sanchez,A.; Aguero,M.; Pastori, A.; Zecchin, B.; Fusaro, A.; Schivo, A.; Salviato, A.; Palumbo, E.; Giussani, E.; Monne, I.; Terregino, C. |
| EPI2291567 | MP | Spain | 2022-Oct-17 | EPI_ISL_16507104 | A/wild_goose/Spain/22VIR12774-21_3814-3/2022 | Istituto Zooprofilattico Sperimentale delle Venezie, EU/OIE/Reference Laboratory and FAO Reference Centre for AI and ND | Istituto Zooprofilattico Sperimentale Delle Venezie | Ruano,M.J.; Rocha,A.; Sanchez,A.; Aguero,M.; Pastori, A.; Zecchin, B.; Fusaro, A.; Schivo, A.; Salviato, A.; Palumbo, E.; Giussani, E.; Monne, I.; Terregino, C. |
| EPI2291573 | NP | Spain | 2022-Sep-01 | EPI_ISL_16507105 | A/northern_gannet/Spain/22VIR12774-29_3076-18/2022 | Istituto Zooprofilattico Sperimentale delle Venezie, EU/OIE/Reference Laboratory and FAO Reference Centre for AI and ND | Istituto Zooprofilattico Sperimentale Delle Venezie | Ruano,M.J.; Rocha,A.; Sanchez,A.; Aguero,M.; Pastori, A.; Zecchin, B.; Fusaro, A.; Schivo, A.; Salviato, A.; Palumbo, E.; Giussani, E.; Monne, I.; Terregino, C. |
| EPI2291574 | NS | Spain | 2022-Sep-01 | EPI_ISL_16507105 | A/northern_gannet/Spain/22VIR12774-29_3076-18/2022 | Istituto Zooprofilattico Sperimentale delle Venezie, EU/OIE/Reference Laboratory and FAO Reference Centre for AI and ND | Istituto Zooprofilattico Sperimentale Delle Venezie | Ruano,M.J.; Rocha,A.; Sanchez,A.; Aguero,M.; Pastori, A.; Zecchin, B.; Fusaro, A.; Schivo, A.; Salviato, A.; Palumbo, E.; Giussani, E.; Monne, I.; Terregino, C. |
| EPI2291575 | MP | Spain | 2022-Sep-01 | EPI_ISL_16507105 | A/northern_gannet/Spain/22VIR12774-29_3076-18/2022 | Istituto Zooprofilattico Sperimentale delle Venezie, EU/OIE/Reference Laboratory and FAO Reference Centre for AI and ND | Istituto Zooprofilattico Sperimentale Delle Venezie | Ruano,M.J.; Rocha,A.; Sanchez,A.; Aguero,M.; Pastori, A.; Zecchin, B.; Fusaro, A.; Schivo, A.; Salviato, A.; Palumbo, E.; Giussani, E.; Monne, I.; Terregino, C. |
| EPI2291580 | NP | Spain | 2022-Sep-19 | EPI_ISL_16507106 | A/Ciconia_ciconia/Spain/22VIR12774-30_3394-2/2022 | Istituto Zooprofilattico Sperimentale delle Venezie, EU/OIE/Reference Laboratory and FAO Reference Centre for AI and ND | Istituto Zooprofilattico Sperimentale Delle Venezie | Ruano,M.J.; Rocha,A.; Sanchez,A.; Aguero,M.; Pastori, A.; Zecchin, B.; Fusaro, A.; Schivo, A.; Salviato, A.; Palumbo, E.; Giussani, E.; Monne, I.; Terregino, C. |
| EPI2291581 | NS | Spain | 2022-Sep-19 | EPI_ISL_16507106 | A/Ciconia_ciconia/Spain/22VIR12774-30_3394-2/2022 | Istituto Zooprofilattico Sperimentale delle Venezie, EU/OIE/Reference Laboratory and FAO Reference Centre for AI and ND | Istituto Zooprofilattico Sperimentale Delle Venezie | Ruano,M.J.; Rocha,A.; Sanchez,A.; Aguero,M.; Pastori, A.; Zecchin, B.; Fusaro, A.; Schivo, A.; Salviato, A.; Palumbo, E.; Giussani, E.; Monne, I.; Terregino, C. |
| EPI2291582 | MP | Spain | 2022-Sep-19 | EPI_ISL_16507106 | A/Ciconia_ciconia/Spain/22VIR12774-30_3394-2/2022 | Istituto Zooprofilattico Sperimentale delle Venezie, EU/OIE/Reference Laboratory and FAO Reference Centre for AI and ND | Istituto Zooprofilattico Sperimentale Delle Venezie | Ruano,M.J.; Rocha,A.; Sanchez,A.; Aguero,M.; Pastori, A.; Zecchin, B.; Fusaro, A.; Schivo, A.; Salviato, A.; Palumbo, E.; Giussani, E.; Monne, I.; Terregino, C. |
| EPI2291588 | NP | Spain | 2022-Sep-25 | EPI_ISL_16507107 | A/northern_gannet/Spain/22VIR12774-31_3453-1/2022 | Istituto Zooprofilattico Sperimentale delle Venezie, EU/OIE/Reference Laboratory and FAO Reference Centre for AI and ND | Istituto Zooprofilattico Sperimentale Delle Venezie | Ruano,M.J.; Rocha,A.; Sanchez,A.; Aguero,M.; Pastori, A.; Zecchin, B.; Fusaro, A.; Schivo, A.; Salviato, A.; Palumbo, E.; Giussani, E.; Monne, I.; Terregino, C. |
| EPI2291589 | NS | Spain | 2022-Sep-25 | EPI_ISL_16507107 | A/northern_gannet/Spain/22VIR12774-31_3453-1/2022 | Istituto Zooprofilattico Sperimentale delle Venezie, EU/OIE/Reference Laboratory and FAO Reference Centre for AI and ND | Istituto Zooprofilattico Sperimentale Delle Venezie | Ruano,M.J.; Rocha,A.; Sanchez,A.; Aguero,M.; Pastori, A.; Zecchin, B.; Fusaro, A.; Schivo, A.; Salviato, A.; Palumbo, E.; Giussani, E.; Monne, I.; Terregino, C. |
| EPI2291590 | MP | Spain | 2022-Sep-25 | EPI_ISL_16507107 | A/northern_gannet/Spain/22VIR12774-31_3453-1/2022 | Istituto Zooprofilattico Sperimentale delle Venezie, EU/OIE/Reference Laboratory and FAO Reference Centre for AI and ND | Istituto Zooprofilattico Sperimentale Delle Venezie | Ruano,M.J.; Rocha,A.; Sanchez,A.; Aguero,M.; Pastori, A.; Zecchin, B.; Fusaro, A.; Schivo, A.; Salviato, A.; Palumbo, E.; Giussani, E.; Monne, I.; Terregino, C. |
| EPI2291596 | NP | Spain | 2022-Oct-03 | EPI_ISL_16507108 | A/northern_gannet/Spain/22VIR12774-33_3497-10/2022 | Istituto Zooprofilattico Sperimentale delle Venezie, EU/OIE/Reference Laboratory and FAO Reference Centre for AI and ND | Istituto Zooprofilattico Sperimentale Delle Venezie | Ruano,M.J.; Rocha,A.; Sanchez,A.; Aguero,M.; Pastori, A.; Zecchin, B.; Fusaro, A.; Schivo, A.; Salviato, A.; Palumbo, E.; Giussani, E.; Monne, I.; Terregino, C. |
| EPI2291597 | NS | Spain | 2022-Oct-03 | EPI_ISL_16507108 | A/northern_gannet/Spain/22VIR12774-33_3497-10/2022 | Istituto Zooprofilattico Sperimentale delle Venezie, EU/OIE/Reference Laboratory and FAO Reference Centre for AI and ND | Istituto Zooprofilattico Sperimentale Delle Venezie | Ruano,M.J.; Rocha,A.; Sanchez,A.; Aguero,M.; Pastori, A.; Zecchin, B.; Fusaro, A.; Schivo, A.; Salviato, A.; Palumbo, E.; Giussani, E.; Monne, I.; Terregino, C. |
| EPI2291598 | MP | Spain | 2022-Oct-03 | EPI_ISL_16507108 | A/northern_gannet/Spain/22VIR12774-33_3497-10/2022 | Istituto Zooprofilattico Sperimentale delle Venezie, EU/OIE/Reference Laboratory and FAO Reference Centre for AI and ND | Istituto Zooprofilattico Sperimentale Delle Venezie | Ruano,M.J.; Rocha,A.; Sanchez,A.; Aguero,M.; Pastori, A.; Zecchin, B.; Fusaro, A.; Schivo, A.; Salviato, A.; Palumbo, E.; Giussani, E.; Monne, I.; Terregino, C. |
| EPI2291604 | NP | Spain | 2022-Sep-29 | EPI_ISL_16507109 | A/Ciconia_ciconia/Spain/22VIR12774-34_3519-1/2022 | Istituto Zooprofilattico Sperimentale delle Venezie, EU/OIE/Reference Laboratory and FAO Reference Centre for AI and ND | Istituto Zooprofilattico Sperimentale Delle Venezie | Ruano,M.J.; Rocha,A.; Sanchez,A.; Aguero,M.; Pastori, A.; Zecchin, B.; Fusaro, A.; Schivo, A.; Salviato, A.; Palumbo, E.; Giussani, E.; Monne, I.; Terregino, C. |
| EPI2291605 | NS | Spain | 2022-Sep-29 | EPI_ISL_16507109 | A/Ciconia_ciconia/Spain/22VIR12774-34_3519-1/2022 | Istituto Zooprofilattico Sperimentale delle Venezie, EU/OIE/Reference Laboratory and FAO Reference Centre for AI and ND | Istituto Zooprofilattico Sperimentale Delle Venezie | Ruano,M.J.; Rocha,A.; Sanchez,A.; Aguero,M.; Pastori, A.; Zecchin, B.; Fusaro, A.; Schivo, A.; Salviato, A.; Palumbo, E.; Giussani, E.; Monne, I.; Terregino, C. |
| EPI2291606 | MP | Spain | 2022-Sep-29 | EPI_ISL_16507109 | A/Ciconia_ciconia/Spain/22VIR12774-34_3519-1/2022 | Istituto Zooprofilattico Sperimentale delle Venezie, EU/OIE/Reference Laboratory and FAO Reference Centre for AI and ND | Istituto Zooprofilattico Sperimentale Delle Venezie | Ruano,M.J.; Rocha,A.; Sanchez,A.; Aguero,M.; Pastori, A.; Zecchin, B.; Fusaro, A.; Schivo, A.; Salviato, A.; Palumbo, E.; Giussani, E.; Monne, I.; Terregino, C. |
| EPI2291612 | NP | Spain | 2022-Oct-07 | EPI_ISL_16507176 | A/northern_gannet/Spain/22VIR12774-36_3583-6/2022 | Istituto Zooprofilattico Sperimentale delle Venezie, EU/OIE/Reference Laboratory and FAO Reference Centre for AI and ND | Istituto Zooprofilattico Sperimentale Delle Venezie | Ruano,M.J.; Rocha,A.; Sanchez,A.; Aguero,M.; Pastori, A.; Zecchin, B.; Fusaro, A.; Schivo, A.; Salviato, A.; Palumbo, E.; Giussani, E.; Monne, I.; Terregino, C. |
| EPI2291613 | NS | Spain | 2022-Oct-07 | EPI_ISL_16507176 | A/northern_gannet/Spain/22VIR12774-36_3583-6/2022 | Istituto Zooprofilattico Sperimentale delle Venezie, EU/OIE/Reference Laboratory and FAO Reference Centre for AI and ND | Istituto Zooprofilattico Sperimentale Delle Venezie | Ruano,M.J.; Rocha,A.; Sanchez,A.; Aguero,M.; Pastori, A.; Zecchin, B.; Fusaro, A.; Schivo, A.; Salviato, A.; Palumbo, E.; Giussani, E.; Monne, I.; Terregino, C. |
| EPI2291614 | MP | Spain | 2022-Oct-07 | EPI_ISL_16507176 | A/northern_gannet/Spain/22VIR12774-36_3583-6/2022 | Istituto Zooprofilattico Sperimentale delle Venezie, EU/OIE/Reference Laboratory and FAO Reference Centre for AI and ND | Istituto Zooprofilattico Sperimentale Delle Venezie | Ruano,M.J.; Rocha,A.; Sanchez,A.; Aguero,M.; Pastori, A.; Zecchin, B.; Fusaro, A.; Schivo, A.; Salviato, A.; Palumbo, E.; Giussani, E.; Monne, I.; Terregino, C. |
| EPI2291625 | MP | Spain | 2022-Oct-04 | EPI_ISL_16507177 | A/Ciconia_ciconia/Spain/22VIR12774-37_3641-1/2022 | Istituto Zooprofilattico Sperimentale delle Venezie, EU/OIE/Reference Laboratory and FAO Reference Centre for AI and ND | Istituto Zooprofilattico Sperimentale Delle Venezie | Ruano,M.J.; Rocha,A.; Sanchez,A.; Aguero,M.; Pastori, A.; Zecchin, B.; Fusaro, A.; Schivo, A.; Salviato, A.; Palumbo, E.; Giussani, E.; Monne, I.; Terregino, C. |
| EPI2291621 | NP | Spain | 2022-Oct-04 | EPI_ISL_16507177 | A/Ciconia_ciconia/Spain/22VIR12774-37_3641-1/2022 | Istituto Zooprofilattico Sperimentale delle Venezie, EU/OIE/Reference Laboratory and FAO Reference Centre for AI and ND | Istituto Zooprofilattico Sperimentale Delle Venezie | Ruano,M.J.; Rocha,A.; Sanchez,A.; Aguero,M.; Pastori, A.; Zecchin, B.; Fusaro, A.; Schivo, A.; Salviato, A.; Palumbo, E.; Giussani, E.; Monne, I.; Terregino, C. |
| EPI2291623 | NS | Spain | 2022-Oct-04 | EPI_ISL_16507177 | A/Ciconia_ciconia/Spain/22VIR12774-37_3641-1/2022 | Istituto Zooprofilattico Sperimentale delle Venezie, EU/OIE/Reference Laboratory and FAO Reference Centre for AI and ND | Istituto Zooprofilattico Sperimentale Delle Venezie | Ruano,M.J.; Rocha,A.; Sanchez,A.; Aguero,M.; Pastori, A.; Zecchin, B.; Fusaro, A.; Schivo, A.; Salviato, A.; Palumbo, E.; Giussani, E.; Monne, I.; Terregino, C. |
| EPI2291624 | MP | Spain | 2022-Oct-04 | EPI_ISL_16507177 | A/Ciconia_ciconia/Spain/22VIR12774-37_3641-1/2022 | Istituto Zooprofilattico Sperimentale delle Venezie, EU/OIE/Reference Laboratory and FAO Reference Centre for AI and ND | Istituto Zooprofilattico Sperimentale Delle Venezie | Ruano,M.J.; Rocha,A.; Sanchez,A.; Aguero,M.; Pastori, A.; Zecchin, B.; Fusaro, A.; Schivo, A.; Salviato, A.; Palumbo, E.; Giussani, E.; Monne, I.; Terregino, C. |
| EPI2292910 | NP | Netherlands | 2022-Dec-17 | EPI_ISL_16524187 | A/Common gull/Netherlands/2/2022 | Erasmus Medical Center | Erasmus Medical Center | |
| EPI2292911 | NS | Netherlands | 2022-Dec-17 | EPI_ISL_16524187 | A/Common gull/Netherlands/2/2022 | Erasmus Medical Center | Erasmus Medical Center | |
| EPI2292912 | MP | Netherlands | 2022-Dec-17 | EPI_ISL_16524187 | A/Common gull/Netherlands/2/2022 | Erasmus Medical Center | Erasmus Medical Center | |
| EPI2292919 | NS | Netherlands | 2022-Dec-23 | EPI_ISL_16524203 | A/Black-headed gull/Netherlands/12/2022 | Erasmus Medical Center | Erasmus Medical Center | |
| EPI2292920 | MP | Netherlands | 2022-Dec-23 | EPI_ISL_16524203 | A/Black-headed gull/Netherlands/12/2022 | Erasmus Medical Center | Erasmus Medical Center | |
| EPI2292918 | NP | Netherlands | 2022-Dec-23 | EPI_ISL_16524203 | A/Black-headed gull/Netherlands/12/2022 | Erasmus Medical Center | Erasmus Medical Center | |

| | | | | | | | | |
|---|---|---|---|---|---|---|---|---|
| EPI2292926 | NP | Netherlands | 2023-Jan-01 | EPI_ISL_16524213 | A/European herring gull/Netherlands/1/2023 | Erasmus Medical Center | Erasmus Medical Center | |
| EPI2292927 | NS | Netherlands | 2023-Jan-01 | EPI_ISL_16524213 | A/European herring gull/Netherlands/1/2023 | Erasmus Medical Center | Erasmus Medical Center | |
| EPI2292928 | MP | Netherlands | 2023-Jan-01 | EPI_ISL_16524213 | A/European herring gull/Netherlands/1/2023 | Erasmus Medical Center | Erasmus Medical Center | |
| EPI2292934 | NP | Netherlands | 2023-Jan-01 | EPI_ISL_16524230 | A/Black-headed gull/Netherlands/2/2023 | Erasmus Medical Center | Erasmus Medical Center | |
| EPI2292935 | NS | Netherlands | 2023-Jan-01 | EPI_ISL_16524230 | A/Black-headed gull/Netherlands/2/2023 | Erasmus Medical Center | Erasmus Medical Center | |
| EPI2292936 | MP | Netherlands | 2023-Jan-01 | EPI_ISL_16524230 | A/Black-headed gull/Netherlands/2/2023 | Erasmus Medical Center | Erasmus Medical Center | |
| EPI2292941 | NP | Netherlands | 2023-Jan-01 | EPI_ISL_16524243 | A/European herring gull/Netherlands/2/2023 | Erasmus Medical Center | Erasmus Medical Center | |
| EPI2292942 | NS | Netherlands | 2023-Jan-01 | EPI_ISL_16524243 | A/European herring gull/Netherlands/2/2023 | Erasmus Medical Center | Erasmus Medical Center | |
| EPI2292943 | MP | Netherlands | 2023-Jan-01 | EPI_ISL_16524243 | A/European herring gull/Netherlands/2/2023 | Erasmus Medical Center | Erasmus Medical Center | |
| EPI2292946 | NP | Netherlands | 2023-Jan-01 | EPI_ISL_16524245 | A/European herring gull/Netherlands/3/2023 | Erasmus Medical Center | Erasmus Medical Center | |
| EPI2292947 | NS | Netherlands | 2023-Jan-01 | EPI_ISL_16524245 | A/European herring gull/Netherlands/3/2023 | Erasmus Medical Center | Erasmus Medical Center | |
| EPI2292948 | MP | Netherlands | 2023-Jan-01 | EPI_ISL_16524245 | A/European herring gull/Netherlands/3/2023 | Erasmus Medical Center | Erasmus Medical Center | |
| EPI2292953 | NP | Netherlands | 2023-Jan-01 | EPI_ISL_16524246 | A/Caspian gull/Netherlands/1/2023 | Erasmus Medical Center | Erasmus Medical Center | |
| EPI2292954 | NS | Netherlands | 2023-Jan-01 | EPI_ISL_16524246 | A/Caspian gull/Netherlands/1/2023 | Erasmus Medical Center | Erasmus Medical Center | |
| EPI2292955 | MP | Netherlands | 2023-Jan-01 | EPI_ISL_16524246 | A/Caspian gull/Netherlands/1/2023 | Erasmus Medical Center | Erasmus Medical Center | |
| EPI2292969 | NP | Netherlands | 2022-Dec-27 | EPI_ISL_16524248 | A/European herring gull/Netherlands/21/2022 | Erasmus Medical Center | Erasmus Medical Center | |
| EPI2292970 | NS | Netherlands | 2022-Dec-27 | EPI_ISL_16524248 | A/European herring gull/Netherlands/21/2022 | Erasmus Medical Center | Erasmus Medical Center | |
| EPI2292971 | MP | Netherlands | 2022-Dec-27 | EPI_ISL_16524248 | A/European herring gull/Netherlands/21/2022 | Erasmus Medical Center | Erasmus Medical Center | |
| EPI2292985 | NP | Spain | 2022-Oct-18 | EPI_ISL_16524591 | A/Ciconia_ciconia/Spain/22VIR12774-40_3803-2/2022 | Istituto Zooprofilattico Sperimentale delle Venezie, EU/OIE/Reference Laboratory and FAO Reference Centre for AI and ND | Istituto Zooprofilattico Sperimentale Delle Venezie | Ruano,M.J.; Rocha,A.; Sanchez,A.; Aguero,M.; Pastori, A.; Zecchin, B.; Fusaro, A.; Schivo, A.; Salviato, A.; Palumbo, E.; Giussani, E.; Monne, I.; Terregino, C. |
| EPI2292986 | NS | Spain | 2022-Oct-18 | EPI_ISL_16524591 | A/Ciconia_ciconia/Spain/22VIR12774-40_3803-2/2022 | Istituto Zooprofilattico Sperimentale delle Venezie, EU/OIE/Reference Laboratory and FAO Reference Centre for AI and ND | Istituto Zooprofilattico Sperimentale Delle Venezie | Ruano,M.J.; Rocha,A.; Sanchez,A.; Aguero,M.; Pastori, A.; Zecchin, B.; Fusaro, A.; Schivo, A.; Salviato, A.; Palumbo, E.; Giussani, E.; Monne, I.; Terregino, C. |
| EPI2292987 | MP | Spain | 2022-Oct-18 | EPI_ISL_16524591 | A/Ciconia_ciconia/Spain/22VIR12774-40_3803-2/2022 | Istituto Zooprofilattico Sperimentale delle Venezie, EU/OIE/Reference Laboratory and FAO Reference Centre for AI and ND | Istituto Zooprofilattico Sperimentale Delle Venezie | Ruano,M.J.; Rocha,A.; Sanchez,A.; Aguero,M.; Pastori, A.; Zecchin, B.; Fusaro, A.; Schivo, A.; Salviato, A.; Palumbo, E.; Giussani, E.; Monne, I.; Terregino, C. |
| EPI2292994 | NS | Spain | 2022-Oct-18 | EPI_ISL_16524593 | A/Anser_anser/Spain/22VIR12774-41_3803-4/2022 | Istituto Zooprofilattico Sperimentale delle Venezie, EU/OIE/Reference Laboratory and FAO Reference Centre for AI and ND | Istituto Zooprofilattico Sperimentale Delle Venezie | Ruano,M.J.; Rocha,A.; Sanchez,A.; Aguero,M.; Pastori, A.; Zecchin, B.; Fusaro, A.; Schivo, A.; Salviato, A.; Palumbo, E.; Giussani, E.; Monne, I.; Terregino, C. |
| EPI2292995 | MP | Spain | 2022-Oct-18 | EPI_ISL_16524593 | A/Anser_anser/Spain/22VIR12774-41_3803-4/2022 | Istituto Zooprofilattico Sperimentale delle Venezie, EU/OIE/Reference Laboratory and FAO Reference Centre for AI and ND | Istituto Zooprofilattico Sperimentale Delle Venezie | Ruano,M.J.; Rocha,A.; Sanchez,A.; Aguero,M.; Pastori, A.; Zecchin, B.; Fusaro, A.; Schivo, A.; Salviato, A.; Palumbo, E.; Giussani, E.; Monne, I.; Terregino, C. |
| EPI2292993 | NP | Spain | 2022-Oct-18 | EPI_ISL_16524593 | A/Anser_anser/Spain/22VIR12774-41_3803-4/2022 | Istituto Zooprofilattico Sperimentale delle Venezie, EU/OIE/Reference Laboratory and FAO Reference Centre for AI and ND | Istituto Zooprofilattico Sperimentale Delle Venezie | Ruano,M.J.; Rocha,A.; Sanchez,A.; Aguero,M.; Pastori, A.; Zecchin, B.; Fusaro, A.; Schivo, A.; Salviato, A.; Palumbo, E.; Giussani, E.; Monne, I.; Terregino, C. |
| EPI2293001 | NP | Spain | 2022-Nov-08 | EPI_ISL_16524594 | A/wild_goose/Spain/22VIR12774-43_4213-1/2022 | Istituto Zooprofilattico Sperimentale delle Venezie, EU/OIE/Reference Laboratory and FAO Reference Centre for AI and ND | Istituto Zooprofilattico Sperimentale Delle Venezie | Ruano,M.J.; Rocha,A.; Sanchez,A.; Aguero,M.; Pastori, A.; Zecchin, B.; Fusaro, A.; Schivo, A.; Salviato, A.; Palumbo, E.; Giussani, E.; Monne, I.; Terregino, C. |
| EPI2293002 | NS | Spain | 2022-Nov-08 | EPI_ISL_16524594 | A/wild_goose/Spain/22VIR12774-43_4213-1/2022 | Istituto Zooprofilattico Sperimentale delle Venezie, EU/OIE/Reference Laboratory and FAO Reference Centre for AI and ND | Istituto Zooprofilattico Sperimentale Delle Venezie | Ruano,M.J.; Rocha,A.; Sanchez,A.; Aguero,M.; Pastori, A.; Zecchin, B.; Fusaro, A.; Schivo, A.; Salviato, A.; Palumbo, E.; Giussani, E.; Monne, I.; Terregino, C. |
| EPI2293003 | MP | Spain | 2022-Nov-08 | EPI_ISL_16524594 | A/wild_goose/Spain/22VIR12774-43_4213-1/2022 | Istituto Zooprofilattico Sperimentale delle Venezie, EU/OIE/Reference Laboratory and FAO Reference Centre for AI and ND | Istituto Zooprofilattico Sperimentale Delle Venezie | Ruano,M.J.; Rocha,A.; Sanchez,A.; Aguero,M.; Pastori, A.; Zecchin, B.; Fusaro, A.; Schivo, A.; Salviato, A.; Palumbo, E.; Giussani, E.; Monne, I.; Terregino, C. |
| EPI2293009 | NP | Spain | 2022-Nov-21 | EPI_ISL_16524595 | A/swan/Spain/22VIR12774-45_4382-2/2022 | Istituto Zooprofilattico Sperimentale delle Venezie, EU/OIE/Reference Laboratory and FAO Reference Centre for AI and ND | Istituto Zooprofilattico Sperimentale Delle Venezie | Ruano,M.J.; Rocha,A.; Sanchez,A.; Aguero,M.; Pastori, A.; Zecchin, B.; Fusaro, A.; Schivo, A.; Salviato, A.; Palumbo, E.; Giussani, E.; Monne, I.; Terregino, C. |
| EPI2293010 | NS | Spain | 2022-Nov-21 | EPI_ISL_16524595 | A/swan/Spain/22VIR12774-45_4382-2/2022 | Istituto Zooprofilattico Sperimentale delle Venezie, EU/OIE/Reference Laboratory and FAO Reference Centre for AI and ND | Istituto Zooprofilattico Sperimentale Delle Venezie | Ruano,M.J.; Rocha,A.; Sanchez,A.; Aguero,M.; Pastori, A.; Zecchin, B.; Fusaro, A.; Schivo, A.; Salviato, A.; Palumbo, E.; Giussani, E.; Monne, I.; Terregino, C. |
| EPI2293011 | MP | Spain | 2022-Nov-21 | EPI_ISL_16524595 | A/swan/Spain/22VIR12774-45_4382-2/2022 | Istituto Zooprofilattico Sperimentale delle Venezie, EU/OIE/Reference Laboratory and FAO Reference Centre for AI and ND | Istituto Zooprofilattico Sperimentale Delle Venezie | Ruano,M.J.; Rocha,A.; Sanchez,A.; Aguero,M.; Pastori, A.; Zecchin, B.; Fusaro, A.; Schivo, A.; Salviato, A.; Palumbo, E.; Giussani, E.; Monne, I.; Terregino, C. |
| EPI2293017 | NP | Spain | 2022-Aug-30 | EPI_ISL_16524596 | A/northern_gannet/Spain/22VIR12774-4_3104-40/2022 | Istituto Zooprofilattico Sperimentale delle Venezie, EU/OIE/Reference Laboratory and FAO Reference Centre for AI and ND | Istituto Zooprofilattico Sperimentale Delle Venezie | Ruano,M.J.; Rocha,A.; Sanchez,A.; Aguero,M.; Pastori, A.; Zecchin, B.; Fusaro, A.; Schivo, A.; Salviato, A.; Palumbo, E.; Giussani, E.; Monne, I.; Terregino, C. |
| EPI2293018 | NS | Spain | 2022-Aug-30 | EPI_ISL_16524596 | A/northern_gannet/Spain/22VIR12774-4_3104-40/2022 | Istituto Zooprofilattico Sperimentale delle Venezie, EU/OIE/Reference Laboratory and FAO Reference Centre for AI and ND | Istituto Zooprofilattico Sperimentale Delle Venezie | Ruano,M.J.; Rocha,A.; Sanchez,A.; Aguero,M.; Pastori, A.; Zecchin, B.; Fusaro, A.; Schivo, A.; Salviato, A.; Palumbo, E.; Giussani, E.; Monne, I.; Terregino, C. |
| EPI2293019 | MP | Spain | 2022-Aug-30 | EPI_ISL_16524596 | A/northern_gannet/Spain/22VIR12774-4_3104-40/2022 | Istituto Zooprofilattico Sperimentale delle Venezie, EU/OIE/Reference Laboratory and FAO Reference Centre for AI and ND | Istituto Zooprofilattico Sperimentale Delle Venezie | Ruano,M.J.; Rocha,A.; Sanchez,A.; Aguero,M.; Pastori, A.; Zecchin, B.; Fusaro, A.; Schivo, A.; Salviato, A.; Palumbo, E.; Giussani, E.; Monne, I.; Terregino, C. |
| EPI2296198 | NP | Belgium | 2022-Dec-08 | EPI_ISL_16547552 | A/Mustela_furo/Belgium/14312-0001/2022 | Sciensano - Animal Infectious Diseases | Sciensano, Department of Animal Infectious Diseases | Van Borm, Steven; Roupie, Virginie; Hostyn, Pierre; Lambrecht, Benedicte; Steensels, Mieke |
| EPI2296201 | MP | Belgium | 2022-Dec-08 | EPI_ISL_16547552 | A/Mustela_furo/Belgium/14312-0001/2022 | Sciensano - Animal Infectious Diseases | Sciensano, Department of Animal Infectious Diseases | Van Borm, Steven; Roupie, Virginie; Hostyn, Pierre; Lambrecht, Benedicte; Steensels, Mieke |

| | | | | | | | | |
|---|---|---|---|---|---|---|---|---|
| EPI2296203 | NS | Belgium | 2022-Dec-08 | EPI_ISL_16547552 | A/Mustela_furo/Belgium/14312-0001/2022 | Sciensano - Animal Infectious Diseases | Sciensano, Department of Animal Infectious Diseases | Van Borm, Steven; Roupie, Virginie; Hostyn, Pierre; Lambrecht, Benedicte; Steensels, Mieke |
| EPI2296212 | MP | Belgium | 2022-Dec-08 | EPI_ISL_16547666 | A/Mustela_furo/Belgium/14312-0002/2022 | Sciensano - Animal Infectious Diseases | Sciensano, Department of Animal Infectious Diseases | Van Borm, Steven; Roupie, Virginie; Hostyn, Pierre; Lambrecht, Benedicte; Steensels, Mieke |
| EPI2296213 | NS | Belgium | 2022-Dec-08 | EPI_ISL_16547666 | A/Mustela_furo/Belgium/14312-0002/2022 | Sciensano - Animal Infectious Diseases | Sciensano, Department of Animal Infectious Diseases | Van Borm, Steven; Roupie, Virginie; Hostyn, Pierre; Lambrecht, Benedicte; Steensels, Mieke |
| EPI2296210 | NP | Belgium | 2022-Dec-08 | EPI_ISL_16547666 | A/Mustela_furo/Belgium/14312-0002/2022 | Sciensano - Animal Infectious Diseases | Sciensano, Department of Animal Infectious Diseases | Van Borm, Steven; Roupie, Virginie; Hostyn, Pierre; Lambrecht, Benedicte; Steensels, Mieke |
| EPI2296227 | MP | Belgium | 2022-Dec-08 | EPI_ISL_16547851 | A/Gallus_gallus/Belgium/14313-001/2022 | Sciensano - Animal Infectious Diseases | Sciensano, Department of Animal Infectious Diseases | Van Borm, Steven; Roupie, Virginie; Hostyn, Pierre; Lambrecht, Benedicte; Steensels, Mieke |
| EPI2296228 | NS | Belgium | 2022-Dec-08 | EPI_ISL_16547851 | A/Gallus_gallus/Belgium/14313-001/2022 | Sciensano - Animal Infectious Diseases | Sciensano, Department of Animal Infectious Diseases | Van Borm, Steven; Roupie, Virginie; Hostyn, Pierre; Lambrecht, Benedicte; Steensels, Mieke |
| EPI2296225 | NP | Belgium | 2022-Dec-08 | EPI_ISL_16547851 | A/Gallus_gallus/Belgium/14313-001/2022 | Sciensano - Animal Infectious Diseases | Sciensano, Department of Animal Infectious Diseases | Van Borm, Steven; Roupie, Virginie; Hostyn, Pierre; Lambrecht, Benedicte; Steensels, Mieke |
| EPI2296239 | NS | Belgium | 2022-Nov-08 | EPI_ISL_16548347 | A/Pelecanus_conspicillatus/Belgium/12905-0001/2022 | Sciensano - Animal Infectious Diseases | Sciensano, Department of Animal Infectious Diseases | Van Borm, Steven; Roupie, Virginie; Hostyn, Pierre; Lambrecht, Benedicte; Steensels, Mieke |
| EPI2296236 | NP | Belgium | 2022-Nov-08 | EPI_ISL_16548347 | A/Pelecanus_conspicillatus/Belgium/12905-0001/2022 | Sciensano - Animal Infectious Diseases | Sciensano, Department of Animal Infectious Diseases | Van Borm, Steven; Roupie, Virginie; Hostyn, Pierre; Lambrecht, Benedicte; Steensels, Mieke |
| EPI2296238 | MP | Belgium | 2022-Nov-08 | EPI_ISL_16548347 | A/Pelecanus_conspicillatus/Belgium/12905-0001/2022 | Sciensano - Animal Infectious Diseases | Sciensano, Department of Animal Infectious Diseases | Van Borm, Steven; Roupie, Virginie; Hostyn, Pierre; Lambrecht, Benedicte; Steensels, Mieke |
| EPI2296244 | NP | Belgium | 2022-Nov-10 | EPI_ISL_16548356 | A/Phasianus_colchicus/Belgium/13027-0001/2022 | Sciensano - Animal Infectious Diseases | Sciensano, Department of Animal Infectious Diseases | Van Borm, Steven; Roupie, Virginie; Hostyn, Pierre; Lambrecht, Benedicte; Steensels, Mieke |
| EPI2296247 | NS | Belgium | 2022-Nov-10 | EPI_ISL_16548356 | A/Phasianus_colchicus/Belgium/13027-0001/2022 | Sciensano - Animal Infectious Diseases | Sciensano, Department of Animal Infectious Diseases | Van Borm, Steven; Roupie, Virginie; Hostyn, Pierre; Lambrecht, Benedicte; Steensels, Mieke |
| EPI2296246 | MP | Belgium | 2022-Nov-10 | EPI_ISL_16548356 | A/Phasianus_colchicus/Belgium/13027-0001/2022 | Sciensano - Animal Infectious Diseases | Sciensano, Department of Animal Infectious Diseases | Van Borm, Steven; Roupie, Virginie; Hostyn, Pierre; Lambrecht, Benedicte; Steensels, Mieke |
| EPI2296257 | NP | Belgium | 2022-Nov-09 | EPI_ISL_16548429 | A/Gallus_gallus/Belgium/13078-0001/2022 | Sciensano - Animal Infectious Diseases | Sciensano, Department of Animal Infectious Diseases | Van Borm, Steven; Roupie, Virginie; Hostyn, Pierre; Lambrecht, Benedicte; Steensels, Mieke |
| EPI2296259 | MP | Belgium | 2022-Nov-09 | EPI_ISL_16548429 | A/Gallus_gallus/Belgium/13078-0001/2022 | Sciensano - Animal Infectious Diseases | Sciensano, Department of Animal Infectious Diseases | Van Borm, Steven; Roupie, Virginie; Hostyn, Pierre; Lambrecht, Benedicte; Steensels, Mieke |
| EPI2296260 | NS | Belgium | 2022-Nov-09 | EPI_ISL_16548429 | A/Gallus_gallus/Belgium/13078-0001/2022 | Sciensano - Animal Infectious Diseases | Sciensano, Department of Animal Infectious Diseases | Van Borm, Steven; Roupie, Virginie; Hostyn, Pierre; Lambrecht, Benedicte; Steensels, Mieke |
| EPI2296309 | MP | Belgium | 2022-Nov-22 | EPI_ISL_16548600 | A/Gallus_gallus/Belgium/12485-0001/2022 | Sciensano - Animal Infectious Diseases | Sciensano, Department of Animal Infectious Diseases | Van Borm, Steven; Roupie, Virginie; Hostyn, Pierre; Lambrecht, Benedicte; Steensels, Mieke |
| EPI2296307 | NP | Belgium | 2022-Nov-22 | EPI_ISL_16548600 | A/Gallus_gallus/Belgium/12485-0001/2022 | Sciensano - Animal Infectious Diseases | Sciensano, Department of Animal Infectious Diseases | Van Borm, Steven; Roupie, Virginie; Hostyn, Pierre; Lambrecht, Benedicte; Steensels, Mieke |
| EPI2296310 | NS | Belgium | 2022-Nov-22 | EPI_ISL_16548600 | A/Gallus_gallus/Belgium/12485-0001/2022 | Sciensano - Animal Infectious Diseases | Sciensano, Department of Animal Infectious Diseases | Van Borm, Steven; Roupie, Virginie; Hostyn, Pierre; Lambrecht, Benedicte; Steensels, Mieke |
| EPI2296317 | MP | Belgium | 2022-Oct-01 | EPI_ISL_16548638 | A/Phasianus_colchicus/Belgium/11704-0003/2022 | Sciensano - Animal Infectious Diseases | Sciensano, Department of Animal Infectious Diseases | Van Borm, Steven; Roupie, Virginie; Hostyn, Pierre; Lambrecht, Benedicte; Steensels, Mieke |
| EPI2296318 | NS | Belgium | 2022-Oct-01 | EPI_ISL_16548638 | A/Phasianus_colchicus/Belgium/11704-0003/2022 | Sciensano - Animal Infectious Diseases | Sciensano, Department of Animal Infectious Diseases | Van Borm, Steven; Roupie, Virginie; Hostyn, Pierre; Lambrecht, Benedicte; Steensels, Mieke |
| EPI2296315 | NP | Belgium | 2022-Oct-01 | EPI_ISL_16548638 | A/Phasianus_colchicus/Belgium/11704-0003/2022 | Sciensano - Animal Infectious Diseases | Sciensano, Department of Animal Infectious Diseases | Van Borm, Steven; Roupie, Virginie; Hostyn, Pierre; Lambrecht, Benedicte; Steensels, Mieke |
| EPI2296774 | NP | Belgium | 2022-Nov-18 | EPI_ISL_16549157 | A/Gallus_gallus/Belgium/13357-0001/2022 | Sciensano - Animal Infectious Diseases | Sciensano, Department of Animal Infectious Diseases | Van Borm, Steven; Roupie, Virginie; Hostyn, Pierre; Lambrecht, Benedicte; Steensels, Mieke |
| EPI2296776 | MP | Belgium | 2022-Nov-18 | EPI_ISL_16549157 | A/Gallus_gallus/Belgium/13357-0001/2022 | Sciensano - Animal Infectious Diseases | Sciensano, Department of Animal Infectious Diseases | Van Borm, Steven; Roupie, Virginie; Hostyn, Pierre; Lambrecht, Benedicte; Steensels, Mieke |
| EPI2296777 | NS | Belgium | 2022-Nov-18 | EPI_ISL_16549157 | A/Gallus_gallus/Belgium/13357-0001/2022 | Sciensano - Animal Infectious Diseases | Sciensano, Department of Animal Infectious Diseases | Van Borm, Steven; Roupie, Virginie; Hostyn, Pierre; Lambrecht, Benedicte; Steensels, Mieke |
| EPI2296782 | NP | Belgium | 2022-Nov-21 | EPI_ISL_16549245 | A/Otus_scops/Belgium/13463-0002/2022 | Sciensano - Animal Infectious Diseases | Sciensano, Department of Animal Infectious Diseases | Van Borm, Steven; Roupie, Virginie; Hostyn, Pierre; Lambrecht, Benedicte; Steensels, Mieke |
| EPI2296785 | NS | Belgium | 2022-Nov-21 | EPI_ISL_16549245 | A/Otus_scops/Belgium/13463-0002/2022 | Sciensano - Animal Infectious Diseases | Sciensano, Department of Animal Infectious Diseases | Van Borm, Steven; Roupie, Virginie; Hostyn, Pierre; Lambrecht, Benedicte; Steensels, Mieke |
| EPI2296784 | MP | Belgium | 2022-Nov-21 | EPI_ISL_16549245 | A/Otus_scops/Belgium/13463-0002/2022 | Sciensano - Animal Infectious Diseases | Sciensano, Department of Animal Infectious Diseases | Van Borm, Steven; Roupie, Virginie; Hostyn, Pierre; Lambrecht, Benedicte; Steensels, Mieke |
| EPI2297244 | NS | Belgium | 2022-Nov-15 | EPI_ISL_16549349 | A/Phasianus_colchicus/Belgium/10993-0003/2022 | Sciensano - Animal Infectious Diseases | Sciensano, Department of Animal Infectious Diseases | Van Borm, Steven; Roupie, Virginie; Hostyn, Pierre; Lambrecht, Benedicte; Steensels, Mieke |
| EPI2297243 | MP | Belgium | 2022-Nov-15 | EPI_ISL_16549349 | A/Phasianus_colchicus/Belgium/10993-0003/2022 | Sciensano - Animal Infectious Diseases | Sciensano, Department of Animal Infectious Diseases | Van Borm, Steven; Roupie, Virginie; Hostyn, Pierre; Lambrecht, Benedicte; Steensels, Mieke |
| EPI2297238 | NP | Belgium | 2022-Nov-15 | EPI_ISL_16549349 | A/Phasianus_colchicus/Belgium/10993-0003/2022 | Sciensano - Animal Infectious Diseases | Sciensano, Department of Animal Infectious Diseases | Van Borm, Steven; Roupie, Virginie; Hostyn, Pierre; Lambrecht, Benedicte; Steensels, Mieke |
| EPI2297249 | NP | Belgium | 2022-Oct-17 | EPI_ISL_16549379 | A/Phasianus_colchicus/Belgium/12352-0002/2022 | Sciensano - Animal Infectious Diseases | Sciensano, Department of Animal Infectious Diseases | Van Borm, Steven; Roupie, Virginie; Hostyn, Pierre; Lambrecht, Benedicte; Steensels, Mieke |
| EPI2297252 | NS | Belgium | 2022-Oct-17 | EPI_ISL_16549379 | A/Phasianus_colchicus/Belgium/12352-0002/2022 | Sciensano - Animal Infectious Diseases | Sciensano, Department of Animal Infectious Diseases | Van Borm, Steven; Roupie, Virginie; Hostyn, Pierre; Lambrecht, Benedicte; Steensels, Mieke |
| EPI2297251 | MP | Belgium | 2022-Oct-17 | EPI_ISL_16549379 | A/Phasianus_colchicus/Belgium/12352-0002/2022 | Sciensano - Animal Infectious Diseases | Sciensano, Department of Animal Infectious Diseases | Van Borm, Steven; Roupie, Virginie; Hostyn, Pierre; Lambrecht, Benedicte; Steensels, Mieke |
| EPI2297257 | NP | Belgium | 2022-Dec-08 | EPI_ISL_16549380 | A/Vulpes_vulpes/Belgium/14775-0001/2022 | Sciensano - Animal Infectious Diseases | Sciensano, Department of Animal Infectious Diseases | Van Borm, Steven; Roupie, Virginie; Hostyn, Pierre; Lambrecht, Benedicte; Steensels, Mieke |

| | | | | | | | | |
|---|---|---|---|---|---|---|---|---|
| EPI2297259 | MP | Belgium | 2022-Dec-08 | EPI_ISL_16549380 | A/Vulpes_vulpes/Belgium/14775-0001/2022 | Sciensano - Animal Infectious Diseases | Sciensano, Department of Animal Infectious Diseases | Van Borm, Steven; Roupie, Virginie; Hostyn, Pierre; Lambrecht, Benedicte; Steensels, Mieke |
| EPI2297260 | NS | Belgium | 2022-Dec-08 | EPI_ISL_16549380 | A/Vulpes_vulpes/Belgium/14775-0001/2022 | Sciensano - Animal Infectious Diseases | Sciensano, Department of Animal Infectious Diseases | Van Borm, Steven; Roupie, Virginie; Hostyn, Pierre; Lambrecht, Benedicte; Steensels, Mieke |
| EPI2300421 | NP | United Kingdom | 2022-Dec-16 | EPI_ISL_16574465 | A/moorhen/Wales/171529/2022 | Animal and Plant Health Agency (APHA) | Animal and Plant Health Agency (APHA) | |
| EPI2300422 | NS | United Kingdom | 2022-Dec-16 | EPI_ISL_16574465 | A/moorhen/Wales/171529/2022 | Animal and Plant Health Agency (APHA) | Animal and Plant Health Agency (APHA) | |
| EPI2300423 | MP | United Kingdom | 2022-Dec-16 | EPI_ISL_16574465 | A/moorhen/Wales/171529/2022 | Animal and Plant Health Agency (APHA) | Animal and Plant Health Agency (APHA) | |
| EPI2300437 | NP | United Kingdom | 2022-Dec-20 | EPI_ISL_16574488 | A/chicken/England/171595/2022 | Animal and Plant Health Agency (APHA) | Animal and Plant Health Agency (APHA) | |
| EPI2300438 | NS | United Kingdom | 2022-Dec-20 | EPI_ISL_16574488 | A/chicken/England/171595/2022 | Animal and Plant Health Agency (APHA) | Animal and Plant Health Agency (APHA) | |
| EPI2300439 | MP | United Kingdom | 2022-Dec-20 | EPI_ISL_16574488 | A/chicken/England/171595/2022 | Animal and Plant Health Agency (APHA) | Animal and Plant Health Agency (APHA) | |
| EPI2300445 | NP | United Kingdom | 2022-Dec-21 | EPI_ISL_16574500 | A/domestic_goose/Scotland/171709/2022 | Animal and Plant Health Agency (APHA) | Animal and Plant Health Agency (APHA) | |
| EPI2300446 | NS | United Kingdom | 2022-Dec-21 | EPI_ISL_16574500 | A/domestic_goose/Scotland/171709/2022 | Animal and Plant Health Agency (APHA) | Animal and Plant Health Agency (APHA) | |
| EPI2300447 | MP | United Kingdom | 2022-Dec-21 | EPI_ISL_16574500 | A/domestic_goose/Scotland/171709/2022 | Animal and Plant Health Agency (APHA) | Animal and Plant Health Agency (APHA) | |
| EPI2300454 | NP | United Kingdom | 2022-Dec-23 | EPI_ISL_16574507 | A/chicken/England/172702/2022 | Animal and Plant Health Agency (APHA) | Animal and Plant Health Agency (APHA) | |
| EPI2300456 | NS | United Kingdom | 2022-Dec-23 | EPI_ISL_16574507 | A/chicken/England/172702/2022 | Animal and Plant Health Agency (APHA) | Animal and Plant Health Agency (APHA) | |
| EPI2300458 | MP | United Kingdom | 2022-Dec-23 | EPI_ISL_16574507 | A/chicken/England/172702/2022 | Animal and Plant Health Agency (APHA) | Animal and Plant Health Agency (APHA) | |
| EPI2300471 | NP | United Kingdom | 2022-Dec-23 | EPI_ISL_16574516 | A/chicken/England/172713/2022 | Animal and Plant Health Agency (APHA) | Animal and Plant Health Agency (APHA) | |
| EPI2300472 | NS | United Kingdom | 2022-Dec-23 | EPI_ISL_16574516 | A/chicken/England/172713/2022 | Animal and Plant Health Agency (APHA) | Animal and Plant Health Agency (APHA) | |
| EPI2300474 | MP | United Kingdom | 2022-Dec-23 | EPI_ISL_16574516 | A/chicken/England/172713/2022 | Animal and Plant Health Agency (APHA) | Animal and Plant Health Agency (APHA) | |
| EPI2300487 | NP | Isle of Man | 2022-Dec-26 | EPI_ISL_16574529 | A/chicken/Isle_of_Man/172945/2022 | Animal and Plant Health Agency (APHA) | Animal and Plant Health Agency (APHA) | |
| EPI2300488 | NS | Isle of Man | 2022-Dec-26 | EPI_ISL_16574529 | A/chicken/Isle_of_Man/172945/2022 | Animal and Plant Health Agency (APHA) | Animal and Plant Health Agency (APHA) | |
| EPI2300490 | MP | Isle of Man | 2022-Dec-26 | EPI_ISL_16574529 | A/chicken/Isle_of_Man/172945/2022 | Animal and Plant Health Agency (APHA) | Animal and Plant Health Agency (APHA) | |
| EPI2300501 | NP | United Kingdom | 2022-Dec-21 | EPI_ISL_16574544 | A/white-fronted_goose/Scotland/173468/2022 | Animal and Plant Health Agency (APHA) | Animal and Plant Health Agency (APHA) | |
| EPI2300503 | NS | United Kingdom | 2022-Dec-21 | EPI_ISL_16574544 | A/white-fronted_goose/Scotland/173468/2022 | Animal and Plant Health Agency (APHA) | Animal and Plant Health Agency (APHA) | |
| EPI2300505 | MP | United Kingdom | 2022-Dec-21 | EPI_ISL_16574544 | A/white-fronted_goose/Scotland/173468/2022 | Animal and Plant Health Agency (APHA) | Animal and Plant Health Agency (APHA) | |
| EPI2300517 | NP | United Kingdom | 2022-Dec-28 | EPI_ISL_16574556 | A/chicken/Scotland/173833/2022 | Animal and Plant Health Agency (APHA) | Animal and Plant Health Agency (APHA) | |
| EPI2300519 | NS | United Kingdom | 2022-Dec-28 | EPI_ISL_16574556 | A/chicken/Scotland/173833/2022 | Animal and Plant Health Agency (APHA) | Animal and Plant Health Agency (APHA) | |
| EPI2300521 | MP | United Kingdom | 2022-Dec-28 | EPI_ISL_16574556 | A/chicken/Scotland/173833/2022 | Animal and Plant Health Agency (APHA) | Animal and Plant Health Agency (APHA) | |
| EPI2300534 | NP | United Kingdom | 2022-Dec-29 | EPI_ISL_16574569 | A/chicken/England/175137/2022 | Animal and Plant Health Agency (APHA) | Animal and Plant Health Agency (APHA) | |
| EPI2300536 | NS | United Kingdom | 2022-Dec-29 | EPI_ISL_16574569 | A/chicken/England/175137/2022 | Animal and Plant Health Agency (APHA) | Animal and Plant Health Agency (APHA) | |
| EPI2300537 | MP | United Kingdom | 2022-Dec-29 | EPI_ISL_16574569 | A/chicken/England/175137/2022 | Animal and Plant Health Agency (APHA) | Animal and Plant Health Agency (APHA) | |
| EPI2300550 | NP | United Kingdom | 2022-Dec-12 | EPI_ISL_16574586 | A/common_buzzard/England/255858/2022 | Animal and Plant Health Agency (APHA) | Animal and Plant Health Agency (APHA) | |
| EPI2300551 | NS | United Kingdom | 2022-Dec-12 | EPI_ISL_16574586 | A/common_buzzard/England/255858/2022 | Animal and Plant Health Agency (APHA) | Animal and Plant Health Agency (APHA) | |
| EPI2300552 | MP | United Kingdom | 2022-Dec-12 | EPI_ISL_16574586 | A/common_buzzard/England/255858/2022 | Animal and Plant Health Agency (APHA) | Animal and Plant Health Agency (APHA) | |
| EPI2300565 | NP | United Kingdom | 2022-Dec-07 | EPI_ISL_16574599 | A/mute_swan/England/119983/2022 | Animal and Plant Health Agency (APHA) | Animal and Plant Health Agency (APHA) | |
| EPI2300566 | NS | United Kingdom | 2022-Dec-07 | EPI_ISL_16574599 | A/mute_swan/England/119983/2022 | Animal and Plant Health Agency (APHA) | Animal and Plant Health Agency (APHA) | |
| EPI2300567 | MP | United Kingdom | 2022-Dec-07 | EPI_ISL_16574599 | A/mute_swan/England/119983/2022 | Animal and Plant Health Agency (APHA) | Animal and Plant Health Agency (APHA) | |

| | | | | | | | | |
|---|---|---|---|---|---|---|---|---|
| EPI2300581 | NP | United Kingdom | 2022-Dec-07 | EPI_ISL_16574612 | A/black-headed_gull/England/120011/2022 | Animal and Plant Health Agency (APHA) | Animal and Plant Health Agency (APHA) | |
| EPI2300582 | NS | United Kingdom | 2022-Dec-07 | EPI_ISL_16574612 | A/black-headed_gull/England/120011/2022 | Animal and Plant Health Agency (APHA) | Animal and Plant Health Agency (APHA) | |
| EPI2300583 | MP | United Kingdom | 2022-Dec-07 | EPI_ISL_16574612 | A/black-headed_gull/England/120011/2022 | Animal and Plant Health Agency (APHA) | Animal and Plant Health Agency (APHA) | |
| EPI2300613 | NP | United Kingdom | 2022-Dec-15 | EPI_ISL_16574657 | A/greylag_goose/England/120308/2022 | Animal and Plant Health Agency (APHA) | Animal and Plant Health Agency (APHA) | |
| EPI2300614 | NS | United Kingdom | 2022-Dec-15 | EPI_ISL_16574657 | A/greylag_goose/England/120308/2022 | Animal and Plant Health Agency (APHA) | Animal and Plant Health Agency (APHA) | |
| EPI2300616 | MP | United Kingdom | 2022-Dec-15 | EPI_ISL_16574657 | A/greylag_goose/England/120308/2022 | Animal and Plant Health Agency (APHA) | Animal and Plant Health Agency (APHA) | |
| EPI2300644 | NP | United Kingdom | 2022-Dec-21 | EPI_ISL_16574682 | A/sparrowhawk/England/120507/2022 | Animal and Plant Health Agency (APHA) | Animal and Plant Health Agency (APHA) | |
| EPI2300646 | NS | United Kingdom | 2022-Dec-21 | EPI_ISL_16574682 | A/sparrowhawk/England/120507/2022 | Animal and Plant Health Agency (APHA) | Animal and Plant Health Agency (APHA) | |
| EPI2300648 | MP | United Kingdom | 2022-Dec-21 | EPI_ISL_16574682 | A/sparrowhawk/England/120507/2022 | Animal and Plant Health Agency (APHA) | Animal and Plant Health Agency (APHA) | |
| EPI2300677 | NP | United Kingdom | 2022-Dec-10 | EPI_ISL_16574697 | A/canada_goose/England/396207/2022 | Animal and Plant Health Agency (APHA) | Animal and Plant Health Agency (APHA) | |
| EPI2300679 | NS | United Kingdom | 2022-Dec-10 | EPI_ISL_16574697 | A/canada_goose/England/396207/2022 | Animal and Plant Health Agency (APHA) | Animal and Plant Health Agency (APHA) | |
| EPI2300680 | MP | United Kingdom | 2022-Dec-10 | EPI_ISL_16574697 | A/canada_goose/England/396207/2022 | Animal and Plant Health Agency (APHA) | Animal and Plant Health Agency (APHA) | |
| EPI2300709 | NP | United Kingdom | 2022-Dec-10 | EPI_ISL_16574723 | A/mute_swan/England/396216/2022 | Animal and Plant Health Agency (APHA) | Animal and Plant Health Agency (APHA) | |
| EPI2300711 | NS | United Kingdom | 2022-Dec-10 | EPI_ISL_16574723 | A/mute_swan/England/396216/2022 | Animal and Plant Health Agency (APHA) | Animal and Plant Health Agency (APHA) | |
| EPI2300712 | MP | United Kingdom | 2022-Dec-10 | EPI_ISL_16574723 | A/mute_swan/England/396216/2022 | Animal and Plant Health Agency (APHA) | Animal and Plant Health Agency (APHA) | |
| EPI2300723 | NP | United Kingdom | 2022-Dec-17 | EPI_ISL_16574736 | A/goose/England/396398/2022 | Animal and Plant Health Agency (APHA) | Animal and Plant Health Agency (APHA) | |
| EPI2300725 | NS | United Kingdom | 2022-Dec-17 | EPI_ISL_16574736 | A/goose/England/396398/2022 | Animal and Plant Health Agency (APHA) | Animal and Plant Health Agency (APHA) | |
| EPI2300727 | MP | United Kingdom | 2022-Dec-17 | EPI_ISL_16574736 | A/goose/England/396398/2022 | Animal and Plant Health Agency (APHA) | Animal and Plant Health Agency (APHA) | |
| EPI2300772 | NP | United Kingdom | 2022-Dec-06 | EPI_ISL_16574769 | A/greylag_goose/Scotland/168361/2022 | Animal and Plant Health Agency (APHA) | Animal and Plant Health Agency (APHA) | |
| EPI2300774 | NS | United Kingdom | 2022-Dec-06 | EPI_ISL_16574769 | A/greylag_goose/Scotland/168361/2022 | Animal and Plant Health Agency (APHA) | Animal and Plant Health Agency (APHA) | |
| EPI2300776 | MP | United Kingdom | 2022-Dec-06 | EPI_ISL_16574769 | A/greylag_goose/Scotland/168361/2022 | Animal and Plant Health Agency (APHA) | Animal and Plant Health Agency (APHA) | |
| EPI2314306 | MP | Belgium | 2022-Dec-20 | EPI_ISL_16676474 | A/Gallus_gallus/Belgium/14733-0001/2022 | Sciensano - Animal Infectious Diseases | Sciensano, Department of Animal Infectious Diseases | Van Borm, Steven; Roupie, Virginie; Hostyn, Pierre; Lambrecht, Benedicte; Steensels, Mieke |
| EPI2314307 | NS | Belgium | 2022-Dec-20 | EPI_ISL_16676474 | A/Gallus_gallus/Belgium/14733-0001/2022 | Sciensano - Animal Infectious Diseases | Sciensano, Department of Animal Infectious Diseases | Van Borm, Steven; Roupie, Virginie; Hostyn, Pierre; Lambrecht, Benedicte; Steensels, Mieke |
| EPI2314304 | NP | Belgium | 2022-Dec-20 | EPI_ISL_16676474 | A/Gallus_gallus/Belgium/14733-0001/2022 | Sciensano - Animal Infectious Diseases | Sciensano, Department of Animal Infectious Diseases | Van Borm, Steven; Roupie, Virginie; Hostyn, Pierre; Lambrecht, Benedicte; Steensels, Mieke |
| EPI2314329 | NS | Belgium | 2022-Jan-17 | EPI_ISL_16676477 | A/Anser_anser/Belgium/00939-0011/2022 | Sciensano - Animal Infectious Diseases | Sciensano, Department of Animal Infectious Diseases | Van Borm, Steven; Roupie, Virginie; Hostyn, Pierre; Lambrecht, Benedicte; Steensels, Mieke |
| EPI2314328 | MP | Belgium | 2022-Jan-17 | EPI_ISL_16676477 | A/Anser_anser/Belgium/00939-0011/2022 | Sciensano - Animal Infectious Diseases | Sciensano, Department of Animal Infectious Diseases | Van Borm, Steven; Roupie, Virginie; Hostyn, Pierre; Lambrecht, Benedicte; Steensels, Mieke |
| EPI2314326 | NP | Belgium | 2022-Jan-17 | EPI_ISL_16676477 | A/Anser_anser/Belgium/00939-0011/2022 | Sciensano - Animal Infectious Diseases | Sciensano, Department of Animal Infectious Diseases | Van Borm, Steven; Roupie, Virginie; Hostyn, Pierre; Lambrecht, Benedicte; Steensels, Mieke |
| EPI2314342 | MP | Belgium | 2022-Jun-16 | EPI_ISL_16676481 | A/Sterna_hirundo/Belgium/08045-0004/2022 | Sciensano - Animal Infectious Diseases | Sciensano, Department of Animal Infectious Diseases | Van Borm, Steven; Roupie, Virginie; Hostyn, Pierre; Lambrecht, Benedicte; Steensels, Mieke |
| EPI2314343 | NS | Belgium | 2022-Jun-16 | EPI_ISL_16676481 | A/Sterna_hirundo/Belgium/08045-0004/2022 | Sciensano - Animal Infectious Diseases | Sciensano, Department of Animal Infectious Diseases | Van Borm, Steven; Roupie, Virginie; Hostyn, Pierre; Lambrecht, Benedicte; Steensels, Mieke |
| EPI2314338 | NP | Belgium | 2022-Jun-16 | EPI_ISL_16676481 | A/Sterna_hirundo/Belgium/08045-0004/2022 | Sciensano - Animal Infectious Diseases | Sciensano, Department of Animal Infectious Diseases | Van Borm, Steven; Roupie, Virginie; Hostyn, Pierre; Lambrecht, Benedicte; Steensels, Mieke |
| EPI2314358 | NP | Belgium | 2022-Jul-26 | EPI_ISL_16676486 | A/Falco_peregrinus/Belgium/09358-0005/2022 | Sciensano - Animal Infectious Diseases | Sciensano, Department of Animal Infectious Diseases | Van Borm, Steven; Roupie, Virginie; Hostyn, Pierre; Lambrecht, Benedicte; Steensels, Mieke |
| EPI2314360 | NS | Belgium | 2022-Jul-26 | EPI_ISL_16676486 | A/Falco_peregrinus/Belgium/09358-0005/2022 | Sciensano - Animal Infectious Diseases | Sciensano, Department of Animal Infectious Diseases | Van Borm, Steven; Roupie, Virginie; Hostyn, Pierre; Lambrecht, Benedicte; Steensels, Mieke |
| EPI2314359 | MP | Belgium | 2022-Jul-26 | EPI_ISL_16676486 | A/Falco_peregrinus/Belgium/09358-0005/2022 | Sciensano - Animal Infectious Diseases | Sciensano, Department of Animal Infectious Diseases | Van Borm, Steven; Roupie, Virginie; Hostyn, Pierre; Lambrecht, Benedicte; Steensels, Mieke |
| EPI2314368 | NS | Belgium | 2022-Sep-16 | EPI_ISL_16676487 | A/Cygnus_olor/Belgium/10811-0003/2022 | Sciensano - Animal Infectious Diseases | Sciensano, Department of Animal Infectious Diseases | Van Borm, Steven; Roupie, Virginie; Hostyn, Pierre; Lambrecht, Benedicte; Steensels, Mieke |
| EPI2314367 | MP | Belgium | 2022-Sep-16 | EPI_ISL_16676487 | A/Cygnus_olor/Belgium/10811-0003/2022 | Sciensano - Animal Infectious Diseases | Sciensano, Department of Animal Infectious Diseases | Van Borm, Steven; Roupie, Virginie; Hostyn, Pierre; Lambrecht, Benedicte; Steensels, Mieke |

| | | | | | | | | |
|---|---|---|---|---|---|---|---|---|
| EPI2314365 | NP | Belgium | 2022-Sep-16 | EPI_ISL_16676487 | A/Cygnus_olor/Belgium/10811-0003/2022 | Sciensano - Animal Infectious Diseases | Sciensano, Department of Animal Infectious Diseases | Van Borm, Steven; Roupie, Virginie; Hostyn, Pierre; Lambrecht, Benedicte; Steensels, Mieke |
| EPI2314383 | MP | Belgium | 2022-Dec-21 | EPI_ISL_16676489 | A/Anas_platyrhynchos_domesticus/Belgium/14735-0001/2022 | Sciensano - Animal Infectious Diseases | Sciensano, Department of Animal Infectious Diseases | Van Borm, Steven; Roupie, Virginie; Hostyn, Pierre; Lambrecht, Benedicte; Steensels, Mieke |
| EPI2314384 | NS | Belgium | 2022-Dec-21 | EPI_ISL_16676489 | A/Anas_platyrhynchos_domesticus/Belgium/14735-0001/2022 | Sciensano - Animal Infectious Diseases | Sciensano, Department of Animal Infectious Diseases | Van Borm, Steven; Roupie, Virginie; Hostyn, Pierre; Lambrecht, Benedicte; Steensels, Mieke |
| EPI2314381 | NP | Belgium | 2022-Dec-21 | EPI_ISL_16676489 | A/Anas_platyrhynchos_domesticus/Belgium/14735-0001/2022 | Sciensano - Animal Infectious Diseases | Sciensano, Department of Animal Infectious Diseases | Van Borm, Steven; Roupie, Virginie; Hostyn, Pierre; Lambrecht, Benedicte; Steensels, Mieke |
| EPI2325487 | NS | France | 2022-Dec-10 | EPI_ISL_16740903 | A/duck/France/22P025647/2022 | Anses (Ploufragan-Plouzané) | ANSES Agence Nationale De Securite Sanitaire De L'alimentation | |
| EPI2325484 | NP | France | 2022-Dec-10 | EPI_ISL_16740903 | A/duck/France/22P025647/2022 | Anses (Ploufragan-Plouzané) | ANSES Agence Nationale De Securite Sanitaire De L'alimentation | |
| EPI2325486 | MP | France | 2022-Dec-10 | EPI_ISL_16740903 | A/duck/France/22P025647/2022 | Anses (Ploufragan-Plouzané) | ANSES Agence Nationale De Securite Sanitaire De L'alimentation | |
| EPI2325492 | NP | France | 2022-Dec-01 | EPI_ISL_16740904 | A/duck/France/22P024591/2022 | Anses (Ploufragan-Plouzané) | ANSES Agence Nationale De Securite Sanitaire De L'alimentation | |
| EPI2325494 | MP | France | 2022-Dec-01 | EPI_ISL_16740904 | A/duck/France/22P024591/2022 | Anses (Ploufragan-Plouzané) | ANSES Agence Nationale De Securite Sanitaire De L'alimentation | |
| EPI2325495 | NS | France | 2022-Dec-01 | EPI_ISL_16740904 | A/duck/France/22P024591/2022 | Anses (Ploufragan-Plouzané) | ANSES Agence Nationale De Securite Sanitaire De L'alimentation | |
| EPI2325503 | NS | France | 2022-Dec-01 | EPI_ISL_16740905 | A/duck/France/22P024657/2022 | Anses (Ploufragan-Plouzané) | ANSES Agence Nationale De Securite Sanitaire De L'alimentation | |
| EPI2325500 | NP | France | 2022-Dec-01 | EPI_ISL_16740905 | A/duck/France/22P024657/2022 | Anses (Ploufragan-Plouzané) | ANSES Agence Nationale De Securite Sanitaire De L'alimentation | |
| EPI2325502 | MP | France | 2022-Dec-01 | EPI_ISL_16740905 | A/duck/France/22P024657/2022 | Anses (Ploufragan-Plouzané) | ANSES Agence Nationale De Securite Sanitaire De L'alimentation | |
| EPI2325511 | NS | France | 2022-Dec-03 | EPI_ISL_16741031 | A/duck/France/22P024701/2022 | Anses (Ploufragan-Plouzané) | ANSES Agence Nationale De Securite Sanitaire De L'alimentation | |
| EPI2325508 | NP | France | 2022-Dec-03 | EPI_ISL_16741031 | A/duck/France/22P024701/2022 | Anses (Ploufragan-Plouzané) | ANSES Agence Nationale De Securite Sanitaire De L'alimentation | |
| EPI2325510 | MP | France | 2022-Dec-03 | EPI_ISL_16741031 | A/duck/France/22P024701/2022 | Anses (Ploufragan-Plouzané) | ANSES Agence Nationale De Securite Sanitaire De L'alimentation | |
| EPI2325516 | NP | France | 2022-Dec-05 | EPI_ISL_16741115 | A/turkey/France/22P024731/2022 | Anses (Ploufragan-Plouzané) | ANSES Agence Nationale De Securite Sanitaire De L'alimentation | |
| EPI2325518 | MP | France | 2022-Dec-05 | EPI_ISL_16741115 | A/turkey/France/22P024731/2022 | Anses (Ploufragan-Plouzané) | ANSES Agence Nationale De Securite Sanitaire De L'alimentation | |
| EPI2325519 | NS | France | 2022-Dec-05 | EPI_ISL_16741115 | A/turkey/France/22P024731/2022 | Anses (Ploufragan-Plouzané) | ANSES Agence Nationale De Securite Sanitaire De L'alimentation | |
| EPI2325604 | NP | France | 2022-Dec-06 | EPI_ISL_16741201 | A/duck/France/22P025078/2022 | Anses (Ploufragan-Plouzané) | ANSES Agence Nationale De Securite Sanitaire De L'alimentation | |
| EPI2325641 | MP | France | 2022-Dec-06 | EPI_ISL_16741201 | A/duck/France/22P025078/2022 | Anses (Ploufragan-Plouzané) | ANSES Agence Nationale De Securite Sanitaire De L'alimentation | |
| EPI2325655 | NS | France | 2022-Dec-06 | EPI_ISL_16741201 | A/duck/France/22P025078/2022 | Anses (Ploufragan-Plouzané) | ANSES Agence Nationale De Securite Sanitaire De L'alimentation | |
| EPI2325708 | MP | France | 2022-Dec-07 | EPI_ISL_16741221 | A/duck/France/22P025252/2022 | Anses (Ploufragan-Plouzané) | ANSES Agence Nationale De Securite Sanitaire De L'alimentation | |
| EPI2325709 | NS | France | 2022-Dec-07 | EPI_ISL_16741221 | A/duck/France/22P025252/2022 | Anses (Ploufragan-Plouzané) | ANSES Agence Nationale De Securite Sanitaire De L'alimentation | |
| EPI2325706 | NP | France | 2022-Dec-07 | EPI_ISL_16741221 | A/duck/France/22P025252/2022 | Anses (Ploufragan-Plouzané) | ANSES Agence Nationale De Securite Sanitaire De L'alimentation | |
| EPI2325718 | NS | France | 2022-Dec-05 | EPI_ISL_16741325 | A/duck/France/22P025303/2022 | Anses (Ploufragan-Plouzané) | ANSES Agence Nationale De Securite Sanitaire De L'alimentation | |
| EPI2325717 | MP | France | 2022-Dec-05 | EPI_ISL_16741325 | A/duck/France/22P025303/2022 | Anses (Ploufragan-Plouzané) | ANSES Agence Nationale De Securite Sanitaire De L'alimentation | |
| EPI2325715 | NP | France | 2022-Dec-05 | EPI_ISL_16741325 | A/duck/France/22P025303/2022 | Anses (Ploufragan-Plouzané) | ANSES Agence Nationale De Securite Sanitaire De L'alimentation | |
| EPI2325725 | MP | France | 2022-Dec-09 | EPI_ISL_16741326 | A/chicken/France/22P025486/2022 | Anses (Ploufragan-Plouzané) | ANSES Agence Nationale De Securite Sanitaire De L'alimentation | |
| EPI2325726 | NS | France | 2022-Dec-09 | EPI_ISL_16741326 | A/chicken/France/22P025486/2022 | Anses (Ploufragan-Plouzané) | ANSES Agence Nationale De Securite Sanitaire De L'alimentation | |
| EPI2325723 | NP | France | 2022-Dec-09 | EPI_ISL_16741326 | A/chicken/France/22P025486/2022 | Anses (Ploufragan-Plouzané) | ANSES Agence Nationale De Securite Sanitaire De L'alimentation | |
| EPI2325827 | NP | France | 2022-Dec-10 | EPI_ISL_16741402 | A/duck/France/22P025498/2022 | Anses (Ploufragan-Plouzané) | ANSES Agence Nationale De Securite Sanitaire De L'alimentation | |
| EPI2325830 | NS | France | 2022-Dec-10 | EPI_ISL_16741402 | A/duck/France/22P025498/2022 | Anses (Ploufragan-Plouzané) | ANSES Agence Nationale De Securite Sanitaire De L'alimentation | |
| EPI2325829 | MP | France | 2022-Dec-10 | EPI_ISL_16741402 | A/duck/France/22P025498/2022 | Anses (Ploufragan-Plouzané) | ANSES Agence Nationale De Securite Sanitaire De L'alimentation | |
| EPI2332837 | MP | Belgium | 2022-Jan-03 | EPI_ISL_16747602 | A/Buteo_buteo/Belgium/00331-0007/2022 | Sciensano - Animal Infectious Diseases | Sciensano, Department of Animal Infectious Diseases | Van Borm, Steven; Roupie, Virginie; Hostyn, Pierre; Lambrecht, Benedicte; Steensels, Mieke |

| | | | | | | | | |
|---|---|---|---|---|---|---|---|---|
| EPI2332835 | NP | Belgium | 2022-Jan-03 | EPI_ISL_16747602 | A/Buteo_buteo/Belgium/00331-0007/2022 | Sciensano - Animal Infectious Diseases | Sciensano, Department of Animal Infectious Diseases | Van Borm, Steven; Roupie, Virginie; Hostyn, Pierre; Lambrecht, Benedicte; Steensels, Mieke |
| EPI2332839 | NS | Belgium | 2022-Jan-03 | EPI_ISL_16747602 | A/Buteo_buteo/Belgium/00331-0007/2022 | Sciensano - Animal Infectious Diseases | Sciensano, Department of Animal Infectious Diseases | Van Borm, Steven; Roupie, Virginie; Hostyn, Pierre; Lambrecht, Benedicte; Steensels, Mieke |
| EPI2334786 | MP | Belgium | 2022-Aug-18 | EPI_ISL_16748778 | A/Aythya_fuligula/Belgium/10064-0004/2022 | Sciensano - Animal Infectious Diseases | Sciensano, Department of Animal Infectious Diseases | Van Borm, Steven; Roupie, Virginie; Hostyn, Pierre; Lambrecht, Benedicte; Steensels, Mieke |
| EPI2334841 | NS | Belgium | 2022-Aug-18 | EPI_ISL_16748778 | A/Aythya_fuligula/Belgium/10064-0004/2022 | Sciensano - Animal Infectious Diseases | Sciensano, Department of Animal Infectious Diseases | Van Borm, Steven; Roupie, Virginie; Hostyn, Pierre; Lambrecht, Benedicte; Steensels, Mieke |
| EPI2334749 | NP | Belgium | 2022-Aug-18 | EPI_ISL_16748778 | A/Aythya_fuligula/Belgium/10064-0004/2022 | Sciensano - Animal Infectious Diseases | Sciensano, Department of Animal Infectious Diseases | Van Borm, Steven; Roupie, Virginie; Hostyn, Pierre; Lambrecht, Benedicte; Steensels, Mieke |
| EPI2335413 | MP | Belgium | 2022-Oct-10 | EPI_ISL_16748957 | A/Corvus_monedula/Belgium/11918_0006/2022 | Sciensano, Department of Animal Infectious Diseases | Sciensano, Department of Animal Infectious Diseases | Van Borm, Steven; Roupie, Virginie; Hostyn, Pierre; Lambrecht, Benedicte; Steensels, Mieke |
| EPI2335411 | NP | Belgium | 2022-Oct-10 | EPI_ISL_16748957 | A/Corvus_monedula/Belgium/11918_0006/2022 | Sciensano, Department of Animal Infectious Diseases | Sciensano, Department of Animal Infectious Diseases | Van Borm, Steven; Roupie, Virginie; Hostyn, Pierre; Lambrecht, Benedicte; Steensels, Mieke |
| EPI2335414 | NS | Belgium | 2022-Oct-10 | EPI_ISL_16748957 | A/Corvus_monedula/Belgium/11918_0006/2022 | Sciensano, Department of Animal Infectious Diseases | Sciensano, Department of Animal Infectious Diseases | Van Borm, Steven; Roupie, Virginie; Hostyn, Pierre; Lambrecht, Benedicte; Steensels, Mieke |
| EPI2351044 | NP | Belgium | 2022-Nov-02 | EPI_ISL_16809999 | A/Anser anser_domesticus/Belgium/12994_0002/2022 | Sciensano - Animal Infectious Diseases | Sciensano, Department of Animal Infectious Diseases | Van Borm, Steven; Roupie, Virginie; Hostyn, Pierre; Lambrecht, Benedicte; Steensels, Mieke |
| EPI2351047 | NS | Belgium | 2022-Nov-02 | EPI_ISL_16809999 | A/Anser anser_domesticus/Belgium/12994_0002/2022 | Sciensano - Animal Infectious Diseases | Sciensano, Department of Animal Infectious Diseases | Van Borm, Steven; Roupie, Virginie; Hostyn, Pierre; Lambrecht, Benedicte; Steensels, Mieke |
| EPI2351046 | MP | Belgium | 2022-Nov-02 | EPI_ISL_16809999 | A/Anser anser_domesticus/Belgium/12994_0002/2022 | Sciensano - Animal Infectious Diseases | Sciensano, Department of Animal Infectious Diseases | Van Borm, Steven; Roupie, Virginie; Hostyn, Pierre; Lambrecht, Benedicte; Steensels, Mieke |
| EPI2351423 | NP | Belgium | 2022-Sep-10 | EPI_ISL_16810069 | A/Morus_bassanus/Belgium/10811_0001/2022 | Sciensano - Animal Infectious Diseases | Sciensano, Department of Animal Infectious Diseases | Van Borm, Steven; Roupie, Virginie; Hostyn, Pierre; Lambrecht, Benedicte; Steensels, Mieke |
| EPI2351484 | NS | Belgium | 2022-Sep-10 | EPI_ISL_16810069 | A/Morus_bassanus/Belgium/10811_0001/2022 | Sciensano - Animal Infectious Diseases | Sciensano, Department of Animal Infectious Diseases | Van Borm, Steven; Roupie, Virginie; Hostyn, Pierre; Lambrecht, Benedicte; Steensels, Mieke |
| EPI2351461 | MP | Belgium | 2022-Sep-10 | EPI_ISL_16810069 | A/Morus_bassanus/Belgium/10811_0001/2022 | Sciensano - Animal Infectious Diseases | Sciensano, Department of Animal Infectious Diseases | Van Borm, Steven; Roupie, Virginie; Hostyn, Pierre; Lambrecht, Benedicte; Steensels, Mieke |
| EPI2351623 | MP | Belgium | 2022-Sep-07 | EPI_ISL_16810105 | A/Larus_argentatus/Belgium/10811_0002/2022 | Sciensano - Animal Infectious Diseases | Sciensano, Department of Animal Infectious Diseases | Van Borm, Steven; Roupie, Virginie; Hostyn, Pierre; Lambrecht, Benedicte; Steensels, Mieke |
| EPI2351620 | NP | Belgium | 2022-Sep-07 | EPI_ISL_16810105 | A/Larus_argentatus/Belgium/10811_0002/2022 | Sciensano - Animal Infectious Diseases | Sciensano, Department of Animal Infectious Diseases | Van Borm, Steven; Roupie, Virginie; Hostyn, Pierre; Lambrecht, Benedicte; Steensels, Mieke |
| EPI2351624 | NS | Belgium | 2022-Sep-07 | EPI_ISL_16810105 | A/Larus_argentatus/Belgium/10811_0002/2022 | Sciensano - Animal Infectious Diseases | Sciensano, Department of Animal Infectious Diseases | Van Borm, Steven; Roupie, Virginie; Hostyn, Pierre; Lambrecht, Benedicte; Steensels, Mieke |
| EPI2351909 | NP | Belgium | 2022-Aug-30 | EPI_ISL_16810562 | A/Mixed_wild_birds/Belgium/10368_0001/2022 | Sciensano - Animal Infectious Diseases | Sciensano, Department of Animal Infectious Diseases | Van Borm, Steven; Roupie, Virginie; Hostyn, Pierre; Lambrecht, Benedicte; Steensels, Mieke |
| EPI2351911 | MP | Belgium | 2022-Aug-30 | EPI_ISL_16810562 | A/Mixed_wild_birds/Belgium/10368_0001/2022 | Sciensano - Animal Infectious Diseases | Sciensano, Department of Animal Infectious Diseases | Van Borm, Steven; Roupie, Virginie; Hostyn, Pierre; Lambrecht, Benedicte; Steensels, Mieke |
| EPI2351912 | NS | Belgium | 2022-Aug-30 | EPI_ISL_16810562 | A/Mixed_wild_birds/Belgium/10368_0001/2022 | Sciensano - Animal Infectious Diseases | Sciensano, Department of Animal Infectious Diseases | Van Borm, Steven; Roupie, Virginie; Hostyn, Pierre; Lambrecht, Benedicte; Steensels, Mieke |
| EPI2351919 | MP | Belgium | 2022-Jul-18 | EPI_ISL_16810698 | A/Bubo_bubo/Belgium/09467_0001/2022 | Sciensano - Animal Infectious Diseases | Sciensano, Department of Animal Infectious Diseases | Van Borm, Steven; Roupie, Virginie; Hostyn, Pierre; Lambrecht, Benedicte; Steensels, Mieke |
| EPI2351920 | NS | Belgium | 2022-Jul-18 | EPI_ISL_16810698 | A/Bubo_bubo/Belgium/09467_0001/2022 | Sciensano - Animal Infectious Diseases | Sciensano, Department of Animal Infectious Diseases | Van Borm, Steven; Roupie, Virginie; Hostyn, Pierre; Lambrecht, Benedicte; Steensels, Mieke |
| EPI2351917 | NP | Belgium | 2022-Jul-18 | EPI_ISL_16810698 | A/Bubo_bubo/Belgium/09467_0001/2022 | Sciensano - Animal Infectious Diseases | Sciensano, Department of Animal Infectious Diseases | Van Borm, Steven; Roupie, Virginie; Hostyn, Pierre; Lambrecht, Benedicte; Steensels, Mieke |
| EPI2352317 | NP | Netherlands | 2022-Dec-31 | EPI_ISL_16811039 | A/Peregrine falcon/Netherlands/1/2022 | Erasmus Medical Center | Erasmus Medical Center | |
| EPI2352318 | NS | Netherlands | 2022-Dec-31 | EPI_ISL_16811039 | A/Peregrine falcon/Netherlands/1/2022 | Erasmus Medical Center | Erasmus Medical Center | |
| EPI2352319 | MP | Netherlands | 2022-Dec-31 | EPI_ISL_16811039 | A/Peregrine falcon/Netherlands/1/2022 | Erasmus Medical Center | Erasmus Medical Center | |
| EPI2352651 | NP | United Kingdom | 2022-Dec-30 | EPI_ISL_16811199 | A/Mute_Swan/England/120693/2022 | Animal and Plant Health Agency (APHA) | Animal and Plant Health Agency (APHA) | |
| EPI2352653 | NS | United Kingdom | 2022-Dec-30 | EPI_ISL_16811199 | A/Mute_Swan/England/120693/2022 | Animal and Plant Health Agency (APHA) | Animal and Plant Health Agency (APHA) | |
| EPI2352655 | MP | United Kingdom | 2022-Dec-30 | EPI_ISL_16811199 | A/Mute_Swan/England/120693/2022 | Animal and Plant Health Agency (APHA) | Animal and Plant Health Agency (APHA) | |
| EPI2352667 | NP | United Kingdom | 2022-Dec-30 | EPI_ISL_16811201 | A/Greylag_Goose/England/120696/2022 | Animal and Plant Health Agency (APHA) | Animal and Plant Health Agency (APHA) | |
| EPI2352669 | NS | United Kingdom | 2022-Dec-30 | EPI_ISL_16811201 | A/Greylag_Goose/England/120696/2022 | Animal and Plant Health Agency (APHA) | Animal and Plant Health Agency (APHA) | |
| EPI2352671 | MP | United Kingdom | 2022-Dec-30 | EPI_ISL_16811201 | A/Greylag_Goose/England/120696/2022 | Animal and Plant Health Agency (APHA) | Animal and Plant Health Agency (APHA) | |
| EPI2352962 | NP | Spain | 2022-Oct-13 | EPI_ISL_16813290 | A/CastillaLaMancha/3869/2022 | Instituto de Salud Carlos III | Instituto de Salud Carlos III | Pozo,F; Iglesias-Caballero, M.; Molinero,M.; Camarero,S.; Mart?n,V.; Reyes,N.; V?zquez-Mor?n,S.; Casas,I. |
| EPI2352963 | NS | Spain | 2022-Oct-13 | EPI_ISL_16813290 | A/CastillaLaMancha/3869/2022 | Instituto de Salud Carlos III | Instituto de Salud Carlos III | Pozo,F; Iglesias-Caballero, M.; Molinero,M.; Camarero,S.; Mart?n,V.; Reyes,N.; V?zquez-Mor?n,S.; Casas,I. |
| EPI2352964 | MP | Spain | 2022-Oct-13 | EPI_ISL_16813290 | A/CastillaLaMancha/3869/2022 | Instituto de Salud Carlos III | Instituto de Salud Carlos III | Pozo,F; Iglesias-Caballero, M.; Molinero,M.; Camarero,S.; Mart?n,V.; Reyes,N.; V?zquez-Mor?n,S.; Casas,I. |

| | | | | | | | | |
|---|---|---|---|---|---|---|---|---|
| EPI2398373 | NS | United Kingdom | 2022-Nov-11 | EPI_ISL_16904550 | A/Turkey/England/150027/2022 | Animal and Plant Health Agency (APHA) | Animal and Plant Health Agency (APHA) | |
| EPI2398374 | MP | United Kingdom | 2022-Nov-11 | EPI_ISL_16904550 | A/Turkey/England/150027/2022 | Animal and Plant Health Agency (APHA) | Animal and Plant Health Agency (APHA) | |
| EPI2398372 | NP | United Kingdom | 2022-Nov-11 | EPI_ISL_16904550 | A/Turkey/England/150027/2022 | Animal and Plant Health Agency (APHA) | Animal and Plant Health Agency (APHA) | |
| EPI2398380 | NP | United Kingdom | 2022-Dec-17 | EPI_ISL_16904551 | A/Chicken/Scotland/170118/2022 | Animal and Plant Health Agency (APHA) | Animal and Plant Health Agency (APHA) | |
| EPI2398381 | NS | United Kingdom | 2022-Dec-17 | EPI_ISL_16904551 | A/Chicken/Scotland/170118/2022 | Animal and Plant Health Agency (APHA) | Animal and Plant Health Agency (APHA) | |
| EPI2398382 | MP | United Kingdom | 2022-Dec-17 | EPI_ISL_16904551 | A/Chicken/Scotland/170118/2022 | Animal and Plant Health Agency (APHA) | Animal and Plant Health Agency (APHA) | |
| EPI2398388 | NP | United Kingdom | 2022-Dec-14 | EPI_ISL_16904552 | A/Chicken/Scotland/172652/2022 | Animal and Plant Health Agency (APHA) | Animal and Plant Health Agency (APHA) | |
| EPI2398389 | NS | United Kingdom | 2022-Dec-14 | EPI_ISL_16904552 | A/Chicken/Scotland/172652/2022 | Animal and Plant Health Agency (APHA) | Animal and Plant Health Agency (APHA) | |
| EPI2398390 | MP | United Kingdom | 2022-Dec-14 | EPI_ISL_16904552 | A/Chicken/Scotland/172652/2022 | Animal and Plant Health Agency (APHA) | Animal and Plant Health Agency (APHA) | |
| EPI2398397 | NS | United Kingdom | 2022-Mar-12 | EPI_ISL_16904553 | A/Harbour_Seal/Scotland/162919/2022 | Animal and Plant Health Agency (APHA) | Animal and Plant Health Agency (APHA) | |
| EPI2398398 | MP | United Kingdom | 2022-Mar-12 | EPI_ISL_16904553 | A/Harbour_Seal/Scotland/162919/2022 | Animal and Plant Health Agency (APHA) | Animal and Plant Health Agency (APHA) | |
| EPI2398396 | NP | United Kingdom | 2022-Mar-12 | EPI_ISL_16904553 | A/Harbour_Seal/Scotland/162919/2022 | Animal and Plant Health Agency (APHA) | Animal and Plant Health Agency (APHA) | |
| EPI2415011 | NP | Italy | 2022-Dec-30 | EPI_ISL_16979817 | A/black-headed_gull/Italy/23VIR1107-4/2022 | Istituto Zooprofilattico Sperimentale delle Venezie, EU/OIE/Reference Laboratory and FAO Reference Centre for AI and ND | Istituto Zooprofilattico Sperimentale Delle Venezie | |
| EPI2414967 | NS | Italy | 2022-Dec-30 | EPI_ISL_16979817 | A/black-headed_gull/Italy/23VIR1107-4/2022 | Istituto Zooprofilattico Sperimentale delle Venezie, EU/OIE/Reference Laboratory and FAO Reference Centre for AI and ND | Istituto Zooprofilattico Sperimentale Delle Venezie | |
| EPI2414968 | MP | Italy | 2022-Dec-30 | EPI_ISL_16979817 | A/black-headed_gull/Italy/23VIR1107-4/2022 | Istituto Zooprofilattico Sperimentale delle Venezie, EU/OIE/Reference Laboratory and FAO Reference Centre for AI and ND | Istituto Zooprofilattico Sperimentale Delle Venezie | |
| EPI2415354 | NP | Belgium | 2022-Dec-28 | EPI_ISL_16984685 | A/Chroicocephalus_ridibundus/Belgium/00132_0002/2022 | Sciensano - Animal Infectious Diseases | Sciensano, Department of Animal Infectious Diseases | Van Borm, Steven; Roupie, Virginie; Hostyn, Pierre; Lambrecht, Benedicte; Steensels, Mieke |
| EPI2415357 | NS | Belgium | 2022-Dec-28 | EPI_ISL_16984685 | A/Chroicocephalus_ridibundus/Belgium/00132_0002/2022 | Sciensano - Animal Infectious Diseases | Sciensano, Department of Animal Infectious Diseases | Van Borm, Steven; Roupie, Virginie; Hostyn, Pierre; Lambrecht, Benedicte; Steensels, Mieke |
| EPI2415356 | MP | Belgium | 2022-Dec-28 | EPI_ISL_16984685 | A/Chroicocephalus_ridibundus/Belgium/00132_0002/2022 | Sciensano - Animal Infectious Diseases | Sciensano, Department of Animal Infectious Diseases | Van Borm, Steven; Roupie, Virginie; Hostyn, Pierre; Lambrecht, Benedicte; Steensels, Mieke |
| EPI2415401 | MP | Belgium | 2022-Dec-15 | EPI_ISL_16984869 | A/ Anser_brachyrhynchus/Belgium/14776_0001/2022 | Sciensano - Animal Infectious Diseases | Sciensano, Department of Animal Infectious Diseases | Van Borm, Steven; Roupie, Virginie; Hostyn, Pierre; Lambrecht, Benedicte; Steensels, Mieke |
| EPI2415402 | NS | Belgium | 2022-Dec-15 | EPI_ISL_16984869 | A/ Anser_brachyrhynchus/Belgium/14776_0001/2022 | Sciensano - Animal Infectious Diseases | Sciensano, Department of Animal Infectious Diseases | Van Borm, Steven; Roupie, Virginie; Hostyn, Pierre; Lambrecht, Benedicte; Steensels, Mieke |
| EPI2415399 | NP | Belgium | 2022-Dec-15 | EPI_ISL_16984869 | A/ Anser_brachyrhynchus/Belgium/14776_0001/2022 | Sciensano - Animal Infectious Diseases | Sciensano, Department of Animal Infectious Diseases | Van Borm, Steven; Roupie, Virginie; Hostyn, Pierre; Lambrecht, Benedicte; Steensels, Mieke |
| EPI2415411 | NS | Belgium | 2022-Aug-29 | EPI_ISL_16984878 | A/Branta_canadensis/Belgium/10598_0003/2022 | Sciensano - Animal Infectious Diseases | Sciensano, Department of Animal Infectious Diseases | Van Borm, Steven; Roupie, Virginie; Hostyn, Pierre; Lambrecht, Benedicte; Steensels, Mieke |
| EPI2415408 | NP | Belgium | 2022-Aug-29 | EPI_ISL_16984878 | A/Branta_canadensis/Belgium/10598_0003/2022 | Sciensano - Animal Infectious Diseases | Sciensano, Department of Animal Infectious Diseases | Van Borm, Steven; Roupie, Virginie; Hostyn, Pierre; Lambrecht, Benedicte; Steensels, Mieke |
| EPI2415410 | MP | Belgium | 2022-Aug-29 | EPI_ISL_16984878 | A/Branta_canadensis/Belgium/10598_0003/2022 | Sciensano - Animal Infectious Diseases | Sciensano, Department of Animal Infectious Diseases | Van Borm, Steven; Roupie, Virginie; Hostyn, Pierre; Lambrecht, Benedicte; Steensels, Mieke |
| EPI2415416 | NP | Belgium | 2022-Dec-24 | EPI_ISL_16985112 | A/Meleagris_gallopavo/Belgium/14826_0002/2022 | Sciensano - Animal Infectious Diseases | Sciensano, Department of Animal Infectious Diseases | Van Borm, Steven; Roupie, Virginie; Hostyn, Pierre; Lambrecht, Benedicte; Steensels, Mieke |
| EPI2415418 | MP | Belgium | 2022-Dec-24 | EPI_ISL_16985112 | A/Meleagris_gallopavo/Belgium/14826_0002/2022 | Sciensano - Animal Infectious Diseases | Sciensano, Department of Animal Infectious Diseases | Van Borm, Steven; Roupie, Virginie; Hostyn, Pierre; Lambrecht, Benedicte; Steensels, Mieke |
| EPI2415419 | NS | Belgium | 2022-Dec-24 | EPI_ISL_16985112 | A/Meleagris_gallopavo/Belgium/14826_0002/2022 | Sciensano - Animal Infectious Diseases | Sciensano, Department of Animal Infectious Diseases | Van Borm, Steven; Roupie, Virginie; Hostyn, Pierre; Lambrecht, Benedicte; Steensels, Mieke |
| EPI2436846 | NS | Belgium | 2022-Aug-28 | EPI_ISL_17071430 | A/Branta_canadensis/Belgium/10393_0025/2022 | Sciensano - Animal Infectious Diseases | Sciensano, Department of Animal Infectious Diseases | Van Borm, Steven; Roupie, Virginie; Hostyn, Pierre; Mathijs, Elisabeth; Lambrecht, Benedicte; Steensels, Mieke |
| EPI2436844 | MP | Belgium | 2022-Aug-28 | EPI_ISL_17071430 | A/Branta_canadensis/Belgium/10393_0025/2022 | Sciensano - Animal Infectious Diseases | Sciensano, Department of Animal Infectious Diseases | Van Borm, Steven; Roupie, Virginie; Hostyn, Pierre; Mathijs, Elisabeth; Lambrecht, Benedicte; Steensels, Mieke |
| EPI2436842 | NP | Belgium | 2022-Aug-28 | EPI_ISL_17071430 | A/Branta_canadensis/Belgium/10393_0025/2022 | Sciensano - Animal Infectious Diseases | Sciensano, Department of Animal Infectious Diseases | Van Borm, Steven; Roupie, Virginie; Hostyn, Pierre; Mathijs, Elisabeth; Lambrecht, Benedicte; Steensels, Mieke |
| EPI2436895 | NS | Belgium | 2022-Dec-18 | EPI_ISL_17071930 | A/Anser_anser/Belgium/14804_0002/2022 | Sciensano - Animal Infectious Diseases | Sciensano, Department of Animal Infectious Diseases | Van Borm, Steven; Roupie, Virginie; Hostyn, Pierre; Mathijs, Elisabeth; Lambrecht, Benedicte; Steensels, Mieke |
| EPI2436893 | MP | Belgium | 2022-Dec-18 | EPI_ISL_17071930 | A/Anser_anser/Belgium/14804_0002/2022 | Sciensano - Animal Infectious Diseases | Sciensano, Department of Animal Infectious Diseases | Van Borm, Steven; Roupie, Virginie; Hostyn, Pierre; Mathijs, Elisabeth; Lambrecht, Benedicte; Steensels, Mieke |
| EPI2436889 | NP | Belgium | 2022-Dec-18 | EPI_ISL_17071930 | A/Anser_anser/Belgium/14804_0002/2022 | Sciensano - Animal Infectious Diseases | Sciensano, Department of Animal Infectious Diseases | Van Borm, Steven; Roupie, Virginie; Hostyn, Pierre; Mathijs, Elisabeth; Lambrecht, Benedicte; Steensels, Mieke |
| EPI2437461 | NP | United Kingdom | 2022-Aug-23 | EPI_ISL_17072388 | A/fox/England/015850/2022 | Animal and Plant Health Agency (APHA) | Animal and Plant Health Agency (APHA) | |
| EPI2437462 | NS | United Kingdom | 2022-Aug-23 | EPI_ISL_17072388 | A/fox/England/015850/2022 | Animal and Plant Health Agency (APHA) | Animal and Plant Health Agency (APHA) | |

| | | | | | | | | |
|---|---|---|---|---|---|---|---|---|
| EPI2437463 | MP | United Kingdom | 2022-Aug-23 | EPI_ISL_17072388 | A/fox/England/015850/2022 | Animal and Plant Health Agency (APHA) | Animal and Plant Health Agency (APHA) | |
| EPI2455221 | NP | Romania | 2022-Nov-15 | EPI_ISL_17165500 | A/cygnus_olor/Romania/17142_23VIR1145-1/2022 | Institute for Diagnosis & Animal Health (IDAH) | Istituto Zooprofilattico Sperimentale Delle Venezie | Barbuceanu, F.; Onita, I.; Neicut, A.; Motiu, R.; Burlacu, R.; Pastori, A.; Zecchin, B.; Fusaro, A.; Schivo, A.; Salviato, A.; Palumbo, E.; Giussani, E.; Monne, I.; Terregino, C. |
| EPI2455222 | NS | Romania | 2022-Nov-15 | EPI_ISL_17165500 | A/cygnus_olor/Romania/17142_23VIR1145-1/2022 | Institute for Diagnosis & Animal Health (IDAH) | Istituto Zooprofilattico Sperimentale Delle Venezie | Barbuceanu, F.; Onita, I.; Neicut, A.; Motiu, R.; Burlacu, R.; Pastori, A.; Zecchin, B.; Fusaro, A.; Schivo, A.; Salviato, A.; Palumbo, E.; Giussani, E.; Monne, I.; Terregino, C. |
| EPI2455223 | MP | Romania | 2022-Nov-15 | EPI_ISL_17165500 | A/cygnus_olor/Romania/17142_23VIR1145-1/2022 | Institute for Diagnosis & Animal Health (IDAH) | Istituto Zooprofilattico Sperimentale Delle Venezie | Barbuceanu, F.; Onita, I.; Neicut, A.; Motiu, R.; Burlacu, R.; Pastori, A.; Zecchin, B.; Fusaro, A.; Schivo, A.; Salviato, A.; Palumbo, E.; Giussani, E.; Monne, I.; Terregino, C. |
| EPI2455229 | NP | Romania | 2022-Nov-17 | EPI_ISL_17165501 | A/cygnus_cygnus/Romania/17198_23VIR1145-2/2022 | Institute for Diagnosis & Animal Health (IDAH) | Istituto Zooprofilattico Sperimentale Delle Venezie | Barbuceanu, F.; Onita, I.; Neicut, A.; Motiu, R.; Burlacu, R.; Pastori, A.; Zecchin, B.; Fusaro, A.; Schivo, A.; Salviato, A.; Palumbo, E.; Giussani, E.; Monne, I.; Terregino, C. |
| EPI2455230 | NS | Romania | 2022-Nov-17 | EPI_ISL_17165501 | A/cygnus_cygnus/Romania/17198_23VIR1145-2/2022 | Institute for Diagnosis & Animal Health (IDAH) | Istituto Zooprofilattico Sperimentale Delle Venezie | Barbuceanu, F.; Onita, I.; Neicut, A.; Motiu, R.; Burlacu, R.; Pastori, A.; Zecchin, B.; Fusaro, A.; Schivo, A.; Salviato, A.; Palumbo, E.; Giussani, E.; Monne, I.; Terregino, C. |
| EPI2455231 | MP | Romania | 2022-Nov-17 | EPI_ISL_17165501 | A/cygnus_cygnus/Romania/17198_23VIR1145-2/2022 | Institute for Diagnosis & Animal Health (IDAH) | Istituto Zooprofilattico Sperimentale Delle Venezie | Barbuceanu, F.; Onita, I.; Neicut, A.; Motiu, R.; Burlacu, R.; Pastori, A.; Zecchin, B.; Fusaro, A.; Schivo, A.; Salviato, A.; Palumbo, E.; Giussani, E.; Monne, I.; Terregino, C. |
| EPI2456197 | NS | Switzerland | 2022-Nov-16 | EPI_ISL_17179652 | A/peacock/Switzerland-Zurich/221192/2022 | Nationales Referenzlabor für Geflügel- und Kaninchenkrankheiten | Institute of Virology and Immunology (IVI) | |
| EPI2456194 | NP | Switzerland | 2022-Nov-16 | EPI_ISL_17179652 | A/peacock/Switzerland-Zurich/221192/2022 | Nationales Referenzlabor für Geflügel- und Kaninchenkrankheiten | Institute of Virology and Immunology (IVI) | |
| EPI2456196 | MP | Switzerland | 2022-Nov-16 | EPI_ISL_17179652 | A/peacock/Switzerland-Zurich/221192/2022 | Nationales Referenzlabor für Geflügel- und Kaninchenkrankheiten | Institute of Virology and Immunology (IVI) | |
| EPI2456205 | NS | Switzerland | 2022-Nov-22 | EPI_ISL_17179653 | A/mute-swan/Switzerland-Ticino/221214/2022 | Nationales Referenzlabor für Geflügel- und Kaninchenkrankheiten | Institute of Virology and Immunology (IVI) | |
| EPI2456204 | MP | Switzerland | 2022-Nov-22 | EPI_ISL_17179653 | A/mute-swan/Switzerland-Ticino/221214/2022 | Nationales Referenzlabor für Geflügel- und Kaninchenkrankheiten | Institute of Virology and Immunology (IVI) | |
| EPI2456202 | NP | Switzerland | 2022-Nov-22 | EPI_ISL_17179653 | A/mute-swan/Switzerland-Ticino/221214/2022 | Nationales Referenzlabor für Geflügel- und Kaninchenkrankheiten | Institute of Virology and Immunology (IVI) | |
| EPI2471929 | NP | United Kingdom | 2022-Nov-18 | EPI_ISL_17257532 | A/Bush_dog/England/037680/2022 | Animal and Plant Health Agency (APHA) | Animal and Plant Health Agency (APHA) | |
| EPI2471930 | NS | United Kingdom | 2022-Nov-18 | EPI_ISL_17257532 | A/Bush_dog/England/037680/2022 | Animal and Plant Health Agency (APHA) | Animal and Plant Health Agency (APHA) | |
| EPI2471931 | MP | United Kingdom | 2022-Nov-18 | EPI_ISL_17257532 | A/Bush_dog/England/037680/2022 | Animal and Plant Health Agency (APHA) | Animal and Plant Health Agency (APHA) | |
| EPI2475286 | NP | United Kingdom | 2022-Nov-14 | EPI_ISL_17268132 | A/chicken/England/152082/2022 | Animal and Plant Health Agency (APHA) | Animal and Plant Health Agency (APHA) | |
| EPI2475287 | NS | United Kingdom | 2022-Nov-14 | EPI_ISL_17268132 | A/chicken/England/152082/2022 | Animal and Plant Health Agency (APHA) | Animal and Plant Health Agency (APHA) | |
| EPI2475288 | MP | United Kingdom | 2022-Nov-14 | EPI_ISL_17268132 | A/chicken/England/152082/2022 | Animal and Plant Health Agency (APHA) | Animal and Plant Health Agency (APHA) | |
| EPI2475294 | NP | United Kingdom | 2022-Oct-21 | EPI_ISL_17268133 | A/Canada_goose/England/394558/2022 | Animal and Plant Health Agency (APHA) | Animal and Plant Health Agency (APHA) | |
| EPI2475295 | NS | United Kingdom | 2022-Oct-21 | EPI_ISL_17268133 | A/Canada_goose/England/394558/2022 | Animal and Plant Health Agency (APHA) | Animal and Plant Health Agency (APHA) | |
| EPI2475296 | MP | United Kingdom | 2022-Oct-21 | EPI_ISL_17268133 | A/Canada_goose/England/394558/2022 | Animal and Plant Health Agency (APHA) | Animal and Plant Health Agency (APHA) | |
| EPI2576137 | NS | Denmark | 2022-Jan-17 | EPI_ISL_17447412 | A/red fox/Denmark/01679-1.01-2/2022 | Statens Serum Institute | Statens Serum Institute | Hjulsager,Charlotte; Liang,Yuan |
| EPI2576134 | NP | Denmark | 2022-Jan-17 | EPI_ISL_17447412 | A/red fox/Denmark/01679-1.01-2/2022 | Statens Serum Institute | Statens Serum Institute | Hjulsager,Charlotte; Liang,Yuan |
| EPI2576136 | MP | Denmark | 2022-Jan-17 | EPI_ISL_17447412 | A/red fox/Denmark/01679-1.01-2/2022 | Statens Serum Institute | Statens Serum Institute | Hjulsager,Charlotte; Liang,Yuan |
| EPI2576142 | NP | Denmark | 2022-Apr-28 | EPI_ISL_17447413 | A/red fox/Denmark/08658-1.01/2022 | Statens Serumm Institute | Statens Serum Institute | Hjulsager,Charlotte; Liang,Yuan |
| EPI2576144 | MP | Denmark | 2022-Apr-28 | EPI_ISL_17447413 | A/red fox/Denmark/08658-1.01/2022 | Statens Serumm Institute | Statens Serum Institute | Hjulsager,Charlotte; Liang,Yuan |
| EPI2576145 | NS | Denmark | 2022-Apr-28 | EPI_ISL_17447413 | A/red fox/Denmark/08658-1.01/2022 | Statens Serumm Institute | Statens Serum Institute | Hjulsager,Charlotte; Liang,Yuan |
| EPI2576153 | NS | Denmark | 2022-Apr-28 | EPI_ISL_17447414 | A/red fox/Denmark/08658-2.02-2/2022 | Statens Serum Institute | Statens Serum Institute | Hjulsager,Charlotte; Liang,Yuan |
| EPI2576150 | NP | Denmark | 2022-Apr-28 | EPI_ISL_17447414 | A/red fox/Denmark/08658-2.02-2/2022 | Statens Serum Institute | Statens Serum Institute | Hjulsager,Charlotte; Liang,Yuan |
| EPI2576152 | MP | Denmark | 2022-Apr-28 | EPI_ISL_17447414 | A/red fox/Denmark/08658-2.02-2/2022 | Statens Serum Institute | Statens Serum Institute | Hjulsager,Charlotte; Liang,Yuan |
| EPI2576158 | NP | Denmark | 2022-Apr-28 | EPI_ISL_17447440 | A/red fox/Denmark/08658-3.02-2/2022 | Statens Serumm Institute | Statens Serum Institute | Hjulsager,Charlotte;Liang,Yuan |
| EPI2576160 | MP | Denmark | 2022-Apr-28 | EPI_ISL_17447440 | A/red fox/Denmark/08658-3.02-2/2022 | Statens Serumm Institute | Statens Serum Institute | Hjulsager,Charlotte;Liang,Yuan |
| EPI2576161 | NS | Denmark | 2022-Apr-28 | EPI_ISL_17447440 | A/red fox/Denmark/08658-3.02-2/2022 | Statens Serumm Institute | Statens Serum Institute | Hjulsager,Charlotte;Liang,Yuan |
| EPI2509900 | NP | United Kingdom | 2022-Jun-19 | EPI_ISL_17465835 | A/harbour_seal/Scotland/054948/2022 | Animal and Plant Health Agency (APHA) | Animal and Plant Health Agency (APHA) | |

| | | | | | | | | |
|---|---|---|---|---|---|---|---|---|
| EPI2509901 | NS | United Kingdom | 2022-Jun-19 | EPI_ISL_17465835 | A/harbour_seal/Scotland/054948/2022 | Animal and Plant Health Agency (APHA) | Animal and Plant Health Agency (APHA) | |
| EPI2509902 | MP | United Kingdom | 2022-Jun-19 | EPI_ISL_17465835 | A/harbour_seal/Scotland/054948/2022 | Animal and Plant Health Agency (APHA) | Animal and Plant Health Agency (APHA) | |
| EPI2525929 | NP | Austria | 2022-Dec-28 | EPI_ISL_17512128 | A/mute_swan/Austria/22155612/2022 | Institute for Veterinary Disease Control Moedling, Austrian Agency for Health and Food Safety | Austrian Agency for Health and Food Safety (AGES) | |
| EPI2525930 | NS | Austria | 2022-Dec-28 | EPI_ISL_17512128 | A/mute_swan/Austria/22155612/2022 | Institute for Veterinary Disease Control Moedling, Austrian Agency for Health and Food Safety | Austrian Agency for Health and Food Safety (AGES) | |
| EPI2525931 | MP | Austria | 2022-Dec-28 | EPI_ISL_17512128 | A/mute_swan/Austria/22155612/2022 | Institute for Veterinary Disease Control Moedling, Austrian Agency for Health and Food Safety | Austrian Agency for Health and Food Safety (AGES) | |
| EPI2536808 | NP | Netherlands | 2022-Jan-16 | EPI_ISL_17580930 | A/Stone Marten/Netherlands/1/2022 | Erasmus Medical Center | Erasmus Medical Center | Vuong,O;Thewessen, S;Bellido-Martin,A.B; Chestakova, I; Sikkema, R;Fouchier, R.A.M. |
| EPI2536809 | NS | Netherlands | 2022-Jan-16 | EPI_ISL_17580930 | A/Stone Marten/Netherlands/1/2022 | Erasmus Medical Center | Erasmus Medical Center | Vuong,O;Thewessen, S;Bellido-Martin,A.B; Chestakova, I; Sikkema, R;Fouchier, R.A.M. |
| EPI2536810 | MP | Netherlands | 2022-Jan-16 | EPI_ISL_17580930 | A/Stone Marten/Netherlands/1/2022 | Erasmus Medical Center | Erasmus Medical Center | Vuong,O;Thewessen, S;Bellido-Martin,A.B; Chestakova, I; Sikkema, R;Fouchier, R.A.M. |
| EPI2536824 | NP | Netherlands | 2022-Jan-16 | EPI_ISL_17583227 | A/Stone Marten/Netherlands/EMC1/2022 | Erasmus Medical Center | Erasmus Medical Center | Vuong,O;Thewessen, S;Bellido-Martin,A.B; Chestakova, I; Sikkema, R;Fouchier, R.A.M. |
| EPI2536825 | NS | Netherlands | 2022-Jan-16 | EPI_ISL_17583227 | A/Stone Marten/Netherlands/EMC1/2022 | Erasmus Medical Center | Erasmus Medical Center | Vuong,O;Thewessen, S;Bellido-Martin,A.B; Chestakova, I; Sikkema, R;Fouchier, R.A.M. |
| EPI2536826 | MP | Netherlands | 2022-Jan-16 | EPI_ISL_17583227 | A/Stone Marten/Netherlands/EMC1/2022 | Erasmus Medical Center | Erasmus Medical Center | Vuong,O;Thewessen, S;Bellido-Martin,A.B; Chestakova, I; Sikkema, R;Fouchier, R.A.M. |
| EPI2536832 | NP | Netherlands | 2022-Sep-13 | EPI_ISL_17583228 | A/Fox/Netherlands/EMC7/2022 | Erasmus Medical Center | Erasmus Medical Center | Vuong,O;Thewessen, S;Bellido-Martin,A.B; Chestakova, I; Sikkema, R;Fouchier, R.A.M. |
| EPI2536833 | NS | Netherlands | 2022-Sep-13 | EPI_ISL_17583228 | A/Fox/Netherlands/EMC7/2022 | Erasmus Medical Center | Erasmus Medical Center | Vuong,O;Thewessen, S;Bellido-Martin,A.B; Chestakova, I; Sikkema, R;Fouchier, R.A.M. |
| EPI2536834 | MP | Netherlands | 2022-Sep-13 | EPI_ISL_17583228 | A/Fox/Netherlands/EMC7/2022 | Erasmus Medical Center | Erasmus Medical Center | Vuong,O;Thewessen, S;Bellido-Martin,A.B; Chestakova, I; Sikkema, R;Fouchier, R.A.M. |
| EPI2536874 | NP | France | 2022-Jan-04 | EPI_ISL_17584742 | A/duck/France/22005/2022 | | Import from public-domain | Croville,G.; Walch,M.; Lebre,L.; Silva,S.; Filaire,F.; Guerin,J.-L. |
| EPI2536898 | MP | France | 2022-Jan-04 | EPI_ISL_17584742 | A/duck/France/22005/2022 | | Import from public-domain | Croville,G.; Walch,M.; Lebre,L.; Silva,S.; Filaire,F.; Guerin,J.-L. |
| EPI2536907 | NS | France | 2022-Jan-04 | EPI_ISL_17584742 | A/duck/France/22005/2022 | | Import from public-domain | Croville,G.; Walch,M.; Lebre,L.; Silva,S.; Filaire,F.; Guerin,J.-L. |
| EPI2540349 | NP | Belgium | 2022-Jun-17 | EPI_ISL_17606776 | A/Larus_argentatus/Belgium/08182_0012/2022 | Sciensano - Animal Infectious Diseases | Sciensano, Department of Animal Infectious Diseases | Van Borm, Steven; Roupie, Virginie; Hostyn, Pierre; Mathijs, Elisabeth; Lambrecht, Benedicte; Steensels, Mieke |
| EPI2540352 | NS | Belgium | 2022-Jun-17 | EPI_ISL_17606776 | A/Larus_argentatus/Belgium/08182_0012/2022 | Sciensano - Animal Infectious Diseases | Sciensano, Department of Animal Infectious Diseases | Van Borm, Steven; Roupie, Virginie; Hostyn, Pierre; Mathijs, Elisabeth; Lambrecht, Benedicte; Steensels, Mieke |
| EPI2540351 | MP | Belgium | 2022-Jun-17 | EPI_ISL_17606776 | A/Larus_argentatus/Belgium/08182_0012/2022 | Sciensano - Animal Infectious Diseases | Sciensano, Department of Animal Infectious Diseases | Van Borm, Steven; Roupie, Virginie; Hostyn, Pierre; Mathijs, Elisabeth; Lambrecht, Benedicte; Steensels, Mieke |
| EPI2540359 | MP | Belgium | 2022-Dec-19 | EPI_ISL_17606777 | A/Gallus_gallus/Belgium/14642_0002/2022 | Sciensano - Animal Infectious Diseases | Sciensano, Department of Animal Infectious Diseases | Van Borm, Steven; Roupie, Virginie; Hostyn, Pierre; Mathijs, Elisabeth; Lambrecht, Benedicte; Steensels, Mieke |
| EPI2540360 | NS | Belgium | 2022-Dec-19 | EPI_ISL_17606777 | A/Gallus_gallus/Belgium/14642_0002/2022 | Sciensano - Animal Infectious Diseases | Sciensano, Department of Animal Infectious Diseases | Van Borm, Steven; Roupie, Virginie; Hostyn, Pierre; Mathijs, Elisabeth; Lambrecht, Benedicte; Steensels, Mieke |
| EPI2540357 | NP | Belgium | 2022-Dec-19 | EPI_ISL_17606777 | A/Gallus_gallus/Belgium/14642_0002/2022 | Sciensano - Animal Infectious Diseases | Sciensano, Department of Animal Infectious Diseases | Van Borm, Steven; Roupie, Virginie; Hostyn, Pierre; Mathijs, Elisabeth; Lambrecht, Benedicte; Steensels, Mieke |
| EPI2540365 | NP | Belgium | 2022-Dec-27 | EPI_ISL_17606778 | A/Gallus_gallus/Belgium/14828_0003/2022 | Sciensano - Animal Infectious Diseases | Sciensano, Department of Animal Infectious Diseases | Van Borm, Steven; Roupie, Virginie; Hostyn, Pierre; Mathijs, Elisabeth; Lambrecht, Benedicte; Steensels, Mieke |
| EPI2540368 | NS | Belgium | 2022-Dec-27 | EPI_ISL_17606778 | A/Gallus_gallus/Belgium/14828_0003/2022 | Sciensano - Animal Infectious Diseases | Sciensano, Department of Animal Infectious Diseases | Van Borm, Steven; Roupie, Virginie; Hostyn, Pierre; Mathijs, Elisabeth; Lambrecht, Benedicte; Steensels, Mieke |
| EPI2540367 | MP | Belgium | 2022-Dec-27 | EPI_ISL_17606778 | A/Gallus_gallus/Belgium/14828_0003/2022 | Sciensano - Animal Infectious Diseases | Sciensano, Department of Animal Infectious Diseases | Van Borm, Steven; Roupie, Virginie; Hostyn, Pierre; Mathijs, Elisabeth; Lambrecht, Benedicte; Steensels, Mieke |
| EPI2550039 | NP | United Kingdom | 2022-Jan-24 | EPI_ISL_17637991 | A/LutraLutra/England/4197OP/2022 | Public Health Wales Microbiology Cardiff | Public Health Wales Microbiology Cardiff | Catherine,Moore; Elizabeth,Chadwick; Kathleen,Pheasant; Tim,Jones; Anastasia,Couzens; Chloe,Farrington; Joanne,Watkins; Sally,Corden and Tom Connor |
| EPI2550037 | MP | United Kingdom | 2022-Jan-24 | EPI_ISL_17637991 | A/LutraLutra/England/4197OP/2022 | Public Health Wales Microbiology Cardiff | Public Health Wales Microbiology Cardiff | Catherine,Moore; Elizabeth,Chadwick; Kathleen,Pheasant; Tim,Jones; Anastasia,Couzens; Chloe,Farrington; Joanne,Watkins; Sally,Corden and Tom Connor |
| EPI2550040 | NS | United Kingdom | 2022-Jan-24 | EPI_ISL_17637991 | A/LutraLutra/England/4197OP/2022 | Public Health Wales Microbiology Cardiff | Public Health Wales Microbiology Cardiff | Catherine,Moore; Elizabeth,Chadwick; Kathleen,Pheasant; Tim,Jones; Anastasia,Couzens; Chloe,Farrington; Joanne,Watkins; Sally,Corden and Tom Connor |
| EPI2580770 | MP | Italy | 2022-Oct-21 | EPI_ISL_17736969 | A/chicken/Italy/22VIR10044-14/2022 | Istituto Zooprofilattico Sperimentale delle Venezie, EU/OIE/Reference Laboratory and FAO Reference Centre for AI and ND | Istituto Zooprofilattico Sperimentale Delle Venezie | Zecchin, B.;Pastori, A.;Fusaro, A.;Schivo, A.;Salviato, A.;Palumbo, E.;Giussani, E.;Monne, I.;Bonfante, F.;Terregino, C. |
| EPI2580768 | NP | Italy | 2022-Oct-21 | EPI_ISL_17736969 | A/chicken/Italy/22VIR10044-14/2022 | Istituto Zooprofilattico Sperimentale delle Venezie, EU/OIE/Reference Laboratory and FAO Reference Centre for AI and ND | Istituto Zooprofilattico Sperimentale Delle Venezie | Zecchin, B.;Pastori, A.;Fusaro, A.;Schivo, A.;Salviato, A.;Palumbo, E.;Giussani, E.;Monne, I.;Bonfante, F.;Terregino, C. |
| EPI2580771 | NS | Italy | 2022-Oct-21 | EPI_ISL_17736969 | A/chicken/Italy/22VIR10044-14/2022 | Istituto Zooprofilattico Sperimentale delle Venezie, EU/OIE/Reference Laboratory and FAO Reference Centre for AI and ND | Istituto Zooprofilattico Sperimentale Delle Venezie | Zecchin, B.;Pastori, A.;Fusaro, A.;Schivo, A.;Salviato, A.;Palumbo, E.;Giussani, E.;Monne, I.;Bonfante, F.;Terregino, C. |
| EPI2589244 | NP | Austria | 2022-Jan-14 | EPI_ISL_17785726 | A/swan/Austria/22004012-001/2022 | Austrian Agency for Health and Food Safety (AGES) | Austrian Agency for Health and Food Safety (AGES) | |
| EPI2589245 | NS | Austria | 2022-Jan-14 | EPI_ISL_17785726 | A/swan/Austria/22004012-001/2022 | Austrian Agency for Health and Food Safety (AGES) | Austrian Agency for Health and Food Safety (AGES) | |
| EPI2589246 | MP | Austria | 2022-Jan-14 | EPI_ISL_17785726 | A/swan/Austria/22004012-001/2022 | Austrian Agency for Health and Food Safety (AGES) | Austrian Agency for Health and Food Safety (AGES) | |

| | | | | | | | | |
|---|---|---|---|---|---|---|---|---|
| EPI2589252 | NP | Austria | 2022-Jan-19 | EPI_ISL_17785727 | A/laying_hen/Austria/22005859-001/2022 | Austrian Agency for Health and Food Safety (AGES) | Austrian Agency for Health and Food Safety (AGES) | |
| EPI2589253 | NS | Austria | 2022-Jan-19 | EPI_ISL_17785727 | A/laying_hen/Austria/22005859-001/2022 | Austrian Agency for Health and Food Safety (AGES) | Austrian Agency for Health and Food Safety (AGES) | |
| EPI2589254 | MP | Austria | 2022-Jan-19 | EPI_ISL_17785727 | A/laying_hen/Austria/22005859-001/2022 | Austrian Agency for Health and Food Safety (AGES) | Austrian Agency for Health and Food Safety (AGES) | |
| EPI2589262 | MP | Austria | 2022-Apr-13 | EPI_ISL_17785728 | A/swan/Austria/22046299-002/2022 | Austrian Agency for Health and Food Safety (AGES) | Austrian Agency for Health and Food Safety (AGES) | |
| EPI2589260 | NP | Austria | 2022-Apr-13 | EPI_ISL_17785728 | A/swan/Austria/22046299-002/2022 | Austrian Agency for Health and Food Safety (AGES) | Austrian Agency for Health and Food Safety (AGES) | |
| EPI2589261 | NS | Austria | 2022-Apr-13 | EPI_ISL_17785728 | A/swan/Austria/22046299-002/2022 | Austrian Agency for Health and Food Safety (AGES) | Austrian Agency for Health and Food Safety (AGES) | |
| EPI2589284 | NP | Austria | 2022-Jan-03 | EPI_ISL_17785731 | A/mute swan/Austria/22000062-001/2022 | Austrian Agency for Health and Food Safety (AGES) | Austrian Agency for Health and Food Safety (AGES) | |
| EPI2589285 | NS | Austria | 2022-Jan-03 | EPI_ISL_17785731 | A/mute swan/Austria/22000062-001/2022 | Austrian Agency for Health and Food Safety (AGES) | Austrian Agency for Health and Food Safety (AGES) | |
| EPI2589286 | MP | Austria | 2022-Jan-03 | EPI_ISL_17785731 | A/mute swan/Austria/22000062-001/2022 | Austrian Agency for Health and Food Safety (AGES) | Austrian Agency for Health and Food Safety (AGES) | |
| EPI2589292 | NP | Austria | 2022-Jan-24 | EPI_ISL_17785732 | A/mute swan/Austria/22007848-001/2022 | Austrian Agency for Health and Food Safety (AGES) | Austrian Agency for Health and Food Safety (AGES) | |
| EPI2589293 | NS | Austria | 2022-Jan-24 | EPI_ISL_17785732 | A/mute swan/Austria/22007848-001/2022 | Austrian Agency for Health and Food Safety (AGES) | Austrian Agency for Health and Food Safety (AGES) | |
| EPI2589294 | MP | Austria | 2022-Jan-24 | EPI_ISL_17785732 | A/mute swan/Austria/22007848-001/2022 | Austrian Agency for Health and Food Safety (AGES) | Austrian Agency for Health and Food Safety (AGES) | |
| EPI2589300 | NP | Austria | 2022-Mar-11 | EPI_ISL_17785733 | A/swan/Austria/22030844-004/2022 | Austrian Agency for Health and Food Safety (AGES) | Austrian Agency for Health and Food Safety (AGES) | |
| EPI2589301 | NS | Austria | 2022-Mar-11 | EPI_ISL_17785733 | A/swan/Austria/22030844-004/2022 | Austrian Agency for Health and Food Safety (AGES) | Austrian Agency for Health and Food Safety (AGES) | |
| EPI2589302 | MP | Austria | 2022-Mar-11 | EPI_ISL_17785733 | A/swan/Austria/22030844-004/2022 | Austrian Agency for Health and Food Safety (AGES) | Austrian Agency for Health and Food Safety (AGES) | |
| EPI2073522 | NP | Netherlands | 2022-May-26 | EPI_ISL_13399697 | A/Greylag Goose/Netherlands/3/2022 | Erasmus Medical Center | Erasmus Medical Center | |
| EPI2073523 | NS | Netherlands | 2022-May-26 | EPI_ISL_13399697 | A/Greylag Goose/Netherlands/3/2022 | Erasmus Medical Center | Erasmus Medical Center | |
| EPI2073524 | MP | Netherlands | 2022-May-26 | EPI_ISL_13399697 | A/Greylag Goose/Netherlands/3/2022 | Erasmus Medical Center | Erasmus Medical Center | |
| EPI2437850 | HA | Netherlands | 2022-February-18 | EPI_ISL_17076095 | A/Netherlands/10355/2023 | Academic Medical Center. University of Amsterdam | RIVM | A, Meijer; L, Wijsman; S, vd Brink |
| EPI2437849 | NA | Netherlands | 2022-February-18 | EPI_ISL_17076095 | A/Netherlands/10355/2023 | Academic Medical Center. University of Amsterdam | RIVM | A, Meijer; L, Wijsman; S, vd Brink |
| EPI2264455 | HA | UK | 2022-December-12 | EPI_ISL_16305454 | A/Barry/9950/2022 | Public Health Wales Microbiology Cardiff | Public Health Wales Microbiology Cardiff | Public Health Wales Microbiology Cardiff |
| EPI2264457 | NA | UK | 2022-December-12 | EPI_ISL_16305454 | A/Barry/9950/2022 | Public Health Wales Microbiology Cardiff | Public Health Wales Microbiology Cardiff | Public Health Wales Microbiology Cardiff |
| EPI2305188 | HA | Australia | 2023-January-5 | EPI_ISL_16613693 | A/Victoria/7/2023 | Austin Health | WHO Collaborating Centre for Reference and Research on Influenza | Deng, Y-M; Barr, I; Aziz, A. |
| EPI2305187 | NA | Australia | 2023-January-5 | EPI_ISL_16613693 | A/Victoria/7/2023 | Austin Health | WHO Collaborating Centre for Reference and Research on Influenza | Deng, Y-M; Barr, I; Aziz, A. |
| EPI1661231 | HA | USA | 2019-December-19 | EPI_ISL_404460 | A/Wisconsin/588/2019 | Wisconsin State Laboratory of Hygiene | CDC | CDC |
| EPI1661230 | NA | USA | 2019-December-19 | EPI_ISL_404460 | A/Wisconsin/588/2019 | Wisconsin State Laboratory of Hygiene | CDC | CDC |
| EPI1617983 | HA | USA | 2019-October-5 | EPI_ISL_397028 | A/Hawaii/70/2019 | State of Hawaii Department of Health | CDC | CDC |
| EPI1617982 | NA | USA | 2019-October-5 | EPI_ISL_397028 | A/Hawaii/70/2019 | State of Hawaii Department of Health | CDC | CDC |
| EPI1542570 | HA | China | 2019-June-17 | EPI_ISL_377080 | A/Guangdong-Moanan/SWL1536/2019 | WHO Chinese National Influenza Center | WHO Chinese National Influenza Center | Xiaoxu, Zeng; Xiyan, Li; Weijuan, Huang; Lei, Yang; Dayan, Wang. |
| EPI1542569 | NA | China | 2019-June-17 | EPI_ISL_377080 | A/Guangdong-Moanan/SWL1536/2019 | WHO Chinese National Influenza Center | WHO Chinese National Influenza Center | Xiaoxu, Zeng; Xiyan, Li; Weijuan, Huang; Lei, Yang; Dayan, Wang. |
| EPI2226437 | HA | Norway | 2022-August-22 | EPI_ISL_15929455 | A/Norway/30483/2022 | WHO National Influenza Centre | Crick Worldwide Influenza Centre | |
| EPI2226438 | NA | Norway | 2022-August-22 | EPI_ISL_15929455 | A/Norway/30483/2022 | WHO National Influenza Centre | Crick Worldwide Influenza Centre | |
| EPI2397499 | HA | Sri Lanka | 2023-January-2 | EPI_ISL_16899227 | A/Sri Lanka/1/2023 | Medical Research Institute | WHO Collaborating Centre for Reference and Research on Influenza | |
| EPI2397498 | NA | Sri Lanka | 2023-January-2 | EPI_ISL_16899227 | A/Sri Lanka/1/2023 | Medical Research Institute | WHO Collaborating Centre for Reference and Research on Influenza | |
| EPI2308489 | HA | Pakistan | 2023-January-4 | EPI_ISL_16630764 | A/Pakistan/GIHSN-ICT-123/2023 | NIH Pakistan | NIH Pakistan | Nazish, Bader; Aemer,Ikram; Muhammad, Salman; HamzaAhmad, Mirza; Abdul, Ahad |

| | | | | | | | | |
|---|---|---|---|---|---|---|---|---|
| EPI23084892 | NA | Pakistan | 2023-January-4 | EPI_ISL_16630764 | A/Pakistan/GIHSN-ICT-123/2023 | NIH Pakistan | NIH Pakistan | Nazish, Bader; Aemer,Ikram; Muhammad, Salman; HamzaAhmad, Mirza; Abdul, Ahad |
| EPI2457821 | HA | Spain | 2023-March-7 | EPI_ISL_17190000 | A/Catalonia/NSAV198269034/2023 | Hospital Universitari Arnau del Vilanova | Hospital Universitari Vall d'Hebron | Andres, C.; Gonzalez-Sanchez, A; Pinana, M; Martin,MC.; Codina, MG.; Pumarola, T; Anton, A. |
| EPI2457820 | NA | Spain | 2023-March-7 | EPI_ISL_17190000 | A/Catalonia/NSAV198269034/2023 | Hospital Universitari Arnau del Vilanova | Hospital Universitari Vall d'Hebron | Andres, C.; Gonzalez-Sanchez, A; Pinana, M; Martin,MC.; Codina, MG.; Pumarola, T; Anton, A. |
| EPI2412687 | HA | Brazil | 2022-December-11 | EPI_ISL_16954869 | A/Mato Grosso do Sul/IAL/C10384/2022 | Instituto Adolfo Lutz National Influenza Centre | Instituto Adolfo Lutz | Sacchi, C.T.; Campos, K.R.; Amarante, A.F.; Santos, M.B.N.; Santos, K.C.O.; Benega, M.A. |
| EPI2412686 | NA | Brazil | 2022-December-11 | EPI_ISL_16954869 | A/Mato Grosso do Sul/IAL/C10384/2022 | Instituto Adolfo Lutz National Influenza Centre | Instituto Adolfo Lutz | Sacchi, C.T.; Campos, K.R.; Amarante, A.F.; Santos, M.B.N.; Santos, K.C.O.; Benega, M.A. |
| EPI12350129 | HA | Netherlands | 2023-January-13 | EPI_ISL_16807930 | A/Netherlands/00390/2023 | Erasmus Medical Center | Erasmus Medical Center | |
| EPI12350128 | NA | Netherlands | 2023-January-13 | EPI_ISL_16807930 | A/Netherlands/00390/2023 | Erasmus Medical Center | Erasmus Medical Center | |
| EPI1647563 | HA | Australia | 2019-November-22 | EPI_ISL_401903 | A/Victoria/2570/2019 | Alfred Hospital | WHO Collaborating Centre for Reference and Research on Influenza | Deng, Y-M; Iannello, P; Lau, H; Todd, A; Spirason, N; Moselen, J; Komadina, N |
| EPI1647562 | NA | Australia | 2019-November-22 | EPI_ISL_401903 | A/Victoria/2570/2019 | Alfred Hospital | WHO Collaborating Centre for Reference and Research on Influenza | Deng, Y-M; Iannello, P; Lau, H; Todd, A; Spirason, N; Moselen, J; Komadina, N |
| EPI2136122 | HA | United Arab Emerites | 2022-July-14 | EPI_ISL_14727205 | A/Abu Dhabi/0000594/2022 | Shaikh Khalifa Medical City | CDC | CDC |
| EPI2136121 | NA | United Arab Emerites | 2022-July-14 | EPI_ISL_14727205 | A/Abu Dhabi/0000594/2022 | Shaikh Khalifa Medical City | CDC | CDC |
| EPI2437894 | HA | Netherlands | 2023-February-13 | EPI_ISL17076101 | A/Netherlands/10368/2023 | RIVM | RIVM | A, Meijer; L, Wijsman; S, vd Brink |
| EPI2437893 | NA | Netherlands | 2023-February-13 | EPI_ISL17076101 | A/Netherlands/10368/2023 | RIVM | RIVM | A, Meijer; L, Wijsman; S, vd Brink |
| EPI2474456 | HA | Norway | 2023-February-20 | EPI_ISL_17266679 | A/Norway/03170/2023 | Molde Hospital Laboratory for Medical Microbiology | Norwegian Institute of Public Health | Bragstad, K; Hunnes, O; Madsen, MP; Rohringer, A; Riis, R |
| EPI2474455 | NA | Norway | 2023-February-20 | EPI_ISL_17266679 | A/Norway/03170/2023 | Molde Hospital Laboratory for Medical Microbiology | Norwegian Institute of Public Health | Bragstad, K; Hunnes, O; Madsen, MP; Rohringer, A; Riis, R |
| EPI2414287 | HA | Pakistan | 2023-February-18 | EPI_ISL_16972586 | A/Pakistan/GIHSN/ICT/969/2023 | NIH Pakistan | NIH Pakistan | Nazish, Bader; Aemer,Ikram; Muhammad, Salman; HamzaAhmad, Mirza; Abdul, Ahad |
| EPI2414291 | NA | Pakistan | 2023-February-18 | EPI_ISL_16972586 | A/Pakistan/GIHSN/ICT/969/2023 | NIH Pakistan | NIH Pakistan | Nazish, Bader; Aemer,Ikram; Muhammad, Salman; HamzaAhmad, Mirza; Abdul, Ahad |
| EPI2467732 | HA | Japan | 2022-December-9 | EPI_ISL_17244493 | A/Okinawa/194/2022 | Okinawa Prefectural Institute of Health and Environment | NIID | Takashita, Emi; Fujisaki, Seiichiro; Watanabe, Shinji; Hasegawa, Hideki |
| EPI2467731 | NA | Japan | 2022-December-9 | EPI_ISL_17244493 | A/Okinawa/194/2022 | Okinawa Prefectural Institute of Health and Environment | NIID | Takashita, Emi; Fujisaki, Seiichiro; Watanabe, Shinji; Hasegawa, Hideki |
| EPI2030270 | HA | Australia | 2022-March-21 | EPI_ISL_12579872 | A/Victoria/50/2022 | Dorevtich Pathology | WHO Collaborating Centre for Reference and Research on Influenza | Deng, Y-M; Iannello, P; Lau, H; Todd, A; Spirason, N; Moselen, J; Komadina, N |
| EPI2030269 | NA | Australia | 2022-March-21 | EPI_ISL_12579872 | A/Victoria/50/2022 | Dorevtich Pathology | WHO Collaborating Centre for Reference and Research on Influenza | Deng, Y-M; Iannello, P; Lau, H; Todd, A; Spirason, N; Moselen, J; Komadina, N |
| EPI1885402 | HA | Australia | 2021-March-16 | EPI_ISL_3534319 | A/Darwin/6/2021 | WHO Collaborating Centre for Reference and Research on Influenza | CDC | |
| EPI1885401 | NA | Australia | 2021-March-16 | EPI_ISL_3534319 | A/Darwin/6/2021 | WHO Collaborating Centre for Reference and Research on Influenza | CDC | |
| EPI1843589 | HA | Cambodia | 2020-July-16 | EPI_ISL_944639 | A/Cambodia/e0826360/2020 | Institute Pasteur du Cambodia | WHO Collaborating Centre for Reference and Research on Influenza | Deng, Y-M; Iannello, P; Lau, H; Todd, A; Spirason, N; Moselen, J; Komadina, N |
| EPI1843588 | NA | Cambodia | 2020-July-16 | EPI_ISL_944639 | A/Cambodia/e0826360/2020 | Institute Pasteur du Cambodia | WHO Collaborating Centre for Reference and Research on Influenza | Deng, Y-M; Iannello, P; Lau, H; Todd, A; Spirason, N; Moselen, J; Komadina, N |
| EPI2293149 | HA | UK | 2023-January-3 | EPI_ISL_16527800 | A/Barry/2781/2023 | Public Health Wales Microbiology Cardiff | Public Health Wales Microbiology Cardiff | Public Health Wales Microbiology Cardiff |
| EPI2293151 | NA | UK | 2023-January-3 | EPI_ISL_16527800 | A/Barry/2781/2023 | Public Health Wales Microbiology Cardiff | Public Health Wales Microbiology Cardiff | Public Health Wales Microbiology Cardiff |
| EPI2213567 | HA | UK | 2022-October-22 | EPI_ISL_15727347 | A/Caerphilly/7012/2022 | Public Health Wales Microbiology Cardiff | Public Health Wales Microbiology Cardiff | Public Health Wales Microbiology Cardiff |
| EPI2213569 | NA | UK | 2022-October-22 | EPI_ISL_15727347 | A/Caerphilly/7012/2022 | Public Health Wales Microbiology Cardiff | Public Health Wales Microbiology Cardiff | Public Health Wales Microbiology Cardiff |
| EPI2468282 | HA | Japan | 2023-February-13 | EPI_ISL_17244658 | A/Kanagawa/AC2206/2023 | NIID | NIID | |
| EPI2468281 | NA | Japan | 2023-February-13 | EPI_ISL_17244658 | A/Kanagawa/AC2206/2023 | NIID | NIID | |
| EPI2341379 | HA | Tunisia | 2022-December-14 | EPI_ISL_16759949 | A/Tunisia/23298/2022 | National Influenza Centre-Tunis | Crick Worldwide Influenza Centre | |
| EPI2359242 | NA | Tunisia | 2022-December-14 | EPI_ISL_16759949 | A/Tunisia/23298/2022 | National Influenza Centre-Tunis | Crick Worldwide Influenza Centre | |
| EPI2392074 | HA | China | 2022-December-11 | EPI_ISL_16873553 | A/Hebei-Shuangqiao/11248/2022 | WHO Chinese National Influenza Center | WHO Chinese National Influenza Center | Wang Dayan. Li Xiyan, Wang Wijuan, Zeng Xiaoxu |
| EPI2392073 | NA | China | 2022-December-11 | EPI_ISL_16873553 | A/Hebei-Shuangqiao/11248/2022 | WHO Chinese National Influenza Center | WHO Chinese National Influenza Center | Wang Dayan. Li Xiyan, Wang Wijuan, Zeng Xiaoxu |

| | | | | | | | |
|---|---|---|---|---|---|---|---|
| EPI2440114 | HA | Spain | 2023-February-3 | EPI_ISL_17092318 | A/Andalucia/PMC-00001/2023 | Servicia de Microbiologia Hospital Universitario Virgen del Rocio | Fundacion Progresso y Salud |
| EPI2440115 | NA | Spain | 2023-February-3 | EPI_ISL_17092318 | A/Andalucia/PMC-00001/2023 | Servicia de Microbiologia Hospital Universitario Virgen del Rocio | Fundacion Progresso y Salud |
| EPI2047284 | HA | United Kingdom | 2022-Apr-15 | EPI_ISL_12943536 | A/Northern Ireland/V22014094/2022 | Regional Virus Laboratory, Microbiology Department, Royal Victoria Hospital | Crick Worldwide Influenza Centre |
| EPI2047285 | NA | United Kingdom | 2022-Apr-15 | EPI_ISL_12943536 | A/Northern Ireland/V22014094/2022 | Regional Virus Laboratory, Microbiology Department, Royal Victoria Hospital | Crick Worldwide Influenza Centre |
| EPI2047286 | HA | United Kingdom | 2022-Apr-25 | EPI_ISL_12943540 | A/Northern Ireland/V22015088/2022 | Regional Virus Laboratory, Microbiology Department, Royal Victoria Hospital | Crick Worldwide Influenza Centre |
| EPI2047287 | NA | United Kingdom | 2022-Apr-25 | EPI_ISL_12943540 | A/Northern Ireland/V22015088/2022 | Regional Virus Laboratory, Microbiology Department, Royal Victoria Hospital | Crick Worldwide Influenza Centre |
| EPI2047288 | HA | United Kingdom | 2022-Apr-22 | EPI_ISL_12943544 | A/Northern Ireland/V32290637/2022 | Regional Virus Laboratory, Microbiology Department, Royal Victoria Hospital | Crick Worldwide Influenza Centre |
| EPI2047289 | NA | United Kingdom | 2022-Apr-22 | EPI_ISL_12943544 | A/Northern Ireland/V32290637/2022 | Regional Virus Laboratory, Microbiology Department, Royal Victoria Hospital | Crick Worldwide Influenza Centre |
| EPI2050519 | HA | United Kingdom | 2022-Mar-31 | EPI_ISL_13017101 | A/Northern Ireland/V22012133/2022 | Regional Virus Laboratory, Microbiology Department, Royal Victoria Hospital | Crick Worldwide Influenza Centre |
| EPI2050520 | NA | United Kingdom | 2022-Mar-31 | EPI_ISL_13017101 | A/Northern Ireland/V22012133/2022 | Regional Virus Laboratory, Microbiology Department, Royal Victoria Hospital | Crick Worldwide Influenza Centre |
| EPI2050521 | HA | United Kingdom | 2022-Apr-04 | EPI_ISL_13017102 | A/Northern Ireland/V22012462/2022 | Regional Virus Laboratory, Microbiology Department, Royal Victoria Hospital | Crick Worldwide Influenza Centre |
| EPI2050522 | NA | United Kingdom | 2022-Apr-04 | EPI_ISL_13017102 | A/Northern Ireland/V22012462/2022 | Regional Virus Laboratory, Microbiology Department, Royal Victoria Hospital | Crick Worldwide Influenza Centre |
| EPI2050524 | HA | United Kingdom | 2022-Apr-01 | EPI_ISL_13017104 | A/Northern Ireland/V32272606/2022 | Regional Virus Laboratory, Microbiology Department, Royal Victoria Hospital | Crick Worldwide Influenza Centre |
| EPI2050525 | NA | United Kingdom | 2022-Apr-01 | EPI_ISL_13017104 | A/Northern Ireland/V32272606/2022 | Regional Virus Laboratory, Microbiology Department, Royal Victoria Hospital | Crick Worldwide Influenza Centre |
| EPI2229355 | HA | United Kingdom | 2022-Oct-28 | EPI_ISL_15937899 | A/Northern Ireland/V32409910 /2022 | Regional Virus Laboratory, Microbiology Department, Royal Victoria Hospital | Crick Worldwide Influenza Centre |
| EPI2229356 | NA | United Kingdom | 2022-Oct-28 | EPI_ISL_15937899 | A/Northern Ireland/V32409910 /2022 | Regional Virus Laboratory, Microbiology Department, Royal Victoria Hospital | Crick Worldwide Influenza Centre |
| EPI2229357 | HA | United Kingdom | 2022-Oct-25 | EPI_ISL_15937900 | A/Northern Ireland/V22039362 /2022 | Regional Virus Laboratory, Microbiology Department, Royal Victoria Hospital | Crick Worldwide Influenza Centre |
| EPI2229358 | NA | United Kingdom | 2022-Oct-25 | EPI_ISL_15937900 | A/Northern Ireland/V22039362 /2022 | Regional Virus Laboratory, Microbiology Department, Royal Victoria Hospital | Crick Worldwide Influenza Centre |
