## Supplementary Figures and Tables for "‘One Health’ Genomic Surveillance of Avian and Human Influenza A Viruses Through Environmental Wastewater Monitoring"

**Table S1. Description of PCR primers and probe used for screening wastewater and sequencing IAV positive samples.**

| Purpose | Primer name | Target | Sequence (5' - 3') | Position | Final reaction concentration (µM) | Reference |
|---|---|---|---|---|---|---|
| IAV Screening | SVIP-MP-F | MP-segment | GGCCCCCTCAAAGCCGA | 77–93 | 1.4 | *Nagy et al., 2021* |
| IAV Screening | SVIP-MP-R | MP-segment | CGTCTACGYTGCAGTCC | 258–242 | 1.4 | |
| IAV Screening | SVIP-MP_P2-MGB | MP-segment | TCACTKGGCACGGTGAGCGT | 237–218 | 0.4 | |
| Genomic Amplification | MBTuni-12 | 3' vRNA Terminus | ACGCGTGATCAGCAAAAGCAGG | | 0.2 | *Zhou et al., 2009* |
| Genomic Amplification | MBTuni-13 | 5' vRNA Terminus | ACGCGTGATCAGTAGAAACAAGG | | 0.2 | |

**Table S2 Thermocycling conditions used for SVIP-MPv2 and MBTuni PCRs.**

| Thermocycling conditions | SVIP-MPv2 (IAV Screening Assay) | | |
|---|---|---|---|
| | **Temperature (°C)** | **Time (mins:secs)** | **Number of cycles** |
| Reverse Transcription (RT) | 50 | 10:00 | |
| RT Denaturation | 95 | 10:00 | |
| Denaturation | 95 | 00:10 | 45 |
| Annealing/Extension | 60 | 00.30 | |
| Hold | 40 | ∞ | |
| **Thermocycling conditions** | **MBTuni (Amplicon Generation for Sequencing)** | | |
| | **Temperature (°C)** | **Time (hr:mins:secs)** | **Number of cycles** |
| Reverse Transcription (RT) | 42 | 1:00:00 | |
| RT Denaturation | 94 | 00:02:00 | |
| Denaturation | 94 | 00:00:30 | 5 |
| Annealing | 45 | 00:00:30 | |
| Extension | 68 | 00:03:00 | |
| Denaturation | 94 | 00:00:30 | 35 |
| Annealing | 57 | 00:00:30 | |
| Extension | 68 | 00:03:00 | |
| Final extension | 68 | 00:02:00 | |
| Hold | 4 | ∞ | |

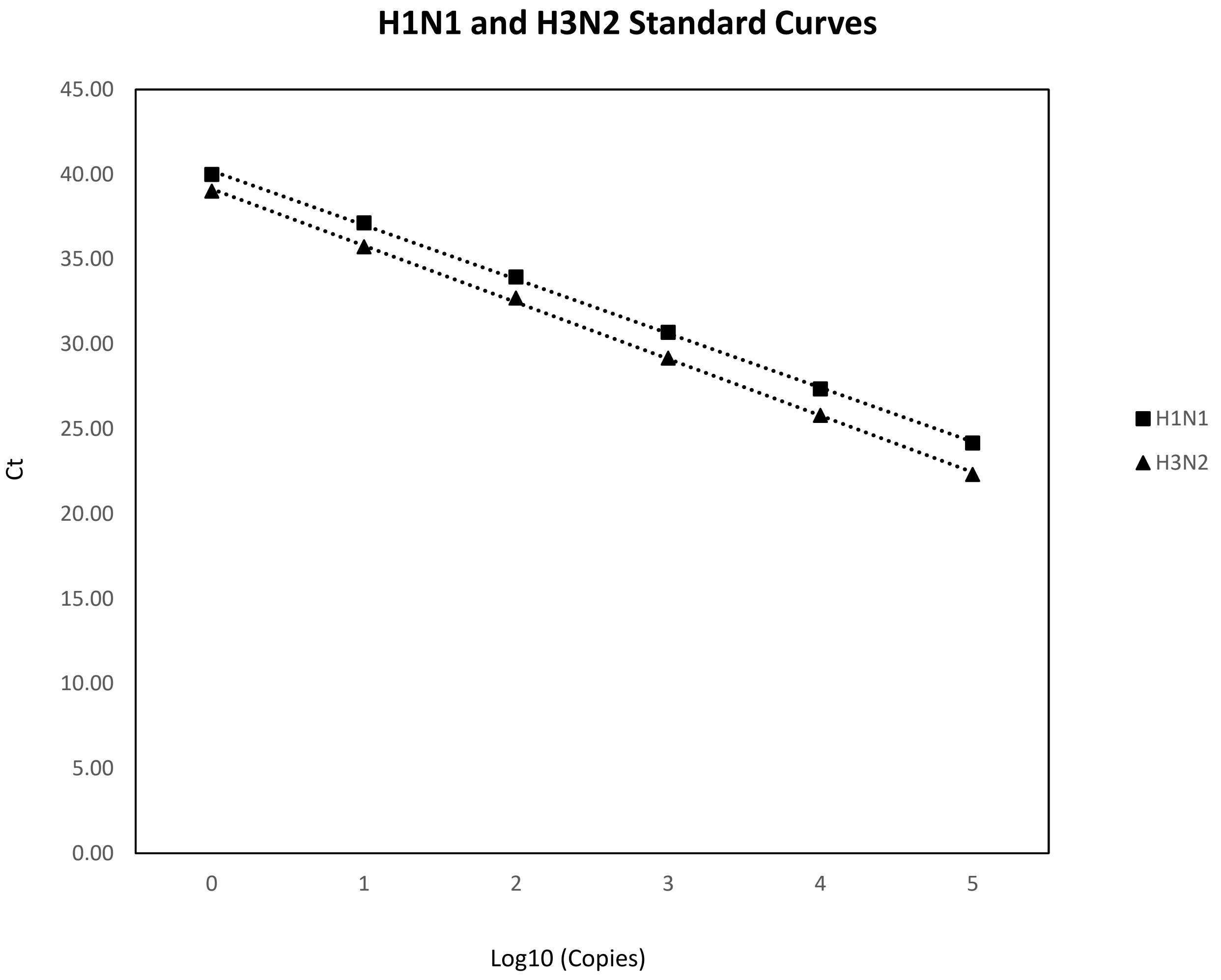


**Figure S1. Standard curve for IAV MP using synthetic H1N1 and H3N2 RNA standards. Amplification efficiency was 105% and 99.3% respectively with R2 >0.999 for both. The LOD95% for the SVIP-MPv2 assay using these standards was approximately 1.5 copies per**

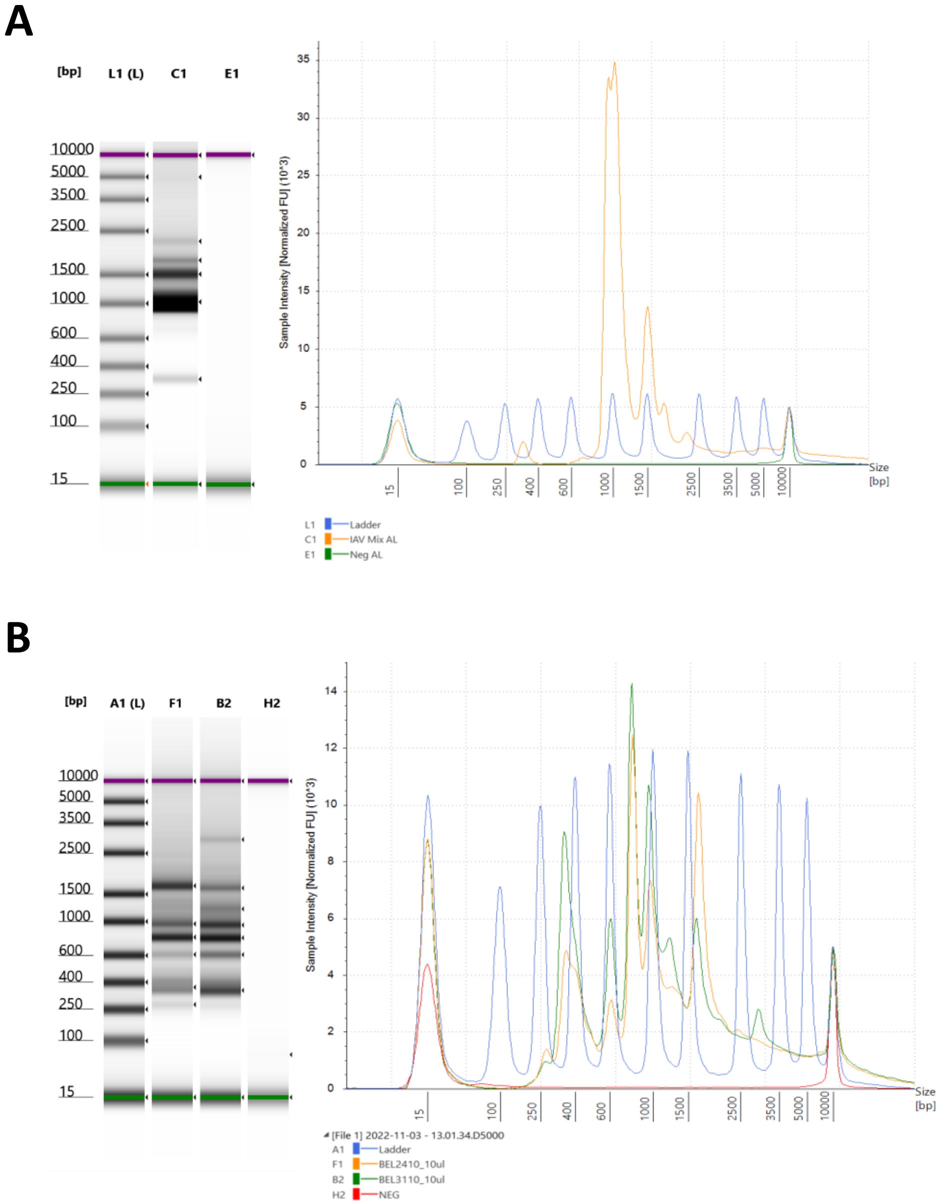


**Figure S2 TapeStation gels and composite electropherograms of purified amplicon mix post multi-segment RT-PCR. (A) Mixed IAV positive control (pH1N1, H1N1, H1N2, H3N2 and H6N1). B) Representative example of wastewater samples: Belfast 24/10/2022 and Belfast 31/10/2022.**

**Table S3 SVIP-MPv2 assay original screening Ct versus Ct value post M-RTPCR amplicon generation for sequencing.**

| WWTW | Sample Date | Original Ct (avg.) | Sequencing QC Ct | Number Of IAV Reads |
|---|---|---|---|---|
| Belfast | 12/09/2022 | 36.7 | 13.8 | 577 |
| Belfast | 10/10/2022 | 35.5 | 17 | 131 |
| Belfast | 24/10/2022 | 36.6 | 13.6 | 1195 |
| Belfast | 31/10/2022 | 37.6 | 16 | 1440 |
| Belfast | 07/11/2022 | 37.6 | 22.5 | 15 |
| Belfast | 28/11/2022 | 35.8 | 15.95 | 671 |
| Ballymena | 12/09/2022 | 36.5 | 15.7 | 169 |
| Ballymena | 28/11/2022 | 35.1 | 13.8 | 818 |
| Craigavon | 12/09/2022 | 36.8 | 13.6 | 610 |
| Derry/Londonderry | 21/11/2022 | 35.2 | 27.4 | None |
| Enniskillen | 12/09/2022 | 36.8 | 14.5 | 624 |
| North Down | 12/09/2022 | 36.4 | 14.6 | 648 |
| North Down | 10/10/2022 | 36.1 | 16.8 | 65 |
| North Down | 17/10/2022 | 35.6 | 31.7 | 1 |
| North Down | 31/10/2022 | 37.4 | 14.7 | 96 |
| North Down | 28/11/2022 | 34.4 | 19.66 | None |

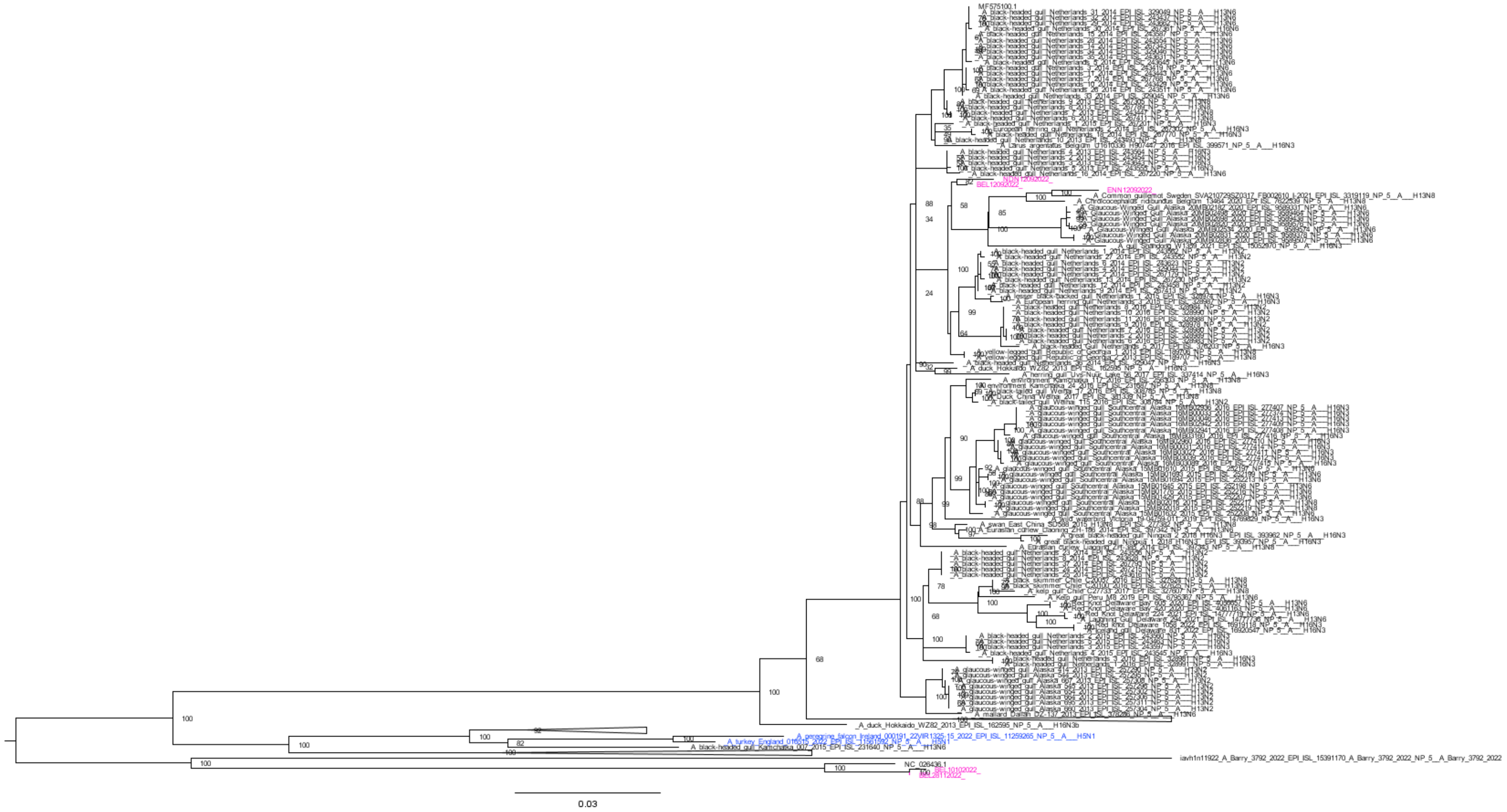


**Figure S3. Maximum-likelihood phylogeny of wastewater-derived avian-like segment 5 sequences.** Phylogenetic tree of the three avian-like segment 7 sequences (in pink; NDN12092022, BEL12092022, ENN12092022) alongside all GISAID gull H13/16-like sequences (black), and 2022 H5N1 European sequences (in blue or in collapsed clades) (accessed January 2023). Relevant bootstrap replicate values are shown on the tree. Human-like segments (BEL10102022 and BEL28112022) are also shown.

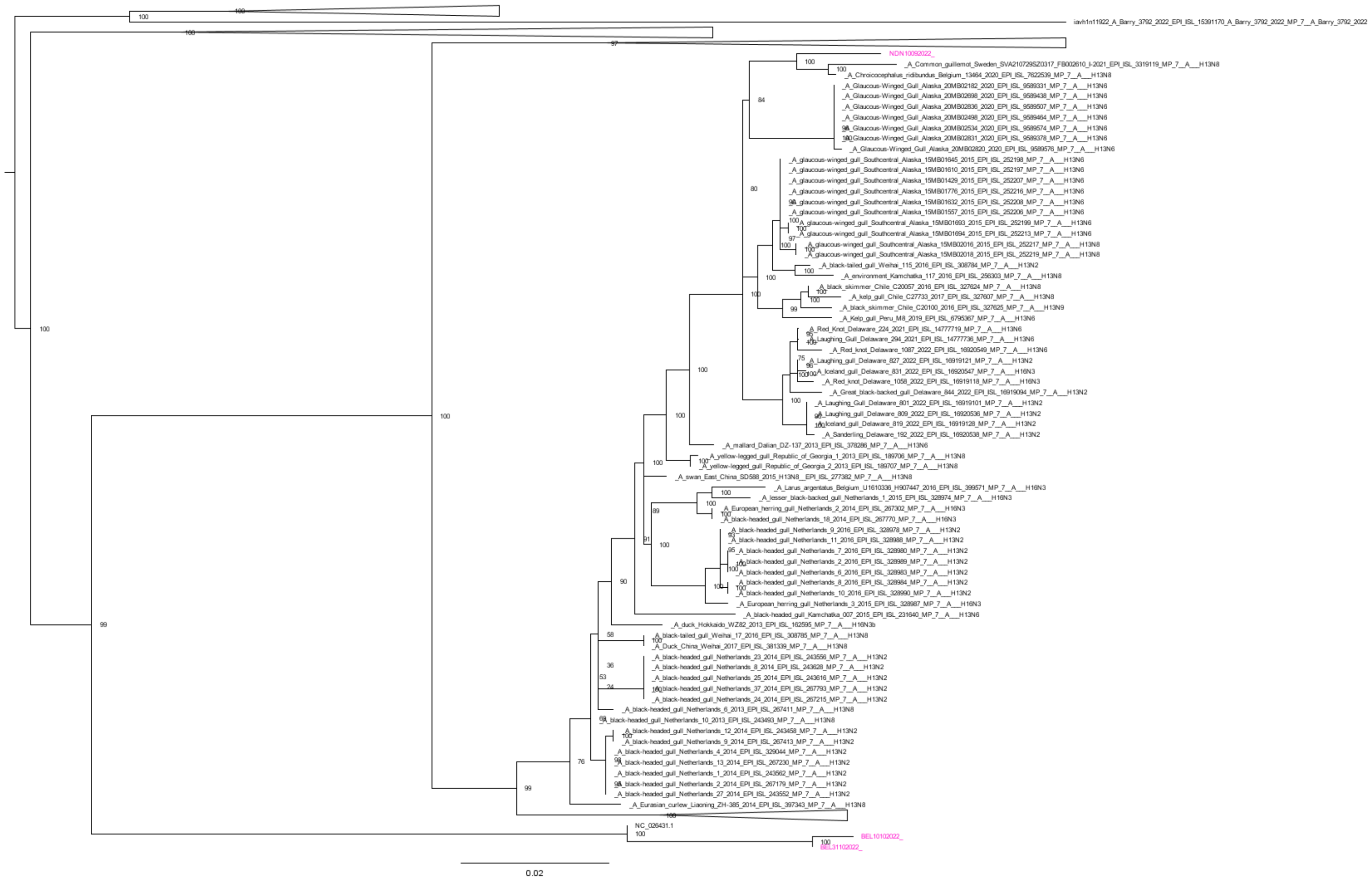


**Figure S4. Maximum-likelihood phylogeny of wastewater-derived avian-like segment 7 sequences.** Phylogenetic tree of the single avian-like segment 7 sequences (in pink; NDN10092022) alongside all GISAID gull H13/16-like sequences (black), and 2022 H5N1 European sequences (in collapsed clades) (accessed January 2023). Relevant bootstrap replicate values are shown on the tree. Human-like segments (BEL10102022 and BEL31102022) are also shown.

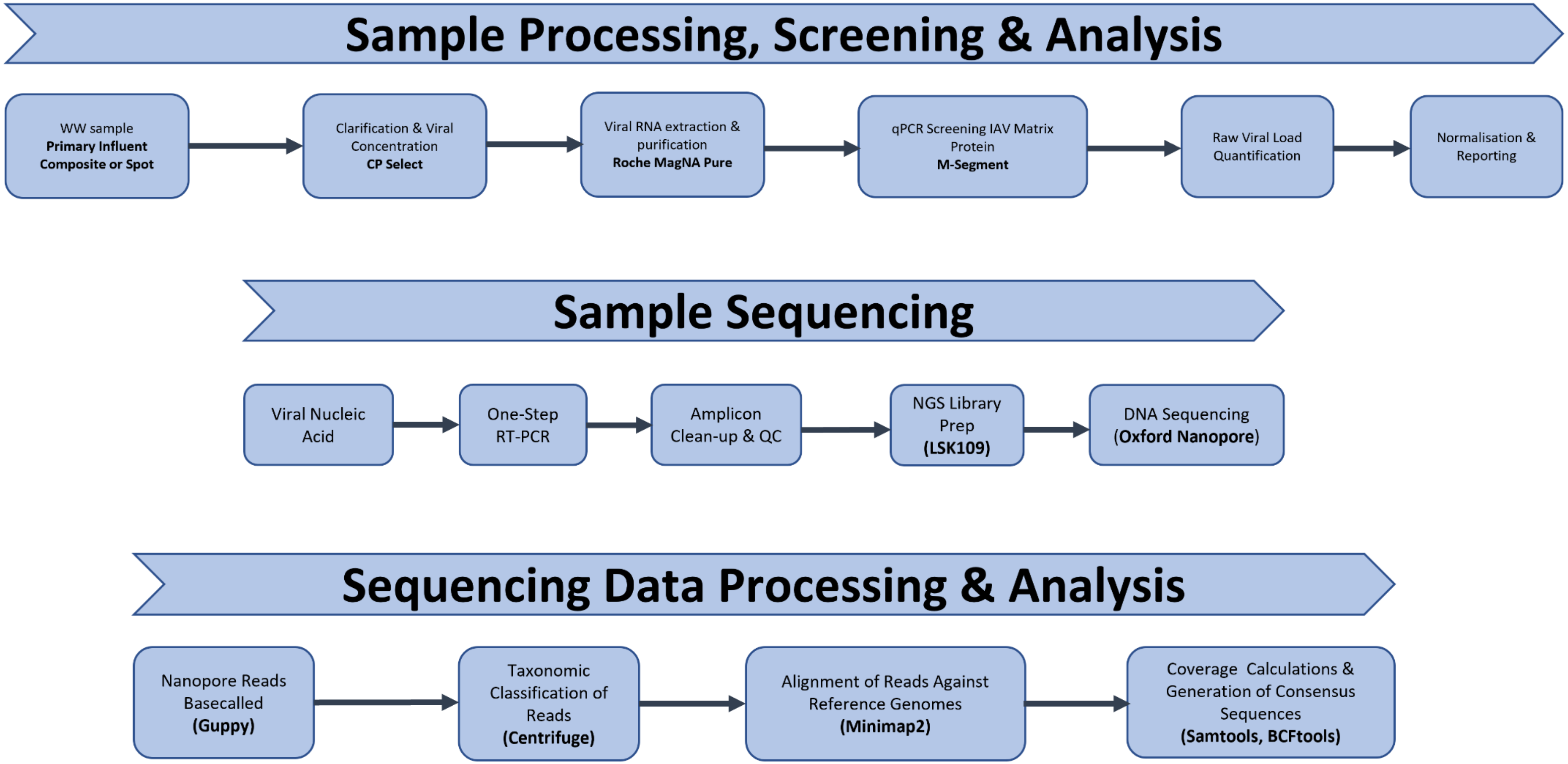


**Figure S5 Summary of workflows used.**

All generated clean consensus sequences

>NC_026438.1 Influenza A virus (A/California/07/2009(H1N1)) segment 1 polymerase PB2 (PB2) gene, complete cds
ATGGAGAGAATAAAAGAACTGAGAGATCTAATGTCGCAGTCCCGCACTCGCGAGATACTCACTAAGACCA
CTGTGGACCATATGGCCATAATCAAAAAGTACACATCAGGAAGGCAAGAGAAGAACCCCGCACTCAGAAT
GAAGTGGATGATGGCAATGAGATACCCAATTACAGCAGACAAGAGAATAATGGACATGATTCCAGAGAGG
AATGAACAAGGACAAACCCTCTGGAGCAAAACAAACGATGCTGGATCAGACCGAGTGATGGTATCACCTC
TGGCCGTAACATGGTGGAATAGGAATGGCCCAACAACAAGTACAGTTCATTACCCTAAGGTATATAAAAC
TTATTTCGAAAAGGTCGAAAGGTTGAAACATGGTACCTTCGGCCCTGTCCACTTCAGAAATCAAGTTAAA
ATAAGGAGGAGAGTTGATACAAACCCTGGCCATGCAGATCTCAGTGCCAAGGAGGCACAGGATGTGATTA
TGGAAGTTGTTTTCCCAAATGAAGTGGGGGCAAGAATACTGACATCAGAGTCACAGCTGGCAATAACAAA
AGAGAAGAAAGAAGAGCTCCAGGATTGTAAAATTGCTCCCTTGATGGTGGCGTACATGCTAGAAAGAGAA
TTGGTCCGTAAAACAAGGTTTCTCCCAGTAGCCGGCGGAACAGGCAGTGTTTATATTGAAGTGTTGCACT
TAACCCAAGGGACGTGCTGGGAGCAGATGTACACTCCAGGAGGAGAAGTGAGAAATGATGATGTTGACCA
AAGTTTGATTATCGCTGCTAGAAACATAGTAAGAAGAGCAGCAGTGTCAGCAGACCCATTAGCATCTCTC
TTGGAAATGTGCCACAGCACACAGATTGGAGGAGTAAGGATGGTGGACATCCTTAGACAGAATCCAACTG
AGGAACAAGCCGTAGACATATGCAAGGCAGCAATAGGGTTGAGGATTAGCTCATCTTTCAGTTTTGGTGG
GTTCACTTTCAAAAGGACAAGCGGATCATCAGTCAAGAAAGAAGAAGAAGTGCTAACGGGCAACCTCCAA
ACACTGAAAATAAGAGTACATGAAGGGTATGAAGAATTCACAATGGTTGGGAGAAGAGCAACAGCTATTC
TCAGAAAGGCAACCAGGAGATTGATCCAGTTGATAGTAAGCGGGAGAGACGAGCAGTCAATTGCTGAGGC
AATAATTGTGGCCATGGTATTCTCACAGGAGGATTGCATGATCAAGGCAGTTAGGGGCGATCTGAACTTT
GTCAATAGGGCAAACCAGCGACTGAACCCCATGCACCAACTCTTGAGGCATTTCCAAAAAGATGCAAAAG
TGCTTTTCCAGAACTGGGGAATTGAATCCATCGACAATGTGATGGGAATGATCGGAATACTGCCCGACAT
GACCCCAAGCACGGAGATGTCGCTGAGAGGGATAAGAGTCAGCAAAATGGGAGTAGATGAATACTCCAGC
ACGGAGAGAGTGGTAGTGAGTATTGACCGATTTTTAAGGGTTAGAGATCAAAGAGGGAACGTACTATTGT
CTCCCGAAGAAGTCAGTGAAACGCAAGGAACTGAGAAGTTGACAATAACTTATTCGTCATCAATGATGTG
GGAGATCAATGGCCCTGAGTCAGTGCTAGTCAACACTTATCAATGGATAATCAGGAACTGGGAAATTGTG
AAAATTCAATGGTCACAAGATCCCACAATGTTATACAACAAAATGGAATTTGAACCATTTCAGTCTCTTG
TCCCTAAGGCAACCAGAAGCCGGTACAGTGGATTCGTAAGGACACTGTTCCAGCAAATGCGGGATGTGCT
TGGGACATTTGACACTGTCCAAATAATAAAACTTCTCCCCTTTGCTGCTGCCCCACCAGAACAGAGTAGG
ATGCAATTTTCCTCATTGACTGTGAATGTGAGAGGATCAGGGTTGAGGATACTGGTAAGAGGCAATTCTC
CAGTATTCAATTACAACAAGGCAACCAAACGACTTACAGTTCTTGGAAAGGATGCAGGTGCATTGACTGA
AGATCCAGATGAAGGCACATCTGGGGTGGAGTCTGCTGTCCTGAGAGGATTTCTCATTTTGGGCAAAGAA
GACAAGAGATATGGCCCAGCATTAAGCATCAATGAACTGAGCAATCTTGCAAAAGGAGAGAAGGCTAATG
TGCTAATTGGGCAAGGGGACGTAGTGTTGGTAATGAAACGAAAACGGGACTCTAGCATACTTACTGACAG
CCAGACAGCGACCAAAAGAATTCGGATGGCCATCAATTAG

>MF575094.1 Influenza A virus (A/black-headed gull/Netherlands/31/2014(H13N6)) segment 1 polymerase PB2 (PB2) gene, complete cds
AGCAAAAGCAGGTCAAATATATTCAATATGGAGAGAATAAAAGAACTAAGAAACCTAATGTCACAATCTC
GCACTCGCGAGATACTGACAAAAACCACAGTAGACCATATGGCCATAATCAAGAAGTACACATCAGGGAG
ACAGGAGAAGAATCCCGCTCTCAGAATGAAGTGGATGATGGCAATGAAATATCCAATCACAGCTGACAAA
AGAATAATGGAGATGATCCCTGAGAGGAATGAACAGGGTCAGATCCTTTGGAGCAAAACTAATGATGCTG
GTTCGGATAGGGTGATGGTGTCACCTCTAGCTGTAACATGGTGGAACAGAAACGGACCAACGGCAAGTAC
GGTCCACTATCCCAAAGTCTACAGAACTTACTTTGAAAAAGTTGAGAGATTGAAACATGGAACATTCGGG
CCCGTTCATTTCCGAAATCAAGTTAAAATACGCCGTAGAGTTGACACAAATCCTGGTCACGCAGATCTCA
GTGCTAAAGAGGCACAGGACGTCATCATGGAAGTTGTATTCCCAAATGAAGTGGGGGCTAGAATACTAAC

ATCAGAGTCACAACTGACAATAACAAAGGAAAAGAAGGAGGAACTTCAGGACTGCAAAATTGCTCCCCTA
ATGGTTGCATACATGTTGGAAAGGGAACTGGTCCGAAAGACTAGATTTTTACCGGTAGCAGGCGGAACAA
GCAGCGTTTATATTGAAGTATTGCATTTAACTCAAGGAACCTGCTGGGAACAAATGTACACTCCAGGAGG
GGAAGTGAGGAATGATGATGTTGACCAGAGCTTAATCATTGCTGCAAGAAATATAGTTAGAAGAGCAACA
GTTTCAGCAGATCCATTAGCATCACTTCTAGAAATGTGCCATAGTACTCAAATAGGCGGAATAAGGATGG
TAGACATCCTTAGACAGAACCCAACAGAAGAACAAGCAGTTGACATTTGTAAGGCAGCGATGGGCCTAAG
AATCAGCTCGTCGTTCAGCTTTGGAGGATTCACCTTTAAAAGGACAAGTGGGTCATCTATCAAAAGAGAA
GAAGAAGTGCTCACGGGCAACCTCCAAACATTGAAAATACGAGTACATGAAGGATACGAGGAATTCACGA
TGGTTGGTCGGAGAGCAACTGCCATATTAAGGAAAGCAACCAGAAGGCTGATTCAGTTAATAGTGAGTGG
GAGAGATGAACAATCCATTGCTGAAGCCATAATTGTAGCGATGGTGTTTTCACAAGAAGATTGCATGATA
AAAGCTGTTCGAGGTGACTTGAATTTCGTAAACAGAGCGAATCAACGGCTGAATCCTATGCACCAGCTTC
TGAGGCACTTCCAAAAAGACGCGAAGGTGTTGTTTCAAAATTGGGGAGTTGAGCCCATTGACAACGTCAT
GGGGATGATAGGAATATTGCCTGACATGACCCCCAACACAGAAATGTCGCTGAGAGGAATAAGAGTTAGC
AAAATGGGGGTAGATGAATACTCCAGCACTGAGAGAGTGGTTGTGAGCATTGATCGCTTCCTAAGGGTTC
GAGACCAGCGGGGGAATGTGCTCTTGTCCCCTGAGGAGGTCAGTGAAACACAGGGAACAGAAAAATTGAC
AATAACGTATTCATCGTCAATGATGTGGGAGATCAACGGTCCGGAGTCAGTCCTAGTAAACACCTATCAA
TGGATCATCAGAAATTGGGAAGTCATAAAGATTCAATGGTCCCAGGATCCTACAATGTTGTACAACAAGA
TGGAATTCGAACCATTCCAATCCTTGGTACCTAAGGCTGCCAGAGGCCAATACAGTGGATTTGTAAGAAC
ATTGTTTCAACAGATGCGCGATGTGTTGGGGACTTTTGATACTGTCCAAATAATCAAATTACTACCGTTT
GCAGCAGCTCCACCGGAACAGAGTAGGATGCAATTTTCTTCTCTAACTGTGAATGTGAGAGGATCAGGGA
TGAGAATACTCGTGAGGGGTAACTCTCCAGTGTTCAACTACAACAAAACAACCAAGAGACTTACAGTTCT
CGGGAAAGATGCAGGCTCCTTAACAGAAGACCCGGATGAGGGTACAGCAGGAGTGGAGTCCGCAGTATTA
AGAGGGTTTCTAATTCTAGGCAAAGAAGACAAAAGATACGGGCCAGCATTGAGCATCAATGAACTGAGCA
ATCTTGCGAAAGGAGAAAAAGCTAATGTGCTAATAGGGCAAGGAGACGTAGTGTTGGTAATGAAACGGAA
ACGGGACTCTAGCATACTTACTGACAGTCAGACAGCGACCAAAAGAATTCGGATGGCCATCAATTAGTGT
CGAGTTGTTTAAAAACGACCTTGTTTCTACT

>iavh1n11922|A/Mute_Swan/Netherlands/2/2022|EPI_ISL_15364797|A/Mute_Swan/Netherlands/2/2022|PB2|1||A/Mute_Swan/Netherlands/2/2022
TCAAATATATTCAATATGGAGAGAATAAAAGAACTAAGAGATTTAATGTCGCAGTCTCGCACTCGCGAGA
TACTGACAAAAACCACTGTGGACCATATGGCCATAATAAAGAAATACACATCAGGAAGACAGGAGAAGAA
CCCTGCCCTCAGGATGAAGTGGATGATGGCAATGAAATATCCTATTACAGCAGACAAAAGAATAATGGAG
ATGATCCCTGAAAGGAATGAGCAAGGTCAGACTCTCTGGAGCAAAACAAATGACGCTGGATCAGACAGAG
TGATGGTGTCACCTCTGGCTGTGACATGGTGGAATAGAAATGGGCCAACGACAAGTACAGTCCATTACCC
AAAGGTTTACAAAACCTACTTTGAAAAAGTGGAAAGATTGAAACATGGAACCTTTGGCCCTGTTCACTTT
CGAAATCAAGTTAAGATACGCCGCAGGGTTGACATAAACCCGGGTCATGCAGATCTCAGTGCCAAAGAAG
CACAGGATGTCATCATGGAAGTCGTTTTCCCAAATGAAGTTGGAGCCAGGATATTGACATCAGAATCACA
ATTAACAATAACAAAGGAAAAGAAGGAGGAACTTCAGGACTGTAAGATTGCTCCTTTGATGGTGGCATAC
ATGTTGGAAAGAGAACTGGTTCGAAAAACCAGATTTTTACCAGTAGCTGGCGGAACAAGCAGCGTATATA
TCGAAGTATTGCATTTGACTCAAGGGACCTGCTGGGAACAAATGTACACACCGGGAGGGNAAGTGAGAAA
TGATGATGTTGATCAGAGTTTGATCATTGCTGCTAGAAATATCGTTAGGAGAGCAACAGTATCAGCAGAC
CCATTGGCTTCGCTCTTGGAAATGTGCCACAGTACACAAATTGGCGGGGTAAGGATGGTAGACATTCTTA
GACAGAACCCAACAGAAGAGCAAGCCGTGGATATATGCAAAGCAGCAATGGGTTTAAGAATCAGTTCATC
CTTCAGCTTTGGAGGTTTCACTTTCAAAAGAACAAGTGGATCGTCTGTCAAAAGAGAAGAAGAAGTGCTC
ACAGGCAACCTCCAAACACTGAAAATAAGAGTACATGAAGGATATGAGGAATTCACAATGGTTGGACGGA
GAGCTACAGCCATTTTAAGGAAAGCAACCAGAAGGCTGATCCAATTAATAGTGAGTGGAAGAGACGAGCA
GTCAATCGCTGAAGCAATCATAGTGGCAATGGTTTTCTCACAAGAGGACTGCATGATAAAAGCAGTACGA

GGTGATCTGAACTTTGTCAATAGAGCAAATCAGCGACTAAATCCTATGCATCAACTTCTGAGGCATTTCC
AAAAGGATGCAAAAGTGCTGTTTCAAAACTGGGGGATTGAACCAATTGACAATGTAATGGGGATGATTGG
AATACTGCCTGACATGACCCCCAGCACGGAGATGTCACTGAGAGGAGTGAGAGTCAGCAAAATGGGAGTG
GATGAATATTCCAGTACTGAGAGAGTGGTCGTGAGCATTGATCGTTTCTTGAGAGTCCGAGATCAAAGAG
GAAACGTGCTCCTGTCTCCTGAGGAAGTTAGTGAAACACAGGGAACAGAGAAACTGACGATAACATATTC
ATCGTCCATGATGTGGGAAATCAATGGTCCGGAATCCATGCTAGTCAACACATATCAATGGATCATCAGA
AATTGGGAGACTGTGAAGATTCAATGGTCCCAAGACCCTACGATGTTGTACAACAAGATGGAATTTGAGC
CCTTCCAATCCTTGGTGCCCAGGGCTGCCAGAGGCCAGTATAGTGGATTTGTGAGGACATTGTTCCAGCA
GATGCGTGATGTGCTGGGAACATTTGATACTGTCCAAATAATAAAGCTCCTGCCTTTTGCTGCAGCCCCA
CCGGAACAGAGTAGGATGCAGTTTTCCTCTCTGACTGTGAACGTAAGAGGTTCAGGAATGAGAATACTTG
TGAGAGGCAACTCCCCTGTGTTCAACTATAACAAGGCCACCAAGAGACTCACAGTTCTTGGAAAGGATGC
AGGTGCCTTGACAGAAGATCCAGATGAGGGAACAGCAGGAGTAGAGTCTGCAGTATTAAGAGGATTTCTA
ATTCTGGGCAAAGAGGACAAAAGATATGGACCAGCATTGAGCATCAACGAACTGAGCAATCTTGCGAAAG
GGGAAAAGGCTAATGTGTTGATAGGGCAAGGAGACGTGGTGTTGGTAATGAAACGGAAACGGGACTCTAG
CATACTTACTGACAGCCAGACAGCGACCAAAAGAATTCGGATGGCCATCAATTAGTGTCGAATTGTTTAA
AAACGA

>iavh1n11922|A/Barry/3792/2022|EPI_ISL_15391170|A/Barry/3792/2022|PB2|1||A/Barry/3792/2022
ATCAGCAAAAGCAGGTCAATTATATTCAGCATGGAAAGAATAAAAGAACTACGGAATCTAATGTCGCAGT
CCCGCACTCGCGAGATACTGACAAAAACCACAGTGGACCATATGGCCATAATTAAGAAGTACACATCGGG
GAGACAGGAAAAGAACCCGTCACTTAGAATGAAATGGATGATGGCAATGAAATATCCAATTACTGCTGAC
AAAAGGATAACAGAAATGGTTCCGGAGAGAAATGAACAAGGACAAACTCTATGGAGTAAAATGAGTGATG
CTGGATCAGACAGAGTGATGGTATCACCTTTGGCTGTAACATGGTGGAATAGGAATGGACCCGTGACAGA
TACGGTCCATTACCCAAAAGTGTACAAAACTTATTTCGACAAAGTCGAAAGGTTAAAACATGGAACCTTT
GGCCCAGTCCATTTTAGAAATCAAGTCAAAATACGAAGAAGAGTAGACATAAATCCTGGTCATGCAGACC
TCAGTGCTAAAGAGGCACAAGATGTAATTATGGAAGTTGTTTTTCCCAATGAAGTGGGAGCCAGAATACT
AACATCAGAATCACAGCTAACAATAACTAAAGAGAAAAAAGAAGAACTCCGAGATTGCAAAATTTCTCCC
TTGATGGTCGCATACATGCTAGAGAGAGAACTTGTGCGGAAAACAAGATTTCTCCCAGTTGCTGGCGGAA
CAAGCAGCATATACATTGAAGTTTTACATTTGACTCAAGGAACGTGTTGGGAACAAATGTACACTCCAGG
TGGAGGAGTGAGGAATGACGATGTTGACCAAAGCTTAATTATTGCGGCCAGGAACATAGTAAGAAGAGCC
GCAGTGTCAGCAGATCCACTAGCATCTTTATTGGAGATGTGCCACAGCACGCAAATTGGCGGAACAAGGA
TGGTGGACATTCTTAGACAGAACCCGACTGAAGAACAGGCTGTGGATATATGCAAAGCTGCAATGGGATT
GAGAATCAGCTCATCTTTCAGCTTTGGTGGCTTCACATTTAAAAGAACGAGCGGGTCGTCAGTCAAAAGA
GAGGAAGAGGTTCTTACAGGCAATCTCCAGACATTGAGAATAAGAGTACATGAGGGGTATGAGGAGTTCA
CAATGGTGGGGAAAAGAGCAACAGCTATACTAAGAAAAGCAACCAGAAGACTGGTTCAACTCATAGTGAG
TGGAAGAGACGAACAGTCAATAGCCGAAGCAATAATCGTGGCCATGGTGTTTTCACAAGAAGATTGCGTG
ATAAAAGCAGTTAGAGGTGACCTGAATTTTGTCAACAGAGCAAATCAGCGGTTGAACCCCATGCATCAGC
TTTTAAGGCATTTTCAGAAAGATGCGAAAGTACTCTTTCAAAATTGGGGAGTTGAACACATCGACAGTGT
GATGGGAATGGTTGGAGTATTACCAGATATGACTCCAAGCACAGAGATGTCAATGAGAGGAATAAGAGTC
AGCAAGATGGGTGTGGATGAATACTCCAGTACAGAGAGGGTGGTGGTTAGCATTGATCGGTTTTTGAGAG
TTCGAGACCAACGTGGGAATGTATTATTATCTCCTGAGGAGGTCAGTGAAACACAGGGAACTGAAAGACT
GACCATAACTTATTCATCATCGATGATGTGGGAGATTAACGGCCCTGAGTCGATTTTGGTCAATACCTAC
CAATGGATCATCAGGAATTGGGAAGCTGTCAAAATTCAATGGTCTCAGAACCCTGCAATGTTGTACAACA
AAATGGAATTTGAACCATTTCAATCTTTAGTCCCCAAGGCCACTAGAAGCCAATACAGTGGGTTTGTCAG
AACTCTATTCCAACAAATGAGAGACGTACTTGGGACATTTGACACTGCCCAGATAATAAAGCTTCTCCCT
TTTGCAGCTGCTCCACCGAAGCAAAGCAGAATGCAGTTCTCTTCACTGACTGTGAATGTGAGGGGATCAG

GGATGAGAATACTTGTAAGGGGCAATTCTCCTGTATTCAACTACAACAAGACCACTAAAAGGCTAACAAT
TCTTGGAAAAGATGCCGGCACTTTAATTGAAGACCCAGATGAAAGCACATCCGGAGTGGAGTCCGCCGTC
TTGAGAGGGTTCCTCATTATAGGTAAGGAAGACAGAAGGTACGGACCAGCATTAAGCATCAATGAACTGA
GTAACCTTGCAAAAGGGGAAAAGGCTAATGTGCTAATTGGGCAAGGAGACGTGGTGTTGGTAATGAAACG
GAAACGGGACTCTAGTATACTTACTGACAGCCAGACAGCGACCAAAAGAATTCGGATGGCCATCAATTAA
TACTGAATAGTTTAAAAACGACCTTGTTTCTACTGATC

>BEL12092022_ H1N1_PB2_1
ATGGAGAGAATAAAAGAACTAAGAGATCTAATGTCGCTGTCTCGCACTCGCGAGATACTCACCAAAACCA
CTGTGGACCACATGGCCATAATCAAAAAATACACATCGGGAAGGCAAGAGAAGAACCCCGCACTAAGGAT
GAAATGGATGATGGCAATGAAATATCCAATCACTGCAGATAAGAGAATAATGGAAATGATTCCTGAAAGG
AATGAGCAGGGACAAACCCTTTGGAGCAAAACAAACGATGCCGGATCAGACCGGGTAATGGTATCTCCTC
TCGCTGTGACGTGGTGGAACAGGAATGGACCAGCAGCAAGTACAGTCCACTACCCAAAGGTATACAAAAC
TTATTTTGAAAAGGTAGAGAGATTGAAAAACGGGACCTTTGGCCCTGTCCACTTCAGAAATCAAGTTAAG
ATAAGACGGAGGGTCGATGTAAACCCAGGCCATGCAGACCTAAGTGCCAAAGAGGCACAGGATGTAATCA
TGGAAGTTGTCTTCCCAAATGAGGTTGGAGCGAGAATATTGACGTCAGAGTCACAACTGACGGTAACAAA
GGAGAAGAAAGAAGAACTCCAGGACTGCCAAATTGCCCCTTTGATGGTTGCATATATGCTAGAGAGGGAA
TTGGTCCGGAAGACAAGGTTCCTCCCAGTGGCTGGTGGAACAAGCAGTGTCTATATTGAGGTGCTACACT
TAACCCAGGGGACATGCTGGGAGCAGATGTATACTCCAGGAGGAGAGGTGAGAAATGATGATGTGGACCA
GAGCTTGATTATTGCTGCCAGGAACATAGTGAGAAGAGCAACAGTATCAGCAGACCCACTAGCATCTCTA
CTGGAAATGTGCCACAGCACACAGATTGGAGGAGTAAGGATGGTCGACATCCTTCGACAAAATCCAACAG
AAGAACAAGCCGTGGACATATGCAAGGCAGCAATGGGATTAAGGATTAGCTCATCTTTCAGTTTTGGTGG
ATTCACTTTTAAAAGAACAAGTGGATCTTCAGTAAAAAGAGAAGAAGAAGTGCTTACGGGCAACCTCAAA
ACACTGAAAATAAGAGTGCATGAGGGGTATGAAGAGTTTACAATGGTAGGAAGACGAGCAACTGCTATCA
TCAGGAAAGCAACCAGGAGGCTGATCCAGCTAATAGTAAGCGGGAGGGACGAGCAGTCAATTGCTGAAGC
AATAATTGTGGCCATGGTGTTCTCACAGGAGGATGGTATGATCAAGGCAGTTCGAGGTGATCTGAATTTT
GTCAATAGGGCGAACTAGAGGCTGAACCCAATGCATCAACTCCTAAGACACTTTCAAAAGGATGCAAAAA
TACTTTTCAAAAACTGGGGGATCGAACCCATTGACAGTGTGATGGGGATGATTGGGATATTGCCCGACAT
GACCCCAAGTACTGAGATGTCGCTGAGGGGGATAAGAGTCAGTAAGACAGGAGTAGATGAATATTCCAGC
ACAGAAAGGGTCGTAGTGAGCATCGACCGATTTTTAAGAGTTCGAGACCAAAGAGGGAACATACTACTGT
CACCTGAAGAAGTCAGCGAGACACAGGGAACAGAGAAATTGACAATCACCTATTCGTCGTCAATGATGTG
GGAGATCAATGGCCTTGAGTCGGTGTTGATAAATACTTATCAGTGGATCATCAGAAACTGGGAAACTGTG
AAAATCCTATGGTCACAGGACCCCACAATGTTATATAACAAGATGGAATTCGAACCATTCCAATCTCTTG
TCCCTAGGGCAGCTAGAGGTCAATACAGTGGATTCGTGAGGACATTATTCCAGCAGATGCGGGATGTGCT
TGGAACATTTGACACTGTTCAGATAATAAAACTCCTCCCATTTGCTGCTGCCCCACCAGAACAAAGTAAG
ATGCAGTTCTCTTCCATGACTGTAAATGTAAGAGGATCAGGAATGAGAATACTGGTAAGAGGCAACTCCC
TGGTGTTCAATTACAATAAGGCCACCAAGAGGCTCACTATTCTCGGGAAAGATGCAGGTGCATTGACCGA
AGATCCAGATGAAGGCACCGCTGGAATAGAGTCTGCTGTTTTAAGAGGATTCCTCATTTTGGGCAAAGAA
GACAAGAGATACGGCCCTGCATTGAGCATCAACGAGCTGAGCAATCTTGCAAAGGGAGGGAAGGCTAATG
TGCTAATTGGGCAAGGAGACGTAGTGTTGGTAATGAAACGGAAACGGGACTTTAGCATACTTACTGACAG
TCAGACAGCGACCAAAAGAATTCGGATGGCCATCAATTAG
>BYM12092022_ H1N1_PB2_1
ATGGAGAGAATAAAAGAACTAAGAGATCTAATGTCACAGTCTCGCACTCGCGAGATACTCACTAAGACCA
CTGTGGACCACATGGCCATAATTAAGAAATACACATCAGGAAGGCAAGAGAAGAACCCCGCACTAAGGAT
GAAATGGATGATGGCAATGAAATACCCAATCACCGCAGATAAGAGAATAATGGAAATGATTCCAGAAAGG
AATGAGCAGGGACAAACCCTTTGGAGTAAAACAAACGATGCCGGATCGGACCGAGTAATGGTATCGCCTC

TCGCCGTGACATGGTGGAACAGGAATGGACCAACAGCAAATACAGTCCACTACCCAAAGGTATACAAAAC
TTATTTTGAAAAAGTTGAGAGATTGAAACACGGGACCTTTGGCCCTGTCCACTTCAGAAATCAAGTTAAG
ATAAGACGGAGGGTTGATGTAAACCCAGGCCATGCAGACCTAAGTGCCAAAGAGGCACGGGATGTAATCA
TGGAAGTTGTCTTCCCAAACGAGGTTGGGGCAAGAATACTGACGTCAGAGTCACAACTGACAGTAACAAA
GGAGAAAAAAGAAGAGCTCCAGGACTGCAAAATTGCCCCTTTGATGGTGGCATATATGCTAGAGAGGGAA
TTGGTCCGGAAGACAAGGTTCCTTCCAGTGGCTGGTGGAACAAGCAGTGTCTATATTGAGGTGCTGCATT
TAACCCAGGGGACATGCTGGGAGCAGATGTACACTCCAGGAGGAGAGGTGAGGAACGATGATGTAGACCA
GAGCTTGATTATAGCTGCCAGAAACATAGTGAGAAGAGCAACAGTATCAGCAGACCCACTAGCATCCCTG
CTGGAAATGTGCCACAGCACACAGATTGGAGGGGTAAGGATGGTCGACATCCTTAGACAAAATCCAACAG
AAGAACAAGCAGTGGACATATGCAAGGCAGCAATGGGGTTAAGGATTAGCTCATCTTTCAGCTTTGGTGG
ATTCACTTTTAAAAGGACAAGTGGATCTTCAGTCAAGAAAGAAGAAGAAGTGCTTACGGGCAACCTTCAA
ACATTGAAAATAAGAGTGCATGAGGGGTATGAAGAGTTCACAATGGTAGGAAGACGAGCAACTGCTATTC
TCAGGAAGGCAACCAGGAGGCTGATCCAGCTAATAGTAAGTGGGAGAGACGAGCAGTCAATTGCTGAAGC
AATAATTGTGGCCATGGTGTTCTCACAGGAGGACTGCATGATCAAGGCAGTTCGAGGTGATCTGAATTTT
GTCAATAGAGCGAACCAGAGGCTGAACCCAATGCATCAGCTCCTAAGACATTTTCAAAAGGATGCAAAAG
TGCTTTTTCAAAACTGGGGAATCGAACCCATTGACAGTGTGATGGGGATGATTGGGATAGTGCCCGGCAC
GGCCCCAAGTACTGAGATGTCGCTGAGGGGGATAAGAGTCAGCAAGACGGGAGTAGATGAATACTCCAGC
ACAGAAAGGGTCGTAGTGAGCATCGACCGATTCTTAAGAGTTCGAGACCAAAGAGGGAACATACTATTGT
CACCTGAGGAAGTCAGCGAGACACAGGGAACAGAGAAATTGACAATCACTTATTCGTCGTCAATGATGTG
GGAAATCAATGGTCCTGAGTCAGTGTTAGTCAATACTTATCAGTGGATCATCAGAAACTGGGAAACTGTG
AAAATTCAATGGTCACAGGATCCCACAGTATTATATAACAAGATGGAATTCGAACCATTCCAATCTCTTG
TCCCTAAGGCAGCTAGGGGTCAATACAGTGGATTCGTGAGGACTTTATTCCAGCAAATGCGGGATGTGCT
TGGAACATTTGACACTGTTCAGATAATAAAACTCCTCCCATTTGCTGCTGCCCCGCCAGAACAAAGTAGG
ATGGAATTCTCCTCCATGACTGTAAATGTAAGAGGGTCAGTGATGAGAATACTGGTGAGAGGCAACTCTC
CAGTATTCAATTACAATAAGGCCACCAAACGACTTACAGTTCTCGGGAAAGATGCAGGTGCATTGACCGA
AGATTCAGATGAAGGCACAGCTTGAATAGAGTCTGCTGTTTCAAGAGGATTCCTCATTTTGGGCAAAGAA
GACAAGAGATATGGCCCGGCATTGAGCATCAATGAGCTGAGCAATCTTGCAAAGGGAGAGAAGGCTAATG
TGCTAATTGGGCAAGGAGACGTAGTGTTGGTAATGAAACGGAAACGGGACTCTAGCATACTTACTGACAG
TCAGACAGCGACCAAAAGAATTCGGATGGCCATCAATTAG
>CRG12092022_ H1N1_PB2_1
NNNNNNNNNATAAAAGAACTAAGAGATCTAATGTCACAGTCTCGCACTCGCGAGATACTCACCAAGACCA
CTGTGGACCACATGGCCATAATTAAGAAATACACATCGGGAAGGCAAGAGAAGAACCCCGCACTCAGAAT
GAAATGGATGATGGCAATGAAATATCCAATCACCGCAGATAAGAGAATAATGGAAATGATTCCTGAAAGG
AATGAGCAGGGACAAACCCTCTGGTGCAAAACAAACGATGCCTGATCAGATCGAGTGACGGCATCGCCTC
TCGCTGCGACGTGGTGGAACAGGAATGGACCAACAGCAAATACAGTCCACTACCCAAAGGTATAAAAAAC
TTATTTTAAAAAAGTTGAGAGATTGAGACACGGTACCTTCGGACCTGTCCACTTCAGAAATCAAGTTAAG
ATAAGAAGGAGGGTCGATGTAAACCCAGGCCATGCAGATCTCAGTGCCAAGGAGGCACAGGATGTAATCA
TGGAAGTTGTCTTCCCAAACGAGGTTGGGGCGAGAATACTGACGTCAGAGTCACAACTGACAGTAACAAA
GGAGAAAAGAGAGGAACTCCAGGATTGCAAAATTGCTCCCTTGATGGTGGCGTATATGCTAGAAAGAGAA
TTGGTCCGGAAGACAAGGTTTCTTCCAGTGGCTGGTGGAACAAGCAGTGTCTATATTGAGGTGCTGCACT
TAACCCAGGGAACATGCTGGGAGCAGATGTACACTCCAGGAGGAGAGGTGAGAAACGATGATGTAGACCA
GAGCTTGATTATAGCTGCCAGAAACATAGTGAGAAGAGCAACAGTATCAGCAGACCCACTAGCATCCCTC
CTGGAAATGTGCCACAGCACACAGATTGGAGGGGTAAGGATGGTCGACATCCTTCGACAAAATCCAACAG
AAGAACAAGCAGTGGACATATGCAAGGCAGCAATGGGGTTAAGGATTAGCTCATCTTTCAGCTTTGGTGG
ATTCACTTTTAAAAGAACAAGTGGATCTTCAGTAAAAAGAGAAGAAGAAGTGCTAACGGGCAACCTTCAA
ACATTGAAAATAAGAGTGCATGAGGGGTATGAAGAGTTCACAATGGTAGGAAGATGAGCAACTGCCATTA

TCAGGAAGGCAACCAGGAGGCTGATCCAGCTAATAGTAAGTGGGAGAGACGAGCAGTCAATTGCTGAAGC
AATAATTGTGGCCATGGTGTTCTCACAGGAGGACTGCATGATCAAGGCAGTTCGAGGTGATCTGAATTTT
GTCAATAGGGCGAACCAGAGGCTGAACCCAATGCATCAACTCCTAAGACATTTTCAAAAGGATGCAAAAG
TGCTTTTTCAAAACTGGGGAATCGAACCCATTGACAGTGTGATGGGGATGATCGGGATATTGCCCGACAT
GACCCCAAGTACTGAGATGTCGCTGAGAGGGATAAGAGTCAGCAAGACGGGAGTAGATGAATACTCCAGC
ACAGAAAGGGTGGTAGTGAGCATCGACCGATTCTTAAGAGTTCGAGACCAAAGAGGGAACATACTATTGT
CACCTGAAGAAGTCAGCGAGACACAAGGAACAGAGAAATTGACAATCACTTATTCGTCATCAATGATGTG
GGAGATCAATGGTCCTGAGTCGGTGTTAGTCAACACATATCATTGGATAATCAGGAACTGGGAAATTGTG
AAAATCCAATGGTCACAAGATCCCACAATGTTATATAACAAAATGGAATTCGAACCATTCCAATCTCTTG
TCCCTAAGGCAGCTAGGGGTCAATACAGTGGATTCGTGAGGACATTATTCCAGCAAATGCGGGATGTGCT
TGGAACATTTGACACTGTTCAGATAATTAAACTTCTCCCATTTGCTGCTGCCCTGCCAGAACAAAGTAGG
ATGCAGTTTTCCTCAATGACTGTAAATGTAAGAGGATCAGGGATGAGAATACTGGTGAGAGGCAACTCTC
CAGTGTTCAATTACAACAAGGCCACCAAAGGACTCACTATTCTGGGAAAAGATGCAGGTGCATTGACCGA
AGATCCAGATGAAGGCACAGCTGGAATAGAGTCTGCTGTTTTAAGAGGATTTCTCATTTTGGGCAAAGAA
GACAAGAGATATGGCCCGGCATTGAGCATCAATGAGCTGAGCAATCTTGCAAAAGGAGAGAAGGCTAATG
TGCTAATTGGGCAAGGAGACGTAGTGTTGGTAATGAAACGGAAACGGGACTCTAGCATACTTACTGACAG
TCAGACAGCGACCAAAAGAATTCGGATGGCCATCAATTAG
>BYM28112022_ H3N2_PB2_1
ATCAGCAAAAGCAGGTCAATTATATTCAGCATGGAAAGAATAAAAGAACTACGGAATCTAATGTCCCGGT
CCCGCACTCGCGAGATACTGACAAAAGCCACAGTGAGCCATATGGCCATAATTAAGAAGTACACATCGGG
GAGACAGGAAAAGAACCCGTCACTTAGAATGAAATGGATGATGGCAATGAAATATCCAATTACTGCTGAC
AAAAGGATAACAGAAATGGTTCCGGAGAGAAATGAACAAGGACAAACTCTATGGAGTAAAATGAGTGATG
CTGGATCAGACAGAGTGATGGTATCACCTTTGGCTGTAACATGGTGGACTAGGAATGGACCCGAGACAGA
TACGGTCCATTACCCAAAAGTGTACAAAACTTATTTCGACAAAGTCGAAAGGTTAAAACATGGAACCTTT
GGCCCTGTCCATTTTAGAAATCAAGTCAAAATACGAAGACGAGTAGACATAAATCCTGGTCATGCAGACC
TCAGTGCCAAAGAGGCACAAGATGTAATTATGGAAGTTGTTTTTCCCAATGAAGTGGGAGCCAGAATACT
AACATCAGAATCACAGCTAACAATAACTAAAGAGAAAAAAGAAGAACTCCGAGATTGCAAAATTTCTCCC
TTGATGGTCGCATACATGCTAGAGAGAGAACTTGTGCGGAAAACAAGGTTTCTCCCAGTTGCTGGCGGAA
CAAGCAGTATATACATTGAAGTTTTACATTTGACTCAAGGAACGTGTTGGGAACAAATGTACACTCCAGG
TGGAGGAGTGAGGAATGACGATGTTGACCAAAGCCTAATTATTGCGGCCAGGAACATAGTAAGAAGAGCC
GCAGTGTCAGCAGACCCACTAGCATCTTTATTGGAGATGTGCCACAGCACGCAAATTGGCGGAACAAGGA
TGGTGGACATTCTTAGACAGAACCCGACTGAAGAACAGGCTGTGGATATATGCAAAGCTGCAATGGGATT
GAGAATCAGCTCATCTTTCAGCTTTGGTGGCTTCACATTTAAAAGAACGAGCGGGTCGTCAGTCAAAAGA
GAGGAAGAGGTTCTTACAGGCAATCTCCAGACATTGAGAATAAGAGTACATGAGGGGTATGAGGAGTTCA
CAATGGTGGGGAAAAGGGCAACAGCTATACTTAGAAAAGCAACCAGAAGACTGGTTCAACTCATAGTGAG
TGGAAGAGACGAACAGTCAATAGCCGAAGCAATAATCGTGGCCATGGTGTTTTCACAAGAAGATTGCGTG
ATAAAAGCAGTTAGAGGTGACCTGAATTTTGTCAACAGAGCAAATCGGCGGTTCAGACCTTTGCATCAGC
TTTTAGAGCATTTTCAGAAAGATGCGAAAGTACTCTTTCAAAATTGGGGAGTTGAACACATCGACAGTGT
GATAGGAATGGTTGGGCTCTTACCAGATATGACTCCAAGCACAGAGATGTCAATGAGAGGAATAAGAGTC
AGCAAAATGGGTGTGGATGAATACTCCAGTACAGAGAGGGTGGTGGTTAGCGATGGTCGGTTTTTGAGAG
TTAGAGACCAACGTGGGAATGTATTATTATCTCCTGAGGAGGTCAGTGAAACACAGGGAACTGAAAGACT
GACCATAACTTATTCATCATCGATGATGTGGGAGATTAACGGCCCTGAGTCGGTTTTGGTCAATACCTAT
CAATGGATCATCAGAAATTGGGAAGCTGTCAAAATTCAATGGTCTCAGAACCCTGCAATGTTGTACAACA
AAATGGAATTTGAACCATTTCAATCTTTAGTCCCCAAGGCCACTAGAAGCCAATACAGTGGGTTTGTCAG
AACTCTATTCCAACAAATGAGAGACTTGCTCGGGACATTTGACACTGCCCAGATAATAAAGCTTCTCCCT
TTTGCAGCTTCTCCACCGAAGCAAAGCAGAATGCAGTTCTCTTTACTGACTGTGAATGTGAGGGGATCAG

GGATGAGAATACTTGTAAAGGGCAATTCTCCTGTATTCAATTACAACAAGACCACTAAAAGGCTAACAAT
TCTTGGAAAAGATGCCGGCACTTTAATTGAAGACCCAGATGAAAGCACATCCGGAGTGGAGTCCGCCGTC
TTGAGAGGGTTCCTCATTATAGGTAAGGAAGACAGAAGATACGGACCAGCATTAAGCATCAATGAACTGA
GTAACGCTGCAAAAGGGGAAAAGGCTAATGTGCTAATTGGGCAAGGAGACGTGGTGTTGGTAATGAAACG
GAAACGGGACTCTAGTATACTTACTGACAGCCAGACAGCGACCAAAAGAATTCGGATGGCCATCAATTAA
TACTGAATAGTTTAAAAACGACCTTGTTTCTACTGATC
>NC_026435.1 Influenza A virus (A/California/07/2009(H1N1)) segment 2 polymerase PB1 (PB1) gene, complete cds; and nonfunctional PB1-F2 protein (PB1-F2) gene, complete sequence
ATGGATGTCAATCCGACTCTACTTTTCCTAAAAATTCCAGCGCAAAATGCCATAAGCACCACATTCCCTT
ATACTGGAGATCCTCCATACAGCCATGGAACAGGAACAGGATACACCATGGACACAGTAAACAGAACACA
CCAATACTCAGAAAAGGGAAAGTGGACGACAAACACAGAGACTGGTGCACCCCAGCTCAACCCGATTGAT
GGACCACTACCTGAGGATAATGAACCAAGTGGGTATGCACAAACAGACTGTGTTCTAGAGGCTATGGCTT
TCCTTGAAGAATCCCACCCAGGAATATTTGAGAATTCATGCCTTGAAACAATGGAAGTTGTTCAACAAAC
AAGGGTAGATAAACTAACTCAAGGTCGCCAGACTTATGATTGGACATTAAACAGAAATCAACCGGCAGCA
ACTGCATTGGCCAACACCATAGAAGTCTTTAGATCGAATGGCCTAACAGCTAATGAGTCAGGAAGGCTAA
TAGATTTCTTAAAGGATGTAATGGAATCAATGAACAAAGAGGAAATAGAGATAACAACCCACTTTCAAAG
AAAAAGGAGAGTAAGAGACAACATGACCAAGAAGATGGTCACGCAAAGAACAATAGGGAAGAAAAAACAA
AGACTGAATAAGAGAGGCTATCTAATAAGAGCACTGACATTAAATACGATGACCAAAGATGCAGAGAGAG
GCAAGTTAAAAAGAAGGGCTATCGCAACACCTGGGATGCAGATTAGAGGTTTCGTATACTTTGTTGAAAC
TTTAGCTAGGAGCATTTGCGAAAAGCTTGAACAGTCTGGGCTCCCAGTAGGGGGCAATGAAAAGAAGGCC
AAACTGGCAAATGTTGTGAGAAAGATGATGACTAATTCACAAGACACAGAGATTTCTTTCACAATCACTG
GGGACAACACTAAGTGGAATGAAAATCAAAATCCTCGAATGTTCCTGGCGATGATTACATATATCACCAG
AAATCAACCCGAGTGGTTCAGAAACATCCTGAGCATGGCACCCATAATGTTCTCAAACAAAATGGCAAGA
CTAGGGAAAGGGTACATGTTCGAGAGTAAAAGAATGAAGATTCGAACACAAATACCAGCAGAAATGCTAG
CAAGCATTGACCTGAAGTACTTCAATGAATCAACAAAGAAGAAAATTGAGAAAATAAGGCCTCTTCTAAT
AGATGGCACAGCATCACTGAGTCCTGGGATGATGATGGGCATGTTCAACATGCTAAGTACGGTCTTGGGA
GTCTCGATACTGAATCTTGGACAAAAGAAATACACCAAGACAATATACTGGTGGGATGGGCTCCAATCAT
CCGACGATTTTGCTCTCATAGTGAATGCACCAAACCATGAGGGAATACAAGCAGGAGTGGACAGATTCTA
CAGGACCTGCAAGTTAGTGGGAATCAACATGAGCAAAAAGAAGTCCTATATAAATAAGACAGGGACATTT
GAATTCACAAGCTTTTTTTATCGCTATGGATTTGTGGCTAATTTTAGCATGGAGCTACCCAGCTTTGGAG
TGTCTGGAGTAAATGAATCAGCTGACATGAGTATTGGAGTAACAGTGATAAAGAACAACATGATAAACAA
TGACCTTGGACCTGCAACGGCCCAGATGGCTCTTCAATTGTTCATCAAAGACTACAGATACACATATAGG
TGCCATAGGGGAGACACACAAATTCAGACGAGAAGATCATTTGAGTTAAAGAAGCTGTGGGATCAAACCC
AATCAAAGGTAGGGCTATTAGTATCAGATGGAGGACCAAACTTATACAATATACGGAATCTTCACATTCC
TGAAGTCTGCTTAAAATGGGAGCTAATGGATGATGATTATCGGGGAAGACTTTGTAATCCCCTGAATCCC
TTTGTCAGTCATAAAGAGATTGATTCTGTAAACAATGCTGTGGTAATGCCAGCCCATGGTCCAGCCAAAA
GCATGGAATATGATGCCGTTGCAACTACACATTCCTGGATTCCCAAGAGGAATCGTTCTATTCTCAACAC
AAGCCAAAGGGGAATTCTTGAGGATGAACAGATGTACCAGAAGTGCTGCAATCTATTCGAGAAATTTTTC
CCTAGCAGTTCATATAGGAGACCGGTTGGAATTTCTAGCATGGTGGAGGCCATGGTGTCTAGGGCCCGGA
TTGATGCCAGGGTCGACTTCGAGTCTGGACGGATCAAGAAAGAAGAGTTCTCTGAGATCATGAAGATCTG
TTCCACCATTGAAGAACTCAGACGGCAAAAATAA
>MF575267.1 Influenza A virus (A/black-headed gull/Netherlands/31/2014(H13N6)) segment 2 polymerase PB1 (PB1) and PB1-F2 protein (PB1-F2) genes, complete cds
AGCAAAAGCAGGCAAACCATTTGAATGGATGTCAATCCGACTTTACTTTTCTTGAAAGTGCCAGCGCAAA
ATGCTATAAGTACTACATTCCCTTATACTGGTGACCCTCCATACAGCCATGGAACAGGAACAGGATATAC
CATGGACACAGTCAACAGAACACACCAGTACTCAGAAAAAGGAACATGGACAACAAACACCGAAACTGGA
GCACCTCAGCTTAACCCGATCGATGGACCACTACCGAAGGATAATGAGCCAAGCGGATATGCACAAACGG
ATTGTGTATTGGAGGCCATGGCTTTCCTCGAAGAATCCCATCCAGGGATCTTTGAGAACTCATGTCTTGA

AACGATGGAAGTTGTTCAGCAAACACGAGTGGACAAGCTGACCCAAGGCCGACAAACATATGATTGGACA
TTGAATAGAAACCAGCCAGCTGCAACGGCTTTAGCCAACACCATAGAGGTATTCAGATCGAACGGTCTAA
CAGCTAATGAATCGGGGAGACTCATTGATTTCCTTAAGGATGTGATGGATTCAATGGATAAAAAAGGAAT
GGAAATAACAACACATTTCCAAAGAAAAAGGAGAGTAAGGGACAACATGACCAAGAAAATGGTCACACAA
AGAACAATAGGGAAAAAGAAACAGAGATTGAGCAAAAAAAGCTACCTAATAAGGGCATTGACATTGAATA
CAATGACCAAAGATGCAGAAAGAGGTAAATTGAAAAGGAGAGCGATTGCAACACCCGGGATGCAGATCAG
AGGTTTCGTCTACTTCGTTGAAACACTTGCAAGGAGCATATGTGAGAAACTTGAACAGTCGGGGCTCCCA
GTTGGAGGAAATGAGAAAAAGGCTAAATTGGCAAACGTTGTAAGAAAGATGATGACTAATTCGCAAGACA
CAGAGCTCTCCTTCACAATTACTGGGGACAACACCAAGTGGAATGAAAATCAAAACCCTCGGATGTTCCT
AGCGATGATAACATACATCACAAGAAACCAGCCTGAATGGTTCAGAAATGTCTTGAGCATTGCCCCTATA
ATGTTCTCAAACAAAATGGCAAGACTTGGGAAAGGATATATGTTCGAAAGTAAGAGCATGAAGCTACGAA
CACAAATACCAGCAGAAATGCTTGCAAGTATTGACCTCAAATATTTCAACGAGTCAACGAGAAAGAAAAT
CGAGAACATAAGACCCCTCTTAATAGATGGCACAGCCTCATTGAGTCCTGGAATGATGATGGGCATGTTC
AACATGTTGAGTACAGTCTTAGGAGTCTCAATCTTAAATCTTGGGCAAAAGAGGTACACCAAGACCACAT
ACTGGTGGGATGGACTCCAATCCTCTGATGACTTTGCACTCATAGTGAATGCACCTAATCATGAGGGAAT
ACAAGCAGGAGTGGACAGATTTTACAGAACCTGCAAATTAGTTGGAATCAACATGAGTAAAAAGAAGTCC
TACATCAATAGAACAGGAACATTTGAATTCACAAGCTTTTTCTATCGCTATGGGTTTGTAGCTAACTTCA
GTATGGAGCTACCAAGCTTTGGCGTATCAGGGATTAATGAGTCAGCTGACATGAGCATTGGAGTAACAGT
GATAAAGAACAACATGATAAACAACGACCTTGGACCAGCAACAGCCCAAATGGCTCTTCAACTGTTCATC
AAGGACTACAGGTACACATATCGATGCCATAGAGGTGATACTCAAATCCAAACAAGGAGATCATTTGAGT
TGAAGAAGCTTTGGGAGCAAACCCGTTCAAAGGCGGGGCTGCTGGTTTCAGATGGAGGACCAAACCTATA
TAATATCAGAAATCTCCATATTCCAGAGGTCTGTCTGAAATGGGAGCTAATGGATGAAGATTATCAGGGC
AGGCTGTGCAACCCTCTGAATCCATTCGTAAGCCATAAGGAAATTGAGTCCGTAAATAATGCTGTGGTAA
TGCCAGCCCATGGTCCAGCCAAAAGTATGGAATATGATGCTGTTGCAACTACACATTCCTGGGTCCCCAA
GAGGAACCGTTCCATCCTCAACACTAGCCAAAGGGGAATCCTTGAGGATGAACAGATGTATCAAAAATGC
TGCAATCTATTCGAAAAATTCTTCCCTAGCAGTTCGTACCGGAGACCAGTTGGAATTTCCAGCATGGTGG
AGGCCATGGTATCCAGGGCCCGAATTGACGCACGGATTGACTTCGAATCTGGAAGGATCAAGAAAGAAGA
GTTTGCTGAGATCATGAAGATCTGTTCCACCATTGAAGAGCTCAGACGGCAAAAGTAGTGAATTTTGCTT
GTCCTTCATGAAAAAATGCCTTGTTTCTACT
>iavh1n11922|A/Mute_Swan/Netherlands/2/2022|EPI_ISL_15364797|A/Mute_Swan/Netherlands/2/2022|PB1|2||A/Mute_Swan/Netherlands/2/2022
CAAACCATTTGAATGGATGTCAATCCGACTTTACTTTTCTTAAAAGTGCCAGCGCAAGATGCCATAAGTA
CCACATTCCCTTACACTGGAGATCCTCCATACAGCCATGGAACAGGGACAGGATACACAATGGACACAGT
CAACAGAACACATCAATACTCAGAGAAGGGGAAATGGACAACAAACACAGAAACCGGAGCACCTCAACTC
AACCCAATTGATGGGCCACTACCTGAGGACAACGAACCGAGCGGATATGCACAAACAGATTGCGTGTTGG
AAGCAATGGCTTTCCTTGAAGAGTCCCATCCAGGGATCTTTGAAAACTCTTGTCTTGAAACGATGGAAGT
CGTTCAGCAAACAAGAGTGGACAAACTAACTCAAGGTCGCCAGACATATGACTGGACACTAAATAGAAAC
CAACCAGCTGCAACTGCCCTGGCCAACACTATAGAGGTCTTCAGATCAAACGGTCTAACAGCCAATGAAT
CGGGGAGACTAATAGATTTCCTCAAGGATGTGATGGACTCAATGGATAAAGAAGAAATGGAAATAACAAC
ACATTTCCAGAGAAAGAGAAGAGTAAGGGACAACATGACCAAGAAAATGGTCACACAAAGAACAATAGGA
AAGAAGAAACAAAGGCTAAACAAGAGGAGCTACTTAATAAGAGCACTGACACTGAATACAATGACAAAAG
ATGCAGAAAGAGGCAAATTGAAGAGACGGGCGATTGCAACACCAGGGATGCAGATTAGAGGATTTGTGTA
CTTTGTCGAAACACTGGCAAGGAGCATCTGTGAAAAACTTGAGCAATCTGGACTCCCCGTTGGAGGGAAT
GAGAAGAAGGCTAAATTGGCAAATGTTGTGAGAAAAATGATGACTAACTCACAAGATACAGAGCTCTCCT
TCACAATTACTGGAGATAACACCAAATGGAATGAGAATCAAAATCCTCGGATGTTTCTGGCAATGATAAC
ATACATTACAAGAAACCAACCTGAATGGTTTAGGAATGTCTTGAGTATTGCCCCTATAATGTTCTCGAAC
AAAATGGCGAGATTGGGAAAAGGGTACATGTTTGAAAGTAAGAGCATGAAGTTACGGACACAAATACCTG

CAGAAATGCTTGCAAACATTGACTTAAAATATTTCAATGAATCAACAAGAAAGAAAATCGAAAAAATAAG
GCCTCTACTAATAGATGGCACTGCCTCATTGAGTCCTGGAATGATGATGGGCATGTTCAATATGCTGAGT
ACAGTATTAGGAGTTTCAATCCTAAATCTTGGGCAAAAGAAGTACACCAAAACCACATACTGGTGGGATG
GACTCCAATCCTCTGATGATTTCGCCCTCATAGTAAATGCACCGAATCATGAGGGAATACAAGCAGGAGT
GGATAGGTTCTATAGGACCTGCAAACTGGTCGGGATCAATATGAGCAAAAAGAAGTCTTACATAAACCGG
ACTGGAACATTTGAGTTCACAAGCTTTTTCTATCGCTATGGATTTGTGGCTAACTTCAGTATGGAGCTGC
CCAGCTTTGGAGTTTCTGGGATCAATGAATCAGCTGACATGAGCATTGGCATCACGGTGATAAAGAACAA
CATGATAAACAATGACCTTGGACCAGCAACAGCTCAAATGGCCCTTCAACTATTCATCAAAGATTACAGG
TACACGTACCGATGCCATAGAGGTGACACACAAATTCAAACGAGGAGATCATTCGAGCTGAAGAAGCTGT
GGGAACAGACCCGTTCAAAGGCAGGACTGTTGGTGTCAGATGGAGGACCAAATCTATACAACATTCGGAA
TCTCCATATCCCAGAGGTCTGCCTGAAATGGGAGCTAATGGACGAAGATTACCAGGGCAGGTTGTGTAAT
CCTCTGAACCCATTTGTCAGCCATAAAGAAATTGAGTCCGTAAACAATGCTGTGGTGATGCCAGCCCACG
GTCCAGCCAAAAGCATGGAATATGATGCCGTTGCGACTACACACTCATGGATTCCTAAAAGGAATCGTTC
CATTCTCAATACCAGTCAAAGGGGAATTCTTGAGGATGAACAGATGTACCAGAAATGCTGCGGTCTATTC
GAGAAATTCTTCCCCAGTAGTTCATACAGGAGACCAGTTGGAATTTCCAGCATGGTGGAGGCCATGGTGT
CTAGGGCCCGAATCGATGCACGCATTGATTTCGAATCTGGAAGGATCAAGAAGGAAGAGTTTGCTGAGAT
CATGAAGATCTGTTCCACCATTGAAGAGCTCAGACGGCAAAAATAGTGAATTTAGCTTGTCCTTCATGAA
AAAATG >iavh1n11922|A/Barry/3792/2022|EPI_ISL_15391170|A/Barry/3792/2022|PB1|2||A/Barry/3792/2022
CAGCAAAAGCAGGCAAACCATTTGAATGGATGTCAATCCGACTCTATTGTTCCTAAAAGTTCCAGCGCAA
AATGCCATAAGCACAACATTCCCTTATACTGGAGATCCTCCATACAGCCATGGAACAGGGACTGGGTACA
CTATGGACACAGTCAACAGAACACACCAATACTCAGAGAGAGGGAAGTGGACGACAAATACAGAAACTGG
GGCTCCCCAGCTCAACCCAATTGATGGACCACTACCTGAGGATAATGAACCAAGTGGATATGCACAAACA
GACTGTGTCCTGGAGGCTATGGCCTTTCTTGAAGAATCCCACCCAGGTATCTTTGAAAACTCCTGCCTTG
AAACAATGGAAGCCGTTCAACAGACAAGGGTAGACAAACTAACCCAAGGTCGCCAGACTTATGATTGGAC
ATTAAACAGGAACCAACCGGCAGCAACTGCATTAGCCAACACCATAGAAGTCTTTAGATCGAACGGATTA
ACAGCTAATGAATCAGGAAGGCTAATAGATTTCCTCAAGGATGTGATGGAATCAATGGATAAGGAGGAAA
TGGAGATAACAACACACTTTCAAAGAAAAAGGAGAGTAAGGGACAACATGACCAAGAAAATGGTCACACA
AAGAACAATAGGGAAGAAAAAACAAAGAGTGAATAAAAGAGGCTACCTAATAAGAGCTCTGACATTGAAC
ACGATGACCAAGGATGCAGAGAGAGGCAAATTAAAAAGAAGGGCTATTGCAACACCCGGGATGCAAATTA
GAGGATTCGTGTATTTCGTTGAAACTTTAGCTAGAAGCATTTGCGAAAAACTTGAACAATCTGGACTTCC
GGTTGGGGGTAATGAAAAGAAGGCCAAACTGGCAAATGTTGTAAGAAAAATGATGACTAATTCACAAGAC
ACAGAGCTTTCTTTCACAATCACTGGAGACAACACTAAGTGGAATGAAAATCAAAACCCCCGAATGTTTT
TGGCAATGATTACATATATCACAAAGAACCAACCTGAATGGTTCAGAAACATCCTGAGCATCGCACCAAT
AATGTTCTCAAACAAAATGGCAAGACTGGGAAAAGGGTACATGTTCGAGAGTAAGAGAATGAAGCTCCGG
ACACAAATACCTGCAGAAATGCTAGCAAGCATTGACCTGAAGTATTTCAATGAATCAACAAGGAAGAAAA
TTGAGAAAATAAGGCCTCTTCTAATAGATGGAACAGCATCATTGAGCCCTGGAATGATGATGGGCATGTT
CAACATGCTAAGTACAGTTTTAGGAGTCTCAATACTGAATCTTGGACAAAAGAAATACACCAAGACAACG
TACTGGTGGGATGGGCTCCAATCCTCAGACGATTTTGCCCTCATAGTGAATGCACCAAATCATGAGGGAA
TACAAGCAGGAGTGGATAGATTCTATAGGACCTGCAAGTTAGTGGGAATCAACATGAGCAAAAAGAAGTC
CTATATAAATAAAACAGGGACATTTGAATTCACTAGCTTCTTTTATCGATATGGATTTGTGGCTAATTTT
AGCATGGAGCTGCCAAGTTTTGGAGTGTCTGGAATAAACGAGTCAGCTGACATGAGCATTGGAGTAACAG
TGATAAAGAACAACATGATAAATAATGACCTTGGACCAGCAACAGCCCAAATGGCTCTCCAATTGTTCAT
CAAAGATTACAGATACACGTATCGGTGCCATAGAGGAGACACACAAATCCAAACAAGAAGATCATTCGAG
ATAAAGAAGCTATGGGATCAAACCCAATCAAAGACAGGATTATTGGTATCAGATGGGGGACCAAACTTAT
ACAATATCCGAAATCTTCACATCCCTGAAGTCTGCTTGAAGTGGGAGCTGATGGACGACAATTATCGGGG
AAGACTTTGTAATCCCCTGAATCCCTTTGTCAGCCATAAAGAAATTGAATCTGTAAACAATGCTGTAGTA

ATGCCAGCTCATGGTCCGGCCAAAAGTATGGAATATGATGCCGTTGCGACTACACACTCCTGGATTCCCA
AAAGGAACCGCTCTATTCTAAACACAAGCCAAAGGGGAATTCTTGAGGATGAACAAATGTACCAGAAATG
CTGCAACTTGTTCGAGAAATTTTTCCCTAGTAGTTCATATAGGAGACCGATTGGAATTTCTAGCATGGTG
GAGGCCATGGTATCTAGGGCCCGGATTGATGCCAGAATTGACTTCGAGTCTGGAAGGATTAAGAAGGAAG
AGTTCTCTGAGATCATGAAGATCTGTTCCACCATTGAAGAACTCAGACGGCAAAAACAATGAATTTAGCT
TGTCCTTCATGAAAAAATGCCTTGTTTCTACTGATC >CRG12092022_ H13N6_PB1_2
NNNNNNNNNNNNNNNNNNNNNNNNNNNNNNNNNNNNNNNNNNNNNNNNNNNNNNNNNNNNNNNNNNNNNN
NNNNNNNNNNNNNNNNNNNNNNNNNNNNNNNNNNNNNNNNNNNNNNNNNNNNNNNNNNNNNNNNNNNNNN
NNNNNNNNNNNNNNNNNNNNNNNNNNNNNNNNNNNNNNNNNNNNNNNNNNNNNNNNNNNNNNNNNNNNNN
NNNNNNNNNNNNNNNNNNNNNNNNNNNNNNNNNNNNNNNNNNNNNNNNNNNNNNNNNNNNNNNNNNNNNN
NNNNNNNNNNNNNNNNNNNNNNNNNNNNNNNNNNNNNNNNNNNNNNNNNNNNNNNNNNNNNNNNNNNNNN
NNNNNNNNNNNNNNNNNNNNNNNNNNNNNNNNNNNNNNNNNNNNNNNNNNNNNNNNNNNNNNNNNNNNNN
NNNNNNNNNNNNNNNNNNNNNNNNNNNNNNNNNNNNNNNNNNNNNNNNNNNNNNNNNNNNNNNNNNNNNN
NNNNNNNNNNNNNNNNNNNNNNNNNNNNNNNNNNNNNNNNNNNNNNNNNNNNNNNNNNNNNNNNNNNNNN
NNNNNNNNNNNNNNNNNNNNNNNNNNNNNNNNNNNNNNNNNNNNNNNNNNNNNNNNNNNNNNNNNNNNNN
NNNNNNNNNNNNNNNNNNNNNNNNNNNNNNNNNNNNNNNNNNNNNNNNNNNNNNNNNNNNNNNNNNNNNN
NNNNNNNNNNNNNNNNNNNNNNNNNNNNNNNNNNNNNNNNNNNNNNNNNNNNNNNNNNNNNNNNNNNNNN
NNNNNNNNNNNNNNNNNNNNNNNNNNNNNNNNNNNNNNNNNNNNNNNNNNNNNNNNNNNNNNNNNNNNNN
NNNNNNNNNNNNNNNNNNNNNNNNNNNNNNNNNNNNNNNNNNNNNNNNNNNNNNNNNNNNNNNNNNNNNN
NNNNNNNNNNNNNNNNNNNNNNNNNNNNNNNNNNNNNNNNNNNNNNNNNNNNNNNNNNNNNNNNNNNNNN
NNNNNNNNNNNNNNNNNNNNNNNNNNNNNNNNNNNNNNNNNNNNNNNNNNNNNNNNNNNNNNNNNNNNNN
NNNNNNNNNNNNNNNNNNNNNNNNNNNNNNNNNNNNNNNNNNNNNNNNNNNNNNNNNNNNNNNNNNNNNN
NNNNNNNNNNNNNNNNNNNNNNNNNNNNNNNNNNNNNNNNNNNNNNNNNNNNNNNNNNNNNNNNNNNNNN
NNNNNNNNNNNNNNNNNNNNNNNNNNNNNNNNNNNNNNNNNNNNNNNNNNNNNNNNNNNNNNNNNNNNNN
NNNNNNNNNNNNNNNNNNNNNNNNNNNNNNNNNNNNNNNNNNNNNNNNNNNNNNNNNNNNNNNNNNNNNN
NNNNNNNNNNNNNNNNNNNNNNNNNNNNNNNNNNNNNNNNNNNNNNNNNNNNNNNNNNNNNNNNNNNNNN
NNNNNNNNNNNNNNNNNNNNNNNNNNNNNNNNNNNNNNNNNNNNNNNNNNNNNNNNNNNNNNNNNNNNNN
NNNNNNNNNNNNNNNNNNNNNNNNNNNNNNNNNNNNNNNNNNNNNNNNNNNNNNNNNNNNNNNNNNNNNN
NNNNNNNNNNNNNNNNNNNNNNNNNNNNNNNNNNNNNNNNNNNNNNNNNNNNNNNNNNNNNNNNNNNNNN
NNNNNNNNNNNNNNNNNNNNNNNNNNNNNNNNNNNNNNNNNNNNNNNNNNNNNNNNNNNNNNNNNNNNNN
NNNNNNNNNNNNNNNNNNNNNNNNNNNNNNNNNNNNNNNNNNNNNNNNAAACAAGGAGATCATTTGAAT
TGAAGAAACTTTGGGAGCAAACCCGTTCAAAAGCGGGGCTACTGGTTTCAGATGGAGGTCCAAATCTATA
TAACATCAGAAATCTCCATATTCCAGAGGTCTGTCTGAAATGGGAGCTAATGGATGAAGATTATCAGGGC
AGGCTGTGCAATCCTCTGAATCCTTTCGTAAGCCATAAGGAAATTGAGTCTGTAAACAATGCTGTGGTAA
TGCCAGCCCATGGTCCAGCCAAAAGTATGGAATATGATGCTGTTGCAACTACACATTCCTGGGTCCCCAA
GAGGAACCGTTCCATTCTCAACACTAGCCAAAGGGGAATCCTTGAGGATGAACAGATGTACCAAAAATGT
TGCAATCTATTCGAAAAATTCTTCCCTAGCAGTTCGTACAGGAGACCAGTTGGAATTTCCAGCATGGTGG
AGGCCATGGTATCCAGGGCCCGAATTGACGCACGGATTGACTTCGAATCTGGAAGGATCAAGAAAGAAGA
GTTTGCTGAGATCATGAAGATCTGTTCCACCATTGAAGAGCTCAGACGGCAAAAGTAGTGAATTTTGCTT
GTCCTTCATGAAAAAATGCCTTGTTTCTACT
>NC_026437.1 Influenza A virus (A/California/07/2009(H1N1)) segment 3 polymerase PA (PA) gene, complete cds
ATGGAAGACTTTGTGCGACAATGCTTCAATCCAATGATCGTCGAGCTTGCGGRAAAGGCAATGAAAGAAT
ATGGGGAAGATCCGAAAATCGAAACTAACAAGTTTGCTGCAATATGCACACATTTGGAAGTTTGTTTCAT
GTATTCGGATTTCCATTTCATCGACGAACGGGGTGAATCAATAATTGTAGAATCTGGTGACCCGAATGCA
CTATTGAAGCACCGATTTGAGATAATTGAAGGAAGAGACCGAATCATGGCCTGGACAGTGGTGAACAGTA
TATGTAACACAACAGGGGTAGAGAAGCCTAAATTTCTTCCTGATTTGTATGATTACAAAGAGAACCGGTT

CATTGAAATTGGAGTAACACGGAGGGAAGTCCACATATATTACCTAGAGAAAGCCAACAAAATAAAATCT
GAGAAGACACACATTCACATCTTTTCATTCACTGGAGAGGAGATGGCCACCAAAGCGGACTACACCCTTG
ACGAAGAGAGCAGGGCAAGAATCAAAACTAGGCTTTTCACTATAAGACAAGAAATGGCCAGTAGGAGTCT
ATGGGATTCCTTTCGTCAGTCCGAAAGAGGCGAAGAGACAATTGAAGAAAAATTTGAGATTACAGGAACT
ATGCGCAAGCTTGCCGACCAAAGTCTCCCACCGAACTTCCCCAGCCTTGAAAACTTTAGAGCCTATGTAG
ATGGATTCGAGCCGAACGGCTGCATTGAGGGCAAGCTTTCCCAAATGTCAAAAGAAGTGAACGCCAAAAT
TGAACCATTCTTGAGGACGACACCACGCCCCCTCAGATTGCCTGATGGGCCTCTTTGCCATCAGCGGTCA
AAGTTCCTGCTGATGGATGCTCTGAAATTAAGTATTGAAGACCCGAGTCACGAGGGGGAGGGAATACCAC
TATATGATGCAATCAAATGCATGAAGACATTCTTTGGCTGGAAAGAGCCTAACATAGTCAAACCACATGA
GAAAGGCATAAATCCCAATTACCTCATGGCTTGGAAGCAGGTGCTAGCAGAGCTACAGGACATTGAAAAT
GAAGAGAAGATCCCAAGGACAAAGAACATGAAGAGAACAAGCCAATTGAAGTGGGCACTCGGTGAAAATA
TGGCACCAGAAAAAGTAGACTTTGATGACTGCAAAGATGTTGGAGACCTTAAACAGTATGACAGTGATGA
GCCAGAGCCCAGATCTCTAGCAAGCTGGGTCCAAAATGAATTCAATAAGGCATGTGAATTGACTGATTCA
AGCTGGATAGAACTTGATGAAATAGGAGAAGATGTTGCCCCGATTGAACATATCGCAAGCATGAGGAGGA
ACTATTTTACAGCAGAAGTGTCCCACTGCAGGGCTACTGAATACATAATGAAGGGAGTGTACATAAATAC
GGCCTTGCTCAATGCATCCTGTGCAGCCATGGATGACTTTCAGCTGATCCCAATGATAAGCAAATGTAGG
ACCAAAGAAGGAAGACGGAAAACAAACCTGTATGGGTTCATTATAAAAGGAAGGTCTCATTTGAGAAATG
ATACTGATGTGGTGAACTTTGTAAGTATGGAGTTCTCACTCACTGACCCGAGACTGGAGCCACACAAATG
GGAAAAATACTGTGTTCTTGAAATAGGAGACATGCTCTTGAGGACTGCGATAGGCCAAGTGTCGAGGCCC
ATGTTCCTATATGTGAGAACCAATGGAACCTCCAAGATCAAGATGAAATGGGGCATGGAAATGAGGCGCT
GCCTTCTTCAGTCTCTTCAGCAGATTGAGAGCATGATTGAGGCCGAGTCTTCTGTCAAAGAGAAAGACAT
GACCAAGGAATTCTTTGAAAACAAATCGGAAACATGGCCAATCGGAGAGTCACCCAGGGGAGTGGAGGAA
GGCTCTATTGGGAAAGTGTGCAGGACCTTACTGGCAAAATCTGTATTCAACAGTCTATATGCGTCTCCAC
AACTTGAGGGGTTTTCGGCTGAATCTAGAAAATTGCTTCTCATTGTTCAGGCACTTAGGGACAACCTGGA
ACCTGGAACCTTCGATCTTGGGGGGCTATATGAAGCAATCGAGGAGTGCCTGATTAATGATCCCTGGGTT
TTGCTTAATGCATCTTGGTTCAACTCCTTCCTCACACATGCACTGAAGTAG
>MF575147.1 Influenza A virus (A/black-headed gull/Netherlands/31/2014(H13N6)) segment 3 polymerase PA (PA) and PA-X protein (PA-X) genes, complete cds
AGCAAAAGCAGGTACTGATCCAAAATGGAAGACTTTGTGCGACAATGCTTCAATCCAATGATTGTCGAGC
TTGCGGAAAAGGCAATGAAAGAATATGGGGAAGATCCGAAAATCGAAACAAACAAATTTGCCGCAATATG
CACACACTTAGAAGTCTGTTTCATGTATTCGGATTTCCACTTTATTGATGAACGGGGCGAATCAATAATT
GTAGAATCTGGCGACCCGAATGCATTATTGAAACACCGATTTGAGATAATTGAAGGAAGAGACCGAACAA
TGGCCTGGACAGTGGTTAATAGTATCTGCAACACCACAGGAGTCGATAAGCCTAAATTCCTCCCAGATTT
ATATGACTACAAAGAGAACCGATTCATTGAAATTGGAGTGACACGAAGGGAAGTCCACATATACTATCTA
GAAAAAGCCAACAAGATAAAATCAGAGAAGACACACATTCACATATTCTCATTCACTGGAGAGGAAATGG
CCACCAAAGCGGACTACACCCTTGATGAAGAGAGCAGAGCAAGAATAAAAACCAGGTTGTTCACTATAAG
ACAAGAAATGGCCAGTAGGGGTCTATGGGATTCCTTTCGTCAGTCCGAGAGAGGCGAAGAGACAATTGAA
GAAAGATTTGAAATCACAGGAACCATGCGCAGGCTTGCCGACCAAAGTCTCCCACCGAACTTCTCCAGCC
TTGAAAACTTTAGAGCCTATGTGGATGGATTCGAACCGAACGGCTGCATTGAGGGCAAGCTTTCTCAAAT
GTCAAAAGAAGTGAACGCCAGAATTGAGCCATTTCTGAAAACAACACCACGCCCTCTCAGGTTACCTGAT
GGGCCTCCCTGCGCTCAGCGGTCAAAGTTCTTGCTGATGGATGCCCTTAAATTAAGCATCGAAGACCCGA
GTCATGAGGGGGAAGGTATACCGCTATATGATGCAATCAAATGCATGAAGACATTCTTCGGCTGGAAAGA
GCCCAACATCGTAAAACCACATGAAAAGGGCATAAACCCCAACTACCTCCTGGCTTGGAAGCAGGTGCTA
GCAGAACTCCAAGATATTGAAAACGAGGAGAAAATCCCAAAAACAAAGAACATGAAGAAGACAAGCCAAT
TGAAGTGGGCACTTGGTGAGAACATGGCACCAGAGAAAGTGGACTTTGAGGACTGCAAAGATGTTAGTGA
TCTAAAACAGTATAACAGTGAAGAACCAGAGTCTAGATCGCTAGCAAGCTGGATCCAGAGTGAATTCAAC
AAGGCATGCGAATTGACAGATTCAAGTTGGATCGAACTTGACGAAATAGGGGAAGACGTTGCTCCAATTG

AGCACATTGCGAGTATGAGGAGAAACTATTTCACAGCGGAAGTATCCCATTGCAGGGCTACTGAATACAT
AATGAAGGGAGTGTATATAAACACAGCCCTGTTGAATGCATCCTGCGCAGCCATGGATGACTTCCAATTG
ATTCCAATGATAAGCAAGTGCAGAACCAAAGAAGGAAGGCGGAAAACAAATCTGTATGGATTCATTGTAA
AAGGCAGATCCCATTTGAGAAACGACACCGATGTGGTTAACTTTGTGAGCATGGAGTTCTCTCTCACTGA
CCCGAGGCTGGAGCCACACAAATGGGAAAAGTACTGTGTTCTTGAGATAGGGGACATGCTCCTACGGACT
GCAATAGGCCAAGTGTCAAGGCCCATGTTCCTGTACGTAAGAACCAACGGGACTTCCAAGATCAAAATGA
AATGGGGCATGGAAATGAGGCGATGCCTTCTTCAGTCCCTTCAACAAATTGAGAGCATGATTGAGGCAGA
GTCTTCTGTCAAAGAGAAGGACATGACCAAGGAATTCTTTGAAAACAAATCAGAAACATGGCCAATTGGA
GAATCGCCCAAAGGGGTGGAGGAGGGCTCCATTGGGAAGGTTTGCAGAACATTACTAGCAAAATCTGTGT
TCAACAGCCTGTATGCATCTCCACAACTCGAAGGGTTTTCAGCTGAATCGAGAAAGTTGCTTCTCATTGT
TCAGGCACTTAGGGACAACCTGGAACCTGGAACCTTCGATCTTGGGGGGCTATATGAAGCAATTGAGGAG
TGCCTGATTAATGATCCCTGGGTTTTGCTTAATGCGTCCTGGTTCAACTCCTTCCTCACACATGCACTGA
AATAGTTGTGGCAATGCTACTATTTGCTATCCATACTGTCCAAAAAAGTACCTTGTTTCTACT
>iavh1n11922|A/Mute_Swan/Netherlands/2/2022|EPI_ISL_15364797|A/Mute_Swan/Netherlands/2/2022|PA|3||A/Mute_Swan/Netherlands/2/2022
TACTGATCCAAAATGGAAGACTTTGTGCGACAATGCTTCAATCCAATGATTGTCGAGCTTGCGGAAAAGG
CAATGAAAGAATATGGGGAAGATCCGAAAATCGAATCAAACAAATTTGCCGCAATATGCACACACCTAGA
AGTCTGCTTCATGTATTCGGATTTCCACTTTATTGATGAACGAGGAGAATCAATAATTGTAGAATCTGGC
GATCCGAATGCATTATTGAAACACCGATTCGAGATAATTGAAGGGAGAGACCGAACCATGGCCTGGACAG
TGGTGAATAGCATCTGCAACACCACAGGAGTCGAAAAGCCCAAATTCCTCCCTGATTTGTATGACTACAA
AGAGAACCGATTCATTGAAATTGGAGTAACGCGAAGGGAAGTTCACATATACTATCTAGAGAAAGCCAAC
AAGATAAAATCAGAGAAGACACACATTCATATATTCTCATTCACTGGAGAGGAAATGGCCACCAAGGCGG
ACTACACCCTTGATGAAGAGAGTAGAGCAAGAATAAAAACCAGGCTATTCACTATAAGACAAGAAATGGC
CAGTAGGGGTCTATGGGATTCCTTTCGTCAGTCCGAGAGAGGCGAAGAGACAATTGAAGAAAGATTTGAA
ATCACAGGAACCATGCGCAGGCTTGCCAACCAAAGTCTCCCACCGAACTTCTCCAGCCTTGAAAACTTTA
GAGCCTATGTGGATGGATTCGAACCGAACGGCTGCATTGAGGGCAAGCTTTCTCAAATGTCAAAAGAAGT
GAATGCCAGAATTGAGCCATTTCTGAAGAAAACACCACGCCCTCTCAGATTACCTGATGGGCCTCCCTGT
TCTCAGCGATCGAAGTTCTTGCTGATGGATGCCCTCAAATTGAGCATCGAAGACCCGAGCCATGAGGGGG
AGGGTATACCGCTGTATGATGCAATCAAATGCATGAAGACATTTTTTGGCTGGAAAGAGCCTAACATCGT
AAAACCGCATGAAAAGGGCATAAACCCTAATTACCTTCTGGCTTGGAAACAGGTGCTGGCAGAACTCCAA
TATATTGAAAATGAGGAGAAAATCCCAAAAACAAAGAACATGAAGAAAACAAGCCAATTGAAGTGGGCAC
TTGGTGAGAACATGGCACCAGAGAAGGTGGACTTTGAGGACTGTAAAGATGTTAGTGATCTAAGACAGTA
CGACAGTGACGAACCAGAGTCTAAATCACTAGCAAGCTGGATCCAGAGTGAATTCAACAAGGCATGCGAA
TTGACAGATTCAAGTTGGATTGAACTTGATGAAATAGGGGAAGACATTGCTCCAATTGAACACATTGCGA
GTATGAGGAGAAACTATTTCACAGCGGAGGTGTCCCATTGCAGGGCTACTGAATACATAATGAAGGGAGT
ATACATAAACACAGCCCTGTTGAATGCATCCTGTGCAGCGATGGATGACTTTCAACTGATTCCTATGATA
AGCAAGTGCAGAACCAAAGAAGGAAGACGGAAGACAAATTTATATGGATTCATTATAAAAGGAAGATCCC
ATTTGAGGAATGACACCGATGTGGTGAACTTTGTGAGCATGGAATTCTCTCTCACTGACCCGAGGCTGGA
GCCACACAAATGGGAAAAGTACTGTGTTCTTGAGATAGGAGACATGCTCCTACGGACTGCAATAGGCCAA
GTGTCAAGGCCCATGTTCTTGTATGTGAGAACCAATGGAACTTCCAAGATCAAAATGAAATGGGGCATGG
AGATGAGACGATGCCTTCTTCAGTCCCTTCAACAAATTGAGAGCATGATTGAGGCCGAGTCTTCTGTCAA
AGAGAAGGACATGACCAAGGAATTCTTTGAAAACAAATCAGAAACATGGCCAATTGGGGAATCACCCAAA
GGGGTGGAAGAGGGCTCCATTGGGAAGGTATGCAGAACATTGCTAGCAAAGTCTGTGTTCAACAGCCTAT
ATGCATCTCCACAACTCGAGGGGTTTTCAGCTGAATCAAGAAAATTGCTTCTCATTGTTCAGGCACTTAG
GGACAACCTGGAACCTGGAACCTTCGATCTTGGGGGGCTATATGAAGCAATTGAGGAGTGCCTGATTAAC
GATCCCTGGGTTTTGCTTAATGCGTCTTGGTTCAACTCCTTCCTCACACATGCACTGAAATAGTTGTGGC
AATGCTACTATTTGCTATCCATACTGTCCAAAAAAGTA

>iavh1n11922|A/Barry/3792/2022|EPI_ISL_15391170|A/Barry/3792/2022|PA|3||A/Barry/3792/2022
CAGCAAAAGCAGGTACTGATTCAAAATGGAAGATTTTGTGCGACAATGCTTCAACCCGAAGCAATGAAAG
AGTATGGGGAGGATCTGAAAATTGAAACCAACAAATTTGCAGCAATATGCACTCACTTGGAGGTGTGTTT
CAGATTTCCATTTCATCAATGAACAAGGCGAATCAATAGTAGTAGAACTTGACGATCCAAATGCACTGTT
AAAACACAGATTTGAAATAATCGAGGGGAGAGACAGAACAATGGCCTGGACAGTAGTAAACAGTATCTGC
AACACTACTGGAGCTGAAAAACCGAAGTTTTTACCGGATTTGTATGATTACAAAGAAAACAGATTCATCG
AAATTGGAGTGACAAGGAGAGAAGTCCACATATATTACCTTGAAAAGGCCAATAAGATTAAATCTGAGAA
AACACACATTCACATTTTTTCATTCACTGGGGAGGAAATGGCCACAAGGGCAGACTACACTCTCGATGAG
GAGAGCAGGGCTAGGATCAAAACCAGGCTGTTTACCATAAGACAAGAAATGGCCAACAGAGGCCTCTGGG
ATTCCTTTCGTCAGTCCGAAAGAGGCGAAGAAACAATTGAAGAAAAATTTGAAATCACAGGGACTATGCG
CAGGCTTGCCGACCAAAGTCTCCCACCGAACTTCTCCTGCCTTGAGAATTTTAGAGCCTATGTGGATGGA
TTCGAACCGAACGGCTGCATTGAGGGCAAGCTTTCTCAAATGTCCAAAGAAGTGAATGCCCAAATTGAAC
CTTTTCTGAAGACAACACCAAGACCGATCAAACTTCCTAGTGGACCTCCTTGTTATCAGCGATCCAAATT
CCTCCTGATGGATGCTTTGAAATTGAGCATTGAAGACCCAAGTCACGAAGGAGAAGGGATCCCATTATAT
GATGCAATCAAGTGCATGAAAACATTCTTTGGATGGAAAGAACCTTGTATAGTCAAACCACACGAAAAGG
GAATAAATTCAAATTACCTGCTGTCATGGAAGCAAGTACTGTCAGAATTGCAGGACATTGAAAATGAGGA
GAAGATTCCAAGAAATAAAAACATGAAGAAAACGAGTCAACTGAAGTGGGCTCTTGGTGAAAACATGGCA
CCAGAGAAGGTAGACTTTGAAAACTGCAGAGACATAAGCGATTTGAAGCAATATGATAGTGAAGAACCTG
AATTAAGGTCACTTTCAAGCTGGATACAGAGTGAGTTCAACAAGGCCTGTGAGCTAACTGATTCAGTCTG
GATAGAACTCGATGAAATTGGAGAGGACGTAGCCCCAATTGAACACATTGCAAGCATGAGAAGGAATTAT
TTCACAGCAGAGGTGTCCCATTGTAGAGCTACTGAATACATAATGAAAGGGGTATACATTAATACTGCCC
TGCTCAATGCATCCTGTGCAGCAATGGACGATTTTCAACTAATTCCCATGATAAGCAAGTGCAGAACTAA
AGAGGGAAGGCGAAAAACCAATTTATATGGATTCATCATAAAGGGAAGATCTCATCTGAGGAATGACACA
GATGTGGTAAACTTTGTGAGCATGGAGTTTTCTCTCACAGATCCTAGACTTGAACCACATAAATGGGAGA
AATATTGTGTCCTTGAGATAGGAGATATGTTACTAAGGAGTGCCATAGGCCAAATTTCAAGGCCAATGTT
CTTGTATGTGAGGACAAACGGAACATCAAAAGTCAAAATGAAATGGGGAATGGAGATGAGACGTTGCCTC
CTTCAGTCACTCCAGCAGATCGAGAGCATGATTGAAGCCGAGTCCTCAGTTAAAGAGAGAGACATGACCA
AAGAGTTTTTTGAGAATAAATCAGAAGCATGGCCCATTGGGGAATCCCCCAAAGGAGTGGAAGAAGGTTC
CATTGGGAAAGTCTGTAGGACTCTATTGGCTAAGTCAGTATTCAATAGCCTATATGCATCACCACAATTG
GAAGGATTTTCAGCAGAGTCAAGAAAACTGCTCCTTATTGTTCAGGCTCTTAGGGACAAACTCGAACCTG
GGACTTTTGATCTTGGGGGGCTATATGAAGCAATTGAGGAGTGCCTGATTAATGATCCCTGGGTTTTGCT
CAATGCGTCTTGGTTCAACTCCTTCCTGACACATGCACTAAAATAGTTATAGCAGTGCTACTATTTGTTA
TCCGTACTGTCCAAAAAAGTACCTTGTTTCTACT
>BYM28112022_ H3N2_PA_3
CAGCAAGAGCAGGTACTGATTCAAAATGGAAGATTTTGTGCGACAATGCTTTAACCCGAAGCAATGAAAG
AGTATGGGGAGGATCTGAAAATTGAAACCAACAAATTTGCAGCAATATGCACTCACTTGGAGGTGTGTTT
CAGATTTCCATTTCATCAATGAACAAGGCGAATCAATAGTAGTAGAACTTGACGATCCAAATGCACTGTT
AAAACACAGATTTGAAATAATCGAGGGGAGAGACAGAACAATGGCCTGGACAGTAGTAAACAGTATCTGC
AACTTTACTGGAGCTGAAAAACCGAAGTTTTTACCGGATTTGTATGATTACAAAGAAAACAGATTCATCG
AAATTGGAGTGACTGGGAGAGAAGTCCACATATATTACCTTGAAAAGGCCAATAAGATTAAATCTGAGAA
AACACACATTCACATTTCTTCATTCACTGGGGAGGAAATGGCCACAAGCCCGAACTACACTCTCGATGAG
GAGAGCAGGGCTAGGATCAAAACCAGACTGTTTACCATAAGACAAGAAATGGCCAACAGAGGCCTCTGAG
ATTCCTTTCGTCAGTCCGAAAGAGGCGAAGAAACAATTGAAGAAAAATTTGAAATCACAGGGACTATGCG
CAGGCTTGCCAACCAAAGTCTCCCACCGAACTTCTCCTGCCTTGAGAATTTTAGAGCCTATGTGGATGGA
TTCGAACCGAACGGCTGCATTGAGGGCAAGCTTTCTCAAATGTCCAAAGAAGTGAATGCCCAAATTGAAC
CTTTTCTGGTGACAACACCAAGACCGATAAAACTTACTCGTGGACATCATATTTATCAGCGATCCAAATT

CCTCCTAATGGATGCTTTGAAATTGAGCATTGAAGACCCAAGTCACGAAGGAGAAGGGATCCCATTATAT
GATGCAATCAAGTGCATGAAAACATTCTTTGGATGGAAAGAACCTTGTATAGTCAAACCACACGAAAAGG
GAATAAATTCAAATTATCTGCTGTCATGGAAGGAAATACTGTCAGAATTGCAGGACATTGAAAATGAAGA
GAAGATTCCAAGAACTAAAAACATGAAGAAAACGAGTCAACTGAAGTGGGCTCTTGGTGAAAACATGGCA
CCAGAGAAGGTAGACTTTGAAAACTACAGAGACATAAGCGATTTGAAGCAATATGATAGTGAAGAACCAG
AATTAAGGTCACTTTCAAACTGGATACAGAGTGAGTTCAACAAGGCCTGTGAGCTAACTGATTCAGTCTG
GATAGAACTCGATGAAATTGGAGAGGACGTAGCCCCAATTGAACACATTGCAAGCATGAGAAGAGATTAT
TTCACAGCAGAGGTGTCCCATTGAAGAGCTACTGAATACATAATGAAAGGGGTATACATTAATACTGCCC
TGCTCAATGCATCCTGTGCAGCAATGGACGATTTTCGACTAATTCCCATGATAAGCAAGTGCAGAACTAA
AGAGGGAAGGCGAAAAACCAATTTATATGGATTCATCATAAAGGGAAGATCCCATCTGAGGAATGACACA
GATGTGGTAAACTTTGTGAGCATGGAGTTTTCTCTCACAGATCCGAGACTTGAACCACATAAATGGGAGA
AATATTGTGTTCTTGAGATAGGAGATATGTTACTAAGGAGTGCCATAGGCCAAATTTCAAGGCCAATGTT
CTTGTATGTGAGGACAAACGGAACATCAAAAGTCAAAATGAAATGGGGAATGGAGATGAGACGTTGCCTC
CTTCAGTCACTCCAGCAGATCGAGAGCATGATTGAAGCCGAGTCCTCAGTTAAAGAGAGAGACATGACCA
AAGAGTTTTTTGAGAATAAATCAGAAGCATGGCCCATTGGGGAATCCCCCAAAGGAGTGGAAGAAGGTTC
CATTGGGAAAGTCTGTAGGACTCTATTGGCTAAGTCAGTATTCAATAGCCTATATGCATCACCACAATTG
GAAGGATTTTCAGCAGAGTCAAGAAAACTGCTCCTTATTGTTCAGGCTCTTAGGGACAAACTCGAACCTG
GGACTTTTGATCTTGGGGGGCTATATGAAGCAATTGAGGAGTGCCTGATTAATGATCCCTGGGTTTTGCT
CAATGCGTCTTGGTTCAACTCCTTCCTGACACATGCACTAAAATAGTTATAGCAGTGCTACTATTTGTTA
TCCGTACTGTCCAAAAAAGTACCTTGTTTCTACT
>NC_026433.1 Influenza A virus (A/California/07/2009(H1N1)) segment 4 hemagglutinin (HA) gene, complete cds
ATGAAGGCAATACTAGTAGTTCTGCTATATACATTTGCAACCGCAAATGCAGACACATTATGTATAGGTT
ATCATGCGAACAATTCAACAGACACTGTAGACACAGTACTAGAAAAGAATGTAACAGTAACACACTCTGT
TAACCTTCTAGAAGACAAGCATAACGGGAAACTATGCAAACTAAGAGGGGTAGCCCCATTGCATTTGGGT
AAATGTAACATTGCTGGCTGGATCCTGGGAAATCCAGAGTGTGAATCACTCTCCACAGCAAGCTCATGGT
CCTACATTGTGGAAACACCTAGTTCAGACAATGGAACGTGTTACCCAGGAGATTTCATCGATTATGAGGA
GCTAAGAGAGCAATTGAGCTCAGTGTCATCATTTGAAAGGTTTGAGATATTCCCCAAGACAAGTTCATGG
CCCAATCATGACTCGAACAAAGGTGTAACGGCAGCATGTCCTCATGCTGGAGCAAAAAGCTTCTACAAAA
ATTTAATATGGCTAGTTAAAAAAGGAAATTCATACCCAAAGCTCAGCAAATCCTACATTAATGATAAAGG
GAAAGAAGTCCTCGTGCTATGGGGCATTCACCATCCATCTACTAGTGCTGACCAACAAAGTCTCTATCAG
AATGCAGATGCATATGTTTTTGTGGGGTCATCAAGATACAGCAAGAAGTTCAAGCCGGAAATAGCAATAA
GACCCAAAGTGAGGGRTCRAGAAGGGAGAATGAACTATTACTGGACACTAGTAGAGCCGGGAGACAAAAT
AACATTCGAAGCAACTGGAAATCTAGTGGTACCGAGATATGCATTCGCAATGGAAAGAAATGCTGGATCT
GGTATTATCATTTCAGATACACCAGTCCACGATTGCAATACAACTTGTCAAACACCCAAGGGTGCTATAA
ACACCAGCCTCCCATTTCAGAATATACATCCGATCACAATTGGAAAATGTCCAAAATATGTAAAAAGCAC
AAAATTGAGACTGGCCACAGGATTGAGGAATATCCCGTCTATTCAATCTAGAGGCCTATTTGGGGCCATT
GCCGGTTTCATTGAAGGGGGGTGGACAGGGATGGTAGATGGATGGTACGGTTATCACCATCAAAATGAGC
AGGGGTCAGGATATGCAGCCGACCTGAAGAGCACACAGAATGCCATTGACGAGATTACTAACAAAGTAAA
TTCTGTTATTGAAAAGATGAATACACAGTTCACAGCAGTAGGTAAAGAGTTCAACCACCTGGAAAAAAGA
ATAGAGAATTTAAATAAAAAAGTTGATGATGGTTTCCTGGACATTTGGACTTACAATGCCGAACTGTTGG
TTCTATTGGAAAATGAAAGAACTTTGGACTACCACGATTCAAATGTGAAGAACTTATATGAAAAGGTAAG
AAGCCAGCTAAAAAACAATGCCAAGGAAATTGGAAACGGCTGCTTTGAATTTTACCACAAATGCGATAAC
ACGTGCATGGAAAGTGTCAAAAATGGGACTTATGACTACCCAAAATACTCAGAGGAAGCAAAATTAAACA
GAGAAGAAATAGATGGGGTAAAGCTGGAATCAACAAGGATTTACCAGATTTTGGCGATCTATTCAACTGT
CGCCAGTTCATTGGTACTGGTAGTCTCCCTGGGGGCAATCAGTTTCTGGATGTGCTCTAATGGGTCTCTA
CAGTGTAGAATATGTATTTAA >MF575089.1 Influenza A virus (A/black-headed gull/Netherlands/31/2014(H13N6)) segment 4 hemagglutinin (HA) gene, complete cds

AGCAAAAGCAGGGGAGAATTCAACAAACCGAAACGAGAGAAATGGAAGTCCCAGTATTCGCACTCTTAGT
GTTAACCAGCATATGCGTACAAGCTGATAGGATTTGTGTTGGGTACTTAAGCACAAACTCATCAGAAAGG
GTTGACACACTGCTAGAGAATAATGTCCCGGTTACAAGCTCTGTTGATTTGGTTGAGACTAACCACACAG
GAACATATTGTTCTTTGGGTGGAATTAGTCCAGTGCACTTGGGAGACTGTAGCTTCGAGGGCTGGATTCT
AGGGAACCCTGCCTGTGCCAGCAACCTGGGGATTAGAGAATGGTCATATTTGATTGAGGACCCTTCTGCT
CCTCATGGATTGTGCTATCCAGGAGAGTTAGACAACAATGGAGAATTGAGGCACTTGTTTAGTGGAATCA
GATCTTTCAGTAGAACGGAATTGTTCGCACCTACCTCTTGGGGGGCAGTGAATGATGGAGTAACGGCTGC
CTGTCAAGACAGAGGAGCTAGCAGCTTTTACCGGAACTTGGTATGGTTTGTGAAGAGAGGGAATAACTAT
CCTGTGATCCGCGGGACCTACAACAACACCACTGGCAGAGATGTCTTGGTTATATGGGGTATACATCACC
CTGTCTCCACAGCCGAAGCGAGGGAACTGTATGCAAAAGACGATCCTTACACATTGGCATCTACCAGTTC
ATGGAGCAAGAAGTACAACCTAGAAACTGGAACCCGACCTGGATACAATGGCCAAAAGAGTTGGATGAAG
ATTTACTGGTATCTGATGCACCCCGGGGATTCAATCAGTTTCGAAAGCAGTGGGGGACTACTGGCTCCCA
GATATGGTTACATTATTGAGGAATATGGAAAAGGGCGAATTTTCCAAAGCCGCATTCGCATTGCTAAATG
CAACACTAAATGCCAGACATCGGTTGGTGGAATAAATACTAACAAAACATTTCAAAACATAGAGAGAAAT
GCACTTGGAGATTGCCCGAAATACATAAAATCTGGACAGCTCAAATTGGCCACTGGACTTAGGAATGTAC
CTGCCATATCAAACAGAGGATTGTTTGGGGCTATTGCAGGCTTCATAGAAGGTGGTTGGCCAGGATTAAT
AAATGGTTGGTATGGATTCCAACATCAGAATGAACAAGGAACGGGCATAGCTGCAGACAAAGAATCAACA
CAAAAGGCTATTGATCAAATAACAACTAAGATAAACAATATAATAGAAAAAATGAATGGGAACTATGACT
CAATACGAGGTGAATTCAGTCAGGTGGAACAAAGAATAAATATGCTTGCAGACAGAATAGATGATGCTGT
AACTGACATATGGTCATACAATGCAAAGCTTCTTGTCTTGCTAGAAAACGACAAGACTCTAGACATGCAC
GACGCTAATGTCAGGAACCTGCATGATCAAGTCCGCAGAGTGCTGAGGACCAATGCAATTGATGAGGGGA
ATGGATGTTTTGAACTCCTCCATAAATGTAATGACTCTTGCATGGAGACAATAAGAAATGGAACGTACAA
TCATATAGAGTATGAAGAGGAATCAAAGTTAAAAAGGCAGGAAATAGAAGGTATAAAGCTGAAGTCAGAC
GACAATGTCTACAAAGCATTGTCAATTTACAGCTGCATTGCAAGCAGTATTGTATTGGTAGGACTCATAC
TTGCATTCATCATGTGGGCATGCAGCAGTGGCAGTTGCCGATTCAATATTTGTATATAAGCAGAAAAAC
ACCCTTGTTTCTACT
>iavh1n11922|A/Mute_Swan/Netherlands/2/2022|EPI_ISL_15364797|A/Mute_Swan/Netherlands/2/2022|HA|4||A/Mute_Swan/Netherlands/2/2022
GTTCACTCTGTCAAAATGGAGAGCATAGTACTTCTTCTTGCAATAGTTAGCCTTGTTAAAAGTGATCAGA
TTTGCATTGGTTACCATGCAAATAATTCGACAGAGCAGGTTGACACGATAATGGAAAAGAACGTCACTGT
TACACATGCCCAAGACATATTGGAAAAAACACACAACGGGAAGCTCTGTGATTTAAATGGGGTGAAGCCT
CTGATTTTAAAGGATTGTAGTGTAGCTGGATGGCTCCTCGGAAACCCAATGTGCGACGAATTCATCAGAG
TGCCGGAATGGTCCTACATAGTGGAGCGGGCTAATCCAGCTAATGACCTCTGTTACCCAGGGAGCCTCAA
TGACTATGAAGAACTGAAACACCTGTTGAGCAGAATAAATCATTTTGAGAAGATTCTTATCATCCCCAAG
AGTTCCTGGCCAAATCATGAAACATCACTAGGGGTGAGCGCAGCTTGTCCATACCAGGGGGCGCCCTCCT
TTTTCAGAAATGTGGTGTGGCTTATCAAAAAGAACGATGCATACCCAACAATAAAGATAAGCTACAATAA
TACCAATCGGGAAGATCTCCTGATACTGTGGGGGATTCATCATTCCAACAATGCAGAAGAACAGACAAAT
CTCTATAAAAACCCAACCACCTACATTTCAGTTGGAACATCAACTTTAAACCAGAGGTTGGTACCAAAAA
TAGCTACTAGATCCCAAGTAAACGGGCAACGTGGAAGAATGGACTTCTTCTGGACAATTTTAAAACCAGA
TGATGCAATCCATTTCGAGAGTAATGGAAATTTCATTGCTCCAGAATATGCATATAAAATTGTCAAGAAA
GGGGACTCAACAATTATGAAAAGTGGAGTGGAATATGGCCACTGCAACACCAAATGTCAAACCCCAGTAG
GAGCGATAAATTCTAGTATGCCATTCCACAACATACATCCTCTCACCATTGGGGAATGCCCCAAATACGT
GAAGTCAAACAAGTTGGTCCTTGCGACTGGGCTCAGAAATAGTCCTCTAAGAGAAAAGAGAAGAAAAAGA
GGCCTGTTTGGGGCGATAGCAGGGTTTATAGAGGGAGGATGGCAGGGAATGGTTGATGGTTGGTATGGGT
ACCATCATAGCAATGAGCAGGGGAGTGGGTACGCTGCAGACAAAGAATCCACCCAAAAGGCAATAGATGG
AGTTACCAATAAGGTCAACTCAATCATTGACAAAATGAACACTCAATTTGAGGCAGTTGGAAGGGAGTTT
AATAACTTAGAAAGGAGGATAGAGAATCTGAACAAGAAAATGGAAGACGGATTCCTAGATGTCTGGACCT

ATAATGCTGAACTTCTAGTTCTCATGGAAAACGAGAGGACTCTAGATTTCCATGATTCAAATGTCAAGAA
CCTTTACGACAAAGTAAGACTACAGCTTAGGGATAATGCAAAGGAGCTTGGTAATGGCTGTTTCGAATTC
TATCACAAATGCGATAATGAATGTATGGAAAGTGTGAGAAATGGGACGTATGACTACCCTCAGTATTCAG
AAGAAGCAAGATTAAAAAGAGAAGAAATAAGCGGAGTGAAATTAGAATCAATAGGAACTTACCAGATACT
GTCAATTTATTCAACAGCGGCGAGTTCCCTAGCACTGGCAATCATGATAGCTGGTCTATCTTTATGGATG
TGCTCCAATGGGTCGTTACAGTGCAGAATTTGCATTTAGATTTGTGAGCTCAGATTGTAGTTAAAAACAC
>iavh1n11922|A/Barry/3792/2022|EPI_ISL_15391170|A/Barry/3792/2022|HA|4||A/Barry/3792/2022
AGCAAAAGCAGGGGATAATTCTATTAACCATGAAGGCTATCATTGCTTTGAGCAACATTCTATGTCTTGT
TTTCGCTCAAAAAATACCTGGAAATGACAATAGCACGGCAACGCTGTGCCTTGGGCACCATGCAGTACCA
AACGGAACGATAGTGAAAACAATCACAAATGACCGAATTGAAGTTACTAATGCTACTGAGTTGGTTCAGA
ATTCATCAATAGGTAAAATATGCAACAGTCCTCATCAGATCCTTGATGGAGGGAACTGCACACTAATAGA
TGCTCTATTGGGGGACCCTCAGTGTGACGGCTTTCAAAATAAGGAATGGGACCTTTTTGTTGAACGAAGC
AGAGCCAACAGCAGCTGTTACCCTTATGATGTGCCGGATTATGCCTCCCTTAGGTCACTAGTTGCCTCAT
CCGGAACACTGGAGTTTAAAAATGAAAGCTTCAATTGGACCGGAGTCAAACAAAACGGAACAAGTTCTGC
GTGCAAAAGGGGATCTAGTAGTAGTTTTTTTAGTAGATTAAATTGGTTGACCAGCTTAAACAACATATAT
CCAGCACAGAACGTGACTATGCCAAACAAGGAACAATTTGACAAATTGTACATTTGGGGGGTTCACCACC
CGGATACGGACAAGAACCAATTCTCCCTGTTTGCTCAATCATCAGGAAGAATCACAGTATCTACCAAAAG
AAGCCAACAAGCTGTAATCCCAAATATCGGATCTAGACCCAGAGTAAGGGATATCCCTAGCAGAATAAGC
ATCTATTGGACAATAGTAAAACCGGGAGACATACTTTTGATTAACAGCACAGGGAATCTAATTGCTCCTA
GGGGTTACTTCAAAATACGAAGTGGGAAAAGCTCAATAATGAGATCAGACGCACCCATTGGCAAATGTAA
GTCTGAATGCATCACTCCAAATGGAAGCATTCCCAATGACAAACCGTTCCAAAATGTAAACAGGATCACA
TACGGGGCCTGTCCCAGATACGTTAAGCAAAGCACCTTGAAATTGGCAACAGGAATGCGAAATGTACCTG
AGAAACAAACCAGAGGCATATTTGGTGCAATAGCGGGTTTCATAGAAAATGGATGGGAGGGAATGGTGGA
TGGTTGGTACGGTTTCAGGCATCAAAATTCTGAGGGAAGAGGACAAGCAGCAGATCTCAAAAGCACTCAA
GCAGCAATCGATCAAATCAGTGGGAAGCTGAATCGATTGATCGGAAAAACCAACGAGAAATTCCATCAGA
TTGAAAAAGAATTCTCAGAAGTAGAAGGAAGAGTTCAAGACCTCGAGAAATATGTTGAGGACACTAAAAT
AGATCTCTGGTCATACAACGCGGAGCTTCTTGTTGCCCTGGAGAACCAACATACGATTGACCTAACTGAC
TCAGAAATGAACAAGCTGTTTGAAAAAACAAAGAAGCAACTGAGGGAAAATGCTGAGGATATGGGAAATG
GTTGTTTCAAAATATACCACAAATGTGACAATGCCTGCATAGGATCAATAAGAAATGAAACTTATGACCA
CAATGTGTACAGGGATGAAGCATTAAACAACCGGTTCCAGATCAAGGGAGTTGAGCTGAAGTCAGGGTAC
AAAGATTGGATCCTATGGATTTCCTTTGCCATGTCATGTTTTTTGCTTTGTATTGCTTTGTTGGGGTTCA
TCATGTGGGCCTGCCAAAAGGGCAACATTAGATGCAACATTTGCATTTGAGTGCATTAATTAAAAACACC
CTTGTTTCTACT
>NDN12092022_ H13N6_HA_4
AGCAAAAGCAGGGGAGGATTCACCAAACCAATACGAGAGAAATGGAAGTCCCGGTATTCGCACTCCTAGT
GTTAACCAGCATATGCGTACAAGCCGATAGGATTTGAGCTGGGTACTCAAGCACAAACTCATCAGAAAGG
GTTTATACACTGTTAGAAAATAATGTCCCGGTTACAAGCTCTGTTGATTTGGTTGAGACTAACCACACAG
GAACATATTGTTCTTTGGGTGGAATTAGTCCAGTGCACTTGGGGGACAGTAGCTTTGAAGGCTGGATTCT
AGGGAACCCTGCCTGTGACAGTAACCTGGGGGTTAGAGAATGGTCATATTTGATTGAAGACCCTTGTGCT
CCTCATGGATTGTGCTACCCAGGAGAGTTAGACAACAATGGAGAATTGAGACATTTGTTTAGTGGGATCA
AATCTTTCAGTAGGACAGAATTGATCGCACCTCCCTCTCTGGGGGAAGTGAATGATGGAGTAACAGCTGC
CTGTGAAGACAAAAGAGCTAGCAGCTTTTACCGAAACTTAGTATGGTTTGTGAAGAGAGGGAGTAACTAC
CCTGTAATCCGCTGGACCTACAACAACACCACTGGCAGAGATGTCTTGGTTATATGGGGTATTCATCACC
CTGTCTCCACAACCGAAACGAGGGACCTGTATGCAAAAGACAACCCATACACATTGGTATCTACCAGTTC
ATGGAGCAAGAAGTACAGCCTAGAAACTGGAACCCGACCTGGATACAATGGCCAAAAGAGTTGGATGAAG
ATTTACTGGCATCTGATGCACCCCGGGGAGTCAATCAGTTTCGAAAGCAATGGGGGATTGCTGGCTCCAA

AATATGGTTACATTGTTGAGGAATATGGCAAAGGGCGAATTTTCCAAAGCCGTATTCGCATTGCTAAATG
TAACACTAAATGCCAGACATCGGTTGGTGGAATAAATACTAACAAAACATTTCAAAACATAGAAAGAAAT
GCACTCGGAGATTGCCCGAAATACATAAAATCTGGACAGCTCAAATTGGTCACTGGACTTAGGAATGTAC
CTGCCATATCAAACAGAGGATTGTTTGGGGCTATTGCAGGCTTCATAGAAGGTGGTTGGCCAGGATTAAT
AAATGGATGGTATGGATTCCAACATCAGAATGAACAGGGAACGGGCATAGCTGCAGACAAAGAATCAACA
CAAAAGGCTATTGACCAAATAACAACTAAAATAAACAATATAATAGAAAAAATGAATGGGAACTATGACT
CAATACGAGGGGAATTCAGTCAAGTGGGGAAGAGGATAAATATGCTGGCAGACAGAATAGATGATGCTGT
AACTGACATATGGTCATACAATGCGAAGCTTCTTGTCTTGTTAGAAAACGATAAGACTCTAGACATGCAC
GACGCTAATGTCAGGAACCTGCATGATCAGGTCCGCAGAGTGCTGAGGGCCAACGCAATCGATGAGGGGA
ATGGGTGTTTTGAACTCCTCCATAAATGTAATGATTCTTGCATGGAGACAATAAGAAATGGAACGTACAA
CCATATAGAGTATGAAGAGGAATCAAAGTTAAAAAGGCAGGAAATAGAAGGTATAAAGCTGAAGTCAGAC
GACGATGTCTGCAAAGCATTGTCAATTTACAGCTGCATTGCAAGCAGTATTGTATTGGTAGGACTCATAC
TTGCATTCATCATGTGGGCATGCAGCAGTGGCAATTGCCGATTCAATATTTGTATATAGGCAGAAAAAAC
ACCCTTGTTTCTACT
>NC_026436.1 Influenza A virus (A/California/07/2009(H1N1)) segment 5 nucleocapsid protein (NP) gene, complete cds
ATGGCGTCTCAAGGCACCAAACGATCATATGAACAAATGGAGACTGGTGGGGAGCGCCAGGATGCCACAG
AAATCAGAGCATCTGTCGGAAGAATGATTGGTGGAATCGGGAGATTCTACATCCAAATGTGCACTGAACT
CAAACTCAGTGATTATGATGGACGACTAATCCAGAATAGCATAACAATAGAGAGGATGGTGCTTTCTGCT
TTTGATGAGAGAAGAAATAAATACCTAGAAGAGCATCCCAGTGCTGGGAAGGACCCTAAGAAAACAGGAG
GACCCATATATAGAAGAGTAGACGGAAAGTGGATGAGAGAACTCATCCTTTATGACAAAGRAGAAATAAG
GAGAGTTTGGCGCCTAGCAAACAATGGCGAAGATGCAACAGCAGGTCTTACTCATATCATGATTTGGCAT
TCCAACCTGAATGATGCCACATATCAGAGAACAAGAGCGCTTGTTCGCACCGGAATGGATCCCAGAATGT
GCTCTCTAATGCAAGGTTCAACACTTCCCAGAAGGTCTGGTGCCGCAGGTGCTGCGGTGAAAGGAGTTGG
AACAATAGCAATGGAGTTAATCAGAATGATCAAACGTGGAATCAATGACCGAAATTTCTGGAGGGGTGAA
AATGGACGAAGGACAAGGGTTGCTTATGAAAGAATGTGCAATATCCTCAAAGGAAAATTTCAAACAGCTG
CCCAGAGGGCAATGATGGATCAAGTAAGAGAAAGTCGAAACCCAGGAAACGCTGAGATTGAAGACCTCAT
TTTCCTGGCACGGTCAGCACTCATTCTGAGGGGATCAGTTGCACATAAATCCTGCCTGCCTGCTTGTGTG
TATGGGCTTGCAGTAGCAAGTGGGCATGACTTTGAAAGGGAAGGGTACTCACTGGTCGGGATAGACCCAT
TCAAATTACTCCAAAACAGCCAAGTGGTCAGCCTGATGAGACCAAATGAAAACCCAGCTCACAAGAGTCA
ATTGGTGTGGATGGCATGCCACTCTGCTGCATTTGAAGATTTAAGAGTATCAAGTTTCATAAGAGGAAAG
AAAGTGATTCCAAGAGGAAAGCTTTCCACAAGAGGGGTCCAGATTGCTTCAAATGAGAATGTGGAAACCA
TGGACTCCAATACCCTGGAACTGAGAAGCAGATACTGGGCCATAAGGACCAGGAGTGGAGGAAATACCAA
TCAACAAAAGGCATCCGCAGGCCAGATCAGTGTGCAGCCTACATTCTCAGTGCAGCGGAATCTCCCTTTT
GAAAGAGCAACCGTTATGGCAGCATTCAGCGGGAACAATGAAGGACGGACATCCGACATGCGAACAGAAG
TTATAAGAATGATGGAAAGTGCAAAGCCAGAAGATTTGTCCTTCCAGGGGCGGGGAGTCTTCGAGCTCTC
GGACGAAAAGGCAACGAACCCGATCGTGCCTTCCTTTGACATGAGTAATGAAGGGTCTTATTTCTTCGGA
GACAATGCAGAGGAGTATGACAGTTGA
>MF575100.1 Influenza A virus (A/black-headed gull/Netherlands/31/2014(H13N6)) segment 5 nucleocapsid protein (NP) gene, complete cds
AGCAAAAGCAGGGTAGATAATCACTCACTGAGTGACATCCACATCATGGCGTCTCAAGGCACCAAACGAT
CCTATGAGCAGATGGAAACTGGTGGTGAACGCCAGAATGCCACTGAGATTCGGGCATCTGTCGGAAGGAT
GGTTGGAGGAATCGGAAGATTCTACATACAGATGTGCACTGAACTCAAACTGAGTGACAATGAAGGAAGG
CTGCTACAAAACAGTATCACAATAGAGAGAATGGTCCTGTCTGCATTTGATGAGAGGAGGAACAGATACT
TGGAGGAGCATCCCAGCGCTGGGAGGGACCCCAAGAAAACTGGTGGACCAATTTACAGGAGGAGAGAGGG
GAAATGGGTGAGAGAATTGGTCCTATATGACAAGGAAGAAATAAGAAGAATCTGGCGACAGGCAAACAAT
GGAGAAGATTCGACTGCTGGCCTCACTCATTTGATGATCTGGCATTCTAATTTGAATGACGCCACATATC
AGAGGACTAGAGCCCTAGTGCGCACCGGGATGGACCCCAGGATGTGCTCCCTCATGCAAGGATCGACACT

CCCAAGAAGGTCTGGAGCGGCTGGTGCAGCTGTAAAAGGAGTTGGGACAATGGTGATGGAGCTCATCAGA
ATGATAAAAAGAGGGGTTAATGACCGTAATTTCTGGAGAGGTGAAAATGGAAGAAGAACAAGAATTGCTT
ATGAGAGAATGTGCAACATCCTCAAAGGGAAATTCCAAACGGCAGCACAACGAGCTATGATGGATCAGGT
CAGAGAAAGCCGTAATCCTGGAAATGCTGAAATTGAAGACCTCATCTTTTTGGCCAGGTCTGCCCTTATT
CTGAGGGGGGCAGTAGCTCATAAATCATGCCTGCCTGCCTGTGTGTATGGACTTGCTGTAGCAAGTGGAT
ATGAATTCGAGAGGGAAGGATATTCCCTTGTTGGGATAGACCCTTTTCGTCTGCTCCAGAACAGCCAAGT
GTTCAGTCTAATCCGACCCAATGAAAATCCGGCACACAAGAGCCAATTGGTATGGATGGCATGTCATTCT
GCTGCATTTGAGGATTTGAGAGTGTCAAGCTTCATCAGAGGAGCAAGAGTGTTACCAAGAGGGCAACTAT
CCACAAGAGGTGTTCAAATTGCGTCCAACGAGAACATGGAGACTATGAGTTCCAGCACCCTTGAATTAAG
GAGCAAATACTGGGCAATAAGGACTAGAAGCGGAGGAAACACCAGCCAACAAAGAGCATCAGCAGGGCAA
ATCAGTGTACAACCTACTTTTTCAGTACAAAGAAACCTCCCTTTTGAAAGAGCGACAATCATGGCTGCAT
TCACAGGAAATGCAGAAGGAAGAACATCCGATATGAGGACTGAGATCATACGGATGATGGAAAGTGCAAG
ACCAGAAGATGTGTCTTTCCAGGGGCGGGGAGTCTTCGAGCTCTCGGACGAAAAGGCCACGAACCCGATC
GTGCCTTCCTTTGACATGAGTAAAGAAGGATCTTATTTCTTCGGAGACAACGCTGAGGAGTTTGACAGTT
GAAGAAAAATACCCTTGTTTCTACT
>iavh1n11922|A/Mute_Swan/Netherlands/2/2022|EPI_ISL_15364797|A/Mute_Swan/Netherlands/2/2022|NP|5||A/Mute_Swan/Netherlands/2/2022
GTAGATAATCACTCACTGAGTGACATCAACATCATGGCGTCTCAAGGCACCAAACGATCTTATGAACAGA
TGGAAACTGGTGGGGAGCGCCAGAATGCCACTGAGATCAGAGCATCTGTTGGGAGAATGGTTGGTGGAAT
TGGGAGGTTCTACATACAGATGTGCACTGAGCTCAAACTCAGCGACTATGAAGGAAGGCTGATCCAGAAC
AGCATAACAATAGAGAGAATGGTTCTCTCTGCATTTGATGAAAGGAGGAACAAATATCTGGAAGAACATC
CCAGTGCGGGGAAGGACCCGAAGAAAACTGGAGGTCCAATTTATCGAAGGAGAGATGGGAAATGGATGAG
AGAACTGATCCTGTATGACAAAGAGGAGATCAGGAGAATCTGGCGTCAAGCGAATAATGGAGAAGACGCA
ACTGCTGGTCTCACTCACCTGATGATCTGGCATTCCAATCTAAATGATGCCACATACCAGAGGACAAGAG
CTCTCGTGCGGACTGGGATGGACCCCAGGATGTGCTCTCTTATGCAAGGATCAACTCTCCCAAGGAGGTC
TGGAGCTGCTGGTGCAGCAGTAAAGGGAGTCGGGACGATGGTGATGGAACTAATTCGGATGATAAAGCGA
GGAATTAATGATCGGAACTTCTGGAGAGGCGAGAACGGGCGAAGGACAAGGATTGCATATGAGAGAATGT
GCAACATCCTCAAAGGGAAATTCCAAACAGCATCACAAAGAGCAATGATGGATCAGGTGCGTGAAAGCAG
GAATCCTGGCAATGCTGAAATTGAAGATCTCATCTTTCTGGCACGGTCTGCACTCATCCTGAGAGGATCA
GTGGCTCATAAGTCCTGCTTGCCTGCTTGTGTGTACGGACTCGCTGTGGCCAGTGGATACGACTTTGAGA
GAGAAGGGTACTCCCTAGTTGGAATAGATCCTTTCCGTCTGCTTCAAAACAGCCAGGTCTTCAGTCTCAT
TAGACCAAATGAGAATCCAGCACATAAGAGTCAATTGGTGTGGATGGCATGTCATTCTGCAGCATTCGAA
GATCTAAGAGTCTCAAGTTTCATCAGAGGAACAAGAGTAGTTCCAAGAGGACAACTATCCACCAGAGGAG
TTCAAATTGCTTCAAATGAGAACATGGAAACAATGGACTCCAGCACTCTTGAACTGAGAAGCAGATATTG
GGCTATAAGAACCAGGAGTGGAGGAAACACCAACCAACAGAGAGCATCTGCAGGACAAATCAGTGTACAG
CCCACTTTTTCTGTACAGAGAAATCTTCCCTTCGAAAGAGCGACCATTATGGCGGCGTTCACAGGGAATA
CTGAGGGCAGAACATCCGACATGAGGACTGAAATCATAAGAATGATGGAAAGTGCCAGACCAGAAGATGT
GTCTTTCCAGGGGCGGGGAGTCTTCGAGCTCTCGGACGAAAAGGCAACGAACCCGATCGTGCCTTCCTTT
GACATGAGTAATGAAGGATCTTATTTCTTCGGAGACAATGCAGAGGAGTATGACAATTAAAGAAAAATAC
>iavh1n11922|A/Barry/3792/2022|EPI_ISL_15391170|A/Barry/3792/2022|NP|5||A/Barry/3792/2022
CAGCAAAAGCAGGGTTGATAATCACTCACTGAGTGACATCAAAATCATGGCGTCCCAAGGCACCAAACGG
TCTTATGAACAGATGGAAACTGATGGAGACCGCCAGAATGCAACTGAGATTAGGGCATCCGTCGGGAAGA
TGATTGATGGGATTGGGAGATTCTACATCCAAATGTGCACTGAACTTAAACTCAGTGATCATGAAGGACG
GTTGATCCAAAATAGCTTGACAATAGAGAAAATGGTACTCTCTGCTTTTGATGAAAGAAGGAATAAATAC
TTGGAAGAACACCCCAGCGCGGGGAAAGATCCCAAGAAAACTGGGGGGCCCATATACAGAAGAGTCGATG
GAAAATGGATGAGGGAACTCGTCCTTTATGACAAAGAGGAAATAAGGCGAATCTGGCGCCAAGCCAACAA
TGGTGAGGATGCTACATCTGGTCTAACCCACATGATGATTTGGCATTCCAATCTGAATGATGCGACATAC

CAGAGGACAAGAGCTCTTGTTCGGACTGGAATGGATCCCAGAATGTGCTCTCTGATGCAGGGGTCGACTC
TCCCTAGAAGGTCCGGAGCTGCAGGTGCTGCAGTCAAAGGAATCGGAACAATGGTGATGGAACTGATCAG
GATGATCAAACGGGGGATCAACGATCGAAATTTTTGGAGAGGTGAGAATGGGCGGAAAACAAGAAGTGCT
TATGATAGAATGTGCAACATTCTTAAAGGAAAATTTCAAACAGCTGCACAAAGAGCAATGGTGGATCAAG
TTAGAGAAAGTCGGAACCCAGGAAACGCTGAGATCGAAGATCTCATATTTTTGGCAAGATCTGCACTGAT
ATTGAGAGGATCAGTTGCTCACAAATCTTGCCTACCTGCCTGTGCATATGGACCTGCAGTATCCAGTGGT
TACGACTTTGAAAAAGAGGGATATTCCTTGGTGGGAATAGACCCTTTCAAACTACTTCAAAATAGCCAAA
TATACAGCTTAATCAGACCTAATGAGAATCCAGCACACAAGAGTCAGCTGGTGTGGATGGCATGCCATTC
TGCTGCATTTGAAGATTTAAGATTGTTAAGTTTCATCAGAGGGACAAAAGTATATCCTCGGGGGAAACTG
TCAACTAGAGGAGTACAAATTGCTTCAAATGAGAACATGGATAATATGGGATCAAGCACTCTTGAACTGA
GAAGCGGGTACTGGGCCATAAGGACCAGGAGCGGAGGAAACACTAATCAACAGAGGGCCTCCGCAGGCCA
AACCAGTGTGCAACCTACATTTTCTGTACAAAGAAACATCCCATTTGAGAAGTCAACCATCATGGCAGCA
TTCACTGGAAATACAGAGGGAAGAACTTCAGACATGAGGGCAGAAATCATAAGGATGATGGAAGGTGCAA
AACCAGAAGAAGTGTCATTCCGGGGGAGGGGAGTTTTCGAGCTCTCAGACGAGAAGGCAGCGAACCCGAT
CGTGCCCTCTTTTGATATGAGCAACGAAGGATCTTATTTCTTCGGAGACAATGCAGAAGAGTATGACAAT
TAAGAAAAAAATACCCTTGTTTCTACTGATC
>NDN12092022_ H13N6_NP_5
AGCAAAAGCAGGGTAGATAATCACTCACTGAGTGACATCCACATCATGGCGTCTCAGGGCACCAAACGAT
CCTATGAGCAGATGGAAACTGGTGGTGAACGCCAGAATGCCACTGAGATTCGAGCATCTGTCGGAAGGAT
GGTTGGAGGAATCGGAAGATTCTACATACAGATGTGCACTGAACTCAAACTGAGTGACAATGAAGGAAGG
CTGATACAAAACAGTATCACAATAGAGAGGATGGTCCTGTCTGCATTTGATGAGAGGAGGAACAGATACT
TGGAGGAGCATCCCAGTGCTGGGAGGGACCCAAAGAAAACTGGTGGACCAATTTACAGAAGGAGAGAAGG
GAAATGGGTGAGAGAATTGGTCCTATATGACAAGGAAGAAATAAGAAGAATCTGGCGACAGGCAAACAAT
GGAGAAGATTCGACTGCTGGCCTCACTCATTTGATGATCTGGCATTCTAATTTGAATGATGCCACATATC
AGAGGACCAGAGCCCTAGTGCGTACCGGGATGGACCCCAGGATGTGCTCCCTCATGCAGGGATCGACACT
CCCAAGAAGGTCTGGAGCGGCTGGTGCAGCTGTGAAAGGAGTTGGGACAATGGTGATGGAGCTCATCAGA
ATGATAAAAAGAGGGGTTAATGACCGTAACTTCTGGAGAGGTGAAAATGGGAGAAGAACAAGAATTGCTT
ATGAGAGGATGTGCAACATCCTCAAAGGGAAATTCCAAACAGCAGCACAACGAGCTATGATGGACCAGGT
CAGAGAAAGCCGTAATCCTGGAAATGCTGAAATTGAAGACCTCATCTTTTTGGCCAGGTCTGCCCTTATT
CTGAGGGGAGCAGTAGCTCATAAATCATGCCTGCCTGCCTGCGTGTATGGACTTGCTGTAGCAAGTGGGT
ATGACTTCGAAAGGGAAGGATATTCCCTTGTTGGGATAGACCCTTTCCGTCTGCTCCAAAACAGCCAAGT
GTTCAGTCTAATCCGACCCAATGAAAACCCAGCACACAAGAGCCAATTGGTATGGATGGCATGTCATTCT
GCTGCATTTGAGGATTTGAGAGTGTCAAGCTTCATCAGGGGAGCAAGAGTGTTGCCAAGAGGGCAACTAT
CCACAAGAGGTGTTCAAATTGCGTCCAACGAGAACATGGAGACTATGAGTTCCAGCACACTTGAATTAAG
GAGCAAATACTGGGCAATAAGGACTAGAAGCGGAGGAAACACCAACCAACAAAGAGCATCAGCAGGGCAA
ATCAGTGTACAACCTACTTTTTCAGTACAAAGAAACCTCCCCTTTGAAAGAGCGACAATCATGGCTGCAT
TCACAGGAAATGCAGAAGGAAGAACATCCGATATGAGGACTGAGATCATACGGATGATGGAAAGTGCAAG
ACCAGAAGATGTGTCTTTCCAGGGGCGGGGAGTCTTCGAGCTCTCAGACGAAAAGGCCACGAACCCGATC
GTGCCTTCCTTTGACATGAGTAAAGAAGGATCTTATTTCTTCGGAGACAATGCTGAGGAGTTTGACAGTT
GAAGAAAAATACCCTTGTTTCTACT
>ENN12092022_ H13N6_NP_5
AGCAAAAGCAGGGTAGATAATCACTCACTGAGTGACATCCACATCATGGCGTCTCAAGGCACCAAACGAT
CTTATGAGCAGATGGAAACTGGTGGAGAACGCCAGAATGCCACTGAGATTCGGGCATCTGTCGGAAGGAT
GGTTGGAGGAATCGGAAGATTCTACATACAGATGTGCACTGAACTCAAATTGAGTGACAATGAGGGAAGG
CTGATACAAAACAGTATCACAATAGAGAGGATGGTCCTGTCTGCATTTGATGAGAGGAGGAACAGATATT
TGGAGGAGCATCCCAGTGCTGGGAGAGACCCCAAGAAAACTGGTGGACCAATTTACAGAAGAAGAGAAGG

GAAATGGGTGAGAGAATTGGTCCTATATGACAAGGAAGAGATAAGACGAATCTGGCGGCAGGCAAACAAT
GGAGAAGATTCGACTGCTGGCCTCACCCATCTGATGATCTGGCATTCTAATTTGAATGATGCCACATACC
AGAGGACCAGAGCCCTAGTGCGCACCGGGATGGACCCCAGGATGTGCTCCCTCATGCAGGGATCGACACT
CCCAAGGAGGTCTGGGGCGGCTGGTGCAGCTGTAAAAGGGGTTGGGACAATGGTAATGGAGCTCATCAGA
ATGATCAAAAGAGGGGTTAATGACCGAAACTTCTGGAGAGGTGAAAATGGGAGAAGAACAAGAATTGCTT
ATGAGAGGATGTGCAACATCCTCAAAGGGAAATTCCAAACAGCAGCACAGCGAGCTATGATGGACCAGGT
CAGAGAAAGCCGTAATCCTGGTAATGCTGAGATTGAAGACCTCATATTTTTGGCAAGGTCTGCCCTTATT
CTAAGGGGAGCTGTGGCTCATAAATCATGTCTGCCTGCCTGCGTGTATGGACTTGCTGTAGCAAGTGGAT
ATGACTTCGAAAGGGAAGGATATTCCCTTGTTGGGATAGACCCTTTCCGTCTGCTCCAAAACAGCCAAGT
GTTTAGTCTAATCCGACCAAATGAAAATCCAGCACACAAGAGCCAATTGGTATGGATGGCGTGTCATTCT
GCTGCATTTGAGGATTTGAGAGTGTCAAGCTTCATCAGGGGAACAAGAGTGTTGCCACGAGGGCAACTAT
CTACAAGAGGGGTTCAAATTGCATCCAACGAGAACATGGAGACTATGAGTTCCAGCACACTTGAACTAAG
GAGCAAGTACTGGGCAATAAGGACTAGAAGCGGAGGAAATACCAACCAACAAAGAGCATCAGCAGGACAA
ATCAGTGTACAACCTACCTTTTCAGTACAAAGGAACCTCCCTTTTGAAAGAGCGACAATCATGGCTGCAT
TCACAGGAAATGCAGAAGGAAGAACATCCGATATGAGGACTGAGATCATACGGATGATGGAAAGTGCAAG
ACCAGAAGATGTGTCTTTCCAGGGGCGGGGAGTCTTCGAGCTCTCAGACGAAAAGGCCACGAACCCGATC
GTGCCTTCCTTTGACATGAGTAAAGAAGGATCTTATTTCTTCGGAGACAATGCTGAGGAGTTTGACAGTT
GAAGAAAAATACCCTTGTTTCTACT
>BEL12092022_ H13N6_NP_5
AGCAAAAGCAGGGTAGATAATCACTCACTGAGTGACATCCACATCATGGCGTCTCAAGGCACCAAACGAT
CTTATGAGCAGATGGAAACTGGTGGTGAACGCCAGAATGCCACTGAGATTCGGGCATCTGTCGGAAGGAT
GGTTGGAGGAATCGGAAGATTCTACATACAGATGTGCACTGAACTCAAACTGAGTGACAATGAAGGAAGG
CTGATACAAAACAGTATCACAATAGAGAGGATGGTCCTGTCTGCATTTGATGAGAGGAGGAACAGATACT
TGGAGGAGCATCCCAGTGCTGGGAGGGACCCCAAGAAAACTGGTGGACCAATTTACAGAAGGAGAGAGGG
GAAATGGGTGAGAGAATTGGTCCTATATGACAAGGAAGAAATAAGAAGAATCTGGCGACAGGCAAACAAT
GGAGAAGATTCGACTGCTGGCCTCACCCATTTGATGATCTGGCATTCTAATTTGAATGATGCCACATATC
AGAGGACCAGAGCCCTAGTGCGCACCGGGATGGACCCCAGGATGTGCTCCCTCATGCAGGGATCGACACT
CCCAAGAAGGTCTGGAGCGGCTGGTGCAGCTGTAAAAGGAGTTGGGACAATGGTGATGGAGCTCATCAGA
ATGATAAAAAGAGGGGTTAATGACCGTAACTTCTGGAGAGGTGAAAATGGGAGAAGAACAAGAATTGCTT
ATGAGAGGATGTGCAACATCCTCAAAGGGAAATTCCAAACAGCAGCACAACGAGCTATGATGGACCAGGT
CAGAGAAAGCCGTAATCCTGGAAATGCTGAAATTGAAGACCTCATCTTTTTGGCCAGGTCTGCCCTTATT
CTAAGGGGAGCTGTAGCTCATAAATCATGCCTGCCTGCCTGTGTGTATGGACTTGCTGTAGCAAGTGGAT
ATGACTTCGAAAGGGAAGGATATTCCCTTGTTGGGATAGACCCTTTCCGTCTGCTCCAAAACAGCCAAGT
GTTCAGTCTAATCCGACCCAATGAAAATCCAGCACACAAGAGCCAATTGGTATGGATGGCATGTCATTCT
GCTGCATTTGAGGATTTGAGAGTGTCAAGCTTCATCAGGGGAGCAAGAGTGTTGCCAAGAGGGCAACTAT
CCACAAGAGGTGTTCAAATTGCGTCCAACGAGAACATGGAGACTATGAGTTCCAGCACACTTGAATTAAG
GAGCAAATACTGGGCAATAAGGACTAGAAGCGGAGGAAACACCAACCAACAAAGAGCATCAGCAGGACAA
ATCAGTGTACAACCTACTTTTTCAGTACAAAGAAACCTCCCTTTTGAAAGAGCGACAATCATGGCTGCAT
TCACAGGAAATGCAGAAGGAAGAACATCCGATATGAGGACTGAGATCATACGGATGATGGAAAGTGCAAG
ACCAGAAGATGTGTCTTTCCAGGGGCGGGGAGTCTTCGAGCTCTCAGACGAAAAGGCCACGAACCCGATC
GTGCCTTCCTTTGACATGAGTAAAGAAGGATCTTATTTCTTCGGAGACAATGCTGAGGAGTTTGACAGTT
GAAGAAAAATACCCTTGTTTCTACT
>BEL10102022_ H1N1_NP_5
ATGGCGTCTCAAGGCACCAAACGATCATATGAACAAATGGAGACTGGTGGGGAGCGCCAGGATACCACAG
AAATCAGAGCATCTGTTGGAAGAATGATTGGTGGAATCGGGAGATTCTATATCCAAATGTGCACTGAACT
AAAACTCAGTGATTATGATGGACGACTAATCCAGAACAGCATAACAATAGAGAGGATGGTGCTTTCTGCT

TTTGATGAGAGAAGAAATAAATACCTAGAAGAGCATCCAAGTGCTGGGAAGGACCCTAAGAAAACAGGAG
GACCCATCTATAGAAGAATAGACGGAAAATGGACAAGAGAACTCATCCTTTATGACAAAGNAGAAATAAG
GAGAGTTTGGCGCCAAGCAAACAATGGCGAAGATGCAACAGCAGGTCTTACTCATATCATGATTTGGCAT
TCCAATCTGAATGATGCCACATATCAGAGGACAAGAGCACTTGTTCGCACTGGAATGGATCCCAGAATGT
GCTCTCTAATGCAAGGTTCAACACTTCCCAGAAGGTCAGGTGCCGCAGGTGCTGCAGTAAAAGGAGTTGG
AACAATAGCTATGGAGTTAATCAGAATGATAAAACGTGGAATCAATGACCGAAATTTCTGGAGGGGTGAA
AATAGACGAAGGACAAGAGTTGCTTATGAAAGAATGTGCAATATCCTCAAAGGAAAGTTTCAAACAGCTG
CTCAGAGGGCAATGATGGATCAAGTAAGGGAAAGCCGAAACCCAGGAAACGCTGAGATTGAAGACCTCAT
TTTCCTGGCGCGGTCAGCACTCATTCTGAGAGGATCAGTTGCACATAAATCCTGCCTGCCTGCTTGTGTG
TATGGGCTTGCAGTAGCAAGTGGCCATGACTTTGAAAGGGAAGGGTACTCATTGGTCGGGATAGACCCGT
TCAAATTACTCCAAAACAGTCAAGTGGTCAGCCTGATGAGACCAAATGAAAATCCAGCTCACAAGAGTCA
ATTGGTATGGATGGCATGCCACTCTGCTGCATTTGAAGATTTAAGAGTATCAAGTTTCATAAGAGGAAAG
AAGGTGATCCCAAGAGGAAAGCTTTCCACAAGAGGGGTTCAGATTGCTTCAAATGAGAATGTGGAAACCA
TGGACTCCAATACCATGGAACTAAGAAGCAGATACTGGGCCATAAGAACCAGGAGTGGAGGAAATACCAA
TCAACAGAAGGCATCTGCAGGCCAGATCAGTGTGCAGCCTACATTCTCAGTGCAGCGAAATCTCCCCTTT
GAAAGAGCAACCATTATGGCAGCATTCAGCGGGAACAATGAAGGACGGACATCCGACATGCGAACAGAAG
TTATAAGAATGATGGAAAGTGCAAAGCCAGAGGATTTGTCCTTCCAGGGGCGGGGAGTCTTCGAGCTCTC
GGACGAAAAGGCAACGAACCCGATCGTGCCTTCCTTTGACATGAGTAACGAAGGGTCTTATTTCTTCGGA
GACAATGCAGAGGAGTATGACAATTGA
>BEL28112022_ H1N1_NP_5
ATGGCGTCTCAAGGCACCAAACGATCATATGAACAAATGGAGACTGGTGGGGAGCGCCAGGATACCACAG
AAATCAGAGCATCTGTTGGAAGAATGATTGGTGGAATCGGGAGATTCTATATCCAAATGTGCACTGAACT
AAAACTCAGTGATTATGATGGACGACTAATCCAGAACAGCATAACAATAGAGAGGATGGTGCTTTCTGCT
TTTGATGAGAGAAGAAATAAATACCTAGAAGAGCATCCAAGTGCTGGGAAGGACCCTAAGAAAACAGGAG
GACCCATCTATAGAAGAATAGACGGAAAATGGACAAGAGAACTCATCCTTTATGACAAAGNAGAAATAAG
GAGAGTTTGGCGCCAAGCAAACAATGGCGAAGATGCAACAGCAGGTCTTACTCATATCATGATTTGGCAT
TCCAATCTGAATGATGCCACATATCAGAGGACAAGAGCACTTGTTCGCACTGGAATGGATCCCAGAATGT
GCTCTCTAATGCAAGGTTCAACACTTCCCAGAAGGTCCGGTGCCGCAGGTGCTGCAGTAAAAGGAGTTGG
AACAATAGCTATGGAGTTAATCAGAATGATAAAACGTGGAATCAATGACCGAAATTTCTGGAGGGGTGAA
AATGGACGAAGGACAAGAGTTGCTTATGAAAGAATGTGCAATATCCTCAAAGGAAAATTTCAAACAGCTG
CTCAGAGGGCAATGATGGATCAAGTAAGGGAAAGCCGAAACCCAGGAAACGCTGAGATTGAAGACCTCAT
TTTCCTGGCGCGGTCAGCACTCATTCTGAGAGGATCAGTTGCACATAAATCCTGCCTGCCTGCTTGTGTG
TATGGGCTTGCAGTAGCAAGTGGCCATGACTTTGAAAGGGAAGGGTACTCATTGGTCGGGATAGACCCGT
TCAAATTACTCCAAAACAGTCAAGTGGTCAGCCTGATGAGACCAAATGAAAATCCAGCTCACAAGAGTCA
ATTGGTATGGATGGCATGCCACTCTGCTGCATTTGAAGATTTAAGAGTATCAAGTTTCATAAGAGGAAAG
AAGGTGATCCCAAGAGGAAAGCTTTCCACAAGAGGGGTTCAGATTGCTTCAAATGAGAATGTGGAAACCA
TGGACTCCAATACCCTGGAACTAAGAAGCAGATACTGGGCCATAAGAACCAGGAGTGGAGGAAATACCAA
TCAACAGAAGGCATCTGCAGGCCAGATCAGTGTGCAGCCTACATTCTCAGTGCAGCGAAATCTCCCCTTT
GAAAGAGCAACCATTATGGCAGCATTCAGCGGGAACAATGAAGGACGGACATCCGACATGCGAACAGAAG
TTATAAGAATGATGGAAAGTGCAAAGCCAGAGGATTTGTCCTTCCAGGGGCGGGGAGTCTTCGAGCTCTC
GGACGAAAAGGCAACGAACCCGATCGTGCCTTCCTTTGACATGAGTAATGAAGGGTCTTATTTCTTCGGA
GACAATGCAGAGGAGTATGACAATTGA
>NC_026434.1 Influenza A virus (A/California/07/2009(H1N1)) segment 6 neuraminidase (NA) gene, complete cds
ATGAATCCAAACCAAAAGATAATAACCATTGGTTCGGTCTGTATGACAATTGGAATGGCTAACTTAATAT
TACAAATTGGAAACATAATCTCAATATGGATTAGCCACTCAATTCAACTTGGGAATCAAAATCAGATTGA
AACATGCAATCAAAGCGTCATTACTTATGAAAACAACACTTGGGTAAATCAGACATATGTTAACATCAGC

AACACCAACTTTGCTGCTGGACAGTCAGTGGTTTCCGTGAAATTAGCGGGCAATTCCTCTCTCTGCCCTG
TTAGTGGATGGGCTATATACAGTAAAGACAACAGTGTAAGAATCGGTTCCAAGGGGGATGTGTTTGTCAT
AAGGGAACCATTCATATCATGCTCCCCCTTGGAATGCAGAACCTTCTTCTTGACTCAAGGGGCCTTGCTA
AATGACAAACATTCCAATGGAACCATTAAAGACAGGAGCCCATATCGAACCCTAATGAGCTGTCCTATTG
GTGAAGTTCCCTCTCCATACAACTCAAGATTTGAGTCAGTCGCTTGGTCAGCAAGTGCTTGTCATGATGG
CATCAATTGGCTAACAATTGGAATTTCTGGCCCAGACAATGGGGCAGTGGCTGTGTTAAAGTACAACGGC
ATAATAACAGACACTATCAAGAGTTGGAGAAACAATATATTGAGAACACAAGAGTCTGAATGTGCATGTG
TAAATGGTTCTTGCTTTACTGTAATGACCGATGGACCAAGTAATGGACAGGCCTCATACAAGATCTTCAG
AATAGAAAAGGGAAAGATAGTCAAATCAGTCGAAATGAATGCCCCTAATTATCACTATGAGGAATGCTCC
TGTTATCCTGATTCTAGTGAAATCACATGTGTGTGCAGGGATAACTGGCATGGCTCGAATCGACCGTGGG
TGTCTTTCAACCAGAATCTGGAATATCAGATAGGATACATATGCAGTGGGATTTTCGGAGACAATCCACG
CCCTAATGATAAGACAGGCAGTTGTGGTCCAGTATCGTCTAATGGAGCAAATGGAGTAAAAGGGTTTTCA
TTCAAATACGGCAATGGTGTTTGGATAGGGAGAACTAAAAGCATTAGTTCAAGAAACGGTTTTGAGATGA
TTTGGGATCCGAACGGATGGACTGGGACAGACAATAACTTCTCAATAAAGCAAGATATCGTAGGAATAAA
TGAGTGGTCAGGATATAGCGGGAGTTTTGTTCAGCATCCAGAACTAACAGGGCTGGATTGTATAAGACCT
TGCTTCTGGGTTGAACTAATCAGAGGGCGACCCAAAGAGAACACAATCTGGACTAGCGGGAGCAGCATAT
CCTTTTGTGGTGTAAACAGTGACACTGTGGGTTGGTCTTGGCCAGACGGTGCTGAGTTGCCATTTACCAT
TGACAAGTAA
>MF575134.1 Influenza A virus (A/black-headed gull/Netherlands/31/2014(H13N6)) segment 6 neuraminidase (NA) gene, complete cds
AGCAAAAGCAGGGTGACAATGAATCCAAATCAGAAGATAATATGCATCTCAGCTACAGGAATGACACTAT
CGGTAGTAAGCCTCCTGATAGGAATTGCCAACTTAGGTTTGAACATCGGACTCCATTACAAGGTTTGTGA
TGCACCAGATTCACCTAGCTCAAATGGGAATGGCACAAACACAACAACAACAATAATCAACAACAATACT
AACAATTTCACAAACATCACCAACATTGTCCAGAATAAAAATGAGGAGAAGACATTTCTAAATTTAACTA
AGCCTCTATGCGCTGTCAACTCATGGCACATCCTGTCAAAGGACAATGCAATAAGAATTGGAGAAAATGC
TCACATATTAGTCACAAGGGAGCCTTACCTATCCTGTGACCCACAAGGGTGCAGAATGTTTGCTCTAAGT
CAAGGCACAACACTCAGGGGACAGCATGCAAATGGAACTATACATGATAGAAGCCCGTTCAGAGCCCTCG
TAAGTTGGGAAATGGGTCAGGCACCCAGTCCATATAATGTCAAAGTCGAATGTATAGGTTGGTCGAGCAC
ATCATGCCATGACGGCAGGTCAAGAATGTCTATATGCATATCAGGACCAAACAATAATGCTTCGGCTGTA
GTATGGTATGGGGGTAGACCAATAACTGAGATTCCATCATGGGCAGGTAATATTCTCAGAACTCAAGAGT
CAGAGTGTGTGTGCCATAAAGGAATTTGCCCGGTAGTCATGACAGATGGCCCAGCAAATAATAAAGCAGC
AACTAAAATAATTTACTTTAAAGAAGGGAAAATTCAAAAAATTGAAGATCTGACAGGGAACGCCCAACAC
ATTGAAGAATGCTCATGCTATGGAGCAAAAGGAGTGATCAAATGCATCTGCAGAGACAATTGGAAGGGAG
CAAATAGGCCAGTAATCACTATAGATCCTGAAATGATGACCCACAGCAGCAAATATTTGTGTTCAAAAGT
CCTAACTGATACTAGTCGTCCCAATGATCCAACTAATGGGAATTGCGACGCACCAATAACAGGGGGAGGC
CCGGACCCTGGAGTCAAAGGATTTGCATTCCTAGATGGGGAAAATTCATGGCTGGGGAGGACAATTAGCA
AAGACTCCAGGTCAGGTTACGAAATGCTAAAAGTCCCAAATGCGGAAACCAGCACTCAATCTGGTCCAAT
CGCAAACCAAGTGATTGTCAACAACCAGAACTGGTCAGGATACTCAGGAGCATTCATAGACTACTGGGCA
AACAAAGAATGCTTTAATCCCTGTTTCTATGTAGAGCTTATTAGAGGGAAACCTAAAGAAAGCAGTGTAT
TGTGGACTTCAAATAGCATTGTAGCCCTTTGCGGATCCAAGGAGCGATTGGGATCATGGTCCTGGCATGA
TGGTGCAGAAATCATCTACTTTAAGTAGCAATGATTTAAGAAAAACACCCTTGTTTCTACT
>iavh1n11922|A/Mute_Swan/Netherlands/2/2022|EPI_ISL_15364797|A/Mute_Swan/Netherlands/2/2022|NA|6||A/Mute_Swan/Netherlands/2/2022
TTCAAAATGAATCCAAATCAAAGGATAATAACCACTGGATCAATCTGTATGGCAATTGGGATAGTCAGCT
TGATGCTGCAAATTGGAAACATAATCTCAATATGGGTTAGCCATTCAATCCAAACAGGGAATCAATACCA
GCCTGAACCATGCAATCAAAGCATCATTACCTATGAGAACAACACCTGGGTAAATCAGACGTATGTCAAC
ATCATCAATACCAATTTTCTTGCTGAGCAGGCTGTTACTTCGGTAACATTAGCGGGCAATTCATCTCTTT
GCCCTATTAGTGGGTGGGCTATATACAGTAAGGACAACGGTATAAGAATTGGGTCCAAGGGGGATGTGTT

TGTTATAAGAGAACCGTTCATCTCTTGCTCCCACTTGGAATGCAGAACCTTTTTNCTGACCCAGGGAGCT
CTGCTGAATGACAAACATTCTAATGGGACCGTTAAGGATAGAAGCCCCTATAGAACTTTGATGAGTTGTC
CCGTGGGTGAGGCTCCTTCCCCGTACAATTCAAGATTTGAGTCTGTTGCTTGGTCGGCAAGTGCTTGTCA
TGATGGCATCAGTTGGCTGACAATCGGTATTTCTGGTCCAGACAATGGAGCTGTGGCTGTATTGAAGTAC
AATGGCATAATAACGGATACTATCAAGAGTTGGAGGAACAACATTTTGAGAACTCAAGAATCTGAATGTG
CGTGCGTAAATGGCTCTTGTTTCACTGTAATGACTGATGGACCAAGCAATGGGCAGGCCTCATATAAAAT
CTTCAAGATAGAGAAAGGGAAAGTAGTCAAATCAGTTGAATTGAATGCCCCTAATTACCACTACGAGGAA
TGCTCCTGTTATCCTGATGCGGGTGAGATTATGTGTGTTTGCAGGGACAATTGGCATGGCTCAAACCGGC
CGTGGGTATCTTTTAACCAAAATCTGGAGTATCAAATAGGATATATATGCAGTGGGGTTTTCGGAGACAA
CCCCCGCCCCAATGACGGAACAGGCAGTTGCAGTCCAATGTCCTCTAACGGGGCATATGGGGTAAAAGGG
TTTTCATTTAAGTACGGTAATGGGGTTTGGATCGGAAGAACAAAAAGCACTAGTTCCAGGAGCGGCTTTG
AGATGATTTGGGATCCGAATGGGTGGACTGAGACGGACAGTAGTTTCTCAGTGAAGCAAGACATCGTAGC
AATAACTGACTGGTCAGGATATAGTGGGACTTTTGTCCAGCACCCAGAACTGACAGGATTAGATTGCATG
AGGCCTTGTTTCTGGGTTGAGCTGATTAGAGGGAGGCCCAAAGAGAACACAATTTGGACTAGCGGGAGCA
GTATATCCTTTTGTGGTGTAAATAGTGACACTGTGGGTTGGTCGTGGCCAGACGGTGCTGAATTGCCATT
CACCATTGACAAGTAGTTTGTTCAAAAAACT
>iavh1n11922|A/Barry/3792/2022|EPI_ISL_15391170|A/Barry/3792/2022|NA|6||A/Barry/3792/2022
AGCAAAAGCAGGAGTAAAGATGAATCCAAATCAAAAGATAATAACGATTGGCTCTGTTTCTCTCACAATT
TCCACAATATGCTTCTTCATGCAAATTGCCATCCTGATAACTACTGTAACATTGCATTTCAAGCAATATG
AATTCAACTCCCCCCCAAATAACCAAGTGATGCTGTGTGAACCAACAATAATAGAAAGAAACATAACAGA
GATAGTGTATTTGACCAACACCACCATAGAGAAGGAAATATGCCCCAAACCAGCAGAATACAGAAATTGG
TCAAAACCGCAATGTGGCATTACAGGATTTGCACCTTTCTCTAAGGACAATTCGATTAGGCTTTCCGCTG
GTGGGGACATATGGGTGACAAGAGAACCGTATGTGTCATGCGATCTTGACAAGTGTTATCAATTTGCCCT
TGGACAGGGAACAACACTAAACAATGTGCATTCAAATAACACAGTACATGATAGAACCCCTTATCGGACT
CTATTGATGAATGAGTTGGGTGTTCCTTTCCATCTGGGGACCAAGCAAGTGTGCATAGCATGGTCCAGCT
CAAGTTGTCACGATGGAAAAGCATGGCTGCATGTTTGTATAACGGGGGATGATAAAAATGCAACTGCTAG
CTTCATTTACAATGGGAGGCTTGTAGATAGTGTTGTTTCATGGTCCAACGATATTCTCAGAACCCAGGAG
TCAGAATGCGTTTGTATCAATGGAACTTGTACAGTAGTAATGACTGATGGAAATGCTACAGGAAAAGCTG
ATACTAAAATACTATTCATTGAGGAGGGGAAAATCGTTCATACTAGCAAATTGTCAGGAAGTGCTCAGCA
TGTCGAAGAGTGCTCTTGCTATCCTCGATATCCTGGTGTCAGATGTGTCTGCAGAGACAACTGGAAAGGA
TCCAACCGGCCCATCATAGATATAAACATAAAGGATCATAGCATTGTTTCCAGGTATGTGTGTTCTGGAC
TTGTTGGAGACACACCCAGAAAAAGCGACAGCTCCAGCAGTAGCCACTGTTTGAACCCTAACAATGAAAA
AGGTGATCATGGAGTGAAAGGCTGGGCCTTTGATGATGGAAATGACGTGTGGATGGGGAGAACAATCAAC
GAGACGTCACGCTTAGGGTATGAAACCTTCAAAGTCGTTGAAGGCTGGTCCAACCCTAAGTCCAAATTGC
AGATAAATAGGCAAGTCATAGTTAATAGAGGCGATAGGTCCGGTTATTCTGGTATTTTCTCTGTTGAAGG
CAAAAGCTGCATCAATCGGTGCTTTTATGTGGAGTTGATTAGGGGAAGAAAAGAGGAAACTGAAGTCTTG
TGGACTTCAAACAGTATTGTTGTGTTTTGTGGCACCTCAGGTACATATGGAACAGGCTCATGGCCTGATG
GGGCGAACCTCAGTCTCATGCATACATAAGCTTTCGCAATTTTAGAAAAAACC
>BEL12092022_ H13N6_NA_6
AGCAAAAGCAGGGTGACAATGAATCCAAATCAGAAGAGCATATGCATCTCAGCTACAGGAATGACACTAT
CGGTAGTAAGCCTTCTGATAGGAATAGCCAACTTAGGTTTGAACATCGGACTCCACTACAAGGTTTGTGA
TATACCAGATTCACCTAGCTCAAATGGGAATGGAACAAACACAACAACAACGATAATCAACAACAATACT
AACAATTTCACAAACATCACTAACATTGTCCAAAACAAAAATCAGGAGAAGACATTTCTAAATTTAACTA
AGCCCCTATGCGCTGTCAACTCATGGCACATCCTGTCAAAGGACAATGCAACAAGAATTGGAGAAAATGC
TCACATATTAGTCACAAGGGAGCCTTACCTATCCTGTGACCCACAAGGGTGTAGAATGTTTGCTCTAAGT
CAAGGCACAACACTCAGAGGGTAACATGCAAATGGGACTATACATGATAGAAGCCCGTTCAGAGCCCTCG

TAAGTTGGGAAATGGGGCAGGCACCCAGTCCATATAATGTCAAAGTAGAATGTATAGGTTGGTCGAGCAC
ATCATGCCATGACGGCAGGTCAAGAATGTCTATATGCATGTCAGGACCAAACAACAATGCTTCTGCTGTA
GTGTGGTATGGAGGTAGACTAATAACTGAGATTCCATCATGGGCAGGTAATATTCTCAGAACTCAAGAGT
CAGAGTGAGTGTGCCATAAAGGGATCTGCCCGGTAGTCATGACAGATGGCCCAGCAAATAATAAGGCAGC
AACTAAAATAATTTACTTTAAAGAAGGGAAAATTCAAAAAATTGAAGATTTGACAGGGAACGCCCAACAC
ATTGAAGAATGCTCATGCTATGGAGCAAAGGGATTAATCAAATGCATCTGCAGGGATAATTGGAAGGGAG
CAAATAGGCCAGTAATCACTATAGATCCAGAAATGATGACCCATAGCAGCAAACATTTGTGTTCAAAGGT
CCTAACTGATACCAGTCGTCCCAATGATCCAACCAATGGGAACTGCGACGCACCAATAACAGGGGGAGGC
CCGGACCCTGGAGTCAAAGGATTTGCGTTCCTAGATGGGGAAAATCCATGGCTAGGAAGGACAATCAGCA
AAGACTCTAGGTCAGGTTACGAAGTGTTAAAAGTTCCAAATGCGGAAACCAGCACTCAATCTGGTCCAAT
CGCACACCAAGTAATTGTCAAGAACCAGAACTGGTCAGGATACTCAGGGGCATTCATAGACTACTGGGCA
AACAAAGAATGCTTTAATCCCTGTTTCTATGTAGAATTAATTAGAGGGAAGCCTAAAGAAAGCAATGTGT
TGTAGACTTCAAATAGCATTGTAGCCCTCGGTGGATCCAAGGAGCGATTGGGATCATGGTCCTGGCATGA
TGGTGCAGAGATCATCTACTTTAAGTAGCAATGATTTAAGAAAAACACCCTTGTTTCTACT
>ENN12092022_ H13N6_NA_6
AGCAAAAGCAGGGTGACAATGAATCCAAATCAGAAGATAATATGCATCTCAGCTACAGGAATGACACTAT
CGGTAGTAAGCCTTTTGATAGGAATAGCCAACTTAGGTTTGAACATCGGACTCCACTACAAGGTTTGTGA
TATACCAGATTCACCTAGCTCAAATGGGAATGGCACAAACACAACAACAACGATAATCAACAACAATACT
AACAATTTCACAAACATCACTAACATTGTCCAAAATAAAAATGAAGAGAAGACATTTCTAAATTTAACTA
AGCCCCTATGCGCTGTCAACTCATGGCACATCCTGTCAAAGGACAATGCAATAAGAATTGGAGAAAATGC
TCACATATTAGTCACAAGGGAGCCTTACCTATCCTGTGACCCACAAGGGTGTAGAATGTTTGCTCTAAGT
CAAGGCACAACACTCAGAGGGCAACATGCAAATGGGACTATACATGATAGAAGCCCGTTCAGAGCCCTTG
TAAGTTGGGAAATGGGGCAGGCACCCAGTCCATATAATGTCAAAGTAGAATGTATAGGTTGGTCGAGCAC
ATCATGCCATGACGGCAGGTCAAGAATGTCTATATGCATGTCAGGACCAAACAACAATGCTTCGGCTGTA
GTGTGGTATGGAGGTAGACCAATAACTGAGATTCCATCATGGGCAGGGAATATTCTCAGAACTCAAGAGT
CAGAGTGTGTGTGCCATAAAGGGATCTGCCCGGTAGTCATGACAGATGGCCCAGCAAATAATAAGGCAGC
AACTAAAATAATTTACTTTAAAGAAGGGAAAATTCAAAAAATTGAAGATTTGACAGGGAACGCCCAACAC
ATTGAAGAATGCTCATGCTATGGAGCAAAAGGATTGATCAAATGCATCTGCAGGGATAATTGGAAGGGAG
CAAATAGGCCAGTAATCACTATAGATCCAGAAATGATGACCCATAGCAGCAAATATTTGTGTTCAAAGGT
CCTAACTGATACCAGTCGTCCCAATGATCCAACTAATGGGAACTGCGACGCACCGATAACAGGGGGAGGC
CCGGACCCTGGAGTCAAAGGATTTGCGTTCCTAGATGGGGAAAATTCATGGCTAGGAAGGACAATCAGCA
AAGACTCTAGGTCAGGTTACGAAGTGTTAAAAGTTCCAAATGCGGAAACCAGCACTCAATCTGGTCCACT
CGCACACCAAGTAATTGTGAACAACCAGAACTGGTCAGGATACTCAGGGGCATTCATAGACTACTGGGCA
AACAAAGAATGCTTTAATCCCTGTTTCTATGTAGAATTAATTAGAGGGAAGCCTAAAGAAAGCAATGTGT
TGTGGACTTCAAATAGCATTGTAGCCCTTTGCGGATCCAAGGAGCGATTGGGATCATGGTCCTGGCATGA
TGGTGCAGAGATCATCTACTTTAAGTAGCAATGATTTAAGAAAAACACCCTTGTTTCTACT
>CRG12092022_ H1N1_NA_6
ATGAATCCAAACCAAAAGATAATAACCATTGGTTCTGTTTGTATGACAATTGGAACGGCTAACTTAATAT
TACAAATTGGAAACATAATCTCAATATGGGTTAGCCACTCAATTCAAATTGGAAATCAAAGCCAGATTGA
AACATGCAATAAAAGCGTCATTACTTATGAAAACAACACTTGGGTAAATCAGACATTTGTTAACATCAGC
AACACTAACTCTGCTGCTAGACAGTCAGTGGCTTCCGTGAAATTAGCGGGCAATTCTTCTCTCTGCCCTG
TTAGTGGATGGGCTATATACAGTAAAGACAACAGTGTAAGAATCGGTTCCAAGGGGGATGTGTTTGTCAT
AAGGGAACCATTCATATCATGCTCCTCCTTGGAATGCAGAACCTTCTTCTTGACTCAAGGGGCTTTGCTA
AATGACAAACATTCCAATGGAACAGTCAAAGACAGAAGCCCATATCGAACCCTAATGAGCTGTCCTATTG
GTGAAGTTCCCTCTCCATACAACTCAAGATTTGAGTCAGTCGCTTGGTCAGCAAGTGCTTGTCATGATGG
CACCAATTGGCTAACAATTGGAATTTCTGGCCCAGACAGTGGGGCAGTGGCTGTGTTAAAATACAATGGC

ATAATAACAGACACTATCAAGAGTTGGAGGAACAAGATATTGAGAACACAAGAGTCTGAATGTGCATGTG
TAAATGGTTCTTGCTTTACCATAATGACTGATGGACCAAGTGATGGACAGGCCTCATACAAAATCTTCAG
AATAGAGAAGGGAAAGATAATCAAATCAGTCGAAATGAAGGCCCCTAATTATCACTATGAAGAATGCTCC
TGTTACCCTGATTCTAGTGAAATCACATGTGTGTGCAGGGATAATTGGCATGGCTCGAATCGACCTTGGG
TGTCTTTCAACCAGAATCTGGAATATCAGATGGGATACATATGCAGTGGGGTTTTCGGAGACAATCCACG
CCCTAATGATAAGACAGGCAGTTGTGGTCCAGTATTGTCTAATGGAGCAAATGGGGTAAAAGGATTTTCA
TTCAAATACGGCAATGGTGTTTGGATAGGGAGAACTAAGAGCATTAGTTCAAGAAAAGGTTTTGAGATGA
TTTGGGATCCGAATGGATGGACTGGGACTGACAATAAATTCTCAAAAAAGCAAGATATTGTAGGGATAAA
TGAGTGGTCAGGGTATAGCGGGAGTTTTGTTCAGCATCCAGAACTAACAGGGCTGAATTGTATAAGACCT
TGCTTCTGGGTTGAACTAATAAGAGGACGACCCGAAGAGAACACGATCTGGACTAGCGGGAGCAGCATAT
CCTTTTGTGGTGTAGACAGTGACATTATGGGTTGGTCTTGGCCAGACGGTGCTGAGTTGCCATTCACCAT
TGACAATTAA
>BEL10102022_ H1N1_NA_6
ATGAATCCAAACCAAAAGATAATAACCATTGGTTCTGTTTGTATGACAATTGGAACGGCTAACTTAATAT
TACAAATTGGAAACATAATCTCAATATGGGTTAGCCACTCAATTCAAATTGGAAATCAAAGCCAGATTGA
AACATGCAATAAAAGCGTCATTACTTATGAAAACAACACTTGGGTAAATCAGACATTTGTTAACATCAGC
AACACTAACTCTGCTGCTAGACAGTCAGTGGCTTCCGTGAAATTAGCGGGCAATTCCTCTCTCTGCCCTG
TTAGTGGATGGGCTATATACAGTAAAGACAACAGTGTAAGAATCGGTTCCAAGGGGGATGTGTTTGTCAT
AAGGGAACCATTCATATCATGCTCCCCCTTGGAATGCAGAACCTTCTTCTTGACTCAAGGGGCTTTGCTA
AATGACAAACATTCCAATGGAACAATCAAAGACAGAAGCCCATATCGAACCCTAATGAGCTGTCCTATTG
GTGAAGTTCCCTCTCCATACAACTCAAGATTTGAGTCAGTCGCTTGGTCAGCAAGTGCTTGTCATGATGG
CACCAATTGGCTAACAATTGGAATTTCTGGCCCAGACAGTGGGGCAGTGGCTGTGTTAAAATACAATGGC
ATAATAACAGACACTATCAAGAGTTGGAGGAACAAGATATTGAGAACACAAGAGTCTGAATGTGCATGTG
TAAATGGTTCTTGCTTTACCATAATGACTGATGGACCAAGTGATGGACAGGCCTCATACAAAATCTTCAG
AATAGAGAAGGGAAAGATAATCAAATCAGTCGAAATGAAGGCCCCTAATTATCACTATGAAGAATGCTCC
TGTTACCCTGATTCTAGTGAAATCACATGTGTGTGCAGGGATAATTGGCATGGCTCGAATCGACCTTGGG
TGTCTTTCAACCAGAATCTGGAATATCAGATGGGATACATATGCAGTGGGGTTTTCGGAGACAATCCACG
CCCTAATGATAAGACAGGCAGTTGTGGTCCAGTATTGTCTAATGGAGCAAATGGGGTAAAAGGATTTTCA
TTCAAATACGGCAATGGTGTTTGGATAGGGAGAACTAAGAGCATTAGTTCAAGAAAAGGTTTTGAGATGA
TTTGGGATCCGAATGGATGGACTGGGACTGACAATAAATTCTCAAAAAAGCAAGATATTGTAGGGATAAA
TGAGTGGTCAGGGTATAGCGGGAGTTTTGTTCAGCATCCAGAACTAACAGGGCTGAATTGTATAAGACCT
TGCTTCTGGGTTGAACTAATAAGAGGACGACCCGAAGAGAACACGATCTGGACTAGCGGGAGCAGCATAT
CCTTTTGTGGTGTAGACAGTGACATTATGGGTTGGTCTTGGCCAGACGGTGCTGAGTTGCCATTCACCAT
TGACAATTAA
>BEL31102022_ H1N1_NA_6
ATGAATCCAAACCAAAAGATAATAACCATTGGTTCTGTTTGTATGACAATTGGAACGGCTAACTTAATAT
TACAAATTGGAAACATAATCTCAATATGGGTTAGCCACTCAATTCAAATTGGAAATCAAAGCCAGATTGA
AACATGCAATAAAAGCGTCATTACTTATGAAAACAACACTTGGGTAAATCAGACATTTGTTAACATCAGC
AACACTAACTCTGCTGCTAGACAGTCAGTGGCTTCCGTGAAATTAGCGGGCAATTCTTCTCTCTGCCCTG
TTAGTGGATGGGCTATATACAGTAAAGACAACAGTGTAAGAATCGGTTCCAAGGGGGATGTGTTTGTCAT
AAGGGAACCATTCATATCATGCTCCCCCTTGGAATGCAGAACCTTCTTCTTGACTCAAGGGGCTTTGCTA
AATGACAAACATTCCAATGGAACAATTAAAGACAGAAGCCCATATCGAACCCTAATGAGCTGTCCTATTG
GTGAAGTTCCCTCTCCATACAACTCAAGATTTGAGTCAGTCGCTTGGTCAGCAAGTGCTTGTCATGATGG
CACCAATTGGCTAACAATTGGAATTTCTGGCCCAGACAGTGGGGCAGTGGCTGTGTTAAAATACAATGGC
ATAATAACAGACACTATCAAGAGTTGGAGGAACAAGATATTGAGAACACAAGAGTCTGAATGTGCATGTG
TAAATGGTTCTTGCTTTACCATAATGACCGATGGACCAAGTGATGGACAGGCCTCATACAAAATCTTCAG

AATAGAGAAGGGAAAGATAATCAAATCAGTCGAAATGAAGGCCCCTAATTATCACTATGAAGAATGCTCC
TGTTACCCTGATTCTAGTGAAATCACATGTGTGTGCAGGGATAATTGGCATGGCTCGAATCGACCTTGGG
TGTCTTTCAATCAGAATCTGGAATATCAGATGGGATACATATGCAGTGGGGTTTTCGGAGACAATCCACG
CCCTAATGATAAGACAGGCAGTTGTGGTCCAGTATTGTCTAATGGAGCAAATGGGGTAAAAGGATTTTCA
TTCAAATACGGCAATGGTGTTTGGATAGGGAGAACTAAAAGCATTAGTTCAAGAAAAGGTTTTGAGATGA
TTTGGGATCCGAATGGATGGACTGGGACTGACAATAAATTCTCAAAAAAGCAAGATATTGTAGGGATAAA
TGAGTGGTCAGGGTATAGCGGGAGTTTTGTTCAGCATCCAGAACTAACAGGGCTGAATTGTATAAGACCT
TGCTTCTGGGTTGAACTAATAAGAGGACGACCCGAAGAGAACACGATCTGGACTAGCGGGAGCAGCATAT
CCTTTTGTGGTGTAGACAGTGACATTATGGGTTGGTCTTGGCCAGACGGTGCTGAGTTGCCATTCACCAT
TGACAATTAA
>BEL24102022_ H1N1_NA_6
ATGAATCCAAACCAAAAGATAATAACCATTGGTTCTATTTGTATGACAATTGGAACGGCTAACTTAATAT
TACAAATTGGAAACATAATCTCAATATGGGTTAGCCACTCAATTCAAATTGGAAATCAAAGCCAGATTGA
AACATGCGATAAAAGCGTCATTACTTATGAAAACAACACTTGGGTAAATCAGACATTTGTTAACATCAGC
AACACTAACTCTGCTGCTAGACAGTCAGTGGCTTCCGTGAAATTAGCGGGCAATTCCTCTCTCTGCCCTG
TTAGTGGATGGGCTATATACAGTAAAGACAACAGTGTAAGAATCGGTTCCAAGGGGGATGTGTTTGTCAT
AAGGGAACCATTCATATCATGCTCTCCCTTGGAATGCAGAACCTTCTTCTTGACTCAAGGGGCTTTGCTA
AATGACAAACATTCCAATGGAACAATTAAAGACAGAAGCCCATATCGAACCCTAATGAGCTGTCCTATTG
GTGAAGTTCCCTCTCCATACAACTCAAGATTTGAGTCAGTCGCTTGGTCAGCAAGTGCTTGTCATGATGG
CACCAATTGGCTAACAATTGGAATTTCTGGCCCAGACAGTGGGGCAGTGGCTGTGTTAAAATACAATGGC
ATAATAACAGACACTATCAAGAGTTGGAGGAACAAGATATTGAGAACACAAGAGTCTGAATGTGCATGTG
TAAATGGTTCTTGCTTTACCATAATGACCGATGGACCAAGTGATGGACAGGCCTCATACAAAATCTTCAG
AATAGAGAAGGGAAAGATAATCAAATCAGTCGAAATGAAGGCCCCTAATTATCACTATGAAGAATGCTCC
TGTTACCCTGATTCTAGTGAAATCACATGTGTGTGCAGGGATAATTGGCATGGCTCGAATCGACCTTGGG
TGTCTTTCAACCAGAATCTGGAATATCAGATGGGATACATATGCAGTGGGGTTTTCGGAGACAATCCACG
CCCTAATGATAAGACAGGCAGTTGTGGTCCAGTATCGTCTAATGGAGCAAATGGGGTAAAAGGATTTTCA
TTCAAATACGGCAATGGTGTTTGGATAGGGAGAACTAAGAGCATTAGTTCAAGAAAAGGTTTTGAGATGA
TTTGGGATCCGAATGGATGGACTGGGACTGACAATAAATTCTCAAAAAAGCAAGATATTGTAGGGATAAA
TGAGTGGTCAGGGTATAGCGGGAGTTTTGTTCAGCATCCAGAACTAACAGGGCTGAATTGTATAAGACCT
TGCTTCTGGGTTGAACTAATAAGAGGACGACCCGGAGAGAACACGATCTGGACTAGCGGGAGCAGCATAT
CCTTTTGTGGTGTAGACAGTGACATTATGGGTTGGTCTTGGCCAGACGGTGCTGAGTTGCCATTCAACAT
TGACAATTAA
>NC_026431.1 Influenza A virus (A/California/07/2009(H1N1)) segment 7 matrix protein 2 (M2) and matrix protein 1 (M1) genes, complete cds
ATGAGTCTTCTAACCGAGGTCGAAACGTACGTTCTTTCTATCATCCCGTCAGGCCCCCTCAAAGCCGAGA
TCGCGCAGAGACTGGAAAGTGTCTTTGCAGGAAAGAACACAGATCTTGAGGCTCTCATGGAATGGCTAAA
GACAAGACCAATCTTGTCACCTCTGACTAAGGGAATTTTAGGATTTGTGTTCACGCTCACCGTGCCCAGT
GAGCGAGGACTGCAGCGTAGACGCTTTGTCCAAAATGCCCTAAATGGGAATGGGGACCCGAACAACATGG
ATAGAGCAGTTAAACTATACAAGAAGCTCAAAAGAGAAATAACGTTCCATGGGGCCAAGGAGGTGTCACT
AAGCTATTCAACTGGTGCACTTGCCAGTTGCATGGGCCTCATATACAACAGGATGGGAACAGTGACCACA
GAAGCTGCTTTTGGTCTAGTGTGTGCCACTTGTGAACAGATTGCTGATTCACAGCATCGGTCTCACAGAC
AGATGGCTACTACCACCAATCCACTAATCAGGCATGAAAACAGAATGGTGCTGGCTAGCACTACGGCAAA
GGCTATGGAACAGATGGCTGGATCGAGTGAACAGGCAGCGGAGGCCATGGAGGTTGCTAATCAGACTAGG
CAGATGGTACATGCAATGAGAACTATTGGGACTCATCCTAGCTCCAGTGCTGGTCTGAAAGATGACCTTC
TTGAAAATTTGCAGGCCTACCAGAAGCGAATGGGAGTGCAGATGCAGCGATTCAAGTGATCCTCTCGTCA
TTGCAGCAAATATCATTGGGATCTTGCACCTGATATTGTGGATTACTGATCGTCTTTTTTTCAAATGTAT
TTATCGTCGCTTTAAATACGGTTTGAAAAGAGGGCCTTCTACGGAAGGAGTGCCTGAGTCCATGAGGGAA

GAATATCAACAGGAACAGCAGAGTGCTGTGGATGTTGACGATGGTCATTTTGTCAACATAGAGCTAGAGT
AA
>MF575114.1 Influenza A virus (A/black-headed gull/Netherlands/31/2014(H13N6)) segment 7 matrix protein 2 (M2) and matrix protein 1 (M1) genes, complete cds
AGCAAAAGCAGGTAGATATTGAAAGATGAGTCTTCTAACCGAGGTCGAAACGTATGTTCTCTCTATCATC
CCGTCAGGCCCCCTCAAAGCCGAGATAGCACAGAAACTTGAAGATGTTTTTGCAGGGAAGAACACAGATC
TGGAGGCTCTTATGGAGTGGTTAAAGACAAGACCAATCTTGTCACCTCTGACTAAGGGGATTTTAGGATT
TGTGTTTACGCTCACCGTGCCCAGTGAGCGAGGACTGCAGCGTAGACGCTTTGTCCAAAACGCCCTAAAT
GGGAATGGAGACCCGAACAACATGGATAGAGCAGTCAAGCTTTATCGAAAGCTGAAAAGAGAGATAACAT
TCCATGGAGCTAAAGAGGTGGCACTTAGCTACTCAACTGGTGCGCTTGCCAGCTGCATGGGTCTCATTTA
CAACAGAATGGGTACAGTGACCACAGAAGTGGCTTTTGGTCTTGTGTGTGCCACTTGCGAGCAGATTGCT
GACTCCCAGCACAGGTCTCACAGACAAATGGTAACCACAACCAATCCACTGATCAGGCATGAGAACAGAA
TGGTGCTGGCAAGCACTACTGCCAAGGCTATGGAGCAGATGGCTGGATCAAGTGAACAAGCAGCTGAGGC
CATGGAGGTTGCTGGTCAGGCCAGACAGATGGTGCAGGCAATGAGAACAATTGGGACTCATCCTAGCTCC
AGTGCTGGTCTAAAAGATGATCTTCTTGAAAACTTGCAGGCCTATCAGAAGCGGATGGGAGTGCAAATGC
AGCGATTCAAGTGACCCTCTCGTCATTGCGGCGAGTGTCATTGGGATCTTGCACTTGATACTGTGGATTC
TTGATCGCCTTTTCTTCAAATGCATTTATCGTCGCATTAAATACGAGTTGAAAAGAGGGCCTTCTACGGA
AGGAGTGCCTGAGTCTATGAGGGAAGAGTATCGGCAGGAGCAGCAGAGTGCTGTGGATGTTGACGATGGT
CATTTTGTCAACATAGAGCTGGAGTAAAAAACTACCTTGTTTCTACT
>iavh1n11922|A/Mute_Swan/Netherlands/2/2022|EPI_ISL_15364797|A/Mute_Swan/Netherlands/2/2022|MP|7||A/Mute_Swan/Netherlands/2/2022
TATTGAAAGATGAGTCTTCTAACCGAGGTCGAAACGTACGTTCTCTCTATCGTCCCGTCGGGCCCCCTCA
AAGCCGAGATCGCGCAGAGACTTGAAGATGTCTTTGCAGGGAAGAACACCGATCTTGAGGCTCTCATGGA
ATGGCTAAAGACAAGACCAATCCTGTCACCTATGACTAAGGGGATTTTGGGATTTGTGTTCACGCTCACC
GTGCCCAGTGAGCGAGGACTGCAGCGTAGACGCTTTGTCCAAAATGCTCTAAATGGAAATGGAGACCCAA
ACAACATGGACAGGGCAGTCAAACTGTACAAGAAATTGAAGAGAGAGATAACATTCCATGGGGCTAAAGA
AGTTGCACTCAGTTACTCAACCGGTGCACTTGCCAGTTGTATGGGTCTCATATACAACAGGATGGGGACG
GTGACCGCAGAAGTGGCATTGGGCCTAGTGTGTGCCACCTGTGAGCAGATTGCTGATTCACAGCATCGGT
CTCACAGGCAGATAGCCACCACCACCAACCCACTAATCAGACATGAAAACAGAATGGTGTTGGCCAGTAC
CACAGCTAAGGCTATGGAGCAGATGGCTGGATCGAGTGAGCAAGCAGCGGAAGCCATGGAGGTTGCTAGT
CAGGCTAGGCAAATGGTGCAGGCGATGAGGACCATTGGAACTCATCCTAGCTCCAGTGCCGGTCTGAGAG
ATGATCTCCTTGAAAATTTGCAGGCCTACCAAAAACGGATGGGAGTGCAACTGCAGCGATTCAAGTGATC
CTCTCGTTATTGCCGCAAGTATCATTGGGATCTTGCACTTGATATTGTGGATTCTTGATCGCCTTTTCTT
CAAATGCGTTTATCGTCGCCTTAAATACGGTTTGAAAAGAGGGCCTTCTACGGAAGGAGTACCTGAGTCC
ATGAGGGAAGAGTACCGGCAGGAACAGCAGAGTGCTGTGGATGTTGACGATGGTCATTTTGTCAACATAG
AGCTGGAGTAAAAAACTA
>iavh1n11922|A/Barry/3792/2022|EPI_ISL_15391170|A/Barry/3792/2022|MP|7||A/Barry/3792/2022
AGCAAAAGCAGGTAGATATTGAAAGATGAGCCTTCTTACCGAGGTCGAAACGTATGTTCTCTCTATCGTT
CCATCAGGCCCCCTCAAAGCCGAGATCGCGCAGAGACTTGAAGATGTCTTTGCTGGGAAAAACACAGATC
TTGAGGCTCTCATGGAATGGTTAAAGACAAGACCAATTCTGTCACCTTTGACTAAGGGGATTTTAGGGTT
TGTTTTCACGCTCACCGTGCCCAGTGAGCGAGGACTGCAGCGTAGACGCTTTGTCCAAAATGCCCTCAAT
GGGAATGGAGACCCAAATAACATGGACAAAGCAGTTAAACTGTATAGGAAACTTAAGAGGGAGATAACGT
TCCACGGGGCCAAAGAAATAGCTCTCAGTTATTCTGCTGGTGCACTTGCCAGTTGCATGGGCCTCATATA
CAATAGGATGGGGGCTGTAACCACTGAAGTGGCATTTGGCCTGGTGTGTGCAACATGTGAGCAGATTGCT
GATTCCCAGCACAGGTCTCATAGGCAGATGGTGGCAACAACCAATCCATTAATAAAACATGAGAACAGAA
TGGTTTTGGCCAGCACTACAGCTAAGGCTATGGAGCAAATGGCTGGCTCAAGTGAGCAAGCAGCAGAGGC
CATGGAGATTGCTAGTCAGGCCAGGCAGATGGTGCAGGCAATGAGAGCCATTGGGACTCATCCTAGTTCC
AGCACTGGTCTAAGAGATGATCTTCTTGAAAATTTGCAGACCTATCAGAAACGAATGGGGGTGCAGATGC

AACGATTCAAGTAACCCACTTGTTGTTGCCGCGAATATCATTGGGATCTTGCACTTGATATTATGGATTC
TTGATCGTCTTTTTTTCAAATGCGTCTATCAACTCTTCAAACACGGCCTTAAAAGAGGCCCTTCTACGGA
AGGTGTGCCTGAGTCTATGAGGGAAGAATACCGAAAGGAACAGCAGAATGCTGTGGATGCTGACGAAAGT
CATTTTGTCAGCATAGAATTGGAGTAAAAAACTACCTTGTTTCTACT
>BEL10102022_ H1N1_M_7
ATGAGTCTTCTAACCGAGGTCGAAACGTACGTTCTTTCTATCATCCCGTCAGGCCCCCTCAAAGCCGAGA
TCGCACAGAGACTGGAAAGTGTCTTTGCAGGAAAGAACACAGATCTTGAGGCTATCATGGAATGGCTAAA
AACAAGACCAATCTTGTCACCTCTGACTAAGGGAATTTTAGGATTTGTGTTCACGCTCACCGTGCCCAGT
GAGCGAGGACTGCAGCGTAGACGCTTTATCCAAAATGCCCTAAATGGAAATGGGGACCCGAACAACATGG
ATAGAGCAGTTAGACTATACAAGAAACTCAAAAGAGAAATAACGTTCCATGGGGCCAAAGAAGTGTCACT
AAGCTATTCAACTGGTGCACTTGCAAGTTGCATGGGCCTCATATACAACAGGATGGGAACAGTGACCACA
GAAGCTGCTTTTGGTCTAGTTTGTGGCACTTGTGAACAGATTGCTGATTCACAGCATCGGTCTCACAGAC
AAATGGCTACTACCACAAATCCACTAATCAGGCATGAAAACAGAATGGTGCTGGCTAGCACTACGGCAAA
GGCTATGGAACAGGTGGCTGGATCGAGTGAACAGGCAGCGGAGGCCATGGAGGTTGCTAATAAGACTAGG
CAGATGGTACAAGCAATGAGAACTATTGGAACTCATCCTAGCTCCAGTGCTGGTCTAAGAGATGACCTTC
TTGAAAATTTACAGGCCTACCAGAAGCGAATGGGAGTGCAGATGCAGCGGTTCAAATGATCCTCTCGTCA
TTGCAGCAAACATCATTGGGATCTTGCACCTGATATTGTGGATTACTGATTGTCTTTTTTTCAAATGCAT
TTATCGTCGCTTTAAATACGGTTTGAAAAGAGGGCCTTCTACGGAAGGAGTGCCTGAGTCCATGAGGGAA
GAATATCAACAGGAGCAGCAGAGTGCTGTGGATGTTGACGATGGTCATTTTGTCAACATAGAGCTAGAGT
AA
>BEL31102022_ H1N1_M_7
ATGAGTCTTCTAACCGAGGTCGAAACGTACGTTCTTTCTATCATCCCGTCAGGCCCCCTCAAAGCCGAGA
TCGCACAGAGACTGGAAAGTGTCTTTGCAGGAAAGAACACAGATCTTGAGGCTATCATGGAATGGCTAAA
GACAAGACCAATCTTGTCACCTCTGACTAAGGGAATTTTAGGATTTGTGTTCACGCTCACCGTGCCCAGT
GAGCGAGGACTGCAGCGTAGACGCTTTATCCAAAATGCCCTAAATGGAAATGGGGACCCGAACAACATGG
ATAGAGCAGTTAGACTATACAAGAAACTCAAAAGAGAAATAACGTTCCATGGGGCCAAAGAAGTGTCACT
AAGCTATTCAACTGGTGCACTTGCAAGTTGCATGGGCCTCATATACAACAGGATGGGAACAGTGACCACA
GAAGCTGCTTTTGGTCTAGTTTGTGCCACTTGTGAACAGATTGCTGATTCACAGCATCGGTCTCACAGAC
AAATGGCTACTACCACAAATCCACTAATCAGGCATGAAAACAGAATGGTGCTGGCTAGCACTACGGCAAA
GGCTATGGAACAGGTGGCTGGATCGAGTGAACAGGCAGCGGAGGCCATGGAGGTTGCTAATAAGACTAGG
CAGATGGTACATGCAATGAGAACTATTGGAACTCATCCTAGCTCCAGTGCTGGTCTAAAAGATGACCTTC
TTGAAAATTTACAGGCCTACCAGAAGCGAATGGGAGTGCAGATGCAGCGGTTCAAATGATCCTCTCGTCA
TTGCAGCAAACATCATTGGGATCTTGCACCTGATATTGTGGATTACTGATCGTCTTTTTTTCAAATGCAT
TTATCGTCGCTTTAAATACGGTTTGAAAAGAGGGCCTTCTACGGAAGGAGTGCCTGAGTCCATGAGGGAA
GAATATCAACAGGAGCAGCAGAGTGCTGTGGATGTTGACGATGGTCATTTTGTCAACATAGAGCTAGAGT
AA
>NDN10092022_ H13N6_M_7
AGCAAAAGCAGGTAGATATTGAAAGATGAGCCTTCTAACCGAGGTCGAAACGTATGTTCTCTCTATCATC
CCATCAGGCCCCCTCAAAGCCGAGATAGCACAGAGACTTGAAGATGTTTTTGCAGGGAAGAACACAGACC
TTGAAGCTCTCATGGAGTGGCTAAAGACAAGACCGATCTTGTCACCTCTGACTAAGGGGATTTTGGGATT
TATTTTTACGCTCACCGTGCCCAGTGAGCGAGGACTGCAGCGTAGACGCTTTGTCCAAAACGCCCTAAAT
GGGAATGGAGACCCGAACAACATGGACAGGGCAGTGAAGCTTTACAGAAAGCTGAAGAGGGAGATAACAT
TCCATGGAGCTAAGGAGGTGGCACTCAGTTACTCGACTGGTGCACTTGCCAGCTGCATGGGTCTCATCTA
CAATAGGATGGGGACAGTAACCACAGAAGTGGCTTTTGGTCTAGTATGTGCCACTTGTGAGCAGATTGCT
GACTCTCAGCACCGATCCCACAGACAGATGGTGACTACAACCAACCCACTAATCAGACATGAAAATAGAA
TGGTGCTTGCAAGCACTACAGCCAAAGCTATGGAGCAGATGGCTGGATCGAGTGAACAAGCAGCAGAGGC

AATGGAGGTTGCAGGTCAGGCTAGACAGATGGTGCAGGCAATGAGAACAATTGGGACCCATCCTAGCTCC
AGTGCTGGTCTGAAAGATGATCTTCTTGAAAATTTACAGGCCTATCAGAAACGAATGGGAGTGCAAATGC
AGCGATTCAAGTGATCCTCTCGTTATTGCAGCAAGTATCATTGGGATCTTGCACTTGATATTGTGGATTC
TTGATCGTATTTTCTTCAAATGCATTTATCGTCGCATTAAATACGAGTTGAAAAGAGGGCCTTCTACGGA
AGGAGTGCCTGAGTCTATGAGGGAAGAATATCGACAGGAACAGCAGAATGCTGTGGATGTTGACGATAGT
CATTTTGTCAACATAGAGCTGGAGTAAAAAACTACCTTGTTTCTACT
>NC_026432.1 Influenza A virus (A/California/07/2009(H1N1)) segment 8 nuclear export protein (NEP) and nonstructural protein 1 (NS1) genes, complete cds
ATGGACTCCAACACCATGTCAAGCTTTCAGGTAGACTGTTTCCTTTGGCATATCCGCAAGCGATTTGCAG
ACAATGGATTGGGTGATGCCCCATTCCTTGATCGGCTCCGCCGAGATCAAAAGTCCTTAAAAGGAAGAGG
CAACACCCTTGGCCTCGATATCGAAACAGCCACTCTTGTTGGGAAACAAATCGTGGAATGGATCTTGAAA
GAGGAATCCAGCGAGACACTTAGAATGACAATTGCATCTGTACCTACTTCGCGCTACCTTTCTGACATGA
CCCTCGAGGAAATGTCACGAGACTGGTTCATGCTCATGCCTAGGCAAAAGATAATAGGCCCTCTTTGCGT
GCGATTGGACCAGGCGATCATGGAAAAGAACATAGTACTGAAAGCGAACTTCAGTGTAATCTTTAACCGA
TTAGAGACCTTGATACTACTAAGGGCTTTCACTGAGGAGGGAGCAATAGTTGGAGAAATTTCACCATTAC
CTTCTCTTCCAGGACATACTTATGAGGATGTCAAAAATGCAGTTGGGGTCCTCATCGGAGGACTTGAATG
GAATGGTAACACGGTTCGAGTCTCTGAAAATATACAGAGATTCGCTTGGAGAAACTGTGATGAGAATGGG
AGACCTTCACTACCTCCAGAGCAGAAATGAAAAGTGGCGAGAGCAATTGGGACAGAAATTTGAGGAAATA
AGGTGGTTAATTGAAGAAATGCGGCACAGATTGAAAGCGACAGAGAATAGTTTCGAACAAATAACATTTA
TGCAAGCCTTACAACTACTGCTTGAAGTAGAACAAGAGATAAGAGCTTTCTCGTTTCAGCTTATTTAATG
ATAAAAAACACCCTTGTTTCTAC
>MF575318.1 Influenza A virus (A/black-headed gull/Netherlands/31/2014(H13N6)) segment 8 nuclear export protein (NEP) and nonstructural protein 1 (NS1) genes, complete cds
AGCAAAAGCAGGGTGACAAAAACATAATGGATTCAAACACTGTATCAAGCTTTCAGGTAGATTGCTTTCT
TTGGCATGTCCGCAAACGATTTGCAGACCAAGAGATGGGTGATGCCCCATTCCTTGACCGGCTTCGCCGA
GATCAGAAGTCTTTAAAGGGGAGGAGCAGCACTCTTGGAATGGATATTGAGGCGGCTACATGCGCTGGGA
AGCGAATAATTGAGAGGATTCTAAAAGAAGAATCCAATGAGGCATTAAAGATGAATATTGCCTCAACGCC
TGCCTCACGCTACATGACTGACCTGACTCCAGAGGAGATGTCAAAAGAATGGTTCATGCTCATGCCAAAA
CAGAAATTTGCTGGGCCTCTCCGCATCAGAATGGATCAGGCGATCATAGATAAAGAAATCACACTGAAAG
CGAATTTCAGTGTAATTTTTGACCGGCTGGAGACTCTCGTACTGTTAAGGGCTTTTACCAATGATGGGGC
TATTGTTGGTGAAATCTCACCATTACCTTCTCTTCCAGGACATACTAACGAGGATGTCAAAAATGCAATT
GGAGTCCTCATCGGTGGACTTGAGTGGAATGATAACACAGTTCGAATCTCTGAAAATTTACAAAGATTCG
CTTGGGGAAGCAGTGATGAGGATGGGCGACCTCCACTCACTTCAAAGCAGGAACAAAAACTGGCGAGAAC
AATTGGGTCAGAAGTTTGAAGAAATAAGGTGGTTGATTGAGGAGGTGAGACACAAATTAAAGACAACAGA
AAACAGTTTTGAACAAATAACATTCATGCAAGCCTTACAACTGTTGCTTGAAGTGGAGCAAGAGATAAGG
ACTTTCTCGTTTCAGCTTATTTAATGATAAAAAACACCCTTGTTTCTACT
>iavh1n11922|A/Mute_Swan/Netherlands/2/2022|EPI_ISL_15364797|A/Mute_Swan/Netherlands/2/2022|NS|8||A/Mute_Swan/Netherlands/2/2022
GACAAAAACATAATGGATTCCAACACTGTGTCAAGCTTTCAGGTAGATTGCTTTCTTTGGCATGTCCGCA
AACGATTTGCAGACCAAGAACTGGGTGATGCCCCATTCCTTGACCGGCTTCGCCGAGATCAGAAATCCCT
GAGAGGAAGAGGCAGCACTCTTGGTCTGGACATCGAAACAGCCACCCGTGCGGGAAAGCAGATAGTGGAG
CTGATTCTGAAAGAAGAATCTGATGAGGCGCTTAAAATGACTATTGCCCCCGTGCCAGCTTCACGCTACC
TAACTGACATGACTCTTGAGGAGATGTCAAGAGACTGGTTCATGCTCATGCCCAAACAGAAAGTGGCAGG
TTCCCTTTGCATCAGAATGGACCAGGCAATAATGGATAAAACCATCATATTGAAAGCAAACTTCAGTGTG
ATTTTTGACCGGCTGGAAACCCTAATACTAATTAGAGCTTTCACAGAAGAAGGAGCAATTGTGGGAGAAA
TCTCACCATTACCTTCTCTTCCAGGACATACTAATGAGGATGTCAAAAATGCAATTGGGGTCCTCATCGG
AGGACTTGAATGGAATAATAACACAGTTCGAGTCTCTGAAACTCTACAGAGATTCGCTTGGAGAAGCAGT
AATGAGAATGGGAGATCTCCACTCCCTCCAAAGCAGAAACGGAAAATGGCGAGGACAATTGAGTCAGAAG
TTTGAAGAAATAAGATGGCTGATTGAAGAAGTGCGGCACAGATTGAAGATTACAGAGAACAGTTTCGAAC

AGATAACTTTCATGCAAGCCTTACAACTATTGCTTGAAGTGGAGCAAGAGATAAGAACTTTCTCGTTTCA
GCTTATTTAATGATAAAAAACAC
>iavh1n11922|A/Barry/3792/2022|EPI_ISL_15391170|A/Barry/3792/2022|NS|8||A/Barry/3792/2022
GATCAGCAAAAGCAGGGTGACAAAGACATAATGGATTCCAACACTGTGTCAAGTTTCCAGGTAGATTGCT
TTCTTTGGCATATCCGGAAACAAGTTGTGGACCAAAAACTGAGTGATGCCCCATTCCTCGATCGGCTTCG
CCGAGATCAGAGGTCCCTAAGGGGAAGAGGCAATACTCTCGGTCTAGACATCAAATCAGCCACCCATGTT
GGAAAGCAAATCGTAGAAAAGATTCTGAAAGGAGAATCTGATGAGGCACTTAAAATGACCATGGTCTCAA
CACCTGCTTCGCGATACATAACTGACATGACTATTGAGGAATTGTCAAGAAACTGGTTCATGCTAATGCC
CAAGCAGAAGGTGGAAGGACCTCTTTGCATCAGAATGGACCAGGCAATCACGGAGAAAAACATCATGTTA
AAAGCGAATTTCAATGTGATTTTTGGCCGGCTAGAGACCATAGTATTGCTAAGAGCTTTCACCGAAGAGG
GAGCAATTGTTGGCGAAATCTCACCATTGCCTTCTTTTCCAGGACATACTATTGAGGATGTCAAAAATGC
AATTGGGGTCCTCATCGGAGGACTTGAATGGAATGATAACACAGTTCGAGTCTCTAAAAATCTACAGAGA
TTCGCTTGGAGAAGCAGTCATGAGAATGGGGGACCTCCACTTACTCCAAAACAGAAACGGGAAATGGCGA
GAACAGCTAGGTCAGAAGTTTGAAGAGATAAGATGGCTGATTGAAGAGGTGAGACACAGATTAAGAACAA
CTGAAAATAGCTTTGAACAAATAACATTCATGCAAGCATTACAACTACTGTTTGAAGTGGAACAGGAGAT
AAGAACTTTCTCATTTCAGCTTATTTAATGATAAAAAACACCCTTGTTTCTACTGATC
>BEL12092022_ H13N6_NS_8
AGCAAAAGCAGGGTGACAAAAACATAATGGATTCAAACACTGTATCAAGCTTTCAGGTAGATTGCTTTCT
TTGGCATGTCCGCAAACGATTTGCAGACCAAGAGATGGGTGATGCCCCATTCCTTGACCGGCTTCGCCGA
GATCAGAAGTCTTTAAAGGGGAGGAGCAGCACTCTTGGAATGGACATTGAGGCGGCTACATGCGCTGGGA
AGCGGATAATTGAGCGGATTCTAAAAGAAGAATCCAATGAGGCATTAAAGATGAATATTGCCTCAACGCC
TGCCTCACGCTACATGACTGACATGACTCCAGAGGAGATGTCAAAAGAATGGTTCATGCTCATGCCAAAA
CAGAAATTTGCTGGGCCTCTCCGCATCAGAATGGATCAGGCAATCATAGATAAAGAAATCACACTGAAAG
CGAATTTCAGTGTAATTTTTGACCGGTTGGAGACTCTCGTGCTGTTAAGGGCTTTTACCAATGATGGGGC
TATTGTTGGTGAAATCTCACCATTGCCTTCTCTTCCAGGACATACTAACGAGGATGTCAAAAATGCAATT
GGAGTCCTCATCGGTGGACTTGAGTGGAATGATAACACAGTTCGAATCTCTGAAAATTTACAGAGATTCG
CTTGGGGAAGCAGTGATGAGAATGGGAGACCTCCATTCACTTCAAAGCAGGAACAAAAACTGGCGAGAAC
AATTGGGTCAGAAGTTTGAAGAAATAAGGTGGTTGATTGAGGAAGTGAGACACAAATTGAAGACAACAGA
AAACAGTTTTGAACAAATAACATTCATGCAAGCCTTACAACTGTTGCTTGAAGTGGAGCAAGAGATAAGG
ACTTTCTCGTTTCAGCTTATTTAATGATAAAAAACACCCTTGTTTCTACT
>CRG12092022_ H13N6_NS_8
AGCAAAAGCAGGGTGACAAAAACATAATGGATTCAAACACTGTATCAAGCTTTCAGGTAGATTGCTTTCT
TTGGCATGTCCGCAAACGATTTGCAGACCAAGAGATGGGTGATGCCCCATTCCTTGACCGGCTTCGCCGA
GATCAGAAGTCTTTAAAGGGGAGGAGCAGCACTCTTGGAATGGACATTGAGGCGGCTACATGCGCTGGGA
AGCGGATAATTGAGCGGATTCTAAAAGAAGAATCCAATGAGGCATTAAAGATGAATATTGCCTCAACGCC
TGCCTCACGCTACATGACTGACATGACTCCAGAGGAGATGTCAAAAGAATGGTTCATGCTCATGCCAAAA
CAGAAATTTGCTGGGCCTCTCCGCATCAGAATGGATCAGGCAATCATAGATAAAGAAATCACACTGAAAG
CGAATTTCAGTGTAATTTTTGACCGGTTGGAGACTCTCGTGCTGTTAAGGGCTTTTACCAATGATGGGGC
TATTGTTGGTGAAATCTCACCATTGCCTTCTCTTCCAGGACATACTAACGAGGATGTCAAAAATGCAATT
GGAGTCCTCATCGGTGGACTTGAGTGGAATGATAACACAGTTCGAATCTCTGAAAATTTACAGAGATTCG
CTTGGGGAAGCAGTGATGAGGATGGGAGACCTCCATTCACTTCAAAGCAGGAACAAAAACTGGCGAGAAC
AATTGGGTCAGAAGTTTGAAGAAATAAGGTGGTTGATTGAGGAAGTGAGACACAAATTGAAGACAACAGA
AAACAGTTTTGAACAAATAACATTCATGCAAGCCTTACAACTGTTGCTTGAAGTGGAGCAAGAGATAAGG
ACTTTCTCGTTTCAGCTTATTTAATGATAAAAAACACCCTTGTTTCTACT
>NDN12092022_ H13N6_NS_8
AGCAAAAGCAGGGTGACAAAAACATAATGGATTCAAACACTGTATCAAGCTTTCAGGTAGATTGCTTTCT

TTGGCATGTCCGCAAACGATTTGCAGACCAAGAGATGGGTGATGCCCCATTCCTTGACCGGCTTCGCCGA
GATCAGAAGTCTTTAAAGGGGAGGAGCAGCACTCTTGGAATGGACATTGAGGCGGCTACATGCGCTGGGA
AGCGGATAATTGAGCGGATTCTAAAAGAAGAATCCAATGAGGCATTAAAGATGAATATTGCCTCAACGCC
TGCCTCACGCTACATGACTGACATGACTCCAGAGGAGATGTCAAAAGAATGGTTCATGCTCATGCCAAAA
CAGAAATTTGCTGGGCCTCTCCGCATCAGAATGGATCAGGCAATCATAGATAAAGAAATCACACTGAAAG
CGAATTTCAGTGTAATTTTTGACCGGTTGGAGACTCTCGTGCTGTTAAGGGCTTTTACCAATGATGGGGC
TATTGTTGGTGAAATCTCACCATTGCCTTCTCTTCCAGGACATACTAACGAGGATGTCAAAAATGCAATT
GGAGTCCTCATCGGTGGACTTGAGTGGAATGATAACACAGTTCGAATCTCTGAAAATTTACAGAGATTCG
CTTGGGGAAGCAGTGATGAGAATGGGAGACCTCCATTCACTTCAAAGCAGGAACAAAAACTGGCGAGAAC
AATTGGGTCAGAAGTTTGAAGAAATAAGGTGGTTGATTGAGGAAGTGAGACACAAATTGAAGACAACAGA
AAACAGTTTTGAACAAATAACATTCATGCAAGCCTTACAACTGTTGCTTGAAGTGGAGCAAGAGATAAGG
ACTTTCTCGTTTCAGCTTATTTAATGATAAAAAACACCCTTGTTTCTACT
>BEL10102022_ H13N6_NS_8
AGCAAAAGCAGGGTGACAAAAACATAATGGATTCAAACACTGTGTCAAGCTTTCAGGTAGACTGCTTTCT
TTGGCATGTCCGCAAACGATTTGCAGACCAAGATATGGGTGATGCCCCATTCCTAGATCGGCTTCGCCGA
GACCAAAAGTCCCTAAAGGGGAGAAGCAGCACTCTTGGTATCGACATTGAGGCAGCTACTTGCTCTGGGA
AGCAGATAATAGAGCGGATTCTGGGTGAAGAATCAGACGAGGCACTAAAAATGACCATTGCCTCTGTGCC
TGCATCCCGCTATATAACTGACATGACCACTGAAGAAATGTCAAGGGACTGGTTTATGCTTATGCCCAAG
CAGAAGTTTGCTGGGCCTCTCTGCATCAAAATGGATCAGGCAATCATAGACAAAGACATCACACTGAAGG
CAAATTTCAGTATAATTTTTAACCGATTGGAAACTCTCGTGCTGCTGAGGGCTTTTACCAATGAGGGGGC
AATAGTGGGCGAAATTTCACAATTGCCTTCTCTTCCAGGACATACTAGCGAGGATGTCAAAAATGCAATT
GGAGTCCTCATCGGGGGACTTGAATGGAATGATAATACAGTTCGAATCTCTGAAAATCTACAGAGATTCG
CTTGGGGAAGCAGTAATGAGAATGGGAGACCTCCATTCACTCCAGAGCAGAAACAGAAACTGGCGAGAAC
AATTGAGTCAGAAGTTTGAAGAAATGAGATGGTTGATTGAAGAAGTGAGACACAAATTGAAGACAACAGA
GAGTAGTTTTGAACAAATAACATTCATGCAAGCATTACAACTATTGCTTGAAGTGGAGCAAGAGATAAGG
ACTTTCTCGTTTCAGCTTATTTAATGATAAAAAACACCCTTGTTTCTACT

Human H1N1 segment 4 and 6 sequences

>BYM28112022_ H1N1_HA_4
ATGAAGGCAATACTAGTAGTTATGCTGTATACATTTACAACCGCAAATGCAGACACATTATGTATAGGTT
ATCATGCGAACAATTCAACAGACACTGTGGACACAGTACTAGAAAAGAATGTAACAGTAACACACTCTGT
CAATCTTCTAGAAGACAAGCATAACGGAAAACTATGCAAACTAAGAGGGGTAGCCCCATTGCATTTGGGT
CAATGTAACATTGCTGGCTGGATCTTGGGAAATCCAGAGTGTGAATCACTCTCCACAGCAAGATCATGGT
CCTACATTGTGGAAACATCTAATTCAGACAATGGAACGTGTTACCCAGGAGATTTCATCAATTATGAGGA
GCTAAGAGAGCAATTGAGCTCAGTGTCATCATTTGAAAGGTTTGAAATATTCCCCAAGACAAGTTCATGG
CCTAATCATGACTCGGACAATGGTGTAACGGCAGCATGTTCTCACGCTGGAGCAAGAAGCTTCTACAAAA
ACTTGATATGGCTGGTTAAAAAAGGAAAATCGTACCCAAAGATCAACCAAACCTACATTAATGATAAAGG
GAAAGAAGTCCTCGTGCTGTGGGGCATTCACCATCCACCCACTATTACTGACCAAGAAAGTCTCTATCAG
AATGCAGATGCATATGTTTTTGTGGGGACATCAAGATACAGCAAGAAGTTCAAGCCGGAAATAGCAGCAA
GACCCAAAGTGAGGGNTCNAGCAGGGAGAATGAACTATTACTGGACACTAGTAGAACCGGGAGACAAAAT
AACATTCGAAGCAACTGGTAATCTAGTGGCACCGAGGTATGCATTCACAATGGAAAAAAAAGCTGGATCT
GGTATTATCATTTCAGATACACCAGTCCACGATTGCAATGCAACTTGTCAGACACCCGAGGGTGCTATAA
ACACCAGCCTCCCATTTCAAAATGTACATCCGATCACGATTGGGAAATGTCCAAAGTATGTAAGAAGCAC
AAAATTGAGACTGGCCACAGGATTGAGGAATGTCCCGTCTATTCAATCTAGAGGCCTATTCGGGGCCATT
GCTGGCTTCATCGAAGGGGGGTGGACAGGGATGGTAGATGGATGGTACGGTTATCACCATCAAAATGATC
AGGGATCAGGATATGCAGCCGATCTGAAGAGCACACAAAATGCCATTGATAAGATTACCAACAAAGTAAA
CTCTGTTATTGAAAAGATGAATACACAGTTCACAGCAGTTGGTAAAGAGTTCAACCACCTTGAAAAAAGA
ATAGAGAATCTAAATAAAAAGGTTGATGATGGTTTCCTGGACGTTTGGACTTACAATGCCGAACTGCTGG
TTCTACTGGAAAATGAAAGAACTTTGGACTATCACGATTCAAATGTGAAGAACTTGTATGAAAAAGTAAG
ACACCAATTAAAAAACAATGCCAAGGAAATTGGAAACGGCTGCTTTGAATTTTACCACAAATGCGACAAC
ACATGCATGGAAAGTGTCAAGAATGGGACTTATGACTACCCAAAATACTCAGAGGAAGCAAAATTAAACA
GAGAAAAAATAGATGGAGTAAAGCTGGACTCAACAAGGATTTACCAGATTTTGGCAATCTATTCAACTGT
TGCCAGTTCATTGGTACTGGTAGTCTCCCTGGGGGCAATCAGCTTCTGGATGTGCTCAAATGGGTCTCTA
CAGTGTAGAATATGTATTTAA
>BEL31102022_ H3N2_HA_4
AGCAAAAGCAGGGGATAATTCTATTAACCATGAAGGCTATCATTGCTTTGAGCAACATTCTATGTCTTGT
TTTCGCTCAAAAAATACCTGGAAATGACAATAGCACGGCAACGCTGTGCCTTGGGCACCATGCAGTACCA
AACGGAACGATAGTGAAAACAATCACAAATGACCGAATTGAAGTTACTAATGCTACTGAGTTGGTTCAGA
ATTCATCAATAGGTAAAATATGCAACAGTCCTCATCAGATCCTTGATGGAGGGAACTGCACACTAATAGA
TGCTCTATTGGGGGACCCTCAGTGTGACGGCTTTCAAAATAAGGAATGGGACCTTTTTGTTGAACGAAGC
AGAGCCAACAGCAGCTGTTACCCTTATGATGTGCCGGATTATGCCTCCCTTAGGTCACTAGTTGCCTCAT
CCGGAACACTGGAGTTTAAAAATGAAAGCTTCAATTGGACCGGAGTCAAACAAAACGGAACAAGTTCTGC
GTGCAAAAGGGGATCTAGTAGTAGTTTTTTTAGTAGATTAAATTGGTTGACCAGCTTAAACAACATATAT
CCAGCACAGAACGTGACTATGCCAAACAAGGAACAATTTGACAAATTGTACATTTGGGGGGTTCACCACC
CGGATACGGACAAGAACCAATTCTCCCTGTTTGCTCAATCATCAGGAAGAATCACAGTATCTACCAAAAG
AAGCCAACAAGCTGTAATCCCAAATATTGGATCTAGACCCAGAGTAAGGGATATCCCTAGCAGAATAAGC
ATCTATTGGACAATAGTAAAACCGGGAGACATACTTTTGATTAACAGCACAGGGAATCTAATTGCTCCTA
GGGGTTACTTCAAAATACGAAGTGGGAGAAGCTCAATAATGAGATCAGACGCACCCATTGGCAAATGTAA
GTCTGAATGCATCACTCCAAATGGAAGCATTCCCAATGACAAACCGTTCCAAAATGTAAACAGGATCACA
TACGGGGCCTGTCCCAGATACGTTAAGCAAAGCACCCTGAAATTGGCAACAGGAATGCGAAATGTACCTG
AGAAACAAACCAGAGGCATATTTGGTGCAATAGCGGGTTTCATAGAAAATGGATGGGAGGGAATGGTGGA
TGGTTGGTACGGTTTCAGGCACCAAAATTCTGAGGGAAGAGGACAAGCAGCAGATCTCAAAAGCACTCAA

GCAGCAATCGATCAAATCAGTGGGAAGCTGAATCGATTGATCGGAAAAACCAACGAGAAATTCCATCAGA
TTGAAAAAGAATTCTCAGAAGTAGAAGGAAGAGTTCAAGACCTCGAGAAATATGTTGAGGACACTAAAAT
AGATCTCTGGTCATACAACGCGGAGCTTCTTGTTGCCCTGGAGAACCAACATACGATTGACCTAACTGAC
TCAGAAATGAACAAGCTGTTTGAAAAAACAAAGAAGCAACTGAGGGAAAATGCTGAGGATATGGGAAATG
GTTGTTTCAAAATATACCACAAATGTGACAATGCCTGCATAGGATCAATAAGAAATGAAACTTATGACCA
CAATGTGTACAGGGATGAAGCATTAAACAACCGGTTCCAGATCAAGGGAGTTGAGCTGAAGTCAGGGTAC
AAAGATTGGATCCTATGGATTTCCTTTGCCATGTCATGTTTTTTGCTTTGTATTGCTTTGTTGGGGTTCA
TCATGTGGGCCTGCCAAAAGGGCAACATTAGATGCAACATTTGCATTTGAGTGCATTAATTAAAAACACC
CTTGTTTCTACT
>BYM28112022_ H3N2_HA_4
AGCAAAAGCAGGGGATAATTCTATTAACCATGAAGACTATCATTGCTTTGAGCAACATTCTATGTCTTGT
TTTCGCTCAAAAAATACCTGGAAATGACAATAGTACGGCAACGCTGTGCCTTGGGCACCATGCAGTACCA
AACGGAACGATAGTGAAAACAATCACAAATGACCGAATTGAAGTTACTAATGCTACTGAGTTGGTTCAGA
ATTCATCAATAGGTGAAATATGCGGCAGTCCTCATCAGATCCTTGATGGAGGGAACTGCACACTAATAGA
TGCTCTATTGGGGGACCCTCAGTGTGACGGCTTTCAAAATAAGGAATGGGACCTTTTTGTTGAAAGAAGC
AGAGCCAACAGCAACTGTTACCCTTATGATGTGCCGGGTTATGCCTCCCTTAGGTCACTAGTTGCCTCAT
CCGGCACACTGGAATTTAAAAATGAAAGCTTCAATTGGACCGGAGTCAAACAAAACGGAACAAGTTCTGC
GTGCAAAAGGGGATCTAGTAGTAGTTTTTTTAGTAGATTAAATTGGTTGACCAGCTTAAACAACATATAT
CCAGCACAGAACGTGACTATGCCAAACAAGGAACAATTTGACAAATTGTACATTTGGGGGGTTCACCACC
CGGATACAGACAAAAACCAAATCTCCCTGTTTGCTCAATCATCAGGAAGAATCACAGTATCTACCAAAAG
AAGCCAACAAGCTGTAATCCCAAATATCGGATCTAGACCCAGAATAAGGGATATCCCTAGCAGAATAAGC
ATCTATTGGACAATAGTAAAACCGGGAGACATACTTTTGATTAACAGCACAGGGAATCTAATTGCTCCTA
GGGGTTACTTCAAAATACGAAGTGGGAAAAGCTCAATAATGAGATCAGATGCACCCATTGGCAGATGTAA
GTCTGAATGCATCACTCCAAATGGAAGCATTCCCAATGACAAACCGTTCCAAAATGTAAACAGGATCACA
TACGGGGCCTGTCCCAGATATGTTAAGCAAAGCACCTTGAAATTGGCAACAGGAATGCGAAATGTACCAG
AGAAACAAACCAGAGGCATATTTGGTGCAATAGCGGGTTTCATAGAAAATGGATGGGAGGGAATGGTGGA
TGGTTGGTACGGTTTCAGGCATCAAAATTCTGAGGGAAGAGGACAAGCAGCAGATCTCAAAAGCACTCAA
GCAGCAATCGATCAAATCAATGGGAAGCTGAATCGATTGATCGGAAAAACCAACGAGAAATTCCATCAGA
TTGAAAAAGAATTCTCAGAAGTAGAAGGAAGAGTTCAAGACCTTGAGAAATATGTTGAGGACACTAAAAT
AGATCTCTGGTCATACAACGCTGAGCTTCTTGTTGCCCTGGAGAACCAACATACGATTGACCTAACTGAC
TCAGAAATGAACAAACTGTTTGAAAAAACAAAGAAGCAACTGAGGGAAAATGCTGAGGATATGGGAAATG
GTTGTTTCAAAATATACCACAAATGTGACAATGCCTGCATAGGATCAATAAGAAATGAAACTTATGACCA
CAATGTGTACAGGGATGAAGCATTAAACAACCGGTTCCAGATCAAGGGAGTTGAGCTGAAATCAGGGTAC
AAAGATTGGATCCTATGGATTTCCTTTGCCATGTCATGTTTTTTGCTTTGTATTGCTTTGTTGGGGTTCA
TCATGTGGGCCTGCCAAAAGGGCAACATTAGATGCAACATTTGCATTTGAGTGCATTAATTAAAAACACC
CTTGTTTCTACT
>BEL28112022_ H3N2_HA_4
AGCAAAAGCAGGGGATAATTCTATTAACCATGAAGACTATCATTGCTTTGAGCAACATTCTATGTCTTGT
TTTCGCTCAAAAAATACCTGGAAACGACAATAGCACGGCAACGCTGTGCCTTGGGTACCATGCAGTACCA
AACGGAACGATAGTGAAAACAATCACAAATGACCGAATTGAAGTTACTAATGCTACTGAGTTGGTTCAGA
ATTCATCAATAGGTAAAATATGCGACAGTCCTCATCAGATCCTTGATGGAGGGAACTGCACACTAATAGA
TGCTCTATTGGGGGACCCTCAGTGTGACGGCGTTCAAAATAAGGAATGGGACCTTTTTGTTGAACGAAGC
AGAGCCAACAGCAACTGTTACCCTTATGATGTGCCGGATTATGCCTCCCTTAGGTCACTAGTTGCCTCAT
CCGGCACACTGGAGTTTAAAAATGAAAGCTTCAATTGGACTGGAGTCAAACAAAACGGAGCAAGTTCTGC
GTGCAAAAGGGGATCTAGTAGTAGTTTTTTTAGTAGATTAAATTGGTTGACCCACTTAAACAACATATAT
CCAGCACAGAACGTGACTATGCCAAACAAGGAACAATTTGACAAATTGTACATTTGGGGGGTTCACCACC

CGGATACGGACAAGAACCAAATCTCCCTGTTCGCCCAATCATCAGGAAGAATCACAGTATCTACCAAAAG
AAGCCAACAAGCTGTAATCCCAAATATCGGATCTAGACCCAGAATAAGGGACATTCCTAGCAGAATAAGC
ATCTATTGGACAATAGTAAAACCGGGAGACATACTTTTGATTAACAGCACAGGGAATCTAATTGCTCCTA
GGGGTTACTTCAAAATACGAAATGGGAAAAGCTCAATAATGAGATCAGATGCACCCATTGGCAAATGCAA
GTCTGAATGCATCACTCCAAATGGAAGCATTCCCAATGACAAACCGTTCCAAAATGTAAACAGGATCACA
TACGGGGCCTGTCCCAGATATGTTAAGCAAAGCACCCTGAAATTGGCAACAGGAATGCGAAATGTACCAG
AGAAACAAACCAGAGGCATATTTGGCGCAATAGCGGGTTTCATAGAAAATGGATGGGAGGGAATGGTGGA
TGGTTGGTACGGTTTCAGGCATCAAAATTCTGAGGGAAGAGGACAAGCAGCAGATCTCAAAAGCACTCAA
GCAGCAATCGATCAAATCAATGGGAAGCTGAATCGATTGATCGGAAAAACCAACGAGAAATTCCATCAGA
TTGAAAAAGAATTCTCAGAGGTAGAAGGAAGAGTTCAAGACCTTGAGAAATATGTTGAGGACACTAAAAT
AGATCTCTGGTCATACAACGCGGAGCTTCTTGTTGCCCTGGAGAACCAACATACGATTGACCTAACTGAC
TCAGAAATGAACAAACTGTTTGAAAAAACAAAGAAGCAACTGAGGGAAAATGCTGAGGATATGGGAAATG
GTTGTTTCAAAATATACCACAAATGTGACAATGCCTGCATAGGATCAATAAGAAATGAAACTTATGACCA
CAATGTGTACAGGGATGAAGCATTAAACAACCGGTTCCAGATCAAGGGAGTTGAGCTGAAGTCAGGGTAC
AAAGATTGGATCCTATGGATTTCCTTTGCCATGTCATGTTTTTTGCTTTGTATTGCTTTGTTGGGGTTCA
TCATGTGGGCCTGCCAAAAGGGCAACATTAGATGCAACATTTGCATTTGAGTGCATTAATTAAAAACACC
CTTGTTTCTACT >BEL10102022_ H1N1_NA_6
ATGAATCCAAACCAAAAGATAATAACCATTGGTTCTGTTTGTATGACAATTGGAACGGCTAACTTAATAT
TACAAATTGGAAACATAATCTCAATATGGGTTAGCCACTCAATTCAAATTGGAAATCAAAGCCAGATTGA
AACATGCAATAAAAGCGTCATTACTTATGAAAACAACACTTGGGTAAATCAGACATTTGTTAACATCAGC
AACACTAACTCTGCTGCTAGACAGTCAGTGGCTTCCGTGAAATTAGCGGGCAATTCCTCTCTCTGCCCTG
TTAGTGGATGGGCTATATACAGTAAAGACAACAGTGTAAGAATCGGTTCCAAGGGGGATGTGTTTGTCAT
AAGGGAACCATTCATATCATGCTCCCCCTTGGAATGCAGAACCTTCTTCTTGACTCAAGGGGCTTTGCTA
AATGACAAACATTCCAATGGAACAATCAAAGACAGAAGCCCATATCGAACCCTAATGAGCTGTCCTATTG
GTGAAGTTCCCTCTCCATACAACTCAAGATTTGAGTCAGTCGCTTGGTCAGCAAGTGCTTGTCATGATGG
CACCAATTGGCTAACAATTGGAATTTCTGGCCCAGACAGTGGGGCAGTGGCTGTGTTAAAATACAATGGC
ATAATAACAGACACTATCAAGAGTTGGAGGAACAAGATATTGAGAACACAAGAGTCTGAATGTGCATGTG
TAAATGGTTCTTGCTTTACCATAATGACTGATGGACCAAGTGATGGACAGGCCTCATACAAAATCTTCAG
AATAGAGAAGGGAAAGATAATCAAATCAGTCGAAATGAAGGCCCCTAATTATCACTATGAAGAATGCTCC
TGTTACCCTGATTCTAGTGAAATCACATGTGTGTGCAGGGATAATTGGCATGGCTCGAATCGACCTTGGG
TGTCTTTCAACCAGAATCTGGAATATCAGATGGGATACATATGCAGTGGGGTTTTCGGAGACAATCCACG
CCCTAATGATAAGACAGGCAGTTGTGGTCCAGTATTGTCTAATGGAGCAAATGGGGTAAAAGGATTTTCA
TTCAAATACGGCAATGGTGTTTGGATAGGGAGAACTAAGAGCATTAGTTCAAGAAAAGGTTTTGAGATGA
TTTGGGATCCGAATGGATGGACTGGGACTGACAATAAATTCTCAAAAAAGCAAGATATTGTAGGGATAAA
TGAGTGGTCAGGGTATAGCGGGAGTTTTGTTCAGCATCCAGAACTAACAGGGCTGAATTGTATAAGACCT
TGCTTCTGGGTTGAACTAATAAGAGGACGACCCGAAGAGAACACGATCTGGACTAGCGGGAGCAGCATAT
CCTTTTGTGGTGTAGACAGTGACATTATGGGTTGGTCTTGGCCAGACGGTGCTGAGTTGCCATTCACCAT
TGACAATTAA >BEL24102022_ H1N1_NA_6
ATGAATCCAAACCAAAAGATAATAACCATTGGTTCTATTTGTATGACAATTGGAACGGCTAACTTAATAT
TACAAATTGGAAACATAATCTCAATATGGGTTAGCCACTCAATTCAAATTGGAAATCAAAGCCAGATTGA
AACATGCGATAAAAGCGTCATTACTTATGAAAACAACACTTGGGTAAATCAGACATTTGTTAACATCAGC
AACACTAACTCTGCTGCTAGACAGTCAGTGGCTTCCGTGAAATTAGCGGGCAATTCCTCTCTCTGCCCTG
TTAGTGGATGGGCTATATACAGTAAAGACAACAGTGTAAGAATCGGTTCCAAGGGGGATGTGTTTGTCAT
AAGGGAACCATTCATATCATGCTCTCCCTTGGAATGCAGAACCTTCTTCTTGACTCAAGGGGCTTTGCTA
AATGACAAACATTCCAATGGAACAATTAAAGACAGAAGCCCATATCGAACCCTAATGAGCTGTCCTATTG
GTGAAGTTCCCTCTCCATACAACTCAAGATTTGAGTCAGTCGCTTGGTCAGCAAGTGCTTGTCATGATGG

CACCAATTGGCTAACAATTGGAATTTCTGGCCCAGACAGTGGGGCAGTGGCTGTGTTAAAATACAATGGC
ATAATAACAGACACTATCAAGAGTTGGAGGAACAAGATATTGAGAACACAAGAGTCTGAATGTGCATGTG
TAAATGGTTCTTGCTTTACCATAATGACCGATGGACCAAGTGATGGACAGGCCTCATACAAAATCTTCAG
AATAGAGAAGGGAAAGATAATCAAATCAGTCGAAATGAAGGCCCCTAATTATCACTATGAAGAATGCTCC
TGTTACCCTGATTCTAGTGAAATCACATGTGTGTGCAGGGATAATTGGCATGGCTCGAATCGACCTTGGG
TGTCTTTCAACCAGAATCTGGAATATCAGATGGGATACATATGCAGTGGGGTTTTCGGAGACAATCCACG
CCCTAATGATAAGACAGGCAGTTGTGGTCCAGTATCGTCTAATGGAGCAAATGGGGTAAAAGGATTTTCA
TTCAAATACGGCAATGGTGTTTGGATAGGGAGAACTAAGAGCATTAGTTCAAGAAAAGGTTTTGAGATGA
TTTGGGATCCGAATGGATGGACTGGGACTGACAATAAATTCTCAAAAAAGCAAGATATTGTAGGGATAAA
TGAGTGGTCAGGGTATAGCGGGAGTTTTGTTCAGCATCCAGAACTAACAGGGCTGAATTGTATAAGACCT
TGCTTCTGGGTTGAACTAATAAGAGGACGACCCGGAGAGAACACGATCTGGACTAGCGGGAGCAGCATAT
CCTTTTGTGGTGTAGACAGTGACATTATGGGTTGGTCTTGGCCAGACGGTGCTGAGTTGCCATTCAACAT
TGACAATTAA >BEL31102022_ H1N1_NA_6
ATGAATCCAAACCAAAAGATAATAACCATTGGTTCTGTTTGTATGACAATTGGAACGGCTAACTTAATAT
TACAAATTGGAAACATAATCTCAATATGGGTTAGCCACTCAATTCAAATTGGAAATCAAAGCCAGATTGA
AACATGCAATAAAAGCGTCATTACTTATGAAAACAACACTTGGGTAAATCAGACATTTGTTAACATCAGC
AACACTAACTCTGCTGCTAGACAGTCAGTGGCTTCCGTGAAATTAGCGGGCAATTCTTCTCTCTGCCCTG
TTAGTGGATGGGCTATATACAGTAAAGACAACAGTGTAAGAATCGGTTCCAAGGGGGATGTGTTTGTCAT
AAGGGAACCATTCATATCATGCTCCCCCTTGGAATGCAGAACCTTCTTCTTGACTCAAGGGGCTTTGCTA
AATGACAAACATTCCAATGGAACAATTAAAGACAGAAGCCCATATCGAACCCTAATGAGCTGTCCTATTG
GTGAAGTTCCCTCTCCATACAACTCAAGATTTGAGTCAGTCGCTTGGTCAGCAAGTGCTTGTCATGATGG
CACCAATTGGCTAACAATTGGAATTTCTGGCCCAGACAGTGGGGCAGTGGCTGTGTTAAAATACAATGGC
ATAATAACAGACACTATCAAGAGTTGGAGGAACAAGATATTGAGAACACAAGAGTCTGAATGTGCATGTG
TAAATGGTTCTTGCTTTACCATAATGACCGATGGACCAAGTGATGGACAGGCCTCATACAAAATCTTCAG
AATAGAGAAGGGAAAGATAATCAAATCAGTCGAAATGAAGGCCCCTAATTATCACTATGAAGAATGCTCC
TGTTACCCTGATTCTAGTGAAATCACATGTGTGTGCAGGGATAATTGGCATGGCTCGAATCGACCTTGGG
TGTCTTTCAATCAGAATCTGGAATATCAGATGGGATACATATGCAGTGGGGTTTTCGGAGACAATCCACG
CCCTAATGATAAGACAGGCAGTTGTGGTCCAGTATTGTCTAATGGAGCAAATGGGGTAAAAGGATTTTCA
TTCAAATACGGCAATGGTGTTTGGATAGGGAGAACTAAAAGCATTAGTTCAAGAAAAGGTTTTGAGATGA
TTTGGGATCCGAATGGATGGACTGGGACTGACAATAAATTCTCAAAAAAGCAAGATATTGTAGGGATAAA
TGAGTGGTCAGGGTATAGCGGGAGTTTTGTTCAGCATCCAGAACTAACAGGGCTGAATTGTATAAGACCT
TGCTTCTGGGTTGAACTAATAAGAGGACGACCCGAAGAGAACACGATCTGGACTAGCGGGAGCAGCATAT
CCTTTTGTGGTGTAGACAGTGACATTATGGGTTGGTCTTGGCCAGACGGTGCTGAGTTGCCATTCACCAT
TGACAATTAA >CRG12092022_ H1N1_NA_6
ATGAATCCAAACCAAAAGATAATAACCATTGGTTCTGTTTGTATGACAATTGGAACGGCTAACTTAATAT
TACAAATTGGAAACATAATCTCAATATGGGTTAGCCACTCAATTCAAATTGGAAATCAAAGCCAGATTGA
AACATGCAATAAAAGCGTCATTACTTATGAAAACAACACTTGGGTAAATCAGACATTTGTTAACATCAGC
AACACTAACTCTGCTGCTAGACAGTCAGTGGCTTCCGTGAAATTAGCGGGCAATTCTTCTCTCTGCCCTG
TTAGTGGATGGGCTATATACAGTAAAGACAACAGTGTAAGAATCGGTTCCAAGGGGGATGTGTTTGTCAT
AAGGGAACCATTCATATCATGCTCCTCCTTGGAATGCAGAACCTTCTTCTTGACTCAAGGGGCTTTGCTA
AATGACAAACATTCCAATGGAACAGTCAAAGACAGAAGCCCATATCGAACCCTAATGAGCTGTCCTATTG
GTGAAGTTCCCTCTCCATACAACTCAAGATTTGAGTCAGTCGCTTGGTCAGCAAGTGCTTGTCATGATGG
CACCAATTGGCTAACAATTGGAATTTCTGGCCCAGACAGTGGGGCAGTGGCTGTGTTAAAATACAATGGC
ATAATAACAGACACTATCAAGAGTTGGAGGAACAAGATATTGAGAACACAAGAGTCTGAATGTGCATGTG
TAAATGGTTCTTGCTTTACCATAATGACTGATGGACCAAGTGATGGACAGGCCTCATACAAAATCTTCAG
AATAGAGAAGGGAAAGATAATCAAATCAGTCGAAATGAAGGCCCCTAATTATCACTATGAAGAATGCTCC

TGTTACCCTGATTCTAGTGAAATCACATGTGTGTGCAGGGATAATTGGCATGGCTCGAATCGACCTTGGG
TGTCTTTCAACCAGAATCTGGAATATCAGATGGGATACATATGCAGTGGGGTTTTCGGAGACAATCCACG
CCCTAATGATAAGACAGGCAGTTGTGGTCCAGTATTGTCTAATGGAGCAAATGGGGTAAAAGGATTTTCA
TTCAAATACGGCAATGGTGTTTGGATAGGGAGAACTAAGAGCATTAGTTCAAGAAAAGGTTTTGAGATGA
TTTGGGATCCGAATGGATGGACTGGGACTGACAATAAATTCTCAAAAAAGCAAGATATTGTAGGGATAAA
TGAGTGGTCAGGGTATAGCGGGAGTTTTGTTCAGCATCCAGAACTAACAGGGCTGAATTGTATAAGACCT
TGCTTCTGGGTTGAACTAATAAGAGGACGACCCGAAGAGAACACGATCTGGACTAGCGGGAGCAGCATAT
CCTTTTGTGGTGTAGACAGTGACATTATGGGTTGGTCTTGGCCAGACGGTGCTGAGTTGCCATTCACCAT
TGACAATTAA >BYM28112022_ H1N1_NA_6
ATGAATCCAAACCAAAAGATAATAACCATTGGTTCTGTTTGTATGACAATTGGAACGGCTAACTTAATAT
TACAAATTGGAAACATAATCTCAATATGGGTTAGCCACTCAATTCAAATTGGAAATCAAAGCCAGATTGA
AACATGCAATAAAAGCGTCATTACTTATGAAAACAACACTTGGGTAAATCAGACATTTGTTAACATCAGC
AACACTAACTCTGCTGCTAGACAGTCAGTGGCTTCCGTGAAATTAGCGGGCAATTCTTCTCTCTGCCCTG
TTAGTGGATGGGCTATATACAGTAAAGACAACAGTGTAAGAATCGGTTCCAAGGGGGATGTGTTTGTCAT
AAGGGAACCATTCATATCATGCTCTCCCTTGGAATGCAGAACCTTCTTCTTGACTCAAGGGGCTTTGCTA
AATGACAAACATTCCAATGGAACAATTAAAGACAGAAGCCCATATCGAACCCTAATGAGCTGTCCTATTG
GTGAAGTTCCCTCTCCATACAACTCAAGATTTGAGTCAGTCGCTTGGTCAGCAAGTGCTTGTCATGATGG
CACCAATTGGCTAACAATTGGAATTTCTGGCCCAGACAGTGGGGCAGTGGCTGTGTTAAAATACAATGGC
ATAATAACAGACACTATCAAGAGTTGGAGGAACAAGATATTGAGAACACAAGAGTCTGAATGTGCATGTG
TAAATGGTTCTTGCTTTACCATAATGACCGATGGACCAAGTGATGGACAGGCCTCATACAAAATCTTCAG
AATAGAGAAGGGAAAGATAATCAAATCAGTCGAAATGAAGGCCCCTAATTATCACTATGAAGAATGCTCC
TGTTACCCTGATTCTAGTGAAATCACATGTGTGTGCAGGGATAATTGGCATGGCTCGAATCGACCTTGGG
TGTCTTTCAACCAGGATCTGGAATATCAGATGGGATACATATGCAGTGGGGTTTTCGGAGACAATCCACG
CCCTAATGATAAGACAGGCAGTTGTGGTCCAGTATTGTCTAATGGAGCAAATGGGGTAAAAGGATTTTCA
TTCAAATACGGCAATGGTGTTTGGATAGGGAGAACTAAGAGCATTAGTTCAAGAAAAGGTTTTGAGATGA
TTTGGGATCCGAATGGATGGACTGGGACTGACAATAAATTCTCAAAAAAACAAGATATTGTAGGGATAAA
TGAGTGGTCAGGGTATAGCGGGAGTTTTGTTCAGCATCCAGAACTAACAGGGCTGAATTGTATAAGACCT
TGCTTCTGGGTTGAACTAATAAGAGGACGACCCGAAGAGAACACGATCTGGACTAGCGGGAGCAGCATAT
CCTTTTGTGGTGTAGACAGTGACATTATGGGTTGGTCTTGGCCAGACGGTGCTGAGTTGCCATTCACCAT
TGACAATTAA >NDN12092022_ H3N2_NA_6
AGCAAAAGCAGGAGTAAAGATGAATCCAAATCAAAAGATAATAACGATTGGCTCTGTTTCTCTCACAATT
TCCACAATATGCTTCTTCATGCAAATTGCCATCCTGATAACTACTGTAACATTGCATTTCAAGCAATATG
AATTCAACTCCCCCCCAAATAACCAAGTGATGCTGTGTGAACCAACAATAATAGAAAGAAACATAACAGA
GATAGTGTATTTGACCAACACCACCATAGAGAAGGAAATATGCCCCAAACCAGCAGAATACAGAAATTGG
TCAAAACCGCAATGTGGCATTACAGGATTTGCACCTTTCTCTAAGGACAATTCGATTAGGCTTTCCGCTG
GTGGGGACATATGGGTGACAAGAGAACCGTATGTGTCATGCGATCTTGACAAGTGTTATCAATTTGCCCT
TGGACAGGGAACAACACTAAACAATGTGCATTCAAATAACACAGTACATGATAGAACCCCTTATCGGACT
CTATTGATGAATGAGTTGGGTGTTCCTTTCCATCTGGGGACCAAGCAAGTGTGCATAGCATGGTCCAGCT
CAAGTTGTCACGATGGAAAAGCATGGCTGCATGTTTGTATAACGGGGGATGATAAAAATGCAACTGCTAG
CTTCATTTACAATGGGAGGCTTGTAGATAGTGTTGTTTCATGGTCCAACGATATTCTCAGAACCCAGGAG
TCAGAATGCGTTTGTATCAATGGAACTTGTACAGTAGTAATGACTGATGGAAATGCTACAGGAAAAGCTG
ATACTAAAATACTATTCATTGAGGAGGGGAAAATCGTTCATACTAGCAAATTGTCAGGAAGTGCTCAGCA
TGTCGAAGAGTGCTCTTGCTATCCTCGATATCCTGGTGTCAGATGTGTCTGCAGAGACAACTGGAAAGGA
TCCAACCGGCCCATCATAGATATAAACATAAAGGATCATAGCATTGTTTCCAGGTATGTGTGTTCTGGAC
TTGTTGGAGACACACCCAGAAAAAGCGACAGCTCCAGCAGTAGCCACTGTTTGAACCCTAACAATGAAAA
AGGTGATCATGGAGTGAAAGGCTGGGCCTTTGATGATGGAAATGACGTGTGGATGGGGAGAACAATCA

>BYM28112022_ H3N2_NA_6
AGCAAAAGCAGGAGTAAAGATGAATCCAAATCAAAAGATAATAACGATTGGCTCTGTTTCTCTCACAATT
TCCACAATATGCTTCTTCATGCAAATTGCCATCCTGATAACTACTGTAACATTGCATTTCAAGCAATATG
AATTCAACTCCCCCCCAAATAACCAAGTGATGCTGTGTGAACCAACAATAATAGAAAGAAACATAACAGA
GATAGTGTATTTGACCAACACCACCATAGAGAAGGAAATATGCCCCAAACCAGCAGAATACAGAAATTGG
TCAAAACCGCAATGTGGCATTACAGGATTTGCACCTTTCTCTAAGGACAATTCGATTAGGCTTTCCGCTG
GTGGGGACATCTGGGTGACAAGAGAACCTTATGTGTCATGCGATCTTGACAAGTGTTATCAATTTGCCCT
TGGACAGGGAACAACACTAAACAATGTGCATTCAAATAACACAGTACGTGATAGAACCCCTTATCGGACT
CTATTGATGAATGAGTTGGGTGTTCCTTTCCATCTGGGGACCAAGCAAGTGTGCATAGCATGGTCCAGCT
CAAGTTGTCACGATGGAAAAGCATGGCTGCATGTTTGTATAACGGGGGATGATAAAAATGCAACTGCTAG
CTTCATTTACAATGGGAGGCTTGTAGATAGTGTTGTTTCATGGTCCAACGATATTCTCAGAACCCAGGAG
TCAGAATGCGTTTGTATCAATGGAACTTGTGCAGTAGTAATGACTGATGGAAATGCTACAGGAAAAGCTG
ATACTAAAATACTATTCATTGAGGAGGGGAAAATCGTTCATACTAGCAAATTGTCAGGAAGTGCTCAGCA
TGTCGAAGAGTGCTCTTGCTATCCTCGATATCCTGGTGTCAGATGTGTCTGCAGAGACAACTGGAAAGGA
TCCAACCGGCCCATCATAGATATAAACATAAAGGATCATAGCATTGTTTCCAGGTATGTGTGTTCTGGAC
TTGTTGGAGACACACCCAGAAAAAGCGACAGCTCCAGCAGTAGCCATTGTTTGAACCCTAACAATGAAAA
AGGTGGTCATGGAGTGAAAGGCTGGGCCTTTGATGATGGAAATGACGTGTGGATGGGGAGAACAATCAAC
GAGACGTCACGCTTAGGGTATGAAACCTTCAAAGTCGTTGAAGGCTGGTCCAACCCTAAGTCCAAATTGC
AGATAAATAGGCAAGTCATAGTTGACAGAGGCGATAGGTCCGGTTATTCTGGTATTTTCTCTGTTGAAGG
CAAAAGCTGCATCAATCGGTGCTTTTATGTGGAGTTGATTAGGGGAAGAAAAGAGGAAACTGAAGTCTTG
TGGACTTCAAACAGTATTGTTGTGTTTTGTGGCACCTCAGGTACATATGGAACAGGCTCATGGCCTGATG
GGGCGAACCTCAGTCTCATGCATATATAAGCTTTTGCAATTTTAGAAAAAACC
